# Therapeutic Anticoagulation in Critically Ill Patients with Covid-19 – Preliminary Report

**DOI:** 10.1101/2021.03.10.21252749

**Authors:** The REMAP-CAP, ACTIV-4a, ATTACC Investigators, Ryan Zarychanski

## Abstract

**Background:** Thrombosis may contribute to morbidity and mortality in Covid-19. We hypothesized that therapeutic anticoagulation would improve outcomes in critically ill patients with Covid-19.

**Methods:** We conducted an open-label, adaptive, multiplatform, randomized, clinical trial. Patients with severe Covid-19, defined as the requirement for organ support with high flow nasal cannula, non-invasive ventilation, invasive ventilation, vasopressors, or inotropes, were randomized to receive therapeutic anticoagulation with heparin or pharmacological thromboprophylaxis as per local usual care. The primary outcome was an ordinal scale combining in-hospital mortality (assigned –1) and days free of organ support to day 21.

**Results:** Therapeutic anticoagulation met the pre-defined criteria for futility in patients with severe Covid-19. The primary outcome was available for 1,074 participants (529 randomized to therapeutic anticoagulation and 545 randomized to usual care pharmacological thromboprophylaxis). Median organ support-free days were 3 days (interquartile range –1, 16) in patients assigned to therapeutic anticoagulation and 5 days (interquartile range –1, 16) in patients assigned to usual care pharmacological thromboprophylaxis (adjusted odds ratio 0.87, 95% credible interval (CrI) 0.70-1.08, posterior probability of futility [odds ratio<1.2] 99.8%). Hospital survival was comparable between groups (64.3% vs. 65.3%, adjusted odds ratio 0.88, 95% CrI 0.67-1.16). Major bleeding occurred in 3.1% of patients assigned to therapeutic anticoagulation and 2.4% of patients assigned to usual care pharmacological thromboprophylaxis.

**Conclusions:** In patients with severe Covid-19, therapeutic anticoagulation did not improve hospital survival or days free of organ support compared to usual care pharmacological thromboprophylaxis.

Trial registration numbers NCT02735707, NCT04505774, NCT04359277, NCT04372589

## Background

Observational studies identified an association between the novel coronavirus disease 2019 (Covid-19), inflammation, hypercoagulability, and thrombosis.^1-4^ Critically ill patients with Covid-19 are at high risk of venous and arterial thrombotic events despite standard dose pharmacological thromboprophylaxis.^5-8^ Higher levels of circulating biomarkers reflecting systemic inflammation and coagulation activation (e.g., D-dimer, C-reactive protein) are independently associated with a greater risk of respiratory failure, thrombosis, and death.^2,9,10^ Thrombotic processes may therefore be an important cause of poor outcome from Covid-19.

Unfractionated and low molecular weight heparins are parenteral anticoagulants with anti-inflammatory properties and possible antiviral properties.^11,12^ On the basis of clinical and pathologic reports of excess thrombotic risk enhanced dose anticoagulation strategies have been incorporated into some Covid-19 guidance statements, especially for critically ill patients.^13,14^ However, the effectiveness and safety of empiric full dose anticoagulation to improve outcomes in Covid-19 has not been established.

To determine whether a pragmatic strategy of therapeutic dose anticoagulation improves survival and reduces the duration of organ support compared to usual care pharmacological thromboprophylaxis in critically ill patients with Covid-19, we conducted an international multiplatform randomized clinical trial (mpRCT).

## Methods

### Trial Design and Oversight

Early in the Covid-19 pandemic, the lead investigators of three international adaptive platform trials harmonized their protocols to study the effect of therapeutic anticoagulation in patients hospitalized for Covid-19 into one integrated mpRCT to accelerate evidence generation and maximize external validity of results (see Protocol Appendix, p. 29). The participating platforms included Randomized, Embedded, Multifactorial Adaptive Platform Trial for Community-Acquired Pneumonia (REMAP-CAP; NCT02735707)^15^, Accelerating Covid-19 Therapeutic Interventions and Vaccines-4 Antithrombotics Inpatient platform trial (ACTIV-4a; NCT04505774 and NCT04359277), and Antithrombotic Therapy to Ameliorate Complications of Covid-19 (ATTACC; NCT04372589).^16^ The three platforms aligned the trial design, eligibility criteria, interventions, outcome measures, and statistical analysis plan *a priori* to execute the mpRCT. Each platform was overseen by independent data and safety monitoring boards (DSMB) and collaboratively guided a cross-platform DSMB interaction plan (see Protocol Appendix, p. 155).

The platforms enrolled patients hospitalized for Covid-19. Although REMAP-CAP enrolled patients with suspected or confirmed Covid-19, only participants with confirmed infection were included in the mpRCT primary analysis. The mpRCT was designed to evaluate the effect of therapeutic anticoagulation in four patient groups: severe Covid-19, or moderate Covid-19 stratified by degree of D-dimer elevation (high, low, or missing). The stopping criteria for a statistical conclusion applied independently to each of the groups except for the group with missing D-dimer.

Severe Covid-19 was defined as the provision of intensive care unit-level respiratory or cardiovascular organ support (high flow nasal oxygen ≥ 20 L/min, non-invasive or invasive mechanical ventilation, extracorporeal life support, vasopressors, or inotropes). Patients were ineligible if they were admitted to the ICU with Covid-19 for more than 48 hours (REMAP-CAP) or to hospital for more than 72 hours (ACTIV-4a, ATTACC) prior to randomization, at imminent risk of death without an ongoing commitment to full organ support, at high risk of bleeding, receiving dual antiplatelet therapy, had a separate clinical indication for therapeutic anticoagulation, or had a history of heparin sensitivity including heparin-induced thrombocytopenia. Detailed exclusion criteria for the platforms are provided in the Protocol Appendix (p. 29).

The mpRCT was conducted in accordance with the principles of the Good Clinical Practice guidelines of the International Conference on Harmonization. Ethics and regulatory approval were obtained at each participating center. The trial was supported by multiple international funding organizations who had no role in the analysis or reporting of the trial result.

### Randomization

Randomization was performed using central web-based systems. Participants were randomized to receive therapeutic anticoagulation with unfractionated or low molecular weight heparin or usual care pharmacological thromboprophylaxis in an open label fashion. ACTIV-4a randomized all participants in a 1:1 ratio. The other two platforms specified response-adaptive randomization; randomization probabilities were updated in the severe patient group within REMAP-CAP and ATTACC during the interim period between the mpRCT interim data cut and the halt of enrollment.

Therapeutic anticoagulation was administered according to local site protocols for the treatment of acute venous thromboembolism for up to 14 days or recovery (defined as hospital discharge, or liberation from supplemental oxygen for at least 24 hours). Usual care pharmacological thromboprophylaxis was administered according to local practice or with guidance from the trial protocol on maximum dosing, which included either standard low dose thromboprophylaxis or enhanced intermediate dose thromboprophylaxis. A subset of participants enrolled in REMAP-CAP were also randomized in the antiplatelet agent domain and in other domains of that trial.

### Outcome measures

The primary outcome, organ support-free days (OSFDs), was an ordinal scale composed of survival to hospital discharge and, in survivors, the number of days free of organ support to day 21. Death in hospital through day 90 was assigned the worst outcome (–1). Among hospital survivors, the number of days free of respiratory organ support (high flow nasal cannula, non-invasive or invasive ventilation, extracorporeal life support) and cardiovascular organ support (vasopressors or inotropes) through day 21 were recorded. A higher value for OSFDs indicates a better outcome. A participant who was discharged from hospital prior to day 21 was assumed to be alive and free of organ support through 21 days. Pre-specified secondary outcomes included survival to day 90, major thrombotic events or death (a composite of myocardial infarction, pulmonary embolism, ischemic stroke, systemic arterial embolism, and in-hospital death) through to 28 days (ACTIV-4a, ATTACC) or through to hospital discharge (REMAP-CAP). Safety outcomes included major bleeding during the treatment period as defined by the International Society of Thrombosis and Haemostasis for non-surgical patients^17^ and laboratory-confirmed heparin-induced thrombocytopenia. Thrombotic and bleeding events were adjudicated by independent platform-specific adjudication committees blinded to treatment assignment (Protocol Appendix, p. 478).

### Statistical Analysis

Although the platforms enrolled participants independently, the mpRCT analyzed combined individual participant-level data from all platforms using a single overarching Bayesian model (see Protocol Appendix, p. 35). Monthly interim analyses of combined data from all platforms were planned within each of the pre-specified patient groups. Randomization continued within each group until a statistical conclusion of superiority (defined as a >99% posterior probability of proportional odds ratio>1) or futility (>95% posterior probability of proportional odds ratio<1.2) was met for a group.

The primary analysis was a Bayesian cumulative logistic model that calculated the posterior probability distribution for the proportional odds ratio for OSFDs. An odds ratio greater than 1 indicates a better outcome with therapeutic anticoagulation. The primary model adjusted for age (categorized into six groups), sex, site, and time period (2-week epochs). The model estimated treatment effects for each of the patient groups (severe, and moderate stratified by D-dimer), utilizing a Bayesian hierarchical approach, which dynamically borrows information between groups if the observed effects are similar between groups.^18^ If, at an interim analysis, a statistical criteria was reached in one group and not the others, only outcomes for participants in that group would be unblinded. For the purposes of this report, the primary analysis was run on all participants enrolled in the mpRCT (including both moderate and severe patient groups) for whom the primary endpoint was available at the date of database lock, January 28, 2021. The analysis of this dataset was pre-specified in a sub-statistical analysis plan for this preliminary report of the results (see Protocol Appendix, p. 120).

The primary model was fit using a Markov Chain Monte Carlo algorithm with 100,000 samples from the joint posterior distribution, allowing calculation of the posterior distributions for the odds ratios, including medians and 95% credible intervals (CrIs) and the posterior probabilities of superiority (odds ratio>1), futility (odds ratio<1.2), or inferiority (odds ratio<1). A similar model was run for survival to hospital discharge (a key subcomponent of OSFDs). The pre-specified sensitivity analyses of the primary model are described in the Statistical Analysis Plan (Supplementary Appendix). To assess the influence of potential prior enthusiasm for therapeutic anticoagulation, a sensitivity analysis was conducted using an enthusiastic prior (prior mean OR 1.75, 95% CrI 0.74-4.15; prior probability of superiority 90%).

For the key secondary endpoints, similar models were restricted to the severe patient group, without borrowing information from moderate patient groups. Per-protocol analyses were conducted restricted to participants who received a dose of study treatment consistent with their treatment assignment in the trial within 24 hours of randomization (see Protocol Appendix for details on this classification). Subgroup analyses assessed whether treatment effect varied according to age, sex, requirement for mechanical ventilation at baseline, and intensity of thromboprophylaxis dosing in the usual care arm (defined based on the pattern of practice at each participating site, see Protocol Appendix).

## Results

The first participant was randomized on April 21, 2020. Enrollment was discontinued in the severe patient group on December 19^th^ 2020 after an interim analysis demonstrated that the statistical criteria for futility was met. At that time a total of 1,205 participants with severe suspected or confirmed Covid-19 were randomized (**Figure 1**). Of these, 25 participants withdrew consent, 91 participants did not have laboratory-confirmed Covid-19, and the primary outcome was not available in 15 participants as of January 28, 2021. The current report presents the results of the primary analysis for 1,074 participants with severe confirmed Covid-19.

**Figure 1.**
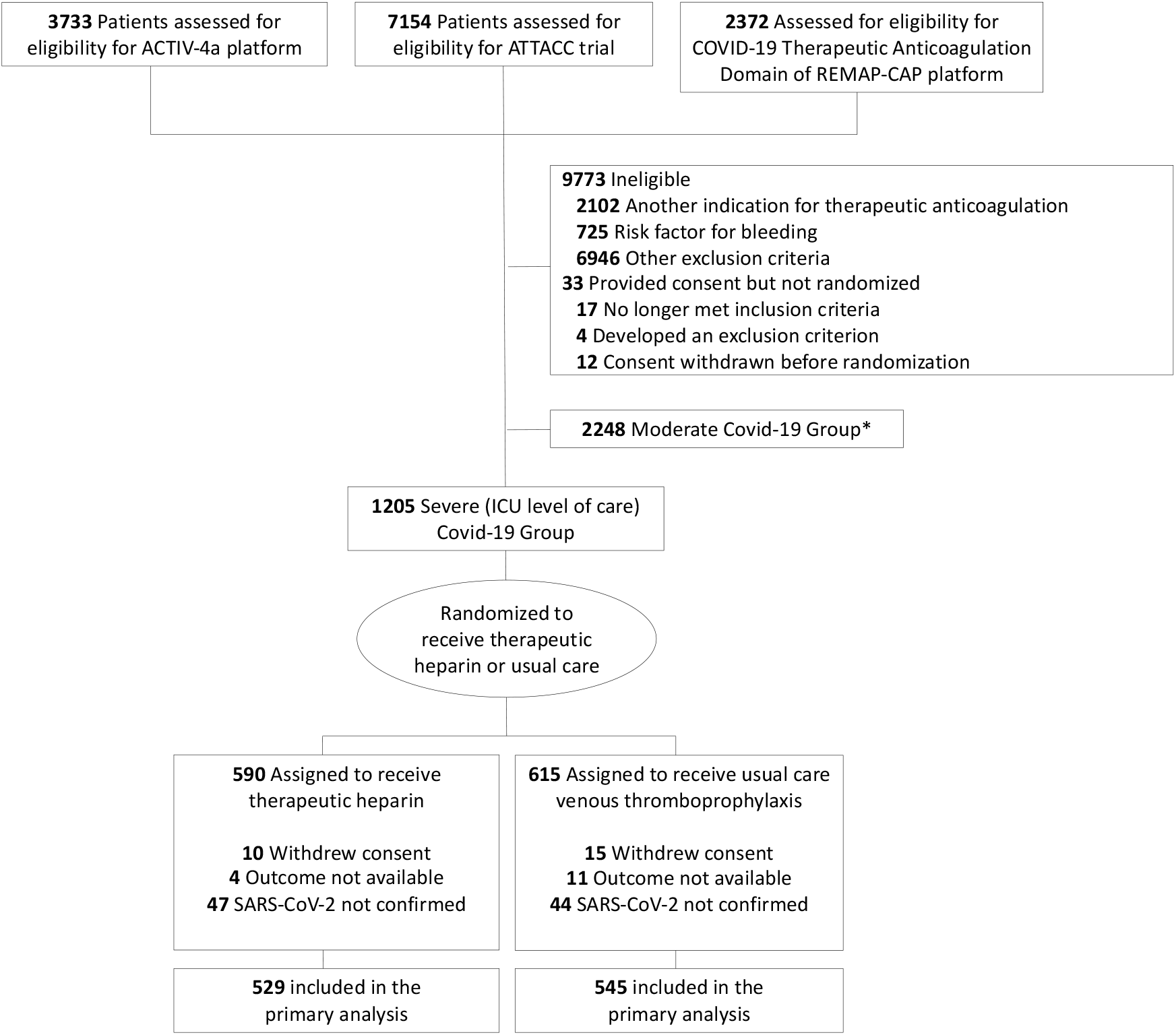
Screening, enrollment, randomization, and inclusion in analysis. *1531 patients from the moderate state were included in the primary statistical model (see Methods for details).

### Participants

Baseline characteristics were comparable between the intervention groups (**Table 1**). The majority of participants were enrolled through the REMAP-CAP platform (n=987, 84%, **Table S1**). The pattern of heparin administration in the intervention groups is described in **Table S1**. In participants randomized to usual care pharmacological thromboprophylaxis, the initial post-randomization dose equivalent corresponded to standard low dose thromboprophylaxis in 41% of participants in whom these data were available and to enhanced intermediate dose thromboprophylaxis in 51% of participants in whom these data were available (**Table S1**).

**Table 1.** Demographic and clinical characteristics of the patients at baseline

|  |  |  |
| --- | --- | --- |
| Characteristic | Therapeutic anticoagulation (N=532) | Usual care pharmacological thromboprophylaxis (N=557) |
| Age – mean (SD), years | 60.2 (13.1) | 61.6 (12.5) |
| Male Sex – n/N (%) | 383/532 (72) | 379/557 (68) |
| Race/Ethnicity – |  |  |
| White n/N (%) | 314/423 (74.2) | 326/443 (73.6) |
| Asian n/N (%) | 69/423 (16.3) | 71/443 (16) |
| Black n/N (%) | 24/423 (5.7) | 20/443 (4.5) |
| Other n/N (%) | 16/423 (3.8) | 26/443 (5.9) |
| Country – n/N (%) |  |  |
| United Kingdom | 388/532 (72.9) | 389/557 (69.8) |
| United States | 78/532 (14.7) | 96/557 (17.2) |
| Canada | 39/532 (7.3) | 53/557 (9.5) |
| Brazil | 12/532 (2.3) | 6/557 (1.1) |
| Other<sup>a</sup> | 15/532 (2.8) | 13/557 (2.3) |
| Body mass index (kg/m<sup>2</sup>) – median (IQR) | 30.4 (26.9, 35.9)<br>(n = 458) | 30.2 (26.4, 34.6)<br>(n = 470) |
| APACHE II score<sup>b</sup> – median (IQR) | 13 (8, 21)<br>(n = 417) | 13 (8, 19)<br>(n = 417) |
| Pre-existing conditions – n/N (%) |  |  |
| Diabetes mellitus (type 1 or 2) | 168/523 (32.1) | 182/546 (33.3) |
| Severe cardiovascular disease<sup>c</sup> | 36/495 (7.3) | 34/517 (6.6) |
| Chronic kidney disease | 56/499 (11.2) | 40/502 (8) |
| Chronic respiratory disease<sup>d</sup> | 121/503 (24.1) | 125/516 (24.2) |
| Chronic liver disease | 6/504 (1.2) | 2/527 (0.4) |
| Baseline treatments<sup>e</sup> – n/N (%) |  |  |
| Antiplatelet agent<sup>f</sup> | 36/488 (7.4) | 42/516 (8.1) |
| Remdesivir | 149/488 (30.5) | 161/517 (31.1) |
| Corticosteroids | 387/488 (79.3) | 410/517 (79.3) |
| Tocilizumab | 9/488 (1.8) | 9/517 (1.7) |
| Baseline organ support – n/N (%) |  |  |
| No oxygen/supplemental oxygen | 8/532 (1.5) | 7/557 (1.3) |
| High flow nasal oxygen | 172/532 (32.3) | 188/557 (33.8) |
| Non-invasive ventilation | 214/532 (40.2) | 200/557 (35.9) |
| Invasive mechanical ventilation | 138/532 (25.9) | 162/557 (29.1) |
| Vasopressors/inotropes | 87/522 (16.7) | 100/548 (18.2) |
| PaO<sub>2</sub>/FIO<sub>2</sub><sup>b</sup> – median (IQR) | 119 (89.5–159.5)<br>(n = 383) | 119 (92–161)<br>(n = 389) |
| Median Laboratory values (IQR) |  |  |
| D-dimer (ng/ml) | 827 (450–1740)<br>(n = 185) | 890 (375–1846)<br>(n = 187) |
| D-dimer greater than or equal 2 times site upper limit of normal – n/N (%) | 88/185 (47.6) | 87/187 (46.5) |
| International normalized ratio | 1.1 (1–1.2)<br>(n = 318) | 1.1 (1–1.3)<br>(n = 314) |
| Neutrophils x10<sup>9</sup>/L | 7.9 (5.5–10.6)<br>(n = 438) | 7.9 (5.6–10.7)<br>(n = 458) |
| Lymphocytes x10<sup>9</sup>/L | 0.7 (0.5–1)<br>(n = 439) | 0.7 (0.5–1)<br>(n = 467) |
| Platelets x10<sup>9</sup>/L | 247 (188–317.5) | 245.5 (185–315) |
|  | (n = 511) | (n = 524) |

- a.** Ireland, Netherlands, Australia, Nepal, Saudi Arabia, and Mexico. - b.** Available in REMAP-CAP only. - c.** Defined in REMAP-CAP as a baseline history of New York Heart Association class IV symptoms; defined in ACTIV-4a and ATTACC as a baseline history of heart failure, myocardial Infarction, coronary artery disease, peripheral arterial disease, or cerebrovascular disease (stroke or transient ischemic attack). - d.** Defined as a baseline history of asthma, chronic obstructive pulmonary disease, bronchiectasis, interstitial lung disease, primary lung cancer, pulmonary hypertension, active tuberculosis, or through the receipt of home oxygen therapy. - e.** Recent or chronic use. - f.** Not included in this summary are 113 patients co-enrolled in the REMAP-CAP Antiplatelet Domain (47 assigned to therapeutic Anticoagulation, 66 to usual care pharmacological thromboprophylaxis).

## Primary Outcome

In participants assigned to therapeutic anticoagulation, the median value for organ support-free days was 3 (interquartile range –1, 16); in participants assigned to usual care pharmacological thromboprophylaxis the median value was 5 (interquartile range –1, 16). In the primary model, the median adjusted proportional odds ratio for the effect of therapeutic anticoagulation on organ support-free days was 0.87 (95% CrI 0.70-1.08), yielding a posterior probability of futility of 99.8% and a posterior probability of inferiority of 89.4% (**Table 2** and **Figure 2**). In-hospital survival was 64.3% in participants assigned to therapeutic anticoagulation and 65.3% in participants assigned to usual care pharmacological thromboprophylaxis (median adjusted odds ratio 0.88, 95% CrI 0.67-1.16; posterior probability of inferiority 81.0%). Survival to day 90 is shown in **Figure 3**.

**Table 2.** Primary and secondary outcomes

| Table 2. Primary and secondary outcomes |  |  |
| --- | --- | --- |
| Outcome | Therapeutic anticoagulation<br>(n=532) | Usual care pharmacological<br>thromboprophylaxis<br>(n=557) |
| Organ support-free days to day 21<sup>a</sup> |  |  |
| No. of patients with known outcome | 529 | 545 |
| Median, IQR | 3 (-1, 16) | 5 (-1, 16) |
| Adjusted proportional odds ratio (95% CrI) | 0.87 (0.70-1.08) | 1 (Reference) |
| Probability of futility, % | 99.8% | - |
| Probability of superiority, % | 10.6% | - |
| Probability of inferiority, % | 89.4% | - |
| Survival to hospital discharge<sup>a</sup> |  |  |
| No. of patients/total no. (%) | 340/529 (64.3%) | 356/545 (65.3%) |
| Adjusted odds ratio (95% CrI) | 0.88 (0.67-1.16) | 1 (Reference) |
| Probability of futility, % | 98.5% | - |
| Probability of superiority, % | 19.0% | - |
| Probability of inferiority, % | 81.0% | - |
| Major thrombotic events or death<sup>b</sup> |  |  |
| No. of patients/total no. (%) |  |  |
| Major thrombotic events | 27/471 (5.7%) | 49/476 (10.3%) |
| Death in hospital | 189/529 (35.7%) | 189/545 (34.7%) |
| Major thrombotic events or death | 200/483 (41.4%) | 211/494 (42.7%) |
| Adjusted odds ratio (95% CrI) | 1.05 (0.79-1.40) | 1 (Reference) |
| Probability of futility, % | 94.5% | - |
| Probability of superiority, % | 37.0% | - |
| Probability of inferiority, % | 63.0% | - |
| Major bleeding<sup>b</sup> |  |  |
| No. of patients/total no. (%) | 15/482 (3.1%) | 12/495 (2.4%) |
| Odds ratio (95% CrI)<sup>a</sup> | 1.19 (0.57-2.49) | 1 (Reference) |
| Probability of futility, % | 82.9% | - |
| Probability of superiority, % | 32.3% | - |
| Probability of inferiority, % | 67.7% | - |

<sup>a</sup> Composite ordinal scale consisting of survival to hospital discharge and days free of organ support to day 21. Odds ratio>1 indicates a benefit from treatment. Probabilities of benefit (proportional odds ratio>1), harm (proportional odds ratio<1), and futility (proportional odds ratio<1.2) are computed from the posterior distribution.
<sup>b</sup> Odds ratio<1 indicates a benefit from treatment. Probabilities of benefit (odds ratio<1), harm (proportional odds ratio>1), and futility (odds ratio>(1/1.2)) are computed from the posterior distribution.

**Figure 2.**
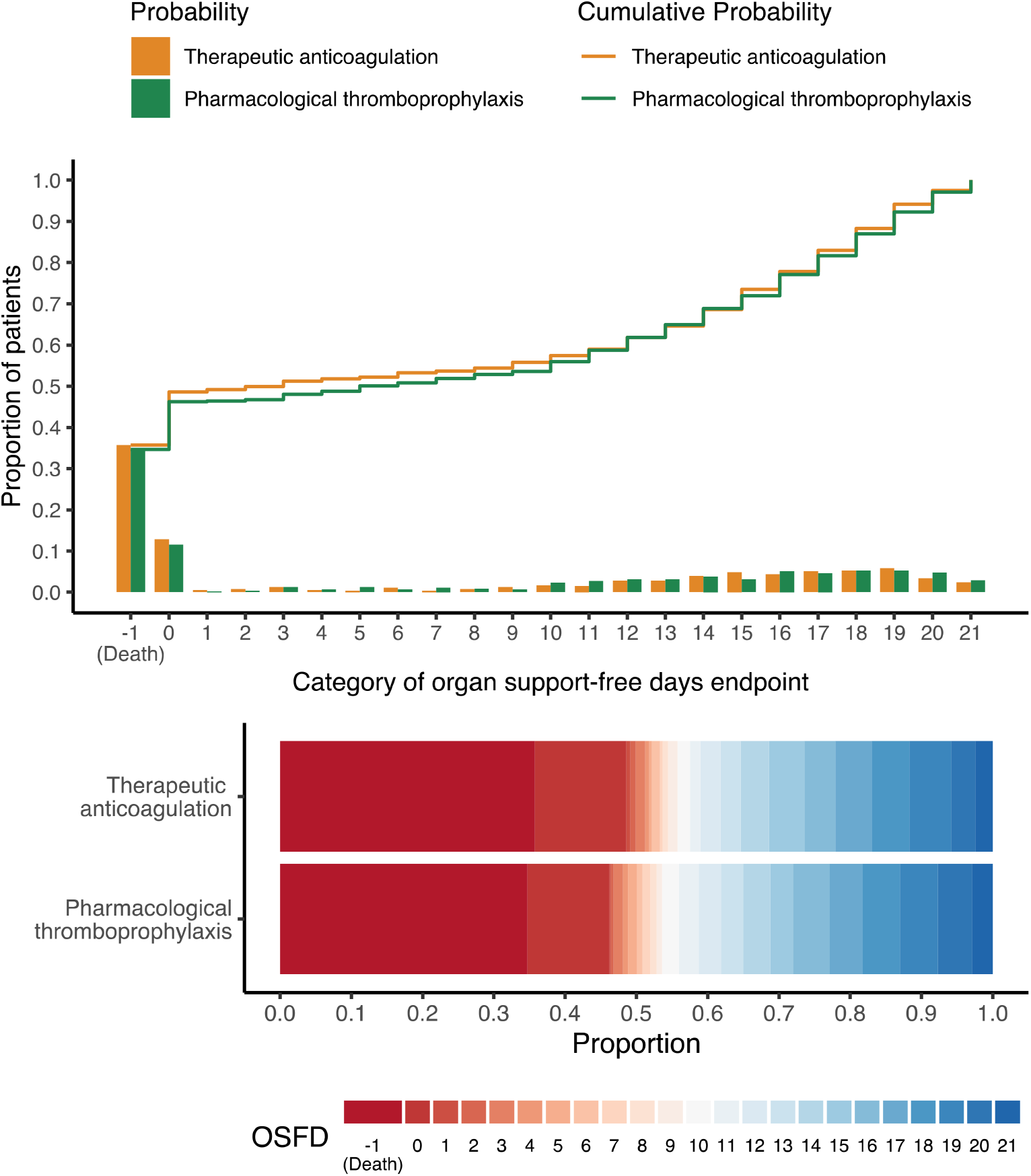
Organ support-free days to day 21. Upper panel) the cumulative proportion (y-axis) for each intervention group by day (x-axis), with death listed first. Curves that rise more slowly indicate a more favorable distribution in the number of days alive and free of organ support. The height of each curve at “-1” indicates the in-hospital mortality rate for each intervention. The height of each curve at any point, for example, at day = 10, indicates the proportion of patients with organ support-free days (OSFD) of 10 or lower (i.e. 10 or worse). The difference in height of the two curves at any point represents the difference in the cumulative probability of having a value for OSFDs less than or equal to that point on the x-axis. Lower panel) Organ support–free days as horizontally stacked proportions by intervention group. Red represents worse outcomes and blue represents better outcomes. The median adjusted odds ratio for the primary analysis was 0.87 (95% credible interval 0.70-1.08, posterior probability of futility 99.8%).

**Figure 3.**
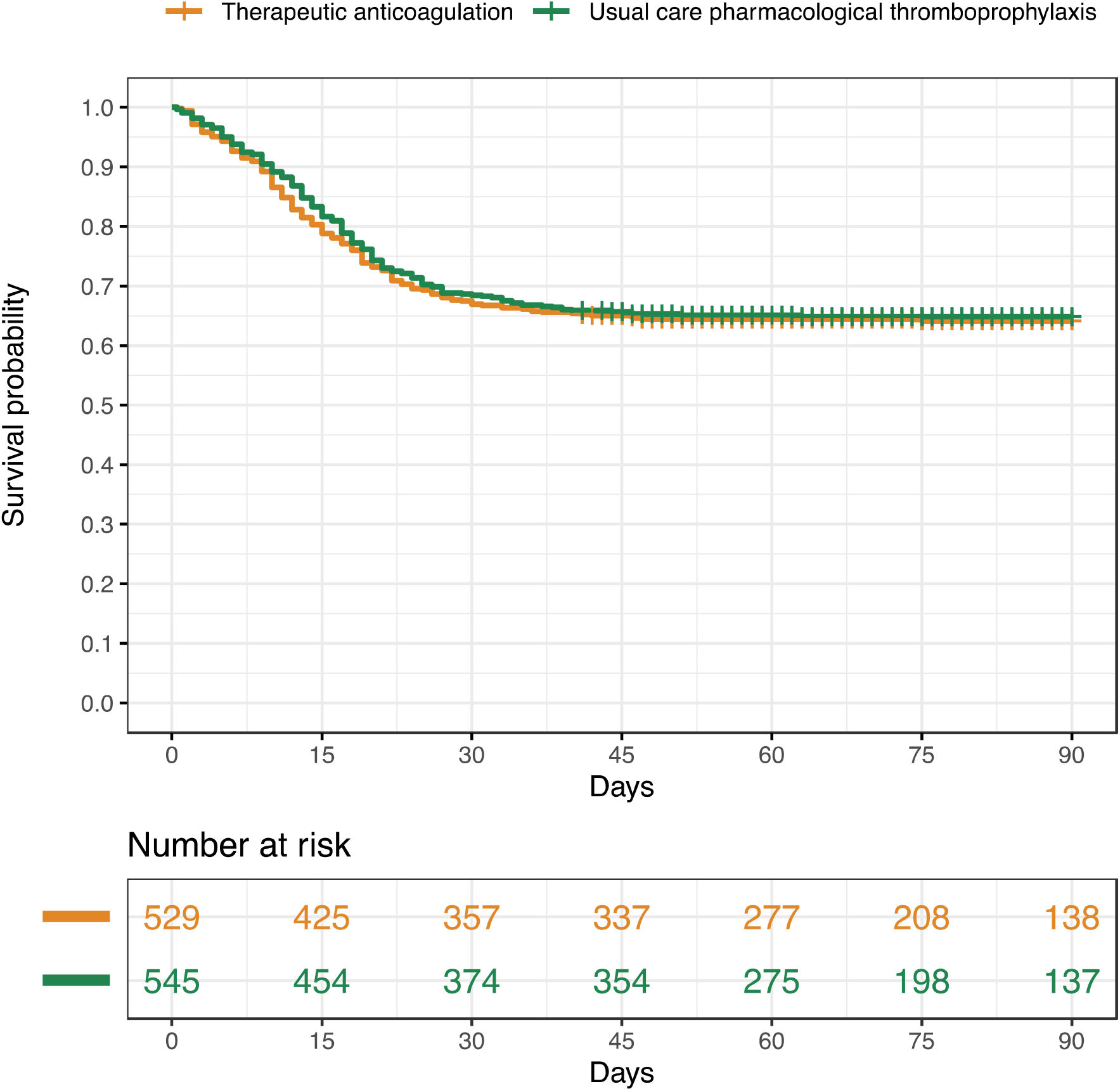
Survival to day 90 according to treatment assignment.

In sensitivity analyses of the primary outcome (**Table S2**), incorporation of prior enthusiasm for therapeutic anticoagulation did not modify the conclusion (median adjusted proportional odds ratio 0.89, 95% CrI 0.72-1.10). Restricting to participants managed per-protocol gave similar results (median adjusted proportional odds ratio 0.94, 95% CrI 0.70-1.25). Including participants with suspected Covid-19 and excluding participants with concomitantly receiving an antiplatelet agent at baseline or those enrolled in the REMAP-CAP antiplatelet agent domain also yielded similar results (**Table S2**). In pre-specified subgroup analyses, the estimated effect did not meaningfully vary according to age, sex, baseline requirement for invasive mechanical ventilation, or the pattern of usual care pharmacological thromboprophylaxis dosing at sites (intermediate vs. low dose) (**Figure S1**).

## Secondary Outcomes

Major thrombotic events or death are reported in **Table 2**. Although there were numerically fewer participants with major thrombotic events among those assigned to therapeutic anticoagulation in comparison to those assigned to usual care pharmacological thromboprophylaxis (5.7% vs. 10.3%), the secondary efficacy outcome of major thrombotic events or death was similar between groups (41.4% vs. 42.7%, median adjusted odds ratio 1.05, 95% CrI 0.79-1.40). A breakdown of major thrombotic events is provided in **Table S3**. A major bleeding event occurred during the treatment period in 3.1% of participants assigned to therapeutic anticoagulation and in 2.4% of participants assigned to usual care pharmacological thromboprophylaxis.

## Discussion

In this international multiplatform randomized clinical trial of over 1,000 critically ill patients with confirmed Covid-19, therapeutic anticoagulation did not improve survival or days free of organ support and had an 89% probability of being inferior to usual care pharmacological thromboprophylaxis. There was an 81% probability that therapeutic anticoagulation reduced survival to hospital discharge in comparison to usual care pharmacological thromboprophylaxis. Bleeding complications were infrequent in both groups. Although therapeutic anticoagulation resulted in a numerical decrease in major thrombotic events, it did not improve organ support-free days or survival to hospital discharge.

Our results refute the hypothesis that, in the absence of a usual clinical indication for therapeutic anticoagulation, empiric administration of therapeutic anticoagulation benefits critically ill patients with Covid-19. This hypothesis was based on observational studies that reported therapeutic anticoagulation was associated with improved outcomes, particularly in critically ill patients.^14,19,20^ Multiple small and moderate-size randomized trials continue to evaluate different anticoagulation strategies in Covid-19.^21^ The results of this study demonstrate that, in critically ill patients with Covid-19, the probability of benefit from a routine full-dose therapeutic anticoagulation strategy is low.

The net effect of enhanced anticoagulation on clinical outcome may depend on the degree of coagulation or inflammation or on the timing of initiation in relation to disease course. As has been reported for other therapies used in Covid-19, the effectiveness of anticoagulation in Covid-19 may vary with the severity of illness when therapy is commenced.^22- 24^ Despite demonstrable activation of coagulation and systemic inflammation in severe Covid-19, our findings suggest that initiating therapeutic anticoagulation once patients develop severe Covid-19 may be too late to reasonably alter the consequences of established disease pathology. The net clinical effect of anticoagulation might also be modified by the use of concomitant immunomodulatory therapies.

In this trial, the probability of inferiority of therapeutic anticoagulation was 89%. Mechanisms that could account for this likely harm are uncertain. Although major bleeding was numerically increased with therapeutic anticoagulation, the occurrence of major bleeding with full-dose therapeutic anticoagulation was low (3.1%) and consistent with previous estimates of bleeding in critically ill patients.^25^ In patients with Covid-19 and severe ARDS, autopsy findings include microthrombosis but also alveolar hemorrhage.^26^ It is possible that, in the presence of marked pulmonary inflammation, therapeutic anticoagulation could exacerbate alveolar hemorrhage leading to worse outcome.

This trial was made possible through a major global collaboration to conduct what is, to our knowledge, the first multiplatform clinical trial whereby a harmonized pragmatic trial protocol was actioned by three platform networks spanning five continents. The interventions evaluated are familiar and widely accessible, rendering the findings highly applicable to critically ill patients with severe Covid-19 disease. Analyses were pre-specified using a Bayesian framework that incorporated frequent interim analyses. Through a combined effort to collaboratively inform practice, we reached a trial conclusion for futility with probable harm more quickly than would have been possible as independent platforms.

One limitation of our trial is the open-label design, although clinician or participant awareness likely had little or no impact on the primary outcome that incorporated mortality and duration of organ support. The open label strategy may also introduce systematic bias in the ascertainment of thrombotic events. Additionally, the pragmatic design of this trial allowed clinicians to employ local site practice in the usual care pharmacological thromboprophylaxis arm. A substantial majority of enrollment in the severe patient group was in the United Kingdom where national practice guidelines changed during the trial to recommend that Covid-19 patients admitted to an ICU receive intermediate dose anticoagulation for thromboprophylaxis. Many participants in the usual care arm therefore received an intermediate dose of thromboprophylaxis. It is possible that the benefit of therapeutic anticoagulation varies according to management of the comparator group. However, in pre-specified subgroup analyses, the treatment effect of therapeutic dose anticoagulation did not vary meaningfully according to site proclivity for low or intermediate dose thromboprophylaxis. Whether intermediate dose thromboprophylaxis is superior to standard low dose thromboprophylaxis in critically ill patients is uncertain. Moreover, the effect of therapeutic anticoagulation in hospitalized, non critically ill patients with Covid-19 remains to be determined.

In conclusion, in critically ill patients with Covid-19, there was no benefit of therapeutic anticoagulation with heparin compared to usual care pharmacological thromboprophylaxis and a high probability of inferiority.

## Supporting information

mpRCT Protocol Appendix

Supplementary Appendix Anticoagulation mpRCT Severe State

## Data Availability

The data is not available

## Acknowledgements

We are grateful for the support of the participants and their families who participated in this trial. We thank those who served on the Data Safety and Monitoring Boards of each platform; we gratefully acknowledge the support of multiple funding organizations for the participating platforms (see Supplementary Appendix).

## Authors

**Executive Writing Committee**

Ewan C. Goligher, M.D., Ph.D.*^1,2^, Charlotte Ann Bradbury, M.B., Ch.B.*^3,4^, Bryan J. McVerry, M.D.*^5,6^, Patrick R. Lawler, M.D. M.P.H.* ^1,7^, Jeffrey S. Berger, M.D.*^8^, Michelle N. Gong, M.D.*^9,10^, Marc Carrier, M.D., M.Sc. ^11,12^, Harmony R. Reynolds, M.D.^8^, Anand Kumar, M.D.^13,^, Alexis F. Turgeon, M.D., M.Sc.^14,15^, Lucy Z. Kornblith, M.D.^16^, Susan R Kahn, M.D., M.Sc.^17^, John C. Marshall, M.D.^18^, Keri S. Kim, Pharm. D.^19^, Brett L Houston, M.D.^13,20^, Lennie P. G. Derde, M.D., Ph.D.^21^, Mary Cushman, M.D., M.Sc.^22^, Tobias Tritschler, M.D., M.Sc.^23^, Derek C. Angus, M.D., M.P.H.^5,6^, Lucas C. Godoy, M.D.^1,7,24^, Zoe McQuilten, Ph.D.^25^, Bridget-Anne Kirwan, Ph.D.^26,27^, Michael E. Farkouh, M.D.^1,7^, Maria M. Brooks, Ph.D.^5^, Roger J. Lewis, M.D., Ph.D. ^28,29^, Anthony C. Gordon, M.B.B.S., M.D.^30,31^, Scott M. Berry, Ph.D.**^28^, Colin J. McArthur, M.B., Ch.B.**^25,32,33^, Matthew D. Neal, M.D.**^5,6^, Judith S. Hochman, M.D.**^8^, Steven A. Webb, M.P.H., Ph.D.**^25,34^, Ryan Zarychanski, M.D., M.Sc.**^13,20^*, ** denotes equal contribution

**Block Writing Committee:** *(In alphabetical order)*

Tania Ahuja, Pharm.D.^35^, Farah Al-Beidh, Ph.D.^30^, Djillali Annane, M.D., Ph.D.^36^, Yaseen M. Arabi, M.D.^37,38^, Diptesh Aryal, M.D.^39,40^, Lisa Baumann Kreuziger, M.D.^41^, Abi Beane, Ph.D.^42,43^, Lindsay

R. Berry, Ph.D.^28^, Zahra Bhimani, M.P.H.^18^, Shailesh Bihari, Ph.D.^44^, Henny H. Billett, M.D., M.Sc.^9,10^, Lindsay Bond ^45^, Marc Bonten, Ph.D.^21^, Frank Brunkhorst ^46^, Meredith Buxton, Ph.D.^47^, Adrian Buzgau ^25^, Lana A. Castellucci, M.D., M.Sc.^11,48^, Sweta Chekuri, M.D.^9^, Jen-Ting Chen, M.D., M.S. ^9^, Allen C. Cheng, Ph.D.^49,50^, Tamta Chkhikvadze, M.D.^8,35^, Benjamin Coiffard, M.D., M.Sc.^51^, Aira Contreras, M.A.^8,35^, Todd W. Costantini, M.D.^52^, Sophie de Brouwer, Ph.D.^26^, Michelle A. Detry, Ph.D.^28^, Abhijit Duggal, M.D., M.P.H., M.Sc.^53^, Vladimír Džavík, M.D.^1,7^, Mark B. Effron, M.D.^54^, Heather F. Eng, BA ^5^, Jorge Escobedo, M.D.^55^, Lise J. Estcourt, M.B.B.Chir., D.Phil.^33, 42^, Brendan M. Everett, M.D., M.P.H.^56^, Dean A. Fergusson, Ph.D.^11,48^, Mark Fitzgerald, Ph.D.^28^, Robert A. Fowler, M.D.^1^, Joshua D. Froess, M.S.^5^, Zhuxuan Fu, M.S., M.P.H.^5^, Jean Philippe Galanaud, M.D.^1,57^, Benjamin T. Galen, M.D.^10^, Sheetal Gandotra, M.D.^58^, Timothy D. Girard, M.D.^59^, Andrew L. Goodman, M.D.^60^, Herman Goossens, M.D.^61^, Cameron Green, M.Sc.^25^, Yonatan Y. Greenstein, M.D.^62^, Peter L. Gross, M.D.^63,64^, M.Sc., Rashan Haniffa, Ph.D.^65,66^, Sheila M. Hegde, M.D., M.P.H.^67^, Carolyn M. Hendrickson, M.D.^16^, Alisa M. Higgins, Ph.D.^25^, Alexander A. Hindenburg, M.D.^68^, Aluko A. Hope, M.D., M.S.C.E.^9,10^, James M. Horowitz, M.D.^35^, Christopher M. Horvat, M.D., M.H.A.^69^, David T. Huang, M.D.^5^, Kristin Hudock, M.D., M.S.T.R.^70^, Beverley J. Hunt, O.B.E.^71^, Mansoor Husain, M.D.^1,2^, Robert C. Hyzy, M.D.^72^, Jeffrey R. Jacobson, M.D.^19^, Devachandran Jayakumar, M.D.^73^, Norma M. Keller, M.D. ^8,74^, Akram Khan, M.D.^75^, Yuri Kim, M.D., Ph.D.^56^, Andrei Kindzelski, M.D., Ph.D.^76^,Andrew J. King, Ph.D.^5^, M. Margaret Knudson, M.D.^16^, Aaron E. Kornblith, M.D.^16^, Matthew E. Kutcher, M.D., M.S.^77^, Michael A. Laffan, D.M.^30^, Francois Lamontagne, M.D.^78^, Grégoire Le Gal, M.D., Ph.D.^11,12^, Christine M. Leeper, M.D., M.Sc.^5^, Eric Leifer, Ph.D.^76^, George Lim, M.D.^79^, Felipe Gallego Lima, M.D.^24^, Kelsey Linstrum, M.S.^5,6^, Edward Litton, Ph.D. ^34,80^, Jose Lopez-Sendon, Ph.D.^81^, Elizabeth Lorenzi, Ph.D.^28^, Sylvain A. Lother, M.D.^13^, Nicole Marten, R.N.^82^, Andréa Saud Marinez, Pharm.D., M.Sc.^83^, Mary Martinez, M.S.^5^, Eduardo Mateos Garcia, M.D., M.Sc.^55^, Stavroula Mavromichalis, M.A.^8,33^, Daniel F. McAuley, M.D.^84,85^, Emily G. McDonald, M.D., M.Sc.^17^, Anna

McGlothlin, Ph.D.^28^, Shay P. McGuinness, M.B., Ch.B.^25,86^, Saskia Middeldorp, M.D., Ph.D.^87^, Stephanie K. Montgomery, M.Sc.^5^, Paul R. Mouncey, M.Sc.^88^, Srinivas Murthy, M.D.^89^, Girish B. Nair, M.D., M.S.^90,91^, Rahul Nair, M.D.^9^, Alistair D. Nichol, M.B., Ph.D.^25,50,92^, Jose C. Nicolau, M.D., Ph.D.^24^, Brenda Nunez-Garcia, BA ^16^, John J. Park, B.S.^16^, Pauline K. Park, M.D.^72^, Rachael L. Parke, Ph.D.^86,93^, Jane C. Parker, B.N.^25^, Sam Parnia, M.D., Ph.D.^8^, Jonathan D. Paul, M.D.^94^, Mauricio Pompilio, Ph.D.^95,96^, John G. Quigley, M.D.^19^, Robert S. Rosenson, M.D.^97,98^, Natalia S. Rost, M.D.^99,100^, Kathryn Rowan, Ph.D.^88^, Fernanda O. Santos ^101^, Marlene Santos, M.D., M.Sc.^18^, Mayler Olombrada Santos, M.Sc.^102^, Lewis Satterwhite, M.D.^103^, Christina T. Saunders, Ph.D.^28^, Jake Schreiber, M.B.A., M.P.H.^104^, Roger E.G. Schutgens, M.D., Ph.D.^21,105^, Christopher W. Seymour, M.D.^5^, Deborah M. Siegal, M.D., M.Sc.^11,48^, Delcio Goncalves Silva Jr., M.Med.^106,107^, Aneesh B. Singhal, M.D.^99,100^, Arthur S. Slutsky, M.D. ^1,18^, Dayna Solvason^13^, Simon J. Stanworth, F.R.C.P., D.Phil.^33,42^, Anne M. Turner, M.P.H.^108^, Wilma van Bentum-Puijk, M.Sc.^21^, Frank L. van de Veerdonk, M.D., Ph.D.^87^, Sean van Diepen, M.D., M.Sc.^109^, Gloria Vazquez-Grande, M.D., M.Sc.^13^, Lana Wahid, M.D.^110^, Vanessa Wareham, H.B.Sc.^45^, R. Jay Widmer, M.D., Ph.D.^111^, Jennifer G. Wilson, M.D.^112^, Eugene Yuriditsky, M.D.^35^, Yongqi Zhong, M.B., M.P.H.^5^.

1. University of Toronto, Toronto, Canada
2. University Health Network, Toronto, Canada
3. University of Bristol, Bristol, United Kingdom
4. University Hospitals Bristol and Weston NHS Foundation Trust, Bristol, United Kingdom
5. University of Pittsburgh, Pittsburgh, United States
6. UPMC, Pittsburgh, United States
7. Peter Munk Cardiac Centre at University Health Network, Toronto, Canada
8. NYU Grossman School of Medicine, New York City, United States
9. Montefiore Medical Center, Bronx, United States
10. Albert Einstein College of Medicine, Bronx, United States
11. Ottawa Hospital Research Institute, Ottawa, Canada
12. Institut du Savoir Montfort, Ottawa, Canada
13. University of Manitoba, Winnipeg, Canada
14. Université Laval, Québec City, Canada
15. CHU de Québec – Université Laval Research Center, Québec City, Canada
16. Zuckerberg San Francisco General Hospital/University of California, San Francisco, United States
17. McGill University, Montreal, Canada
18. St. Michael’s Hospital Unity Health, Toronto, Canada
19. University of Illinois, Chicago, United States
20. CancerCare Manitoba, Winnipeg, Canada
21. University Medical Center Utrecht, Utrecht University, Utrecht, The Netherlands
22. Larner College of Medicine at the University of Vermont, Burlington, United States
23. Inselspital, Bern University Hospital, University of Bern, Switzerland
24. Instituto do Coração (InCor), Hospital das Clínicas HCFMUSP, Universidade de São Paulo, São Paulo, Brazil
25. Australian and New Zealand Intensive Care Research Centre, Monash University, Melbourne, Australia
26. SOCAR Research SA, Nyon, Switzerland
27. London School of Hygiene and Tropical Medicine, London, UK
28. Berry Consultants, LLC, Austin, United States
29. Harbor-UCLA Medical Center, Torrance, United States
30. Imperial College London, London, United Kingdom
31. Imperial College Healthcare NHS Trust, St. Mary’s Hospital, London, United Kingdom
32. Auckland City Hospital, Auckland, New Zealand
33. NHS Blood and Transplant, Oxford, United Kingdom
34. St John of God Hospital, Subiaco, Australia
35. NYU Langone Health, NYU Langone Hospital, New York City, United States
36. Fédération Hospitalo Universitaire SEPSIS, Garches, France
37. King Saud bin Abdulaziz University for Health Sciences, Riyadh, Kingdom of Saudi Arabia
38. King Abdullah International Medical Research Center, Riyadh, Kingdom of Saudi Arabia
39. Nepal Mediciti Hospital, Lalitpur, Nepal
40. Nepal Intensive Care Research Foundation, Kathmandu, Nepal
41. Versiti Blood Research Institute, Milwaukee, United States
42. Oxford University, Oxford, United Kingdom
43. NICS-MORU, Colombo, Sri Lanka
44. Flinders University, Bedford Park, Australia
45. Ozmosis Research Inc., Toronto, Ontario
46. Jena University Hospital, Jena, Germany
47. Global Coalition for Adaptive Research, Los Angeles, United States
48. University of Ottawa, Ottawa, Canada
49. Monash University, Melbourne, Australia
50. Alfred Health, Melbourne, Australia
51. Aix-Marseille University, Marseille, France
52. University of California San Diego School of Medicine, San Diego, United States
53. Cleveland Clinic, Cleveland, Ohio
54. Ochsner Medical Center, University of Queensland-Ochsner Clinical School, New Orleans, United States
55. Instituto Mexicano del Seguro Social, Mexico City, Mexico
56. Harvard Medical School and Brigham and Women’s Hospital, Boston, United States
57. Sunnybrook Health Sciences Centre, Toronto, Canada
58. University of Alabama, Birmingham, United States
59. The Clinical Research, Investigation, and Systems Modeling of Acute Illness (CRISMA) Center, University of Pittsburgh, Pittsburgh, United States
60. TriStar Centennial Medical Center, Nashville, United States
61. University of Antwerp, Wilrijk, Belgium
62. Rutgers New Jersey Medical School, Newark, United States
63. McMaster University, Hamilton, Canada
64. Thrombosis and Atherosclerosis Research Institute, Hamilton, Canada
65. University of Oxford, Bangkok, Thailand
66. University College London Hospital, London, United Kingdom
67. Brigham and Women’s Hospital, Boston, United States
68. NYU Langone Long Island, Mineola, United States
69. UPMC Children’s Hospital of Pittsburgh, Pittsburgh, United States
70. University of Cincinnati, Cincinnati, United States
71. Kings Healthcare Partners, London, United Kingdom
72. University of Michigan, Ann Arbor, United States
73. Apollo Speciality Hospital - OMR, Chennai, India
74. Bellevue Hospital, New York City, United States
75. Oregon Health & Science University, Portland, United States
76. National Heart Lung and Blood Institute, National Institutes of Health, Bethesda, MD, United States
77. University of Mississippi Medical Center, Jackson, United States
78. Université de Sherbrooke, Sherbrooke, Canada
79. University of California Los Angeles, Los Angeles, United States
80. Fiona Stanley Hospital, Perth, Australia
81. IdiPaz Research Institute, Universidad Autonoma, Madrid, Spain
82. St Boniface Hospital, Winnipeg, Canada
83. Avanti Pesquisa Clínica, Sao Paulo, Brazil
84. Queen’s University Belfast, Belfast, Northern Ireland
85. Royal Victoria Hospital, Belfast, Northern Ireland
86. Auckland City Hospital, Auckland, New Zealand
87. Radboud University Medical Center, Nijmegen, The Netherlands
88. Intensive Care National Audit & Research Centre (ICNARC), London, United Kingdom
89. University of British Columbia, Vancouver, Canada
90. Beaumont Health, Royal Oak, United States
91. OUWB School of Medicine, Auburn Hills, United States
92. University College Dublin, Dublin, Ireland
93. The University of Auckland, Auckland, New Zealand
94. University of Chicago, Chicago, United States
95. Hospital do Coração de Mato Grosso do Sul (HCMS), Campo Grande, Brazil
96. Federal University of Mato Grosso do Sul (UFMS), FAMED, Campo Grande, Brazil
97. Icahn School of Medicine at Mount Sinai, New York City, United States
98. Mount Sinai Heart, New York City, United States
99. Massachusetts General Hospital, Boston, United States
100. Harvard Medical School, Boston, United States
101. Hospital 9 de Julho, São Paulo, Brazil
102. INGOH, Clinical Research Center, Goiânia, Brazil.
103. University of Kansas School of Medicine, Kansas City, United States
104. The Chartis Group, Chicago, United States
105. University Utrecht, Utrecht, The Netherlands
106. Hospital Universitário Maria Aparecida Pedrossia, Campo Grande, Brazil
107. Hospital Unimed Campo Grande, Campo Grande, Brazil
108. Medical Research Institute of New Zealand, Wellington, New Zealand
109. University of Alberta, Edmonton, Canada
110. Duke University Hospital, Durham, North Carolina
111. Baylor Scott and White Health, Temple, United States
112. Stanford University School of Medicine, Palo Alto, United States

