## Supplementary material for "Therapeutic Anticoagulation in Critically Ill Patients with Covid-19 – Preliminary Report": mpRCT Protocol Appendix

### Appendix 1

#### Multi-Platform Randomized Controlled Trial (mpRCT) Protocols and Cross-Trial Comparisons

### Appendix 1 - Table of Contents

**Introduction** **19**

**Protocol Synopses** **20**

*REMAP-CAP* *20*

*ATTACC* *21*

*ACTIV-4a* *26*

**Cross-Platform Protocol Comparison Tables** **29**

*Eligibility Criteria* *29*

*Interventions* *31*

*Endpoints* *33*

**Multi-platform Randomized-Controlled Trial (mpRCT) Analysis Plan for ACTIV-4, ATTACC, and REMAP-CAP (AARC)** **35**

**Statistical Analysis Plan for the Multi-Platform Randomized Clinical Trial of Therapeutic Anticoagulation for COVID-19** **41**

*Version history* *41*

*SAP Authors* *41*

*SAP Introduction* *42*

mpRCT status at time of SAP preparation 42

Prior analysis plan documents 42

Reporting strategy 43

*Design Considerations* *43*

Patient Subgroups 43

Interventions 44

Primary Endpoint 44

Adaptive Design 45

Endpoint adjudication 46

*Unblinding* *46*

*Analysis Populations* *46*

*Endpoints* *47*

*Graphical Data Summaries* *51*

*Descriptive Statistics* *51*

*Baseline Characteristics and Co-interventions* *51*

*Adherence* *51*

*Analytic Approach* *51*

Primary Analysis of Primary Endpoint 52

Proportional Odds Assumption 52

Analytic Approach for Secondary Dichotomous Endpoints 52

Analytic Approach for Secondary Time-To-Event Endpoints 53
Analytical Approach for Subgroup Analyses 53
Markov Chain Monte Carlo (MCMC) Model Stability 53
Model Outputs 54
Exploratory Analyses 54
Handling of missing data 54
Definition of times 54
Post-randomization analyses 55

*List of Pre-Specified Analyses* *55*

*Models for urgent reporting* *60*
Primary analysis of OSFDs 60
Primary analysis of in-hospital mortality 61
Sensitivity analysis for OSFDs – proportional odds assumptions 62
Sensitivity analysis of OSFDs in unblinded population 63
Sensitivity analysis of in-hospital mortality in unblinded population 64
Sensitivity analysis for OSFDs – include suspected but not confirmed patients 65
Sensitivity analysis for in-hospital mortality – include suspected but not confirmed patients 66
Sensitivity analysis for OSFDs – site and time effects removed 67
Sensitivity analysis for in-hospital mortality – site and time effects removed 68
Sensitivity analysis excluding patients on antiplatelet agents at baseline 69
Sensitivity analysis excluding patients on antiplatelet agents at baseline or during treatment 70
Sensitivity analysis of OSFDs specifying a prior describing enthusiasm or skepticism for benefit 71
Sensitivity analysis of OSFDs treating missing OSFDs based on last known status carried forward 72
Secondary analysis of major thrombotic events or death 73
Secondary analysis of all thrombotic events or death 74
Secondary analysis of deep venous thrombosis 75
Secondary analysis of pulmonary embolism 76
Secondary analysis of 90-day mortality 77
A secondary analysis of days free of vasopressors or inotropes 78
A secondary analysis of days free of invasive or non-invasive ventilation 79
Days free of renal-replacement therapy 80
ECMO utilization 81
A secondary analysis of ICU length-of-stay 82
A secondary analysis of hospital length-of-stay 83
ICU readmission 84
Ischemic cerebrovascular event 85
Systemic arterial thromboembolism 86
Acute myocardial infarction 87
World Health Organization Ordinal Scale 88
Safety analysis of major bleeding 89
Sensitivity analysis of major bleeding – exclude patients on antiplatelet therapy at baseline 90
Safety analysis of laboratory-confirmed heparin-induced thrombocytopenia 91
Safety analysis of fatal bleeding 92
Safety analysis of symptomatic bleeding into critical organ 93
Safety analysis of intracranial hemorrhage 94
Safety analysis of bleeding leading to a ≥20 g/L drop in hemoglobin 95
Safety analysis of bleeding leading to ≥2 red blood cell unit transfusion 96
Per protocol analysis of OSFDs 97
Per protocol analysis of in-hospital mortality 98

Per protocol analysis of major thrombotic events 99
Per protocol analysis of all thrombotic events 100
Per protocol analysis of deep venous thrombosis 101
Per protocol analysis of pulmonary embolism 102
Per protocol analysis of 90-day mortality 103
Per protocol analysis of days free of vasopressors or inotropes 104
Per protocol analysis of days free of invasive or non-invasive ventilation 105
Per protocol analysis of days free of renal-replacement therapy 106
ECMO utilization 107
Per protocol analysis of ICU length-of-stay 108
Per protocol analysis of hospital length-of-stay 109
ICU readmission 110
Per protocol analysis of ischemic cerebrovascular event 111
Per protocol analysis of systemic arterial thromboembolism 112
Per protocol analysis of acute myocardial infarction 113
Per protocol analysis of World Health Organization Ordinal Scale 114

*Prospectively defined subgroup analyses* *115*
Age 115

*Other exploratory subgroups of interest* *119*

*Graphical summaries* *119*

**Categorization of frequently used low- and intermediate-heparin doses in the ATTACC, ACTIV-4a, and REMAP-CAP mpRCT** **120**

*Subcutaneous Enoxaparin* *120*
Low dose 120
Intermediate dose 120

*Subcutaneous Dalteparin* *120*
Low dose 120
Intermediate dose 120

*Subcutaneous Tinzaparin* *120*
Low dose 120
Intermediate dose 120

*Subcutaneous Unfractionated heparin* *121*
Low dose 121
Intermediate dose 121

**Statistical Analysis Plan for Preliminary Report of Severe Subtype of mpRCT of Therapeutic Anticoagulation in Covid-19** **122**

*Version* *122*

*Background* *122*

*Unblinded Population* *122*

*Data* *122*

*Planned Analyses* *122*
Primary 122
Sensitivity analyses of the primary models 122

Key secondary and safety endpoints 123
Per protocol analyses 123
Subgroup analyses 123

*Required Variables for Severe State Patients* *124*

**Simulation Report for Multiplatform Randomized-Controlled Trial (mpRCT) for ACTIV4, ATTACC, and REMAP-CAP 127**

**CLINICAL TRIAL SIMULATION REPORT VERSION 130**

***Version History*** *130*

**INTRODUCTION 130**

***Primary Endpoint*** *130*

***Arms*** *130*

***Patient Subgroups*** *130*

***Trial Status*** *131*

*o* ***Trial Design and Simulation Assumptions*** *132*

***Simulation Operating Characteristics*** *133*

**OPERATING CHARACTERISTICS 135**

***Same Treatment Effect Across Subgroups*** *135*

***Effective in Severe Subgroup Only*** *137*

***Effective in Severe and Moderate/High Subgroups*** *139*

***Escalating Effects Across Subgroups*** *141*

• ***Severe State Power by Scenario*** *143*

***a. Moderate/Low Subgroup Power by Scenario*** *144*

***b. Moderate/High Subgroup Power by Scenario*** *145*

**Current State of the Statistical Model for the AARC Multi-Platform Randomized Clinical Trial Analysis Plan 146**

**Multiple Platform RCT DSMB Interaction Plan 157**

ACTIV-4a, ATTACC, and REMAP-CAP Multiplatform Randomized Controlled Trial Data Flow Plan, Interim Analysis Plan and DSMB Coordination Plan 157

*Background* *157*

*Study Synopses* *157*

*DSMB Interaction Protocol* *158*
Report submission 158
DSMB Report review for each platform 158
Report communication between DSMB 159
DSMB Recommendations for each Trial 159

*SUBSECTIONS* *162*
Subsection A: Data Flow and Report Generation Process 162

Subsection B: Joint Efficacy Analysis Plan 163
Subsection C: Joint Safety Analysis Plan 168
Subsection D: Timeline 172
Subsection E: Masking/Blinding 172

**Protocol 1: Randomized, Embedded, Multifactorial Adaptive Platform Trial for Community-Acquired Pneumonia (REMAP-CAP) 173**

**REMAP-CAP Core Protocol 173**

**TABLE OF FIGURES 178**

***5.1. Severe Community-Acquired Pneumonia*** *198*
5.1.1. Introduction 198
5.1.2. Epidemiology 199
5.1.3. Standard care for patients with severe CAP 199
5.1.4. Treatment guidelines 201
5.1.5. Variation in care and compliance with guidelines 201
5.1.6. An unmet need for better evidence 202

***5.2. Influenza pandemics and emerging pathogens*** *202*

***5.3. Randomized Embedded Multifactorial Adaptive Platform Trials*** *202*
5.3.1. Generating clinical evidence 202
5.3.2. Underlying Principles of the Study Design 203
5.3.3. Nomenclature 204
5.3.4. Randomization and Response Adaptive Randomization 204
5.3.5. Embedding 206
5.3.6. Multifactorial 207
5.3.7. Adaptive 208
5.3.8. Nesting of the REMAP within a Registry 213
5.3.9. Platform 214

**6. OBJECTIVES 214**

***6.1. Primary objective*** *214*

***6.2. Secondary objectives*** *215*

**7. SUMMARY OF TRIAL DESIGN 215**

***7.1. Introduction*** *215*

***7.2. Nomenclature*** *216*

***7.3. Study setting and participating regions*** *216*

***7.4. Eligibility criteria*** *217*
7.4.1. REMAP Inclusion Criteria 218
7.4.2. REMAP Exclusion Criteria 218
7.4.3. Domain-Specific Entry criteria 218

***7.5. Interventions*** *219*
7.5.1. Domain-Specific Information 219
7.5.2. Treatment allocation and Response Adaptive Randomization 219
7.5.3. Adaptation of Domains and Interventions 221

*7.6. Endpoints* *221*

**REMAP-CAP Pandemic Appendix to the Core Protocol** **267**

**1. ABBREVIATIONS** **273**

**2. PROTOCOL APPENDIX STRUCTURE** **274**

**3. PANDEMIC APPENDIX TO THE CORE PROTOCOL VERSION** **275**

*3.1. Version History* *275*

**4. PANDEMIC APPENDIX TO THE CORE PROTOCOL GOVERNANCE** **275**

*4.1. Pandemic Working Group* *275*

*4.2. Contact Details* *276*

**5. PANDEMIC WORKING GROUP AUTHORISATION** **277**

**6. BACKGROUND AND RATIONALE** **277**

*6.1. Introduction* *277*

*6.2. Pandemic research preparedness* *278*

6.2.1. Introduction 278

6.2.2. Pre-planned 279

6.2.3. Pre-approved 279

6.2.4. Practiced 280

6.2.5. Implications of REMAP design during a pandemic 280

6.2.6. Setting of research priorities 282

*6.3. WHO endorsement* *282*

**7. ADAPTATION OF REMAP-CAP DURING A PANDEMIC** **283**

*7.1. Objectives* *283*

*7.2. Study setting: definition of an ICU and relationship of settingto severity of illness* *283*

*7.3. Eligibility criteria* *284*

*7.4. Pandemic stratum* *285*

7.4.1. Introduction 285

7.4.2. Activation and deactivation of the PAtC and PISOP stratum 286

*7.5. The pandemic statistical model* *286*

7.5.1. Introduction 286

7.5.2. Pre-specification of trial parameter options 288

7.5.3. Application of other strata specified in the Core Protocol in the pandemic model 288

7.5.4. Strata within the PISOP stratum 289

7.5.5. States within the PISOP stratum 290

**REMAP-CAP Domain Specific Appendix: COVID-19 Therapeutic Anticoagulation** **306**

Summary 308

**TABLE OF CONTENTS** **311**

ABBREVIATIONS 8 311
PROTOCOL APPENDIX STRUCTURE 10 311
COVID-19 THERAPEUTIC ANTICOAGULATION DOMAIN-SPECIFIC APPENDIX VERSION 11 311
COVID-19 THERAPEUTIC ANTICOAGULATION THERAPY DOMAIN GOVERNANCE 11 311
COVID-19 THERAPEUTIC ANTICOAGULATION THERAPY DOMAIN-SPECIFIC WORKING GROUP AUTHORIZATION 13 311
BACKGROUND AND RATIONALE 13 311
DOMAIN OBJECTIVES 21 311
TRIAL DESIGN 22 311
APPENDIX 1. OVERVIEW OF DESIGN AND INITIAL RESULTS FOR THE THERAPEUTIC ANTICOAGULATION DOMAIN 38 314

**PROTOCOL APPENDIX STRUCTURE 317**

**COVID-19 THERAPEUTIC ANTICOAGULATION DOMAIN-SPECIFIC APPENDIX VERSION 319**

*3.1. Version history 319*

**COVID-19 THERAPEUTIC ANTICOAGULATION THERAPY DOMAIN GOVERNANCE 319**

***4.1.*** *Domain members 319*
Members: 319

***4.2.*** *Contact Details 322*

***4.3.*** *Interaction with ATTACC trial 323*

**COVID-19 THERAPEUTIC ANTICOAGULATION THERAPY DOMAIN-SPECIFIC WORKING GROUP AUTHORIZATION 323**

**BACKGROUND AND RATIONALE 323**

***6.1.*** *Domain definition 323*

***6.2.*** *Domain-specific background 325*

**DOMAIN OBJECTIVES 339**

**TRIAL DESIGN 341**

***8.1.*** *Population 341*

***8.2.*** *Eligibility criteria 341*

***8.3.*** *Anticoagulant Interventions 343*

***8.4.*** *Concomitant care 348*

***8.5.*** *Endpoints 348*

**TRIAL CONDUCT 350**

***9.1.*** *Microbiology 350*

***9.2.*** *Domain-specific data collection 350*

***9.3.*** *Criteria for discontinuation 351*

***9.4.*** *Blinding* *351*

**STATISTICAL CONSIDERATIONS 351**

***10.1.*** *Domain-specific stopping rules* *351*

***10.2.*** *Unit-of-analysis and strata* *351*

***10.3.*** *Timing of revealing of randomization status* *353*

***10.4.*** *Interactions with interventions in other domains* *353*

***10.5.*** *Nesting of interventions* *355*

***10.6.*** *Threshold probability for superiority and inferiority* *355*

***10.7.*** *Threshold odds ratio delta for equivalence* *355*

***10.8.*** *Informative priors* *355*

***10.9.*** *Post-trial sub-groups* *357*

**ETHICAL CONSIDERATIONS 357**

***11.1.*** *Data Safety and Monitoring Board* *357*

***11.2.*** *Potential domain-specific adverse events* *357*

***11.3.*** *Domain-specific consent issues* *359*

***11.4.*** *Relationship to Antiplatelet Domain* *359*

**GOVERNANCE ISSUES 359**

***12.1.*** *Funding of domain* *359*

***12.2.*** *Funding of domain interventions and outcome measures* *360*

***12.3.*** *Domain-specific declarations of interest* *361*

**REFERENCES 362**

**APPENDIX 1. OVERVIEW OF DESIGN AND INITIAL RESULTS FOR THE THERAPEUTIC ANTICOAGULATION DOMAIN 369**

***14.1.*** *Introduction* *369*

***14.2.*** *Primary Analysis Model* *369*

***14.3.*** *Simulation Details* *371*

***14.4.*** *Operating Characteristics* *371*

***14.5.*** *Summary* *372*

**Protocol 2: Antithrombotic Therapy to Ameliorate Complications of COVID-19 (ATTACC) 373**

*ANTITHROMBOTIC THERAPY TO AMELIORATE COMPLICATIONS OF COVID-19 (ATTACC), IN COLLABORATION WITH ACCELERATING COVID-19 THERAPEUTIC INTERVENTIONS AND VACCINES (ACTIV-4)* *373*

**Investigator Initiated Sponsor's Agreement to Protocol Version 3.0, 29-Sep-2020 376**

**1 SYNOPSIS 377**

2 LIST OF ABBREVIATIONS 386

3 BACKGROUND 388

4 STUDY RATIONALE 389

4.1 ATTACC Trial Rationale Overview 389

4.2 Risk Stratification in Patients with COVID-19 389

4.3 Hypercoagulability in Patients with COVID-19 390

4.4 Managing Hypercoagulability in Patients with COVID-19 391

4.5 Other Potential Therapeutic Benefits of Heparin in COVID-19: Anti-viral and Anti-inflammatory Effects 391

4.6 Safety of Heparin 392

4.7 Background Summary 393

5 THE TRIAL 395

5.1 Study Design 395

5.2 Patient Population 395

5.3 Primary Endpoint 395

5.4 Secondary Endpoints 395

6 PATIENT ELIGIBILITY 397

6.1 Inclusion Criteria 397

6.2 Exclusion Criteria 397

6.3 Patient Consent 399

6.4 Patient Enrollment 399

7 STUDY PLAN 400

7.1 Anticoagulation Administration 400

7.2 Study Schedule 400

8 CONCOMITANT MEDICATIONS 404

8.1 Drug Interactions 404

9 MEDICINAL PRODUCT 405

9.1 Characterization of Investigation Medicinal Product 405

9.2 Accountability and Destruction 406

9.3 Dose Adjustments 406

9.4 Subject Compliance 406

9.5 Premature Withdrawal/ Discontinuation Criteria 406

10 OPTIONAL BIOREPOSITORY 408

11 SAFETY AND REPORTING REQUIREMENTS 409

11.1 Adverse Event (ONLY study drug related AEs will be reported) 409

11.2 Adverse Event Documentation 409

11.3 Serious Adverse Event (SAE) due to the study drug 409

(we will only report study drug related SAEs): 409

11.4 Reporting Serious Adverse Events (Only Study Drug Related SAEs will be reported) 411

11.5 Procedure for Expedited Reporting 411

11.6 Study Management and Governance 411

12 STATISTICAL ANALYSIS 413

12.1 Study Population 413

12.2 Evaluation of Safety 413

12.3 Trial Design Introduction 413

12.4 Primary Endpoint 413

12.5 Primary Analysis 415

13 ADAPTIVE DESIGN 417

13.1 Subgroup Conclusions 417

14 PUBLICATION POLICES AND DISCLOSURE OF DATA 418

14.1 Intellectual Property 418

14.2 Data Sharing 418

15 ETHICS 419

Ethics Board Approval 419

Each sub study will be reviewed by IRB/REB for determination if full board review is needed or not. 419

16 RECORD ACCESS AND MAINTENANCE OF STUDY RECORDS 420

16.1 Documentation of Subject's Participation 420

16.2 Regulatory Requirements 420

16.3 Subject Confidentiality and Access to Source Data/Documents 420

16.4 Confidentiality of the Study 422

16.5 Registration of Clinical Trial 422

16.6 Data Reporting 422

16.7 Maintenance of Study Records 422

17 QUALITY ASSURANCE AND QUALITY CONTROL 424

17.1 Monitoring / Auditing 424

18 ADMINISTRATIVE PROCEDURES 425

18.2 Protocol Deviations and Violations 425

18.3 Premature Discontinuation of the Study 425

19 REFERENCES 426

20 APPENDIX A – FDA GUIDANCE ON CLINICAL TRIAL CONDUCT DURING COVID-19 PANDEMIC 430

22 APPENDIX C – WHO ORDINAL SCALE FOR CLINICAL IMPROVEMENT 432

Protocol 3: A Multicenter, Adaptive, Randomized Controlled Platform Trial of the Safety and Efficacy of Antithrombotic Strategies in Hospitalized Adults with COVID-19 (ACTIV-4 Acute) 433

*A Multicenter, Adaptive, Randomized Controlled Platform Trial of the Safety and Efficacy of Antithrombotic Strategies in Hospitalized Adults with COVID-19* 433

*Supported by:* 433

*U.S. Department of Health & Human Services (HHS) Version Number: 1.0* 433

Statement of Compliance 435

Table of Contents 436

Master Protocol Summary 441

1 Introduction, Background Information and Scientific Rationale 443

1.1 *Background Information, Significance and Relevant Literature* 443
1.1.1 *Adaptive Design* *444*
1.2 *Potential Risks & Benefits* 445

**2 Study Design** **445**

2.1 *Overall Study Design* 445
2.2 *Randomization* 445

**3 Objectives and Purpose** **445**

**4 Study Design and Endpoints** **446**

4.1 *Description of Study Design* 446
4.2 *Study Endpoints* 446
4.2.1 *Primary Study Endpoint* *446*
4.2.2 *Secondary Endpoints* *446*
*Other Secondary Endpoints:* *446*
4.2.3 *All-cause mortality at 90 days Additional Study Endpoints* *447*
4.2.4 *Safety Endpoints* *447*

**5 Study Enrollment** **447**

5.1 *Inclusion Criteria* 447
5.2 *Exclusion Criteria* 447
5.3 *Vulnerable Subjects* 447
5.4 *Strategies for Recruitment and Retention* 448
5.5 *Duration of Study Participation* 448
5.6 *Participant Withdrawal or Termination* 448
*5.6.1 Reasons for Withdrawal or Termination* *448*
5.7 *Premature Termination or Suspension of Study* 448

**6 Study Agent and Procedural Intervention** **449**

6.1 *Study Agents* 449
6.2 *Duration of Therapy* 449

**7 Study Procedures and Schedule** **449**

7.1 *Study Schedule* 449
7.1.1 *Visit 1 and Hospitalization Visits (see arm-specific appendices for details)* *450*
*Visit 1 (Screening and Randomization)* 450
*Hospitalization Visits* 451
7.1.2 *28 days and/or Date of Hospital Discharge* *451*
7.2 *Concomitant Medications, Treatments, and Procedures* 451
7.3 *Expedited Critical and Major Event Reporting* 451
7.4 *Data and Safety Monitoring Plan and Study Halting Rules* 451

**8 Statistical Considerations** **451**

8.1 *Statistical and Analytical Plans (SAP)* 451
8.2 *Statistical Modeling for the Primary Analysis* 452
8.3 *Model Priors* 453
8.4 *Assessing Effectiveness* 454
8.5 *Analysis Datasets* 455
*8.5.1 Safety Analyses 455*
*8.5.2 Adherence and Retention Analyses 455*
*8.5.3 Baseline Descriptive Statistics 455*
*8.5.4 Planned Interim Analysis 455*
*8.5.5 Safety Review 456*
*8.5.6 Tabulation of Individual Response Data 456*
*8.5.7 Exploratory Analyses 456*
8.6 *Sample Size* 456

**9 Measures to Minimize Bias 456**

9.1 *Enrollment/Randomization* 456

**10 Randomization 457**

**11 Source Documents and Access to Source Data/Documents 457**

**12 Quality Assurance and Quality Control 457**

**13 Ethics/Protection of Human Subjects 457**

13.1 *Ethical Standard* 457
13.2 *Institutional Review Board* 457
13.3 *Informed Consent Process* 457
*13.3.1 Consent/Assent and Other Informational Documents Provided to Participants 457*
*13.3.2 Consent Procedures and Documentation 458*
13.4 *Posting of Clinical Trial Consent Form* 458
13.5 *Participant and Data Confidentiality* 458

**14 Data Handling and Record Keeping 458**

14.1 *Data Collection and Management Responsibilities* 458
14.2 *Study Records Retention* 458
14.3 *Protocol Deviations* 458
14.4 *Publication and Data Sharing Policy* 459

**15 Study Finances 459**

15.1 *Funding Source* 459
15.2 *Costs to the Participant* 459

**16 Conflict of Interest Policy 459**

**17 References 459**

**Appendix 1: Criteria for Addition and Replacement of Arms** **462**

**Appendix 2: Definition and Determination of Outcomes** **463**

*A2.1 Approach to ascertainment and verification of outcomes* 463
*A2.2 Outcome definitions* 463

*Primary Endpoint* *463*

*Deep vein thrombosis* *464*

*Pulmonary embolism* *464*

*Stroke/ Peripheral Arterial Systemic Thromboembolism* *464*

*ICU Level of care disease state* *464*

*Myocardial infarction* *465*

*Acute Kidney Injury* *465*

*Disseminated Intravascular Coagulation (DIC) (Overt) – DIC score ≥ 5* *465*

*ISTH Defined Major Bleeding* *465*

*Symptomatic Intracranial or Intracerebral Hemorrhage (sICH)* *465*
*A3.1 Therapeutic Dose Anticoagulation*** 467
*A3.2 Discontinuation of study intervention:* 467
*A3.3 Study Schedule* 468
*A3.4 Potential Risks & Benefits* 469

*A3.4.1 Known Potential Risks* *469*

*A3.4.2 Known Potential Benefits* *469*

*A3.5.1 Inclusion Criteria* *469*

*A3.5.2 Exclusion Criteria* *469*

*A3.6 Event Adjudication* *470*
A3.8 *Statistical Analyses* 470

*A3.8.1 Interim Analysis Schedule* *470*
A3.9 *Adaptive Decision Rules* 470
A3.10 *Operating Characteristics* 471

*A3.11 References* *471*

**Appendix 4: Prophylactic Dose Anticoagulation (Arm B)** **472**

A4.1 *Prophylactic Dose Anticoagulation** 472
A4.2 *Discontinuation of study intervention* 472
A4.4 *Potential Risks & Benefits* 474

*A4.4.1 Known Potential Risks* *474*

*A4.4.2 Known Potential Benefits* *474*

*A4.5.1 Inclusion Criteria* *474*

*A4.5.2 Exclusion Criteria* *474*

*A4.6 Event Adjudication* *475*

A4.8 *Statistical Analyses* 475

*A4.9 Number of Participants* *475*

*A4.10 References* *475*

**Appendix 5: ACTIV-4 Blood Sampling – proposed samples and times for sites participating in mechanistic studies and biorepository** **476**

*Blood collection times for inpatients:* 476
*Standard samples to be collected & volumes at each time point:* 476
A5.3 *Biorepository/Central Lab* 477

**Appendix 6. Additional data inclusion from other trials merged under activ-4 platform** **479**

**Adjudication Charters** **480**

**REMAP-CAP** **480**

Introduction **482**

Interaction with ATTACC and ACTIV-IV **482**

Endpoints for adjudication **482**

Secondary endpoint definitions **483**

*Domain-specific secondary endpoints* *483*
Major bleeding 483
Acute Myocardial Infarction (AMI) 483
Confirmed deep vein thrombosis 484
Confirmed pulmonary embolus 484
Confirmed ischemic cerebrovascular event 484
Mesenteric ischemia 484
Limb ischemia 485
Heparin induced thrombocytopenia (HIT) 485

*Derived endpoints* *485*
Systemic arterial thrombosis or embolism 486
Major thrombotic event 486
All thrombotic events 486

Data items to be used for adjudication **486**

Adjudication process **486**

*Data extraction* *486*

*Blinded Adjudication* *487*

*Additional data collection* *487*

*Resolution of discordant adjudication assessments* *488*

**ATTACC** **489**

**Clinical Events Committee (CEC) Charter** **489**

**ABBREVIATIONS** **494**

3.1 Primary efficacy endpoint 497
3.2 Secondary efficacy endpoint 497
7.1 CEC Chairperson 499
7.2 CEC Adjudicators 499
7.2.1 CEC Adjudicator Qualifications 500
7.2.2 CEC Adjudicator Membership 500
8.1 CEC Adjudicator Confidentiality 500
8.2 SOCAR CEC Coordination Team 500
8.3 Disbandment of CEC 501
9.1 Event Identification 501
9.1.1 Primary Method 501
9.1.2 Supplementary Methods 502
9.2 Multiple Events occurring in The Same Subject 502
9.3 Redaction 503
9.4 CEC Coordination Team Review 503
9.5 Dossier Creation and Event Assignment 504
9.6 Adjudication Timeline 505
9.7 CEC Adjudicator Review Process 506
9.7.1 Phase I Review 506
9.7.2 Phase II Review 509
9.7.3 Data Changes after the Event Adjudication 510
9.8 CEC Quality Assessment 510
9.8.1 Quality Assessment review and documentation 511
9.9 Committee Meetings 511
11.1 Administrative Documents 512
11.2 Documents for Events Adjudication 512
11.3 Document archival and study closure 512

***12.1*** *Deep Venous Thromboembolism* *513*

***12.2*** *Pulmonary Embolism* *514*

***12.3*** *Systemic arterial thrombosis or embolism* *515*

***12.4*** *Myocardial Infarction* *515*

***12.5*** *Stroke* *521*

***12.6*** *Major Bleeding* *524*

***12.7*** *Heparin-Induced Thrombocytopenia* *524*

*Appendix A: ATTACC Events Adjudication Committee Membership* *525*

*Appendix B: Conflict of Interest* *526*

*Appendix C: Source Documents Requested from Site, by Event Type* *527*

**ACTIV-4a** **530**

**Signature Page** **531**

**INTRODUCTION** **532**

**1.0. MISSION OF THE CLINICAL ENDPOINT COMMITTEE** **532**

**2.0. MEMBERS OF THE CLINICAL ENDPOINT COMMITTEE** **533**

**3.0. RESPONSIBILITIES LIST** **534**

**4.0. PROCEDURES OF THE CLINICAL ENDPOINT COMMITTEE** **535**

*5.1. Identification and Reporting of Clinical Events to the CEC* *535*

*5.1. Assembly of Clinical Dossiers for CEC Review* *535*

*5.2. CEC Adjudication Process* *536*

**5.0. CLINICAL EVENT DEFINITIONS** **537**

*6.1. Deep Venous Thromboembolism* *537*

*6.2. Pulmonary Embolism* *538*

*6.3. Arterial Thromboembolism* *538*

*6.4. Myocardial Infarction* *539*

*6.5. Stroke* *543*

*6.6. Major Bleeding* *545*

*Major bleeding is defined as acute clinically overt bleeding associated with one or more of the following (as per ISTH guidelines):[8, 9]* *545*

*6.7. Definitions of Cardiovascular, Non-cardiovascular, and Undetermined Cases of Death* *545*

Definitions of Cardiovascular, Non-Cardiovascular, and Undetermined Cases of Death 545

**7.0. REFERENCES** **547**

**8.0. EXPERT SIGNATURE PAGE** **549**

### Introduction

The pages that follow detail the three study protocols that comprise the multi-platform randomized controlled trial (mp-RCT) formed by the REMAP CAP, ACTIV4a, and ATTACC platforms. The mpRCT represents an unprecedented display of global collaboration whereby three organizationally distinct platforms harmonized protocols and worked together to answer the important question of whether a strategy of empiric therapeutic anticoagulation benefits patients with Covid-19 compared to usual care thromboprophylaxis.

The concept of the mp-RCT was borne out of discussion and shared interest among clinical trialists and leaders of three major international platforms to evaluate the efficacy of therapeutic anticoagulation for inpatients with Covid-19. To answer the question as quickly as possible and with maximal generalizability, three protocols were harmonized to create a single mpRCT with common eligibility criteria, study intervention details, and key primary and secondary outcomes. Data collection, leadership, and oversight are shared across the platforms, and agreement was established to federate the data collected within each platform into one overarching analysis. Trial data providers are SOCAR (ATTACC, ACTIV IV) and SPIRAL/UPMC (REMAP-CAP), and statisticians from Berry Consultants serve as trial statisticians for all three platforms with an independent group comprising the Statistical Analysis Committee (SAC) for the mpRCT. With agreed upon stopping rules for efficacy and futility for the primary endpoint, the mpRCT operates under a unified multiplatform analysis plan with monthly interim analyses (See attached Statistical Analysis Plan – Appendix 1 pages 21-98). The mpRCT investigators collaborate on making all major decisions.

Oversight was executed collaboratively across the three platforms, each with its own independent DSMB (See Pages 113-128 for detailed description of DSMB oversight). Each trial DSMB maintained its responsibilities within its platform. The interim efficacy and safety analyses were reported to the DSMBs for each of the trials. Since safety data are not aggregated across trials, safety analysis reports for each platform were shared with the DSMBs of other platforms. Upon viewing safety reports, each DMC reached a consensus on safety topics, and the DSMB chairs or designees from each of the platforms communicated with one another to discuss the efficacy and safety results across trials. Platform leadership and DSMB chairs agreed that individual platforms would not make public disclosures of efficacy without the agreement of all three oversight boards. A pre-defined publication plan was established by platform investigators for the mpRCT.

The results presented in the manuscript reflect the highly collaborative effort of the network of networks efficiently working together to rapidly answer a question of high public health importance. In effort to simplify the data review process, this protocol appendix includes individual protocol summaries, three tables that outline the harmonization across the three platforms, the unified mpRCT statistical analysis plan, and detailed versions of each individual platform protocol.

### Protocol Synopses

#### REMAP-CAP

| REMAP-CAP: COVID-19 Therapeutic Anticoagulation Domain Summary | |
|---|---|
| Interventions | • Local standard venous thromboprophylaxis<br>• Therapeutic anticoagulation with intravenous unfractionated heparin or subcutaneous low molecular weight heparin |
| Unit of Analysis, Strata, and State | This domain is analyzed only in the pandemic statistical model.<br><br>The pandemic statistical model includes only patients who are in the Pandemic Infection Suspected or Proven (PISOP) stratum. Within this stratum, the unit-of-analysis is defined by illness severity state at time of enrollment, defined as either Moderate State or Severe State. Unit-of-analysis may also be defined by SARS-CoV-2 infection or d-dimer strata or both. Borrowing is permitted between states and strata. If the SARS-CoV-2 strata is applied in analysis, Response Adaptive Randomization will be applied to all PISOP patients, in each illness severity state, using probabilities derived from the SARS-CoV-2 confirmed stratum. Response Adaptive Randomization may also be applied according to D-dimer strata status. |
| Evaluable treatment-by-treatment Interactions | No interaction will be evaluated with any other domain of REMAP-CAP. |
| Nesting | None |
| Timing of Reveal | Randomization with Immediate Reveal and Initiation or Randomization with Deferred Reveal if prospective agreement to participate is required. |
| Inclusions | Patients will be eligible for this domain if:<br>• COVID-19 infection is suspected by the treating clinician or has been confirmed by microbiological testing<br>• Microbiological testing for SARS-CoV-2 infection of upper or lower respiratory tract secretions or both has occurred or is intended to occur |
| Domain-Specific Exclusions | Patients will be excluded from this domain if they have any of the following:<br>• More than 48 hours has elapsed since ICU admission (noting that this may be operationalized as more than 48 hours has elapsed since commencement of organ failure support)<br>• Clinical or laboratory bleeding risk or both that is sufficient to contraindicate therapeutic anticoagulation, including intention to continue or commence dual anti-platelet therapy<br>• Therapeutic anticoagulation is already present due to prior administration of any anticoagulant agent that is known or likely to still be active or a clinical decision has been made to commence therapeutic anticoagulation<br>• Enrolment in a trial evaluating anticoagulation for proven or suspected COVID-19 infection, where the protocol of that trial requires continuation of the treatment assignment specified in that trial<br>• Known or suspected previous adverse reaction to UFH or LMWH including heparin induced thrombocytopenia (HIT).<br>• The treating clinician believes that participation in the domain would not be in the best interests of the patient |
| Intervention-Specific Exclusions | None |

#### ATTACC

| | |
|---|---|
| Study Title: | **A**nti**T**hrombotic **T**herapy to **A**meliorate **C**omplications of **COVID**-19 (**ATTACC**) in collaboration with Accelerating COVID-19 Therapeutic Interventions and Vaccines (ACTIV-4) |
| Study Design: | A phase III prospective, open-label, adaptive multi- platform randomized controlled trial |
| Primary Objective/ Endpoint: | The primary endpoint in the trial is days alive and free of organ support at day 21. This endpoint is defined as the number of days that a patient is alive and free of organ support through the first 21 days after trial entry. |
| Secondary Objectives: | Secondary Safety Endpoints:<br>- Laboratory confirmed heparin induced thrombocytopenia **(HIT)**<br>- **Major bleeding**, defined according to the International Society on Thrombosis and Haemostasis (ISTH)/Scientific and Standardization Committee (SSC) definitions and bleeding assessment tool in non-surgical patients (Schulman *J Thromb Haemost* 2005): fatal bleeding; and/or symptomatic bleeding in a critical area or organ, such as intracranial, intraspinal, intraocular, retroperitoneal, intraarticular or pericardial, or intramuscular with compartment syndrome; and/or bleeding causing a fall in hemoglobin level of ≥20 g/L, or leading to transfusion of 2 or more units of whole blood or red cells.<br><br>Secondary Efficacy Endpoints:<br>- A composite endpoint of death, deep vein thrombosis, pulmonary embolism, systemic arterial thromboembolism, myocardial infarction, or ischemic stroke collected during hospitalization or at 28 days and 90 days after enrollment (whichever is earlier)<br>- Ordered categorical endpoint with three possible outcomes based on the worst status of each patient through day 30 following randomization: no invasive mechanical ventilation, invasive mechanical ventilation, or death. |

| | |
|---|---|
| | - All cause mortality assessed at 28 and 90 days following randomization<br>- All cause mortality during initial hospitalization (includes death after 28 days)<br>- Intubation assessed at 30 days following randomization<br>- Ventilator-free days (days alive not on a ventilator) assessed at 28 days following randomization<br>- Hospital-free days (days alive outside hospital assessed at 28 days following randomization)<br>- Vasopressor-free days (days alive not on a vasopressor) assessed at 28 days following randomization<br>- Renal replacement free days (days alive not on renal replacement) assessed at 28 days following randomization<br>- Hospital re-admission within 28 days<br>- Symptomatic proximal venous thromboembolism (DVT or PE) assessed at 28 and 90 days following randomization<br>- Myocardial infarction assessed at 28 and 90 days following randomization<br>- Ischaemic stroke assessed at 28 and 90 days following randomization<br>- Acute kidney injury as defined by KDIGO criteria<br>- Systemic arterial thrombosis or embolism assessed at 28 and 90 days following randomization<br>- Use of extracorporeal membrane oxygenation (ECMO) support<br>- Mechanical circuit (dialysis or ECMO) thrombosis<br>- WHO ordinal scale (peak scale over 28 days, scale at 14 days, and proportion with improvement by at least 2 categories compared to enrollment, at 28 days) |
| Duration: | The duration of accrual on this study will be ongoing in nature during the COVID-19 pandemic, following outcomes for each patient up to a maximum of 90 days. |

| | |
|---|---|
| Planned Total Sample Size: | The trial is a Bayesian adaptive design and as such is not predicated on a fixed *a priori* sample size. This design was chosen given uncertainty regarding anticipated event rates and potential treatment effect sizes. Approximately 350 to a maximum of 3000 evaluable patients are anticipated to be enrolled in this adaptive trial in combination with the ACTIV 4 and REMAP-CAP trials, with anticipated reprioritization of key subgroups (including D-dimer defined) as the trial is undertaken. |
| Drug Administration: | Participants randomized to the <u>investigational arm</u> will receive therapeutic anticoagulation for 14 days (or until hospital discharge or liberation from the need for supplemental oxygen, whichever comes first) with preference for low-molecular weight heparin (LMWH), or alternative unfractionated heparin (UFH).<br><br>Participants randomized to the <u>control arm</u> will receive usual care, which is anticipated to include thromboprophylactic dose anticoagulation according to local practice. |
| Inclusion/Exclusion Criteria: | Inclusions:<br>1. Patients ≥18 years of age providing (possibly through a substitute decision maker) informed consent who require hospitalization anticipated to last ≥72 hours, for microbiologically-confirmed COVID-19, enrolled < 72 hours of hospital admission **or** of COVID-19 confirmation<br><br>Exclusions:<br>1. Requirement for chronic mechanical ventilation via tracheostomy prior to hospitalization<br>2. Patients for whom the intent is to not use pharmacologic thromboprophylaxis<br>3. Active bleeding<br>4. Risk factors for bleeding, including:<br>a. intracranial surgery or stroke within 3 months;<br>b. history of intracerebral arteriovenous malformation;<br>c. cerebral aneurysm or mass lesions of the central nervous system; |

| | |
|---|---|
| | d. intracranial malignancy<br>e. history of intracranial bleeding<br>f. history of bleeding diatheses (e.g., hemophilia)<br>g. history of gastrointestinal bleeding within previous 3 months<br>h. thrombolysis within the previous 7 days<br>i. presence of an epidural or spinal catheter<br>j. recent major surgery <14 days<br>k. uncontrolled hypertension (sBP >200 mmHg, dBP >120 mmHg)<br>l. other physician-perceived contraindications to anticoagulation<br>5. Platelet count <50 x$10^9$/L, INR >2.0, or baseline aPTT >50<br>6. Hemoglobin <80 g/L (to minimize the likelihood of requiring red blood cell transfusion if potential bleeding were to occur)<br>7. Acute or subacute bacterial endocarditis<br>8. History of heparin induced thrombocytopenia (HIT) or other heparin allergy including hypersensitivity<br>9. Current use of dual antiplatelet therapy<br>10. Patients with an independent indication for therapeutic anticoagulation<br>11. Patients in whom imminent demise is anticipated and there is no commitment to active ongoing intervention<br>12. Anticipated transfer to another hospital that is not a study site within 72 hours<br>13. Enrollment in other trials related to anticoagulation or antiplatelet therapy |
| Study Assessments: | Study assessments are depicted in the study schedule. |
| Safety Variables & Analysis: | The safety of therapeutic anticoagulation with LWMH or intravenous UFH infusion will be evaluated by AE reports. Treatment-related AEs include bleeding and HIT. |
| Efficacy Assessments & Analysis | The efficacy of therapeutic-dose parenteral anticoagulation with subcutaneous LMWH or |

| | |
|---|---|
| | intravenous UFH will be evaluated in comparison to usual care. |
| Reasons for premature discontinuation of therapy: | Treatment will continue until any of the following occurs:<br>• Heparin induced thrombocytopenia or other heparin allergy/hypersensitivity<br>• Thrombocytopenia (platelet count <50 x$10^9$/L)<br>• Major bleeding, defined based closely on the International Society on Thrombosis and Haemostasis (ISTH)/Scientific and Standardization Committee (SSC) definitions and bleeding assessment tool in non-surgical patients<br>• Coagulopathy associated with an elevated INR (e.g., >2.0) or hypofibrinogemia<br>• Following invasive procedures where heparin is deemed unsafe to re-institute<br>• Patients requiring systemic fibrinolytic therapy<br>• Treating physician discretion |
| Statistical Analysis: | Data will be analyzed by an intention to treat analysis for the primary analysis; a per-protocol analysis will also be completed as a secondary analysis. Patients who receive at least one dose of drug will be evaluable for safety and efficacy. Response-adaptive randomization based on D-dimer subgroups is embedded. |

#### ACTIV-4a

| | |
|---|---|
| Title | A Multicenter, Adaptive, Randomized, Open Label Controlled Platform Trial of the Safety and Efficacy of Antithrombotic Strategies in Hospitalized Adults with COVID-19 |
| Short Title | ACTIV-4 ACUTE |
| Brief Summary | This is a randomized, open label, adaptive platform trial to compare the effectiveness of antithrombotic strategies for prevention of adverse outcomes in COVID-19 positive inpatients |
| Objectives | 1. To determine the most effective antithrombotic strategy for increasing the number of days free of organ support and reducing death.<br><br>2. To determine the most effective antithrombotic strategy on the composite endpoint of death, deep vein thrombosis (DVT), pulmonary embolism (PE), myocardial infarction (MI), ischemic stroke, or other systemic arterial thrombosis (AT).<br><br>3. To assess the safety of antithrombotic strategies through the endpoint of major bleeding as defined by ISTH.<br><br>4. To compare the effect of antithrombotic strategies on the endpoint of all-cause mortality in the study population.<br><br>Assessment of efficacy and safety will yield information of the net clinical benefit of different antithrombotic strategies in the study population. It will also yield information on outcomes specific to under-represented minority populations, specifically African- and Hispanic- descent persons. |
| Methodology | Adaptive Randomized Platform Trial |

| | |
|---|---|
| Endpoints | Primary Endpoint: 21 Day Organ Support Free Days, which is defined as the number of days that a patient is alive and free of organ support through the first 21 days after trial entry. Organ Support is defined as receipt of invasive or non-invasive mechanical ventilation, high flow nasal oxygen, vasopressor therapy, or ECMO support, with death at any time (including beyond 21 days) during the index hospitalization assigned -1 days.<br><br>Key Secondary Endpoint: Composite endpoint of death, pulmonary embolism, systemic arterial thromboembolism, myocardial infarction, or ischemic stroke at hospital discharge or 28 days, whichever occurs first.<br><br>Other Secondary Endpoints: Composite endpoint of death, deep vein thrombosis, pulmonary embolism, systemic arterial thromboembolism, myocardial infarction, or ischemic stroke at hospital discharge or 28 days, whichever occurs first. Acute kidney injury defined by KDIGO criteria, Individual endpoints comprising the key secondary endpoint, death during hospitalization, 28 Day Ventilator-Free Days, 28 Day Vasopressor Free Days, 28 Day Renal Replacement Free Days, WHO clinical scale, 28 Day Hospital Free Days, 28 day organ support free days, and all-cause mortality at 90 days.<br><br>Primary Safety Endpoint: Major bleeding (as defined by the ISTH) Secondary Safety Endpoint: Confirmed heparin induced thrombocytopenia (HIT) |
| Study Duration | Approximately 1 year |
| Participant Duration | Hospital duration with periodic contact at post-discharge, including at 90 days, with potential contact up to 1 year |
| Duration of assigned treatment strategy | During hospitalization (unless otherwise specified in description of arm) |
| Population | Adult patients hospitalized for COVID-19 |
| Study Sites | Approximately 150 sites |
| Number of participants | The sample size is described in each arm-specific appendix. |
| Description of Study Agents | Randomized arms- see appendix<br><br>This platform trial allows for multiple therapies to be investigated in this trial over time. The trial is governed by a Master Protocol that describes the trial design, endpoint collection, primary endpoint, and inclusion/exclusion criteria. Different therapies, referred to as arms, are detailed in arm-specific appendices. These arm-specific appendices work in a modular fashion as arms are removed and added to the platform trial. |
| Key Procedures | Observation during hospitalization, contact at 90 days post-enrollment, and collection of standard of care laboratory results. Ancillary biobanking will be completed in consenting patients at capable centers. |

| | |
|---|---|
| Statistical Analysis | Inferences in this trial are based on a Bayesian statistical model, which considers the variation in outcomes by site, disease state, time, and arm of the trial. The specific analyses for each arm, including interim<br>analysis schedule, are specified in each arm-specific appendix. |

### Cross-Platform Protocol Comparison Tables

#### Eligibility Criteria

| | **REMAP-CAP** | **ACTIV-4a** | **ATTACC** |
|---|---|---|---|
| *Inclusion Criteria* | | | |
| Age | • Adult (age not specifically listed) | • ≥ 18 years of age | |
| Duration of hospitalization for COVID-19 | • Expected hospital LOS > 48 hours (i.e. not expected to be discharged today or tomorrow) | • Expected hospital LOS ≥ 72 hours | |
| SARS-CoV-2 infection | • Suspected or confirmed with intent to test for COVID-19* | • Confirmed | |
| Enrollment Window | • Unspecified | • <72 hours from admission OR COVID-19 confirmation | |

| | | | |
|---|---|---|---|
| *Exclusion Criteria* | | | |
| Platelet Count | | • < 50x $10^9$/L | • <50 x$10^9$/L, INR >2.0, or baseline aPTT >50 seconds |
| Hemoglobin | | • Hemoglobin <80 g/L (8 g/dL) (to minimize the likelihood of requiring red blood cell transfusion if potential bleeding were to occur) | |
| Heparin Induced Thrombocytopenia (HIT) | • Known or suspected previous adverse reaction to unfractionated heparin or low molecular weight heparin including HIT | • History of heparin-induced thrombocytopenia (HIT) or other heparin allergy including hypersensitivity | |
| Dual Antiplatelet Therapy | • Intention to continue or commence dual antiplatelet therapy | • Patient on dual antiplatelet therapy, when one of the agents cannot be stopped safely | • Current use of dual antiplatelet therapy |
| Mechanical Ventilation | | • Chronic mechanical ventilation via tracheostomy prior to hospitalization | |
| Prognosis | • Death is deemed to be imminent and inevitable during the next 24 hours AND<br>• One or more of the patient, substitute decision maker, or attending physician are not committed to full active treatment | • Imminent death | • Patients in whom imminent demise is anticipated and there is no commitment to active ongoing intervention |
| Co-Enrollment | • Enrollment in a trial evaluating anticoagulation for proven or suspected COVID-19 infection, where the protocol of that trial requires continuation of the treatment assignment specified in that trial | • Co-enrollment in other trials is permitted as long as the other trial does not test agents with antithrombotic properties and there | • Enrollment in other trials related to anticoagulation or antiplatelet therapy |

| | | | |
|---|---|---|---|
| | | is no other scientific contraindication | |
| Bleeding Risk | • Clinical and/or laboratory bleeding risk or both that is sufficient to contraindicate therapeutic anticoagulation | Contraindication to anticoagulation, including but not limited to:<br>• known bleeding within the last 30 days requiring emergency room presentation or hospitalization<br>• known history of an inherited or active acquired bleeding disorder<br>• recent ischemic stroke | • Intracranial surgery or stroke within 3 months<br>• history of intracerebral arteriovenous malformation<br>• cerebral aneurysm or mass lesions of the central nervous system<br>• intracranial malignancy<br>• history of intracranial bleeding<br>• history of bleeding diatheses (e.g., hemophilia)<br>• history of gastrointestinal bleeding within previous 3 months<br>• thrombolysis within the previous 7 days<br>• presence of an epidural or spinal catheter<br>• recent major surgery <14 days<br>• uncontrolled hypertension (sBP >200 mmHg, dBP >120 mmHg)<br>• other physician-perceived contraindications to anticoagulation |
| Miscellaneous | • Treating physician does not feel trial participation is in the best interest of the patient | • Pregnancy | |

#### Interventions

| | REMAP-CAP | ACTIV-4a | ATTACC |
|---|---|---|---|
| *Intervention arm management* | | | |
| Anticoagulant drug | • Unfractionated heparin or low molecular weight heparin<br>• Patients may be switched between unfractionated heparin and low molecular weight heparin | • Unfractionated heparin or low molecular weight heparin<br>• Patients may be switched between unfractionated heparin and low molecular weight heparin<br>• Patients with impaired renal function were stipulated to received unfractionated heparin | • Unfractionated heparin or low molecular weight heparin<br>• Either agent permitted and patients may be switched between unfractionated heparin and low molecular weight heparin |
| Dose | • Dosed according to local hospital policy, practice, and guidelines for treatment of venous thromboembolism<br>• For UFH, suggested target for aPTT of 1.5 to 2.5 times the upper limit of normal or therapeutic anti-Xa levels<br>• Low molecular weight heparin dosed according to patient weight | • Low molecular weight heparin dosed according to patient weight and creatinine clearance<br>• For UFH, suggested target of anti-Xa of 0.3-0.7 IU/ml or aPTT 1.5 to 2.5 times the upper limit of normal | • Low molecular weight heparin dosed according to patient weight and creatinine clearance according to local practice and policy<br>• For UFH, suggested target of aPTT 1.5 to 2.5 times the upper limit of normal or therapeutic anti-Xa levels |
| Duration of intervention | • Up to 14 days or to hospital discharge, whichever comes first<br>• For ICU patients, therapeutic anticoagulation could be discontinued at ICU discharge | • Up to 14 days or to hospital discharge, whichever comes first | • Up to 14 days or until hospital discharge or recovery (defined as liberation from supplemental oxygen>24 hours, provided oxygen was required), whichever comes first |
| *Usual care arm management* | | | |
| Thromboprophylaxis agent | • Standard venous thromboprophylaxis according to local guidelines or usual practice | • Any one of enoxaparin, dalteparin, tinzaparin, fondaparinux, or heparin according to local preference | • Standard venous thromboprophylaxis according to local guidelines or usual practice |
| Thromboprophylaxis dose | • Dose of chosen agent should not be sufficient | • Dose of agent specified to be | • Dose of chosen agent should not be more |

| | | | |
|---|---|---|---|
| | to result in therapeutic anticoagulation | consistent with guidelines for low dose thromboprophylaxis | than half of the approved therapeutic dose for the treatment of venous thromboembolism |
| Duration of thromboprophylaxis | • Up to 14 days or hospital discharge, whichever comes first<br>• After this period, decisions regarding thromboprophylaxis are at discretion of treating clinician | • Up to 14 days or hospital discharge, whichever comes first<br>• After this period, decisions regarding thromboprophylaxis are at discretion of treating clinician | • Up to 14 days or hospital discharge, whichever comes first<br>• After this period, decisions regarding thromboprophylaxis are at discretion of treating clinician |

#### Endpoints

| | REMAP-CAP | ACTIV-4a | ATTACC |
|---|---|---|---|
| Primary endpoint | • Days alive and free of organ support to day 21<br>• An ordered categorical endpoint ranging between 0 and 21; patients who die at any time in hospital are assigned a value of -1 | | |
| Secondary efficacy endpoints | • All-cause mortality at 90 days<br>• Hospital length-of-stay censored at day 90 | • All-cause mortality at 28 days<br>• All-cause mortality in hospital<br>• Hospital re-admission within 28 days<br>• Hospital length-of-stay<br>• WHO ordinal scale at 14 days, proportion with improvement by at least 2 levels compared to enrolment at 28 days | • All-cause mortality at 28 days and 90 days<br>• Hospital re-admission within 28 days<br>• WHO ordinal scale at 14 days, proportion with improvement by at least 2 levels compared to enrolment at 28 days<br>• Intubation at day 30<br>• Ordered categorical endpoint with three possible outcomes based on the worst status of each patient through day 30 (not invasively ventilated, invasively ventilated, or death) |
| Secondary ICU outcomes | • ICU readmission<br>• ICU mortality censored at day 90<br>• ICU length-of-stay censored at day 90<br>• Ventilator-free days at 28 days<br>• Tracheostomy censored at 28 days | • Ventilator-free days at 28 days<br>• Hospital-free days at 28 days<br>• Vasopressor-free days at 28 days<br>• Renal replacement-free days at 28 days<br>• Use of ECMO in hospital | |
| Secondary thrombosis endpoints | • Confirmed deep venous thrombosis in hospital<br>• Confirmed pulmonary embolism<br>• Confirmed ischemic cerebrovascular event<br>• Confirmed acute myocardial infarction<br>• Other thrombotic event including mesenteric ischemia and limb ischemia<br>• Peak troponin between randomization to day 15 | • Major thrombotic events (death, deep vein thrombosis, pulmonary embolism, systemic arterial thromboembolism, myocardial infarction, or ischemic stroke) at 28 days<br>• Symptomatic proximal venous thromboembolism (DVT or PE) assessed at 28 days<br>• Myocardial infarction at 28 days<br>• Ischemic stroke at 28 days<br>• Acute kidney injury as defined by KDIGO criteria<br>• Systemic arterial thrombosis or embolism at 28 days<br>• Mechanical circuit (dialysis or ECMO) thrombosis | |
| Safety endpoints | • Major bleeding, defined according to International Society on Thrombosis and Haemostasis (ISTH) definitions | | |

| | |
|---|---|
| | • Fatal bleeding<br>• Symptomatic bleeding in a critical area or organ<br>• Bleeding causing a fall in hemoglobin level of ≥20 g/L<br>• Requiring a transfusion of 2 or more units of whole blood or red cells<br>• Laboratory-confirmed heparin-induced thrombocytopenia |

CONFIDENTIAL

### Multi-platform Randomized-Controlled Trial (mpRCT) Analysis Plan for ACTIV-4, ATTACC, and REMAP-CAP (AARC)

**Version 1.0**
**September 17, 2020**

**Background**
Three operationally distinct trial platforms – ACTIV-4, ATTACC, and REMAP-CAP (*i.e.,* AARC) – are undertaking a jointly-developed, single multiplatform randomized controlled trial (mpRCT) evaluating the efficacy of antithrombotic strategies in patients hospitalized with COVID-19. Each of these platforms is randomizing a control arm (prophylactic-dose anticoagulation) against a therapeutic-dose anticoagulation through a common mpRCT. This document describes the agreed analytic plan across the three platforms for the question of efficacy related to therapeutic-dose anticoagulation.

**Operational Separation**
Each of the three platforms will operate individually but in coordination. The three trials will use closely harmonized protocols, and overlapping data collection, leadership, and oversight approaches. The decisions each platform makes based on the efficacy analyses may vary, but they agree on the joint efficacy analysis plan and will collaborate on making major decisions.

**Inferential Synergy**
The three platforms have agreed to prospectively federate patient-level data together to form a single mpRCT to evaluate the efficacy and safety of therapeutic-dose anticoagulation. This document describes the agreed analytic plan and an overview of the analysis implementation for the anticoagulation efficacy.

**Interim Analysis Strategy**
Interim analyses for anticoagulation efficacy will be conducted monthly with the first interim occurring upon finalization of this plan. At least 100 must be randomized across AARC between interims or the monthly interim will be skipped. The interim analyses will commence the first working day of each month. This will determine the time at which data are transferred for the interim analysis.

External events, such as other trials reporting positive/negative results of similar questions, may also trigger an analysis of the anticoagulation efficacy results. The same procedures, models, and analyses will take place, but at times dictated by the trials' leadership.

**Implementation Plan**
The efficacy analyses will be conducted by a Statistical Analysis Committee (SAC). The SAC is an independent group chartered to conduct interim analyses. The SAC members are unblinded to trial results. At the time of an interim analysis, each trial platform will send the necessary data for the interim analysis to the SAC. The format for this data delivery and the specifications for the data file necessary for the interim analyses will be set by the SAC.

Each platform is responsible for the monitoring and quality of their data. The SAC receives this data and performs appropriate data investigation but does not change data nor have the responsibility to verify data or perform data cleaning or monitoring.

The SAC will conduct the efficacy analyses (described below) and will report the results of the interim analyses to the DSMBs for each of the trials (Figure 1). The DSMBs are then responsible for the communication to each of the platforms' leadership/steering committees. The three platforms agree to jointly present and publicly disclose the outcome of the anticoagulation results.

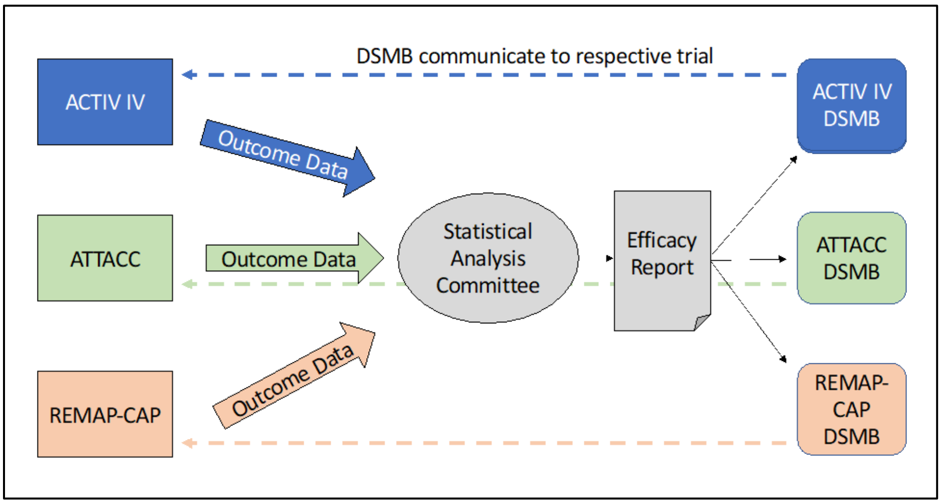

Figure 1: The communication plan across trials for the interim analyses

**Response Adaptive Randomization**

The REMAP-CAP and ATTACC trials utilize response adaptive randomization. At the interim analyses the response adaptive randomization proportions for the respective trials will be provided to their randomization services for their continued uses of response adaptive randomization. The ACTIV-4 trial is not currently utilizing response adaptive randomization.

The model run for REMAP-CAP for the response adaptive randomization will be different than the model run for the joint efficacy analysis. The REMAP-CAP model is conducted on all randomized patients within the REMAP-CAP trial, accounting for other interventions, to evaluate potential platform conclusions and set the randomization proportions. This *larger model* is not used to trigger success or futility, as the analysis detailed in this report is jointly used and agree upon to label superiority and futility of the anticoagulation domain. The RAR for REMAP-CAP will be updated at a REMAP-CAP specific interim.

The adaptive randomization proportions for the ATTACC platform will vary by d-dimer group and will be based on the mpRCT analysis outlined below.

**Joint Efficacy Analysis Plan**

**Arms**
Control Arm: The prophylactic-dose anticoagulation arm.
Treatment Arm: The therapeutic-dose anticoagulation arm.

**Analysis Population**
The joint efficacy analyses will be conducted on the joint anticoagulation PCR positive populations. This includes each patient who was randomized to either the control arm or the treatment arm who had one or more PCR positive test results.

If new arms are added to this joint analysis, this definition would include patients randomized to that additional arm. For clarity: REMAP-CAP is enrolling patients to other domains; a patient randomized to other domains but not the anticoagulation domain would not be included in this analysis. The alternative randomization to other arms will not be included in this joint analysis of efficacy for anticoagulation.

**Primary Endpoint**
The primary endpoint is an ordinal scale that is a composite of in-hospital mortality and organ support-free days (OSFD) through 21 days. Organ support is defined as delivery of one or more of the following while admitted to an ICU:

1. Receipt of invasive or non-invasive mechanical ventilatory support
2. High-flow nasal oxygen
3. Vasopressor therapy

The time on organ support is defined as the time from the start of organ support in ICU to the last use of organ support (intervening time off organ support between the first and last use are considered on organ support) during any and each ICU admission in the 21 days after randomization. Time out of ICU prior to the end of study day 21 is counted as organ support-free. The organ support "free-days" is defined as 21 days minus the time on organ support in ICU. If a patient dies in their acute hospital stay, then they are assigned the worst outcome value of –1. For patients that survive the hospital stay, they are assigned the number of organ support-free days, rounded to the nearest integer number of days. The best possible outcome of 22 is reserved for patients who survive and never have organ support in ICU. There are 24 possible outcomes for this ordinal outcome: all integers from – 1 to 22. Higher values correspond to better patient outcomes.

**Patient Subtypes**
There are four patient subgroups – denoted *subtypes* – that have separate inferences for anticoagulation in this analysis. The subtypes are based on a cross classification of the baseline disease state severity (receiving organ support with ICU level of care vs not) and for those not receiving ICU organ support, the baseline d-dimer group, defined as low or high. Patients missing d-dimer level will be included in a separate subtype that will help inform the overall model through the Bayesian hierarchical structure, but no adaptive triggers for superiority or futility will be actionable for this subtype. For notation purposes we label the disease severity state, as s=1, for patients not receiving organ support in ICU at baseline,

and s=2 for patients receiving organ support in ICU at baseline. For s=1, we have three d-dimer levels, labeled, d=1 (d-dimer low), d=2 (d-dimer high), and d=3 (d-dimer is missing). We refer to these using the s:d notation, where the 4 subtypes are 1:1, 1:2, 1:3, and 2:0 (we use the d-dimer notation d=0 for the organ support in ICU disease state). We use the term "severe" for the patients receiving organ support in ICU at baseline and "moderate" for the patients not receiving organ support eg ICU level of care at baseline.

**Adaptive Decision Rules**

The following adaptive decision rules have been specified as labels for the efficacy outcomes. These rules are in place to determine the statistical triggers to label the treatment arm as superior to the control arm or to label the treatment arm as futile on the primary endpoint compared to the control arm. Each rule is separately evaluated within the three subtypes: moderate state: low d-dimer, moderate state: high d-dimer, and severe state. Efficacy conclusions will not be made for the subgroup of moderate patients with missing d-dimer levels.

1. **Superiority**

If the posterior probability of superiority within a subtype is greater than 99% then the treatment arm will be labeled as superior to the control arm for OSFDs.

2. **Futility**

If the posterior probability of at least an odds-ratio benefit of 1.2 is less than 5% then the treatment arm will be considered futile compared to the control arm.

**Modeling of the Primary Endpoint**

Let $Y_i = \{-1, 0, 1, \ldots, 21, 22\}$ denote the ordinal outcome (OSFD) for patient $i$. The probability of patient $i$, in state $s$, observing $y$ OSFD or less is denoted as $\pi_{isy} = \Pr(Y_i \leq y)$. The model is a proportional odds model, where the log odds-ratio parameters in the model are structured so that a value > 0 implies treatment benefit, and an odds-ratio > 1 implies treatment benefit. The primary analysis model is formulated as follows:

$$\log\left(\frac{\pi_{isy}}{1 - \pi_{isy}}\right) = \alpha_{y,s} - \left[\gamma_P + \nu_{Site,s} + \lambda_{Time,s} + \theta_{a,s:d} + \beta_{Age,s} + \beta_{Sex,s} + \beta_d\right]$$

1. Each "platform," *P*, has a covariate adjustment in the model, *P*=1 is REMAP-CAP, *P*=2 is ATTACC, and *P*=3 is ACTIV-4.
2. The "site" variable is the clinical site within the trial. These will be set as distinct sites across all three trials. The site effects are estimated separately within the severe and moderate disease states, but do not vary by d-dimer level.
3. The "time" variable is an indicator of the time epoch in which a patient was enrolled in the trial, numbered increasing from the first most recent epoch to the earliest time for the analysis. Time epochs are two-week time periods, Time=1, and then every 2-week epoch, moving back in time throughout the enrollment in the trial, Time=2,3,4,…. The first two time periods are combined to form a 4-week category that serves as the referent. The time effects are modeled separately within the severe and moderate disease states, but do not vary by d-dimer level. The time epochs are defined to be the same time periods for both moderate and severe.

4. The "arm" to which a patient is randomized is labeled as *a* where *a*=1 is the control and *a*=2 is the treatment arm. The effects of arm vary by the disease state and the d-dimer level. The treatment effects for arm *a* within subtype *s*:*d* are modeled with the $\theta_{a,s:d}$ parameters.
5. The "age" variable is a categorical classification of age as ≤ 39, 40-49, 50-59, 60-69, 70-79, and 80+. The age effects will be estimated separately within the moderate and severe disease states, but do not vary by d-dimer level.
6. The "sex" variable is sex at birth. The sex effects will be estimated separately within the moderate and severe disease states, but do not vary by d-dimer level.
7. The additive effects of d-dimer levels are modeled with the $\beta_d$ for d = 0, 1, 2, 3.
8. The $\alpha_{y,s}$ parameters determine the baseline rates of the ordinal outcome, which are modeled separately by disease state but not d-dimer level.

**Model Priors**

The ordinal endpoint rates are modeled using a Dirichlet prior where the individual probabilities for the 24 outcomes are based on one patient's weight on real-world evidence-based outcomes (details of the vector *P* in the Anticoagulation Domain Current State).

$$logit(\alpha_{y,s}) \sim Dirichlet(P)$$

For identifiability, the platform effect for ATTACC is defined such that $\gamma_2 \equiv 0$. The platform effects, $\gamma_P,\ P = 1, 3$, are modeled with independent N(0, 1) priors.

The site effects, $\nu_{Site}$, are modeled using a hierarchical model where site is nested within the country of the site:

$$\nu_{Site,s} \sim N(\mu_{country,s}, \tau^2_{country,s}), site = 1, \ldots, N_{Site}$$

$$\mu_{country,s} \sim N(0,1);\ \tau^2_{country,s} \sim IG(0.25, 0.1), s = 1,2$$

A referent country, the United States, will be set such that $\mu_{country,1} \equiv \mu_{country,2} \equiv 0$. The hyper-parameters of the site hierarchical model vary by disease state *s*.

The effect of age category is modeled with independent, normally distributed priors for each disease state. The prior mean for each age category effect is based on emerging data from external sources (specific details for the age prior in the Anticoagulation Domain Current State).

The effect of time (*Time=T*) is modeled using a second-order normal dynamic linear model separately by disease state, *s*. The two most recent time epochs are modeled as the referent time epochs with the time parameters set to 0. The preceding time epochs are modeled as a normal dynamic linear model as:

$$\lambda_1 = \lambda_2 \equiv 0$$

$$\lambda_3 \sim N(0, 0.15^2)$$

$$\lambda_T - 2\lambda_{T-1} + \lambda_{T-2} \sim N(0, \tau_{Time}^2), T \geq 4$$

The treatment effect parameters are relative to a control arm, which will be labeled in the arm-specific appendix. The treatment effect for the control arm, labeled as arm *a* = 1, will be set to 0 for each of the disease subtypes and d-dimer level:

$$\theta_{1,s:d} \equiv 0$$

The effects of the treatment arm are modeled hierarchically across disease subtype and d-dimer level. The treatment effects for each arm, *a*, are modeled as:

$$\theta_{a,2:0} \sim N(\mu_{a,1}, \tau_{a,1}^2),$$

$$\theta_{a,1} \sim N(\mu_{a,1}, \tau_{a,1}^2),$$

$$\theta_{a,s:d} \sim N(\theta_{a,1}, \tau_{a,2}^2), \qquad s:d = 1:1, 1:2, 1:3,$$

$$\mu_{a,1} \sim N(0, 1);\ \tau_{a,1}^2 \sim IG(0.25, 0.1),$$

$$\tau_{a,2}^2 \sim IG(0.25, 0.0025)$$

The effect of d-dimer is modeled with a N(0, 1) prior for d = 2,3, and the parameters $\beta_0$ and $\beta_1$ are set to 0 for identifiability. Any additional covariates included in the model will have independent N(0, 1) priors unless otherwise specified.

### Statistical Analysis Plan for the Multi-Platform Randomized Clinical Trial of Therapeutic Anticoagulation for COVID-19

#### Version history

Version 1: Initiated December 21, 2020; Finalized January 5, 2021

#### SAP Authors

Ewan Goligher, University of Toronto, Toronto, CA

Patrick Lawler, University of Toronto, Toronto, CA

Scott Berry, Berry Consultants, Austin, TX, USA

Judith Hochman, NYU Grossman School of Medicine/NYU Langone Health, New York, NY

Matthew D. Neal, University of Pittsburgh, PA, USA

Bridget-Anne Kirwan, Socar Research, Nyon, Switzerland

Lindsay Berry, Berry Consultants, Austin, TX, USA

Elizabeth Lorenzi, Berry Consultants, Austin, TX, USA

Steven Webb, Monash University, Melbourne, Victoria, Australia

Colin McArthur, University of Auckland, Auckland, NZ

Derek C. Angus, University of Pittsburgh and UPMC Health System, PA, USA

Charlotte Bradbury, University Hospitals Bristol, Bristol, UK

Michael Farkouh, University of Toronto, Toronto, CA

Bryan McVerry, University of Pittsburgh and UPMC Health System, PA, USA

Jeff Berger, NYU Grossman School of Medicine/NYU Langone Health, NY, NY, USA

Michelle Ng Gong, Albert Einstein College of Medicine, NY, NY. USA

Anthony Gordon, Imperial College, London, UK

Ryan Zarychanski, University of Manitoba, Winnipeg, CA

#### SAP Introduction

This is the statistical plan for the analysis of the Covid-19 Therapeutic Anticoagulation Multi-Platform RCT (mpRCT). This plan has been prespecified by the investigators prior to unblinding of the data for the Severe state of the mpRCT.

##### mpRCT status at time of SAP preparation

Enrollment in the severe state was halted across platforms on December 18, 2020. The severe state-specific conclusion was publicly disclosed on December 22, 2020. The predefined statistical trigger for futility was met with data available to the Statistical Analytic Committee (SAC), and hence the results for the severe state will be unblinded and made public. This document prespecifies the analysis plan for this unblinding as well as future unblinding of the remaining subtypes. This SAP is relevant for the potential conclusions for the severe state and moderate states for comparing therapeutic anticoagulation to pharmacological venous thrombophrophylaxis. The first trigger for the severe state will utilize this analysis plan for reporting.

The authors of this document are blinded to all individual data. The authors are aware that the statistical trigger for futility has been reached for therapeutic anticoagulation in the severe state, have been advised of the primary model result for the interim analysis that prompted the trigger, along with the raw mortality rates and major bleeding event rates in the severe state. The moderate state is continuing randomization.

##### Prior analysis plan documents

There are three analysis plan documents that are the precursor to this statistical analysis plan:

1. **Multi-platform Randomized-Controlled Trial (mpRCT) Analysis Plan for ACTIV-4, ATTACC, and REMAP-CAP (AARC)** was created on September 17, 2020. This document described the creation of the AARC mpRCT and the agreed analysis plan.
2. Simulation Report for **Multi-platform Randomized-Controlled Trial (mpRCT) Analysis Plan for ACTIV-4, ATTACC, and REMAP-CAP** was created September 21, 2020. This document describes the clinical trial simulations conducted to characterize the operating characteristics of the analysis plan for the mpRCT.
3. The **Current State Document** dated November 19, 2020 is a complete specification of the statistical model and thresholds for the interim analyses to be conducted. This is a document created by the blinded joint steering committees as instructions to the Statistical Analysis Committee (SAC) for running interim analyses. This document can evolve as the mpRCT evolves incorporating the adaptations taking place in the three adaptive platform trials. The current state document referenced here is the current state document in place when the statistical trigger for futility in the severe state was met.

#### Reporting strategy

This SAP describes the planned analyses for the entire mpRCT. As outlined below, various subtypes within the mpRCT (defined below) are expected to report at different times. Moreover, given the complex process of federating data for analysis across platforms, multiple reports describing the results of analysis may be prepared and published to ensure expedited dissemination of mpRCT findings. It is anticipated that for each subtype, preliminary reports will describe key primary and secondary findings and limited subgroup analyses. Endpoints to be reported in the preliminary reports include:

- Organ support-free days to day 21 (primary endpoint)
- In-hospital mortality (co-primary endpoint)
- Major thrombotic events (key secondary endpoint)
- Major bleeding (key safety endpoint)

Comprehensive reports providing complete characterization of trial results will subsequently be prepared and published.

#### Design Considerations

The mpRCT is designed with a Bayesian analysis as the primary analysis method for the trial. There is one overarching Bayesian model, prespecified in the SAP, driving all adaptations, statistical triggers, and result summaries. The decision to use a Bayesian analysis was driven in part by the uncertainty of the extent of the pandemic. The required sample size could have been small or large. Given the expected evolution of the design and uncertain sample size, a Bayesian approach is more appropriate.

The mpRCT SAP is a joint analysis plan for three separate adaptive platform trials. The three platforms are ACTIV-4a, ATTACC, and REMAP-CAP. The three platforms have agreed to analyze the results of the comparison between two interventions, therapeutic anticoagulation and prophylactic anticoagulation, in a combined effort – labeled a multi-platform randomized-controlled trial (mpRCT). The three platform do not report their results separately–they have agreed this joint effort would be the primary method for analyzing these two interventions across the severe and moderate states. The analysis methods take the philosophy that the three platforms are essentially three multicenter randomized trials and hence combining the results together is a larger multicenter randomized trial. In this section we describe the basic defined structure of the analysis plan. This includes definitions for the patient subgroups, referred to as subtypes, the interventions, the primary endpoint, and the adaptive design. Italicized terms are defined terms by the mpRCT for these analyses.

#### Patient Subgroups

There are two *disease states* defined for this analysis; the severe and moderate disease states. The *severe state* is defined as a hospitalized patient on ICU-level organ-support at time of randomization. The *moderate state* is defined as a hospitalized patient that is not in the severe state.

The primary analysis for the mpRCT creates four distinct populations to analyze potential differential benefit of the interventions. In this mpRCT these distinct groups are labeled as *subtypes*. The four subtypes in this mpRCT are: 1) patients in the severe state; 2) patients in the moderate state with a high baseline d-dimer; 3) patients in the moderate state with a low baseline d-dimer; 4) patients in the moderate state with unknown baseline d-dimer.

A baseline d-dimer level more than 2 times the upper limit of normal is defined as being a *high* d-dimer. Any value less than two times the upper limit of normal is defined as a *low* d-dimer.

#### Interventions

The two interventions for this mpRCT are labeled as *therapeutic anticoagulation* (TAC), which plays the inferential role of the investigational treatment and *venous thromboprophylaxis* (VTP) (referred to as Control), which plays the inferential role of the control arm. These interventions are defined as:

1. **TAC**: systemic anticoagulation using unfractionated or low molecular weight heparin to achieve the degree of anticoagulation generally required for the treatment of venous thromboembolism in accordance with usual local practice.
2. **VTP**: usual care strategies, including anticoagulation with lower doses of unfractionated or low molecular weight heparin or other anticoagulant as prophylaxis against the development of venous thromboembolism in accordance with usual local practice.

#### Primary Endpoint

The primary endpoint for the mpRCT is *organ support-free days* (OSFD). The endpoint is the number of days, out of the first 21 days after randomization, that a patient is alive and free of ICU-level organ support. For the purposes of the calculation of OSFDs, if a patient dies during their index hospitalization they will be considered the status of dead even if the death occurs after the first 21 days. If a patient dies during their acute hospitalization they are coded as having –1 OSFDs, which is the worst outcome for the measure. Any patient on ICU-level organ support for the first 21 days that does not die, will be labeled as 0 OSFDs, the second worst outcome. The number of days is rounded to the nearest day, creating an integer valued outcome. The values of 1, …, 21 refer to the number of days alive and free of organ support, with smaller values being worse outcomes. The value of *22* is reserved for a patient that is alive and never on ICU-level organ support in the first 21 days. Therefore, the primary outcome OSFDs is an ordinal outcome with 24 possible outcomes for each patient, –1, 0, 1, 2, …, 21, 22. The details for the calculation of the endpoint is detailed in the mpRCT SAC data dictionaries versions 20201210A (ATTACC and ACTIV) and 10.1 (REMAP-CAP).

ICU-level of organ support is defined as high flow nasal cannula, non-invasive ventilation, invasive ventilation, extracorporeal life support, vasopressors, and/or inotropes delivered in an ICU or repurposed critical care area. In REMAP-CAP and ATTACC, high flow nasal cannula is considered to be ICU-level organ support when applied inspiratory flow is 30 L/min or higher at an $FiO_2 \geq 0.4$. In ACTIV-4, high flow nasal cannula is considered to be organ support when applied inspiratory flow is ≥20 L/min and $FiO_2 \geq 0.4$. Due to the varying provision of organ-support in potentially repurposed areas during the pandemic, ACTIV-4 defines any hospitalized area able to deliver the above organ-support as an ICU. ATTACC and REMAP-CAP specifically define regions as ICUs and non-ICUs. These matters are documented in detail in the SAC data dictionaries (see ATTACC/ACTIV version 20201210A).

This primary endpoint is an ordinal outcome and the primary analysis model analyzes the outcome as ordinal, with a cumulative logistic proportional effects model. The details of the primary analysis model are presented in Section 12.1. The measure of relative efficacy for the interventions is an odds ratio (OR) which captures the effect of having improved outcomes in OSFDs across the scale between the two interventions. The model is structured so that for TAC an OR > 1 implies improved outcomes on OSFDs for the TAC intervention compared to the VTP intervention.

#### Adaptive Design

There is a prospective adaptive analysis plan for the mpRCT. The plan is to have approximately monthly analyses of the mpRCT for potential adaptive conclusions. There are two potential prospective adaptive conclusions that can be reached for the comparison of the therapeutic anticoagulation to the prophylactic anticoagulation: *superiority* and *futility*. Superiority of TAC to VTP is defined as a high probability of an OR greater than 1 for TAC and hence improved outcomes in OSFDs for the TAC intervention. Futility will be declared if there is a high probability that the effect of TAC is below a small relative effect of a 1.20 OR. Prospective analyses have been created where statistical thresholds for claiming superiority or futility have been defined. These statistical thresholds are referred to as *statistical triggers*.

The mpRCT defines two statistical triggers within the trial that, at any analysis of the trial, would result in a declaration of superiority or futility as multi-platform conclusions.

The following statistical triggers were defined at the onset of the trial before unblinding:

1. *Superiority*. If TAC has at least a 99% posterior probability of OR>1 for organ support-free days this would trigger a claim of superiority for TAC.
2. *Futility*. If therapeutic anticoagulation is deemed to have a less than 5% posterior probability of at least a 1.20 OR compared to VTP for organ support-free days, then a claim of futility of that intervention would be declared.

For the purpose of this analysis plan, *inferiority* for TAC is defined as an OR<1.

CONFIDENTIAL

At each analysis of the mpRCT each statistical trigger would be checked for three of the subtypes:

1. Severe state subtype
2. Moderate state, high d-dimer subtype
3. Moderate state, low d-dimer subtype

The fourth subtype, moderate state with unknown d-dimer, the same conclusions will be investigated when the two moderate state subtypes have reached a conclusion but would not trigger a declaration at an interim point. The fourth analysis subtype is part of the primary analysis model but does not have adaptive statistical triggers.

#### Endpoint adjudication

Thrombosis and bleeding endpoints will be centrally adjudicated in all platforms. Preliminary reports may describe data available prior to completion of adjudication where necessary.

#### Unblinding

On December 18-19, 2020, the DSMBs advised that the statistical trigger for futility was met in the Severe state. The investigators have been unblinded to this statistical trigger in the Severe state with respect to the OR for the primary endpoint, organ support-free days, mortality rates, and rates of major bleeding.

#### Analysis Populations

For the purpose of this SAP, several analysis populations are defined.

1. **mpRCT confirmed (Primary)**. The mpRCT primary analysis population includes all enrolled patients with laboratory-confirmed COVID-19 randomized to either intervention and analyzed according to the intention-to-treat principle (i.e. according to randomly assigned treatment status, irrespective of actual treatment receipt).

It is recognized that the primary analysis includes all patients from all four subtypes. This analysis population will remain primary even if one subtype has a trigger before the other subtypes. The primary analysis will be conducted by the unblinded statistical analysis committee and the results for the subtypes with statistical triggers will be reported.

The following analysis groups will be defined for each public disclosure corresponding to each subtype being unblinded.

2. **mpRCT confirmed unblinded.** The subset of patients in the mpRCT confirmed population that belong to the subtype(s) being reported (i.e. those specific subtype(s) that have been unblinded for reporting). This population consists entirely of patients with laboratory-confirmed COVID-19 randomized to therapeutic anticoagulation or prophylactic anticoagulation and analyzed according to the intention-to-treat principle.

3. **mpRCT confirmed and suspected unblinded** (exploratory sensitivity analyses only)**.** The mpRCT severe state population including patients in REMAP-CAP with suspected but unconfirmed Covid-19 who belong to the subtypes that are unblinded for reporting.
4. **mpRCT per protocol.** This consists of the patients in the mpRCT confirmed unblinded population who have been treated as per protocol. In this analysis that is defined as 1) patients randomized to TAC and who received at least 1 dose of anticoagulation at therapeutic doses on the first full study after randomization **or** within 48 hours of randomization, **and** 2) patients randomized to VTP and who did not receive anticoagulants at therapeutic doses on the first or second full study day after randomization.

#### Endpoints

The following endpoints will be analyzed, displayed graphically, and summarized with descriptive statistics.

1. **Organ Support-Free Days** (OSFDs)
   - This is the primary endpoint for the mpRCT, and is a composite ordinal endpoint reflecting the number of days alive and off organ support, with in-hospital mortality from any cause as the worst possible outcome. Organ support considered is cardiovascular (vasopressor/inotrope support) and respiratory support (high flow nasal cannula, invasive or non-invasive ventilation, or ECMO). In-hospital death is considered a –1 and may occur after study day 21 as long as it occurs during the index hospitalization.
   - Detailed definitions for OSFDs are specified in the mpRCT SAC data dictionary
   - Missing values for organ support-free days will be treated as "missing and ignored". We will conduct a sensitivity analysis on the primary endpoint treating missing values using the "last known status carried forward" approach.

2. **In-Hospital Mortality**
   - A dichotomous endpoint of in-hospital death from any cause where the death component corresponds to a –1 on the OSFD endpoint. The measurement of in-hospital mortality is truncated at 90 days.

3. **Mortality**
   - This is a time-to-event endpoint through 28 days and 90 days.
   - Any patient currently in the hospital or transferred on organ support to an alternative care facility will be censored at their last known status alive.
   - Any patient successfully discharged from hospital, alive, without organ support, will be censored at the date of discharge if 90-day mortality data are not yet recorded.

4. **Vasopressor/inotrope-free days to day 28**
   - An ordinal outcome of the number of days alive and free of vasopressor/inotropes. This is the exact calculation of OSFD, with vasopressor/inotropes as the only organ support category. In-hospital death is considered a 0.
   - All platforms compute vasopressor-free days based on integer days on which vasopressors/inotropes were not received at any time
   - Vasopressor/inotrope-free days will be computed based on the duration of time from the initiation of vasopressors/inotropes to the final cessation of vasopressors/inotropes during the 28-day period; intervening days on which patients are not on vasopressors/inotropes will be ignored

5. **Ventilator-free days to day 28**
   - An ordinal outcome of the number of days alive and free of ventilation. This is the exact calculation of OSFD, with invasive or non-invasive ventilation as the only organ support category. In-hospital death is considered a 0.
   - All platforms compute ventilator-free days based on integer days on which invasive or non-invasive ventilation were not received at any time.
   - Ventilator-free days will be computed based on the duration of time from the initiation of invasive or non-invasive ventilation to the final cessation of invasive or non-invasive ventilation during the 28-day period; intervening days on which patients are not on ventilatory support will be ignored.

6. **Renal replacement-free days to day 28**
   - An ordinal outcome of the number of days free of renal replacement therapy.
   - All platforms compute renal replacement-free days based on integer days on which renal replacement therapy was not received at any time
   - Renal replacement-free days will be computed based on the duration of time from the initiation of renal replacement therapy to the final cessation of renal replacement therapy during the 28-day period; intervening days on which patients are not on renal replacement therapy will be ignored

7. **ECMO utilization on or before day 28**
   - A dichotomous endpoint of use of extracorporeal membrane oxygenation.

8. **Duration of ICU stay**
   - A time-to-event endpoint of leaving the ICU alive. If a patient is known to leave the ICU and return to the ICU within 21 days that intervening time will be ignored.

CONFIDENTIAL

- This variable will be truncated at 90 days.
- Patients who die in ICU at any time will be considered censored at 90 days.
- Patients still in the ICU at data snapshot will be considered censored at the time of exposure.

9. **Duration of hospital stay**
   - A time-to-event endpoint of leaving the hospital alive. If a patient is known to leave and return to the hospital within 21 days that intervening time will be ignored.
   - This variable will be truncated at 90 days.
   - Patients who die in hospital at any time will be considered censored at 90 days.
   - Patients still in the hospital at data snapshot will be considered censored at the time of exposure.

10. **ICU readmission**
    - A dichotomous endpoint of readmission to ICU.

11. **Major bleeding on or before day 14**
    - A dichotomous endpoint of major bleeding as defined according to International Society of Thrombosis and Hemostasis (ISTH) criteria in non-surgical patients.
    - The endpoint is censored at 14 days to correspond with the intervention duration.

12. **Fatal bleeding on or before day 14**
    - A dichotomous endpoint of fatal bleeding defined as death attributable to bleeding according to the site investigator reporting or as judged via central adjudication.
    - The endpoint is censored at 14 days to correspond with the intervention duration.

13. **Heparin-induced thrombocytopenia (HIT)**
    - A dichotomous endpoint of laboratory-confirmed HIT.
    - The endpoint is censored at 14 days to correspond with the intervention duration.

14. **Deep venous thrombosis**
    - A dichotomous endpoint of clinically detected deep venous thrombosis diagnosed at any time during the index hospitalization.

15. **Pulmonary embolism**
    - A dichotomous endpoint of clinically detected pulmonary embolism diagnosed at any time during the index hospitalization.

16. **Ischemic cerebrovascular event**

- A dichotomous endpoint of ischemic cerebrovascular event (stroke).

17. **Acute myocardial infarction**
    - A dichotomous endpoint of acute myocardial infarction defined according to the universal definition of myocardial infarction.

18. **Systemic arterial thromboembolism**
    - A dichotomous endpoint of clinically diagnosed systemic arterial thrombosis or embolism
    - In REMAP-CAP, this endpoint is captured as “other thrombotic event including mesenteric ischemia and limb ischemia”
    - In ACTIV IV, this endpoint is captured as “systemic arterial thromboembolism”
    - In ATTACC, this endpoint was not specified in the original protocol (v1.0 April 27, 2020); the updated protocol (v3.0, September 29, 2020) specifies this endpoint as “systemic arterial thromboembolism”

19. **Major thrombotic event or death**
    - A composite dichotomous endpoint of pulmonary embolism, ischemic cerebrovascular event, myocardial infarction, or systemic arterial thromboembolism diagnosed at any time during the index hospitalization or death in hospital

20. **All thrombotic events or death**
    - A composite dichotomous endpoint of deep vein thrombosis, pulmonary embolism, ischemic cerebrovascular event, myocardial infarction, or systemic arterial thromboembolism diagnosed at any time during the index hospitalization or death in hospital

21. **Intracranial hemorrhage**
    - A dichotomous endpoint of ischemic cerebrovascular event (stroke).

22. **Hospital re-admission**
    - A dichotomous endpoint of readmission to hospital.
    - This endpoint will be reported descriptively using proportions.

23. **The World Health Organization (WHO) 8-point ordinal scale, value on day 14.**
    - A modified WHO ordinal scale will be used:
      0 + 1 + 2 = No longer hospitalized
      3 = Hospitalized, no oxygen therapy
      4 = Oxygen by mask or nasal prongs
      5 = Non-invasive ventilation or high-flow oxygen
      6 = Intubation and mechanical ventilation
      7 = Ventilation + additional organ support: vasopressors, renal replacement therapy (RRT), ECMO
      8 = Death

#### Graphical Data Summaries

1. All ordinal endpoints will be plotted using stacked cumulative bar plots and cumulative probability plots.
2. All time-to-event endpoints will be plotted using Kaplan-Meier plots. Positive clinical event outcomes will be plotted as the cumulative rate of event, and negative events will be plotted as the cumulative rate of event-free.

#### Descriptive Statistics

1. Ordinal endpoints will be summarized by the cumulative frequency of each outcome. The 25th, 50th, and 75th percentiles will be summarized.
2. Dichotomous endpoints will be summarized by the proportion in each category.
3. Time-to-event outcomes will be summarized by the 2.5th, 10th, 25th, 50th, 75th, 90th, and 97.5th percentiles from the Kaplan-Meier estimates, as available.

#### Baseline Characteristics and Co-interventions

The following demographics will be summarized across arms. More may be added as baseline summaries: Age, sex, BMI, race, ethnicity, illness severity at admission, pre-existing conditions, baseline use of high-flow nasal oxygenation, non-invasive ventilation, invasive mechanical ventilation, ECMO, vasopressors/inotropes, renal replacement therapy, and miscellaneous physiological values and inflammatory biomarker laboratory values (see Appendix A). Anticoagulation dosing on day 1 will be compared between groups. Additionally, exposure to relevant drugs (e.g., antiplatelet agents, steroids, immunomodulatory therapies) prior to hospitalization, at baseline, and during the treatment period will be compared between groups.

#### Adherence

Adherence will be assessed based on the proportion of patients receiving anticoagulants at doses consistent with their randomly assigned anticoagulation strategy (TAC vs VTP) on the first full study day after randomization.

- In REMAP-CAP this will be study day 2
- In ACTIV-4 and ATTACC this will be study day 1

Treatment will be classified as adherent or non-adherent based on the following dosing equivalents categorization: (1) standard prophylactic, (2) intermediate prophylactic, (3) subtherapeutic, and (4) therapeutic; dosing equivalents (1) and (2) will be considered “prophylactic,” and (3) and (4) will be considered “therapeutic” for the purposes of per-protocol analyses. The criteria for each categorization for each anticoagulant are given in Appendix B.

#### Analytic Approach

Each inferential analysis will be done using a Bayesian model. Some default frequentist methods are used for exploration and description. A summary of the analyses methods is provided below. Events that occur at low frequency will be reported descriptively and not modelled.

#### Primary Analysis of Primary Endpoint

The **primary analysis model** is a Bayesian cumulative logistic model for the ordinal primary endpoint. The model is described below.

The primary endpoint for the severe state has 22 possible ordered outcomes. Let the outcome for a patient by labeled as $Y_i$, with possible values, –1 (death), 0, 1, …, 21, 22. The outcome of 22 for the severe state (never received organ support) is not possible. A cumulative logistic model is specified. The model is structured so that an OR >1 implies clinical benefit. The full details of the model are specified in the Current State Document, Version 1.1 dated November 19, 2020. The model has factors for:

1. Each level of the ordinal endpoint
2. Each global site, nested within country
3. Age; ≤39, 40-49, 50-59, 60-69, 70-79, 80+
4. Sex
5. Time: 2-week epoch binds of time working backwards from the last enrolled patient, with the most recent epoch being 4 weeks.
6. Within moderate state an effect for d-dimer level
7. An effect for each intervention; the effects for TAC are nested across subtypes
8. All sites within a country that have <5 patients randomized will be combined into a single site within that country.
9. For the primary outcome, if there is an outcome in the ordinal scale that did not occur in the data, then that outcome will be combined with a neighboring outcome (the worse outcome). This is done for model stability. For example, if the outcome 11 never occurred, then a combined outcome of 10 & 11 will be modeled for the analysis.
10. If there is a single subtype or a set of subtypes that don't allow the hierarchical structure in the model, the hierarchical structure of the treatment parameters will be replaced by a standard normal prior unless otherwise specified.

The primary analysis model will be referenced with certain model assumptions for sensitivity analyses. For example, the "time effects" in the model could be assumed to be 0.

#### Proportional Odds Assumption

The primary analysis model is based on an assumption of a proportional effect of treatment across the scale of the ordinal outcome. In order to assess the robustness of the results to this assumption, a dichotomous model is fit to every level of the ordinal outcome across the scale and the OR for each dichotomous break is presented. If the probabilities for the tails of the ordinal endpoint have small probabilities (<5%) they may not be conducted. No statistical test of proportional odds is conducted.

#### Analytic Approach for Secondary Dichotomous Endpoints

A Bayesian logistic regression model will be used for each dichotomous outcome. The model will always specify the "event" as the negative outcome and be parameterized so that an OR >1 implies benefit to patients. The model is the standard logistic link function model:

$$log\left(\frac{\pi}{1-\pi}\right) = \alpha - [factors]$$

References will be made to the factors in the model and their prior distribution. Many of these factors will be the same as the primary analysis model, with the same priors, as the parameters have similar interpretation. For example, all in-hospital mortality models should use the Beta prior distribution implied by the Dirichlet prior in the OSFD model. If not otherwise specified, the prior distribution for the main effect is $\alpha \sim N(0, 1.82^2)$ (similar to a uniform prior on the probability scale).

#### Analytic Approach for Secondary Time-To-Event Endpoints

All inferential time-to-event analyses will be done using a Bayesian piecewise exponential model. The Bayesian time-to-event model is intended to mirror a Cox proportional hazards model, with the underlying hazard rate modeled with a piecewise exponential model. The underlying hazard will be modeled with a hazard rate for each 10-day period in the model. The prior distribution for the hazard rate for each day is a gamma distribution with 1 day of exposure and a mean equal to the total exposure divided by the total number of events. This prior will have very little weight but will provide numerical stability to the model. Each factor is incorporated as a proportional hazard rate through an additive linear model of the log-hazard. The default prior for each factor is the same as for the log-odds in the ordinal model. If other non-specified variables are added to the model, then a normal distribution with mean 0 and standard deviation 10 will be utilized.

#### Analytical Approach for Subgroup Analyses

The analyses for subgroup use the same analysis for the primary models (ordinal, dichotomous, and time-to-event) with the following differences. For each model the treatment effect is modeled separately and independently in each defined subgroup. A single group will be selected as the group to have a main effect treatment effect, modeled with a normal distribution with mean 0 and standard deviation 10 (for the log-odds or log-hazard ratio). This group is either the largest group or the first subgroup. Each additional group will have an additive effect on the log parametrization scale with independent normal distribution priors with mean 0 and standard deviation 10.

If multiple subtypes are reported in a single analysis, each group within each subtype will be modeled independently without Bayesian borrowing across subtypes for the treatment effect.

#### Markov Chain Monte Carlo (MCMC) Model Stability

The Bayesian models have many parameters and there may be risk of poor model stability, including convergence and mixing behavior of the MCMC sampler. These instabilities may be based on sparse data on the outcome or covariates. The statisticians running the model may make changes that do not affect the overall interpretation but provide reliable model diagnostics and scientific rigor. Any alterations will be noted.

#### Model Outputs

The standard model outputs for each treatment effect will be the mean, standard deviation, median, and 95% credible intervals (all credible intervals will be equal-tailed intervals, so 95% credible intervals will range from the 2.5$^{th}$ percentile to the 97.5$^{th}$ percentile of the posterior distribution). For the ordinal endpoints, the odds ratios will be summarized. For the dichotomous endpoints, the odds ratios will be summarized. For the time-to-event endpoints, the hazard ratios will be summarized. **For consistency, all models will be parameterized so that an odds ratio or hazard ratio greater than 1 indicates clinical benefit.**

For each inferential model, a posterior probability that one arm is superior will be provided for each comparison between arms. This posterior probability has been identified as the primary analysis metric between arms. A posterior probability greater than 99% of superiority or inferiority has been identified as statistically significant. For futility a threshold of 95% has been specified.

#### Exploratory Analyses

Exploratory analyses after unblinding will not be considered inferential and no p-values will be presented. Any post-hoc exploratory analyses will use the following methods:

1. Ordinal endpoints will be compared using a cumulative proportional odds model with summaries of the OR, 95% confidence intervals, and Wilcoxon tests for robustness against a lack of proportional odds.
2. Time-to-Event analyses will utilize a Cox proportional hazards model, summarizing the hazard ratios and 95% confidence intervals.
3. Continuous endpoints will compare means with 95% confidence intervals based on two-sample t-test procedures.
4. Dichotomous proportions will be compared using logistic regression summarizing the OR and 95% confidence intervals. Differences between proportions will be summarized using observed differences and normal approximations for the 95% credible intervals.

#### Handling of missing data

For the primary endpoint of OSFDs missing primary outcomes will be ignored. A sensitivity analysis is conducted where last status carried forward is used for imputation. Patients with missing age, date of randomization, sex or treatment assignment will be ignored. For additional endpoints those patients missing the endpoint will be ignored (for time to event analyses censoring will be used and aren't considered missing). For the subgroup analyses patients with missing subgroup variables will be lumped into a single group of "missing" in addition to the subgroup classifications.

#### Definition of times

Adherence and per protocol analyses will rely on assessment of drug administration on post-randomization day 1. In REMAP-CAP, post-randomization day 1 is referred to as study day 2; the 24-hour period defining study day 2 (e.g., midnight to midnight, 7 am to 7 am) is defined according to local site practice. In ATTACC and ACTIV-4, post-randomization day 1 is referred to as study day 1 and constitutes the 24-hour period commencing at midnight of the day after randomization.

#### Post-randomization analyses

Participants who are randomized to receive one strategy may in fact be treated with another strategy based on health status and provider discretion. Co-interventions during the treatment period (e.g., antiplatelet agents, corticosteroids, IL6 antagonists) may modify the benefit or harm of therapeutic anticoagulation. Exploratory analyses will estimate the causal effect of the treatment for these patients using marginal structural modelling techniques. These techniques use inverse probability weighting methods that are based on patient-level covariates to create comparable groups for the analysis.

#### List of Pre-Specified Analyses

Prospectively defined primary, sensitivity, secondary, and safety analyses are summarized in Table 1. Prospectively planned subgroup analyses are summarized in Table 2. All models are described in detail below.

**Table 1.** Primary, sensitivity, secondary, safety, and per protocol analyses

| # | Status | Population | Endpoint | Notes |
|---|---|---|---|---|
| 14.1 | Primary | mpRCT confirmed | OSFDs | Primary ordinal model |
| 14.2 | Primary | mpRCT confirmed | In-hospital mortality | Primary dichotomous model |
| 14.3 | Sensitivity | mpRCT confirmed | Dichotomized OSFD | Primary dichotomous model for each dichotomization of OSFDs as a robustness check. |
| 14.4 | Sensitivity | mpRCT confirmed unblinded | OSFDs | Primary ordinal model |
| 14.5 | Sensitivity | mpRCT confirmed unblinded | In-hospital mortality | Primary dichotomous model |
| 14.6 | Sensitivity | mpRCT confirmed and suspected unblinded | OSFDs | Include REMAP-CAP suspected but not proven COVID-19 patients |
| 14.7 | Sensitivity | mpRCT confirmed and suspected unblinded | In-hospital mortality | Include REMAP-CAP suspected but not proven COVID-19 patients |
| 14.8 | Sensitivity | mpRCT confirmed unblinded | OSFDs | Remove site and time effects |
| 14.9 | Sensitivity | mpRCT confirmed unblinded | In-hospital mortality | Remove site and time effects |
| 14.10 | Sensitivity | mpRCT confirmed unblinded | OSFDs | Excluding patients who received antiplatelet agents at baseline or who are randomized in the antiplatelet |

CONFIDENTIAL

| | | | | |
|---|---|---|---|---|
| | | | | domain in REMAP-CAP |
| 14.11 | Sensitivity | mpRCT confirmed unblinded | In-hospital mortality | Excluding patients who received antiplatelet agents at baseline or who are randomized in the antiplatelet domain in REMAP-CAP |
| 14.12 | Exploratory sensitivity analysis: Severe State only | mpRCT confirmed unblinded | OSFDs | Specifies prior for TAC for enthusiasm [N(0.56,0.44)] and prior for skepticism [N(0, 0.44)] |
| 14.13 | Sensitivity | mpRCT confirmed unblinded | OSFDs | Treats missing data for OSFDs using “last known status carried forward” instead of “missing and ignored” |
| 14.14 | Secondary | mpRCT confirmed unblinded | Major thrombotic events or death | Primary dichotomous model |
| 14.15 | Secondary | mpRCT confirmed unblinded | All thrombotic events (major + DVT) or death | Primary dichotomous model |
| 14.16 | Secondary | mpRCT confirmed unblinded | Deep venous thrombosis | Primary dichotomous model |
| 14.17 | Secondary | mpRCT confirmed unblinded | Pulmonary embolism | Primary dichotomous model |
| 14.18 | Secondary | mpRCT confirmed unblinded | 90-day mortality | Primary time to event model |
| 14.19 | Secondary | mpRCT confirmed unblinded | Vasopressor/inotrope-free days to day 28 | Primary ordinal model |
| 14.20 | Secondary | mpRCT confirmed unblinded | Ventilator-free days to day 28 | Primary ordinal model |
| 14.21 | Secondary | mpRCT confirmed unblinded | Renal replacement-free days to day 28 | Primary ordinal model |
| 14.22 | Secondary | mpRCT confirmed unblinded restricted to patients not on ECMO at baseline | ECMO utilization on or before day 28 | Primary dichotomous model |
| 14.23 | Secondary | mpRCT confirmed unblinded | ICU length-of-stay | Primary time to event model |
| 14.24 | Secondary | mpRCT confirmed unblinded | Hospital length-of-stay | Primary time to event model |
| 14.25 | Secondary | mpRCT confirmed unblinded | ICU readmission | Primary dichotomous model |
| 14.26 | Secondary | mpRCT confirmed unblinded | Ischemic cerebrovascular event | Primary dichotomous model |
| 14.27 | Secondary | mpRCT confirmed unblinded | Systemic arterial thromboembolism | Primary dichotomous model |
| 14.28 | Secondary | mpRCT confirmed unblinded | Acute myocardial infarction | Primary dichotomous model |
| 14.29 | Secondary | mpRCT confirmed unblinded | WHO scale | Primary ordinal model |
| 14.30 | Safety | mpRCT confirmed unblinded | Major bleeding | Primary dichotomous model |
| 14.31 | Safety sensitivity analysis | mpRCT confirmed unblinded | Major bleeding | Primary dichotomous model |

| | | | | |
|---|---|---|---|---|
| | | | | Excluding patients who received antiplatelet agents at baseline or who are randomized into the antiplatelet domain |
| 14.32 | Safety | mpRCT confirmed unblinded | Laboratory-confirmed heparin-induced thrombocytopenia | Primary dichotomous model |
| 14.33 | Safety* | mpRCT confirmed unblinded | Fatal bleeding | Primary dichotomous model |
| 14.34 | Safety | mpRCT confirmed unblinded | Symptomatic bleeding into critical organ | Primary dichotomous model |
| 14.35 | Safety | mpRCT confirmed unblinded | Intracranial hemorrhage | Primary dichotomous model |
| 14.36 | Safety | mpRCT confirmed unblinded | Bleeding causing a ≥20 g/L drop in hemoglobin | Primary dichotomous model |
| 14.37 | Safety | mpRCT confirmed unblinded | Bleeding leading to ≥2 RBC unit transfusion | Primary dichotomous model |
| 14.38 | Sensitivity | mpRCT per protocol | OSFDs | Primary ordinal model |
| 14.39 | Sensitivity | mpRCT per protocol | In-hospital mortality | Primary dichotomous model |
| 14.40 | Sensitivity | mpRCT per protocol | Major thrombotic events or death | Primary dichotomous model |
| 14.41 | Sensitivity | mpRCT per protocol | All thrombotic events (major + DVT) or death | Primary dichotomous model |
| 14.42 | Secondary | mpRCT per protocol | Deep venous thrombosis | Primary dichotomous model |
| 14.43 | Secondary | mpRCT per protocol | Pulmonary embolism | Primary dichotomous model |
| 14.44 | Secondary | mpRCT per protocol | 90-day mortality | Primary time to event model |
| 14.45 | Secondary | mpRCT per protocol | Vasopressor/inotrope-free days to day 28 | Primary ordinal model |
| 14.46 | Secondary | mpRCT per protocol | Ventilator-free days to day 28 | Primary ordinal model |
| 14.47 | Secondary | mpRCT per protocol | Renal replacement-free days to day 28 | Primary ordinal model |
| 14.48 | Secondary | mpRCT per protocol restricted to patients not on ECMO at baseline | ECMO utilization on or before day 28 | Primary dichotomous model |
| 14.49 | Secondary | mpRCT per protocol | ICU length-of-stay | Time to event model |
| 14.50 | Secondary | mpRCT per protocol | Hospital length-of-stay | Time to event model |
| 14.51 | Secondary | mpRCT per protocol | ICU readmission | Primary dichotomous model |
| 14.52 | Secondary | mpRCT per protocol | Ischemic cerebrovascular event | Primary dichotomous model |
| 14.53 | Secondary | mpRCT per protocol | Systemic arterial thromboembolism | Primary dichotomous model |
| 14.54 | Secondary | mpRCT per protocol | Acute myocardial infarction | Primary dichotomous model |
| 14.55 | Secondary | mpRCT per protocol | WHO scale | Primary ordinal model |

**Table 2.** Prospectively defined subgroup analyses

| Subgroup | | Endpoint – Model # |
|---|---|---|

CONFIDENTIAL

| | **Specification of subgroup** | OSFDs (efficacy) | Hospital mortality (efficacy) | Major thrombotic event or death (efficacy) | Major bleeding (safety) |
|---|---|---|---|---|---|
| Age | Categorical (<50 years, 50-70 years, and >70 years) | 15.1.1 | 15.1.2 | 15.1.3 | 15.1.4 |
| Sex | Dichotomous | 15.2.1 | 15.2.2 | 15.2.3 | 15.2.4 |
| Invasive mechanical ventilation at baseline* | Dichotomous | 15.3.1 | 15.3.2 | 15.3.3 | 15.3.4 |
| Antiplatelet agent use at baseline in hospital at time of randomization | Dichotomous | 15.4.1 | 15.4.2 | 15.4.3 | 15.4.4 |

*Pre-specified in REMAP-CAP domain specific appendix; DSA also specifies "all remaining potentially evaluable treatment-by-treatment interactions with other domains"

**Table 3.** Other exploratory subgroups of interest

| **Subgroup** | **Specification of subgroup** | **Endpoint – Model #** | | | |
|---|---|---|---|---|---|
| | | OSFDs (efficacy) | Hospital mortality (efficacy) | Major thrombotic event or death (efficacy) | Major bleeding (safety) |
| Race | Categorical | 16.1.1 | 16.1.2 | 16.1.3 | 16.1.4 |
| D-dimer | Quintiles | 16.2.1 | 16.2.2 | 16.2.3 | 16.2.4 |
| Baseline troponin* | Terciles | 16.3.1 | 16.3.2 | 16.3.3 | 16.3.4 |
| Therapeutic anticoagulation strategy: LMWH vs. UFH (site classification strategy)* | Dichotomous | 16.4.1 | 16.4.2 | 16.4.3 | 16.4.4 |
| Therapeutic anticoagulation strategy: LMWH vs. UFH (day 1 patient classification strategy)* | Dichotomous | 16.5.1 | 16.5.2 | 16.5.3 | 16.5.4 |
| Proven concomitant bacterial co-infection* | Dichotomous | 16.6.1 | 16.6.2 | 16.6.3 | 16.6.4 |
| Randomization status in steroid domain (within REMAP only) | Dichotomous (steroid or control) | 16.7.1 | 16.7.2 | 16.7.3 | 16.7.4 |
| Randomization status in immunomodulatory domain (within REMAP only, restricted to patients with unblinded data in IM domain) | Dichotomous (immunomodulatory therapy or control) | 16.8.1 | 16.8.2 | 16.8.3 | 16.8.4 |
| Body mass index | Terciles | 16.9.1 | 16.9.2 | 16.9.3 | 16.9.4 |

| | | | | | |
|---|---|---|---|---|---|
| Severity of illness | Terciles | 16.10.1 | 16.10.2 | 16.10.3 | 16.10.4 |
| Shock (use of vasopressors or inotropes at baseline)* | Dichotomous | 16.11.1 | 16.11.2 | 16.11.3 | 16.11.4 |
| Baseline chronic kidney disease | Dichotomous | 16.12.1 | 16.12.2 | 16.12.3 | 16.12.4 |
| Steroid administration for COVID-19 at baseline | Dichotomous | 16.13.1 | 16.13.2 | 16.13.3 | 16.13.4 |
| Statin use at baseline | Dichotomous | 16.14.1 | 16.14.2 | 16.14.3 | 16.14.4 |
| Usual care practice: low vs intermediate (patient classification strategy)[1] | Dichotomous | 16.15.1 | 16.15.2 | 16.15.3 | 16.15.4 |
| Usual care practice: low vs intermediate (day 1 site classification strategy)[2] | Dichotomous | 16.16.1 | 16.16.2 | 16.16.3 | 16.16.4 |

*Pre-specified in REMAP-CAP domain specific appendix; DSA also specifies "all remaining potentially evaluable treatment-by-treatment interactions with other domains"

[1]Patients will be classified as receiving low dose VTP if they receive a low dose anticoagulant (according to criteria given in Appendix B) on both of the first two full study days following randomization. Patients will be classified as receiving intermediate dose VTP if they receive an intermediate dose anticoagulant (according to criteria given in Appendix B) on either of the first two full study days following randomization.

[2]Sites will be classified as "low dose" usual practice if >50% of patients randomized to the VTP at that site received low dose VTP; otherwise, sites will be classified as "intermediate dose" usual practice.

#### Models for urgent reporting

#### Primary analysis of OSFDs

| | |
|---|---|
| Population | mpRCT confirmed |
| Endpoint | Organ support-free days |
| Model | Primary analysis ordinal model |
| Factors | Intervention, age, sex, site, time, d-dimer (moderate) |
| Analysis | Conducted by the unblinded SAC |

The following posterior probabilities will be reported for each subtype being reported:

| **Quantity of Interest** | **Posterior Probability** |
|---|---|
| Therapeutic anticoagulation is superior to control | |
| Therapeutic anticoagulation is futile | |
| Therapeutic anticoagulation is inferior | |

The following will be reported:

| **Odds Ratio Parameter** | **Mean** | **SD** | **Median** | **95% Credible Interval** |
|---|---|---|---|---|
| Age < 39 | | | | |
| Age 40, 49 | | | | |
| Age 50, 59 | | | | |
| Age 70-79 | | | | |
| Age 80+ | | | | |
| D-dimer high (for moderate) | | | | |
| D-dimer missing (for moderate) | | | | |
| Female | | | | |
| Time epoch 1 | | | | |
| … | | | | |
| Time epoch k-1 | | | | |
| Therapeutic anticoagulation | | | | |

#### Primary analysis of in-hospital mortality

| | |
|---|---|
| Population | mpRCT confirmed |
| Endpoint | Hospital mortality (censored at day 90 or time of snapshot) |
| Model | Primary analysis dichotomous model |
| Factors | Intervention, age, sex, site, time, d-dimer (moderate) |
| Analysis | Conducted by the unblinded SAC |

The following posterior probabilities will be reported for each subtype being reported:

| **Quantity of Interest** | **Posterior Probability** |
|---|---|
| Therapeutic anticoagulation is superior to control | |
| Therapeutic anticoagulation is futile | |
| Therapeutic anticoagulation is inferior | |

The following will be reported:

| **Odds Ratio Parameter** | **Mean** | **SD** | **Median** | **95% Credible Interval** |
|---|---|---|---|---|
| Age < 39 | | | | |
| Age 40, 49 | | | | |
| Age 50, 59 | | | | |
| Age 70-79 | | | | |
| Age 80+ | | | | |
| D-dimer high (for moderate) | | | | |
| D-dimer missing (for moderate) | | | | |
| Female | | | | |
| Time epoch 1 | | | | |
| … | | | | |
| Time epoch k-1 | | | | |
| Therapeutic anticoagulation | | | | |

CONFIDENTIAL

#### Sensitivity analysis for OSFDs – proportional odds assumptions

| | |
|---|---|
| Population | mpRCT confirmed |
| Endpoint | Organ support-free days |
| Model | Primary dichotomous model |
| Factors | Intervention, age, sex, site, time |
| Analysis | Conducted by the unblinded SAC |

**Notes**

1. For this analysis, the primary dichotomous model will be fit to each dichotomization of OSFDs and the summaries of the odds ratio of therapeutic anticoagulation will be reported.

The following summaries will be reported for the therapeutic anticoagulation odds ratios:

| **OSFD Dichotomization** | **Mean** | **SD** | **Median** | **95% Credible Interval** |
|---|---|---|---|---|
| -1 vs ≥0 | | | | |
| ≤0 vs ≥1 | | | | |
| ≤1 vs ≥2 | | | | |
| ≤2 vs ≥3 | | | | |
| ≤3 vs ≥4 | | | | |
| ≤4 vs ≥5 | | | | |
| ≤5 vs ≥6 | | | | |
| ≤6 vs ≥7 | | | | |
| ≤7 vs ≥8 | | | | |
| ≤8 vs ≥9 | | | | |
| ≤9 vs ≥10 | | | | |
| ≤10 vs ≥11 | | | | |
| ≤11 vs ≥12 | | | | |
| ≤12 vs ≥13 | | | | |
| ≤13 vs ≥14 | | | | |
| ≤14 vs ≥15 | | | | |
| ≤15 vs ≥16 | | | | |
| ≤16 vs ≥17 | | | | |
| ≤17 vs ≥18 | | | | |
| ≤18 vs ≥19 | | | | |
| ≤19 vs ≥20 | | | | |
| ≤20 vs 21 | | | | |

CONFIDENTIAL

#### Sensitivity analysis of OSFDs in unblinded population

| | |
|---|---|
| Population | mpRCT confirmed unblinded |
| Endpoint | Organ support-free days |
| Model | Primary analysis ordinal model |
| Factors | Intervention, age, sex, site, time, d-dimer (moderate) |
| Analysis | Conducted by the mpRCT analysis team |

The following posterior probabilities will be reported for each subtype being reported:

| **Quantity of Interest** | **Posterior Probability** |
|---|---|
| Therapeutic anticoagulation is superior to control | |
| Therapeutic anticoagulation is futile | |
| Therapeutic anticoagulation is inferior | |

The following will be reported:

| **Odds Ratio Parameter** | **Mean** | **SD** | **Median** | **95% Credible Interval** |
|---|---|---|---|---|
| Age < 39 | | | | |
| Age 40, 49 | | | | |
| Age 50, 59 | | | | |
| Age 70-79 | | | | |
| Age 80+ | | | | |
| D-dimer high (for moderate) | | | | |
| D-dimer missing (for moderate) | | | | |
| Female | | | | |
| Time epoch 1 | | | | |
| … | | | | |
| Time epoch k-1 | | | | |
| Therapeutic anticoagulation | | | | |

#### Sensitivity analysis of in-hospital mortality in unblinded population

| | |
|---|---|
| Population | mpRCT confirmed unblinded |
| Endpoint | Hospital mortality (censored at day 90 or time of snapshot) |
| Model | Primary analysis dichotomous model |
| Factors | Intervention, age, sex, site, time, d-dimer (moderate) |
| Analysis | Conducted by the mpRCT analysis team |

The following posterior probabilities will be reported for each subtype being reported:

| **Quantity of Interest** | **Posterior Probability** |
|---|---|
| Therapeutic anticoagulation is superior to control | |
| Therapeutic anticoagulation is futile | |
| Therapeutic anticoagulation is inferior | |

The following will be reported:

| **Odds Ratio Parameter** | **Mean** | **SD** | **Median** | **95% Credible Interval** |
|---|---|---|---|---|
| Age < 39 | | | | |
| Age 40, 49 | | | | |
| Age 50, 59 | | | | |
| Age 70-79 | | | | |
| Age 80+ | | | | |
| D-dimer high (for moderate) | | | | |
| D-dimer missing (for moderate) | | | | |
| Female | | | | |
| Time epoch 1 | | | | |
| … | | | | |
| Time epoch k-1 | | | | |
| Therapeutic anticoagulation | | | | |

#### Sensitivity analysis for OSFDs – include suspected but not confirmed patients

| | |
|---|---|
| Population | mpRCT confirmed and suspected unblinded |
| Endpoint | Organ support-free days |
| Model | Primary analysis ordinal model |
| Factors | Intervention, age, sex, site, time, d-dimer (moderate) |
| Analysis | Conducted by the mpRCT analysis team |

The following posterior probabilities will be reported for each subtype being reported:

| **Quantity of Interest** | **Posterior Probability** |
|---|---|
| Therapeutic anticoagulation is superior to control | |
| Therapeutic anticoagulation is futile | |
| Therapeutic anticoagulation is inferior | |

The following will be reported:

| **Odds Ratio Parameter** | **Mean** | **SD** | **Median** | **95% Credible Interval** |
|---|---|---|---|---|
| Age < 39 | | | | |
| Age 40, 49 | | | | |
| Age 50, 59 | | | | |
| Age 70-79 | | | | |
| Age 80+ | | | | |
| D-dimer high (for moderate) | | | | |
| D-dimer missing (for moderate) | | | | |
| Female | | | | |
| Time epoch 1 | | | | |
| … | | | | |
| Time epoch k-1 | | | | |
| Therapeutic anticoagulation | | | | |

#### Sensitivity analysis for in-hospital mortality – include suspected but not confirmed patients

| | |
|---|---|
| Population | mpRCT confirmed and suspected unblinded |
| Endpoint | Hospital mortality (censored at day 90 or time of snapshot) |
| Model | Primary analysis dichotomous model |
| Factors | Intervention, age, sex, site, time, d-dimer (moderate) |
| Analysis | Conducted by the mpRCT analysis team |

The following posterior probabilities will be reported for each subtype being reported:

| **Quantity of Interest** | **Posterior Probability** |
|---|---|
| Therapeutic anticoagulation is superior to control | |
| Therapeutic anticoagulation is futile | |
| Therapeutic anticoagulation is inferior | |

The following will be reported:

| **Odds Ratio Parameter** | **Mean** | **SD** | **Median** | **95% Credible Interval** |
|---|---|---|---|---|
| Age < 39 | | | | |
| Age 40, 49 | | | | |
| Age 50, 59 | | | | |
| Age 70-79 | | | | |
| Age 80+ | | | | |
| D-dimer high (for moderate) | | | | |
| D-dimer missing (for moderate) | | | | |
| Female | | | | |
| Time epoch 1 | | | | |
| … | | | | |
| Time epoch k-1 | | | | |
| Therapeutic anticoagulation | | | | |

#### Sensitivity analysis for OSFDs – site and time effects removed

| | |
|---|---|
| Population | mpRCT confirmed unblinded |
| Endpoint | Organ support-free days |
| Model | Primary analysis ordinal model |
| Factors | Intervention, age, sex, d-dimer (moderate) |
| Analysis | Conducted by the mpRCT analysis team |

The following posterior probabilities will be reported for each subtype being reported:

| **Quantity of Interest** | **Posterior Probability** |
|---|---|
| Therapeutic anticoagulation is superior to control | |
| Therapeutic anticoagulation is futile | |
| Therapeutic anticoagulation is inferior | |

The following will be reported:

| **Odds Ratio Parameter** | **Mean** | **SD** | **Median** | **95% Credible Interval** |
|---|---|---|---|---|
| Age < 39 | | | | |
| Age 40, 49 | | | | |
| Age 50, 59 | | | | |
| Age 70-79 | | | | |
| Age 80+ | | | | |
| D-dimer high (for moderate) | | | | |
| D-dimer missing (for moderate) | | | | |
| Female | | | | |
| Therapeutic anticoagulation | | | | |

#### Sensitivity analysis for in-hospital mortality – site and time effects removed

| | |
|---|---|
| Population | mpRCT confirmed unblinded |
| Endpoint | Hospital mortality (censored at day 90 or time of snapshot) |
| Model | Primary analysis dichotomous model |
| Factors | Intervention, age, sex, d-dimer (moderate) |
| Analysis | Conducted by the mpRCT analysis team |

The following posterior probabilities will be reported for each subtype being reported:

| **Quantity of Interest** | **Posterior Probability** |
|---|---|
| Therapeutic anticoagulation is superior to control | |
| Therapeutic anticoagulation is futile | |
| Therapeutic anticoagulation is inferior | |

The following will be reported:

| **Odds Ratio Parameter** | **Mean** | **SD** | **Median** | **95% Credible Interval** |
|---|---|---|---|---|
| Age < 39 | | | | |
| Age 40, 49 | | | | |
| Age 50, 59 | | | | |
| Age 70-79 | | | | |
| Age 80+ | | | | |
| D-dimer high (for moderate) | | | | |
| D-dimer missing (for moderate) | | | | |
| Female | | | | |
| Therapeutic anticoagulation | | | | |

#### Sensitivity analysis excluding patients on antiplatelet agents at baseline

| | |
|---|---|
| Population | mpRCT confirmed unblinded excluding patients on antiplatelet agents at baseline or who are randomized in the antiplatelet domain in REMAP-CAP |
| Endpoint | Organ support-free days |
| Model | Primary analysis ordinal model |
| Factors | Intervention, age, sex, site, time, d-dimer (moderate) |
| Analysis | Conducted by the mpRCT analysis team |

The following posterior probabilities will be reported for each subtype being reported:

| **Quantity of Interest** | **Posterior Probability** |
|---|---|
| Therapeutic anticoagulation is superior to control | |
| Therapeutic anticoagulation is futile | |
| Therapeutic anticoagulation is inferior | |

The following will be reported:

| **Odds Ratio Parameter** | **Mean** | **SD** | **Median** | **95% Credible Interval** |
|---|---|---|---|---|
| Age < 39 | | | | |
| Age 40, 49 | | | | |
| Age 50, 59 | | | | |
| Age 70-79 | | | | |
| Age 80+ | | | | |
| D-dimer high (for moderate) | | | | |
| D-dimer missing (for moderate) | | | | |
| Female | | | | |
| Time epoch 1 | | | | |
| … | | | | |
| Time epoch k-1 | | | | |
| Therapeutic anticoagulation | | | | |

CONFIDENTIAL

#### Sensitivity analysis excluding patients on antiplatelet agents at baseline or during treatment

| | |
|---|---|
| Population | mpRCT confirmed unblinded excluding patients on antiplatelet agents at baseline or who are randomized in the antiplatelet domain in REMAP-CAP |
| Endpoint | Hospital mortality (censored at day 90 or time of snapshot) |
| Model | Primary analysis dichotomous model |
| Factors | Intervention, age, sex, site, time, d-dimer (moderate) |
| Analysis | Conducted by the mpRCT analysis team |

The following posterior probabilities will be reported for each subtype being reported:

| **Quantity of Interest** | **Posterior Probability** |
|---|---|
| Therapeutic anticoagulation is superior to control | |
| Therapeutic anticoagulation is futile | |
| Therapeutic anticoagulation is inferior | |

The following will be reported:

| **Odds Ratio Parameter** | **Mean** | **SD** | **Median** | **95% Credible Interval** |
|---|---|---|---|---|
| Age < 39 | | | | |
| Age 40, 49 | | | | |
| Age 50, 59 | | | | |
| Age 70-79 | | | | |
| Age 80+ | | | | |
| D-dimer high (for moderate) | | | | |
| D-dimer missing (for moderate) | | | | |
| Female | | | | |
| Time epoch 1 | | | | |
| … | | | | |
| Time epoch k-1 | | | | |
| Therapeutic anticoagulation | | | | |

#### Sensitivity analysis of OSFDs specifying a prior describing enthusiasm or skepticism for benefit

| | |
|---|---|
| Population | mpRCT confirmed unblinded |
| Endpoint | Hospital mortality (censored at day 90 or time of snapshot) |
| Model | Primary analysis ordinal model |
| Factors | Intervention, age, sex, site, time, d-dimer (moderate) |
| Analysis | Conducted by the mpRCT analysis team |

**Notes**

1. The enthusiastic prior distribution for the intervention effect will be set to N(0.56, 0.44) (equivalent a 50% posterior probability of OR≥1.75 and 10% posterior probability of OR<1). OR=1.75 is equivalent to an approximately 10% absolute risk reduction in mortality assuming a baseline mortality rate of 35%. This is deemed to represent reasonable enthusiasm for the effect of treatment.
2. The skeptical prior distribution for the intervention effect will be set to N(0, 0.44) (equivalent to a 50% posterior probability of OR≤1 and a 66% posterior probability of futility (OR≤1.2). This is deemed to represent reasonable skepticism for the effect of treatment.

The following posterior probabilities will be reported for each subtype being reported:

| **Quantity of Interest** | **Posterior Probability** |
|---|---|
| Therapeutic anticoagulation is superior to control | |
| Therapeutic anticoagulation is futile | |
| Therapeutic anticoagulation is inferior | |

The following will be reported:

| **Odds Ratio Parameter** | **Mean** | **SD** | **Median** | **95% Credible Interval** |
|---|---|---|---|---|
| Age < 39 | | | | |
| Age 40, 49 | | | | |
| Age 50, 59 | | | | |
| Age 70-79 | | | | |
| Age 80+ | | | | |
| D-dimer high (for moderate) | | | | |
| D-dimer missing (for moderate) | | | | |
| Female | | | | |
| Time epoch 1 | | | | |
| … | | | | |
| Time epoch k-1 | | | | |
| Therapeutic anticoagulation | | | | |

CONFIDENTIAL

#### Sensitivity analysis of OSFDs treating missing OSFDs based on last known status carried forward

| | |
|---|---|
| Population | mpRCT confirmed unblinded |
| Endpoint | Organ support-free days where missing data are handled based on last known status carried forward |
| Model | Primary analysis ordinal model |
| Factors | Intervention, age, sex, site, time, d-dimer (moderate) |
| Analysis | Conducted by the mpRCT analysis team |

The following posterior probabilities will be reported for each subtype being reported:

| Quantity of Interest | Posterior Probability |
|---|---|
| Therapeutic anticoagulation is superior to control | |
| Therapeutic anticoagulation is futile | |
| Therapeutic anticoagulation is inferior | |

The following will be reported:

| Odds Ratio Parameter | Mean | SD | Median | 95% Credible Interval |
|---|---|---|---|---|
| Age < 39 | | | | |
| Age 40, 49 | | | | |
| Age 50, 59 | | | | |
| Age 70-79 | | | | |
| Age 80+ | | | | |
| D-dimer high (for moderate) | | | | |
| D-dimer missing (for moderate) | | | | |
| Female | | | | |
| Time epoch 1 | | | | |
| … | | | | |
| Time epoch k-1 | | | | |
| Therapeutic anticoagulation | | | | |

CONFIDENTIAL

#### Secondary analysis of major thrombotic events or death

| | |
|---|---|
| Population | mpRCT confirmed unblinded |
| Endpoint | Major thrombotic events or in-hospital death |
| Model | Primary analysis dichotomous model |
| Factors | Intervention, age, sex, site, time, d-dimer (moderate) |
| Analysis | Conducted by the mpRCT analysis team |

The following posterior probabilities will be reported for each subtype being reported:

| **Quantity of Interest** | **Posterior Probability** |
|---|---|
| Therapeutic anticoagulation is superior to control | |
| Therapeutic anticoagulation is futile | |
| Therapeutic anticoagulation is inferior | |

The following will be reported:

| **Odds Ratio Parameter** | **Mean** | **SD** | **Median** | **95% Credible Interval** |
|---|---|---|---|---|
| Age < 39 | | | | |
| Age 40, 49 | | | | |
| Age 50, 59 | | | | |
| Age 70-79 | | | | |
| Age 80+ | | | | |
| D-dimer high (for moderate) | | | | |
| D-dimer missing (for moderate) | | | | |
| Female | | | | |
| Time epoch 1 | | | | |
| … | | | | |
| Time epoch k-1 | | | | |
| Therapeutic anticoagulation | | | | |

#### Secondary analysis of all thrombotic events or death

| | |
|---|---|
| Population | mpRCT confirmed unblinded |
| Endpoint | All thrombotic events or death |
| Model | Primary analysis dichotomous model |
| Factors | Intervention, age, sex, site, time, d-dimer (moderate) |
| Analysis | Conducted by the mpRCT analysis team |

The following posterior probabilities will be reported for each subtype being reported:

| **Quantity of Interest** | **Posterior Probability** |
|---|---|
| Therapeutic anticoagulation is superior to control | |
| Therapeutic anticoagulation is futile | |
| Therapeutic anticoagulation is inferior | |

The following will be reported:

| **Odds Ratio Parameter** | **Mean** | **SD** | **Median** | **95% Credible Interval** |
|---|---|---|---|---|
| Age < 39 | | | | |
| Age 40, 49 | | | | |
| Age 50, 59 | | | | |
| Age 70-79 | | | | |
| Age 80+ | | | | |
| D-dimer high (for moderate) | | | | |
| D-dimer missing (for moderate) | | | | |
| Female | | | | |
| Time epoch 1 | | | | |
| … | | | | |
| Time epoch k-1 | | | | |
| Therapeutic anticoagulation | | | | |

#### Secondary analysis of deep venous thrombosis

| | |
|---|---|
| Population | mpRCT confirmed unblinded |
| Endpoint | Deep venous thrombosis |
| Model | Primary analysis dichotomous model |
| Factors | Intervention, age, sex, site, time, d-dimer (moderate) |
| Analysis | Conducted by the mpRCT analysis team |

The following posterior probabilities will be reported for each subtype being reported:

| **Quantity of Interest** | **Posterior Probability** |
|---|---|
| Therapeutic anticoagulation is superior to control | |
| Therapeutic anticoagulation is futile | |
| Therapeutic anticoagulation is inferior | |

The following will be reported:

| **Odds Ratio Parameter** | **Mean** | **SD** | **Median** | **95% Credible Interval** |
|---|---|---|---|---|
| Age < 39 | | | | |
| Age 40, 49 | | | | |
| Age 50, 59 | | | | |
| Age 70-79 | | | | |
| Age 80+ | | | | |
| D-dimer high (for moderate) | | | | |
| D-dimer missing (for moderate) | | | | |
| Female | | | | |
| Time epoch 1 | | | | |
| … | | | | |
| Time epoch k-1 | | | | |
| Therapeutic anticoagulation | | | | |

CONFIDENTIAL

#### Secondary analysis of pulmonary embolism

| | |
|---|---|
| Population | mpRCT confirmed unblinded |
| Endpoint | Acute pulmonary embolism |
| Model | Primary analysis dichotomous model |
| Factors | Intervention, age, sex, site, time, d-dimer (moderate) |
| Analysis | Conducted by the mpRCT analysis team |

The following posterior probabilities will be reported for each subtype being reported:

| Quantity of Interest | Posterior Probability |
|---|---|
| Therapeutic anticoagulation is superior to control | |
| Therapeutic anticoagulation is futile | |
| Therapeutic anticoagulation is inferior | |

The following will be reported:

| Odds Ratio Parameter | Mean | SD | Median | 95% Credible Interval |
|---|---|---|---|---|
| Age < 39 | | | | |
| Age 40, 49 | | | | |
| Age 50, 59 | | | | |
| Age 70-79 | | | | |
| Age 80+ | | | | |
| D-dimer high (for moderate) | | | | |
| D-dimer missing (for moderate) | | | | |
| Female | | | | |
| Time epoch 1 | | | | |
| … | | | | |
| Time epoch k-1 | | | | |
| Therapeutic anticoagulation | | | | |

#### Secondary analysis of 90-day mortality

| | |
|---|---|
| Population | mpRCT confirmed unblinded |
| Endpoint | Mortality at 90 days |
| Model | Time to event model |
| Factors | Intervention, age, sex, site, time, d-dimer (moderate) |
| Analysis | Conducted by the mpRCT analysis team |

The following posterior probabilities will be reported for each subtype being reported:

| **Quantity of Interest** | **Posterior Probability** |
|---|---|
| Therapeutic anticoagulation is superior to control | |
| Therapeutic anticoagulation is futile | |
| Therapeutic anticoagulation is inferior | |

The following will be reported:

| **Odds Ratio Parameter** | **Mean** | **SD** | **Median** | **95% Credible Interval** |
|---|---|---|---|---|
| Age < 39 | | | | |
| Age 40, 49 | | | | |
| Age 50, 59 | | | | |
| Age 70-79 | | | | |
| Age 80+ | | | | |
| D-dimer high (for moderate) | | | | |
| D-dimer missing (for moderate) | | | | |
| Female | | | | |
| Time epoch 1 | | | | |
| … | | | | |
| Time epoch k-1 | | | | |
| Therapeutic anticoagulation | | | | |

#### A secondary analysis of days free of vasopressors or inotropes

| | |
|---|---|
| Population | mpRCT confirmed unblinded |
| Endpoint | Days free of vasopressors and inotropes to day 28 |
| Model | Primary analysis ordinal model |
| Factors | Intervention, age, sex, site, time, d-dimer (moderate) |
| Analysis | Conducted by the mpRCT analysis team |

The following posterior probabilities will be reported for each subtype being reported:

| **Quantity of Interest** | **Posterior Probability** |
|---|---|
| Therapeutic anticoagulation is superior to control | |
| Therapeutic anticoagulation is futile | |
| Therapeutic anticoagulation is inferior | |

The following will be reported:

| **Odds Ratio Parameter** | **Mean** | **SD** | **Median** | **95% Credible Interval** |
|---|---|---|---|---|
| Age < 39 | | | | |
| Age 40, 49 | | | | |
| Age 50, 59 | | | | |
| Age 70-79 | | | | |
| Age 80+ | | | | |
| D-dimer high (for moderate) | | | | |
| D-dimer missing (for moderate) | | | | |
| Female | | | | |
| Time epoch 1 | | | | |
| … | | | | |
| Time epoch k-1 | | | | |
| Therapeutic anticoagulation | | | | |

CONFIDENTIAL

#### A secondary analysis of days free of invasive or non-invasive ventilation

| | |
|---|---|
| Population | mpRCT confirmed unblinded |
| Endpoint | Days free of invasive or non-invasive ventilation to day 28 |
| Model | Primary analysis ordinal model |
| Factors | Intervention, age, sex, site, time, d-dimer (moderate) |
| Analysis | Conducted by the mpRCT analysis team |

The following posterior probabilities will be reported for each subtype being reported:

| Quantity of Interest | Posterior Probability |
|---|---|
| Therapeutic anticoagulation is superior to control | |
| Therapeutic anticoagulation is futile | |
| Therapeutic anticoagulation is inferior | |

The following will be reported:

| Odds Ratio Parameter | Mean | SD | Median | 95% Credible Interval |
|---|---|---|---|---|
| Age < 39 | | | | |
| Age 40, 49 | | | | |
| Age 50, 59 | | | | |
| Age 70-79 | | | | |
| Age 80+ | | | | |
| D-dimer high (for moderate) | | | | |
| D-dimer missing (for moderate) | | | | |
| Female | | | | |
| Time epoch 1 | | | | |
| … | | | | |
| Time epoch k-1 | | | | |
| Therapeutic anticoagulation | | | | |

CONFIDENTIAL

#### Days free of renal-replacement therapy

| | |
|---|---|
| Population | mpRCT confirmed unblinded |
| Endpoint | Days free of renal replacement therapy to day 28 |
| Model | Primary analysis ordinal model |
| Factors | Intervention, age, sex, site, time, d-dimer (moderate) |
| Analysis | Conducted by the mpRCT analysis team |

The following posterior probabilities will be reported for each subtype being reported:

| **Quantity of Interest** | **Posterior Probability** |
|---|---|
| Therapeutic anticoagulation is superior to control | |
| Therapeutic anticoagulation is futile | |
| Therapeutic anticoagulation is inferior | |

The following will be reported:

| **Odds Ratio Parameter** | **Mean** | **SD** | **Median** | **95% Credible Interval** |
|---|---|---|---|---|
| Age < 39 | | | | |
| Age 40, 49 | | | | |
| Age 50, 59 | | | | |
| Age 70-79 | | | | |
| Age 80+ | | | | |
| D-dimer high (for moderate) | | | | |
| D-dimer missing (for moderate) | | | | |
| Female | | | | |
| Time epoch 1 | | | | |
| … | | | | |
| Time epoch k-1 | | | | |
| Therapeutic anticoagulation | | | | |

CONFIDENTIAL

#### ECMO utilization

| | |
|---|---|
| Population | mpRCT confirmed unblinded excluding patients on ECMO at baseline |
| Endpoint | ECMO utilization to day 28 |
| Model | Primary analysis dichotomous model |
| Factors | Intervention, age, sex, site, time, d-dimer (moderate) |
| Analysis | Conducted by the mpRCT analysis team |

The following posterior probabilities will be reported for each subtype being reported:

| **Quantity of Interest** | **Posterior Probability** |
|---|---|
| Therapeutic anticoagulation is superior to control | |
| Therapeutic anticoagulation is futile | |
| Therapeutic anticoagulation is inferior | |

The following will be reported:

| **Odds Ratio Parameter** | **Mean** | **SD** | **Median** | **95% Credible Interval** |
|---|---|---|---|---|
| Age < 39 | | | | |
| Age 40, 49 | | | | |
| Age 50, 59 | | | | |
| Age 70-79 | | | | |
| Age 80+ | | | | |
| D-dimer high (for moderate) | | | | |
| D-dimer missing (for moderate) | | | | |
| Female | | | | |
| Time epoch 1 | | | | |
| … | | | | |
| Time epoch k-1 | | | | |
| Therapeutic anticoagulation | | | | |

#### A secondary analysis of ICU length-of-stay

| | |
|---|---|
| Population | mpRCT confirmed unblinded |
| Endpoint | ICU length-of-stay (event=ICU discharge); patients who die in hospital are assigned a value of 90 days |
| Model | Primary time-to-event model |
| Factors | Intervention, age, sex, site, time, d-dimer (moderate) |
| Analysis | Conducted by the mpRCT analysis team |

The following posterior probabilities will be reported for each subtype being reported:

| **Quantity of Interest** | **Posterior Probability** |
|---|---|
| Therapeutic anticoagulation is superior to control | |
| Therapeutic anticoagulation is futile | |
| Therapeutic anticoagulation is inferior | |

The following will be reported:

| **Odds Ratio Parameter** | **Mean** | **SD** | **Median** | **95% Credible Interval** |
|---|---|---|---|---|
| Age < 39 | | | | |
| Age 40, 49 | | | | |
| Age 50, 59 | | | | |
| Age 70-79 | | | | |
| Age 80+ | | | | |
| D-dimer high (for moderate) | | | | |
| D-dimer missing (for moderate) | | | | |
| Female | | | | |
| Time epoch 1 | | | | |
| … | | | | |
| Time epoch k-1 | | | | |
| Therapeutic anticoagulation | | | | |

CONFIDENTIAL

#### A secondary analysis of hospital length-of-stay

| | |
|---|---|
| Population | mpRCT confirmed unblinded |
| Endpoint | Hospital length-of-stay (event=hospital discharge); patients who die in hospital are assigned a value of 90 days |
| Model | Primary time-to-event model |
| Factors | Intervention, age, sex, site, time, d-dimer (moderate) |
| Analysis | Conducted by the mpRCT analysis team |

The following posterior probabilities will be reported for each subtype being reported:

| **Quantity of Interest** | **Posterior Probability** |
|---|---|
| Therapeutic anticoagulation is superior to control | |
| Therapeutic anticoagulation is futile | |
| Therapeutic anticoagulation is inferior | |

The following will be reported:

| **Odds Ratio Parameter** | **Mean** | **SD** | **Median** | **95% Credible Interval** |
|---|---|---|---|---|
| Age < 39 | | | | |
| Age 40, 49 | | | | |
| Age 50, 59 | | | | |
| Age 70-79 | | | | |
| Age 80+ | | | | |
| D-dimer high (for moderate) | | | | |
| D-dimer missing (for moderate) | | | | |
| Female | | | | |
| Time epoch 1 | | | | |
| … | | | | |
| Time epoch k-1 | | | | |
| Therapeutic anticoagulation | | | | |

#### ICU readmission

| | |
|---|---|
| Population | mpRCT confirmed unblinded |
| Endpoint | ICU readmission during index hospitalization |
| Model | Dichotomous model |
| Factors | Intervention, age, sex, site, time, d-dimer (moderate) |
| Analysis | Conducted by the mpRCT analysis team |

The following posterior probabilities will be reported for each subtype being reported:

| **Quantity of Interest** | **Posterior Probability** |
|---|---|
| Therapeutic anticoagulation is superior to control | |
| Therapeutic anticoagulation is futile | |
| Therapeutic anticoagulation is inferior | |

The following will be reported:

| **Odds Ratio Parameter** | **Mean** | **SD** | **Median** | **95% Credible Interval** |
|---|---|---|---|---|
| Age < 39 | | | | |
| Age 40, 49 | | | | |
| Age 50, 59 | | | | |
| Age 70-79 | | | | |
| Age 80+ | | | | |
| D-dimer high (for moderate) | | | | |
| D-dimer missing (for moderate) | | | | |
| Female | | | | |
| Time epoch 1 | | | | |
| … | | | | |
| Time epoch k-1 | | | | |
| Therapeutic anticoagulation | | | | |

CONFIDENTIAL

#### Ischemic cerebrovascular event

| | |
|---|---|
| Population | mpRCT confirmed unblinded |
| Endpoint | Ischemic cerebrovascular event |
| Model | Dichotomous model |
| Factors | Intervention, age, sex, site, time, d-dimer (moderate) |
| Analysis | Conducted by the mpRCT analysis team |

The following posterior probabilities will be reported for each subtype being reported:

| **Quantity of Interest** | **Posterior Probability** |
|---|---|
| Therapeutic anticoagulation is superior to control | |
| Therapeutic anticoagulation is futile | |
| Therapeutic anticoagulation is inferior | |

The following will be reported:

| **Odds Ratio Parameter** | **Mean** | **SD** | **Median** | **95% Credible Interval** |
|---|---|---|---|---|
| Age < 39 | | | | |
| Age 40, 49 | | | | |
| Age 50, 59 | | | | |
| Age 70-79 | | | | |
| Age 80+ | | | | |
| D-dimer high (for moderate) | | | | |
| D-dimer missing (for moderate) | | | | |
| Female | | | | |
| Time epoch 1 | | | | |
| … | | | | |
| Time epoch k-1 | | | | |
| Therapeutic anticoagulation | | | | |

CONFIDENTIAL

#### Systemic arterial thromboembolism

| | |
|---|---|
| Population | mpRCT confirmed unblinded |
| Endpoint | Systemic arterial thromboembolism |
| Model | Dichotomous model |
| Factors | Intervention, age, sex, site, time, d-dimer (moderate) |
| Analysis | Conducted by the mpRCT analysis team |

The following posterior probabilities will be reported for each subtype being reported:

| **Quantity of Interest** | **Posterior Probability** |
|---|---|
| Therapeutic anticoagulation is superior to control | |
| Therapeutic anticoagulation is futile | |
| Therapeutic anticoagulation is inferior | |

The following will be reported:

| **Odds Ratio Parameter** | **Mean** | **SD** | **Median** | **95% Credible Interval** |
|---|---|---|---|---|
| Age < 39 | | | | |
| Age 40, 49 | | | | |
| Age 50, 59 | | | | |
| Age 70-79 | | | | |
| Age 80+ | | | | |
| D-dimer high (for moderate) | | | | |
| D-dimer missing (for moderate) | | | | |
| Female | | | | |
| Time epoch 1 | | | | |
| … | | | | |
| Time epoch k-1 | | | | |
| Therapeutic anticoagulation | | | | |

#### Acute myocardial infarction

| | |
|---|---|
| Population | mpRCT confirmed unblinded |
| Endpoint | Acute myocardial infarction |
| Model | Dichotomous model |
| Factors | Intervention, age, sex, site, time, d-dimer (moderate) |
| Analysis | Conducted by the mpRCT analysis team |

The following posterior probabilities will be reported for each subtype being reported:

| **Quantity of Interest** | **Posterior Probability** |
|---|---|
| Therapeutic anticoagulation is superior to control | |
| Therapeutic anticoagulation is futile | |
| Therapeutic anticoagulation is inferior | |

The following will be reported:

| **Odds Ratio Parameter** | **Mean** | **SD** | **Median** | **95% Credible Interval** |
|---|---|---|---|---|
| Age < 39 | | | | |
| Age 40, 49 | | | | |
| Age 50, 59 | | | | |
| Age 70-79 | | | | |
| Age 80+ | | | | |
| D-dimer high (for moderate) | | | | |
| D-dimer missing (for moderate) | | | | |
| Female | | | | |
| Time epoch 1 | | | | |
| … | | | | |
| Time epoch k-1 | | | | |
| Therapeutic anticoagulation | | | | |

#### World Health Organization Ordinal Scale

| | |
|---|---|
| Population | mpRCT confirmed unblinded |
| Endpoint | WHO scale on day 14 |
| Model | Ordinal model |
| Factors | Intervention, age, sex, site, time, d-dimer (moderate) |
| Analysis | Conducted by the mpRCT analysis team |

The following posterior probabilities will be reported for each subtype being reported:

| **Quantity of Interest** | **Posterior Probability** |
|---|---|
| Therapeutic anticoagulation is superior to control | |
| Therapeutic anticoagulation is futile | |
| Therapeutic anticoagulation is inferior | |

The following will be reported:

| **Odds Ratio Parameter** | **Mean** | **SD** | **Median** | **95% Credible Interval** |
|---|---|---|---|---|
| Age < 39 | | | | |
| Age 40, 49 | | | | |
| Age 50, 59 | | | | |
| Age 70-79 | | | | |
| Age 80+ | | | | |
| D-dimer high (for moderate) | | | | |
| D-dimer missing (for moderate) | | | | |
| Female | | | | |
| Time epoch 1 | | | | |
| … | | | | |
| Time epoch k-1 | | | | |
| Therapeutic anticoagulation | | | | |

CONFIDENTIAL

#### Safety analysis of major bleeding

| | |
|---|---|
| Population | mpRCT confirmed unblinded |
| Endpoint | Major bleeding by ISTH criteria |
| Model | Dichotomous model |
| Factors | Intervention, age, sex, site, time, d-dimer (moderate) |
| Analysis | Conducted by the mpRCT analysis team |

The following posterior probabilities will be reported for each subtype being reported:

| **Quantity of Interest** | **Posterior Probability** |
|---|---|
| Therapeutic anticoagulation is superior to control | |
| Therapeutic anticoagulation is futile | |
| Therapeutic anticoagulation is inferior | |

The following will be reported:

| **Odds Ratio Parameter** | **Mean** | **SD** | **Median** | **95% Credible Interval** |
|---|---|---|---|---|
| Age < 39 | | | | |
| Age 40, 49 | | | | |
| Age 50, 59 | | | | |
| Age 70-79 | | | | |
| Age 80+ | | | | |
| D-dimer high (for moderate) | | | | |
| D-dimer missing (for moderate) | | | | |
| Female | | | | |
| Time epoch 1 | | | | |
| … | | | | |
| Time epoch k-1 | | | | |
| Therapeutic anticoagulation | | | | |

#### Sensitivity analysis of major bleeding – exclude patients on antiplatelet therapy at baseline

| | |
|---|---|
| Population | mpRCT confirmed unblinded excluding patients on antiplatelet therapy at baseline or who are randomized in the antiplatelet domain of REMAP-CAP |
| Endpoint | Major bleeding by ISTH criteria |
| Model | Dichotomous model |
| Factors | Intervention, age, sex, site, time, d-dimer (moderate) |
| Analysis | Conducted by the mpRCT analysis team |

The following posterior probabilities will be reported for each subtype being reported:

| **Quantity of Interest** | **Posterior Probability** |
|---|---|
| Therapeutic anticoagulation is superior to control | |
| Therapeutic anticoagulation is futile | |
| Therapeutic anticoagulation is inferior | |

The following will be reported:

| **Odds Ratio Parameter** | **Mean** | **SD** | **Median** | **95% Credible Interval** |
|---|---|---|---|---|
| Age < 39 | | | | |
| Age 40, 49 | | | | |
| Age 50, 59 | | | | |
| Age 70-79 | | | | |
| Age 80+ | | | | |
| D-dimer high (for moderate) | | | | |
| D-dimer missing (for moderate) | | | | |
| Female | | | | |
| Time epoch 1 | | | | |
| … | | | | |
| Time epoch k-1 | | | | |
| Therapeutic anticoagulation | | | | |

CONFIDENTIAL

#### Safety analysis of laboratory-confirmed heparin-induced thrombocytopenia

| | |
|---|---|
| Population | mpRCT confirmed unblinded |
| Endpoint | Heparin-induced thrombocytopenia |
| Model | Dichotomous model |
| Factors | Intervention, age, sex, site, time, d-dimer (moderate) |
| Analysis | Conducted by the mpRCT analysis team |

The following posterior probabilities will be reported for each subtype being reported:

| **Quantity of Interest** | **Posterior Probability** |
|---|---|
| Therapeutic anticoagulation is superior to control | |
| Therapeutic anticoagulation is futile | |
| Therapeutic anticoagulation is inferior | |

The following will be reported:

| **Odds Ratio Parameter** | **Mean** | **SD** | **Median** | **95% Credible Interval** |
|---|---|---|---|---|
| Age < 39 | | | | |
| Age 40, 49 | | | | |
| Age 50, 59 | | | | |
| Age 70-79 | | | | |
| Age 80+ | | | | |
| D-dimer high (for moderate) | | | | |
| D-dimer missing (for moderate) | | | | |
| Female | | | | |
| Time epoch 1 | | | | |
| … | | | | |
| Time epoch k-1 | | | | |
| Therapeutic anticoagulation | | | | |

#### Safety analysis of fatal bleeding

| | |
|---|---|
| Population | mpRCT confirmed unblinded |
| Endpoint | Fatal bleeding (subcomponent of major bleeding event definition) |
| Model | Dichotomous model |
| Factors | Intervention, age, sex, site, time, d-dimer (moderate) |
| Analysis | Conducted by the mpRCT analysis team |

The following posterior probabilities will be reported for each subtype being reported:

| **Quantity of Interest** | **Posterior Probability** |
|---|---|
| Therapeutic anticoagulation is superior to control | |
| Therapeutic anticoagulation is futile | |
| Therapeutic anticoagulation is inferior | |

The following will be reported:

| **Odds Ratio Parameter** | **Mean** | **SD** | **Median** | **95% Credible Interval** |
|---|---|---|---|---|
| Age < 39 | | | | |
| Age 40, 49 | | | | |
| Age 50, 59 | | | | |
| Age 70-79 | | | | |
| Age 80+ | | | | |
| D-dimer high (for moderate) | | | | |
| D-dimer missing (for moderate) | | | | |
| Female | | | | |
| Time epoch 1 | | | | |
| … | | | | |
| Time epoch k-1 | | | | |
| Therapeutic anticoagulation | | | | |

CONFIDENTIAL

#### Safety analysis of symptomatic bleeding into critical organ

| | |
|---|---|
| Population | mpRCT confirmed unblinded |
| Endpoint | Symptomatic bleeding into critical organ (subcomponent of major bleeding event definition) |
| Model | Dichotomous model |
| Factors | Intervention, age, sex, site, time, d-dimer (moderate) |
| Analysis | Conducted by the mpRCT analysis team |

The following posterior probabilities will be reported for each subtype being reported:

| **Quantity of Interest** | **Posterior Probability** |
|---|---|
| Therapeutic anticoagulation is superior to control | |
| Therapeutic anticoagulation is futile | |
| Therapeutic anticoagulation is inferior | |

The following will be reported:

| **Odds Ratio Parameter** | **Mean** | **SD** | **Median** | **95% Credible Interval** |
|---|---|---|---|---|
| Age < 39 | | | | |
| Age 40, 49 | | | | |
| Age 50, 59 | | | | |
| Age 70-79 | | | | |
| Age 80+ | | | | |
| D-dimer high (for moderate) | | | | |
| D-dimer missing (for moderate) | | | | |
| Female | | | | |
| Time epoch 1 | | | | |
| … | | | | |
| Time epoch k-1 | | | | |
| Therapeutic anticoagulation | | | | |

#### Safety analysis of intracranial hemorrhage

| | |
|---|---|
| Population | mpRCT confirmed unblinded |
| Endpoint | Intracranial hemorrhage (subcomponent of major bleeding event definition) |
| Model | Dichotomous model |
| Factors | Intervention, age, sex, site, time, d-dimer (moderate) |
| Analysis | Conducted by the mpRCT analysis team |

The following posterior probabilities will be reported for each subtype being reported:

| **Quantity of Interest** | **Posterior Probability** |
|---|---|
| Therapeutic anticoagulation is superior to control | |
| Therapeutic anticoagulation is futile | |
| Therapeutic anticoagulation is inferior | |

The following will be reported:

| **Odds Ratio Parameter** | **Mean** | **SD** | **Median** | **95% Credible Interval** |
|---|---|---|---|---|
| Age < 39 | | | | |
| Age 40, 49 | | | | |
| Age 50, 59 | | | | |
| Age 70-79 | | | | |
| Age 80+ | | | | |
| D-dimer high (for moderate) | | | | |
| D-dimer missing (for moderate) | | | | |
| Female | | | | |
| Time epoch 1 | | | | |
| … | | | | |
| Time epoch k-1 | | | | |
| Therapeutic anticoagulation | | | | |

CONFIDENTIAL

#### Safety analysis of bleeding leading to a ≥20 g/L drop in hemoglobin

| | |
|---|---|
| Population | mpRCT confirmed unblinded |
| Endpoint | Bleeding to a ≥20 g/L drop in hemoglobin (subcomponent of major bleeding event definition) |
| Model | Dichotomous model |
| Factors | Intervention, age, sex, site, time, d-dimer (moderate) |
| Analysis | Conducted by the mpRCT analysis team |

The following posterior probabilities will be reported for each subtype being reported:

| **Quantity of Interest** | **Posterior Probability** |
|---|---|
| Therapeutic anticoagulation is superior to control | |
| Therapeutic anticoagulation is futile | |
| Therapeutic anticoagulation is inferior | |

The following will be reported:

| **Odds Ratio Parameter** | **Mean** | **SD** | **Median** | **95% Credible Interval** |
|---|---|---|---|---|
| Age < 39 | | | | |
| Age 40, 49 | | | | |
| Age 50, 59 | | | | |
| Age 70-79 | | | | |
| Age 80+ | | | | |
| D-dimer high (for moderate) | | | | |
| D-dimer missing (for moderate) | | | | |
| Female | | | | |
| Time epoch 1 | | | | |
| … | | | | |
| Time epoch k-1 | | | | |
| Therapeutic anticoagulation | | | | |

#### Safety analysis of bleeding leading to ≥2 red blood cell unit transfusion

| | |
|---|---|
| Population | mpRCT confirmed unblinded |
| Endpoint | Bleeding leading to a ≥2 red blood cell unit transfusion (subcomponent of major bleeding event definition) |
| Model | Dichotomous model |
| Factors | Intervention, age, sex, site, time, d-dimer (moderate) |
| Analysis | Conducted by the mpRCT analysis team |

The following posterior probabilities will be reported for each subtype being reported:

| **Quantity of Interest** | **Posterior Probability** |
|---|---|
| Therapeutic anticoagulation is superior to control | |
| Therapeutic anticoagulation is futile | |
| Therapeutic anticoagulation is inferior | |

The following will be reported:

| **Odds Ratio Parameter** | **Mean** | **SD** | **Median** | **95% Credible Interval** |
|---|---|---|---|---|
| Age < 39 | | | | |
| Age 40, 49 | | | | |
| Age 50, 59 | | | | |
| Age 70-79 | | | | |
| Age 80+ | | | | |
| D-dimer high (for moderate) | | | | |
| D-dimer missing (for moderate) | | | | |
| Female | | | | |
| Time epoch 1 | | | | |
| … | | | | |
| Time epoch k-1 | | | | |
| Therapeutic anticoagulation | | | | |

#### Per protocol analysis of OSFDs

| | |
|---|---|
| Population | mpRCT confirmed per protocol |
| Endpoint | Organ support-free days |
| Model | Primary analysis ordinal model |
| Factors | Intervention, age, sex, site, time, d-dimer (moderate) |
| Analysis | Conducted by the mpRCT analysis team |

The following posterior probabilities will be reported for each subtype being reported:

| **Quantity of Interest** | **Posterior Probability** |
|---|---|
| Therapeutic anticoagulation is superior to control | |
| Therapeutic anticoagulation is futile | |
| Therapeutic anticoagulation is inferior | |

The following will be reported:

| **Odds Ratio Parameter** | **Mean** | **SD** | **Median** | **95% Credible Interval** |
|---|---|---|---|---|
| Age < 39 | | | | |
| Age 40, 49 | | | | |
| Age 50, 59 | | | | |
| Age 70-79 | | | | |
| Age 80+ | | | | |
| D-dimer high (for moderate) | | | | |
| D-dimer missing (for moderate) | | | | |
| Female | | | | |
| Time epoch 1 | | | | |
| … | | | | |
| Time epoch k-1 | | | | |
| Therapeutic anticoagulation | | | | |

#### Per protocol analysis of in-hospital mortality

| | |
|---|---|
| Population | mpRCT confirmed per protocol |
| Endpoint | Hospital mortality (censored at day 90 or time of snapshot) |
| Model | Dichotomous model |
| Factors | Intervention, age, sex, site, time, d-dimer (moderate) |
| Analysis | Conducted by the mpRCT analysis team |

The following posterior probabilities will be reported for each subtype being reported:

| **Quantity of Interest** | **Posterior Probability** |
|---|---|
| Therapeutic anticoagulation is superior to control | |
| Therapeutic anticoagulation is futile | |
| Therapeutic anticoagulation is inferior | |

The following will be reported:

| **Odds Ratio Parameter** | **Mean** | **SD** | **Median** | **95% Credible Interval** |
|---|---|---|---|---|
| Age < 39 | | | | |
| Age 40, 49 | | | | |
| Age 50, 59 | | | | |
| Age 70-79 | | | | |
| Age 80+ | | | | |
| D-dimer high (for moderate) | | | | |
| D-dimer missing (for moderate) | | | | |
| Female | | | | |
| Time epoch 1 | | | | |
| … | | | | |
| Time epoch k-1 | | | | |
| Therapeutic anticoagulation | | | | |

CONFIDENTIAL

#### Per protocol analysis of major thrombotic events

| | |
|---|---|
| Population | mpRCT confirmed per protocol |
| Endpoint | Major thrombotic events or death |
| Model | Dichotomous model |
| Factors | Intervention, age, sex, site, time, d-dimer (moderate) |
| Analysis | Conducted by the mpRCT analysis team |

The following posterior probabilities will be reported for each subtype being reported:

| **Quantity of Interest** | **Posterior Probability** |
|---|---|
| Therapeutic anticoagulation is superior to control | |
| Therapeutic anticoagulation is futile | |
| Therapeutic anticoagulation is inferior | |

The following will be reported:

| **Odds Ratio Parameter** | **Mean** | **SD** | **Median** | **95% Credible Interval** |
|---|---|---|---|---|
| Age < 39 | | | | |
| Age 40, 49 | | | | |
| Age 50, 59 | | | | |
| Age 70-79 | | | | |
| Age 80+ | | | | |
| D-dimer high (for moderate) | | | | |
| D-dimer missing (for moderate) | | | | |
| Female | | | | |
| Time epoch 1 | | | | |
| … | | | | |
| Time epoch k-1 | | | | |
| Therapeutic anticoagulation | | | | |

#### Per protocol analysis of all thrombotic events

| | |
|---|---|
| Population | mpRCT confirmed per protocol |
| Endpoint | All thrombotic events or death |
| Model | Dichotomous model |
| Factors | Intervention, age, sex, site, time, d-dimer (moderate) |
| Analysis | Conducted by the mpRCT analysis team |

The following posterior probabilities will be reported for each subtype being reported:

| **Quantity of Interest** | **Posterior Probability** |
|---|---|
| Therapeutic anticoagulation is superior to control | |
| Therapeutic anticoagulation is futile | |
| Therapeutic anticoagulation is inferior | |

The following will be reported:

| **Odds Ratio Parameter** | **Mean** | **SD** | **Median** | **95% Credible Interval** |
|---|---|---|---|---|
| Age < 39 | | | | |
| Age 40, 49 | | | | |
| Age 50, 59 | | | | |
| Age 70-79 | | | | |
| Age 80+ | | | | |
| D-dimer high (for moderate) | | | | |
| D-dimer missing (for moderate) | | | | |
| Female | | | | |
| Time epoch 1 | | | | |
| … | | | | |
| Time epoch k-1 | | | | |
| Therapeutic anticoagulation | | | | |

#### Per protocol analysis of deep venous thrombosis

| | |
|---|---|
| Population | mpRCT confirmed per protocol |
| Endpoint | Deep venous thrombosis |
| Model | Dichotomous model |
| Factors | Intervention, age, sex, site, time, d-dimer (moderate) |
| Analysis | Conducted by the mpRCT analysis team |

The following posterior probabilities will be reported for each subtype being reported:

| **Quantity of Interest** | **Posterior Probability** |
|---|---|
| Therapeutic anticoagulation is superior to control | |
| Therapeutic anticoagulation is futile | |
| Therapeutic anticoagulation is inferior | |

The following will be reported:

| **Odds Ratio Parameter** | **Mean** | **SD** | **Median** | **95% Credible Interval** |
|---|---|---|---|---|
| Age < 39 | | | | |
| Age 40, 49 | | | | |
| Age 50, 59 | | | | |
| Age 70-79 | | | | |
| Age 80+ | | | | |
| D-dimer high (for moderate) | | | | |
| D-dimer missing (for moderate) | | | | |
| Female | | | | |
| Time epoch 1 | | | | |
| … | | | | |
| Time epoch k-1 | | | | |
| Therapeutic anticoagulation | | | | |

CONFIDENTIAL

#### Per protocol analysis of pulmonary embolism

| | |
|---|---|
| Population | mpRCT confirmed per protocol |
| Endpoint | Acute pulmonary embolism |
| Model | Dichotomous model |
| Factors | Intervention, age, sex, site, time, d-dimer (moderate) |
| Analysis | Conducted by the mpRCT analysis team |

The following posterior probabilities will be reported for each subtype being reported:

| **Quantity of Interest** | **Posterior Probability** |
|---|---|
| Therapeutic anticoagulation is superior to control | |
| Therapeutic anticoagulation is futile | |
| Therapeutic anticoagulation is inferior | |

The following will be reported:

| **Odds Ratio Parameter** | **Mean** | **SD** | **Median** | **95% Credible Interval** |
|---|---|---|---|---|
| Age < 39 | | | | |
| Age 40, 49 | | | | |
| Age 50, 59 | | | | |
| Age 70-79 | | | | |
| Age 80+ | | | | |
| D-dimer high (for moderate) | | | | |
| D-dimer missing (for moderate) | | | | |
| Female | | | | |
| Time epoch 1 | | | | |
| … | | | | |
| Time epoch k-1 | | | | |
| Therapeutic anticoagulation | | | | |

#### Per protocol analysis of 90-day mortality

| | |
|---|---|
| Population | mpRCT per protocol |
| Endpoint | Mortality at 90 days |
| Model | Time to event model |
| Factors | Intervention, age, sex, site, time, d-dimer (moderate) |
| Analysis | Conducted by the mpRCT analysis team |

The following posterior probabilities will be reported for each subtype being reported:

| **Quantity of Interest** | **Posterior Probability** |
|---|---|
| Therapeutic anticoagulation is superior to control | |
| Therapeutic anticoagulation is futile | |
| Therapeutic anticoagulation is inferior | |

The following will be reported:

| **Odds Ratio Parameter** | **Mean** | **SD** | **Median** | **95% Credible Interval** |
|---|---|---|---|---|
| Age < 39 | | | | |
| Age 40, 49 | | | | |
| Age 50, 59 | | | | |
| Age 70-79 | | | | |
| Age 80+ | | | | |
| D-dimer high (for moderate) | | | | |
| D-dimer missing (for moderate) | | | | |
| Female | | | | |
| Time epoch 1 | | | | |
| … | | | | |
| Time epoch k-1 | | | | |
| Therapeutic anticoagulation | | | | |

#### Per protocol analysis of days free of vasopressors or inotropes

| | |
|---|---|
| Population | mpRCT per protocol |
| Endpoint | Days free of vasopressors and inotropes to day 28 |
| Model | Primary analysis ordinal model |
| Factors | Intervention, age, sex, site, time, d-dimer (moderate) |
| Analysis | Conducted by the mpRCT analysis team |

The following posterior probabilities will be reported for each subtype being reported:

| **Quantity of Interest** | **Posterior Probability** |
|---|---|
| Therapeutic anticoagulation is superior to control | |
| Therapeutic anticoagulation is futile | |
| Therapeutic anticoagulation is inferior | |

The following will be reported:

| **Odds Ratio Parameter** | **Mean** | **SD** | **Median** | **95% Credible Interval** |
|---|---|---|---|---|
| Age < 39 | | | | |
| Age 40, 49 | | | | |
| Age 50, 59 | | | | |
| Age 70-79 | | | | |
| Age 80+ | | | | |
| D-dimer high (for moderate) | | | | |
| D-dimer missing (for moderate) | | | | |
| Female | | | | |
| Time epoch 1 | | | | |
| … | | | | |
| Time epoch k-1 | | | | |
| Therapeutic anticoagulation | | | | |

#### Per protocol analysis of days free of invasive or non-invasive ventilation

| | |
|---|---|
| Population | mpRCT per protocol |
| Endpoint | Days free of invasive or non-invasive ventilation to day 28 |
| Model | Primary analysis ordinal model |
| Factors | Intervention, age, sex, site, time, d-dimer (moderate) |
| Analysis | Conducted by the mpRCT analysis team |

The following posterior probabilities will be reported for each subtype being reported:

| **Quantity of Interest** | **Posterior Probability** |
|---|---|
| Therapeutic anticoagulation is superior to control | |
| Therapeutic anticoagulation is futile | |
| Therapeutic anticoagulation is inferior | |

The following will be reported:

| **Odds Ratio Parameter** | **Mean** | **SD** | **Median** | **95% Credible Interval** |
|---|---|---|---|---|
| Age < 39 | | | | |
| Age 40, 49 | | | | |
| Age 50, 59 | | | | |
| Age 70-79 | | | | |
| Age 80+ | | | | |
| D-dimer high (for moderate) | | | | |
| D-dimer missing (for moderate) | | | | |
| Female | | | | |
| Time epoch 1 | | | | |
| … | | | | |
| Time epoch k-1 | | | | |
| Therapeutic anticoagulation | | | | |

#### Per protocol analysis of days free of renal-replacement therapy

| | |
|---|---|
| Population | mpRCT per protocol |
| Endpoint | Days free of renal replacement therapy to day 28 |
| Model | Primary analysis ordinal model |
| Factors | Intervention, age, sex, site, time, d-dimer (moderate) |
| Analysis | Conducted by the mpRCT analysis team |

The following posterior probabilities will be reported for each subtype being reported:

| **Quantity of Interest** | **Posterior Probability** |
|---|---|
| Therapeutic anticoagulation is superior to control | |
| Therapeutic anticoagulation is futile | |
| Therapeutic anticoagulation is inferior | |

The following will be reported:

| **Odds Ratio Parameter** | **Mean** | **SD** | **Median** | **95% Credible Interval** |
|---|---|---|---|---|
| Age < 39 | | | | |
| Age 40, 49 | | | | |
| Age 50, 59 | | | | |
| Age 70-79 | | | | |
| Age 80+ | | | | |
| D-dimer high (for moderate) | | | | |
| D-dimer missing (for moderate) | | | | |
| Female | | | | |
| Time epoch 1 | | | | |
| … | | | | |
| Time epoch k-1 | | | | |
| Therapeutic anticoagulation | | | | |

CONFIDENTIAL

#### ECMO utilization

| | |
|---|---|
| Population | mpRCT per protocol excluding patients on ECMO at baseline |
| Endpoint | ECMO utilization to day 28 |
| Model | Primary analysis dichotomous model |
| Factors | Intervention, age, sex, site, time, d-dimer (moderate) |
| Analysis | Conducted by the mpRCT analysis team |

The following posterior probabilities will be reported for each subtype being reported:

| Quantity of Interest | Posterior Probability |
|---|---|
| Therapeutic anticoagulation is superior to control | |
| Therapeutic anticoagulation is futile | |
| Therapeutic anticoagulation is inferior | |

The following will be reported:

| Odds Ratio Parameter | Mean | SD | Median | 95% Credible Interval |
|---|---|---|---|---|
| Age < 39 | | | | |
| Age 40, 49 | | | | |
| Age 50, 59 | | | | |
| Age 70-79 | | | | |
| Age 80+ | | | | |
| D-dimer high (for moderate) | | | | |
| D-dimer missing (for moderate) | | | | |
| Female | | | | |
| Time epoch 1 | | | | |
| … | | | | |
| Time epoch k-1 | | | | |
| Therapeutic anticoagulation | | | | |

#### Per protocol analysis of ICU length-of-stay

| | |
|---|---|
| Population | mpRCT per protocol |
| Endpoint | ICU length-of-stay (event=ICU discharge); patients who die in hospital are assigned a value of 90 days |
| Model | Primary time-to-event model |
| Factors | Intervention, age, sex, site, time, d-dimer (moderate) |
| Analysis | Conducted by the mpRCT analysis team |

The following posterior probabilities will be reported for each subtype being reported:

| Quantity of Interest | Posterior Probability |
|---|---|
| Therapeutic anticoagulation is superior to control | |
| Therapeutic anticoagulation is futile | |
| Therapeutic anticoagulation is inferior | |

The following will be reported:

| Odds Ratio Parameter | Mean | SD | Median | 95% Credible Interval |
|---|---|---|---|---|
| Age < 39 | | | | |
| Age 40, 49 | | | | |
| Age 50, 59 | | | | |
| Age 70-79 | | | | |
| Age 80+ | | | | |
| D-dimer high (for moderate) | | | | |
| D-dimer missing (for moderate) | | | | |
| Female | | | | |
| Time epoch 1 | | | | |
| … | | | | |
| Time epoch k-1 | | | | |
| Therapeutic anticoagulation | | | | |

CONFIDENTIAL

#### Per protocol analysis of hospital length-of-stay

| | |
|---|---|
| Population | mpRCT per protocol |
| Endpoint | Hospital length-of-stay (event=hospital discharge); patients who die in hospital are assigned a value of 90 days |
| Model | Primary time-to-event model |
| Factors | Intervention, age, sex, site, time, d-dimer (moderate) |
| Analysis | Conducted by the mpRCT analysis team |

The following posterior probabilities will be reported for each subtype being reported:

| Quantity of Interest | Posterior Probability |
|---|---|
| Therapeutic anticoagulation is superior to control | |
| Therapeutic anticoagulation is futile | |
| Therapeutic anticoagulation is inferior | |

The following will be reported:

| Odds Ratio Parameter | Mean | SD | Median | 95% Credible Interval |
|---|---|---|---|---|
| Age < 39 | | | | |
| Age 40, 49 | | | | |
| Age 50, 59 | | | | |
| Age 70-79 | | | | |
| Age 80+ | | | | |
| D-dimer high (for moderate) | | | | |
| D-dimer missing (for moderate) | | | | |
| Female | | | | |
| Time epoch 1 | | | | |
| … | | | | |
| Time epoch k-1 | | | | |
| Therapeutic anticoagulation | | | | |

#### ICU readmission

| | |
|---|---|
| Population | mpRCT per protocol |
| Endpoint | ICU readmission during index hospitalization |
| Model | Dichotomous model |
| Factors | Intervention, age, sex, site, time, d-dimer (moderate) |
| Analysis | Conducted by the mpRCT analysis team |

The following posterior probabilities will be reported for each subtype being reported:

| Quantity of Interest | Posterior Probability |
|---|---|
| Therapeutic anticoagulation is superior to control | |
| Therapeutic anticoagulation is futile | |
| Therapeutic anticoagulation is inferior | |

The following will be reported:

| Odds Ratio Parameter | Mean | SD | Median | 95% Credible Interval |
|---|---|---|---|---|
| Age < 39 | | | | |
| Age 40, 49 | | | | |
| Age 50, 59 | | | | |
| Age 70-79 | | | | |
| Age 80+ | | | | |
| D-dimer high (for moderate) | | | | |
| D-dimer missing (for moderate) | | | | |
| Female | | | | |
| Time epoch 1 | | | | |
| … | | | | |
| Time epoch k-1 | | | | |
| Therapeutic anticoagulation | | | | |

CONFIDENTIAL

#### Per protocol analysis of ischemic cerebrovascular event

| | |
|---|---|
| Population | mpRCT per protocol |
| Endpoint | Ischemic cerebrovascular event |
| Model | Dichotomous model |
| Factors | Intervention, age, sex, site, time, d-dimer (moderate) |
| Analysis | Conducted by the mpRCT analysis team |

The following posterior probabilities will be reported for each subtype being reported:

| **Quantity of Interest** | **Posterior Probability** |
|---|---|
| Therapeutic anticoagulation is superior to control | |
| Therapeutic anticoagulation is futile | |
| Therapeutic anticoagulation is inferior | |

The following will be reported:

| **Odds Ratio Parameter** | **Mean** | **SD** | **Median** | **95% Credible Interval** |
|---|---|---|---|---|
| Age < 39 | | | | |
| Age 40, 49 | | | | |
| Age 50, 59 | | | | |
| Age 70-79 | | | | |
| Age 80+ | | | | |
| D-dimer high (for moderate) | | | | |
| D-dimer missing (for moderate) | | | | |
| Female | | | | |
| Time epoch 1 | | | | |
| … | | | | |
| Time epoch k-1 | | | | |
| Therapeutic anticoagulation | | | | |

#### Per protocol analysis of systemic arterial thromboembolism

| | |
|---|---|
| Population | mpRCT per protocol |
| Endpoint | Systemic arterial thromboembolism |
| Model | Dichotomous model |
| Factors | Intervention, age, sex, site, time, d-dimer (moderate) |
| Analysis | Conducted by the mpRCT analysis team |

The following posterior probabilities will be reported for each subtype being reported:

| **Quantity of Interest** | **Posterior Probability** |
|---|---|
| Therapeutic anticoagulation is superior to control | |
| Therapeutic anticoagulation is futile | |
| Therapeutic anticoagulation is inferior | |

The following will be reported:

| **Odds Ratio Parameter** | **Mean** | **SD** | **Median** | **95% Credible Interval** |
|---|---|---|---|---|
| Age < 39 | | | | |
| Age 40, 49 | | | | |
| Age 50, 59 | | | | |
| Age 70-79 | | | | |
| Age 80+ | | | | |
| D-dimer high (for moderate) | | | | |
| D-dimer missing (for moderate) | | | | |
| Female | | | | |
| Time epoch 1 | | | | |
| … | | | | |
| Time epoch k-1 | | | | |
| Therapeutic anticoagulation | | | | |

#### Per protocol analysis of acute myocardial infarction

| | |
|---|---|
| Population | mpRCT per protocol |
| Endpoint | Acute myocardial infarction |
| Model | Dichotomous model |
| Factors | Intervention, age, sex, site, time, d-dimer (moderate) |
| Analysis | Conducted by the mpRCT analysis team |

The following posterior probabilities will be reported for each subtype being reported:

| **Quantity of Interest** | **Posterior Probability** |
|---|---|
| Therapeutic anticoagulation is superior to control | |
| Therapeutic anticoagulation is futile | |
| Therapeutic anticoagulation is inferior | |

The following will be reported:

| **Odds Ratio Parameter** | **Mean** | **SD** | **Median** | **95% Credible Interval** |
|---|---|---|---|---|
| Age < 39 | | | | |
| Age 40, 49 | | | | |
| Age 50, 59 | | | | |
| Age 70-79 | | | | |
| Age 80+ | | | | |
| D-dimer high (for moderate) | | | | |
| D-dimer missing (for moderate) | | | | |
| Female | | | | |
| Time epoch 1 | | | | |
| … | | | | |
| Time epoch k-1 | | | | |
| Therapeutic anticoagulation | | | | |

CONFIDENTIAL

#### Per protocol analysis of World Health Organization Ordinal Scale

| | |
|---|---|
| Population | mpRCT per protocol |
| Endpoint | WHO scale on day 14 |
| Model | Ordinal model |
| Factors | Intervention, age, sex, site, time, d-dimer (moderate) |
| Analysis | Conducted by the mpRCT analysis team |

The following posterior probabilities will be reported for each subtype being reported:

| **Quantity of Interest** | **Posterior Probability** |
|---|---|
| Therapeutic anticoagulation is superior to control | |
| Therapeutic anticoagulation is futile | |
| Therapeutic anticoagulation is inferior | |

The following will be reported:

| **Odds Ratio Parameter** | **Mean** | **SD** | **Median** | **95% Credible Interval** |
|---|---|---|---|---|
| Age < 39 | | | | |
| Age 40, 49 | | | | |
| Age 50, 59 | | | | |
| Age 70-79 | | | | |
| Age 80+ | | | | |
| D-dimer high (for moderate) | | | | |
| D-dimer missing (for moderate) | | | | |
| Female | | | | |
| Time epoch 1 | | | | |
| … | | | | |
| Time epoch k-1 | | | | |
| Therapeutic anticoagulation | | | | |

#### Prospectively defined subgroup analyses

##### Age

*OSFDs by age subgroup*

| | |
|---|---|
| Population | mpRCT confirmed unblinded |
| Endpoint | Organ support-free days |
| Model | Primary analysis ordinal model |
| Subgroups | Age categories (<50, 50-70, >70 years) |
| Factors | Age, sex, site, time, d-dimer (moderate) |
| Analysis | Conducted by the mpRCT analysis team |

The following posterior probabilities will be reported

| **Quantity of Interest** | **Posterior Probability** | | |
|---|---|---|---|
| | Superiority | Futility | Inferiority |
| Therapeutic anticoagulation in 1st age subgroup | | | |
| Therapeutic anticoagulation in 2nd age subgroup | | | |
| Therapeutic anticoagulation in 3rd age subgroup | | | |
| Probability effect in 1st group > 2nd group | | | |
| Probability effect in 1st group > 3rd group | | | |
| Probability effect in 2nd group > 3rd group | | | |

The following will be reported:

| **Odds ratio Parameter** | **Mean** | **SD** | **Median** | **95% Credible Interval** |
|---|---|---|---|---|
| Age < 39 | | | | |
| Age 40, 49 | | | | |
| Age 50, 59 | | | | |
| Age 70-79 | | | | |
| Age 80+ | | | | |
| Female | | | | |
| Time Epoch 1 | | | | |
| … | | | | |
| Time Epoch k-1 | | | | |
| Therapeutic anticoagulation in 1st age subgroup | | | | |
| Therapeutic anticoagulation in 2nd age subgroup | | | | |
| Therapeutic anticoagulation in 3rd age subgroup | | | | |

*In-hospital mortality by age subgroup*

| | |
|---|---|
| Population | mpRCT confirmed unblinded |
| Endpoint | In-hospital mortality |
| Model | Dichotomous model |
| Subgroups | Age categories (<50, 50-70, >70 years) |
| Factors | Age, sex, site, time, d-dimer (moderate) |
| Analysis | Conducted by the mpRCT analysis team |

The following posterior probabilities will be reported

| **Quantity of Interest** | **Posterior Probability** | | |
|---|---|---|---|
| | Superiority | Futility | Inferiority |
| Therapeutic anticoagulation in 1st age subgroup | | | |
| Therapeutic anticoagulation in 2nd age subgroup | | | |
| Therapeutic anticoagulation in 3rd age subgroup | | | |
| Probability effect in 1st group > 2nd group | | | |
| Probability effect in 1st group > 3rd group | | | |
| Probability effect in 2nd group > 3rd group | | | |

The following will be reported:

| **Odds ratio Parameter** | **Mean** | **SD** | **Median** | **95% Credible Interval** |
|---|---|---|---|---|
| Age < 39 | | | | |
| Age 40, 49 | | | | |
| Age 50, 59 | | | | |
| Age 70-79 | | | | |
| Age 80+ | | | | |
| Female | | | | |
| Time Epoch 1 | | | | |
| … | | | | |
| Time Epoch k-1 | | | | |
| Therapeutic anticoagulation in 1st age subgroup | | | | |
| Therapeutic anticoagulation in 2nd age subgroup | | | | |
| Therapeutic anticoagulation in 3rd age subgroup | | | | |

CONFIDENTIAL

*Major thrombotic events by age subgroup*

| | |
|---|---|
| Population | mpRCT confirmed unblinded |
| Endpoint | Major thrombotic events or death |
| Model | Dichotomous model |
| Subgroups | Age categories (<50, 50-70, >70 years) |
| Factors | Age, sex, site, time, d-dimer (moderate) |
| Analysis | Conducted by the mpRCT analysis team |

The following posterior probabilities will be reported

| **Quantity of Interest** | **Posterior Probability** | | |
|---|---|---|---|
| | Superiority | Futility | Inferiority |
| Therapeutic anticoagulation in 1st age subgroup | | | |
| Therapeutic anticoagulation in 2nd age subgroup | | | |
| Therapeutic anticoagulation in 3rd age subgroup | | | |
| Probability effect in 1st group > 2nd group | | | |
| Probability effect in 1st group > 3rd group | | | |
| Probability effect in 2nd group > 3rd group | | | |

The following will be reported:

| **Odds ratio Parameter** | **Mean** | **SD** | **Median** | **95% Credible Interval** |
|---|---|---|---|---|
| Age < 39 | | | | |
| Age 40, 49 | | | | |
| Age 50, 59 | | | | |
| Age 70-79 | | | | |
| Age 80+ | | | | |
| Female | | | | |
| Time Epoch 1 | | | | |
| … | | | | |
| Time Epoch k-1 | | | | |
| Therapeutic anticoagulation in 1st age subgroup | | | | |
| Therapeutic anticoagulation in 2nd age subgroup | | | | |
| Therapeutic anticoagulation in 3rd age subgroup | | | | |

CONFIDENTIAL

##### *Major bleeding by age subgroup*

| | |
|---|---|
| Population | mpRCT confirmed unblinded |
| Endpoint | Major bleeding |
| Model | Dichotomous model |
| Subgroups | Age categories (<50, 50-70, >70 years) |
| Factors | Age, sex, site, time, d-dimer (moderate) |
| Analysis | Conducted by the mpRCT analysis team |

The following posterior probabilities will be reported

| **Quantity of Interest** | **Posterior Probability** | | |
|---|---|---|---|
| | Superiority | Futility | Inferiority |
| Therapeutic anticoagulation in 1st age subgroup | | | |
| Therapeutic anticoagulation in 2nd age subgroup | | | |
| Therapeutic anticoagulation in 3rd age subgroup | | | |
| Probability effect in 1st group > 2nd group | | | |
| Probability effect in 1st group > 3rd group | | | |
| Probability effect in 2nd group > 3rd group | | | |

The following will be reported:

| **Odds ratio Parameter** | **Mean** | **SD** | **Median** | **95% Credible Interval** |
|---|---|---|---|---|
| Age < 39 | | | | |
| Age 40, 49 | | | | |
| Age 50, 59 | | | | |
| Age 70-79 | | | | |
| Age 80+ | | | | |
| Female | | | | |
| Time Epoch 1 | | | | |
| … | | | | |
| Time Epoch k-1 | | | | |
| Therapeutic anticoagulation in 1st age subgroup | | | | |
| Therapeutic anticoagulation in 2nd age subgroup | | | | |
| Therapeutic anticoagulation in 3rd age subgroup | | | | |

#### Other exploratory subgroups of interest

Each model output for the additional subgroup analyses in Section 15, Tables 2 and 3 follow the forms in 15.1.1, 15.1.2, 15.1.3, and 15.1.4.

#### Graphical summaries

1. All ordinal endpoints will be graphed using stacked cumulative bar plots
2. All time-to-event endpoints will be plotted using Kaplan-Meier plots
3. All dichotomous endpoints will be plotted using bar plots
4. Thrombotic events will be plotted using Kaplan-Meier plots

### Categorization of frequently used low- and intermediate-heparin doses in the ATTACC, ACTIV-4a, and REMAP-CAP mpRCT

Drugs/doses for prophylaxis in the mpRCT falling outside of those below are manually categorized, considering participant body weight/BMI and renal function, informed by American Society of Hematology and NICE guidance.

#### Subcutaneous Enoxaparin

##### Low dose

- *Standard dose*: 40mg once daily
- *Possible*: If BMI ≥ 40 kg/m2 (or weight ≥ 120 kg) and CrCl ≥ 30 mL/min: 40mg BID
- *Possible*: If CrCl < 30 mL/min: 30 mg once daily

##### Intermediate dose

###### *Twice daily*

- *Standard dose* up to and including (a) 0.5 mg/kg twice daily + 10% (rounding factor), or (b) 40 mg twice daily (whichever is higher)
- *Possible*: If BMI ≥ 40 kg/m2 (or weight ≥ 120 kg) and CrCl ≥ 30 mL/min: up to 60 mg twice daily
- *Possible*: If CrCl < 30 mL/min: intermediate twice daily dose not defined

###### *Daily*

- *Standard dose*: up to 1.0 mg/kg once daily + 10% (rounding factor)
- *Possible*: If BMI ≥ 40 kg/m2 (or weight ≥ 120 kg) and CrCl ≥ 30 mL/min: up to 0.8 mg/kg once daily + 10%
- *Possible*: If CrCl < 30 mL/min (and weight ≥ 60 kg): up to 0.5 mg/kg once daily +10%

#### Subcutaneous Dalteparin

##### Low dose

- *Standard dose*: 5,000 units once daily
- *Possible*: If BMI ≥ 40 kg/m2 (or weight ≥120 kg) and CrCl ≥ 30 mL/min: 7,500 units once daily

##### Intermediate dose

- *Standard dose*: 5,000 units twice daily
- *Possible*: If BMI ≥ 40 kg/m2 (or weight ≥ 120 kg) and CrCl ≥ 30 mL/min: 7,500 units twice daily

#### Subcutaneous Tinzaparin

##### Low dose

- *Standard dose*: up to and including (a) 75 anti-Xa units/kg + 10% (rounding factor) once daily or (b) 4,500 units once daily (whichever is higher)
- *Possible*: If BMI ≥ 40 kg/m2 (or weight ≥ 120 kg) and CrCl ≥ 30 mL/min: 8,000 units once daily

##### Intermediate dose

- *Standard dose*: 4,500 units twice daily

- o *Possible*: If BMI ≥ 40 kg/m2 (or weight ≥ 120 kg) and CrCl ≥ 30 mL/min: 8,000 units twice daily

#### Subcutaneous Unfractionated heparin

##### Low dose

- o *Standard dose*: 5,000 units twice or three times daily
- o *Possible*: If BMI ≥ 40 kg/m2 (or weight ≥ 120 kg) and CrCl ≥ 30 mL/min: 7,500 units twice daily

##### Intermediate dose

- o *Standard dose*: 7,500 units three times daily or 10,000 units two times daily
- o *Possible*: If BMI ≥ 40 kg/m2 (or weight ≥ 120 kg) and CrCl ≥ 30 mL/min: 10,000 units twice daily

### Statistical Analysis Plan for Preliminary Report of Severe Subtype of mpRCT of Therapeutic Anticoagulation in Covid-19

#### Version

Version 1.0, initialized January 5, 2021, finalized January 12, 2021

#### Background

This document is an ancillary document to the statistical analysis protocol (Version 1.0, dated January 5, 2020) for the mpRCT of therapeutic anticoagulation in Covid-19. It outlines the planned analyses required for the urgent preliminary reporting of trial results in the severe Covid-19 subtype.

Enrolment in the mpRCT severe state was halted on December 19, 2020 following a recommendation from the data safety and monitoring boards of the three platforms based on a statistical trigger reached during interim analysis. The investigators aim to prepare a preliminary report that details key findings of the trial in the severe state for broad dissemination followed by a subsequent comprehensive report.

The statistical analysis protocol for the mpRCT stipulates that primary endpoints and key secondary endpoints (major thrombotic events, major bleeding) must be included in preliminary reports. Limited subgroup analyses may also be reported depending on the available data. This supplementary statistical analysis plan lists the planned analyses for this preliminary report; all of these analyses are listed and described in detail in the mpRCT SAP.

#### Unblinded Population

mpRCT Covid-19 severe subtype will be reported in this sub-SAP report. The unblinded population defined in the SAP is the severe subtype.

#### Data

Data used for this preliminary report will be available for the set of patients analyzed by the unblinded SAC for interim analysis of the primary mpRCT statistical model on January 4, 2021. This analysis will only include patients randomized through the stop of randomization to the TAC arm in the severe state (on December 19, 2020) for whom the primary endpoint was available on January 4, 2021. Information on baseline characteristics, secondary endpoints, and subgroup classification for this set of patients becoming available after January 4, 2021 will be included in the preliminary report where possible.

#### Planned Analyses

##### Primary

| # | Status | Population | Endpoint | Notes |
|---|---|---|---|---|
| 14.1 | Primary | mpRCT confirmed | OSFDs | Primary ordinal model |
| 14.2 | Primary | mpRCT confirmed | In-hospital mortality | Primary dichotomous model |

##### Sensitivity analyses of the primary models

| # | Status | Population | Endpoint | Notes |
|---|---|---|---|---|
| 14.3 | Sensitivity | mpRCT confirmed | Dichotomized OSFD | Primary dichotomous model for each dichotomization of OSFDs as a robustness check. |
| 14.4 | Sensitivity | mpRCT confirmed unblinded | OSFDs | Primary ordinal model |
| 14.5 | Sensitivity | mpRCT confirmed unblinded | In-hospital mortality | Primary dichotomous model |

| | | | | |
|---|---|---|---|---|
| 14.6 | Sensitivity | mpRCT confirmed and suspected unblinded | OSFDs | Include REMAP-CAP suspected but not proven COVID-19 patients |
| 14.7 | Sensitivity | mpRCT confirmed and suspected unblinded | In-hospital mortality | Include REMAP-CAP suspected but not proven COVID-19 patients |
| 14.8 | Sensitivity | mpRCT confirmed unblinded | OSFDs | Remove site and time effects |
| 14.9 | Sensitivity | mpRCT confirmed unblinded | In-hospital mortality | Remove site and time effects |
| 14.10 | Sensitivity | mpRCT confirmed unblinded | OSFDs | Excluding patients who received antiplatelet agents at baseline or who are randomized in the antiplatelet domain in REMAP-CAP |
| 14.11 | Sensitivity | mpRCT confirmed unblinded | In-hospital mortality | Excluding patients who received antiplatelet agents at baseline or who are randomized in the antiplatelet domain in REMAP-CAP |
| 14.12 | Exploratory sensitivity analysis: Severe State only | mpRCT confirmed unblinded | OSFDs | Specifies prior for TAC for enthusiasm [N(0.56,0.44)] and prior for skepticism [N(0, 0.44)] |

#### Key secondary and safety endpoints

| # | Status | Population | Endpoint | Notes |
|---|---|---|---|---|
| 14.14 | Secondary | mpRCT confirmed unblinded | Major thrombotic events or death | Primary dichotomous model |
| 14.30 | Safety | mpRCT confirmed unblinded | Major bleeding | Primary dichotomous model |
| 14.31 | Safety sensitivity analysis | mpRCT confirmed unblinded | Major bleeding | Primary dichotomous model<br><br>Excluding patients who received antiplatelet agents at baseline or who are randomized into the antiplatelet domain |

- Will report type of major thrombotic event descriptively

#### Per protocol analyses

| # | Status | Population | Endpoint | Notes |
|---|---|---|---|---|
| 14.38 | Sensitivity | mpRCT per protocol | OSFDs | Primary ordinal model |
| 14.39 | Sensitivity | mpRCT per protocol | In-hospital mortality | Primary dichotomous model |
| 14.40 | Sensitivity | mpRCT per protocol | Major thrombotic events or death | Primary dichotomous model |

#### Subgroup analyses

Reporting of these subgroup analyses will be contingent on the completeness of data on the subgroup variable. This decision will be at the discretion of the investigators and contingent on the completeness of data on the subgroup variable.

| Subgroup | Specification of covariate | Endpoint – Model # | | | |
|---|---|---|---|---|---|
| | | OSFDs (efficacy) | Hospital mortality (efficacy) | Major thrombotic event or death (efficacy) | Major bleeding (safety) |

| | | | | | |
|---|---|---|---|---|---|
| Age | Categorical (<50 years, 50-70 years, and >70 years) | 15.1.1 | 15.1.2 | 15.1.3 | 15.1.4 |
| Sex | Dichotomous | 15.2.1 | 15.2.2 | 15.2.3 | 15.2.4 |
| Invasive mechanical ventilation at baseline* | Dichotomous | 15.3.1 | 15.3.2 | 15.3.3 | 15.3.4 |
| Antiplatelet agent use at baseline in hospital at time of randomization | Dichotomous | 15.4.1 | 15.4.2 | 15.4.3 | 15.4.4 |
| Usual care practice: low vs intermediate (site classification strategy) | Dichotomous | 16.15.1 | 16.15.2 | 16.15.3 | 16.15.4 |
| Usual care practice: low vs intermediate (day 1 patient classification strategy) | Dichotomous | 16.16.1 | 16.16.2 | 16.16.3 | 16.16.4 |

#### Required Variables for Severe State Patients

The following list of required variables is derived from the endpoints and subgroup variables listed above and the covariates listed in the Statistical Analysis Plan. Endpoints are defined in the Statistical Analysis Plan. Further work is required to define how these variables are defined in each platform. The following is the list of data needed for completing this sub-SAP.

The following data would be provided to Berry Consultants blinded Analysis Team for all analyses except 14.1, 14.2, and 14.3, which will be conducted by the Statistical Analysis Committee (SAC).

The following outcomes would be provided for every patient randomized to either VTP or TAC in the severe state that has not removed consent for data.

**Table 1.** Patient-level variables required for analysis

| Variable | Format | Variable Name by Platform | | |
|---|---|---|---|---|
| | | REMAP-CAP | ACTIV-4a | ATTACC |
| Age | Numeric | | | |
| Sex | M/F | | | |
| Site | Label/number | | | |
| Date of randomization | Any date format | | | |
| Randomization arm (TAC vs. VTP) | Any code for TAC/VTP | | | |
| Laboratory-confirmed Covid-19 status (proven vs. suspected) | 1 = proven; 0 = suspected | | | |
| Organ support-free days to day 21 | Ordinal | | | |
| Major Pulmonary Embolism | Y/N | | | |
| If Major Pulmonary Embolism, date of event | Date format | | | |

| | | | | |
|---|---|---|---|---|
| Major ischemic cerebrovascular event | Y/N | | | |
| If Major ischemic cerebrovascular, date of event | Date format | | | |
| Major myocardial infarction event | Y/N | | | |
| If Major myocardial infarction, date of event | Date format | | | |
| Major systemic arterial thromboembolism event | Y/N | | | |
| If Major systemic arterial thromboembolism, date of event | Date format | | | |
| Major bleeding event | Y/N | | | |
| If Major Bleed, date of bleeding event | Date format | | | |
| Randomized in REMAP-CAP antiplatelet domain? | Y/N | | | |
| Antiplatelet agent administration in hospital prior to or at the time of randomization | Y/N | | | |
| Classification of anticoagulant dosing administered on each of first two full study days following randomization for each patient randomized to VTP arm (low vs. intermediate) (See statistical analysis Table 3 footnotes for definitions of classification) | NA/low/Inter | | | |
| Invasive mechanical ventilation at time of randomization (yes vs. no) | Y/N | | | |
| Race | Hispanic or latino/Caucasian/black/Asian/First Nations | | | |
| Heart failure | Yes/No | n/a | | |
| Severe cardiovascular disease | Yes/No | | n/a | n/a |
| Diabetes mellitus (Type 1 or Type 2) | Yes/No | | | |
| Chronic kidney disease or end-stage renal disease | Yes/No | | | |
| Chronic respiratory disease | Yes/No | | | |
| Current tobacco use | Yes/No | | | |
| Immunosuppressive treatment | Yes/No | | | |
| Liver disease or cirrhosis | Yes/No | | | |

| | | | | |
|---|---|---|---|---|
| Acute respiratory support at time of randomization | None/supplemental O2/high flow nasal O2/non-invasive ventilation/invasive ventilation/ECMO | | | |
| PaO2/FiO2 in ventilated patients only | Numeric | | n/a | n/a |
| D-dimer | Numeric (fold increase relative to upper limit of normal) | | | |
| Fibrinogen | Numeric | | | |
| INR | Numeric | | | |
| Neutrophils (x$10^9$/L) | Numeric | | | |
| Lymphocytes (x$10^9$/L) | Numeric | | | |
| Platelets (x$10^9$/L) | Numeric | | | |
| Creatinine (mg/dL) | Numeric | | | |
| Troponin (units?) | Numeric | | | |
| Pre-hospital use of anti-platelet agent | Yes/No | n/a | | |
| Dexamethasone exposure at baseline | Yes/No | | | |
| Remdesivir exposure at baseline | Yes/No | | | |
| Anti-platelet agent (aspirin, clopidogrel, ticagrelor, dipyridamole) | Yes/No | | | |

**Table 2.** Site-level variables required for analysis

| **Variable** | **Format** | **Variable Name by Platform** | | |
|---|---|---|---|---|
| | | REMAP-CAP | ACTIV-4a | ATTACC |
| Country | Numeric | | | |
| Standard VTP strategy | Intermediate/low | | | |

### Simulation Report for Multiplatform Randomized-Controlled Trial (mpRCT) for ACTIV4, ATTACC, and REMAP-CAP

CONFIDENTIAL

### CLINICAL TRIAL SIMULATION REPORT VERSION

#### *Version History*

Version 1.0: Draft by Scott Berry, Elizabeth Lorenzi, and Lindsay Berry; September 21, 2020

### INTRODUCTION

This report details the clinical trial simulations that have been conducted for the multiplatform randomized-controlled trial (mpRCT) analysis plan for ACTIV4, ATTACC, and REMAP-CAP. The goal of the simulations is to understand the operating characteristics of the joint analysis of the three platform trials. The analysis model is described briefly in this report, with the "mpRCT Analysis Plan" document providing the full details.

#### *Primary Endpoint*

The primary endpoint for this mpRCT is organ support-free days (OSFD). This endpoint is a composite of in-hospital mortality and the number of days free of organ support in the 21-day period following the time of enrollment. A value of –1 is for any patient that dies within the first 21 days or during their in-hospital stay. For patients that do not die, the outcome is the number of days (rounded to the nearest integer) out of the ICU with 0 as the worst non-death outcome. The best possible outcome of 22 is reserved for patients who survive and never have organ support in ICU. We label the outcome for a patient as Y. This endpoint is an ordinal outcome with 24 possible outcomes (integers –1 to 22) where higher values correspond to better patient outcomes.

#### *Arms*

This analysis includes a control and treatment arm. The control arm represents the respective control from each platform: prophylactic-dose anticoagulation. The treatment arm is a therapeutic dose anticoagulation arm.

#### *Patient Subgroups*

There are four patient subgroups in this analysis based on a cross classification of the baseline disease severity (severe or moderate) and the baseline d-dimer group (low, high, or missing). The four subgroups are: severe, moderate/low, moderate/high, moderate/missing. We label the disease severity as s = 1 (moderate) and s = 2 (severe). We label the d-dimer group as d = 1, 2, 3 for low, high, and missing, respectively. We refer to the subgroups with s:d notation, where the 4 subgroups are 1:1, 1:2, 1:3, and 2:0. Since the severe state is defined as receiving organ support at baseline, the outcome of 22 is not possible in this state.

#### *Trial Status*

In this report, we do not simulate the fixed effects in the model for platform, sex, age, site, or time. Let $Y_{i,s} \in \{-1, 0, 1, \ldots, 21, 22\}$ denote the ordinal outcome for subject $i$ in state $s$. We denote the probability of subject $i$ observing $y$ number of ICU free days or less as $\pi_{iys} = \Pr(Y_{i,s} \leq y)$. Then, for $y = \{-1, 0, 1, \ldots, 21\}$, the primary analysis model is formulated as follows for this report:

$$\log\left(\frac{\pi_{iy}}{1 - \pi_{iy}}\right) = \alpha_{y,s} - (\theta_{a,s:d} + \beta_{\mathrm{d}})$$

The model is a proportional odds model, where the log odds-ratio parameters in the model are structured so that a value > 0 implies treatment benefit, and an odds-ratio > 1 implies treatment benefit.

9. The "arm" to which a patient is randomized is labeled as *a* where *a*=1 is the control and *a*=2 is the treatment arm. The effects of arm vary by the disease state and the d-dimer level. The treatment effects for arm *a* within subtype *s*:*d* are modeled with the $\theta_{a,s:d}$ parameters.
10. The additive effects of d-dimer levels are modeled with the $\beta_d$ for d = 0, 1, 2, 3.
11. The $\alpha_{y,s}$ parameters determine the baseline rates of the ordinal outcome, which are modeled separately by disease state but not d-dimer level.

The treatment effect parameters are relative to a control arm. The treatment effect for the control arm, labeled as arm *a* = 1, will be set to 0 for each of the disease subtypes and d-dimer levels:

$$\theta_{1,s:d} \equiv 0$$

The effects of the treatment arms are modeled hierarchically across disease subtype and d-dimer level. The treatment effects for arm a=2 are modeled as:

$$\theta_{\mathrm{a},2:0} \sim N(\mu_{\mathrm{a},1}, \tau^2_{\mathrm{a},1}),$$

$$\theta_{a,1} \sim N(\mu_{a,1}, \tau^2_{a,1}),$$

$$\theta_{\mathrm{a},s:\mathrm{d}} \sim N(\theta_{a,1}, \tau^2_{\mathrm{a},2}), \qquad s:d = 1:1, 1:2, 1:3,$$

$$\mu_{a,1} \sim N(0, 1);\ \tau^2_{a,1} \sim IG(0.25, 0.1),$$

$$\tau^2_{a,2} \sim IG(0.25, 0.0025)$$

The effects of d-dimer levels are modeled with a N(0,1) prior for d = 2,3, and we define $\beta_d = 0$ for d=0,1 for identifiability.

The ordinal endpoint rates are modeled using a Dirichlet prior where the individual probabilities for the 24 outcomes are based on 1 patient's weight on real-world evidence-based outcomes.

$$logit(\alpha_{y,s}) \sim Dirichlet(P)$$

For the severe state, the model is constrained so that the probability of an OSFD of 22 is zero (*i.e.,* $\alpha_{21,s} = \infty$).

##### ○ *Trial Design and Simulation Assumptions*

The following are the trial design rules and thresholds as simulated:

- If the posterior probability of superiority within a subgroup is greater than 99% then the treatment arm will be labeled as superior to the control arm for OSFDs.
- If the posterior probability of at least an odds-ratio benefit of 1.2 is less than 5% then the treatment arm will be considered futile compared to the control arm.
- Response adaptive randomization (RAR) is used for allocation of each intervention within the ATTACC and REMAP-CAP platforms. The randomization of each patient is proportional to the probability that each regimen is the optimal regimen for a patient within that patient strata. The minimum RAR probability for any intervention is 10% across all regimens.

There are additional assumptions made in the simulations that are not specific to the trial design. The following assumptions are made for the simulations:

- We assume there are no dropouts or missing data besides the subgroup with missing d-dimer level.
- In this mpRCT, interims will occur monthly. For these simulations, interim analyses are timed by the number of patients with complete endpoint data and we assume that interims occur every 200 subjects. For these simulations, the first interim analysis is conducted at 300 then every 200 subjects up to a maximum sample size of 3000. When a conclusion of superiority or futility is made for a subgroup, accrual to that subgroup stops within the simulated trials. Since no conclusions are made for the missing d-dimer group, RAR to that subtype continues until conclusions are made on the remaining subgroups.
- We simulate the ordinal outcome with five levels: death (–1), 0 OSFD, 1-10 OSFD, 11-21 OSFD, and 22 OSFD. This simplification was made for both computational efficiency and statistical model stability, and we believe the impact on operating characteristics is minimal. We retain the proportional odds assumption made in the primary analysis model.

- We assume that 50% of patients are within the severe state and 50% are in the moderate state. Within the moderate state, we assume that 60% of subjects are in the low d-dimer group, 30% are in high d-dimer, and 10% are missing their baseline d-dimer level.
- Table 2.5.1 includes the baseline scenarios for the distribution of the ordinal outcome in the control groups. These control rates correspond to odds-ratios of 0.75 and 0.875 for the moderate/high and moderate/missing subtypes relative to the moderate/low subtype.

Table 2.5.1: Control Rate Scenarios

| Outcome | State/D-Dimer Subgroup | | | |
|---|---|---|---|---|
| | Severe | Moderate/Low | Moderate/High | Moderate/Missing |
| –1 (Death) | 0.295 | 0.090 | 0.117 | 0.102 |
| 0 | 0.224 | 0.071 | 0.087 | 0.078 |
| 1-10 | 0.150 | 0.040 | 0.047 | 0.043 |
| 11-21 | 0.330 | 0.099 | 0.112 | 0.105 |
| 22 | 0.000 | 0.700 | 0.636 | 0.671 |

#### *Simulation Operating Characteristics*

The remaining sections of the report summarize the trial characteristics for different patterns of treatment effects. The operating characteristics present the probability of statistical triggers for superiority and futility.

Figures entitled “Cumulative Power”, such as Figure 3.1.1, report the cumulative probability that superiority is demonstrated in one or more subgroups. At each interim sample size, we compute the proportion of simulations that have demonstrated superiority in at least one subgroup. This provides an estimate of power cumulatively up to a maximum of 3,000 subjects.

Figures entitled “Cumulative Power by Subgroup”, such as Figure 3.1.2, report the cumulative probability of each subgroup meeting the statistical trigger for superiority throughout the trial. In the null scenario (labeled with and odds ratio of 1), this quantity represents the type I error rate of declaring superiority for a subgroup where the intervention is ineffective.

Figures entitled “Cumulative Futility”, such as Figure 3.1.3, report the cumulative probability of meeting the statistical trigger for futility for each subgroup. For each interim sample size, we average across all simulations to find the probability of futility and report the cumulative probability of futility up to a total sample size of 3,000.

Figures entitled “Cumulative Power by Scenario”, such as Figure 3.5.1, report the cumulative probability of a subgroup meeting the statistical trigger for superiority throughout the trial for various treatment effect patterns. For these sections, we consider the treatment effect patterns shown in Table 2.6.1 where X = 1 (null), 1.3, 1.5, 1.75, and 2.

Table 2.6.1: Treatment Effect Scenarios

| State/D-Dimer Subgroup | Treatment Effect Scenario | | |
|---|---|---|---|
| | Works in all subgroups | Works in Severe Only | Works in Severe, Mod/High |
| Severe | X | X | X |
| Moderate/Low | X | 1 | 1 |
| Moderate/High | X | 1 | X |
| Moderate/Missing | X | 1 | (X+1)/2 |

### OPERATING CHARACTERISTICS

#### *Same Treatment Effect Across Subgroups*

This section shows the operating characteristics when the active treatment has the same effect across subgroups. For this pattern of treatment effects, we consider odds-ratio effects of: 1 (no effect), 1.3, 1.5, 1.75, and 2.

Figure 3.1.1: Same Treatment Effect Across Subgroups, Cumulative Power

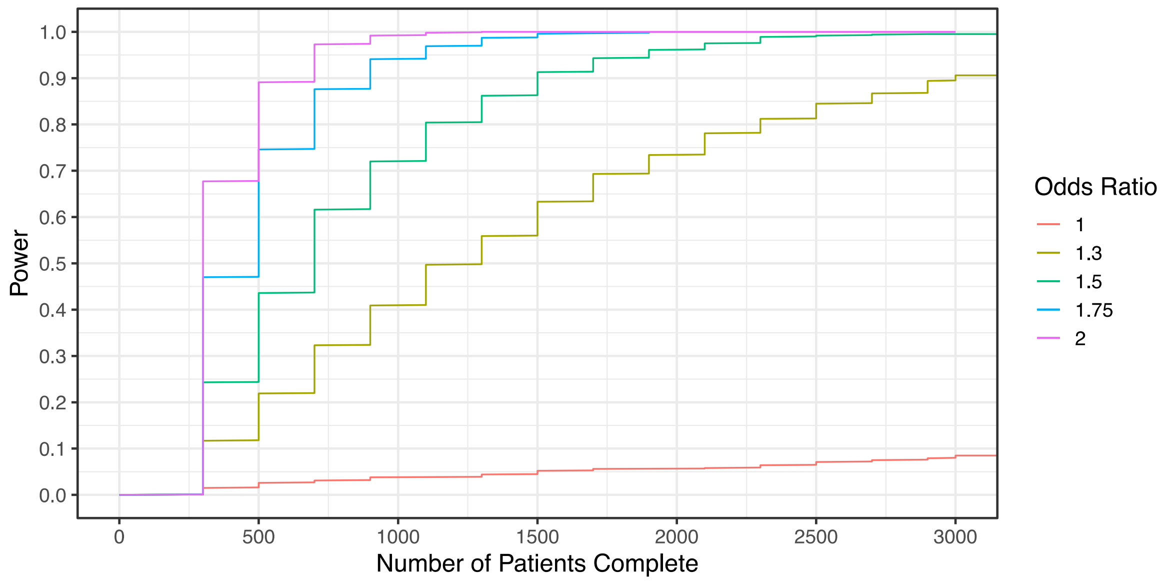

Figure 3.1.2: Same Treatment Effect Across Subgroups, Cumulative Power by Subgroup

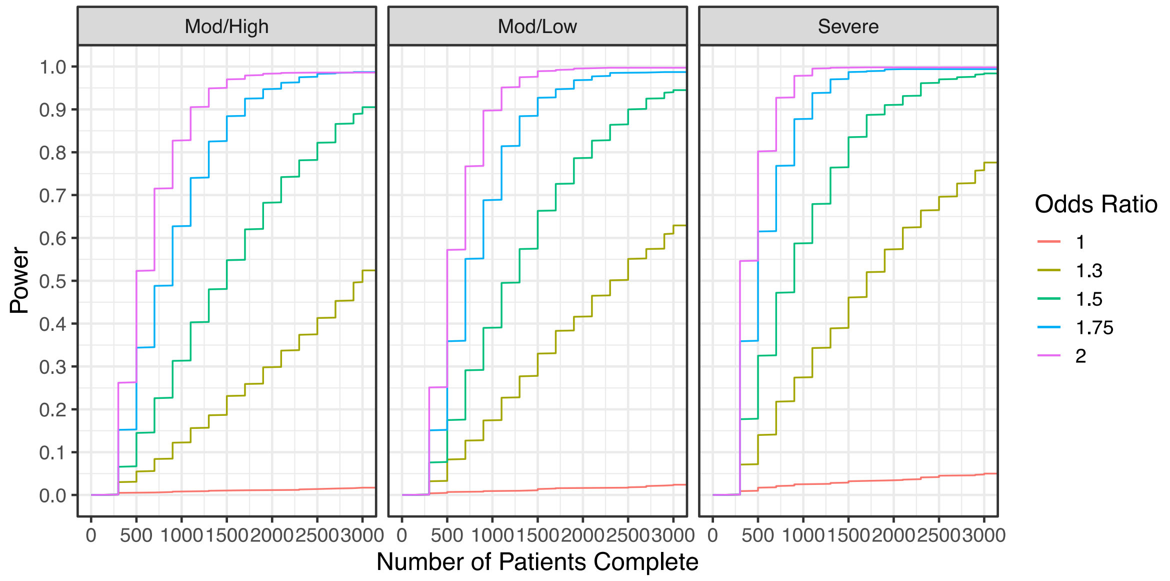

Figure 3.1.3: Same Treatment Effect Across Subgroups, Cumulative Futility

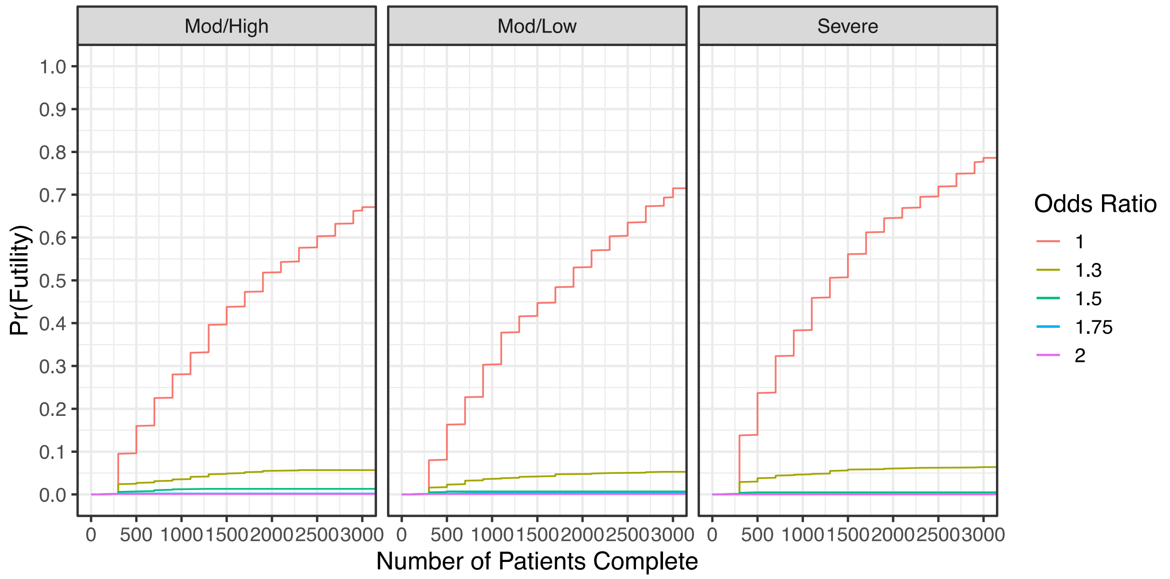

#### *Effective in Severe Subgroup Only*

This section shows the operating characteristics when the active treatment is only effective in the "Severe" subgroup. For this pattern of treatment effects, we consider odds-ratio effects of 1 (no effect), 1.3, 1.5, 1.75, and 2 in the Severe subgroup. In this scenario, the moderate subgroups have odds-ratio effects of 1.

Figure 3.2.4: Effective in Severe Subgroup, Cumulative Power

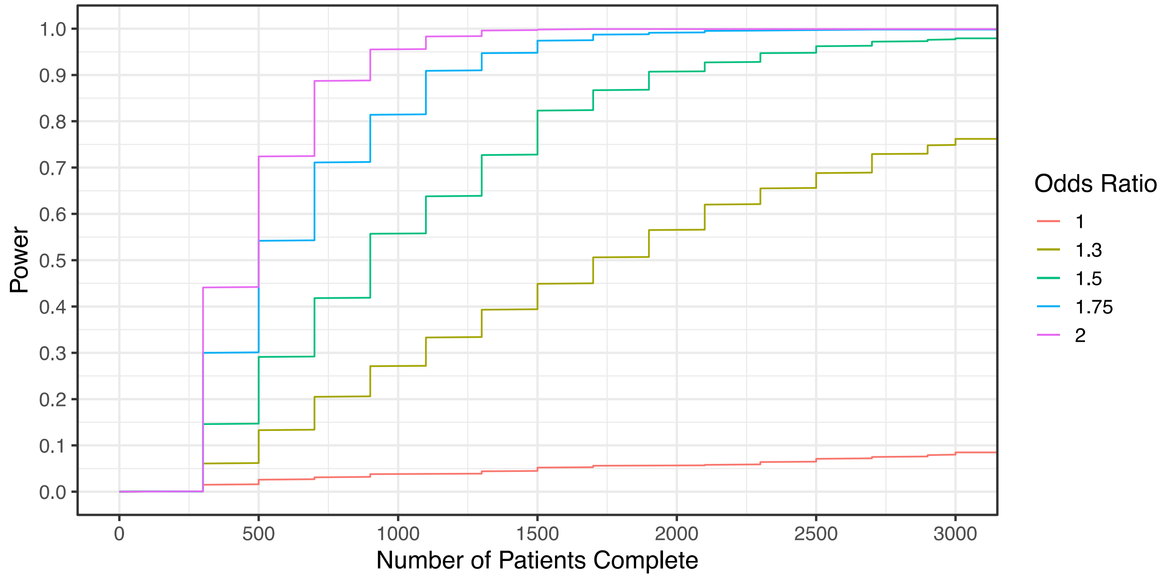

Figure 3.2.5: Effective in Severe Subgroup, Cumulative Power by Subgroup

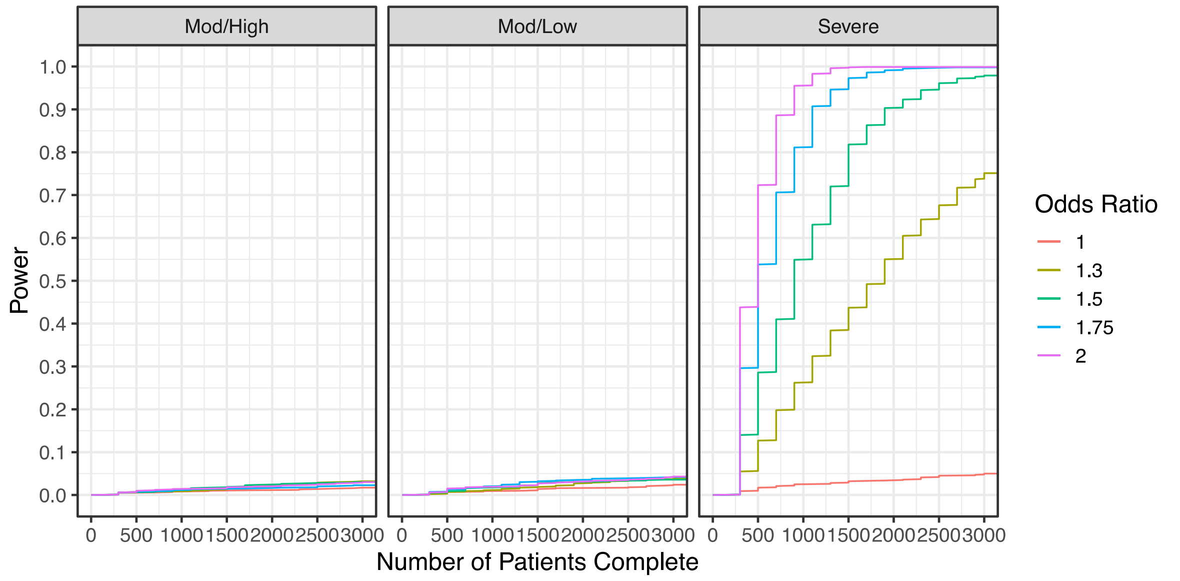

Figure 3.2.6: Effective in Severe Subgroup, Cumulative Futility

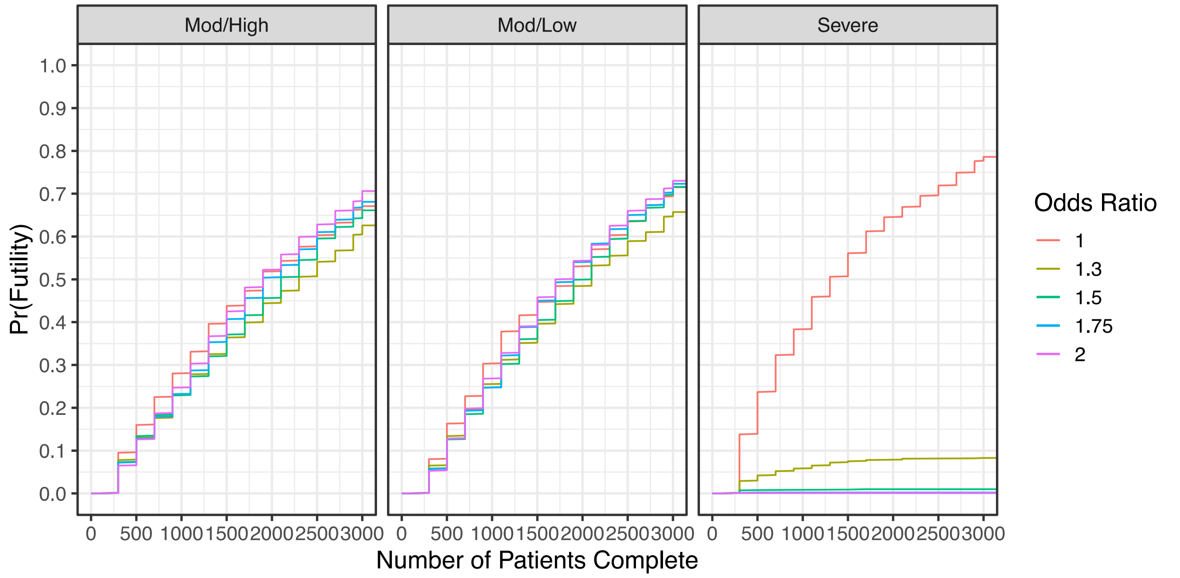

#### *Effective in Severe and Moderate/High Subgroups*

This section shows the operating characteristics when the active treatment has the same effect in the Severe and Moderate/High subgroups. For this pattern of treatment effects, we consider odds-ratio effects of 1 (no effect), 1.3, 1.5, 1.75, and 2. In this scenario, the moderate/low subgroup has an odd-ratio effect of 1, and the moderate/missing subgroup has an odd-ratio effect that is an average of moderate/high and moderate/low odds-ratios (*i.e.,* if mod/high OR = 2 and mod/low OR =1, then mod/missing OR = 1.5).

Figure 3.3.7: Effective in Severe and Moderate/High Subgroups, Cumulative Power

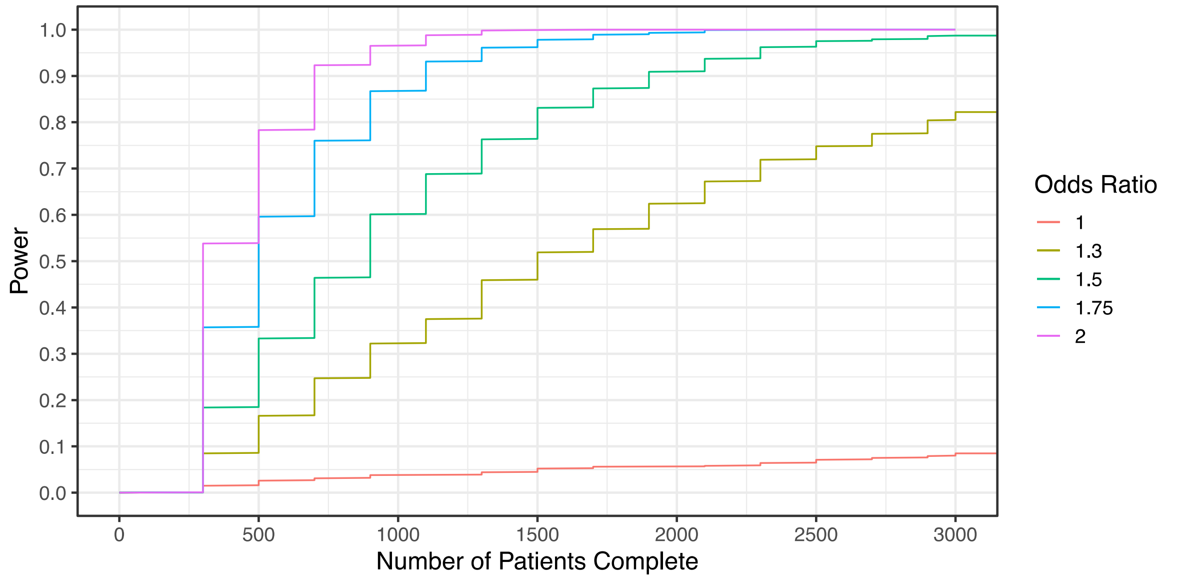

Figure 3.3.8: Effective in Severe and Moderate/High Subgroups, Cumulative Power by Subgroup

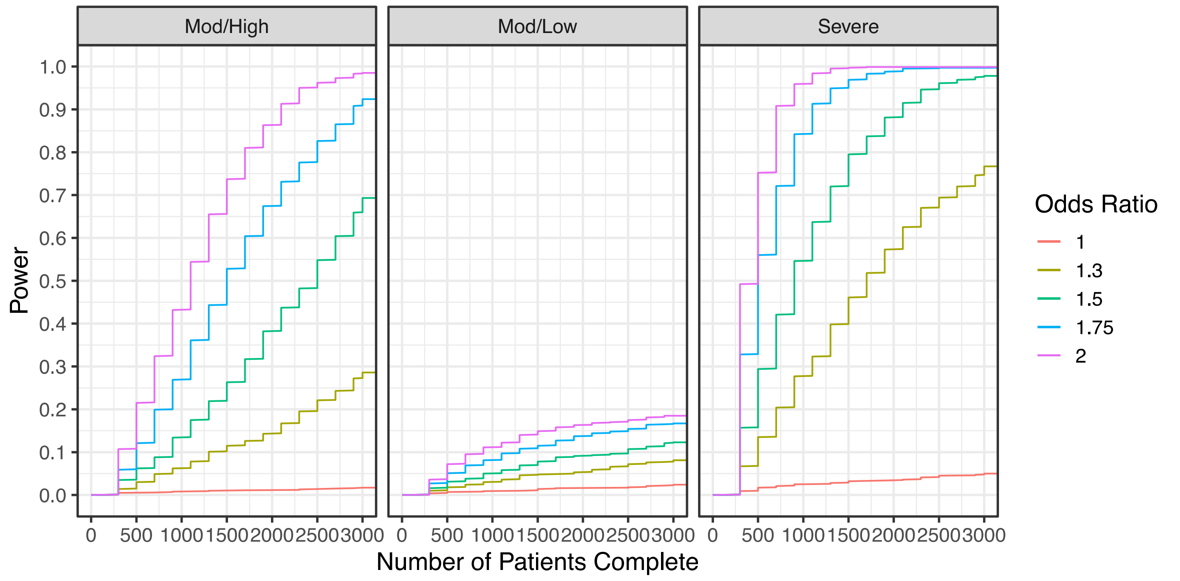

Figure 3.3.9: Effective in Severe and Moderate/High Subgroups, Cumulative Futility

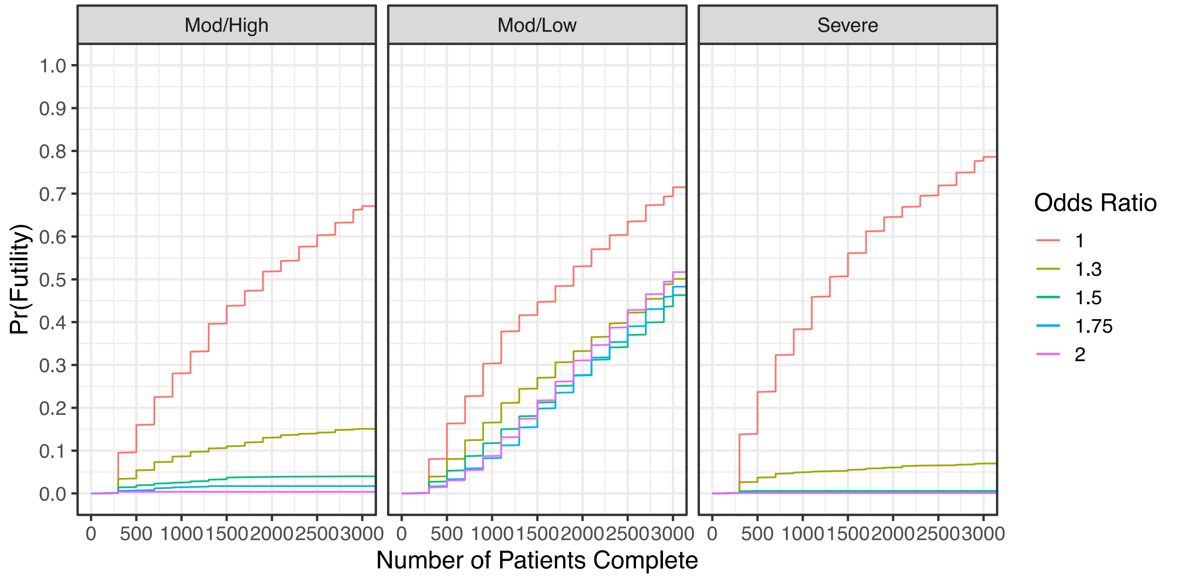

#### *Escalating Effects Across Subgroups*

This section shows the operating characteristics when the active treatment has escalating effects across the patient subgroups. We present two scenarios for this pattern of treatment effects, shown in the table below. These scenarios are based on an assumption that the "missing d-dimer" subgroup is 50% low d-dimer and 50% high d-dimer patients, and that the treatment effect is an average of the two subgroup treatment effects.

| Treatment Effect Scenario | State/D-Dimer Subgroup | | | |
|---|---|---|---|---|
| | Severe | Moderate/Low | Moderate/High | Moderate/Missing |
| 1 | 2 | 1.3 | 1.75 | 1.5 |
| 2 | 2 | 1.5 | 2 | 1.75 |

Figure 3.4.10: Escalating Effects Across Subgroups, Cumulative Power

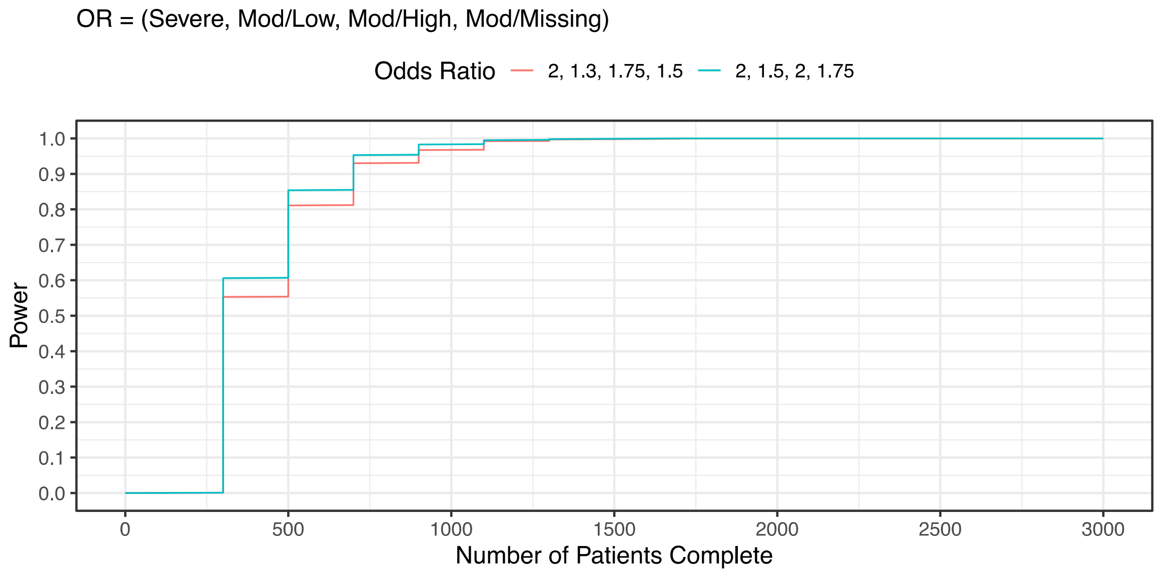

Figure 3.4.11: Escalating Effects Across Subgroups, Cumulative Power by Subgroup

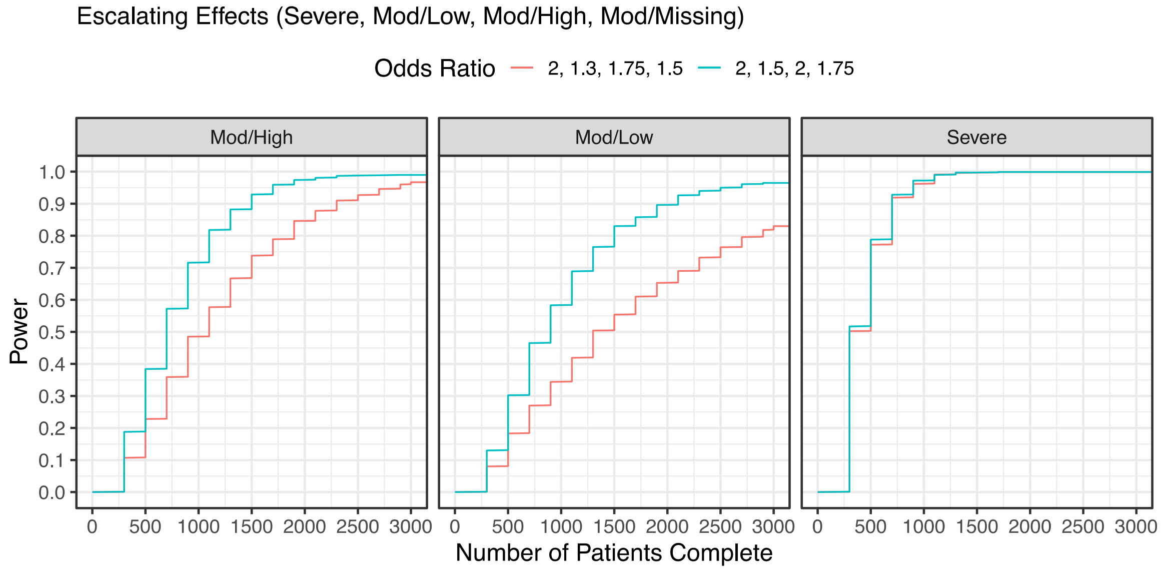

Figure 3.4.12: Escalating Effects Across Subgroups, Cumulative Futility

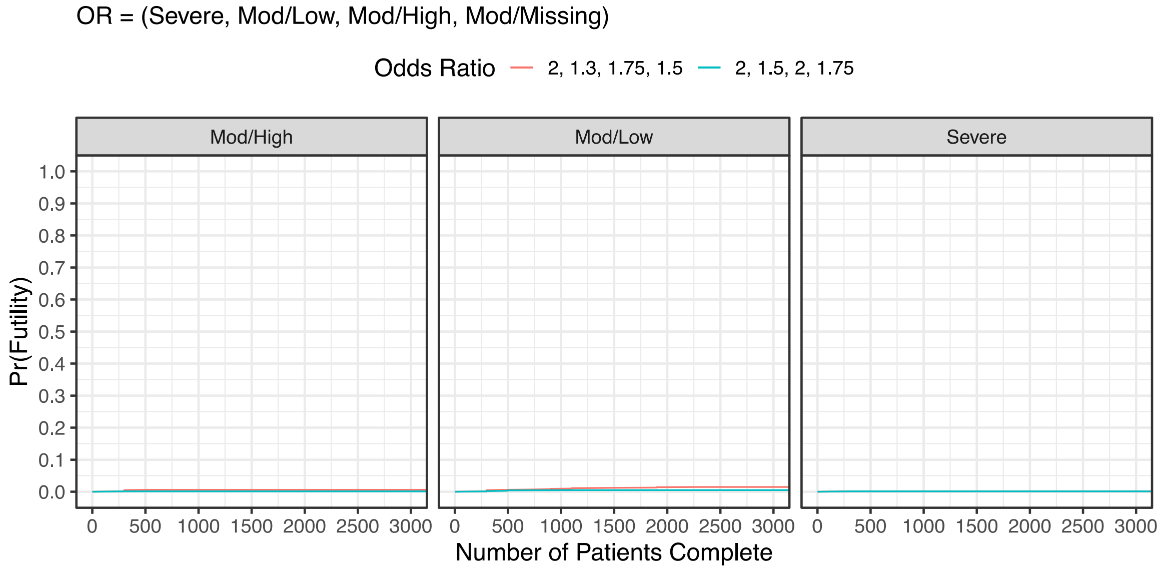

- ***Severe State Power by Scenario***

This section shows the cumulative power in the severe state across the pattern of treatment effect scenarios shown in Table 2.6.1.

Figure 3.5.1: Power in Severe State, Cumulative Power by Scenario

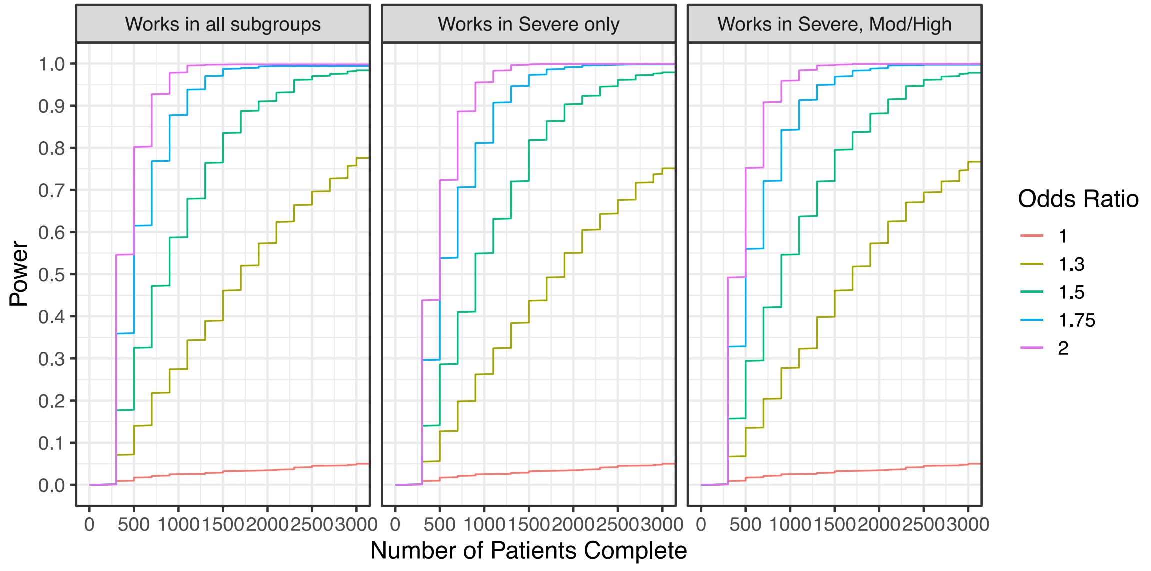

#### a. *Moderate/Low Subgroup Power by Scenario*

This section shows the cumulative power in the moderate/low subgroup across the pattern of treatment effect scenarios shown in Table 2.6.1.

Figure 3.6.1: Power in Moderate/Low Subgroup, Cumulative Power by Scenario

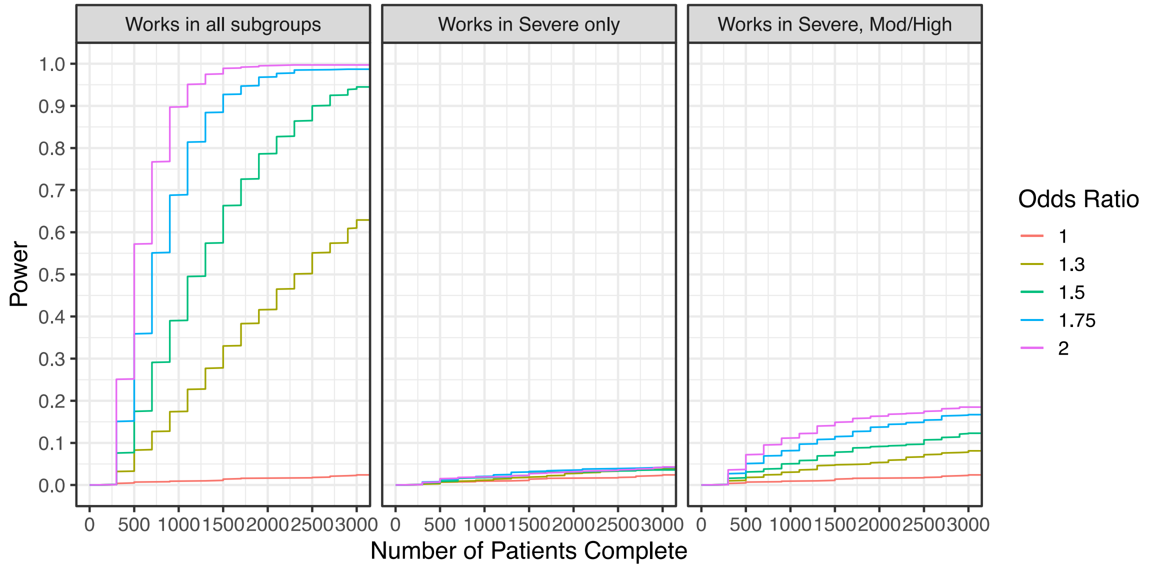

##### *b. Moderate/High Subgroup Power by Scenario*

This section shows the cumulative power in the moderate/high subgroup across the pattern of treatment effect scenarios shown in Table 2.6.1.

Figure 3.7.1: Power in Moderate/High Subgroup, Cumulative Power by Scenario

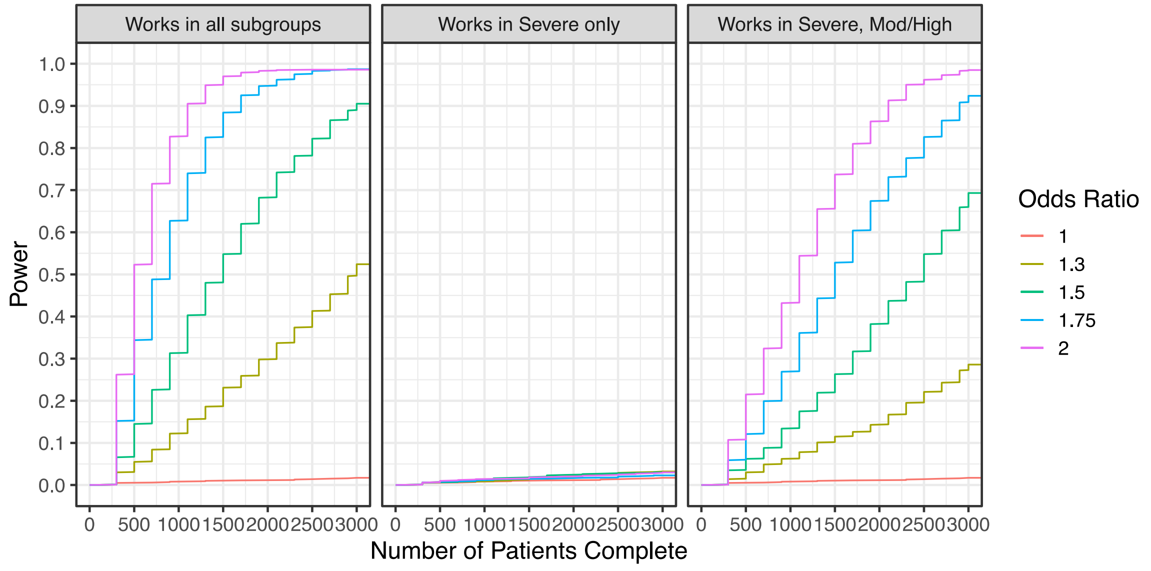

### Current State of the Statistical Model for the AARC Multi-Platform Randomized Clinical Trial Analysis Plan

Version 1.1 dated 19 November 2020

**Version History:**

**Version 1.1** – Updated by Lindsay Berry; November 19, 2020

Version 1.1 is updated to replace Version 1.0 as the Current State document for the first analysis of the AARC mpRCT. The following edits were made to Version 1.0:

- The platform effects have been removed from the statistical model due to the possibility of non-identifiability of platform effects and site effects. On page 9, the table of parameters for platform effect has been deleted. On page 11 on Model Specification, the gamma parameters have been removed from the list of variables and the moderate and severe state likelihoods.

**Version 1.0** - Written by Elizabeth Lorenzi, Lindsay Berry, and Scott Berry; September 26, 2020

Version 1.0 of this document was intended to be used for the first analysis of the anticoagulation mpRCT.

#### Terminology

**Severe State** is a patient that is receiving organ failure support in an ICU at baseline

**Moderate State** is defined by not being admitted to an ICU, or admitted to an ICU but not receiving organ failure support at baseline.

**d-dimer** is a baseline measure that will be classified into three groups: low, high, and missing. The definition of **high** is a value greater than or equal to 2 times the upper limit of normal.

**Subtype** is the smallest unit-of-analysis subgroup for this analysis

#### Modeling Conventions

**Primary Endpoint**

| **Composite of:** | **Definition** | **Notes** |
|---|---|---|
| 22 | No organ support in an ICU | Not a possible outcome in the Severe state |
| 0, 1, …, 21 | Number of study days for which the subject is alive and not receiving organ support in an ICU up until the end of study day 21 | Rounded to the nearest integer |
| −1 | Death before discharge from an acute hospital | |

Derivation Details

- Note that the SAC will receive the derived endpoint from data providers, and the derivation details below are provided solely for clarification purposes.
- Deriving this endpoint from hours
    - 0 organ support hours, OSFD = 22
    - Less than 12 hours on organ support, OSFD = 21
    - Off organ support for more than 12 hours, OSFD = 1
    - Less than 12 hours free of organ support, OSFD = 0
- Time of first organ support to time of last organ support within an ICU admission (intervening time free of organ support does not count)

#### Anticoagulation

- **2 Interventions**
- **Subgroups:** The anticoagulation domain includes pre-specified subtypes based on baseline d-dimer status. The subtypes are:
  - moderate state, low d-dimer;
  - moderate state, high d-dimer;
  - moderate state, missing d-dimer;
  - severe state
- **Statistical Triggers:** The domain allows superiority of the therapeutic dose anticoagulation over venous thromboprophylaxis and futility of therapeutic dose anticoagulation compared to the venous thromboprophylaxis. These conclusions apply individually to three of the four subtypes:
  - moderate state, low d-dimer;
  - moderate state, high d-dimer;
  - severe state

| Intervention Name | Input Data File Code | Notes |
|---|---|---|
| Venous thromboprophylaxis | 1 | H1 |
| Therapeutic anticoagulation | 2 | H2 |

**Response Adaptive Randomization**

The response adaptive randomization for the ATTACC trial should be set for the moderate/low d-dimer, and the moderate/high d-dimer groups. ATTACC does not "enroll" in a severe group, even if some are classified as 'severe' in the model. The Moderate group with missing d-dimer should remain equal randomization.

Adaptive randomization is based on the posterior probability that the odds-ratio for H2 is greater than 1 within each subtype. This probability should be truncated between 0.10 and 0.90.

### MODEL TERMS SPECIFICATION

| Model States and Notation | |
|---|---|
| State | Note |
| Moderate | Parameter s = 1 |
| Severe | Parameter s = 2 |

| Control rate probabilities by state | | | |
|---|---|---|---|
| Term | State | Prior | Note |
| $\alpha_{-1,s=1} \cdots \alpha_{22,s=1}$ | Moderate | $\alpha_{k,s=1} = logit\left(\sum_{j=-1}^{k} p_{j,s=1}\right), k = -1, \ldots, 22$ where $(p_{-1,s=1}, \ldots, p_{22,s=1}) \sim Dirichlet(\hat{p}_{s=1})$ and $\hat{p}_{s=1}$ is a vector of the prior probabilities reported below. | Intercepts in ordinal model for moderate state |
| $\alpha_{-1,s=2} \cdots \alpha_{21,s=2}$ | Severe | $\alpha_{k,s=2} = logit\left(\sum_{j=-1}^{k} p_{j,s=2}\right), k = -1, \ldots, 21$ where $(p_{-1,s=2}, \ldots, p_{21,s=2}) \sim Dirichlet(\hat{p}_{s=2})$ and $\hat{p}_{s=2}$ is a vector of the prior probabilities reported below. The severe model is constrained such that $p_{22,s=2} = 0$ and $\alpha_{21,s=2} = \infty$. | Intercepts in ordinal model for severe state |

Notes:

- If there is an outcome in the ordinal scale that did not occur in the data for a state, then that outcome will be combined to a single outcome with a neighboring outcome for that state (the worse outcome).
- For example, if the outcome 11 never occurred in the moderate state, then a combined outcome of 10 & 11 will be modeled for the moderate state in this analysis.
- For the severe state, $\hat{p}_{s=2}$has 29.5% probability on –1, 22.5% on 0, 1.5% on each outcome from 1-10, and 3.0% on each outcome from 11-21. The sum of the Dirichlet prior concentration parameter is 1.
- For the moderate state, $\hat{p}_{s=1}$has 9.0% probability on –1, 7.1% on 0, 0.4% on each outcome from 1-10, 0.9% on each outcome from 11-21, and 70% probability on 22. The sum of the Dirichlet prior concentration parameter is 1.

| Main Effects of Era by State | | | |
|---|---|---|---|
| Term | Description | Prior | Note |

| | | | |
|---|---|---|---|
| $\lambda_{T=1,s}$ | Most recent 4 weeks** | 0 | Set to 0 |
| $\lambda_{T=2,s} - \lambda_{T=1,s}$ | Difference for 2-week period after 1 months from most recent | N(0, $0.15^2$) | |
| $\lambda_{T,s} - 2\lambda_{T-1,s} + \lambda_{T-2,s}$ | Difference for each additional 2-week bucket | $N(0, \tau_\lambda^2)$<br>$\tau_\lambda^2 \sim IG(0.25, 0.00562)$ | |

| **Definition of Era Terms by State** | |
|---|---|
| Term | Description |
| $\lambda_{T=1,s}$ | Most recent 4 weeks** in state *s* |
| $\lambda_{T=2,s}$ | 4-6 week from time 0 in state *s* |
| $\lambda_{T=3,s}$ | 6-8 week from time 0 in state *s* |
| … | Continuation of buckets as time progresses |

Notes:

- **Time 0: Time of randomization of the most recent subject with a complete outcome in the analysis dataset. Time 0 is the same for the severe and moderate state.
- The time era buckets are the same for each state, but we estimate time effects by state.
- Time buckets with <5 subjects in a state randomized within the bucket may be combined with a neighboring bucket for that state.

| **Main Effects of Site by state. Site modeled within Parent Country $c_R$ where $c_R = 1, \ldots, C$** | | |
|---|---|---|
| Term | Description | Prior |
| $\nu_{R=1,s}$ | Site 1 | $[\nu_{R,s}] \sim N(\mu_{C_R,s}, \tau^2_{C_R,s})$ $R = 1, \ldots, N_R$,<br>$[\mu_{C_R,s}] \sim N(0,1)$; $[\tau^2_{C_R,s}] \sim IG(0.25,0.1)$,<br>where c=2,…,C. s=1,2<br>$[\mu_{1,s}] = 0$ |
| $\nu_{R=2,s}$ | Site 2 | |
| $\nu_{R=3,s}$ … | Site 3 | |
| $\nu_{R=N_R,s}$ | Site $N_R$ | |
| Note: c=1 will represent the US. | | |

Notes:

- All sites within a country that have <5 subjects randomized in a state in the analysis population will be combined into a single site within that country.

| **Main Effects of Age Category** | | | |
|---|---|---|---|
| Term | Description | Prior | Note |
| $\beta_{age=1,s}$ | 39 years old or less | N(1.5300, 1) | |

| | | | |
|---|---|---|---|
| $\beta_{age=2,s}$ | 40-49 | N(1.2959, 1) | |
| $\beta_{age=3,s}$ | 50-59 | N(0.6054, 1) | |
| $\beta_{age=4,s}$ | 60-69 | 0 | Set to 0 |
| $\beta_{age=5,s}$ | 70-79 | N(−0.4837, 1) | |
| $\beta_{age=6,s}$ | 80+ | N(−0.3900, 1) | |

| **Main Effects of Sex at Birth Category by State** | | | |
|---|---|---|---|
| Term | Description | Prior | Note |
| $\beta_{Sex=1,s}$ | Male | Set to 0 | Referent |
| $\beta_{Sex=2,s}$ | Female | N(0, 1) | |

| **Main Effects of D-Dimer Subgroup in Moderate State** | | | |
|---|---|---|---|
| Term | Description | Prior | Note |
| $\beta_{D-Dimer=0}$ | Missing baseline d-dimer | N(0, 1) | |
| $\beta_{D-Dimer=1}$ | Low baseline d-dimer | Set to 0 | Referent |
| $\beta_{D-Dimer=2}$ | High baseline d-dimer | N(0, 1) | |

**Model Treatment Parameters:**

| Model Parameters | State (s) | Prior Distribution | Description of Parameter |
|---|---|---|---|
| $\theta_{1,s}$ | 1, 2 | 0 | Effect of No Therapeutic Anticoagulation |
| $\theta_{2,2}$ | 2 | $N(\mu_{\theta,1}, \tau^2_{\theta,1})$ | Effect of Therapeutic Anticoagulation in severe state |
| $\theta_{2,1}$ | 1 | $N(\mu_{\theta,1}, \tau^2_{\theta,1})$ | Effect of Therapeutic Anticoagulation in moderate state |
| $\theta_{2,1:0}$ | 1 | $N(\theta_{2,1}, \tau^2_{\theta,2})$ | Effect of Therapeutic Anticoagulation in moderate state, missing baseline D-Dimer |
| $\theta_{2,1:1}$ | 1 | $N(\theta_{2,1}, \tau^2_{\theta,2})$ | Effect of Therapeutic Anticoagulation in moderate state, low baseline D-Dimer |
| $\theta_{2,1:2}$ | 1 | $N(\theta_{2,1}, \tau^2_{\theta,2})$ | Effect of Therapeutic Anticoagulation in moderate state, high baseline D-Dimer |
| $\mu_{\theta,1}$ | 1,2 | N(0, 1) | Shared mean for nested parameters |
| $\tau^2_{\theta,1}$ | 1,2 | IG(0.25, 0.1) | Shared variance for nested state parameters |
| $\tau^2_{\theta,2}$ | 1 | IG(0.25, 0.0025) | Shared variance for nested moderate parameters |

### MODEL SPECIFICATION

#### Modeling Strategy for Disease States

- There are separate, independent covariate effects for the moderate and severe states. This includes the effects of site, age, sex, and time.
- The $\alpha$ parameters control the distribution of the ordinal outcome. There will two separate, independent vectors of $\alpha$ parameters: one for the moderate state and one for the severe state. In the severe state, it is not possible for a subject to have an outcome of 22 (no organ support). To adjust for this, we set the probability of a 22 equal to 0, effectively forcing $\alpha_{21,s=2} = \infty$.
- Intervention effects will vary by subtype and there will be borrowing between the subtypes.

Let the following variables be assigned for subject $i$:

- $s_i$ = disease state
- $r_i$ = site
- $age_i$ = age group
- $sex_i$ = biological sex at birth
- $T_i$ = randomization era
- $D_i$= d-dimer subgroup. Possible values are 0 (missing), 1 (low), and 2 (high) for subjects in the moderate state. Severe patients have no baseline d-dimer subgroup.
- $a_i$ = arm (intervention)

The ordinal outcomes are labeled $Y_{i,s} \in \{-1,0,1,\ldots,21,22\}$ for the OSFD outcome of patient $i$ in state $s$. We model the probability of an outcome *y* or worse for patient $i$ in state $s$ as $\pi_{iys} = Pr(Y_i \leq y | s_i = s)$.

**Severe State Likelihood:**

For subjects that are in the Severe state, we model the ordinal outcome $Y_i$ as:

$$log\left(\frac{\pi_{iy2}}{1-\pi_{iy2}}\right) = \alpha_{y,s=2} - (v_{r_i,s=2} + \beta_{age_i,s=2} + \beta_{sex_i,s=2} + \lambda_{T_i,s=2} + \theta_{a_i,2})$$

**Moderate State Likelihood:**

For subjects that are in the Moderate state, we model the ordinal outcome $Y_{i,s=1}$as follows:

$$log\left(\frac{\pi_{iy1}}{1-\pi_{iy1}}\right) = \alpha_{y,s=1} - (v_{r_i,s=1} + \beta_{D_i} + \beta_{age_i,s=1} + \beta_{sex_i,s=1} + \lambda_{T_i,s=1} + \theta_{a_i,1:D_i})$$

- The moderate state adjusts for the baseline d-dimer subgroup through the parameter $\beta_D$.

**Transition from Moderate to Severe State:**

This mpRCT analysis does not include any modeling of patients that transition from the moderate to severe state. A patient who transitions would only be included in the disease state in which they were randomized to an anticoagulation intervention.

#### Statistical Triggers

- For each intervention, we list all possible statistical triggers
- Triggers are made by subtype

| Intervention | Subtype | Conclusion | Comparator | Decision quantity | | Threshold |
|---|---|---|---|---|---|---|
| Therapeutic anticoagulation | S=2<br>Severe | Superiority | Venous thromboprophylaxis | Pr($exp(\theta_{2,2})$ >1) | > | 0.99 |
| Therapeutic anticoagulation | S=2<br>Severe | Futility | Venous thromboprophylaxis | Pr($exp(\theta_{2,2})$ >1.2) | < | 0.05 |
| Therapeutic anticoagulation | S=1,D=1<br>Moderate, Low D-Dimer | Superiority | Venous thromboprophylaxis | Pr($exp(\theta_{2,1:1})$ >1) | > | 0.99 |
| Therapeutic anticoagulation | S=1,D=1<br>Moderate, Low D-Dimer | Futility | Venous thromboprophylaxis | Pr($exp(\theta_{2,1:1})$ >1.2) | < | 0.05 |
| Therapeutic anticoagulation | S=1,D=2<br>Moderate, High D-Dimer | Superiority | Venous thromboprophylaxis | Pr($exp(\theta_{2,1:2})$ >1) | > | 0.99 |
| Therapeutic anticoagulation | S=1,D=2<br>Moderate, High D-Dimer | Futility | Venous thromboprophylaxis | Pr($exp(\theta_{2,1:2})$ >1.2) | < | 0.05 |

### Multiple Platform RCT DSMB Interaction Plan

#### ACTIV-4a, ATTACC, and REMAP-CAP Multiplatform Randomized Controlled Trial Data Flow Plan, Interim Analysis Plan and DSMB Coordination Plan

##### Background

ACTIV-4A, ATTACC, REMAP-CAP have jointly developed a single multiplatform randomized controlled trial (mpRCT) evaluating the efficacy of antithrombotic strategies in patients hospitalized with COVID-19. PROTECT was a vanguard/pilot study which was approved by the NHLBI COVID Protocol Review Committee and approved by a separate DSMB (that currently oversee NCATS funded CONTAIN trial) which is incorporated into ACTIV4A). PROTECT data is being combined into ACTIV 4A and that DSMB will no longer have oversight. Each of these separate trials (or platforms) is randomizing a control arm of prophylactic-dose anticoagulation against a therapeutic-dose anticoagulation through a common mpRCT. This document describes the interim analysis reporting plans and procedure for interactions between DSMBs providing governance for three of the trials for the question of efficacy related to therapeutic- dose anticoagulation.

The three platforms use closely harmonized protocols, overlapping data collection, leadership, and oversight approaches. Trial data coordinating centers are the University of Pittsburgh (in collaboration with SOCAR) for ACTIV-4a, SOCAR (ATTACC), and SPIRAL/UPMC (REMAP-CAP). PROTECT data are combined into ACTIV-4a and the role of that DSMB will cease. Statisticians from Berry Consultants serve as trial design statisticians for the three platforms: ACTIV-4a, ATTACC, and REMAP-CAP. This group remains blinded to trial data and continue to work on the evolutions and design of these adaptive platforms. There is an independent unblinded Statistical Analysis Committee (SAC) for the mpRCT. The SAC is composed of statisticians from Berry Consultants, but is an entirely separate group with firewalls between the group and the design teams. With agreed upon stopping rules for efficacy and futility for the primary endpoint, the mpRCT operates under a multiplatform analysis plan with monthly interims. The mpRCT investigators will collaborate on making major decisions.

##### Study Synopses

The multiplatform randomized controlled trial (mpRCT) is a collaboration of three platforms. The ACTIV-4a, ATTACC, and REMAP-CAP platform trials have very closely harmonized protocols and shared endpoints (see cross-trial comparison table in subsection G). Each of these platforms will contribute data from patients already enrolled prior to the creation of the mpRCT as well as continue to contribute patients enrolled in these platforms internationally. The ACTIV-4a Trial is enrolling at sites established through a “network of networks” that includes numerous NIH funded networks (eg. PETAL, ISCHEMIA, STROKENet). In addition to these sites, there are 4 additional sources of patients for ACTIV-4a. First, PROTECT was an NHLBI protocol review committee approved trial that serves as a vanguard trial for ACTIV-4a. PROTECT has closed and the patients are now considered part of ACTIV-4a as described below. Next, REMAP and ATTACC, although continuing as platform trials internationally, have moved their US sites into ACTIV-4a by adopting/synergizing protocols and eCRFs. These US sites are thus referred to at the REMAP-CAP *Network* and ATTACC *Network* to distinguish them from the international platform trials of the same name.  Fourth, the RAPID Trial is a separate trial of anticoagulation in COVID-19. Although the RAPID Trial has not combined efforts into the mpRCT, they have converted US sites into the RAPID *Network* and are enrolling patients into ACTIV-4a using the ACTIV4a protocol and eCRF (adopting the identical model of the REMAP-CAP and ATTACC *Networks*).

A synosis of each study is included in subsection F.

A chart showing the similarities of eligibility criteria and outcomes is presented in subsection G.

#### DSMB Interaction Protocol

The following protocol offers general principles and operational guidelines for the DSMB of each platform within the mpRCT.

##### Report submission

The interim efficacy and safety analyses will be reported to the DSMBs for each of the trials (Figure 1). PROTECT is closing, its data is being folded into ACTIV-4a and will no longer have its own DSMB. Each DSMB (ACTIV-4A, REMAP-CAP, ATTACC) will receive a mpRCT efficacy analysis report from the SAC and a safety analysis report for the individual platform from the platform's data coordinating center. Since safety data are not aggregated across trials, safety analysis reports for each platform will be shared with the DSMBs of other platforms. Each DSMB will receive:

- Combined efficacy interim analysis
- ACTIV-4a safety report (including PROTECT)
- REMAP-CAP safety report
- ATTACC safety report

Efficacy and safety reports will be produced and delivered monthly, using data cutoff of the first working day of the month. Reports will be completed and delivered to the DSMB one to two weeks after the data transfer. It is expected that each DSMB will schedule monthly review meetings of the interim reports once reports are complete.

##### DSMB Report review for each platform

Each DSMB will review all delivered efficacy and safety interim analysis reports, and will reach a consensus on safety topics, if any, and the efficacy analysis (if an efficacy or futility trigger has been met), and risk/benefit of efficacy/safety.

#### Report communication between DSMB

The successful integration of the data and analyses of the three platforms will require communication between the DSMBs of the three platforms.

To facilitate this process the project teams from each study recommend that the NIH invite the Chairs of the ATTACC and REMAP DSMBs (or a designee) to participate as non-voting members of the ACTIV-4a DSMB, and that the chair (or a designee) of the ACTIV-4a DSMB be invited to participate as a non-voting member of the ATTACC DSMB and the anticoagulation component of the REMAP-CAP DSMB. Additional communication between individual DSMBs will occur based on expressed need and will be at the discretion of each DSMB. This will allow each of the DSMBs to communicate on the efficacy triggers that have been defined in the mpRCT analysis plan as well as any safety concerns.

In the event that one DSMB decides to terminate the study or an arm of the study, it will be up to the other DSMBs to decide what to recommend to their sponsor.

In the event that either ATTACC or REMAP is terminated, a member of the ACTIV-4a team (Dr. Matthew Neal) may be unmasked to participate in the discussion of the impact on ACTIV-4a. In the event that Dr. Neal is unmasked, he participation in further decision making will be limited in a fashion similar to that of the unmasked biostatistician, Dr. Maria Brooks.

#### DSMB Recommendations for each Trial

Each trial DSMB maintains its responsibilities within its trial. This includes any one trial DSMB recommending halting enrollment to any arms for their platform for efficacy or safety reasons. Efficacy will be determined according to the mpRCT joint statistical analysis plan and not individually within each platform.

It is generally agreed that individual trials will not make public disclosures of efficacy without the agreement of all three DSMBs and the respective study sponsors (NHLBI for ACTIV4A).

If a stopping recommendation is reached, each DSMB is responsible for the communication to each of the platforms' leadership/steering committees which should be done with similar timing.

Figure 1: Communication Plan Across Trials for Interim Analyses

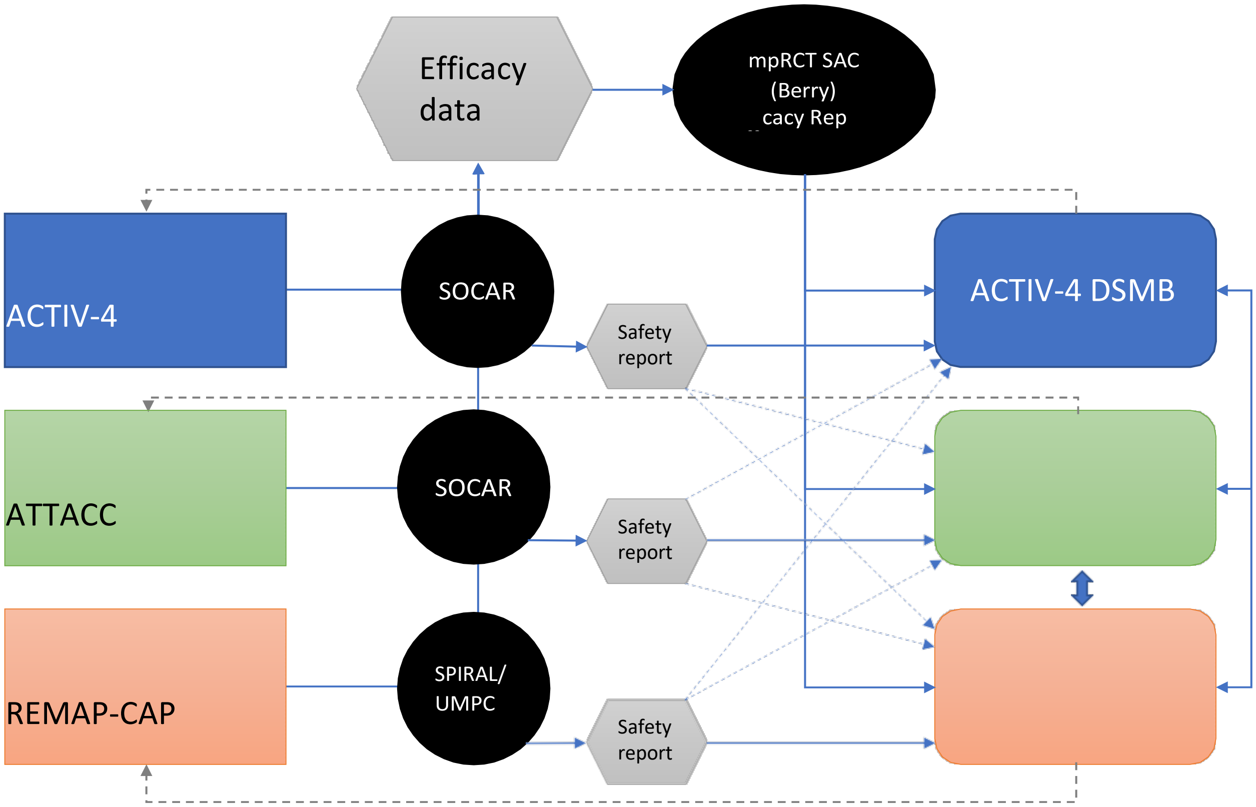

We, the multi-platform chairs are in agreement with the publication plan outlined in this document, Version 1.0, 18 December 2020.

Ryan Zarychanski Date: 18 December 2020
Platform Chair, ATTACC

Judith Hochman Date: 19 January 2021
Platform Chair, ACTIV4a

Steve Webb Date: 20 January 2021
Platform Chair, REMAP-CAP

#### SUBSECTIONS

##### Subsection A: Data Flow and Report Generation Process

The table below summarizes the completion of the management of study data.

| | Platform | | |
|---|---|---|---|
| | ACTIV4A | ATTACC | REMAP |
| Who is responsible for data cleaning | SOCAR/PITT | SOCAR | Each Data Provider (SPIRAL, Research Online, and UPMC) and the DCC (Monash/ICNARC) |
| Who is responsible for calculation of the efficacy and safety outcomes | PITT | SOCAR | Each Data Provider |
| QC process for identifying data issues/errors | SOCAR/PITT | SOCAR | Data Providers/DCC |
| QC of the score program calculations | PITT | SOCAR | Data Providers |
| How are the data physically transferred to the SAC (who, security measures, firewalls between study team and SAC) | SOCAR has set-up a secure sFTP server | SOCAR has set-up a secure sFTP server | sFTP from SPIRAL, Research Online, UPMC. SAC has internal Firewall, secure folders |
| How are the data combined by the SAC | There is a data format controlled by the SAC, which is how the data are submitted. There is internal data mapping by the SAC. | | |
| QC process of SAC for interim efficacy analysis and feedback mechanism | Internal processes for the interim analysis process. Programs validated, verification of data, to programs, to report. | | |
| QC process of each platform for interim safety analysis | | Validation is done by an independent programmer | |

#### Subsection B: Joint Efficacy Analysis Plan

*Arms*

Control Arm: The prophylactic-dose anticoagulation arm.
Treatment Arm: The therapeutic-dose anticoagulation arm.

*Analysis Population*

The efficacy analyses will be conducted on the joint anticoagulation PCR positive populations. This includes each patient who was randomized to either the control arm or the treatment arm who had one or more PCR positive test results.

REMAP-CAP randomizes patients for the different domains based on proven or suspected COVID-19. They have been randomizing COVID-19 patients since March, where testing was not routine or reliable. Over time testing has become much more reliable. Only REMAP-CAP patients that have positive COVID-19 will be included in this combined analysis.

If new arms are added to this joint analysis, this definition would include patients randomized to that additional arm. For clarity: REMAP-CAP is enrolling patients to other domains; a patient randomized to other domains but not the anticoagulation domain would not be included in this analysis. Patients randomized in the anticoagulation domain may be randomized within other domains. The mean number of doamins for each patient is between 2-3, with a maximum of 7 domains thus far. The status of randomization to other domains will not be included in this joint analysis of efficacy for anticoagulation.

*Primary Endpoint*

The primary endpoint is an ordinal scale that is a composite of in-hospital mortality and organ support-free days (OSFD) through 21 days. Organ support is defined as delivery of one or more of the following while admitted to an ICU:

1. Receipt of invasive or non-invasive mechanical ventilatory support
2. High-flow nasal oxygen
3. Vasopressor therapy

The time on organ support is defined as the time from the start of organ support in ICU to the last use of organ support (intervening time off organ support between the first and last use are considered on organ support) during any and each ICU admission in the 21 days after randomization. Time out of ICU prior to the end of study day 21 is counted as organ support-free. The organ support "free-days" is defined as 21 days minus the time on organ support in ICU. If a patient dies in their acute hospital stay, then they are assigned the worst outcome value of –1. For patients that survive the hospital stay, they are assigned the number of organ support-free days, rounded to the nearest integer number of days. The best possible outcome of 22 is reserved for patients who survive and never have organ support in ICU. There are 24 possible outcomes for this ordinal outcome: all integers from –1 to 22. Higher values correspond to better patient outcomes.

##### *Patient Subtypes*

There are four patient subgroups – denoted *subtypes* – that have separate inferences for anticoagulation in this analysis. The subtypes are based on a cross classification of the baseline disease state severity (receiving organ support with ICU level of care vs not) and for those not receiving ICU organ support, two baseline d-dimer groups defined as low or high. Patients missing d-dimer level will be included in a separate subtype that will help inform the overall model through the Bayesian hierarchical structure, but no adaptive triggers for superiority or futility will be actionable for this subtype. For notation purposes we label the disease severity state, as s=1, for patients not receiving organ support in ICU at baseline, and s=2 for patients receiving organ support in ICU at baseline. For s=1, we have three d-dimer levels, labeled, d=1 (d-dimer low), d=2 (d-dimer high), and d=3 (d-dimer is missing). We refer to these using the s:d notation, where the 4 subtypes are 1:1, 1:2, 1:3, and 2:0 (we use the d-dimer notation d=0 for the organ support in ICU disease state). We use the term "severe" for the patients receiving organ support in ICU at baseline and "moderate" for the patients not receiving organ support eg ICU level of care at baseline.

Table: Stratification of Random Assignment

| ACTIV4A | ATTACC | REMAP |
|---|---|---|
| Randomization assignments are at the participant level and are assigned at baseline. Randomization will be stratified by enrolling site and may also be stratified by severity of illness and/or other arm-specific criteria. In general, allocation will be equally distributed across arms for which the participant is eligible, but may be altered with future arm-specific appendices. | The adaptive aspects of the trial include response adaptive randomization within each of the 2 D-dimer subgroups as well as any potential conclusions (superiority, futility) for the effect of therapeutic anticoagulation within each of these 2 subgroups. | There are three units-of- analysis for this domain, each applying to patients who received an allocation status in this domain. They are: (1) all enrolled patients, (2) only patients in a moderate state at enrollment, and (3) only patients in a severe state at enrollment. Moderate and severe states are defined in Inclusions below. Units of analysis may be modified to allow analysis to be stratified by SARS-CoV-2 infection confirmed or not confirmed with borrowing permitted. If this occurs, Response Adaptive Randomization will be applied to patients in REMAP-COVID using probabilities derived from SARS-CoV-2 confirmed stratum. Additionally, the units of analysis may be modified to allow analysis by strata defined by D-dimer level. |

*Adaptive Decision Rules*

The following adaptive decision rules have been specified as labels for the efficacy outcomes. These rules are in place to determine the statistical triggers to label the treatment arm as superior to the control arm or to label the treatment arm as futile on the primary endpoint compared to the control arm. Each rule is separately evaluated within the three subtypes:

- moderate state: low d-dimer,
- moderate state: high d-dimer,
- severe state.

Efficacy conclusions will not be made for the subgroup of moderate patients with missing d-dimer levels.

Superiority

If the posterior probability of <u>superiority</u> within a subtype is greater than 99% then the treatment arm will be labeled as superior to the control arm for OSFDs.

Futility

If the posterior probability of at least an odds-ratio benefit of 1.2 is less than 5% then the treatment arm will be considered <u>futile</u> compared to the control arm.

*Report Contents*

The report will consist of the following displays:

- Consort diagram of samples contributing from each study
- descriptive statistics for the primary ordinal outcome (which statistics?) by treatment group – overall and by the 4 strata (including the unknown d-dimer group) once for the combined trials,
- posterior probability of success and posterior probability of futility for each of the 3 strata for the combined protocols , with indication of meeting the stopping criteria flagged.

*Modeling of the Primary Endpoint*

Let $Y_{\text{[illegible]}} = \{-1, 0, 1, \ldots, 21, 22\}$ denote the ordinal outcome (OSFD) for patient $i$. The probability of patient $i$, in state $s$, observing $y$ OSFD or less is denoted as $\pi_{\text{[illegible]}} = \Pr(Y_{\text{[illegible]}} \leq y)$. The model is a proportional odds model, where the log odds-ratio parameters in the model are structured so that a value > 0 implies treatment benefit, and an odds-ratio > 1 implies treatment benefit. The primary analysis model is formulated as follows:

$$\log\left(\frac{\pi_{\text{[illegible]}}}{1-\pi_{\text{[illegible]}}}\right) = \alpha_{\text{[illegible]}} - \left(\gamma_{\text{[illegible]}} + \nu_{\text{[illegible]}} + \lambda_{\text{[illegible]}} + \theta_{\text{[illegible]}} + \beta_{\text{[illegible]}} + \beta_{\text{[illegible]}} + \beta_{\text{[illegible]}}\right.$$

1. Each "platform," *P*, has a covariate adjustment in the model, *P*=1 is REMAP-CAP, *P*=2 is ATTACC, and *P*=3 is ACTIV-4.

2. The “site” variable is the clinical site within the trial. These will be set as distinct sites across all three trials. The site effects are estimated separately within the severe and moderate disease states, but do not vary by d-dimer level.
3. The “time” variable is an indicator of the time epoch in which a patient was enrolled in the trial, numbered increasing from the first most recent epoch to the earliest time for the analysis. Time epochs are two-week time periods, Time=1, and then every 2-week epoch, moving back in time throughout the enrollment in the trial, Time=2,3,4,…. The first two time periods are combined to form a 4-week category that serves as the referent. The time effects are modeled separately within the severe and moderate disease states, but do not vary by d-dimer level. The time epochs are defined to be the same time periods for both moderate and severe.
4. The “arm” to which a patient is randomized is labeled as *a* where *a*=1 is the control and *a*=2 is the treatment arm. The effects of arm vary by the disease state and the d-dimer level. The treatment effects for arm *a* within subtype *s*:*d* are modeled with the $\theta_{+,":-}$ parameters.
5. The “age” variable is a categorical classification of age as ≤ 39, 40-49, 50-59, 60-69, 70-79, and 80+. The age effects will be estimated separately within the moderate and severe disease states, but do not vary by d-dimer level.
6. The “sex” variable is sex at birth. The sex effects will be estimated separately within the moderate and severe disease states, but do not vary by d-dimer level.
7. The additive effects of d-dimer levels are modeled with the $\beta_{!}$ for d = 0, 1, 2, 3.
8. The $\alpha_{",\$}$ parameters determine the baseline rates of the ordinal outcome, which are modeled separately by disease state but not d-dimer level.

*Model Priors*

The ordinal endpoint rates are modeled using a Dirichlet prior where the individual probabilities for the 24 outcomes are based on one patient’s weight on real-world evidence-based outcomes (details of the vector *P* in the Anticoagulation Domain Current State).

$$logit\text{G}\alpha_{\#,"}\text{H}{\sim}Dirichlet(P)$$

For identifiability, the platform effect for ATTACC is defined such that $\gamma_1 \equiv 0$. The platform effects, $\gamma_{\%}$, $P = 1, 3$, are modeled with independent N(0, 1) priors.

The site effects, $\nu_{\&!'(}$, are modeled using a hierarchical model where site is nested within the country of the site:

$$\nu_{\&!'(,"}{\sim}N\text{G}\mu_{2345'6\#,"},\tau^{1}{}_{2345'6\#,}\text{H}, site = 1, \dots, N_{\&!'(}$$

$$\mu_{2345'6\#,"}{\sim}N(0,1);\ \ \tau^{1}{}_{2345'6\#,}{\sim}IG(0.25,0.1), s = 1,2$$

A referent country, the United States, will be set such that $\mu_{2345'6\#,7} \equiv \mu_{2345'6\#,1} \equiv 0$. The hyper-parameters of the site hierarchical model vary by disease state *s*.

The effect of age category is modeled with independent, normally distributed priors for each disease state. The prior mean for each age category effect is based on emerging data from external sources (specific details for the age prior in the Anticoagulation Domain Current State).

The effect of time (*Time=T*) is modeled using a second-order normal dynamic linear model separately by disease state, *s*. The two most recent time epochs are modeled as the referent time epochs with the time parameters set to 0. The preceding time epochs are modeled as a normal dynamic linear model as:

$$\lambda_7 = \lambda_1 \equiv 0$$

$$\lambda_8 \sim N(0, 0.15^1)$$

$$\lambda_) - 2\lambda_{)97} + \lambda_{)91} \sim N(0, \tau^1_{)!*}), T \geq 4$$

The treatment effect parameters are relative to a control arm, which will be labeled in the arm-specific appendix. The treatment effect for the control arm, labeled as arm *a* = 1, will be set to 0 for each of the disease subtypes and d-dimer level:

$$\theta_{7,":-} \equiv 0$$

The effects of the treatment arm are modeled hierarchically across disease subtype and d-dimer level. The treatment effects for each arm, *a*, are modeled as:

$$\theta_{:,1:;} \sim N\mathrm{G}\mu_{:,7}, \tau^1\ \mathrm{H}_{:,}$$

$$\theta_{+,7} \sim N\mathrm{G}\mu_{+,7}, \tau^1\ \mathrm{H}_{+,}$$

$$\theta_{:,<:=} \sim N\mathrm{G}\theta_{+,7}, \tau^1_{:,}\ \mathrm{H}, \qquad s{:}\,d = 1{:}\,1, 1{:}\,2, 1{:}\,3,$$

$$\mu_{+,7} \sim N(0, 1); \tau^1 \sim_{+,} IG(0.25, 0.1),$$

$$\tau^1 \sim_{+,} IG(0.25, 0.0025)$$

The effect of d-dimer is modeled with a N(0, 1) prior for d = 2,3, and the parameters $\beta_;$ and $\beta_7$ are set to 0 for identifiability. Any additional covariates included in the model will have independent N(0, 1) priors unless otherwise specified.

#### Subsection C: Joint Safety Analysis Plan

Two key Safety Outcome analyses for the multiplatform randomized-controlled trial (mpRCT) consortium, stratified by Trial (ACTIV4A, REMAP-CAP, and ATTACC) , will be presented to the DSMBs in addition to the efficacy analyses of the primary endpoint. The expected safety events for the current treatment strategies are major bleeding events, defined by the International Society on Thrombosis and Haemostasis (ISTH) criteria, and heparin induced thrombocytopenia (HIT) that occur within 15 days (24 hours after the assigned treatment stops) after randomization. Below are the definitions of the two specified safety endpoints.

##### *ISTH Defined Major Bleeding*

Bleeding that:

1. Resulted in death,
2. Intracranial hemorrhage
3. Occurred in a critical location (intraspinal, intraocular, retroperitoneal, intraarticular, intramuscular with compartment syndrome, or pericardial), or
4. Was associated with either a decrease in the hemoglobin level of at least 2 g per deciliter or transfusion of at least 2 units of packed red cells

##### *Heparin induced thrombocytopenia (HIT)*

HIT that is laboratory confirmed by anti-PF4 test or Serotonin Release Assay (SRA)

*Population*:

The mpRCT safety analyses will be conducted on the anticoagulation PCR positive participants that have had the opportunity to be followed for 21 days or more to be consistent with the efficacy analyses.

*Analyses*:

For each of these two safety endpoints, each trial will present the estimated event rates 15 days after randomization and the associated 95% confidence intervals for patients assigned to the Prophylactic-Dose Anticoagulation treatment group and for those assigned to the Therapeutic-Dose Anticoagulation treatment group. We will also compute the absolute risk difference and the relative risk ratio with their respective 95% confidence intervals. Results will be presented for the overall sample and stratified by the patient subtypes consistent with the mpRCT efficacy analyses. Subtypes are defined based on a cross classification of the baseline disease state severity and baseline d-dimer level. Those who are classified as having “severe” severity level (s=2), defined as receiving organ support in the ICU at baseline, comprise one stratum. Those who are classified as having “moderate” severity (s=1), defined as not receiving organ support in ICU at baseline, are further categorized by d- dimer level: low (d=1), high (d=2) and missing (d=3). As with the efficacy analysis, we will refer to these strata using the s:d notation, where the 4 subtypes are 1:1, 1:2, 1:3, and 2:0 (we use the d-dimer notation d=0 to indicate any d-dimer value with the severe disease state).

The table shown below will be completed by each trial and sent to each of the three trial DSMBs. Stopping rules for major bleeding and HIT are not specified. These analyses will be presented along with the pooled multiplatform analyses of the primary efficacy endpoint. A decision to stop a treatment arm would need to balance the bleeding and HIT risks with any observed benefit on the primary endpoint. We leave decisions to recommend stopping a treatment arm for safety reasons to the DSMB given the overall risk/benefit profile.

*Safety Analyses*

**ISTH Major Bleeding and Heparin Induced Thrombocytopenia (HIT) Events at 15-days[@] Overall and Stratified by Patient Subtype defined by Disease Severity and D-Dimer**

| Safety Endpoints at 15-days | Quantity | Prophylactic-Dose Anticoagulation Event Rate | Therapeutic-Dose Anticoagulant Event Rate | Risk Difference* | Risk Ratio** |
|---|---|---|---|---|---|
| **Overall** | | | | | |
| ISTH Major Bleeding | Number of Patients with Events | X1 | X2 | | |
| | Total Number of Patients | N1 | N2 | | |
| | Estimate | X1/N1 | X2/N2 | Risk Diff | Risk Ratio |
| | 95% Conf Interval | LB1, UB1 | LB2, UB2 | 95% CI | 95% CI |
| HIT | Number of Patients with Events | X1 | X2 | | |
| | Total Number of Patients | N1 | N2 | | |
| | Estimate | X1/N1 | X2/N2 | Risk Diff | Risk Ratio |
| | 95% Conf Interval | LB1, UB1 | LB2, UB2 | 95% CI | 95% CI |
| **Moderate Disease Severity / Low D-Dimer (1:1)** | | | | | |
| ISTH Major Bleeding | Number of Patients with Events | X1 | X2 | | |
| | Total Number of Patients | N1 | N2 | | |
| | Estimate | X1/N1 | X2/N2 | Risk Diff | Risk Ratio |
| | 95% Conf Interval | LB1, UB1 | LB2, UB2 | 95% CI | 95% CI |
| HIT | Number of Patients with Events | X1 | X2 | | |
| | Total Number of Patients | N1 | N2 | | |
| | Estimate | X1/N1 | X2/N2 | Risk Diff | Risk Ratio |
| | 95% Conf Interval | LB1, UB1 | LB2, UB2 | 95% CI | 95% CI |
| **Moderate Disease Severity / High D-Dimer (1:2)** | | | | | |
| ISTH Major Bleeding | Number of Patients with Events | X1 | X2 | | |
| | Total Number of Patients | N1 | N2 | | |
| | Estimate | X1/N1 | X2/N2 | Risk Diff | Risk Ratio |
| | 95% Conf Interval | LB1, UB1 | LB2, UB2 | 95% CI | 95% CI |
| HIT | Number of Patients with Events | X1 | X2 | | |
| | Total Number of Patients | N1 | N2 | | |
| | Estimate | X1/N1 | X2/N2 | Risk Diff | Risk Ratio |
| | 95% Conf Interval | LB1, UB1 | LB2, UB2 | 95% CI | 95% CI |
| **Moderate Disease Severity / Missing D-Dimer (1:3)** | | | | | |

| | | | | | |
|---|---|---|---|---|---|
| ISTH Major Bleeding | Number of Patients with Events | X1 | X2 | | |
| | Total Number of Patients | N1 | N2 | | |
| | Estimate | X1/N1 | X2/N2 | Risk Diff | Risk Ratio |
| | 95% Conf Interval | LB1, UB1 | LB2, UB2 | 95% CI | 95% CI |
| HIT | Number of Patients with Events | X1 | X2 | | |
| | Total Number of Patients | N1 | N2 | | |
| | Estimate | X1/N1 | X2/N2 | Risk Diff | Risk Ratio |
| | 95% Conf Interval | LB1, UB1 | LB2, UB2 | 95% CI | 95% CI |
| **Severe Disease Severity (2:0)** | | | | | |
| ISTH Major Bleeding | Number of Patients with Events | X1 | X2 | | |
| | Total Number of Patients | N1 | N2 | | |
| | Estimate | X1/N1 | X2/N2 | Risk Diff | Risk Ratio |
| | 95% Conf Interval | LB1, UB1 | LB2, UB2 | 95% CI | 95% CI |
| HIT | Number of Patients with Events | X1 | X2 | | |
| | Total Number of Patients | N1 | N2 | | |
| | Estimate | X1/N1 | X2/N2 | Risk Diff | Risk Ratio |
| | 95% Conf Interval | LB1, UB1 | LB2, UB2 | 95% CI | 95% CI |

@ Safety endpoints are defined as events occurring within 15 days of randomization. The population is defined as participants with at least 21 days of follow-up so that the sample population presented in this table is consistent with the sample used for the corresponding mpRCT efficacy analyses.

* Risk Difference = (X1/N1) – (X2/N2)
** Risk Ratio = (X1/N1) / (X2/N2)

#### Subsection D: Timeline

Interim Analyses

- Interim analyses will be sent monthly
- Data lock will occur on the first working day of the month
- Efficacy
    - Safety data from ATTACC, REMAP, and ACTIV4A will be sent to the Statistical Analysis Committee to complete the interim efficacy analyses
    - Interim efficacy results will be sent to the appointed representative of each DSMB
- Safety
    - Safety analyses will be tabulated by each platform
    - Platform-specific results will be sent from each platform to the appointed representative of each DSMB

Full Standard Open and Closed Reports of Patient enrollment, data quality, and complete patient safety including SAEs for ACTIV 4A Meetings will occur quarterly.

- The initial report will take place in January, 2021.
- Reports will include data from ACTIV 4A participants
- Tables will be compiled by the unblinded analysis team within the ACTIV 4A Data Coordinating Center and distributed to the members of the DSMB

Note that the data quality of the PROTECT, ATTACC, and REMAP were reviewed by their respective DSMBs.

#### Subsection E: Masking/Blinding

Masked/blinded: Drs. Berry, Hochman, Neal

Unmasked/unblinded: Dr. Brooks

Protocol 1: Randomized, Embedded, Multifactorial Adaptive Platform Trial for Community-Acquired Pneumonia (REMAP-CAP)

REMAP-CAP Core Protocol

### Randomized, Embedded, Multifactorial Adaptive Platform trial for Community-Acquired Pneumonia (REMAP-CAP): CORE PROTOCOL

REMAP-CAP Core Protocol Version 3 dated 10 July 2019

Not for RB submission

Not for IRB Submission

5.3.1. Amendments ... 87
**5.4. Confidentiality ... 88**
**5.5. Declarations of interest ... 88**
**5.6. Post-trial care ... 88**
**5.7. Communication ... 88**
5.7.1. Reporting ... 88
5.7.2. Communication of trial results ... 88
**5.8. Publication policy ... 89**
**5.9. Data access and ownership ... 89**
5.9.1. Data ownership ... 89
5.9.2. Access to Data ... 89
**5.10. Consent form ... 90**
**6. REFERENCES ... 91**

#### TABLE OF TABLES

Table 1: Potential targets of interventions to reduce mortality in patients with CAP ... 28
Table 2: Features of a REMAP that contribute to advantages of the design ... 42

#### TABLE OF FIGURES

Figure 1: Protocol Structure ... 15
Figure 2: REMAP-CAP Organization Chart ... 20
Figure 3: Adaptive Analyses ... 38
Figure 4: REMAP Evolution Over Time ... 41
Figure 5: REMAP Structure ... 44
Figure 6: Study Procedures ... 74

Not for IRB Submission

### 1. ABBREVIATIONS AND GLOSSARY

#### 1.1. Abbreviations

| | |
|---|---|
| ANZ | Australia and New Zealand |
| APACHE | Acute Physiology and Chronic Health Evaluation |
| ARDS | Acute Respiratory Distress Syndrome |
| BHM | Bayesian Hierarchical Model |
| CAP | Community-Acquired Pneumonia |
| CIHR | Canadian Institutes of Health Research |
| CIHR-SPOR | Canadian Institutes of Health Research Strategy for Patient-Oriented Research |
| CRF | Case Report Form |
| DSA | Domain-Specific Appendix |
| DSMB | Data Safety and Monitoring Board |
| DSWG | Domain-Specific Working Group |
| eCIS | Electronic Clinical Information System |
| eCRF | Electronic Case Report Form |
| EMA | European Medicines Agency |
| EU | European |
| FDA | Food and Drug Administration (United States) |
| GCP | Good Clinical Practice |
| HDU | High Dependency Unit |
| HRC | Health Research Council |
| HRQoL | Health Related Quality of Life |
| ICMJE | International Committee of Medical Journal Editors |
| ICU | Intensive Care Unit |
| IEIG | International Embedding Interest Group |
| IIG | International Interest Group |
| ILTOHEIG | International Long-term Outcomes and Health Economics Interest Group |
| IPWG | International Pandemic Working Group |
| ISIG | International Statistics Interest Group |

Not for IRB submission

| | |
|---|---|
| ITSC | International Trial Steering Committee |
| ITT | Intention-To-Treat |
| LOS | Length of Stay |
| NHMRC | National Health and Medical Research Council |
| OFFD | Organ Failure Free Days |
| P:F Ratio | Ratio of Partial Pressure of Oxygen in Arterial Blood and Fraction of Inspired Oxygen Concentration |
| PEEP | Positive End-Expiratory Pressure |
| PREPARE | Platform for European Preparedness Against (Re-)emerging Epidemics |
| RAR | Response Adaptive Randomization |
| REMAP | Randomized, Embedded, Multifactorial, Adaptive Platform trial |
| REMAP-CAP | Randomized, Embedded, Multifactorial, Adaptive Platform trial for Community-Acquired Pneumonia |
| RCC | Regional Coordinating Center |
| RCT | Randomized Controlled Trial |
| RMC | Regional Management Committee |
| RSA | Region-Specific Appendix |
| SAC | Statistical Analysis Committee |
| SAE | Serious Adverse Event |
| SARS | Severe Acute Respiratory Syndrome |
| SOPs | Standard Operating Procedures |
| VFD | Ventilator Free Days |
| WG | Working Group |
| WHODAS | World Health Organization Disability Assessment Schedule |

Not for IRB Submission

##### 1.2. Glossary

***Borrowing*** is the process within the statistical model, whereby, when the treatment effect is similar in different strata, evidence relating to the effectiveness of an intervention in one stratum contributes to the estimation of the posterior probability in another stratum.

***Core Protocol*** is a module of the protocol that contains all information that is generic to the Randomized, Embedded, Multifactorial, Adaptive Platform trial (REMAP), irrespective of the regional location in which the trial is conducted and the domains or interventions that are being tested.

***Domain-Specific Appendix*** is an appendix to the Core Protocol. These appendices are modules of the protocol that contain all information about the interventions, which are nested within a domain that will be a subject of this REMAP. Each domain will have its own Domain-Specific Appendix (DSA). The information contained in each DSA includes criteria that determine eligibility of patients to that domain, the features of the interventions and how they are delivered, and any additional endpoints and data collection that are not covered in the Core Protocol.

***Domain-Specific Working Group*** is a sub-committee involved in trial management, the members of which take responsibility for the development and management of a current or proposed new domain.

***Domain*** consists of a specific set of competing alternative interventions within a common clinical mode, which, for the purposes of the platform, are mutually exclusive and exhaustive. Where there is only a single intervention option within a domain the comparator is all other usual care in the absence of the intervention. Where multiple interventions exist within a domain, comparators are the range of interventions either with or without a no intervention option, depending on whether an intervention, within the domain, is provided to all patients as part of standard care. Within the REMAP every patient will be assigned to receive one and only one of the available interventions within every domain for which they are eligible.

***International Trial Steering Committee*** is the committee that takes overall responsibility for the management and conduct of the REMAP with oversight over all regions and all domains.

***Intervention*** is a treatment option that is subject to variation in clinical practice (comparative effectiveness intervention) or has been proposed for introduction into clinical practice (experimental intervention) and also is being subjected to experimental manipulation within the design of a

Not for IRB Submission

REMAP. For the purposes of the REMAP an intervention can include an option in which no treatment is provided.

***Monte-Carlo Simulations*** are computational algorithms that employ repeated random sampling to obtain a probability distribution. They are used in the design of the study to anticipate trial performance under a variety of potential states of 'truth' (e.g., to test the way in which a particular trial design feature will help or hinder the ability to determine whether a 'true' treatment effect will be discovered by the trial). Monte Carlo methods are also used to provide updated posterior probability distributions for the ongoing analyses of the trial.

***Pandemic Appendix*** describes an appendix to the Core Protocol that includes the modifications to the Core Protocol that will occur during a pandemic of respiratory infection that results in severe CAP.

***Platform Conclusion*** describes when a Statistical Trigger has been reached and, following evaluation by the Data Safety and Monitoring Board (DSMB) +/- the International Trial Steering Committee (ITSC), there is a *decision* to conclude that superiority, inferiority or equivalence has been demonstrated. Under all circumstances a Platform Conclusion leads to implementation of the result within the REMAP and under almost all circumstances a Platform Conclusion leads, immediately, to Public Disclosure of the result by presentation and publication. Where the Statistical Trigger is for superiority or inferiority, so long as the DSMB is satisfied that the Statistical Trigger has truly been met a Platform Conclusion will be automatic in almost all circumstances. Where the Statistical Trigger is for equivalence the DSMB, in conjunction with the ITSC, may decide to not reach a Platform Conclusion at that time but, rather, to continue recruitment, for example, to allow a conclusion to be reached regarding clinically important secondary endpoints. There are situations in which the need to evaluate interactions may also result in a Statistical Trigger not leading, immediately, to a Platform Conclusion, although if superiority or inferiority has been demonstrated all patients in the REMAP will receive the superior intervention or no longer be exposed to inferior intervention(s), respectively.

***Platform Trial*** is a type of clinical trial that studies multiple interventions simultaneously. Common features of a platform trial include frequent adaptive analyses using Bayesian statistical analysis, Response Adaptive Randomization (RAR), evaluation of treatment effect in pre-specified strata, and evaluation of multiple research questions simultaneously that can be perpetual with substitution of answered research questions with new questions as the trial evolves.

Not for IRB Submission

***Public Disclosure*** is the communication of a Platform Conclusion to the broad medical community by means of presentation, publication or both.

***Regimen*** consists of the unique combination of interventions, within multiple domains, (including no treatment options) that a patient receives within a REMAP.

***Region-Specific Appendix*** is an appendix to the Core Protocol. These appendices are modules of the protocol that contain all information about the trial specific to the conduct of the trial in that region. Each region will have its own Regional-Specific Appendix (RSA). A region is defined as a country or collection of countries with study sites for which a Regional Management Committee (RMC) is responsible.

***Regional Management Committee*** is a sub-committee involved in trial management. The members of the RMC take responsibility for the management of trial activities in a specified region. The role, responsibilities, and composition of each RMC are specified in each region's RSA.

***REMAP*** is a variant of a platform trial that targets questions that are relevant to routine care and relies heavily on embedding the trial in clinical practice. Like other platform trials, the focus is on a particular disease or condition, rather than a particular intervention, and it is capable of running perpetually, adding new questions sequentially.

***Response Adaptive Randomization*** is a dynamic process in which the analysis of accrued trial data is used to determine the proportion of future patients who are randomized to each intervention within a domain.

***State*** a state is a set of mutually exclusive and exhaustive categories, defined by characteristics of a patient within the REMAP, that are capable of changing over time for a single patient at different time-points during the patient's participation in the REMAP (i.e. they can be dynamic). States are used to define eligibility for domains and this can include defining eligibility that occurs after the time of enrollment. State is used as an additive covariate within the Bayesian statistical model.

***Statistical Analysis Committee*** takes responsibility for the conduct of the preplanned adaptations in the trial. This task generally consists of running predetermined statistical models at each adaptive analysis and providing this output to the DSMB. It is not a trial sub-committee. Rather, it will usually comprise individuals who are employed by the organization that undertakes statistical analysis, and from a trial governance perspective is under the supervision of the DSMB.

Not for IRB Submission

***Statistical Model*** is a computational algorithm that is used to estimate the posterior probability of the superiority, inferiority or equivalence of the regimens and interventions that are being evaluated within the REMAP.

***Statistical Trigger*** within the REMAP two or more interventions within a domain are evaluated and statistical models are used to determine if one or more interventions are superior, inferior or equivalent. A Statistical Trigger occurs when the statistical models used to analyze the REMAP indicate that the *threshold* for declaring superiority, inferiority, or equivalence for one or more interventions within a domain has been crossed. A Statistical Trigger applies to a stratum but may be reached in more than one stratum for the same intervention at the same adaptive analysis.

***Strata*** comprise a set of mutually exclusive and exhaustive categories (stratum), defined by baseline characteristics of a patient within the REMAP, in which the relative effects of interventions may be differential. These possibly differential effects of interventions are reflected in the statistical model, the randomization probabilities, and the Platform Conclusions. The criteria that define a stratum must be present at or before the time of enrollment.

***Unit-of-analysis*** is the group of patients who are analyzed together within the model for a particular domain. The unit-of-analysis can be all patients who have received an allocation status in that domain or a sub-group of patients who received an allocation status determined by their status with respect to one or more strata. Within a domain, the RAR is applied to the unit-of-analysis.

Not for IRB Submission

### 2. INTRODUCTION

#### 2.1. Synopsis

**Background**: Community-acquired pneumonia (CAP) that is of sufficient severity to require admission to an Intensive Care Unit (ICU) is associated with substantial mortality. All patients with severe pneumonia who are treated in an ICU will receive therapy that consists of a combination of multiple different treatments. For many of these treatments, different options are available currently. For example, several antibiotics exist that are active against the microorganisms that cause pneumonia commonly but it is not known if one antibiotic strategy is best or whether all suitable antibiotic strategies have similar levels of effectiveness. Of all the treatments that clinicians use for patients with severe CAP, only a small minority have been tested in randomized controlled trials to determine their comparative effectiveness. As a consequence, the standard treatments that are administered vary between and within countries. Current conventional clinical trials methods to assess the efficacy of treatments for pneumonia generally compare two treatment options (either two options for the same treatment modality, where both are in common use; or a new treatment against no treatment or placebo where the effectiveness of the new treatment is not known). Using this approach, in a series of separate and sequential trials, it will take an inordinate length of time to study all the treatment options. Additionally, with conventional trial designs it is not possible to evaluate interactions between treatment options.

**Aim**: The primary objective of this REMAP is, for patients with severe CAP who are admitted to an ICU, to identify the effect of a range of interventions to improve outcome as defined by all-cause mortality at 90 days.

**Methods**: The study will enroll adult patients with severe CAP who are admitted to ICUs using a design known as a REMAP, which is a type of platform trial. Within this REMAP, eligible participants will be randomized to receive one intervention in each of one or more domains (a domain is a category of treatment that contains one or more options, termed interventions, with each intervention option being mutually exclusive). The primary outcome is all-cause mortality at 90 days. There will also be both general and domain-specific secondary outcome measures.

In a conventional trial, enrollment continues until a pre-specified sample size is obtained, at which time enrollment ceases, and the trial data are analyzed to obtain a result. The possible results are

Not for IRB Submission

that a difference is detected or no that no difference is detected. However, when the conclusion of the statistical test is “no difference”, this could be that there truly is no meaningful difference, or that the result is indeterminate (i.e. it is possible that if more patients had been enrolled a clinically relevant difference may have been detected).

In comparison to a conventional trial, this REMAP uses an adaptive design, relying on pre-specified criteria for adaptation, that: avoids indeterminate results; concludes an answer to a question when sufficient data have accrued (not when a pre-specified sample is reached); evaluates the effect of treatment options in pre-defined subgroups of patients (termed strata); utilizes already accrued data to increase the likelihood that patients within the trial are randomized to treatments that are more likely to be beneficial; is multifactorial, evaluating multiple questions simultaneously; is intended to be perpetual (or at least open-ended), substituting new questions in series as initial questions are answered; and can evaluate the interaction between interventions in different domains. Bayesian statistical methods will be used to establish the superiority, inferiority, or equivalence of interventions within a domain. Interventions determined to be superior will be incorporated into standard care within the ongoing REMAP. Interventions determined to be inferior will be discontinued. While a limited number of initial treatments and treatment domains have been specified at initiation, it is planned that this REMAP will continue to evaluate other treatments in the future. Furthermore, in the event of a future epidemic of a novel or re-emerging respiratory pathogen (which typically present as severe CAP), this REMAP would be adapted to evaluate the most relevant treatment options. Each new treatment that is proposed to be evaluated within the REMAP will be submitted for prospective ethical review.

#### 2.2. Protocol Structure

The structure of this protocol is different to that used for a conventional trial because this trial is highly adaptive and the description of these adaptations is better understood and specified using a ‘modular’ protocol design. While all adaptations are pre-specified, the structure of the protocol is designed to allow the trial to evolve over time, for example by the introduction of new domains or interventions or both (see glossary for definitions of these terms), by changing aspects of the trial during a pandemic, and commencement of the trial in new regions. The structure of the protocol is outlined in Figure 1.

Not for IRB submission

*Figure 1: Protocol Structure*

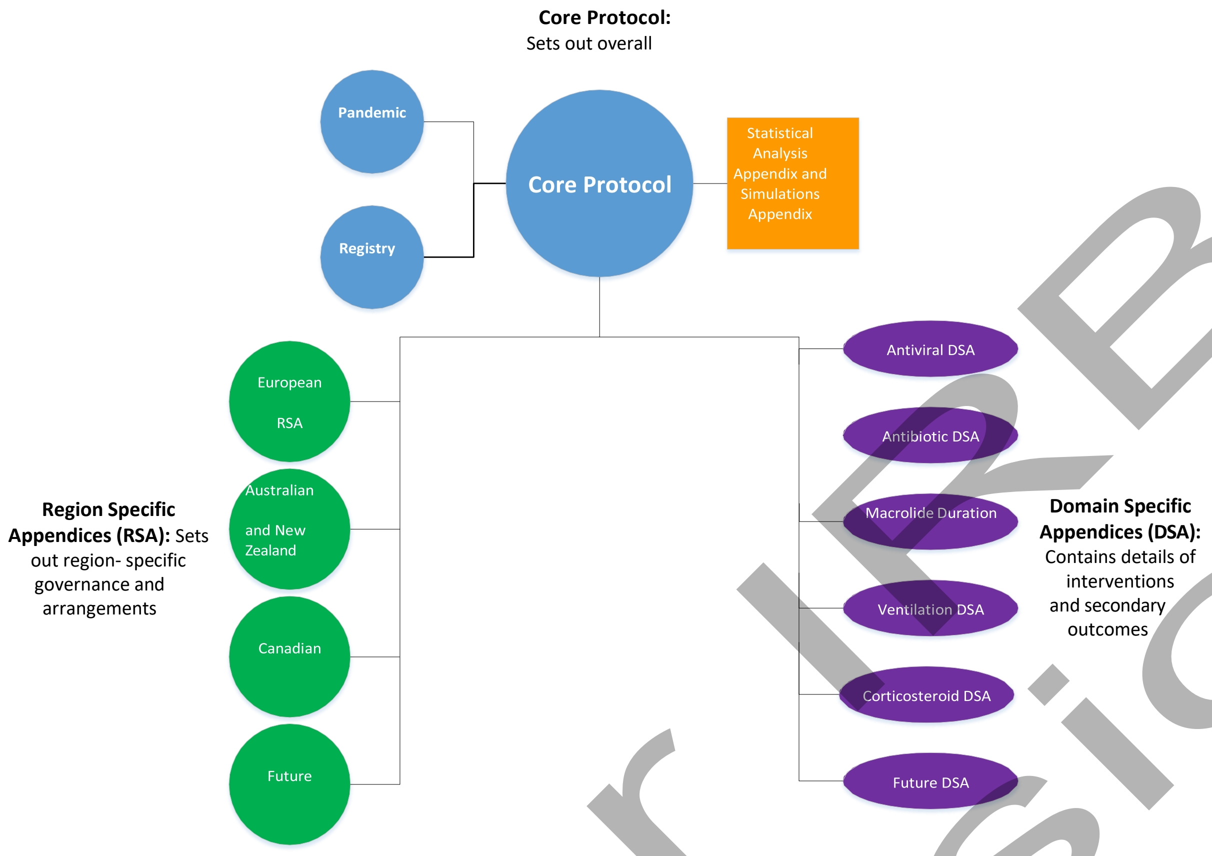

The protocol has multiple modules, comprising a Core Protocol, Pandemic Appendix to the Core Protocol, multiple DSAs, multiple RSAs, and a Statistical Analysis Appendix. A Pandemic Appendix to the Core Protocol is intended to be added subsequently. A Simulations Appendix is updated periodically as an operational document.

##### 2.2.1. Core Protocol

The Core Protocol contains all information that is generic to the trial, irrespective of the regional location in which the trial is conducted and the domains or interventions that are being tested. The Core Protocol may be amended but it is anticipated that such amendments will be infrequent. The Core Protocol has the following structure:

- The background and rationale for studying severe CAP
- The background and rationale for the research approach
- The trial design including study setting, the criteria that define eligibility for the REMAP, treatment allocation, strata (see glossary for a definition of this term), principles of application of trial interventions, trial endpoints, methods to control bias, principles of statistical analysis, and criteria for termination of the trial

Not for IRB submission

- The trial conduct including recruitment methods, time-lines for sites, delivery of trial interventions, data collection, data management, and management of participant safety
- The overall / international trial governance structures and ethical considerations

##### 2.2.2. Domain-Specific Appendices

DSAs contain all information about the interventions that will be the subject of the REMAP, which are nested within domains. As such, the Core Protocol does not include information about the intervention(s) that will be evaluated within the REMAP, but rather provides the framework on which multiple different interventions, within domains, can exist within this trial. Each new DSA or addition of one or more interventions to an existing DSA will be submitted for ethical approval prior to commencement. It is anticipated that the DSAs will change over time with removal and addition of interventions within an existing domain, as well as removal and addition of entire domains. Each DSA has the following structure:

- background on the interventions within that domain
- criteria that determine eligibility of patients to that domain
- the features of the interventions and how they are delivered
- any endpoints and data collection that are specific to the domain and additional to those specified in the Core Protocol
- any ethical issues specific to the domain
- the organization of management of the domain

##### 2.2.3. Region-Specific Appendices

This REMAP is intended to be a global trial, conducted in multiple different geographical regions. The RSAs contain all information about the REMAP that is specific to the conduct of the trial in a particular region. This allows additional regions to be added or changes to each region to be made without needing to make major amendments to the Core Protocol in other regions. It is planned that, within each region, the documents submitted for ethical review will comprise the Core Protocol, DSAs, and the RSA for that region (but not other regions). Each RSA has the following structure:

- the definition of the region
- the organization of trial management and administration within the region
- information about availability of domains and interventions
- data management and randomization procedures

Not for IRB Submission

- ethical issues that are specific to a region.

If there is information that applies to one or more sub-areas of a region (e.g. a country within Europe or a state or territory within a country) and it is necessary to incorporate this information in the protocol, this information will be included within the RSA. Unless otherwise specified, the RSA will apply to all locations within that region.

##### 2.2.4. Statistical Analysis Appendix and Simulations Appendix

The Statistical Analysis Appendix contains a detailed description of how the statistical analysis will be conducted for reporting treatment effects and reporting interaction between treatments, as well as the RAR. The Statistical Analysis Appendix will be amended when new interventions are added to a domain or when a new domain is added, but will not be updated when interventions are removed from a domain because of inferiority.

The Simulations Appendix is an operational document that contains the results of Monte Carlo simulations that are conducted to describe and understand the operating characteristics of the REMAP across a range of plausible assumptions regarding outcomes, treatment effects, and interactions between interventions in different domains. The statistical power of the study (likelihood of type II error) and the likelihood of type I error are evaluated using these simulations. As the trial adapts, with, for example, the introduction of new interventions, the trial simulations are updated and the Simulations Appendix is amended. The Simulations Appendix is not part of the formal protocol but the conclusions from the Simulations Appendix will be included in protocol documents which will be updated as required. The Simulations Appendix will be maintained as a publicly accessible document on the study website.

##### 2.2.5. Pandemic Appendix

The Pandemic Appendix (to the Core Protocol) contains information about how the core elements of the REMAP will be modified during a pandemic of severe acute respiratory infection that results in CAP. The Pandemic Appendix has the following structure:

- The background and rationale for studying severe CAP caused by a pandemic
- The procedure that will determine activation of the Pandemic Appendix
- How the trial design adapts during a pandemic, including changes to one or more of study setting, treatment allocation, strata, trial endpoints, and principles of statistical analysis that

Not for IRB Submission

will operate during a pandemic, as well as how the platform resets following a resolution of a pandemic

##### 2.2.6. Version History

Version 1: Approved by the ITSC on 20 November 2016

Version 1.1: Approved by the ITSC on 10 April 2017

Version 2: Approved by the ITSC on 12 December 2017

Version 2.1: Approved by the ITSC on 26 March 2019

Version 3: Approved by the ITSC on 10 July 2019

#### 2.3. Lay Description

Pneumonia, or infection involving the lungs, is a common reason for admission to an ICU. Severe pneumonia is associated not only with failure of lungs supplying oxygen to the body, but also failure of other organ systems that is due to an uncontrolled immune response to infection.

Patients with severe pneumonia routinely receive multiple treatments at the same time – medications to treat the infection (antibiotics), medications that may modify the immune system (immunomodulators) and supportive treatments to support failing organs, such as mechanical ventilation (organ support) and prevention of complications of critical illness or its treatment. For many categories of treatment there are many treatment options that are in widespread use, are believed or known to be safe and effective, but it is not known which option is best. This REMAP aims to determine the best treatment in each category of treatment, for example, the best antibiotic, the best immunomodulation strategy, and the best method to support each failing organ system.

In a conventional clinical trial, selected patients are allocated to receive one treatment from a short list of alternatives, typically one or two. This trial differs from conventional clinical trials by being randomized, embedded, multifactorial, adaptive, and a platform (a "REMAP"). (Angus, 2015) In this type of trial, we will test many alternative treatments ("multifactorial") by replacing *ad hoc* treatment decisions with "randomized" treatment allocation ("embedded"). Although treatments will be allocated randomly, patients will preferentially be allocated to treatments that statistical models derived from trial data indicate are more likely to be the most effective treatments. The trial will "adapt" in multiple ways including answering questions as soon as sufficient data have accrued

Not for IRB Submission

to answer the question of the effectiveness of each treatment and by changing the treatments that are being tested over-time so as to progressively determine the best package of treatments for pre-defined categories of patients with severe pneumonia. Once a treatment is identified as being optimal it is subsequently routinely provided to all eligible patients within the REMAP. The REMAP is also designed to adapt to test relevant interventions during a pandemic caused by lung infection that results in severe pneumonia.

##### 2.4. Trial registration

This is a single trial conducted in multiple regions, but will, as a minimum, be registered with ClinicalTrials.gov. The trial registration number is: NCT02735707.

The Universal Trial Number is: U1111-1189-1653.

##### 2.5. Funding of the trial

At initiation, the trial had funding from the following sources.

The Platform for European Preparedness Against (Re-)emerging Epidemics (PREPARE) consortium is funded by the European Union (FP7-HEALTH-2013-INNOVATION-1, grant number 602525). Within the PREPARE consortium, the trial has funding for the recruitment of approximately 4000 patients.

In Australia, the trial has been funded by the National Health and Medical Research Council (NHMRC) (APP1101719) for AUD $4,413,145, for the recruitment of 2000 patients.

In New Zealand, the trial has been funded by the Health Research Council (HRC) (16/631) for NZD $4,814,924, for the recruitment of 800 patients.

In Canada, the trial has been funded by the Canadian Institute of Health Research, Strategy for Patient-Oriented Research (CIHR-SPOR) Innovative Clinical Trials Program Grant (no. 158584) for CAD $1,497,200, for the recruitment of 300 patients.

Funding is being sought for other regions and countries.

#### 3. STUDY ADMINISTRATION STRUCTURE

The study administration structure is designed to provide appropriate management of all aspects of the study, taking into account multiple factors including representation from regions that are participating in the trial, availability of skills and expertise related to trial conduct and statistical

Not for IRB submission

analysis, and content knowledge regarding pneumonia and the interventions that are being evaluated. The administration model is designed to provide effective operational and strategic management of the REMAP that operates in multiple regions, is supported by multiple funding bodies and sponsors, and will evolve with addition of further regions and funding bodies as well as changes in the domains and interventions that are being evaluated.

The ITSC takes overall responsibility for the trial design and conduct. Each participating region has a RMC that takes primary responsibility for trial execution in that region. An internationally based Domain-Specific Working Group (DSWG) exists for each domain (or for several domains that are closely related) and has responsibility for design and oversight of each domain. Internationally based Interest Groups exist to allow discussion and development of particular aspects of the REMAP related to statistical analysis, embedding, and health economic analysis of results from the trial.

The organizational chart for REMAP-CAP is outlined in Figure 2.

*Figure 2: REMAP-CAP Organization Chart*

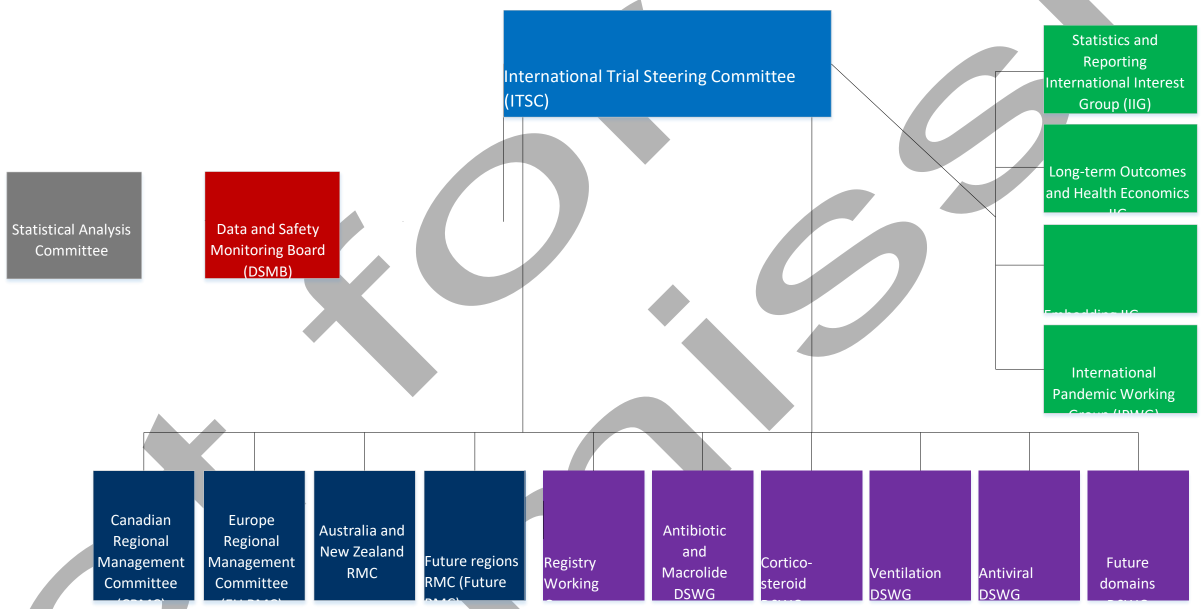

#### 3.1. International Trial Steering Committee

The ITSC comprises the investigators who initially conceived and designed the trial (Foundation members) and representatives from each (funded and active) region. The intent of the ITSC is to have both theoretical and practical experience and knowledge regarding overall design, domain-specific expertise, and regional-specific expertise. As such, the ITSC will include clinical trialists, biostatisticians, regional lead investigators, domain lead investigators, and regional project managers, and must include one individual who is a Research Coordinator.

Not for IRB Submission

##### 3.1.1. Responsibilities

The responsibilities of the ITSC are:

- development and amendment of the Core Protocol
- recruitment and approval of new regions to the REMAP
- liaison with the DSMB including, where appropriate, decisions regarding Platform Conclusions
- consideration of requests and approval of the addition of domains and their nested interventions to the REMAP including prioritization of new domains, new interventions within a domain or both
- liaison with the academic community including the International Committee of Medical Journal Editors (ICMJE) regarding issues such as data sharing and reporting of platform trials including REMAPs
- in conjunction with DSWGs, the analysis and reporting of results from domains
- approval of manuscripts reporting results that are submitted by DSWGs
- coordination of the REMAP during a pandemic
- obtaining funding for the REMAP
- determine the strategic direction of the REMAP

##### 3.1.2. Members

Membership of the ITSC comprises at least 3 investigators from each funded location, the project manager or trial physician in each funded location, at least 1 investigator from Berry Consultants, at least one individual who is a research coordinator, and the chairs of active DSWGs. The operation of the ITSC will be specified by Terms of Reference that will be developed and modified, as required, by the ITSC. The members of the ITSC are:

Professor Derek Angus, Chair Corticosteroid DSWG and Foundation member Ms.

Wilma van Bentum-Puijk, European (EU) Project Manager

Dr. Scott Berry, President and Senior Statistical Scientist of Berry Consultants, and Foundation member

Ms. Zahra Bhimani, Canadian Project Manager

Professor Marc Bonten, European Executive Director, Chair European RMC, and PREPARE Work Package 5 co-lead (specific issues)

Professor Frank Brunkhorst, member EU RMC

Not for IRB Submission

Professor Allen Cheng, Chair Antibiotic Domain and Macrolide Duration DSWG Professor

Menno De Jong, member Antiviral DSWG

Dr. Lennie Derde, European Coordinating Investigator, PREPARE Work Package 5 co-lead (specific issues)

Professor Herman Goossens, Principal Investigator for PREPARE Professor

Anthony Gordon, member EU RMC

Mr. Cameron Green, Global Project Manager

Professor Roger Lewis, Foundation member (will step down when SAC is convened) Dr. Ed

Litton, member Australian and New Zealand (ANZ) RMC

Professor John Marshall, Canadian Executive Director

Dr. Colin McArthur, ANZ Deputy Executive Director and Chair Registry WG Dr. Shay

McGuinness, Chair ANZ RMC

Associate Professor Srinivas Murthy, Canadian Deputy Executive Director and Chair Antiviral DSWG

Professor Alistair Nichol, Chair Ventilation DSWG Associate

Professor Rachael Parke, member ANZ RMC Ms. Jane Parker,

Australian Project Manager Professor Kathy Rowan, member EU

RMC

Ms. Anne Turner, New Zealand Project Manager

Professor Steve Webb, ANZ Executive Director and Foundation member

###### 3.1.3. Contact Details

The secretariat functions of the ITSC will rotate among the Regional Coordinating Centers (RCC).

#### *3.2. Regional Management Committees*

The operation of the REMAP in each region is undertaken by that region's RMC, the composition of which is be determined by investigators in each region with membership listed in each RSA. Cross-representation between RMCs is strongly encouraged.

Not for IRB Submission

##### 3.2.1. Responsibilities

The responsibilities of each RMC are:

- development and amendment of the RSA for that region
- identification and management of sites in that region
- obtaining funding for that region
- liaison with regional funding bodies
- consideration of the feasibility and suitability of interventions (and domains) for that region
- liaison with the sponsor(s) for that region
- management of systems for randomization and data management for that region

#### 3.3. Domain-Specific Working Groups

Each active and future planned domain (or closely related set of domains) will be administered by a DSWG.

##### 3.3.1. Responsibilities

The responsibilities of each DSWG are:

- development and amendment of the DSA
- proposal and development of new interventions within a domain
- in conjunction with the ITSC, analyzing and reporting results from the domain
- obtaining funding to support the domain, with a requirement that, if such funds are obtained, that an appropriate contribution to the conduct of the REMAP is also made.

##### 3.3.2. Members

Membership of each DSWG is set out in the corresponding DSA but should comprise individuals that provide broad international representation, content knowledge of the domain, and expertise of trial conduct and design.

#### 3.4. International Interest Groups

The following International Interest Groups (IIG) contribute to the trial:

- REMAP-CAP International Statistics Interest Group (ISIG)
- REMAP-CAP International Embedding Interest Group (IEIG)

Not for IRB submission

- REMAP-CAP International Long-term Outcomes and Health Economics Interest Group (ILTOHEIG)
- REMAP-CAP International Pandemic Working Group (IPWG)

###### 3.4.1. Role

The role of the interest groups is to provide advice to the ITSC and DSWGs about trial design and conduct as well as advance academic aspects of the conduct, analysis, and reporting of platform trials including REMAPs.

##### 3.5. *Sponsors*

In relation to recruitment that occurs in:

- countries in Europe the sponsor is University Medical Center Utrecht.
- Australia the sponsor is Monash University.
- New Zealand the sponsor is the Medical Research Institute of New Zealand.
- Canada the sponsor is Unity Health Toronto.

###### 3.5.1. Role of sponsor

The role of the sponsor in each region is specified in each RSA.

###### 3.5.2. Insurance

The provision of insurance is specified in each RSA.

#### 4. INTERNATIONAL TRIAL STEERING COMMITTEE AUTHORIZATION

The ITSC have read the appendix and authorize it as the official Core Protocol for the study entitled REMAP-CAP. Signed by the ITSC,

**EU Executive Director** ____ 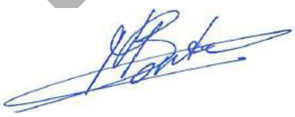
Marc Bonten

**ANZ Executive Director** ____ 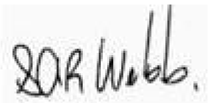
Steve Webb

Not for IRB Submission

**ANZ Deputy Director**
Colin McArthur

**ITSC Member**
Derek Angus

**ITSC Member**
Wilma van Bentum-Puijk

**ITSC Member**
Scott Berry

**ITSC Member**
Zahra Bhimani

**ITSC Member**
Frank Brunkhorst

**ITSC Member**
Allen Cheng

**ITSC Member**
Menno De Jong

**ITSC Member**
Lennie Derde

**ITSC Member**
Herman Goossens

**ITSC Member**
Anthony Gordon

**ITSC Member**
Cameron Green

**ITSC Member**
Roger Lewis

Not for IRB submission

**ITSC Member**
Ed Litton

**ITSC Member**
John Marshall

**ITSC Member**
Shay McGuinness

**ITSC Member**
Srinivas Murthy

**ITSC Member**
Alistair Nichol

**ITSC Member**
Rachael Parke

**ITSC Member**
Jane Parker

**ITSC Member**
Kathy Rowan

**ITSC Member**
Anne Turner

Not for IRB Submission

### 5. BACKGROUND & RATIONALE

#### *5.1. Severe Community-Acquired Pneumonia*

##### 5.1.1. Introduction

This section, within the Core Protocol, provides background on the epidemiology, causes, treatment categories, and evidence base for the management of patients with severe community pneumonia. Detailed information regarding the rationale for specific interventions to which patients will be randomized within the REMAP can be found in a corresponding DSA. As the trial is intended to be perpetual, if background information changes, appropriate amendments to the protocol documents will occur periodically, but it is anticipated that this will occur predominantly by amendment of DSAs.

##### 5.1.2. Epidemiology

CAP is a syndrome in which acute infection of the lungs develops in persons who have neither been hospitalized recently nor had regular exposure to the healthcare system. (Musher and Thorner, 2014) A wide range of micro-organisms are capable of causing pneumonia but bacteria and viruses are responsible for the vast majority of cases where a cause is identified. Severe CAP is defined as pneumonia of sufficient severity to be an immediate threat to life. In developed countries, patients with severe CAP are often admitted to an ICU or a High Dependency Unit (HDU). Throughout the remainder of this protocol, we will use the term ICU for units that provide specialized care for critically ill patients, including HDU, Critical Care Units, and Intensive Treatment Units. Although admission criteria may vary, the occurrence of admission to an ICU or a HDU can be used as an operational definition of severe CAP.

CAP is an important health problem and a common cause of death from infection globally, with lower respiratory tract infection, implicated in 3.1 million deaths in 2012, ranked as the 4th most common cause of death, although most of these deaths occur in low and middle-income countries. (Bjerre et al., 2009, Musher et al., 2013, Singanayagam et al., 2009) In developed countries, around half of patients with CAP are treated successfully without admission to hospital. (Almirall et al., 2000) Among patients who are admitted to hospital around 10 to 20% are admitted to an ICU. (Alvarez-Lerma and Torres, 2004, Ewig et al., 2011) The population incidence of CAP that involves admission to an ICU is about 0.4 cases per 1000 per year. (Finfer et al., 2004) Among patients admitted to an ICU with CAP, case-fatality is reported to be in the range from 20 to 50%. (Alvarez- Lerma and Torres, 2004, Leroy et al., 1995, Sligl and Marrie, 2013) In low and middle-income countries, the overlapping syndromes of CAP, bronchiolitis, and bronchitis are a major public health problem and represent the world's most important cause of disability-adjusted life years lost and the third most important cause of death. (World Health Organization, 2008)

##### 5.1.3. Standard care for patients with severe CAP

All patients admitted to an ICU with severe CAP will receive multiple different component therapies and many of these therapies will be administered concurrently. These therapies can be grouped into the following categories: treatment of the underlying infection (including antibacterial and antiviral

Not for IRB submission

agents); the optional use of agents, such as corticosteroids, that modulate the host immune response to infection; and multiple supportive therapies that are used to manage organ systems that have failed or prevent complications of critical illness and its treatment (Table 1).

The choice of empiric antimicrobial therapy is generally made before a microbiologic etiology is established, both because of the lag between collection of specimens and the availability of results from microbiological tests, and because microbiological tests lack sensitivity, particularly when samples are collected after initiation of antimicrobial therapy. It is recommended that antimicrobial treatment be initiated promptly and at the point of care where the diagnosis of pneumonia is first made. (Musher and Thorner, 2014)

Examples of commonly used therapies that support failed organ systems or prevent the complications of critical illness and its treatment include oxygen therapy, invasive and non-invasive mechanical ventilation, intravenous fluid resuscitation, vasoactive drugs, dialysis, provision of nutrition, sedation, physiotherapy including mobilization, diuretic medications, suppression of gastric acid production, and mechanical or pharmacological interventions to prevent venous thromboembolism. The exact combination of supportive therapies is influenced by the spectrum of organ failures that occurs in any individual patient. (Dellinger et al., 2013)

*Table 1: Potential targets of interventions to reduce mortality in patients with CAP*

| Target of intervention | Examples |
| --- | --- |
| **Eradication of pathogens** | Antibiotics (agents, route, dose)<br>Antivirals (agents, route, dose)<br>Microbiological diagnostic strategies |
| **Modulation of the host immune response** | Corticosteroid<br>Macrolides |
| **Methods to support failing organ systems and prevention of complications** | Lung ventilation strategies and respiratory salvage modalities (e.g. extra-corporeal membrane oxygen, prone positioning)<br>Renal replacement therapy<br>Inotropic/vasopressor support<br>Fluid resuscitation strategies |

| | |
|---|---|
| | Nutrition<br>Mobilization<br>Sedation<br>Venous thromboembolism prophylaxis<br>Stress ulcer prophylaxis |

##### 5.1.4. Treatment guidelines

A range of different guidelines have been published that are relevant to the care of critically ill patients with CAP. (Eccles et al., 2014, Lim et al., 2009, Mandell et al., 2007, Wiersinga et al., 2012, Wilkinson and Woodhead, 2004, Woodhead et al., 2011) These guidelines generally focus on recommendations related to assessment of severity, diagnostic evaluation, and empiric and guided antimicrobial therapy. Guidelines from the Surviving Sepsis Campaign are relevant to many aspects of the supportive care of the critically ill patients with CAP. (Dellinger et al., 2013)

There is a stark contrast between the substantial public health impact of severe CAP and the low quality of evidence that guides therapy. The number of treatment recommendations in guidelines that are supported by high quality randomized controlled trial (RCT) evidence is 4 of 44 for treatment recommendations in the European guidelines (Eccles et al., 2014, Lim et al., 2009, Woodhead et al., 2011), 11 of 43 in the United States guidelines (Mandell et al., 2007), and 7 of 93 in the Surviving Sepsis Campaign Guidelines. (Rhodes et al., 2017) As a consequence of the limited evidence-base there are a number of inconsistencies and even complete contradictions among international guidelines.

##### 5.1.5. Variation in care and compliance with guidelines

Several observational studies report substantial variation in care with, for example, compliance with administration of antibiotics recommended by guidelines occurring in between 40% and 75% of patients. (Bodi et al., 2005, Frei et al., 2010, Lee et al., 2014, Shorr et al., 2006) These and other studies also report better clinical outcomes for patients who received antibiotics that were recommended by guidelines. (McCabe et al., 2009, Mortensen et al., 2004, Mortensen et al., 2005) However, it remains unclear if adherence to guideline recommendations is due to a direct causal link, or whether it is a surrogate for better quality care generally. There is also widely reported variation in compliance with many supportive therapies for patients with severe CAP, such as use of

Not for IRB submission

low tidal volume ventilation, type of resuscitation fluid, and thresholds for the administration of transfusion for anemia. (Bellani et al., 2016, Finfer et al., 2010, Blood Observational Study Investigators of Anzics-Clinical Trials Group et al., 2010, Cecconi et al., 2015)

##### 5.1.6. An unmet need for better evidence

Many factors contribute to the substantial unmet need for better evidence to determine the optimal treatment for patients with severe CAP. Severe CAP is common, case-fatality is high, the strength of current evidence is limited, and there is evidence of substantial variation in existing standard care. The combination of these factors provides a strong rationale for the need for better quality evidence about the impact of the different treatment options that are in existing practice, the impact of different combinations of treatment options, and the timely and effective evaluation of new candidate interventions to improve outcomes.

#### *5.2. Influenza pandemics and emerging pathogens*

A pandemic of severe CAP caused by a known (e.g., influenza) or unknown virus, as occurred during the Severe Acute Respiratory Syndrome (SARS) outbreak, can rapidly change the etiological spectrum of severe CAP in patients who require admission to an ICU. This necessitates adaptation of empiric treatment protocols or diagnostic procedures or both. Naturally, there will be no evidence base for the medical management of such a disease at the time of its emergence, and medical decisions will be mostly based on expert opinion with extrapolation from evidence derived from the treatment of analogous clinical syndromes. There is substantial unmet need to generate evidence about the most effective treatment approaches during a pandemic or regional outbreak.
Furthermore, to have impact on patient outcomes during an outbreak, evidence must be available during the pandemic. As a consequence, such evidence must be capable of being generated, disseminated, and implemented rapidly. More detailed background information about pandemics of respiratory infection, together with challenges associated with the clinical research response are outlined in the Pandemic Appendix.

#### *5.3. Randomized Embedded Multifactorial Adaptive Platform Trials*

##### 5.3.1. Generating clinical evidence

Angus has noted several problems encountered when generating robust clinical evidence, including barriers to conducting clinical trials, the generalizability of data from populations that are too broad or too narrow, the issue of equipoise especially when comparing different types of existing care, and

Not for IRB Submission

the delay in translating results into clinical practice. (Angus, 2015) A REMAP provides a strategy to address many of these problems by gaining economies of scale from a common platform, which allows for broad enrollment but retaining the ability to examine for heterogeneity of treatment effects between defined subgroups. A REMAP focuses predominantly on the evaluation of treatment options for the disease of interest that are variations within the spectrum of standard care (although testing of novel or experimental therapies is not precluded) and does so by embedding the trial within routine healthcare delivery. In this regard the REMAP seeks to replace random variation in treatment with randomized variation in treatment allowing causal inference to be generated about the comparative effectiveness of different existing treatment options. The use of RAR, which allows the allocation ratios to change over time based on accruing outcomes data, maximizes the chance of good outcomes for trial participants. The embedding of such a platform within the day-to-day activities of ICUs facilitates the translation of outcomes to clinical practice as a "self-learning" system. As such, it also functions as an embedded and automated continuous quality-improvement program. A final advantage of a REMAP for pneumonia is the ability to rapidly adapt to generate evidence if new respiratory pathogens emerge, avoiding the inevitable delays associated with conventional trials in an outbreak of a new infectious diseases. (Burns et al., 2011)

##### 5.3.2. Underlying Principles of the Study Design

A REMAP applies novel and innovative trial adaptive design and statistical methods to evaluate a range of treatment options as efficiently as possible. The broad objective of a REMAP is, over time, to determine and continuously update the optimal set of treatments for the disease of interest. The set of treatments that may be tested within a REMAP comprise the set of all treatments that are used currently or may be developed in the future and used or considered for use in the disease of interest. The design maximizes the efficiency with which available sample size is applied to evaluate treatment options as rapidly as possible. A REMAP has the capacity to identify differential treatment effects in defined sub-groups (termed strata), address multiple questions simultaneously, and can evaluate interactions among selected treatment options. Throughout the platform, patients who are enrolled in the trial are treated as effectively as possible. (Angus, 2015, Berry et al., 2015, Carey and Winer, 2016, Harrington and Parmigiani, 2016, Park et al., 2016, Rugo et al., 2016)

A conventional RCT (i.e. a non-platform trial) makes a wide range of assumptions at the time of design. These assumptions include the plausible size of the treatment effect, the incidence of the primary outcome, the planned sample size, the (typically, small number of) treatments to be tested, and that treatment effects are not influenced by concomitant treatment options. These assumptions are held constant until the trial completes recruitment and is analyzed. (Barker et al., 2009, Berry,

Not for IRB submission

2012, Connor et al., 2013) Participants who are enrolled in a conventional RCT are not able to benefit from knowledge accrued by the trial because no results are made available until the trial completes. A REMAP uses five approaches to minimize the impact of assumptions on trial efficiency and also maximizes the benefit of participation for individuals who are enrolled in the trial. (Angus, 2015, Berry et al., 2015, Aikman et al., 2013, Carey and Winer, 2016, Harrington and Parmigiani, 2016, Park et al., 2016, Rugo et al., 2016)

These design features are:

- frequent adaptive analyses using Bayesian statistical methods
- RAR
- evaluation of differential treatment effects in pre-specified sub-groups (strata)
- evaluation of specified intervention-intervention interactions
- testing of multiple interventions in parallel and, subsequently, in series

This creates a ‘perpetual trial’ with no pre-defined sample size, the objective of which is to define and continuously update best treatment over the life-time of the REMAP. The design aspects, including the risk of type I and type II error, are optimized prior to the commencement of the trial by the conduct of extensive pre-trial Monte Carlo simulations, modification of the trial design, and re- simulation in an iterative manner. The methods related to the application of the design features and the statistical analysis of this trial are outlined in the methods section of the protocol (Section 7).
The following sections describe the background, rationale, and potential advantages of each of the design features of a REMAP (Section 5.3.4).

##### 5.3.3. Nomenclature

A specific set of nomenclature is used to categorize potential treatments evaluated and populations within a REMAP as well as other aspects of the trial design and statistical analysis. A detailed glossary can be found in Section 1.2. Please see the glossary for the definition and explanations for the following terms: domain, intervention, regimen, stratum, state, Statistical Trigger, Platform Conclusion, and Public Disclosure.

##### 5.3.4. Randomization and Response Adaptive Randomization

The study will randomly allocate participants to one or more interventions, with each intervention nested within a domain. In this regard, a platform trial is no different to other forms of RCT in that randomization provides the basis for causal inference. However, unlike a conventional RCT, the proportion of participants who are randomized to each available intervention within a domain will

Not for IRB Submission

not be fixed. Rather, the trial will incorporate RAR. RAR utilizes random allocation with a weighted probability for each intervention, with the weighted probability being proportional to the extent to which similar participants recruited earlier in the trial benefited or not from each particular intervention. (Angus, 2015, Berry, 2012, Connor et al., 2013, Aikman et al., 2013, Carey and Winer, 2016, Harrington and Parmigiani, 2016, Park et al., 2016, Rugo et al., 2016) RAR will result in participants in each particular stratum being randomized with greater probability to interventions that are performing better within that stratum. At the initiation of a new domain or when a new intervention is added to a domain the randomization proportion of all new interventions is balanced and only changes, with the application of RAR, that takes into account uncertainty about treatment effect so as to avoid excessive variability in proportions generated by RAR until sufficient sample size has accrued.

The major consequence of RAR is that better therapies move through the evaluation process faster, resulting in trial efficiency gains. (Berry, 2012, Connor et al., 2013) The platform “learns” more quickly about the treatments we ultimately care about, i.e. those that work best. Moreover, as data accrues, newly randomized participants are more likely to receive interventions from which they benefit. (Berry, 2012, Connor et al., 2013, Meurer et al., 2012, Angus, 2015, Carey and Winer, 2016, Harrington and Parmigiani, 2016, Park et al., 2016, Rugo et al., 2016) This is a highly ethical fusion of trial science with continuous quality improvement and a learning healthcare system. (Institute of Medicine, 2013) Assuming at least some interventions are better than others, the total mortality within the trial population will be lower than would have occurred with a fixed randomization proportion. It is also particularly relevant to the ethical conduct of trials that enroll critically ill patients where unanticipated increases in mortality have been seen (Dellinger et al., 2013) and to the conduct of trials during a pandemic in which there is in-built implementation of the therapies that are more likely to be beneficial during the trial. The simulations underpinning REMAP-CAP demonstrate that, in instances where particular interventions are indeed superior to others, the use of RAR will, on average, increase the odds of discovering the superiority not only with lower sample size, but with fewer participants exposed to the less efficacious therapies and, thus, fewer deaths.

There are potential disadvantages associated with RAR. It is intended that participating sites and trial investigators will be blind to the RAR proportions. One disadvantage is that, for interventions that are provided without blinding, the treating clinicians may be able to draw inference about the RAR proportions and, as a consequence, draw inference about the interim standing of interventions that are being tested in the REMAP. This could have adverse consequences including that clinicians are influenced to not enroll participants within a domain but rather directly prescribe the treatment that

Not for IRB Submission

they believe to be doing better outside the trial. However, a number of factors mitigate this potential concern. First, it can be difficult to distinguish between patterns of sequential allocation status that are derived from fixed versus RAR. Second, extreme proportions will not be used (except where a Statistical Trigger but not a Platform Conclusion has been reached, see later). Finally, for many conditions, team-based management means that an individual clinician will directly observe only a small proportion of all participants enrolled within the trial at each participating site. Another disadvantage of RAR is that, under certain allocation rules, statistical power can be reduced. This concern is mitigated via pre-trial simulation to test the effects of different allocation rules. Furthermore, a REMAP that comprises multiple domains with multiple interventions within each domain will generally have higher, rather than lower, power as a consequence of the use of RAR. Finally, by deploying RAR rules to minimize the odds of exposure to inferior interventions, the design is intended to motivate embedding in clinical practice, thereby resulting in more rapid recruitment.

Within each domain, RAR will be implemented for participants who are eligible to receive two or more interventions within a domain. Where a participant is eligible for only one option within a domain, this will be the treatment allocation for such a participant. In these circumstances, the provision of a treatment allocation status is made, predominantly, so as to provide a process that enhances the effectiveness of embedding, i.e. wherever possible the platform provides the treatment allocation.

##### 5.3.5. Embedding

A trial is most efficient when all eligible participants are recognized and enrolled. Achieving universal enrollment of eligible participants increases the speed with which new knowledge is generated, maximizes internal and external validity, and minimizes operational complexity at the bedside (there is no need to distinguish between trial and non-trial patients, because all patients are trial patients). A number of strategies will be utilized to very tightly "nest" or embed trial processes in daily clinical care operations. The effectiveness of strategies to achieve embedding will be evaluated, updated, and shared with sites, taking into account different clinical processes at different sites. Wherever possible trial treatment allocations will be integrated with electronic customized order sets, produced at the point of delivery of care that also includes each site's local care standards for concomitant therapies. This allows clinical staff to follow their typical workflow using protocolized order sheets to govern many aspects of patient care and serves to enhance compliance with the interventions allocated by the trial. The intention of embedding is that recruitment occurs 24/7 and is dependent on the usual medical staff who are responsible for patient care. Where possible electronic health records will be utilized to enhance screening and recruitment and specify the

Not for IRB submission

‘order set’ for participants, including those orders that are determined by allocation status within the REMAP. While screening and recruitment for a REMAP can be conducted by research staff, it is not intended that recruitment should be dependent on research staff, particularly as such staff are typically only present during office hours. In addition to the facilitation of recruitment and high-fidelity delivery of the intervention, a further advantage is that the results of the trial can be translated rapidly within the ongoing REMAP so that all appropriate participants receive a treatment declared to be superior with continued allocation to that treatment option within the REMAP used to ensure implementation.

##### 5.3.6. Multifactorial

If the trial randomizes in more than one domain of care it is multifactorial. The number of domains, at any time, is determined by a combination of the interventions that are appropriate and amenable for evaluation within the REMAP and the available statistical power, as determined by the conduct of simulations. It is intended that this REMAP will increase the number of domains, progressively, as the number of sites and rate of recruitment increases over time. The Bayesian models evaluate treatment effects (superiority, inferiority, equivalence) within each regimen but then, by isolating the effect of each intervention across all regimens in which that intervention is included, the independent effect of each intervention is estimated. The capacity to evaluate interventions within multiple domains, in parallel, increases trial efficiency substantially.

An additional advantage of the trial being multifactorial is the capacity to evaluate interactions between selected interventions in different domains. Where pre-specified, on the basis of clinical plausibility, statistical models will evaluate whether there is interaction between interventions in different domains. Where no interaction is suspected, interactions will not be evaluated as part of the *a priori* statistical model.

Although participants within a REMAP will, typically, receive treatment allocations for multiple domains the decision-making regarding concomitant therapies will be made by the treating clinician in other domains of care. Treatment decisions in other domains of care will be recorded and may be analyzed, using observational methods, to evaluate candidate interventions for evaluation by randomization within the REMAP.

Not for IRB Submission

##### 5.3.7. Adaptive

###### *5.3.7.1. Frequent adaptive analyses*

Frequent adaptive analyses using Bayesian statistical methods will be undertaken using Markov Chain Monte Carlo (MCMC) estimates of the Bayesian posterior probability distributions. The trial will utilize a set of pre-specified rules to reach conclusions regarding the effectiveness of interventions that are being evaluated. It is these pre-specified rules that determines how the trial "adapts" to the information contained in accumulating participant data. An analogy is that the 'routes' that a trial can take are pre-specified, within the protocol, but the exact route that the trial takes is determined by the data that accrues. Such adaptation improves statistical efficiency substantially.

###### *5.3.7.2. Analysis of data to reach conclusions*

The following structure and sequence of events will be used to reach conclusions from data as it accrues and is analyzed. This document, the Core Protocol, sets out the pre-specified rules for interpreting the results of analyses. These rules include pre-specified threshold levels of probability for achieving superiority, inferiority or equivalence of interventions within a domain. At each adaptive analysis the Statistical Analysis Committee (SAC) evaluates whether one or more probability thresholds that are derived from the trial's statistical model have been exceeded. When the model indicates one or more of superiority, inferiority, or equivalence has occurred this is termed a Statistical Trigger. A Statistical Trigger may be reached for one or more strata at any given adaptive analysis.

The occurrence of a Statistical Trigger is communicated immediately to the trial DSMB by the SAC. The DSMB has primary responsibility for determining if a Statistical Trigger should lead to a Platform Conclusion. The declaration of a Platform Conclusion results in the removal of inferior intervention from randomization options or removal of all other interventions if an intervention is declared as superior. A Platform Conclusion will be communicated to the ITSC who have responsibility for immediate dissemination of the result by presentation and publication of the result.

The algorithm by which a Platform Conclusion is reached is different for Statistical Triggers of superiority or inferiority, compared to those triggers that arise because of equivalence. Where the Statistical Trigger is for superiority or inferiority, so long as the DSMB is satisfied that the Statistical Trigger has been met validly, the default position is that the DSMB will declare this result as a Platform Conclusion. The only exception to this situation is if there is a need to evaluate potential interactions between treatments in different domains. In this circumstance the randomization

Not for IRB submission

schedule will be adapted (all participants receive the superior intervention or randomization to one or more inferior interventions is removed) but Public Disclosure may be delayed until evaluation of the interaction is completed.

Where the Statistical Trigger is for equivalence the DSMB will evaluate clinically relevant secondary endpoints. The results, in relation to both primary and secondary endpoints, will be communicated to the ITSC. The DSMB, in conjunction with the ITSC, may declare a Platform Conclusion (for equivalence) or may opt to continue recruitment and randomization to the 'equivalent' interventions, for example, to allow a conclusion to be reached regarding clinically important secondary endpoints, to allow additional accrual to narrow the margin of equivalence (for example where health economic issues are relevant), or to allow evaluation of an interaction).

The pathway for and potential outcomes from each adaptive analysis is displayed in Figure 3.

Not for IRB Submission

*Figure 3: Adaptive Analyses*

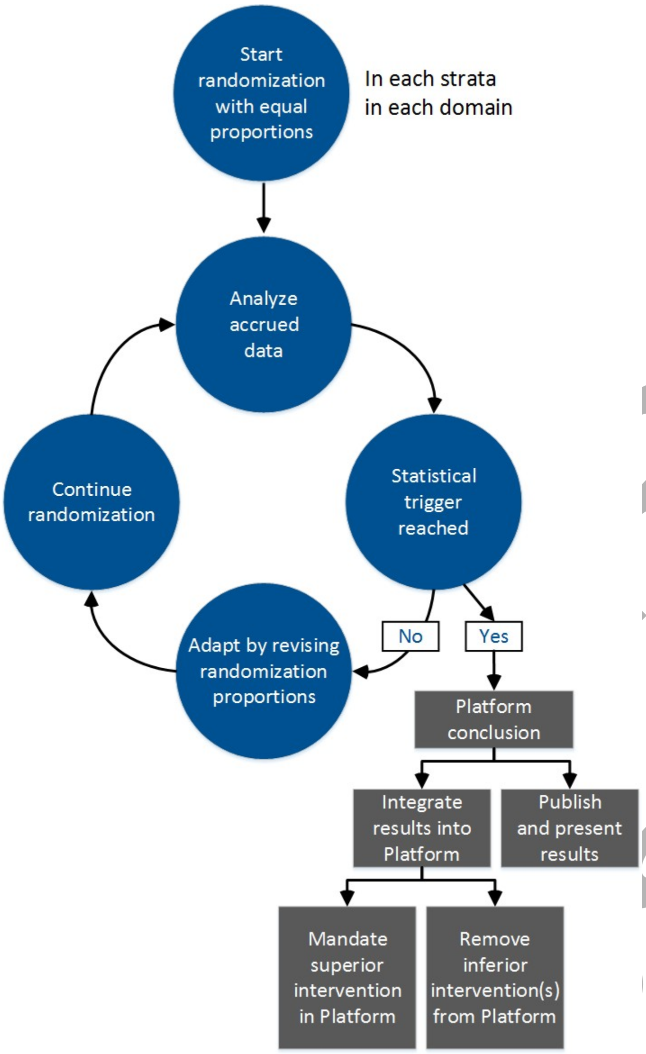

##### 5.3.7.3. *Probability thresholds*

In this REMAP the pre-specified rules are that, at any adaptive analysis, an intervention will be declared "superior," if it is has at least a 0.99 posterior probability of being the best intervention within its domain. An intervention will be declared "inferior" if it has a less than 0.01 probability of being the best intervention within its domain. Intervention equivalence is declared between two factors when there is at least a 0.90 posterior probability of the rate of the primary endpoint falls within a pre-specified delta.

##### 5.3.7.4. *Analysis within and between strata*

The frequent adaptive analyses will evaluate the primary endpoint, *within one or more stratum*. Where specified, the statistical models for each strata will be able to 'borrow' information from adjacent strata leading to the declaration of a Statistical Trigger in one, more, or all strata. The extent to which borrowing occurs is dependent on the pre-specified structure of the model and the degree of statistical congruence of treatment effect between stratum. Where treatment effects are divergent between stratum there is less ‘borrowing’. The capacity to evaluate strata is particularly important for interventions that might plausibly have differential, including opposite, treatment effects in different strata. (Dellinger et al., 2013, Finfer et al., 2004, The Acute Respiratory Distress Syndrome Network, 2000) In traditional trial designs, divergent treatment effects among sub-groups may cancel each other out and this is one plausible explanation for the trials that report no overall difference in outcome. It should be noted that strata can be different for different domains and that strata can be changed over time (in conjunction with amendment of the protocol).

If a Platform Conclusion is reached just within a single stratum, this leads to cessation of randomization within that stratum, while continuing to randomize in other strata. It is acknowledged that a Platform Conclusion in one strata may rely on ‘borrowing’ from adjacent strata and that analysis just within a strata may yield a result that is different. Nevertheless, a Platform Conclusion is still regarded as valid if it relies upon borrowing from adjacent strata and will be reported and published including the extent to which it relies on borrowing.

###### *5.3.7.5 Frequency of adaptive analyses*

Adaptive analyses will occur frequently, with the frequency being approximately proportional to the rate of recruitment, and will be a largely automatic process; the frequency is chosen to balance logistical demands with the goal of learning rapidly from accumulating data. While this process will be overseen by an independent DSMB, the DSMB will not make design decisions unless the trial’s algorithms are no longer acceptable from an ethical, safety, or scientific point of view. The DSMB, in conjunction with the ITSC, having reached a Platform Conclusion, and in deciding to terminate an intervention or domain (in conjunction with a Public Disclosure), may take into account one or more issues such as the value of continuing randomization so as to evaluate additional clinically relevant endpoints or to evaluate potential interactions, as well as take into account the opportunity cost associated with not moving to introduce new domains or interventions.

###### *5.3.7.6 Advantages of adaptive analysis*

The major advantage of this type of analysis approach is that a conclusion is reached when there is sufficient information to support the conclusion, rather than when enrollment reaches a predetermined sample size. This approach allows a result to be obtained as quickly as possible with appropriate sample size. It also avoids indeterminate results by continuing randomization until either superiority, inferiority, or equivalence is concluded. (Barker et al., 2009, Berry, 2012, Connor

Not for IRB Submission

et al., 2013, Meurer et al., 2012, Carey and Winer, 2016, Harrington and Parmigiani, 2016, Park et al., 2016, Rugo et al., 2016) An additional advantage is that dissemination of such results does not interrupt the conduct of the platform. In a single REMAP, there is no need for the “start-and-stop” periods that would typically occur under the alternative approach of multiple separate trials. These “downtime” periods can be quite extensive and carry a number of disadvantages. First, there is a lot of duplicative effort every time a near-identical treatment protocol goes through the appropriate development and approval processes. Second, clinical investigation units must maintain a certain infrastructure, and that infrastructure can be expensive to maintain during periods when participants are not being enrolled or expensive to recreate if the infrastructure degrades. Third, downtime is simply one more contributor to delay in the production of scientific knowledge.
Participants at large benefit from earlier production of knowledge regardless of whether new information demonstrates a therapy is effective or ineffective. Finally, the inevitable start up delay before a trial can “go live” can wipe out any possibility of conducting effective research during time-critical situations such as a pandemic.

###### *5.2.7.7 Substitution of new domains and interventions within the REMAP*

It is intended that the REMAP will be ‘perpetual’. In conjunction with a Platform Conclusion being reached, the ITSC takes responsibility for determining what new questions will be introduced to the REMAP including adding one or more new interventions to a domain or adding one or more new domains. In a REMAP, the sample size is not fixed, rather maximum use is made of the available sample and more questions may be asked for the same monetary investment. (Barker et al., 2009, Berry, 2012, Connor et al., 2013, Meurer et al., 2012, Aikman et al., 2013, Bhatt and Mehta, 2016, Park et al., 2016) The only limit on the duration of a platform trial is the availability of ongoing funding, the availability of new interventions to evaluate, and that the disease continues to be a public health problem. The ITSC responsible for the REMAP will develop appropriate processes for identifying and prioritizing the selection of new interventions and domains that are introduced progressively into the REMAP over time.

How the domains and interventions within a REMAP might evolve over time is depicted in Figure 4.

Not for IRB submission

*Figure 4: REMAP Evolution Over Time*

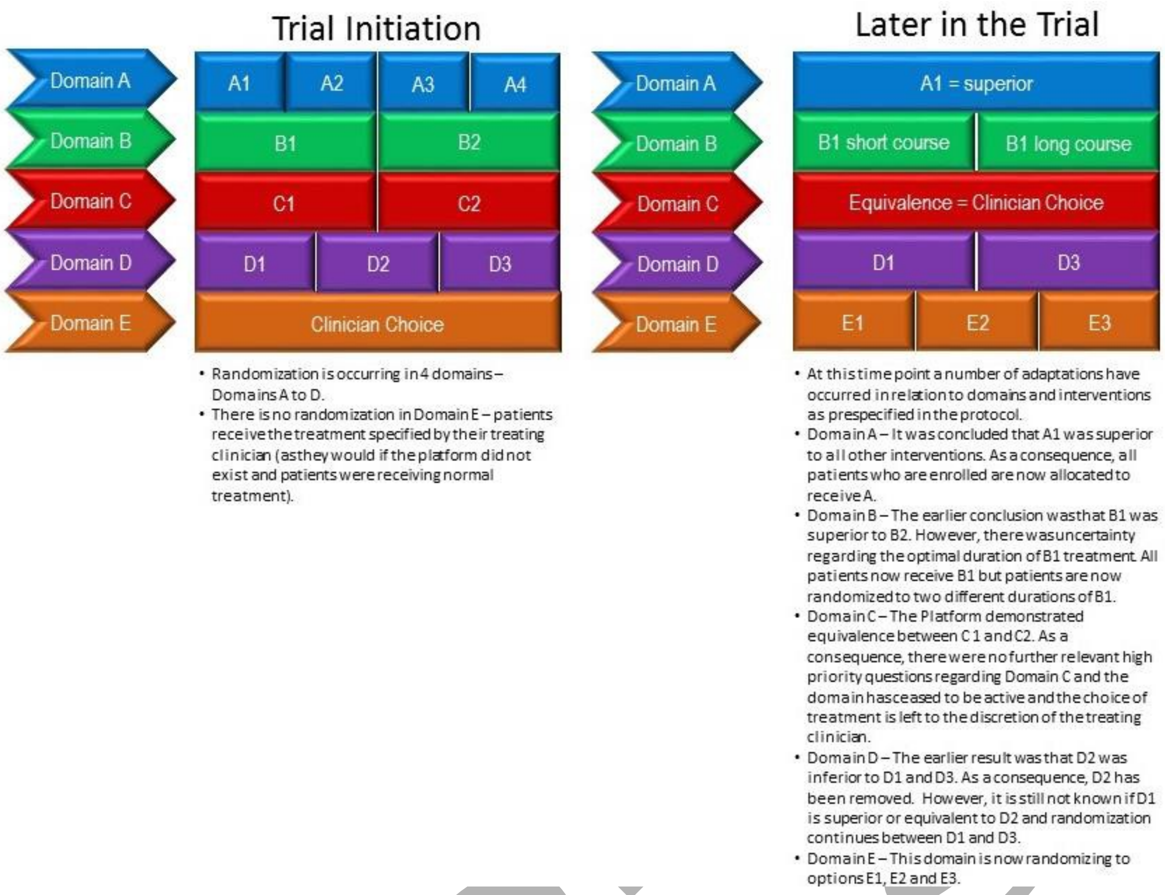

##### 5.3.8. Nesting of the REMAP within a Registry

The REMAP can also be nested within a registry, with the registry recording information (typically a subset of the trial Case Report Form (CRF)) in all participants who met the REMAP entry criteria, or an expanded set of entry criteria, but who, for any reason, were not randomized. Information obtained from eligible but not randomized participants can be useful for evaluating the external validity of results and optimizing recruitment. Evaluation of non-randomized treatments received by all participants, both randomized and non-randomized, can be used to identify the consequences of natural variation in care so as to identify interventions that should be prioritized for evaluation by randomization within the REMAP. (Byrne and Kastrati, 2013) The design features of the trial and the conceptual advantages associated with each design feature are summarized in Table 2.

If a registry component is included the operation of the registry will be specified in a DSA that applies only to the registry aspects of the study.

Not for Submission

##### 5.3.9. Platform

Platform trials simultaneously evaluate multiple potential therapies, where the focus is on finding the best treatment for the disease, rather than precisely characterizing the effect of each intervention in isolation. (Angus, 2015, Berry et al., 2015, Bhatt and Mehta, 2016, Carey and Winer, 2016, Park et al., 2016, Rugo et al., 2016, Harrington and Parmigiani, 2016) Thus the goals of a platform trial are much more aligned with the goals of clinical care than a traditional, narrowly focused phase III RCT of a single agent. All of the component design features of a REMAP have been used previously and have accepted validity. What is innovative and novel, for a REMAP, is the combination of all of these design features within a single platform combined with their use for phase III evaluations and by using embedding to integrate the trial within routine clinical care.

*Table 2: Features of a REMAP that contribute to advantages of the design*

| | Efficient use of information | Safety of trial participants | Avoiding trial down-time | Fusing research with care | Determining optimal disease management | Self-learning healthcare system |
|---|---|---|---|---|---|---|
| Multifactorial | ✓ | | ✓ | ✓ | ✓ | |
| Response Adaptive Randomization | ✓ | ✓ | | ✓ | | ✓ |
| Embedding | | | | ✓ | | ✓ |
| Frequent adaptive analyses | ✓ | ✓ | | | ✓ | ✓ |
| Analysis of strata | ✓ | ✓ | | | ✓ | |
| Evaluation of interaction | | ✓ | | | ✓ | |
| Substitution of new interventions | ✓ | | ✓ | | ✓ | |

### 6. OBJECTIVES

#### *6.1. Primary objective*

The primary objective of this REMAP is, for adult patients with severe CAP who are admitted to an ICU, to identify the effect of a range of interventions to improve outcome as defined by all-cause mortality at 90 days.

##### 6.2. *Secondary objectives*

The secondary objectives are to determine, for adult patients with severe CAP who are admitted to an ICU, the effect of interventions on ICU mortality, ICU length of stay (LOS), hospital LOS, ventilator free days (VFDs) censored at 28 days, organ failure free days (OFFDs) censored at 28 days, other endpoints as indicated for specific domains, and, where feasible or specified in a DSA, survival at 6 months, health related quality of life (HRQoL) assessed after 6 months using the EQ5D and disability assessed after 6 months using the World Health Organization Disability Assessment Schedule (WHODAS).

#### 7. SUMMARY OF TRIAL DESIGN

##### 7.1. *Introduction*

This is a REMAP that aims to test many interventions in a number of domains with the primary outcome being the all-cause mortality at 90 days. Frequent adaptive analyses will be performed to determine if an intervention is superior, inferior, or equivalent to one or more other interventions to which it is being compared, within a domain. A Bayesian analysis method will be used to evaluate superiority, inferiority, or equivalence, as well as to inform the adaptive randomization strategy within each domain. Where it is anticipated that interactions between interventions in different domains may be likely the statistical models will allow evaluation of such interactions. Where the statistical models evaluate such an interaction the models can incorporate the relative likelihood of such interactions, but with possibly low prior probability in cases where it is biologically implausible for interactions to occur. Each intervention within each domain will be evaluated within prospectively defined and mutually exclusive strata (sub-groups) of participants but information from one stratum may be used (via 'borrowing') to contribute to the analysis of the effect of that intervention in other strata. Interventions that are found to be inferior, for a specific stratum, are removed from use in that stratum, and will, typically, be removed from the REMAP allowing new interventions or domains or both to be introduced. An RAR algorithm will be used to preferentially randomize participants to interventions that appear to be performing better. Extensive simulation studies have been performed to define the type I error, power to detect specified differences, and demonstration of equivalence as well as a broad range of operating characteristics. It is planned that further simulation studies will be conducted in conjunction with consideration of the introduction of new interventions or domains or both into the REMAP. The intention-to-treat (ITT) principle will be used for all primary analyses.

Not for IRB Submission

The key structure of the REMAP is outlined in Figure 5.

*Figure 5: REMAP Structure*

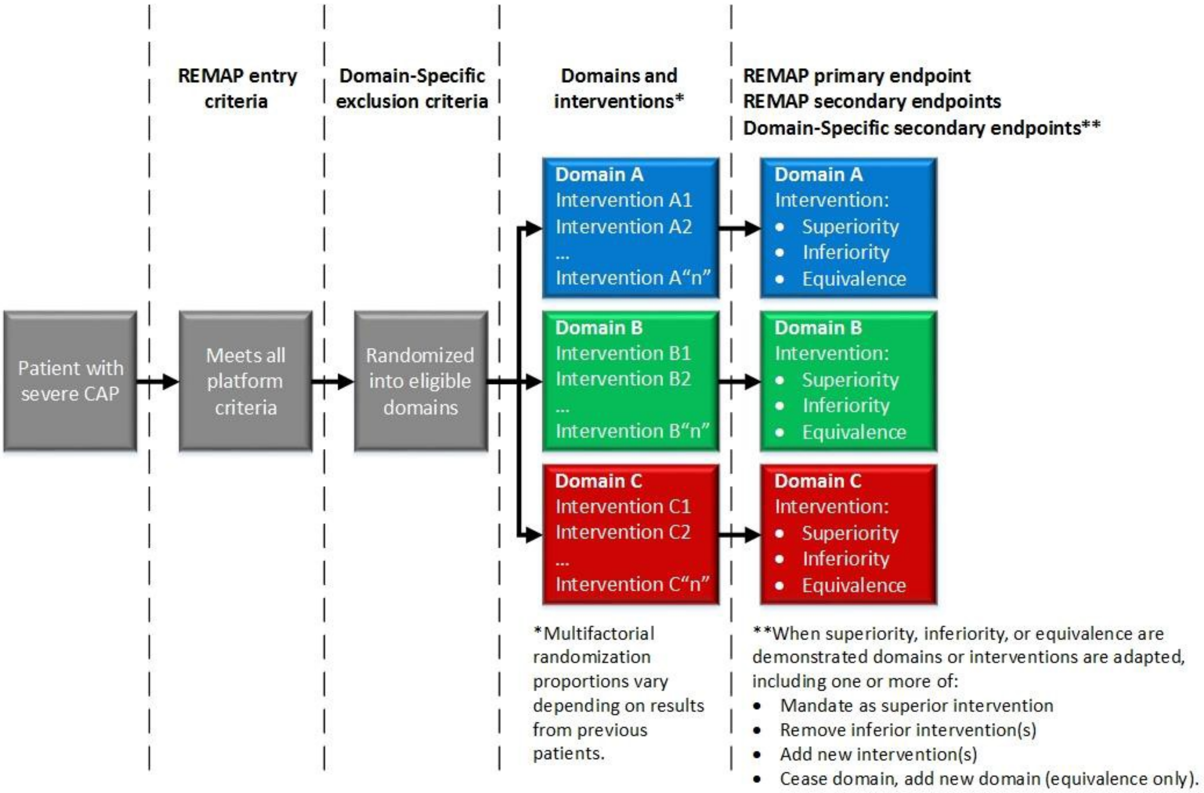

#### 7.2. Nomenclature

A specific set of nomenclature is used to categorize potential treatments evaluated and populations within a platform trial as well as other aspects of the trial design and statistical analysis. A detailed glossary can be found in Section 1.2. Please see the glossary for the definition and explanations for the following terms: domain, intervention, regimen, stratum, state, Statistical Trigger, Platform Conclusion, and Public Disclosure. The following section can only be understood in the context of an understanding of the definition and meaning of these specific terms.

#### 7.3. Study setting and participating regions

The trial will recruit only participants who are admitted to an ICU. An ICU is defined as a location that identifies itself as an ICU (or HDU) and is able to provide at least non-invasive ventilation and continuous administration of vasoactive medications. By agreement with the RMC, the definition of an ICU may include a general ward in which a patient is under the care of an Intensive Care Specialist (Intensivist), but resource limitations prevent the immediate delivery of care occurring in the ICU. It is intended that the trial will be conducted in multiple regions. A region is defined as a country or

Not for Submission

collection of countries with study sites for which a RMC is responsible. The country or countries for which a RMC are responsible, as well as all aspects of trial conduct that are specific to each region, are described in the RSAs.

Participating ICUs will be selected by a RMC based on response to an expression of interest and fulfilling pre-specified criteria including number of beds in the ICU, annual admissions for severe CAP, resources available to support research activities, and track record in conducting investigator-initiated multicenter trials.

The current regions are:

- Europe, with funding from a European Union FP7 grant (FP7-HEALTH-2013-INNOVATION-1, grant number 602525), to support the enrollment of 4000 participants. This funding terminates in 2021.
- Australia and New Zealand. In Australia the project has received funding from a NHMRC Project Grant (APP1101719), to support the enrollment of 2000 participants. This funding terminates in December 2021, although some extension may be feasible. In New Zealand the project has received funding from a HRC Programme Grant (16/631), to support the enrollment of 800 participants. This funding terminates in November 2021.
- Canada. In Canada the project has received funding for a CIHR grant (158584), to support the enrollment of 300 participants. This funding terminates in 2022.

It is intended that additional regions will be added if funding can be secured in other locations. It is desirable that the REMAP is active in as many locations as possible. There is no upper limit to the number of regions and the number of participating sites.

#### *7.4. Eligibility criteria*

The eligibility criteria for the REMAP are applied at two levels. One level is that there are inclusion and exclusion criteria that determine eligibility for randomization within the REMAP. The other level is that, once eligible for inclusion within the REMAP, additional criteria, typically exclusion criteria, are applied that are specific to the level of the domain. A patient is eligible for inclusion within a domain when:

- all REMAP inclusion criteria are present
- none of the REMAP exclusion criteria are present
- Domain-Specific criteria are met

Not for IRB Submission

As such, the key "inclusion criteria" for being eligible for a domain are that the patient is eligible for the REMAP. Criteria for inclusion in the registry, in which patients do not receive any randomized intervention, may be broader than the entry criteria for the REMAP (i.e. it is only a subset of registry eligible patients who are eligible for randomization within the REMAP).

##### 7.4.1. REMAP Inclusion Criteria

In order to be eligible to participate in this trial, a patient must meet both of the following criteria:

1. Adult patient admitted to an ICU for acute severe CAP within 48 hours of hospital admission with
    a. symptoms or signs or both that are consistent with lower respiratory tract infection (for example, acute onset of dyspnea, cough, pleuritic chest pain) AND
    b. Radiological evidence of new onset infiltrate of infective origin (in patients with pre-existing radiological changes, evidence of new infiltrate)
2. Up to 48 hours after ICU admission, receiving organ support with one or more of:
    a. Non-invasive or invasive ventilatory support;
    b. Receiving infusion of vasopressor or inotropes or both

##### 7.4.2. REMAP Exclusion Criteria

A potentially eligible patient who meets any of the following criteria will be excluded from participation in this trial:

1. Healthcare-associated pneumonia:
    a. Prior to this illness, is known to have been an inpatient in any healthcare facility within the last 30 days
    b. Resident of a nursing home or long-term care facility.
2. Death is deemed to be imminent and inevitable during the next 24 hours AND one or more of the patient, substitute decision maker or attending physician are not committed to full active treatment.
3. Previous participation in this REMAP within the last 90 days

##### 7.4.3. Domain-Specific Entry criteria

Each domain may have additional, domain-specific eligibility criteria, typically just exclusion criteria, although a combination of inclusion and exclusion criteria can be specified. Patients who fulfill the Overall REMAP Eligibility Criteria will be assessed for enrollment into all domains that are active at a

Not for IRB Submission

site. A participant enrolled in the trial will receive the number of REMAP-specific interventions equivalent to the number of Domains to which they are enrolled. The additional eligibility criteria that are specific to a domain are provided in each DSA.

Where a participant has an exclusion criterion to one or more interventions within a domain, but there are at least two interventions within that domain to which the participant is eligible the patient will be randomized to receive one of the interventions to which the participant is eligible.

#### 7.5. Interventions

##### 7.5.1. Domain-Specific Information

All information related to the background, rationale, and specification of interventions that will be administered within the trial are located in the DSAs. The minimum number of interventions within a domain is two and the maximum number is limited only by statistical power. Each RMC will select the interventions that will be available within a domain that will be offered to participating sites in that region but the default position is that all interventions that are available and feasible in that region or country should be offered to sites. Individual participating sites will select the interventions within a domain that will be available at their site with the default position being all available interventions. The randomization program will only provide treatment allocations that are permitted at each participating site. This allows interventions that are not necessarily available in all regions, for example because of licensing reasons, to be included within the REMAP. Within the context of comparative effectiveness research, this also allows sites to determine the interventions that are within their usual or reasonable spectrum of care. However, the viability of a domain is dependent on at least one intervention being available in all regions and being available at a substantial majority of participating sites. This level of 'connectedness' is necessary for the validity of the statistical models that are used to analyze trial results.

##### 7.5.2. Treatment allocation and Response Adaptive Randomization

Random allocation of treatment status forms the basis of all evaluations of causal inference. RAR will be used to vary the proportion of participants who are allocated randomly to each available intervention. Randomization is done at the regimen level, where a regimen is a selection of one intervention from each domain. The proportion of participants who receive a specified regimen will be determined by a weighted probability, with that probability being determined by the probability, taking into account all accrued data, of that regimen being the optimal regimen. RAR will result in participants being randomized with higher probability to interventions that are performing better.

Not for IRB Submission

The proportions that are specified by RAR are determined only by analysis of the primary outcome measure in participants who have completed 90 days of follow-up from the time of enrollment. Although outcome may be known before 90 days (death in hospital) the time at which these alternate events occur may be different. By only including participants in the analysis models that determine the RAR proportions potential bias that arises from different events occurring with different patterns of timing within the 90 day follow up period is avoided. The same statistical model will be used to both analyze the results of the REMAP as well as specify the randomization proportions.

RAR weights reflect the probability each particular regimen is the most effective over all possible regimens within each stratum. The probability a regimen is optimal reflects not just the point estimate of difference in outcomes, but also the uncertainty around that estimate. At initiation of a new domain, the proportion of participants allocated to each intervention is balanced (i.e. all interventions have equal proportions). The RAR proportions are then updated at the first adaptive analysis and at all subsequent adaptive analyses. When sample sizes are small, such as at the initiation of a domain, credible (probability) intervals are wide, and therefore randomization proportions remain close to being balanced among all regimens (i.e. randomization weights are weak and allocation remains close to balanced). When a new intervention is added to an existing domain it will commence with balanced randomization and the randomization weights will be updated with each adaptive analysis but will remain weak until sample size for the new intervention accrues.

As the data accrues and sample sizes increase, if the probability an intervention is part of the optimal regimen becomes large, but not large enough to claim superiority, the randomization proportions will be capped. This is done because interventions are provided on an open-label basis and extreme ratios would be at risk of allowing clinicians who recruit participants to draw inference about the effectiveness of individual interventions or regimens.

Some domains may have more than two interventions and it is possible that participant- or site-level characteristics may result in one or more interventions within a domain not being appropriate for an individual participant (for example, known intolerance to one of the interventions or a machine that is necessary to deliver an intervention not being available). Where a participant is unable to receive one or more interventions, but there are still two or more available interventions, random allocation will still be performed using RAR. However, interventions that are not available will be ‘blocked’ and the remaining RAR proportions will be divided by one minus the sum of the unavailable proportions and applied to the available interventions.

Not for IRB Submission

A detailed description of the statistical models and the application of RAR is outlined in the Statistical Analysis Appendix.

##### 7.5.3. Adaptation of Domains and Interventions

Over the lifetime of this REMAP, it is anticipated that new interventions will be added to the starting domains and new domains initiated, including domains that are planned for activation in the event of a pandemic. The addition of interventions within existing domains, and the creation of new domains, will be considered according to a set of priorities and contingencies developed by the ITSC and are dependent on existing or new clinical need and there being sufficient statistical power available within the REMAP. All new interventions and domains will be subject to ethics and regulatory approval prior to initiation.

A domain in which an intervention is identified as being superior and for which there are no new interventions that are appropriate to be introduced will continue as a domain within the REMAP but with all participants allocated to receive the superior intervention. Interventions that are identified as being inferior will be removed from a domain, with or without replacement, as appropriate. If all interventions are identified to have equivalence the ITSC will consider options that include cessation of the domain or continuation of the domain with a smaller delta.

The implementation of adaptations that occurs as a consequence of declaration of a Platform Conclusion may be limited by availability of an intervention in some locations. For example, if a superior intervention was not available (for licensing or site-specific reasons) all inferior options would be removed only at the sites where the superior option is available. Randomization to remaining interventions would likely continue at those sites until the superior intervention is available at those sites.

#### 7.6. Endpoints

The primary outcome for this REMAP will apply to all domains. Secondary outcomes generic to all Domains are provided in this Core Protocol below. Secondary outcomes specific to individual domains are provided in the relevant DSAs. The Primary Endpoint (or the end-point that is used for RAR) may be modified during a pandemic and will be outlined in the Pandemic Appendix.

##### 7.6.1. Primary Endpoint

The primary endpoint for all domains will be all-cause mortality at 90 days.

Not for IRB submission

##### 7.6.2. Secondary Endpoints

A set of generic secondary endpoints will be evaluated in all domains. Additional secondary endpoints may be specified for a domain within the DSA. Some domain-specific secondary endpoints may be specified as Key Domain-Specific Endpoints and will be interpreted in conjunction with the primary endpoint in determining the overall effectiveness of interventions.

The generic secondary endpoints for the trial are:

ICU outcomes:

- ICU mortality censored at 90 days;
- ICU LOS censored at 90 days;
- VFDs censored at 28 days;
- OFFDs censored at 28 days;
- Proportion of intubated participants who receive a tracheostomy censored at 28 days;

Ventilator- and organ failure-free days will be calculated by counting the number of days that the participant is not ventilated or has no organ failure. If a participant dies during the hospitalization during which enrollment occurred, the number of VFDs or OFFDs will be set to zero. If the participant is discharged alive from hospital, the remainder of days censored at 90 days are counted as ventilator- or organ failure-free days.

Hospital outcomes:

- Hospital LOS censored 90 days after enrollment;
- Destination at time of hospital discharge (characterized as home, rehabilitation hospital, nursing home or long-term care facility, or another acute hospital);
- Readmission to the index ICU during the index hospitalization in the 90 days following enrollment;

The index hospital admission is defined as continuing while the participant is admitted to any healthcare facility or level of residence that provides a higher level of care than that corresponding to where the participant was residing prior to the hospital admission. (Huang et al., 2016) This definition is used commonly in ICU trials. Participants who have been and still are admitted to a healthcare facility 90 days after enrollment are coded as being alive.

Not for IRB Submission

Day 90 all-cause mortality will be collected in all regions. Additional outcomes will be collected, where feasible, may be mandated in a DSA or a RSA, may be collected by central trial staff or site staff, and will comprise:

- Survival at 6 months after enrollment (where feasible, refer to relevant regional RSA)
- HRQoL at 6 months after enrollment using the EQ5D-5L (where feasible, refer to relevant regional RSA)
- Disability status measured at 6 months after enrollment using the WHODAS 2.0, 12-item instrument (where feasible, refer to relevant regional RSA)

#### 7.7. *Bias Control*

##### 7.7.1. Randomization

Randomization will be conducted through a password-protected, secure website using a central, computer-based randomization program. Randomization will be at the patient level and occur after data necessary to implement the inclusion and exclusion criteria have been entered into the secure randomization website. The RAR will occur centrally as part of the computerized randomization process. Sites will receive the allocation status and will not be informed of the randomization proportions. Each region will maintain its own computer-based randomization program that is accessed by sites in that region but the RAR proportions will be determined by a SAC and provided monthly to the administrator of each region's randomization program who will update the RAR proportions.

##### 7.7.2. Allocation concealment

Allocation concealment will be maintained by using centralized randomization that is remote from study sites.

##### 7.7.3. Blinding of treatment allocation

The default position within the REMAP is that treatments determined by randomization will be provided on an open-label basis. However, the blinding of treatment status is not precluded within the REMAP. If required, details related to blinding of interventions will be specified in the DSAs.

Not for IRB submission

###### 7.7.4. Blinding of outcome adjudication

The primary outcome of all-cause mortality censored at 90 days is not subject to ascertainment bias. Wherever possible, trial management personnel, who are blinded to allocation status, will conduct any follow up after discharge.

###### 7.7.5. Follow up and missing data

Regional trial management personnel will perform timely validation of data, queries and corrections. Any common patterns of errors found during quality control checks will be fed back to all sites. Data management center study personnel performing site checks will be blind to the study allocation. Missing data will be minimized through a clear and comprehensive data dictionary with online data entry including logical consistency rules. If values necessary for the Bayesian modelling of the primary endpoint and the RAR are missing they may be imputed, using available data. For example, if strata or state is missing, it will be multiply imputed based on the available variables and a prior distribution on the relative prevalence of each strata or state. Values for the primary endpoint will not be imputed. Additional details are provided in the Statistical Analysis Appendix.

##### *7.8. Principles of Statistical Analysis*

###### 7.8.1. Preface

The purpose of this section of the protocol is to introduce and summarize the statistical methods that will be used to analyze data within the REMAP. This section duplicates some of the information provided in the Statistical Analysis Appendix but this section is intended to be accessible to individuals with an understanding of common clinical trial designs and classical frequentist analytical methods but without necessarily having training in Bayesian statistics. Interpretation of this section also requires an understanding of the meaning of specific terms for which definitions are provided in the glossary (see Section 1.2).

A formal description of the adaptive Bayesian data analysis methods fundamental to the REMAP design, which assumes substantial familiarity with Bayesian calculation of posterior distributions conditioned on observed data, is located in the Statistical Analysis Appendix. There is some limited overlap between these two sections of the protocol so that each may serve an appropriate audience as a standalone description of the statistical methods.

Not for IRB submission

##### 7.8.2. Introduction

Within the REMAP, two or more interventions within a domain are evaluated and sequential Bayesian statistical analyses are used over time to incorporate new trial outcome information to determine if an intervention is superior, if one or more interventions are inferior in comparison to all other interventions, or if one or more pairs of interventions are equivalent, with respect to the primary endpoint. Every participant will be assigned a set of interventions, comprising one intervention from each domain for which the participant is eligible. The combination of interventions to which a participant is assigned comprises the regimen and the regimens are the available arms in the trial. Participants will be classified by membership in different populations defined by one or more strata. The unit-of-analysis for a domain is the most granular level, defined by one or more stratum, or a state, within which the treatment effect of interventions within that domain may vary in the statistical model. Participants are also classified by the criteria that determine eligibility for each domain.

Inference in this REMAP is determined by analyses using pre-specified statistical models that incorporate region, country, time periods, age, and disease severity to adjust for heterogeneity of enrolled participants that might influence risk of death. These models incorporate variables that represent each intervention assigned to participants and possible interactions between interventions in different domains. The efficacy of each intervention within a domain may be modeled as not varying in any of the strata, or possibly varying in one or more of the different strata in the REMAP. Where the efficacy of each intervention within a domain is modeled as possibly varying, borrowing between strata is permitted. The unit-of-analysis that will be modeled may comprise the entire population (i.e. no categorization by strata is applied) or may be defined by one or more stratum. The unit-of-analysis and whether borrowing can occur between strata is pre- specified for each domain. At each analysis the current active statistical model (or models) is (are) used, and may include patients who were enrolled when previous versions of the model were being used. The current model is described in an operational document, maintained by the SAC. Unless otherwise specified (see Section 8.12) modifications and implementation of modifications to the model require the approval of the ITSC and do not require a protocol amendment.

Whenever a model hits a predefined threshold for any of superiority, inferiority, or equivalence for an intervention with respect to the primary endpoint, this is termed a Statistical Trigger. At any given adaptive analysis, a Statistical Trigger may be reached for all participants or for one or more stratum and will be reviewed immediately by the DSMB. When a Statistical Trigger is confirmed by the DSMB, based on a thorough review of the data including an evaluation of the proportion of patients

Not for IRB Submission

for whom monitoring of variables that contribute to the model has been completed, and totality of evidence, and where no compelling reason exists not to reach a conclusion (see Section 7.8.9) regarding that question the result that has led to a Statistical Trigger will be specified to be a Platform Conclusion. The declaration of a Platform Conclusion will lead to appropriate modification of the interventions available within that domain and a Public Disclosure of the result. A Statistical Trigger can be considered as a mathematical threshold, whereas a Platform Conclusion is a decision regarding one or more interventions within a domain.

##### 7.8.3. Target populations (strata and states) and implications for evaluation of treatment-by-treatment and treatment-by-strata interactions

###### *Introduction* 7.8.3.1.

In a clinical trial there are many different potential participant-level covariates. A covariate can be a demographic variable that remains unchanged throughout the trial (i.e. age or gender) or a variable representing the severity or course of the disease that can vary over time (i.e. it can be assessed at the time of enrollment and at other times after enrollment during the course of the illness). In this REMAP, there are two special roles for a subset of these potentially time-varying covariates.

First, covariates determined at the time of enrollment that are identified in the design as possibly having differential treatment effect (i.e. interventions may have differential efficacy for the different levels of the covariate) are referred to as strata. Strata are used to define the unit-of-analysis for a domain within a model. Strata are a recognized element in Platform Trials.

Second, within this REMAP, there is interest in studying domains that are relevant for a target population or defined disease state that, while it may be present at the time of enrollment for some participants, may only occur after enrollment for other participants and may never occur for another set of participants. This disease state could be identified by the same covariate that might also have been used to define a strata (but doesn't have to have been). In this regard, the concept of 'state' is used to define participants with characteristics that define a target population that will be evaluated by a domain, analyzed within the REMAP, and for which the characteristics can be present at the time of enrollment or may develop after the time of enrollment. State can also be used to define the unit-of-analysis for a domain within the model.

The appropriate statistical handling of the analysis of patients who become eligible for a domain as a consequence of entering a state, after the time of enrollment, requires the use of models that take into account that the likelihood of entering the state after enrollment may have been influenced by

Not for IRB Submission

the allocation status for other domains that specified the initiation of interventions that commenced at the time prior to entry into the state.

This evolution of Platform Trial design, to include 'state' is a new extension that has not been considered within Platform Trials conducted previously.

##### *Stratum* 7.8.3.2.

A covariate in the REMAP that can be used as a unit-of-analysis within a Bayesian statistical model that allows for the possibility of differential treatment effects for different levels of the variable is referred to as a strata. The covariate is classified into mutually exclusive and exhaustive sets for analysis of treatment effect, as well as for defining separate RAR. The criteria that define a stratum are based on a characteristic that is present at or before the time of enrollment.

The simplest structure for strata is a single dichotomous stratum variable, which divides participants in the REMAP into two stratum. More complex arrangements are possible, such as a single strata variable that is ordinal or two (or more) dichotomous or ordinal strata variables the combination of which defines a single stratum (i.e. there are $2^N$ stratum when there are N dichotomous stratum variables).

The number of strata variables and the number of strata within the REMAP may be varied, depending on the impact of such decisions on statistical power, as determined by simulations. The modeling of strata may assume no differential effect for some domains. This may occur in two ways. Firstly, when the strata structure defines the entry criteria for a domain. Secondly, when two or more stratum are combined within a single unit-of-analysis (i.e. the unit-of-analysis comprises two or more stratum). If the unit-of-analysis comprises less than all available strata the analysis that is performed assumes that treatment effect does not vary between stratum combined within a common unit-of-analysis. The RAR is applied according to the model. So, the RAR applies to the patients that comprise the unit-of-analysis, irrespective of whether the unit-of-analysis comprises a single stratum or two or more stratum.

A strata variable can be set that is maintained as a silent or 'sleeping' strata which becomes active under pre-defined circumstances, such as the occurrence of a pandemic. In this situation, during the inter-pandemic period, all participants are categorized as non-pandemic but, during a pandemic, a distinction is made between patient with proven or suspected pandemic infection and patients in whom pandemic infection is neither proven nor suspected.

Not for IRB Submission

The *a priori* defined strata that are used for determination of results and for RAR may be changed during the life of the REMAP as knowledge is accumulated and, if this occurs, will result in amendment of one or both of the Core Protocol and DSAs. Data from patients enrolled before the change in the strata can be used to determine priors that are incorporated into the model at the outset of the incorporation of the new strata into the model.

###### *7.8.3.3. Treatment-by-strata interactions: borrowing between strata*

Where specified in the statistical model, the treatment effect of an intervention is allowed to vary between different strata. A Bayesian Hierarchical Model (BHM) is used for all treatment-by-strata interactions. In the BHM a hyperprior is used for the differing treatment effects across strata. The standard deviation of the hyperprior, gamma, is a modeling starting estimate for the variation in the magnitude of the difference in treatment effects between strata. By default, the starting estimate of the difference is zero. The gamma parameter influences the extent to which the treatment effect of different interventions is permitted to vary between strata. At the commencement of a model, the gamma parameter must be set, for each domain-strata pair.

In this REMAP, only three options are permitted with respect to specifying the gamma parameter for each domain-strata pair. Firstly, gamma may be set to zero. The effect of this is that treatment effect of an intervention is not permitted to differ between specified strata. The unit-of-analysis is not sub-divided according to the stratum variable. If gamma is set to zero for all strata for a domain, the unit of analysis is all patients randomized in that domain. Secondly, and at the opposite extreme, gamma can be set to infinity. In this situation treatment effect is evaluated separately and independently in each stratum (with no borrowing between stratum). Thirdly, gamma may be set to a defined number between zero and infinity. This parameter value cannot be varied for different domain-strata pairs, a global REMAP value has been selected. This specified value for gamma places a constraint on the variance of the difference in treatment effect in different stratum but permits the model to estimate treatment effect in one stratum by borrowing from other stratum. Borrowing occurs to the extent that it is supported by the accumulated data, but the setting of gamma influences the amount of borrowing and how quickly borrowing is able to occur. The value of gamma that has been chosen has been determined by simulations to achieve a compromise between type I and type II error in baseline scenarios that assume either equivalence or superiority. Where a value for gamma is specified in the model, in this REMAP the value of gamma will be 0.15.

Not for IRB submission

The specification of gamma determines the unit of analysis in the model and the extent of borrowing. For each domain-strata pair, the unit of analysis can be all patients (gamma = zero), each stratum with borrowing (gamma = 0.15), or each stratum separately (gamma = infinity).

The gamma that will be set, and hence the unit-of-analysis, for each domain-strata pair is specified in each DSA.

###### 7.8.3.4. *Analysis set for strata, timing of enrollment and timing of information regarding strata membership*

It has already been specified that the criteria that define a stratum must be present at or before the time of enrollment. In some situations, the information necessary to determine membership of a stratum may become available after the time of enrollment or may be acquired from information derived after enrollment where the understanding of biology of a disease makes it reasonable to assume that the criteria was met at the time of enrollment. This situation might apply to status with respect to a particular pathogen where results of microbiological testing are not available until after enrollment or when the sample that is tested is not collected until after enrollment.

In this situation randomization is permitted within patients where the criteria is suspected or proven at the time of randomization. With regards to possible infection with a specified pathogen, suspected or proven infection at the time of randomization is sufficient to allow an allocation status to be made. For a patient with suspected infection, membership within the strata is defined by the final test results, but a patient who is suspected but is never tested is analyzed as a positive. If a Platform Conclusion is reached for one or more stratum, analyses will also be done on patients with suspected infection who receive the intervention but who turn out to be negative. Whether borrowing between strata is permitted will be specified in the DSA.

###### 7.8.3.5. *State*

A state is a clinical condition of a participant that may change during the course of their treatment. The different states within the REMAP are used to define possible eligibility of the participant for different domains at different times in the trial. A state is a set of mutually exclusive categories, defined by characteristics of a participant, that are dynamic in that they can change for a single participant, at different time-points, during the participant's participation in the REMAP.

The number of state variables and the number of states within the REMAP may be varied, depending on the impact of such decisions on statistical power, as determined by simulations. The same state may be shared by one or more domains but may be different in different domains. The *a priori*

Not for IRB submission

defined states that are used for determination of results and for RAR may be changed during the life of the REMAP as knowledge is accumulated or as domains change and, if this occurs, will result in amendment of one or both of the Core Protocol or DSAs. Data from patients enrolled before the change in the state can be used to determine priors that are incorporated into the model at the outset of the incorporation of the new state into the model.

##### *7.8.3.6. Timing of randomization and revealing of allocation status*

Several different scenarios are recognized that represent different combinations of randomization within a stratum or a state and by the options for the time (at enrollment or later) at which administration of the allocated intervention is commenced.

At the time of enrollment, all participants, are randomized to one intervention in every domain for which the participant is eligible for at enrollment or might become eligible for depending on the progression of the state of their illness (i.e. randomization occurs once and only once at the time of enrollment).

For participants, who at the time of enrollment are eligible for a domain and for which the intervention will be commenced immediately, the allocation status is revealed immediately and the participant then commences treatment according to their allocated intervention. This is referred to as **Randomization with Immediate Reveal and Initiation**.

In circumstances where the participant is eligible for inclusion in the REMAP but is not eligible for a domain at the time of enrollment but might become eligible if the participant's state changes, the participant's allocation status is revealed only if and when the patient enters the state that confers eligibility. This is referred to as **Randomization with Delayed Reveal**.

Another situation applies when eligibility is determined by information that relates to the condition of the patient at the time of initial assessment of eligibility and is relevant to determination of eligibility but is not known until later. In this circumstance, the participant's allocation status can be revealed when the additional information becomes available. Examples of this type of information include the results of microbiological tests and the outcome of a request for consent. Information related to the safety of an intervention in individuals that may change between the time of initial assessment of eligibility and initiation of an intervention may also be reassessed and be used to determine if an allocation status will be revealed. Where initiation of the intervention is deferred pending availability of this additional information, this is referred to as **Randomization with**

Not for IRB Submission

**Deferred Reveal.** It is noted that submission of information regarding microbiological results, consent, or safety information occurs without knowledge of allocation status.

Variation in relation to the timing of revealing and initiation of an intervention has implications to the treatment-by-treatment interactions that are potentially evaluable. Analysis of participants who are enrolled in one or more domains on the basis of Randomization with Immediate Reveal can be conducted within a state, for which membership occurs for at least some participants at the time of enrollment. However, the analysis within this state will also include participants who are enrolled in the same domain on the basis of Randomization with Delayed Reveal with their eligibility for the act of revealing allocation status being defined by progression to the same state at some time-point after enrollment. Participants who are randomized within such a domain, at time of enrollment, but never enter a state that corresponds to eligibility for a domain never have their allocation status revealed and do not contribute to the analysis of treatment effect for interventions in that domain. In this regard, the ITT principle is not violated as the allocation status of such participants is never revealed. The models that are used to provide statistical analysis of the effect of an intervention within a domain that is contained wholly within one state are not able to evaluate interactions with interventions in domains that are defined in different states.

The final scenario to consider involves participants who are enrolled in one or more domains on the basis of Randomization with Deferred Reveal within a stratum. For such participants, their allocation status is revealed at, or close to, the time of deferred initiation of the intervention, when additional information necessary to establish eligibility has become available but relates to information that applies at baseline. Participants in this category are analyzed within baseline stratum in an ITT fashion. As such, the model allows evaluation of interactions with treatments in other domains that share the same stratum. Within such a domain, it can be assumed that there will be some participants who are never eligible to commence receiving the intervention (for example, due to death, or never reaching the defined criteria for the intervention to be commenced) and do not receive the intervention. However, all participants who have an allocation status revealed, even if the intervention is never administered, are analyzed according to and in compliance with the ITT principle.

###### 7.2.3.7 Treatment-by-treatment interactions

Where specified in the statistical model, the treatment effect of an intervention is allowed to vary depending on treatment allocation in another domain (i.e. allow evaluation of treatment-by-treatment interaction). A BHM is used for all treatment-by-treatment interactions. In the BHM, a

Not for IRB Submission

hyperprior is used for the differing treatment-by-treatment interaction effects. The standard deviation of the hyperprior, lambda, is a modeling starting estimate for the variation in the magnitude of the difference in treatment effect dependent on an intervention assignment in another domain. By default, the starting estimate of the difference is zero (i.e. no interaction). The lambda parameter influences the extent to which the treatment effect of different interventions is permitted to vary dependent on intervention assignment in other domains. At the commencement of a model, the lambda parameter must be set, for each domain by domain pair.

In this REMAP, only three options are permitted with respect to specifying the lambda parameter for each domain-domain pair. Firstly, lambda may be set to zero. The effect of this is that there are no treatment-by-treatment interactions being evaluated between interventions in those two domains. Alternatively, lambda may be set to a defined number between zero and infinity. This parameter value cannot be varied for different domain-domain pairs; a global REMAP value has been selected. This specified value for lambda places a constraint on the variance of the difference in treatment-by-treatment interaction. Borrowing occurs to the extent that it is supported by the accumulated data, but the setting of lambda influences the initial amount of borrowing and the degree of borrowing as data accumulates. The value of lambda that has been chosen has been determined by simulations to achieve a compromise between type I and type II error in baseline scenarios that assume either no interactions or moderate interactions exist. Where a value for gamma is specified in the model, in this REMAP the value of gamma will be 0.075. The third choice is to allow no borrowing of the treatment-by-treatment interactions. This is equivalent to selecting a lambda of infinity. This choice would be the most aggressive choice in estimating treatment-by-treatment interactions.

The lambda that will be set for each domain-domain pair is specified in each DSA.

###### 7.8.3.8. *Nested analysis of interventions within a domain*

Within domains in which there are three or more interventions, some interventions may be more likely to have a similar treatment effect. There are several examples of such similarity. For example, the interventions within a domain may comprise a no intervention option and two doses or strategy of administration of the same intervention, or two or more interventions within a domain may belong to the same class of drug than one or more other interventions in that domain.

In situations in which interventions may be more similar than others, the model may nest the more similar interventions within a higher-level intervention category that comprises all the interventions deemed similar. In this situation, and to evaluate the occurrence of a Statistical Trigger, there are two models for analysis. Firstly, all patients receiving the nested interventions, treated as a single

Not for IRB submission

combined intervention, are compared with all other interventions in the domain. Secondly, all interventions are modeled individually. In this analysis, the interventions within a nest are modeled using a BHM incorporating the nesting structure. The BHM has a hyperprior specified for the shrinkage across interventions within the nest. This analysis will compare all interventions within a domain to all other interventions. This BHM analysis is used for the RAR assignments.

Whether nested analysis will be performed and, if so, the membership of category of more similar interventions will be specified in the DSA.

###### 7.8.3.8. *Current strata and states*

The strata are defined, at the time of enrollment, by:

- Shock, defined in 2 categories, present or absent, with present defined as the patient is receiving continuous infusion of intravenous vasopressor or inotrope medications at the time of enrollment
- Influenza defined in two categories, present or absent, based on the results of microbiological tests for influenza. Any patient with suspected influenza who is not tested will be deemed positive. Any patient who is not suspected of having influenza and is not tested will be deemed negative. The availability and interpretation of microbiological tests are likely to change during the REMAP and an operational document will be used to specify how different tests are interpreted. Eligibility for a domain that tests antiviral medications active against influenza will be based on status with respect to influenza being proven or suspected at time of enrollment but it is noted that strata status is defined by the final results of influenza testing which may not be known at time of enrollment and may include analysis of samples collected after enrollment where it is reasonable to presume that the sample reflected influenza status at time of enrollment.
- Pandemic infection defined in two categories, proven or suspected pandemic infection or neither proven nor suspected pandemic infection. This is a ‘sleeping strata’ and will not be active before or after a pandemic but may be activated during a pandemic. The decision to activate a pandemic infection strata is specified in the Pandemic Appendix to the Core Protocol.

The default states are defined by the occurrence of:

- Hypoxemia, defined in 3 categories, comprising participants who are not receiving invasive mechanical ventilation; participants who are receiving invasive mechanical ventilation and

Not for IRB Submission

have a ratio of arterial partial pressure of oxygen to fractional inspired concentration of oxygen (P:F ratio) of ≥ 200 mmHg or are receiving invasive mechanical ventilation with the Positive End-Expiratory Pressure (PEEP) set to less than 5 cm of water (irrespective of the P:F ratio); and participants who are receiving invasive mechanical ventilation with a PEEP of 5 cm of water or more and have a P:F ratio of <200 mmHg.

The domains to which each strata or state applies, the unit-of-analysis (which determines which if any treatment-by-strata interactions are evaluated in the model), the relationship between the timing of domain eligibility and the revealing of allocation status, whether nested analysis will occur, and what treatment-by-treatment interactions will be evaluated are specified in each DSA.

###### 7.8.3.10. *Pre-specified subgroup analysis after achievement of a Platform Conclusion*

Following the achievement of a Platform Conclusion it is permissible for additional sub-group analyses to be conducted. The variables that specify such sub-groups are outlined *a priori* in each DSA. These variables are different to those that define strata or states in the model and are not used in determination of a Statistical Trigger or RAR for that domain. In a domain in which the unit-of- analysis comprises two or more stratum, additional sub-group analyses can be conducted for variables that do specify stratum that have been combined to determine the unit-of-analysis.

All such analyses will only be conducted following the determination of a Platform Conclusion and, although reported, such analyses are always regarded as preliminary. Following a Platform Conclusion, the results of a pre-specified subgroup analysis may be used to make changes to the model and, where appropriate and to an appropriate degree, data derived from the REMAP can be used to set the prior distribution at the commencement of the new model.

###### 7.8.4. Bayesian Statistical modeling

Inferences in this trial are based on a Bayesian statistical model, that will calculate the probability of superiority, inferiority, and equivalence of the interventions (known as a posterior probability distribution) within a unit-of-analysis that is defined by one or more stratum, taking into account the evidence accumulated during the trial (based on data on the outcomes of participants) and on assumed prior knowledge (known as a prior distribution). For the evaluation of the main effects of interventions within a domain (and evaluation of regimens) the default design assumes that parameters in the model have uninformative prior distributions at the first adaptive analysis. This means that any subsequent Platform Conclusion is not capable of being influenced by any discretionary choice regarding the pre-trial choice of prior distribution (i.e. it is the most

Not for IRB submission

conservative approach, making no assumptions regarding the prior distribution). At each subsequent adaptive analysis, the prior distribution is determined by all accumulated data available at the time of the adaptive analysis. The Bayesian approach is seen as continually updating the distribution of the model parameters.

It is not precluded that, under certain circumstances, such as during a pandemic and where there was strong prior evidence along with an ethical imperative to evaluate a particular choice of therapy, that the design could allow an informative prior to be used for the analysis of results from the trial. It may also be permitted to use an informative prior when data that is incorporated in the informative prior is derived from patients already randomized within this REMAP. If informative priors are used this will be specified in the relevant DSA.

The study design can use informed priors to guide some elements of the design, such as for the evaluation of interaction terms, and will be described in the Statistical Analysis Appendix. As outlined above, gamma will be set to allow and influence the evaluation of treatment-by-strata interactions and lambda will be set to allow and influence the evaluation of treatment-by-treatment interactions.

This method of statistical analysis differs from conventional (frequentist) trials. Frequentist statistics calculate the probability of seeing patterns in the data from a trial if a hypothesis is true (including patterns not observed). This approach relies on assumptions about frequency distributions of trial results that would arise if the same trial were repeated *ad infinitum*. Thus, it requires specific sample sizes, which in turn requires pre-experiment assumptions regarding plausible effect sizes and outcome rates. Although many clinicians are comfortable with this approach, the pre-trial assumptions are frequently incorrect, and the design lacks the flexibility either to easily address the complex questions more reflective of clinical practice or to make mid-trial corrections when the pre- trial assumptions are wrong without concern that the integrity of the final analysis is violated. To allow increased flexibility and yet still generate robust statistical inferences, REMAP relies on an overarching Bayesian, rather than frequentist, framework for statistical inference.

A Bayesian approach calculates the probability a hypothesis is true, given the observed data and, optionally, prior information and beliefs. The advantage of this approach is that, as more data are accrued, the probability can be continually updated (the updated probability is called the posterior probability). In this trial, frequent adaptive analyses will be performed, creating a very complicated sample space, and hence the Bayesian approach is a very natural one for these adaptive designs. The characterization of the risk of false positive error, or power, are done through Monte Carlo trial

Not for IRB Submission

simulation. In contrast to frequentist confidence intervals which have awkward direct interpretation, Bayesian analyses return probability estimates that are directly interpretable as probabilities that statements are true (like the probability that one intervention is superior to another).

A number of variables are incorporated into the statistical model so as to provide ‘adjustment’. The variables for which such adjustment will be made will be the country in which a participant is treated, changes in outcome that occur over time (era), stratum and state at enrollment (shock and hypoxemia as measures of severity of illness), and age.

The main effect in the model is the treatment effect of each intervention. Each stratum, combination of stratum, or state (where eligibility is defined by a state) is analyzed separately but the model captures the commonalities across such sub-groups. Additionally, and where specified, the statistical model allows evidence relating to the effectiveness of an intervention in one stratum to contribute (via ‘borrowing’) to the estimation of the posterior probability in other strata, but this only occurs to the extent that treatment effect is similar in different strata.

When a Platform Conclusion is achieved, the results derived from the model, including any contribution from borrowing, will be reported. It is acknowledged that the estimate of treatment effect for a stratum may be contributed to by borrowing from adjacent strata but the results from the strata that have contributed to borrowing will not be reported. The results of these analyses are used to achieve the primary objective of the trial which is to determine the effectiveness of interventions and, where specified, the extent to which that effectiveness varies between strata (intervention-stratum interaction). Additionally, but only where specified *a priori*, the model is able to estimate the effectiveness of an intervention in one domain contingent on the presence of an intervention in another domain (treatment-by-treatment interaction). Although the model can identify an optimal regimen this is not the primary objective of the trial.

Greater detail of the methods within the Bayesian model to be applied in this REMAP are provided in the Statistical Analysis Appendix. The adaptive analyses will use data submitted from participating sites to their regional database. Each provider of regional data management will provide regular updates of data to the SAC for utilization in the adaptive analyses. The frequency of adaptive analyses will occur approximately monthly, unless the amount of data in a month is deemed insufficient. The timely provision of outcome data from participating sites is critically important to the conduct of frequent adaptive analyses.

Not for IRB Submission

##### 7.8.5. Statistical Handling of Ineligible Participants

The goal of this REMAP is to enroll as wide a participant population as possible. Because of this and the desire to explore multifactorial regimens it will not be uncommon that a participant will be ineligible for single interventions or entire domains, or interventions may be temporarily unavailable for use. In this section we present the details for how this REMAP deals with these possible circumstances.

If an intervention is unavailable at the time of randomization due to site restrictions (for example, exhausted supply or unavailable machinery) then the participant will be randomized to all remaining interventions and this participant will be included in the primary analysis set as though they were randomized unrestricted to their assigned intervention.

If a participant is ineligible for an entire domain then that participant will not be randomized to an intervention from that domain. The participant will be randomized to a regimen from all remaining domains. As long as the participant is randomized within at least one domain they will be included in the primary analysis. For the ineligible domain the participant will be assigned a covariate for that domain reflecting the ineligibility for the domain. This allows the model to learn about the relative efficacy of the remaining interventions in the domains in which the participant has been randomized. If there is a domain with only two interventions and participant is ineligible for one of the two then the participant will be treated as though they are ineligible for the domain. If there is a domain with more than two interventions but a participant is ineligible for all but one then the participant will be deemed ineligible for the domain. If a participant is only eligible for one intervention within a domain the allocation process may still provide a recommendation that the only available intervention should be provided to the participant (but this is so as to reinforce trial processes associated with successful embedding and such patients will not be included within any analysis of the relevant domain).

If there is a domain with more than two interventions and the participant is ineligible for at least one due to a patient-level factor (for example known intolerance to an intervention), but eligible for at least two, then the participant will be randomized among those interventions that the participant is eligible to receive. The participant will have their assignment included in the primary Bayesian model with an appropriate covariate identifying their ineligibility status that takes into account that a patient-level factor that determines partial eligibility could be associated independently with outcome. The impact of participants with partial eligibility will be taken into consideration by the

Not for IRB submission

DSMB at the time of consideration of whether a Platform Decision is appropriate following a Statistical Trigger.

###### 7.8.6. Intervention Superiority Statistical Trigger

At any adaptive analysis, if a single intervention has at least a 0.99 posterior probability of being a member of the optimal regimen, for that unit-of-analysis, then that intervention will be deemed as being superior to all other interventions in that domain in that target population. This Statistical Trigger may also be applied for a state that defines the target population for a domain.

###### 7.8.7. Intervention Inferiority Statistical Trigger

At any adaptive analysis, if a single intervention has less than a 0.01 posterior probability of being a member of the optimal regimen, for a unit-of-analysis, then that intervention will be deemed as being inferior for that target population. If superiority and inferiority were to be discovered simultaneously (for example when there are two interventions), the result will be interpreted as demonstrating superiority. This Statistical Trigger may also be applied for a state that defines the target population for a domain.

###### 7.8.8. Intervention Equivalence Statistical Trigger

If two interventions within a domain, for a unit-of-analysis, have at least a 0.90 probability of being within a pre-specified delta for the primary endpoint then these interventions will be deemed as being equivalent. The size of the pre-specified odds ratio delta is 0.20, meaning equivalence is reached with at least a 90% probability of neither intervention increasing the odds ratio of mortality by more than 0.20. An odds ratio delta of 0.2 has been chosen on the basis that it is consistent with guidance from the Food and Drug Administration (FDA) (U.S. Department of Health and Human Services, 2016) and the European Medicines Agency (EMA) (European Medicines Agency, 2005), as well as discussed in academic literature, and the magnitude of treatment effect that has been specified in published superiority trials that enroll patients who are critically ill (Aberegg et al., 2010, Ware and Antman, 1997, European Medicines Agency, 2005, U.S. Department of Health and Human Services, 2016). A measure of relative treatment effect (odds ratio) is specified, rather than an absolute difference in treatment effect. This choice is made because it is reasonable to expect the mortality rates to vary between strata, and the relative effect is a more robust analysis method across these differences.

Not for IRB Submission

In a domain with two interventions equivalence is evaluated between the single pair of interventions. In a domain with more than two interventions, equivalence is evaluated for every possible pairwise comparison.

A DSA may define levels of delta for equivalence that are different from the default delta. This includes the possibilities of specifying a delta that may be asymmetrical for some or all pair-wise comparisons or both. The DSA will set out the rationale for any variation in delta and may include, but are not limited to, cost or burden.

This Statistical Trigger for equivalence may also be applied for a state that defines the target population for a domain.

##### 7.8.9. Action when a Statistical Trigger is achieved

###### *Introduction* 7.8.9.1.

If a Statistical Trigger is achieved this will be communicated by the SAC to the DSMB. Subject to the DSMB confirming that a Statistical Trigger has been reached validly, the DSMB will oversee a range of actions, as follows.

###### 7.8.9.2. *Actions following Statistical Trigger for superiority*

If an intervention triggers a threshold for superiority and the DSMB declares this as a Platform Conclusion, the intervention is deemed as being superior. At that point randomization to all other remaining interventions in the domain in that unit-of-analysis will be halted at sites at which the superior intervention is available (randomization to the non-superior interventions may continue at sites at which the superior intervention is not available pending its availability). The result will be communicated to the ITSC who will take responsibility to undertake Public Disclosure as soon as practicable with the dissemination of the research result via presentation or publication or both.

Within the REMAP and at sites with access to the superior intervention, all participants will be allocated to the superior intervention (while still being randomized to interventions from the other domains). In this regard the domain remains active with what can be considered as 100% RAR to the superior intervention, pending the addition of any new interventions to be evaluated against the current superior intervention. It is also possible that a superior intervention will be retained but subject to further evaluation, by randomization, to refine the optimal characteristics of the superior intervention (for example duration of therapy or optimal dose).

Not for IRB Submission

###### 7.8.9.3. *Actions following Statistical Trigger for inferiority*

If the trial triggers a threshold for inferiority and the DSMB declares this as a Platform Conclusion, the intervention is deemed as being inferior. At that point the intervention will not be randomized to any more participants in that unit-of-analysis. The result will be communicated to the ITSC who will take responsibility to undertake Public Disclosure as soon as practicable with the dissemination of the research result via presentation or publication or both.

Where a Platform Conclusion is reached for superiority or inferiority, the DSMB may recommend that Public Disclosure should be delayed until additional results are available, so as to allow further recruitment to evaluate interactions between interventions in different domains or for other clinically or statistically valid reasons. However, declaration of a Platform Conclusion will always result in the removal of inferior interventions from a domain and that all eligible participants within the REMAP receive a superior intervention.

###### 7.8.9.4. *Actions following Statistical Trigger for equivalence*

If a Statistical Trigger arises because one or more pairs of interventions are deemed as being equivalent within a unit-of-analysis, this will be communicated to the ITSC by the DSMB. The ITSC in conjunction with the DSMB may undertake additional analyses, for example, of clinically relevant secondary endpoints.

The approach to a Statistical Trigger for equivalence is different depending on the number of interventions within a domain.

For domains with only two interventions a valid Statistical Trigger for equivalence will be reported as a Platform Conclusion. With respect to the adaptation of the domain, the following actions are possible:

- Removal of the domain from the Platform
- Switching the allocation status to deterministically assign one of the Interventions, for example the less burdensome or less expensive intervention
- No change to the interventions within the domain with continuation of RAR. This could be to further evaluate secondary endpoints, a smaller delta of equivalence, or interest in interactions with other Interventions. Such changes would require amendment to the DSA.

Not for IRB submission

Factors that should be taken into account by the DSMB and the ITSC include the results of the primary analysis, analysis of clinically relevant secondary end-points, the possibility of treatment-by- treatment interactions, the relative burden and cost of the two interventions, the clinical interpretation of the adequacy of the delta, and the possibility that ongoing randomization with a smaller delta might also allow a Statistical Trigger for superiority (with a small effect size).

The options following a Statistical Trigger for a pair of Interventions in a Domain with three or more Interventions are more complex. Within a domain with three or more interventions the information provided by the DSMB to the ITSC may include specification of the ordinal rank of the equivalent interventions within the domain. With respect to reporting of Platform Conclusions and adaptations of the domain the following actions are possible:

- A pair of equivalent interventions may be compressed into a single group for the purposes of ongoing analysis. Both interventions continue to be interventions that are available within the domain for allocation, but the primary analysis considers the effect of the two interventions as a single group, where a balanced randomization will be assigned to each of the intervention pair within this compressed group. Secondary analyses can continue to be conducted to determine if equivalence is maintained with the possibility of the intervention being restored as individual interventions if results no longer support equivalence. It is acknowledged that re-analysis of the domain immediately following compression of one (or more) pairs of equivalent interventions may result in the occurrence of other Statistical Triggers (e.g. a compressed pair may be superior or inferior to all remaining interventions). Any statistical Trigger that results from compression of one or more pairs will be responded to as outlined in this section with reporting of the cascade of Statistical Triggers. Compression of a pair of interventions can occur with or without reporting of a Platform Conclusion.
- Removal of one of the pair of equivalent interventions from the domain, for example the more burdensome or more expensive intervention, which will result in a reporting of a Platform Conclusion.
- No change to the interventions within the domain with continuation of RAR. This could be to further evaluate secondary endpoints, a smaller delta of equivalence, or interest in interactions with other interventions. Such changes would require amendment to the DSA. This could occur with or without reporting a Platform Conclusion.

Not for IRB Submission

Factors that should be taken into account by the DSMB and the ITSC include the results of the primary analysis, analysis of clinically relevant secondary end-points, the possibility of treatment-by- treatment interactions, the relative burden and cost of the two interventions, the clinical interpretation of the adequacy of the delta, the possibility that ongoing randomization with a smaller delta might also allow a Statistical Trigger for superiority (with a small effect size) and the ordinal position of the equivalent pair within the domain.

In a domain that comprises three or more interventions, but in which two or more interventions are analyzed in a nested manner, the nested group may be combined for analyses of equivalence. Where compression converts a domain with three or more interventions into a domain with two interventions (and data continues to support equivalence of the compressed interventions) such a domain will be regarded as a two-intervention domain for the purposes of evaluation of Statistical Triggers for superiority, inferiority, and equivalence.

If a Platform Conclusion is reached, the ITSC will take responsibility to undertake Public Disclosure as soon as practicable with the dissemination of the research result via presentation or publication or both. There is no automated adaptation when equivalence is deemed to have occurred. Where appropriate each DSWG will produce an operational document, that is publicly accessible, that considers a range of plausible scenarios and provides guidance as to the actions that should occur in the event of a Statistical Trigger for equivalence for different pairs of interventions. If any of these documents are updated, previous versions will be archived but continue to be publicly accessible.

##### 7.8.10. Analysis set for reporting

The primary analysis set that will be used for reporting a Public Disclosure will comprise all participants who are analyzed at the time the adaptive analysis results in the occurrence of a Statistical Trigger. As such, there will be some participants who have been randomized but are not included within this analysis, either because participants have not yet completed 90 days of follow up or because data for a participant who has completed 90 days of follow up has not yet been submitted. At the time of Public Disclosure, a secondary analysis will also be reported that comprises all participants who are evaluable through to the point at which there was cessation of randomization to the relevant comparator arms.

##### 7.8.11. Simulations and statistical power

The design of the trial, at initiation, and in conjunction with the planning of the introduction of new interventions within a domain or of new domains, will be informed by the conduct of extensive

Not for IRB Submission

simulations using standard Monte Carlo methods. Simulations will be updated whenever a new intervention is added within a domain or whenever a new domain is added to the REMAP. However, simulations will not be updated when an intervention is removed from a domain because of the declaration of a Platform Conclusion that the intervention is inferior. These simulations will evaluate the impact of a range of plausible scenarios on the statistical properties of the trial.

Existing simulations indicate that when a single intervention in a domain with two interventions is beneficial, with a constant benefit for all participants, the power to be determined superior to the complement intervention as a function of its odds-ratio benefit is greater than 90% when there is at least a 25% odds-ratio decrease in the probability of mortality for the funded sample size of 6800 participants. The timing of these conclusions of superiority have a median time of less than 2000 participants. The probability that an intervention will be deemed superior to a complementary intervention when in truth the two are equal (a type I error) is typically less than 2.5%.

The results of detailed simulations of current domains is located in the Simulations Appendix which is maintained as an operational document that is publicly accessible and updated as required.

###### 7.8.12. Updating model after monitoring

If any variable that contributes to the model is identified to be inaccurate at a monitoring visit, the data will be corrected and utilized for the next interim analysis. Any change to a previous statistical trigger will be reviewed by the DSMB to determine the implications. The DSMB will advise the ITSC if there is any material change in a Platform Conclusion which, if published, will be reported to the journal as an erratum.

##### 7.9. Co-enrollment with other trials

Co-enrollment of participants in other research studies, including interventional trials, is strongly encouraged. The principle is that co-enrollment should always occur and is only not permitted when there is a clear threat to the validity of either study or it would materially influence the risk to participants. Decisions regarding co-enrollment with other trials will be made on a trial-by-trial basis. Where a potentially co-enrolling trial is being conducted in more than one region in which the REMAP is being conducted the decision regarding co-enrollment will lie with the ITSC. Where a potentially co-enrolling trial is being conducted only in one region in which the REMAP is being conducted the decision regarding co-enrollment will lie with the RMC. In all circumstances the ITSC and RMCs should liaise regarding decisions about co-enrollment. Decisions regarding co-enrollment

Not for IRB Submission

with other trials will be distributed to participating sites as an operational document and will not require or involve amendment of this protocol.

#### 7.10. Cooperation between the REMAP and other trials with overlapping populations or interventions

During the life-time of the REMAP it is likely that there will be many other clinical trials that will have inclusion and exclusion criteria which would include participants who are eligible for this REMAP. This would include, obviously, trials with a primary interest in patients with CAP, but could also include patients with the Acute Respiratory Distress Syndrome (ARDS) and patients with severe sepsis or septic shock. Such trials will likely test a range of interventions, some of which may also be intervention options within this REMAP. This REMAP seeks to cooperate and coordinate maximally with other trials. Examples of such cooperation and coordination would include, but not be limited to, utilization of REMAP infrastructure for screening and recruitment to other trials, sharing of data collected by the REMAP, and sharing of allocation status so as to allow incorporation of allocation status within analysis models.

Where another trial is evaluating an intervention that is also included within this REMAP each site (or region) would need to establish rules that determine circumstances in which each trial has preference for recruitment. Where another trial and this REMAP are evaluating different interventions the extent to which cooperation is possible will also be determined by the extent to which the interventions are compatible, i.e. capable of having their effect evaluated independently within each trial.

##### 7.11. Registry of non-randomized patients

In some locations, the REMAP may be nested within a registry. Where this occurs the operation of the registry, including eligibility criteria, ethical issues, and variables that will be collected, will be described in a separate Registry Appendix.

##### 7.12. Criteria for termination of the trial

This trial is designed as a platform, allowing for continued research in patients with CAP admitted to an ICU. The platform allows for the study to be perpetual, with multiple different domains that can be evaluated at any one time, and over time. Frequent adaptive analyses are performed to determine whether the interventions under evaluation are still eligible for further testing or randomization should be stopped due to demonstrated inferiority, superiority or equivalence.

Not for IRB submission

It is anticipated that after inclusion of the initially planned sample size, the study would continue to include additional participants and test additional domains and/or interventions until one of the following occurs:

- CAP is no longer deemed to be a public health problem
- The effectiveness and/or cost-effectiveness of all interventions are known and there are no new plausible interventions to test

Should the whole study be stopped, the end of trial is the date of the last scheduled follow up for any participant.

### 8. TRIAL CONDUCT

#### *8.1. Site time-lines*

##### 8.1.1. Initiation of participation at a site

A range of options are available for the sequence of activities by which a site commences participation. The following outlines the default sequence of participation. The first level of participation is termed 'observational only'. During this stage eligible participants will be identified, preferably using a process of embedding with recognition by clinical staff and registration on the study website as soon as eligibility is recognized. Treatment decisions will be made by that site's clinical staff, and observational data using the study CRF or a sub-set of the CRF will be collected. The next level of participation is termed 'single domain'. During this time period, eligible participants are identified and randomized, but only within a single domain. The next level of participation is termed 'multiple domains' although this would typically include only the addition of a single domain at any one time-point with staggered introduction of additional domains. Decisions about transition through levels would be made by the site, in conjunction with the RCC, and would be influenced by factors including speed and accuracy of identification of eligible participants, accuracy of information provided at time of randomization, compliance with allocated treatment status, and timeliness of reporting of outcome variables that are used to determine RAR algorithms. It is also permissible to commence the trial with multiple domains being active at initiation.

##### 8.1.1. Vanguard sites

In each region or at the initiation of a new domain or both, the trial may consider commencing with only a small number of vanguard sites. The purpose of commencing the trial at vanguard sites is to learn about the effectiveness of different options for trial processes so that this information about

Not for IRB Submission

the most effective trial processes can be shared with subsequent non-vanguard sites. If a site is acting as a vanguard site this will be specified in any application for ethical approval at that site.

#### 8.2. Summary of time-lines for recruited participants

A summary of the study and follow up schedule is outlined in Figure 6.

*Figure 6: Study Procedures*

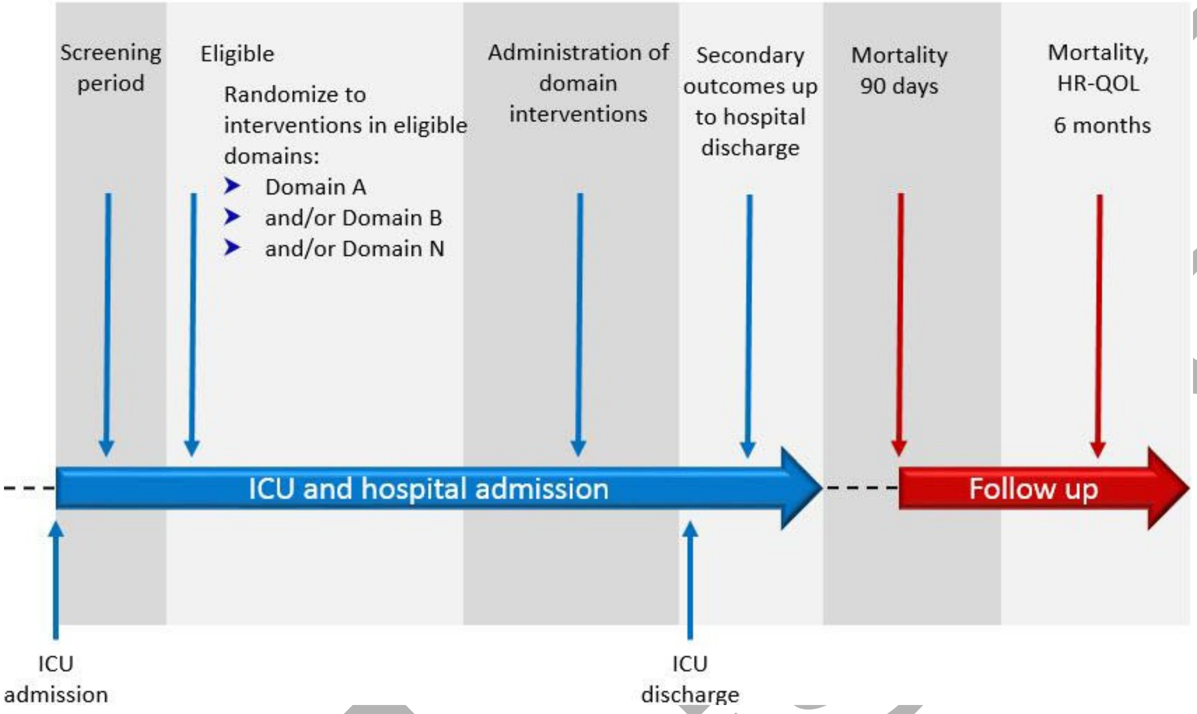

#### 8.3. Recruitment of participants including embedding

##### 8.3.1. Embedding

The trial is designed to substitute allocation of treatment status by randomization where otherwise a treatment decision would have been made by clinical staff (where it is clinically and ethically appropriate to do so), and for this to occur at the time that the treatment decision would have otherwise been made. It is not essential that embedding is used to achieve recruitment and randomization but it is preferable and it is encouraged that participating sites work in conjunction with the trial team to achieve embedding wherever possible and as soon as possible.

The success of embedding can be evaluated by the proportion of eligible participants who are recruited and randomized, that recruitment and randomization occurs as soon as possible after eligibility occurs, and that there is compliance with the allocated intervention. Successful embedding will enhance the internal and external validity of the results generated by the trial.

Each site, taking into account its own clinical work practices, will be asked to develop internal processes that will be used to achieve successful embedding. Wherever possible the RCC will advise and assist sites to achieve successful embedding. In brief, each participating site will identify their ICU admission procedures that occur with each new patient and then align these procedures to facilitate assessment of eligibility by clinical staff who provide routine care for each patient. This can be achieved through several methods including checklists on electronic Clinical Information Systems (eCIS).

##### 8.3.2. Participant recruitment procedures at participating units

Once screened and identified as eligible the clinical staff (medical or nursing) or research staff will randomize the participant. Standard Operating Procedures (SOPs) will be developed to guide staff who undertake randomization. For example, in ICUs with an eCIS, an integrated website link may be used to allow direct access to the trial randomization webpage and, where possible, provide a summary (or direct population from the eCIS) of information that is required to be entered into the randomization web-site. To complement this system the research staff in each ICU will review patients admitted each day to assess the suitability of patients deemed not eligible out of hours, either because they were missed on screening or because the clinical situation has changed.

#### *8.4. Treatment allocation*

An eligible participant will receive a treatment allocation that is determined for all domains for which the participant is eligible to receive at least one of the available interventions. The management of the randomization process in each region is specified in each RSA. Information related to RAR is presented in the Interventions section of the Trial Design (Section 7.5.2) and in the Statistical Analysis Appendix. As noted elsewhere, all randomized allocation will be determined at the time of initial enrollment, but allocation status will not be made known for domains that operate using Randomization with Delayed Reveal (see Section 7.8.3.4). If the participants clinical condition changes and enters the state that confers eligibility this information will be provided to the randomization web-site and the allocation status will be revealed to the site.

Not for IRB submission

#### 8.5. *Delivery of interventions*

##### 8.5.1. Treatment allocation and protocol adherence at participating units

In conjunction with participating sites, trial management staff will develop generic and site-specific documents that outline processes for implementation of and facilitate adherence with participant's allocated treatment status. Wherever possible these will seek to integrate trial processes with existing routine treatment processes to allow seamless adoption of the allocated treatments. For example, after randomization the clinical staff will be directed to use a pre-populated order sheet, necessary for the treating clinicians to authorize and for a bedside nursing staff to follow allocated treatment processes for that individual participant. It is intended that this process will not only reduce the complexity of ordering the study treatments but also reduce errors and increase adherence to the allocated protocol.

With respect to blinding, the default position within the REMAP is that treatments determined by randomization will be provided on an open-label basis. Where interventions are conducted on an open-label basis, all members of the ITSC and all other staff associated with a RCC of the trial will remain blinded until a Platform Conclusion is reported by the DSMB. Although the default is the provision of open-label treatments the blinding of treatment status is not precluded within the REMAP. Whether interventions are open-label or blinded will be specified in DSAs.

#### 8.6. *Unblinding of allocation status*

Unblinding of any blinded treatment by site research staff or the treating clinician should only occur only in when it is deemed that knowledge of the actual treatment is essential for further management of the participant. A system for emergency unblinding will be provided in the DSA of any domain that includes interventions that are administered in a blinded fashion. Any unblinding process will ensure that the investigator can directly and rapidly unblind in an emergency situation. All unblindings and reasons as they occur will be documented in the CRF. Unblinding should not necessarily be a reason for study drug discontinuation.

#### 8.7. *Criteria for discontinuation of a participant in the trial*

Trial participants may be discontinued from the trial entirely or from one or more domain-specific interventions according to predefined criteria for discontinuation. The criteria for discontinuation specific to each domain are specified in the relevant DSA.

Not for IRB submission

Criteria for discontinuation from the REMAP interventions entirely include:

1. The treating clinician considers continued participation in the REMAP interventions are not deemed to be in the best interests of the patient
2. The participant or their Legal Representative requests withdrawal from ongoing participation in all REMAP interventions

In the case of discontinuation, the reasons for withdrawal will be documented. Consent to the use of study data, including data collected until the time of discontinuation and data to inform primary and secondary outcome data will be requested specifically from participants or their Legal Representative who request discontinuation. Following discontinuation of a REMAP intervention, participants will be treated according to standard ICU management. Participants who are withdrawn will not be replaced. All data will be analyzed using the ITT principle.

#### 8.8. Concomitant care and co-interventions

All treatment decisions outside of those specified within the REMAP will be at the discretion of the treating clinician. Prespecified co-interventions related to specific domains will be recorded in the CRF and are outlined in the relevant DSAs.

#### 8.9. Data collection

##### 8.9.1. Principles of data collection

Streamlined data collection instruments and procedures will be used to minimize the workload in study sites. The CRF will be developed by the ITSC and made available to the participating sites as a paper and electronic CRF (eCRF) for ease of data collection. Data may be entered directly into the eCRF or first entered onto a paper copy of the CRF and entered subsequently into the eCRF. All data will be collected by trained staff who will have access to a comprehensive data dictionary. Information recorded in the CRF should accurately reflect the subject’s medical/ hospital notes, must be completed as soon as it is made available, and must be collected from source data. The intent of this process is to improve the quality of the clinical study including being able to provide prompt feedback to the site staff on the progress, accuracy, and completeness of the data submitted. The eCRF will be web-based and accessible by a site or investigator specific password protected.

Not for IRB submission

##### 8.9.2. Variables to be collected

The generic variables to be collected for all domains in this REMAP are as detailed, indicatively, in the Core Protocol, below. Additional domain-specific variables are outlined in the relevant DSAs. Baseline variables are defined as at or before the time of randomization.

###### 8.9.2.1. *Baseline and required for randomization*

- Overall REMAP Inclusion / exclusion check list
- Date and time of hospital admission
- Date and time of first ICU admission
- Domain-specific exclusion checklist
- Shock status
- Hypoxemia status
- Influenza status
- Pandemic status

###### 8.9.2.2. *Baseline but not required for randomization*

- Demographic data (date of birth, age, sex, estimated body weight and height)
- Co-existing illnesses and risk factors for pneumonia
- Source of ICU admission
- Acute Physiology and Chronic Health Evaluation (APACHE) II variables
- Sequential Organ Failure Assessment (SOFA) variables
- Intervention allocation status within domains and randomization number
- Results of microbiological testing

###### 8.9.2.3. *Daily from randomization until discharge from ICU or Day-28 whichever comes first*

- Hypotension and administration of vasopressors/inotropes
- Administration of dialysis
- Administration of invasive or non-invasive ventilation
- P:F ratio components

###### 8.9.2.4. *ICU Outcome data*

- Date and time of ICU discharge
- Survival status at ICU discharge

Not for IRB Submission

- Dates of ICU readmission and discharge

###### 8.9.2.5. *Hospital outcome data*

- Date and time of hospital discharge
- Survival status at hospital discharge
- Discharge destination
- Results of microbiological testing

###### 8.9.2.6. *Antimicrobial Administration*

- Administration of antibiotic medications
- Administration of antiviral medications

###### 8.9.2.7. *Outcome data*

At the discretion of the site, unless specified otherwise in a RSA or DSA, and collected by phone:

- Survival status at 90 days
- Survival status at 6 months
- HRQoL measured by EQ-5D at 6 months
- Disability status measured by WHODAS at 6 months and baseline information to interpret disability
- Opinions and beliefs regarding participation in research (reported at 6 months)

###### 8.9.2.8. *Process-related outcomes*

- Time from index hospital admission to ICU admission
- Time from ICU admission to randomization
- Selected co-interventions
- Compliance with allocated intervention(s).

##### 8.9.3. Data required to inform Response Adaptive Randomization

This REMAP will use frequent adaptive analyses and incorporate RAR. All variables used to inform RAR will be pre-specified. The key variables include:

2. Baseline and allocation status
    a. Unique trial-specific number
    b. Location (Country and Site code)

Not for IRB Submission

c. Date and time of randomization

d. Eligibility for each domain

e. Intervention allocation for each domain

f. Reveal status for each intervention allocation for each domain

g. Age category

h. Strata

   i. Shock or no shock

   iii. Influenza status

   iv. Pandemic strata

i. State

   i. Hypoxemia

3. Outcome

   a. All-cause mortality at 90 days

   b. Date of hospital discharge

Data fields required to inform the adaptive randomization process and Statistical Trigger will be pre-specified and will be required to be entered into the eCRF within 7 days of death and within 97 days of enrollment into the REMAP if the participant is alive at 90 days.

###### 8.9.4. Blinding of outcome assessment

Wherever feasible outcome assessment will be undertaken by research staff who are blinded to allocation status. Such blinding will not be feasible for many outcomes, particularly those that occur while the participant is still admitted to an ICU or the hospital. However, the primary endpoint and key secondary endpoints are not variables that are open to interpretation and so accuracy will not be affected by outcome assessors not being blinded to allocation status.

##### *8.10. Data management*

###### 8.10.1. Source Data

Source documents are where data are first recorded, and from which participants' eCRF data are obtained. These include, but are not limited to, hospital records (from which medical history and previous and concurrent medication may be summarized into the eCRF), clinical and office charts, laboratory and pharmacy records, radiographs, and correspondence.

Not for IRB submission

##### 8.10.2. Confidentiality

All documents will be stored safely in confidential conditions. On all trial-specific documents, other than the signed consent, the participant will be referred to by a unique trial-specific number and/or code in any database, not by name. Information linking the participant's medical data to database materials will be maintained in a secure location at the participating site. This information will not be transmitted to the members of the ITSC, any DSWG, or RMC. The key to code and recode participant identifiers will only be accessible to local site investigators (research nurse and principal investigator) but not to members of the central study team. ICU and coded individual subject data and records will be held in strictest confidence by the site investigator and healthcare staff and by all central research staff, as permitted by law.

#### *8.11. Quality assurance and monitoring*

The trial will be conducted in accordance with the current approved protocol, Good Clinical Practice (GCP), relevant regulations and SOPs.

##### 8.11.1. Plans for improving protocol adherence and complete data

Data entry and data management will be coordinated by the Regional Project Manager and the RCC, including programming and data management support.

Several procedures to ensure data quality and protocol standardization will help to minimize bias. These include:

- Start-up meeting for all research coordinators and investigators will be held prior to study commencement to ensure consistency in procedures;
- A detailed dictionary will define the data to be collected on the CRF;
- The data management center will perform timely validation of data, queries and corrections if errors are found during quality control checks;
- Data monitoring will occur as described below.

##### 8.11.2. Data Monitoring

The study will be monitored by a representative of the RCC. A site initiation teleconference or visit will be conducted before site activation. Routine monitoring visits will be conducted the frequency of which will be determined by each site's rate of recruitment. Email and telephone communication will supplement site visits.

Not for IRB submission

A monitoring report will be prepared following each visit and reviewed by the RMC if appropriate. A follow up letter will be sent to the principal investigator and research coordinator at the site and will be filed in the site investigator file.

Medical records, any other relevant source documents and the site investigator files must be made available to the representative of the RCC for these monitoring visits during the course of the study and at the completion of the study as needed.

Domain-specific monitoring and protocol adherence issues are addressed in each DSA.

#### 8.12. Data safety and monitoring board

A single DSMB will take responsibility for the trial in all regions in which it is conducted. The DSMB compiled for this study will consist of 5-7 members; the chair has been selected to have expertise in clinical trial methodology, and to have experience with adaptive clinical trial design. Additional medical, statistical, and other experts will be selected to ensure all necessary expertise to oversee a trial of this complexity and scope. The DSMB will conduct its activities in accordance with a separate Charter; the Charter must be approved by the DSMB, and ITSC prior to the initiation of the trial. The DSMB will be unblinded to ensure the highest quality oversight of the trial, in accordance with current recommendations of regulatory authorities.

The DSMB will review received frequent updates of the trial's adaptive analyses from the SAC. The role of the DSMB will be to ensure that the pre-specified trial algorithm is being implemented as designed, that the design remains appropriate from a scientific and ethical point of view, to confirm when a Statistical Trigger has been reached, and to either reach or recommend that a Platform Conclusion has been reached, as outlined in Section 7.8.9. Trial enrollment and conduct will be continuous.

The DSMB will not make design decisions. If the DSMB believes the trial's algorithms are no longer acceptable from an ethical, safety, or scientific point of view it will make recommendations to the ITSC which has ultimate decision-making authority regarding the trial design. Where the DSMB and the SAC agree on a temporary deviation from the study protocol for safety reasons, they are not required to inform the ITSC of this decision. If the DSMB and SAC agree that a permanent change is necessary, the chairs of the DSMB, SAC and ITSC will meet to discuss the best way to proceed to ensure patient safety and the scientific integrity of the trial. Where the SAC and DSMB disagree on the need to deviate from the pre-specified trial design, the DSMB must inform the ITSC of their recommendations and the rationale for these.

Not for IRB Submission

#### 8.13. *Safety monitoring and reporting*

##### 8.13.1. Principles

The principles used in the conduct of safety monitoring and reporting in this trial are those outlined by Cook *et al.* in the manuscript "Serious adverse events in academic critical care research". (Cook et al., 2008) A high proportion of critically ill patients who will be enrolled in this trial will experience mortality or substantial morbidity. The case-fatality proportion for critically ill patients with CAP is likely to be in the order of 20 to 30% and high proportions of patients will have one or both of laboratory abnormalities or complications of critical illness and its treatment. Patients who are critically ill, irrespective of whether or not they are enrolled in a trial, will typically experience multiple events that would meet the conventional definition of a Serious Adverse Event (SAE).

Trials involving vulnerable populations must have research oversight that protects patient safety and patient rights and also ensures that there can be public trust that the trial is conducted in a manner that safeguards the welfare of participants. The strategy outlined for the definition, attribution, and reporting of SAEs in this trial is designed to achieve these goals but does so in a way that seeks to avoid the reporting of events that are likely to be part of the course of the illness or events that are recognized as important by their incorporation as trial endpoints.

##### 8.13.2. Definition

In accordance with accepted standards a SAE is defined as an event that is fatal, life-threatening, results in (or may result) in disability that is long-lasting and significant, or results in a birth defect or congenital anomaly.

##### 8.13.3. Reporting Procedures for Serious Adverse Events

The trial endpoints, as outlined in the Core Protocol and as specified in DSAs, are designed to measure the vast majority of events that might otherwise constitute an SAE. In particular, SAEs that might be attributable to specific interventions are included as secondary endpoints in each DSA but are recorded only for participants who are enrolled in that domain. If required, additional clarification of issues related to the identification of SAEs that are relevant to a specific domain will be described in the DSA. Generally, only SAEs that are not trial-end points require reporting.
However, any SAE that is considered by the site-investigator to be attributable to a study intervention or study participation should be reported (Section 8.13.4). Where an SAE is not a trial end point it should be reported only where, in the opinion of the site-investigator, the event might

Not for IRB Submission

reasonably have occurred as consequence of a study intervention or study participation (Section 8.13.4).

Events that meet the definition of an SAE, require reporting in accordance with the criteria outlined above, and occur between trial enrollment but before hospital discharge will be reported to a RCC. These SAEs should be reported to a RCC within 72 hours of trial staff becoming aware of the event, unless otherwise specified in a RSA. The minimum information that will be reported will comprise:

- Unique trial-specific number
- Date(s) of the event
- Nature of the event, including its outcome, and the rationale for attribution to a trial intervention
- Whether treatment was required for the event and, if so, what treatment was administered

##### 8.13.4. Attribution of serious events to study interventions

It is likely that many participants within the trial will experience events that could be attributed to one or more study interventions. However, it will often be difficult to distinguish, in real-time, between events that occur as a consequence of critical illness and treatments that are not specified by the trial, and interventions specified by the trial. Site investigators should exercise caution in attributing events to study interventions. However, the standard that should be applied to determine whether SAEs are attributable to study interventions in this trial is that it is possible, probable, or certain that there is a direct link between a trial intervention and the SAE or the SAE is not considered to be a normal feature of the evolution of critical illness and its treatment.

##### 8.13.5. Attribution of a death to study interventions or study participation

Critically ill patients who will be enrolled in this trial are at high risk of death. The primary endpoint of the trial is mortality and the objective of the trial is to identify differences in the primary endpoint that can be attributed to treatment allocation which will often include treatments that are believed to be or known to be safe and effective but for which it is not known whether some treatments are more effective than others. Where the trial evaluates interactions that are novel and not part of usual standard care the threshold for considering attribution to the novel experimental intervention should be lower than if an intervention is already in widespread use and its safety profile has already been established.

Not for IRB Submission

### 9. GOVERNANCE AND ETHICAL CONSIDERATIONS

#### 9.1. Management of participating sites and trial coordination

Each region will have a RCC. Each RCC will take primary responsibility for the management of participating sites, data management for those sites, and provide web-based randomization for sites in its region. The processes by which each RCC will provide trial management and coordination is set out in each RSA.

#### 9.2. Ethics and regulatory issues

##### 9.2.1. Guiding principles

The study will be conducted according to the principles of the latest version of the Declaration of Helsinki (version Fortaleza 2013) and in accordance with all relevant local ethical, regulatory, and legal requirements as specified in each RSA.

##### 9.2.2. Ethical issues relevant to this study

Patients who will be eligible for this study are critically ill, and many eligible patients will be receiving sedative medications for comfort, safety and to facilitate standard life saving ICU procedures. In patients who are not necessarily receiving sedative medications, the presence of critical illness, itself, leads commonly to an altered mental state that will affect the patient's mental capacity. The presence of these factors will mean that most patients who are eligible for the study will not be able to provide prospective consent for participation. Additionally, many interventions within this trial must be initiated urgently, either because there is an immediate time critical imperative to initiate the intervention or because the most valid evaluation of the intervention occurs if the trial intervention is initiated at the same time-point as would occur in clinical practice.

The broad approach regarding consent that will be used in this study are as follows:

- Patients who, in the opinion of the treating clinician, are competent to consent will be provided with information about the trial and invited to participate
- The vast majority of patients who are eligible for the REMAP will not be competent to consent. For such patients, and as permitted by local laws and requirements for ethical approval:
    - For domains in which all interventions available at the participating site are regarded as being part of the spectrum of acceptable standard care by the

Not for IRB Submission

clinicians at that site, entry to the study is preferred to be via waiver-of- consent or some form of delayed consent. If required by local laws or ethical requirements and alternative to this pathway will be participation in conjunction with the agreement of an authorized representative of the participant.

- For domains in which at least one intervention available at the participating site is regarded as experimental or not part of the spectrum of acceptable standard care then prospective agreement by an authorized representative will be required. An exception to this principle is recognized when there is a time-imperative to commence the intervention which would routinely preclude obtaining the prospective agreement by an authorized representative.
- For domains in which eligibility may develop after initial enrollment in the trial it is permissible to obtain contingent consent from the participant or contingent agreement from an authorized representative, i.e. there is contingent approval to randomize the participant if the participant meets eligibility criteria for a domain subsequently.
- Where any participant is enrolled without having provided their own consent, the participant's authorized representative will be informed as soon as appropriate and informed of processes to cease trial participation. If required by local laws or processes for ethical approval, the authorized representative will be asked to provide agreement to on-going participation. In undertaking these trial processes research staff will be cognizant of the need to avoid unnecessary distress or create unnecessary confusion for authorized representatives and all other persons who have an interest in the participant's welfare.
- Where any participant is enrolled without having provided their own consent, the participant should be informed of their enrollment after regaining competency, in accordance with local practice and jurisdictional requirements. Where any participant is enrolled and does not regain competency (due to their death or neurological impairment) the default position, subject to local laws and ethical review processes, will be that the enrolled person will continue to be a participant in the trial.

Not for IRB submission

It should be noted that once RAR is initiated, participants within the REMAP, on average, derive benefit from participation. As a consequence of RAR participants are more likely to be allocated to the interventions within each domain that are more likely to result in better outcomes.

###### 9.2.3. Approvals

The protocol, consent form(s) and participant and/or authorized representative information sheet(s) will be submitted to an appropriate ethical review body at each participating institution and, as required, to any additional regulatory authorities. Written approval to commence the study is required for all relevant ethical and regulatory bodies.

##### *9.3. Protocol modifications*

###### 9.3.1. Amendments

A “substantial amendment” is defined as an amendment to one or more of the Core Protocol, DSA, or RSA that is likely to affect to a significant degree:

- the safety or physical or mental integrity of the subjects of the trial;
- the scientific value of the trial;
- the conduct or management of the trial;
- the quality or safety of any intervention used in the trial;
- cessation of any intervention or domain for any reason;
- the addition of any new intervention within a domain; or
- the addition of new interventions within a new domain

All substantial amendments to the original approved documents, including all modifications of interventions available within a domain and the addition of interventions within a new domain will be submitted for approval to all relevant ethical and regulatory review bodies that were required for original approvals. Non-substantial amendments will not be notified to such review bodies, but will be recorded and filed by the trial sponsors.

Where the cessation of any intervention or any domain occurs for any reason, this is an operational issue and randomization to that intervention or domain will no longer be available. Cessation of an intervention or domain, either entirely, or within a prespecified subgroup, will be reported to all relevant regulatory bodies.

Not for IRB Submission

#### 9.4. Confidentiality

The principles of confidentiality that will apply to this trial, are that all trial staff will ensure that the confidentiality of all participants information will be maintained and preserved at all times. The participants will be identified only by a unique trial-specific number on all documents and electronic databases that contain any information specific to the participating individual. Each site will maintain a separate file that links each participant's unique trial-specific number to the participant's name and other identifying information such as date of birth, address, and other contact information. No other information will be maintained in the file that links the participant unique trial-specific number to participant identifying information.

#### 9.5. Declarations of interest

All trial staff will be required to declare and update all interests that might or might be seen to influence one or both of the conduct of the trial or the interpretation of results. All investigators involved in REMAP-CAP maintain a registry of interests on the REMAP-CAP website. These are updated periodically and publicly accessible on the study website.

#### 9.6. Post-trial care

The trial has no responsibility for the ongoing management or care of participants following the cessation of all trial specified interventions.

#### 9.7. Communication

##### 9.7.1. Reporting

Each participating site will comply with all local reporting requirements, as specified by that site's institution.

Should the entire trial be terminated, all relevant local ethical and regulatory bodies will be informed within 90 days after the end of the study. The end of the study is defined as the last participant's last follow-up.

##### 9.7.2. Communication of trial results

Trial results will be communicated by presentation and publication.

Not for IRB Submission

#### 9.8. Publication policy

Manuscript(s) and abstract(s) resulting from the data collected during this study will be prepared by the corresponding DSWG. Where results are influenced by interaction between domains, the DSWG for both domains will take responsibility for preparation of manuscripts and abstracts. All manuscripts and abstracts reporting trial results that are prepared by one or more DSWGs must be submitted to and approved by the ITSC before submission.

Site investigators will not publish or present interim or definite results, including but not restricted to oral presentations. The role of site investigators and research coordinators at participating sites will be acknowledged by their names being listed as collaborators. Where required publications will comply with the publication policies of clinical trials groups that have endorsed or supported the study.

#### 9.9. Data access and ownership

##### 9.9.1. Data ownership

All data are owned by the responsible sponsor under the custodianship of the ITSC. As the trial is intended to be perpetual, all data will be retained indefinitely.

##### 9.9.2. Access to Data

Direct access will be granted to authorized representatives from ITSC, sponsors, host institution and the regulatory authorities to permit trial-related monitoring, audits and inspections. The trial will comply with all relevant jurisdictional and academic requirements relating to access to data, as apply at the time that the data are generated. Ownership and access to data where a commercial organization is involved in the trial (for example by provision of goods or services that are tested within a domain) will be set out in a contract between trial sponsors and that commercial organization.

The trial will not enter into a contract with a commercial organization unless the contract specifies that:

- There is complete academic independence with regard to the design and conduct of all aspects of the trial including analysis and reporting of trial results

Not for IRB submission

- May agree to provide a pre-publication version of presentations or manuscripts to a commercial organization but that the commercial organization has no authority to prevent or modify presentation or publication
- That all data are owned by the trial and the commercial organization has no authority to access data

##### 9.10. Consent form

Template information and consent forms will be provided to participating sites as an operational document.

Not for IRB Submission

### 10.REFERENCES

ABEREGG, S. K., RICHARDS, D. R. & O'BRIEN, J. M. 2010. Delta inflation: a bias in the design of randomized controlled trials in critical care medicine. *Crit Care,* 14**,** R77.

AIKMAN, K. L., HOBBS, M. R., TICEHURST, R., KARMAKAR, G. C., WILSHER, M. L. & THOMAS, M. G. 2013. Adherence to guidelines for treating community-acquired pneumonia at a New Zealand hospital. *J Pharm Pract Res,* 43**,** 272-275.

ALMIRALL, J., BOLIBAR, I., VIDAL, J., SAUCA, G., COLL, P., NIKLASSON, B., BARTOLOME, M. & BALANZO, X. 2000. Epidemiology of community-acquired pneumonia in adults: a population- based study. *Eur.Respir.J.,* 15**,** 757-763.

ALVAREZ-LERMA, F. & TORRES, A. 2004. Severe community-acquired pneumonia. *Curr.Opin.Crit Care,* 10**,** 369-374.

ANGUS, D. C. 2015. Fusing Randomized Trials With Big Data: The Key to Self-learning Health Care Systems? *JAMA,* 314**,** 767-8.

BARKER, A. D., SIGMAN, C. C., KELLOFF, G. J., HYLTON, N. M., BERRY, D. A. & ESSERMAN, L. J. 2009. I-SPY 2: an adaptive breast cancer trial design in the setting of neoadjuvant chemotherapy. *Clin.Pharmacol.Ther.,* 86**,** 97-100.

BELLANI, G., LAFFEY, J. G., PHAM, T., FAN, E., BROCHARD, L., ESTEBAN, A., GATTINONI, L., VAN HAREN, F., LARSSON, A., MCAULEY, D. F., RANIERI, M., RUBENFELD, G., THOMPSON, B. T., WRIGGE, H., SLUTSKY, A. S., PESENTI, A., INVESTIGATORS, L. S. & GROUP, E. T. 2016. Epidemiology, Patterns of Care, and Mortality for Patients With Acute Respiratory Distress Syndrome in Intensive Care Units in 50 Countries. *JAMA,* 315**,** 788-800.

BERRY, D. A. 2012. Adaptive clinical trials in oncology. *Nat.Rev.Clin.Oncol.,* 9**,** 199-207.

BERRY, S. M., CONNOR, J. T. & LEWIS, R. J. 2015. The platform trial: an efficient strategy for evaluating multiple treatments. *JAMA,* 313**,** 1619-1620.

BHATT, D. L. & MEHTA, C. 2016. Adaptive Designs for Clinical Trials. *N Engl J Med,* 375**,** 65-74.

BJERRE, L. M., VERHEIJ, T. J. & KOCHEN, M. M. 2009. Antibiotics for community acquired pneumonia in adult outpatients. *Cochrane.Database.Syst.Rev.***,** CD002109.

BLOOD OBSERVATIONAL STUDY INVESTIGATORS OF ANZICS-CLINICAL TRIALS GROUP, WESTBROOK, A., PETTILA, V., NICHOL, A., BAILEY, M. J., SYRES, G., MURRAY, L., BELLOMO, R., WOOD, E., PHILLIPS, L. E., STREET, A., FRENCH, C., ORFORD, N., SANTAMARIA, J. & COOPER, D. J. 2010. Transfusion practice and guidelines in Australian and New Zealand intensive care units. *Intensive Care Med,* 36**,** 1138-46.

BODI, M., RODRIGUEZ, A., SOLE-VIOLAN, J., GILAVERT, M. C., GARNACHO, J., BLANQUER, J., JIMENEZ, J., DE LA TORRE, M. V., SIRVENT, J. M., ALMIRALL, J., DOBLAS, A., BADIA, J. R., GARCIA, F., MENDIA, A., JORDA, R., BOBILLO, F., VALLES, J., BROCH, M. J., CARRASCO, N., HERRANZ, M. A. & RELLO, J. 2005. Antibiotic prescription for community-acquired pneumonia in the intensive care unit: impact of adherence to Infectious Diseases Society of America guidelines on survival. *Clin.Infect.Dis.,* 41**,** 1709-1716.

BURNS, K. E., CHANT, C., SMITH, O., CUTHBERTSON, B., FOWLER, R., COOK, D. J., KRUGER, P., WEBB, S., ALHASHEMI, J., DOMINGUEZ-CHERIT, G., ZALA, C., RUBENFELD, G. D. & MARSHALL, J. C. 2011. A Canadian Critical Care Trials Group project in collaboration with the international

Not for IRB submission

forum for acute care trialists - Collaborative H1N1 Adjuvant Treatment pilot trial (CHAT): study protocol and design of a randomized controlled trial. *Trials,* 12**,** 70.

BYRNE, R. A. & KASTRATI, A. 2013. Unmet aspirations--where to now for catheter thrombectomy? *N Engl J Med,* 369**,** 1649-50.

CAREY, L. A. & WINER, E. P. 2016. I-SPY 2--Toward More Rapid Progress in Breast Cancer Treatment. *N Engl J Med,* 375**,** 83-4.

CECCONI, M., HOFER, C., TEBOUL, J. L., PETTILA, V., WILKMAN, E., MOLNAR, Z., DELLA ROCCA, G., ALDECOA, C., ARTIGAS, A., JOG, S., SANDER, M., SPIES, C., LEFRANT, J. Y., DE BACKER, D., INVESTIGATORS, F. & GROUP, E. T. 2015. Fluid challenges in intensive care: the FENICE study: A global inception cohort study. *Intensive Care Med,* 41**,** 1529-37.

CONNOR, J. T., ELM, J. J. & BROGLIO, K. R. 2013. Bayesian adaptive trials offer advantages in comparative effectiveness trials: an example in status epilepticus. *J.Clin.Epidemiol.,* 66**,** S130- S137.

COOK, D., LAUZIER, F., ROCHA, M. G., SAYLES, M. J. & FINFER, S. 2008. Serious adverse events in academic critical care research. *CMAJ,* 178**,** 1181-4.

DELLINGER, R. P., LEVY, M. M., RHODES, A., ANNANE, D., GERLACH, H., OPAL, S. M., SEVRANSKY, J. E., SPRUNG, C. L., DOUGLAS, I. S., JAESCHKE, R., OSBORN, T. M., NUNNALLY, M. E., TOWNSEND, S. R., REINHART, K., KLEINPELL, R. M., ANGUS, D. C., DEUTSCHMAN, C. S., MACHADO, F. R., RUBENFELD, G. D., WEBB, S. A., BEALE, R. J., VINCENT, J. L., MORENO, R. & SURVIVING SEPSIS CAMPAIGN GUIDELINES COMMITTEE INCLUDING THE PEDIATRIC, S. 2013.
Surviving sepsis campaign: international guidelines for management of severe sepsis and septic shock: 2012. *Crit Care Med,* 41**,** 580-637.

ECCLES, S., PINCUS, C., HIGGINS, B. & WOODHEAD, M. 2014. Diagnosis and management of community and hospital acquired pneumonia in adults: summary of NICE guidance. *BMJ,* 349**,** g6722.

EUROPEAN MEDICINES AGENCY 2005. Guideline on the choice of the non-inferiority margin. *In:* COMMITTEE FOR MEDICINAL PRODUCTS FOR HUMAN USE (CHMP) (ed.). London.

EWIG, S., WOODHEAD, M. & TORRES, A. 2011. Towards a sensible comprehension of severe community-acquired pneumonia. *Intensive Care Med.,* 37**,** 214-223.

FINFER, S., BELLOMO, R., BOYCE, N., FRENCH, J., MYBURGH, J. & NORTON, R. 2004. A comparison of albumin and saline for fluid resuscitation in the intensive care unit. *N.Engl.J.Med.,* 350**,** 2247- 2256.

FINFER, S., LIU, B., TAYLOR, C., BELLOMO, R., BILLOT, L., COOK, D., DU, B., MCARTHUR, C., MYBURGH, J. & INVESTIGATORS, S. T. 2010. Resuscitation fluid use in critically ill adults: an international cross-sectional study in 391 intensive care units. *Crit Care,* 14**,** R185.

FREI, C. R., ATTRIDGE, R. T., MORTENSEN, E. M., RESTREPO, M. I., YU, Y., ORAMASIONWU, C. U., RUIZ, J. L. & BURGESS, D. S. 2010. Guideline-concordant antibiotic use and survival among patients with community-acquired pneumonia admitted to the intensive care unit.
*Clin.Ther.,* 32**,** 293-299.

HARRINGTON, D. & PARMIGIANI, G. 2016. I-SPY 2--A Glimpse of the Future of Phase 2 Drug Development? *N Engl J Med,* 375**,** 7-9.

HUANG, D. T., ANGUS, D. C., MOSS, M., THOMPSON, B. T., FERGUSON, N. D., GINDE, A., GONG, M. N., GUNDEL, S., HAYDEN, D. L., HITE, R. D., HOU, P. C., HOUGH, C. L., IWASHYNA, T. J., LIU, K. D., TALMOR, D. S., YEALY, D. M., COMMITTEE, R. P. & THE, N. I. H. N. P. N. I. 2016. Design

Not for IRB Submission

and Rationale of the Reevaluation of Systemic Early Neuromuscular Blockade (ROSE) Trial for Acute Respiratory Distress Syndrome. *Ann Am Thorac Soc*.

INSTITUTE OF MEDICINE 2013. *Best Care at Lower Cost: The Path to Continuously Learning Health Care in America,* Washington D.C, The National Academies Press.

LEE, J. S., NSA, W., HAUSMANN, L. R., TRIVEDI, A. N., BRATZLER, D. W., AUDEN, D., MOR, M. K., BAUS, K., LARBI, F. M. & FINE, M. J. 2014. Quality of care for elderly patients hospitalized for pneumonia in the United States, 2006 to 2010. *JAMA Intern.Med.,* 174**,** 1806-1814.

LEROY, O., SANTRE, C., BEUSCART, C., GEORGES, H., GUERY, B., JACQUIER, J. M. & BEAUCAIRE, G. 1995. A five-year study of severe community-acquired pneumonia with emphasis on prognosis in patients admitted to an intensive care unit. *Intensive Care Med.,* 21**,** 24-31.

LIM, W. S., BAUDOUIN, S. V., GEORGE, R. C., HILL, A. T., JAMIESON, C., LE, J., I, MACFARLANE, J. T., READ, R. C., ROBERTS, H. J., LEVY, M. L., WANI, M. & WOODHEAD, M. A. 2009. BTS guidelines for the management of community acquired pneumonia in adults: update 2009. *Thorax,* 64 Suppl 3**,** iii1-55.

MANDELL, L. A., WUNDERINK, R. G., ANZUETO, A., BARTLETT, J. G., CAMPBELL, G. D., DEAN, N. C., DOWELL, S. F., FILE, T. M., JR., MUSHER, D. M., NIEDERMAN, M. S., TORRES, A. & WHITNEY, C. G. 2007. Infectious Diseases Society of America/American Thoracic Society consensus guidelines on the management of community-acquired pneumonia in adults. *Clin Infect Dis,* 44 Suppl 2**,** S27-S72.

MCCABE, C., KIRCHNER, C., ZHANG, H., DALEY, J. & FISMAN, D. N. 2009. Guideline-concordant therapy and reduced mortality and length of stay in adults with community-acquired pneumonia: playing by the rules. *Arch Intern Med* 169**,** 1525-1531.

MEURER, W. J., LEWIS, R. J. & BERRY, D. A. 2012. Adaptive clinical trials: a partial remedy for the therapeutic misconception? *JAMA,* 307**,** 2377-2378.

MORTENSEN, E. M., RESTREPO, M., ANZUETO, A. & PUGH, J. 2004. Effects of guideline-concordant antimicrobial therapy on mortality among patients with community-acquired pneumonia. *Am J Med,* 117**,** 726-731.

MORTENSEN, E. M., RESTREPO, M. I., ANZUETO, A. & PUGH, J. 2005. The impact of empiric antimicrobial therapy with a beta-lactam and fluoroquinolone on mortality for patients hospitalized with severe pneumonia. *Crit Care,* 10**,** R8.

MUSHER, D. M., ROIG, I. L., CAZARES, G., STAGER, C. E., LOGAN, N. & SAFAR, H. 2013. Can an etiologic agent be identified in adults who are hospitalized for community-acquired pneumonia: results of a one-year study. *J Infect,* 67**,** 11-8.

MUSHER, D. M. & THORNER, A. R. 2014. Community-acquired pneumonia. *N.Engl.J.Med.,* 371**,** 1619-1628.

PARK, J. W., LIU, M. C., YEE, D., YAU, C., VAN 'T VEER, L. J., SYMMANS, W. F., PAOLONI, M., PERLMUTTER, J., HYLTON, N. M., HOGARTH, M., DEMICHELE, A., BUXTON, M. B., CHIEN, A. J., WALLACE, A. M., BOUGHEY, J. C., HADDAD, T. C., CHUI, S. Y., KEMMER, K. A., KAPLAN, H. G., ISAACS, C., NANDA, R., TRIPATHY, D., ALBAIN, K. S., EDMISTON, K. K., ELIAS, A. D., NORTHFELT, D. W., PUSZTAI, L., MOULDER, S. L., LANG, J. E., VISCUSI, R. K., EUHUS, D. M., HALEY, B. B., KHAN, Q. J., WOOD, W. C., MELISKO, M., SCHWAB, R., HELSTEN, T., LYANDRES, J., DAVIS, S. E., HIRST, G. L., SANIL, A., ESSERMAN, L. J., BERRY, D. A. & INVESTIGATORS, I. S. 2016. Adaptive Randomization of Neratinib in Early Breast Cancer. *N Engl J Med,* 375**,** 11-22.

RHODES, A., EVANS, L. E., ALHAZZANI, W., LEVY, M. M., ANTONELLI, M., FERRER, R., KUMAR, A., SEVRANSKY, J. E., SPRUNG, C. L., NUNNALLY, M. E., ROCHWERG, B., RUBENFELD, G. D., ANGUS, D. C., ANNANE, D., BEALE, R. J., BELLINGHAN, G. J., BERNARD, G. R., CHICHE, J. D.,

Not for IRB submission

COOPERSMITH, C., DE BACKER, D. P., FRENCH, C. J., FUJISHIMA, S., GERLACH, H., HIDALGO, J. L., HOLLENBERG, S. M., JONES, A. E., KARNAD, D. R., KLEINPELL, R. M., KOH, Y., LISBOA, T. C., MACHADO, F. R., MARINI, J. J., MARSHALL, J. C., MAZUSKI, J. E., MCINTYRE, L. A., MCLEAN, A. S., MEHTA, S., MORENO, R. P., MYBURGH, J., NAVALESI, P., NISHIDA, O., OSBORN, T. M., PERNER, A., PLUNKETT, C. M., RANIERI, M., SCHORR, C. A., SECKEL, M. A., SEYMOUR, C. W., SHIEH, L., SHUKRI, K. A., SIMPSON, S. Q., SINGER, M., THOMPSON, B. T., TOWNSEND, S. R., VAN DER POLL, T., VINCENT, J. L., WIERSINGA, W. J., ZIMMERMAN, J. L. & DELLINGER, R.P. 2017. Surviving Sepsis Campaign: International Guidelines for Management of Sepsis and Septic Shock: 2016. *Intensive Care Med,* 43**,** 304-377.

RUGO, H. S., OLOPADE, O. I., DEMICHELE, A., YAU, C., VAN 'T VEER, L. J., BUXTON, M. B., HOGARTH, M., HYLTON, N. M., PAOLONI, M., PERLMUTTER, J., SYMMANS, W. F., YEE, D., CHIEN, A. J., WALLACE, A. M., KAPLAN, H. G., BOUGHEY, J. C., HADDAD, T. C., ALBAIN, K. S., LIU, M. C., ISAACS, C., KHAN, Q. J., LANG, J. E., VISCUSI, R. K., PUSZTAI, L., MOULDER, S. L., CHUI, S. Y., KEMMER, K. A., ELIAS, A. D., EDMISTON, K. K., EUHUS, D. M., HALEY, B. B., NANDA, R., NORTHFELT, D. W., TRIPATHY, D., WOOD, W. C., EWING, C., SCHWAB, R., LYANDRES, J., DAVIS, S. E., HIRST, G. L., SANIL, A., BERRY, D. A., ESSERMAN, L. J. & INVESTIGATORS, I. S. 2016. Adaptive Randomization of Veliparib-Carboplatin Treatment in Breast Cancer. *N Engl J Med,* 375**,** 23-34.

SHORR, A. F., BODI, M., RODRIGUEZ, A., SOLE-VIOLAN, J., GARNACHO-MONTERO, J. & RELLO, J. 2006. Impact of antibiotic guideline compliance on duration of mechanical ventilation in critically ill patients with community-acquired pneumonia. *Chest,* 130**,** 93-100.

SINGANAYAGAM, A., CHALMERS, J. D. & HILL, A. T. 2009. Severity assessment in community-acquired pneumonia: a review. *QJM.,* 102**,** 379-388.

SLIGL, W. I. & MARRIE, T. J. 2013. Severe community-acquired pneumonia. *Crit Care Clin.,* 29**,** 563-601.

THE ACUTE RESPIRATORY DISTRESS SYNDROME NETWORK 2000. Ventilation with lower tidal volumes as compared with traditional tidal volumes for acute lung injury and the acute respiratory distress syndrome. The Acute Respiratory Distress Syndrome Network. *N.Engl.J.Med.,* 342**,** 1301-1308.

U.S. DEPARTMENT OF HEALTH AND HUMAN SERVICES 2016. Non-inferiority clinical trials to establish effectiveness: guidance for industry. *In:* FOOD AND DRUG ADMINISTRATION (ed.).

WARE, J. H. & ANTMAN, E. M. 1997. Equivalence trials. *N Engl J Med,* 337**,** 1159-61.

WIERSINGA, W. J., BONTEN, M. J., BOERSMA, W. G., JONKERS, R. E., ALEVA, R. M., KULLBERG, B. J., SCHOUTEN, J. A., DEGENER, J. E., JANKNEGT, R., VERHEIJ, T. J., SACHS, A. P. & PRINS, J. M. 2012. SWAB/NVALT (Dutch Working Party on Antibiotic Policy and Dutch Association of Chest Physicians) guidelines on the management of community-acquired pneumonia in adults. *Neth.J.Med.,* 70**,** 90-101.

WILKINSON, M. & WOODHEAD, M. A. 2004. Guidelines for community-acquired pneumonia in the ICU. *Curr.Opin.Crit Care,* 10**,** 59-64.

WOODHEAD, M., BLASI, F., EWIG, S., GARAU, J., HUCHON, G., IEVEN, M., ORTQVIST, A., SCHABERG, T., TORRES, A., VAN DER, H. G., READ, R. & VERHEIJ, T. J. 2011. Guidelines for the management of adult lower respiratory tract infections--full version. *Clin.Microbiol.Infect.,* 17 Suppl 6**,** E1-59.

WORLD HEALTH ORGANIZATION 2008. The Global burden of disease, 2004 update. Geneva, Switzerland: WHO Press.

Not for IRB submission

### Randomized, Embedded, Multifactorial Adaptive Platform trial for Community-Acquired Pneumonia (REMAP-CAP):

### PANDEMIC APPENDIX TO THE CORE PROTOCOL (REMAP-COVID)

REMAP-CAP Pandemic Appendix to the Core Protocol Version 2.0 dated 18 May 2020

Not for IRB Submission

**Summary**

**Background**: REMAP-CAP is an adaptive platform trial that evaluates multiple aspects of care of patients who are admitted to an Intensive Care Unit (ICU) with severe Community Acquired Pneumonia. It is reasonable to presume that any pandemic respiratory infection of major significance to public health will manifest as severe Community Acquired Pneumonia with concomitant admission to an ICU. Previous pandemics and more localized outbreaks of respiratory emerging infections have resulted in severe Community Acquired Pneumonia (CAP) and admission to an Intensive Care Unit[1-3]. Admission to an ICU may occur at the time of first presentation to a hospital or may be preceded by admission to a non-ICU ward or floor. For patients admitted to a non-ICU ward there is an opportunity to intervene to prevent the development of severe CAP. A pandemic of respiratory infection is much more likely to be caused by a virus than a bacterium. Differences in trial design may be required for influenza, viruses which are known to result in periodic but unpredictable pandemics, in comparison with other viruses, such as Coronaviruses that may also have pandemic potential.

Previous pandemics and outbreaks of emerging infectious diseases have outlined the urgent need for evidence, preferably from Randomized Clinical Trials, to guide best treatment. However, there are substantial challenges associated with being able to organize such trials when the time of onset of a pandemic and its exact nature are unpredictable[4-6]. As an adaptive platform trial that enrolls patients during the interpandemic period, REMAP-CAP is ideally positioned to adapt, in the event of a respiratory pandemic, to evaluate existing potential as well as novel treatment approaches.

The precise nature of a respiratory pandemic cannot be known in advance. The Pandemic Appendix to the Core Protocol lists potential adaptations to trial design and management that are generic, in that they will occur irrespective of the nature of the pandemic, as well as adaptations that are possible, depending on the nature of the pandemic, and the process for determining which adaptations will be applied.

The Pandemic Appendix to the Core Protocol also achieves alignment with a separate document, REMAP-COVID Core Protocol, which comprises only those elements of the Core Protocol of REMAP-CAP and the Pandemic Appendix that applies to the COVID-19 pandemic. For the COVID-19 pandemic, a site can utilize either the REMAP-CAP Core Protocol combined with the Pandemic Appendix to the Core Protocol, or REMAP-COVID Core Protocol. Both sets of documents specify identical methods and data requirements. Data derived from sites using either set of documents is

Not for IRB Submission

analyzed in the same pandemic statistical model. A single site must use either REMAP-COVID Core Protocol or REMAP-CAP Core Protocol with this associated pandemic appendix.

The objective of the Pandemic Appendix to the Core Protocol (PAtC) is to describe the adaptations to the Core Protocol that would apply during a pandemic, including how analyses of domains already operative during the interpandemic period as well as domains that are pandemic-specific, will be integrated during a pandemic. This includes scientific, as well as governance and logistic aspects.

**Aim**: The primary objective of the REMAP during a pandemic is to identify the effect of a range of interventions to improve outcome for patients admitted to a hospital with acute illness due to suspected or proven pandemic infection, as defined by the pandemic primary end-point.

**Methods:** The methods that will be utilized during a pandemic are those in the Core Protocol but with potential for changes to the primary end-point, frequency and process for adaptive analyses, and determination of which domains will be analyzed using a statistical model that includes data from patients with proven or suspected pandemic infection. During a pandemic, patients who are neither suspected nor proven to have pandemic infection and for certain pre-existing domains, will continue to be analyzed using the statistical model that is outlined in the Core Protocol that was operating during the pre-pandemic period. Depending on the characteristics of a pandemic, one or more interpandemic domains may be analyzed within the pandemic statistical model and one or more pandemic-specific domains may be commenced for patients with suspected or proven pandemic infection.

**Lay description**

REMAP-CAP is a global trial examining the best treatments for community-acquired pneumonia. In the setting of a pandemic that causes life-threatening respiratory infection, some key aspects of the study will be changed to integrate new interventions into the trial, evaluate existing interventions within the trial specifically in patients with pandemic infection, alter trial governance, and provide time-critical data for public health. This will allow the platform to identify which treatments work best for patients during a pandemic.

Not for IRB submission

#### TABLE OF FIGURES

Figure 1. The multifactorial structure of REMAP-CAP........................................................................ 15
Figure 2. Diagram of the interpandemic and pandemic models .......................................................... 21

Not for IRB Submission

#### 1. ABBREVIATIONS

| | |
|---|---|
| CAP | Community-Acquired Pneumonia |
| CRF | Case Report Form |
| DSA | Domain-Specific Appendix |
| DSMB | Data Safety and Monitoring Board |
| ICU | Intensive Care Unit |
| ISIG | International Statistics Interest Group |
| ITSC | International Trial Steering Committee |
| MERS-CoV | Middle-Eastern Respiratory Syndrome Coronavirus |
| NAI | Neuraminidase inhibitors |
| PAtC | Pandemic Appendix to the Core Protocol |
| PINSNP | Pandemic infection is neither suspected nor proven |
| PISOP | Pandemic infection is either suspected or proven |
| PWG | Pandemic Working Group |
| RAR | Response Adaptive Randomization |
| REMAP | Randomized, Embedded, Multifactorial, Adaptive Platform trial |
| REMAP-CAP | Randomized, Embedded, Multifactorial, Adaptive Platform trial for Community-Acquired Pneumonia |
| RCC | Regional Coordinating Center |
| RCT | Randomized Controlled Trial |
| RSA | Region Specific Appendix |
| SAC | Statistical Analysis Committee |
| SARS | Severe Acute Respiratory Syndrome |
| WHO | World Health Organization |

Not for IRB Submission

#### 2. PROTOCOL APPENDIX STRUCTURE

The structure of this protocol is different to that used for conventional trials because this trial is highly adaptive and the description of these adaptations is better understood and specified using a 'modular' protocol design. While all adaptations are pre-specified, the structure of the protocol is designed to allow the trial to evolve over time, for example by the introduction of new domains or interventions or both (see glossary, Section 1.2 Core Protocol for definitions of these terms) and commencement of the trial in new geographical regions.

The protocol has multiple modules, in brief, comprising a Core Protocol (overview and design features of the study), a Statistical Analysis Appendix (details of the current statistical analysis plan and models), multiple Domain-Specific Appendices (DSA) (detailing all interventions currently being studied in each domain), a Registry Appendix, this Pandemic Appendix to the Core Protocol, and multiple Regions-Specific Appendices (RSA) (detailing regional management and governance).

The Core Protocol contains all information that is generic to the trial, irrespective of the regional location in which the trial is conducted and the domains or interventions that are being tested. The Core Protocol may be amended but it is anticipated that such amendments will be infrequent.

The Core Protocol does not contain information about the intervention(s), within each domain, because one of the trial adaptations is that domains and interventions will change over time. Information about interventions, within each domain, is covered in a DSA. These Appendices are anticipated to change over time, with removal and addition of options within an existing domain, at one level, and removal and addition of entire domains, at another level. Each modification to a DSA will be subject of a separate ethics application for approval.

The Core Protocol does not contain detailed information about the statistical analysis, because the analysis model will change over time in accordance with the domain and intervention trial adaptations, but this information is contained in the Statistical Analysis Appendix. These Appendices are anticipated to change over time, as trial adaptations occur. Each modification will be subject to approval from the International Trial Steering Committee (ITSC) in conjunction with advice from the International Statistics Interest Group (ISIG) and the Data Safety and Monitoring Board (DSMB).

The Core Protocol also does not contain information that is specific to a particular region in which the trial is conducted, as the locations that participate in the trial are also anticipated to increase over time. Information that is specific to each region that conducts the trial is contained within an RSA. This includes information related to local management, governance, and ethical and regulatory

Not for IRB Submission

aspects. It is planned that, within each region, only that region's RSA, and any subsequent modifications, will be submitted for ethical review in that region.

### 3. PANDEMIC APPENDIX TO THE CORE PROTOCOL VERSION

The version of the Pandemic Appendix to the Core Protocol is in this document's header and on the cover page.

#### 3.1. Version History

Version 1: Approved by the Pandemic Working Group on 31st January, 2020

Version 1.1: Approved by the Pandemic Working Group on 12th February, 2020

Version 2.0: Approved by the Pandemic Working Group on 18th May, 2020

### 4. PANDEMIC APPENDIX TO THE CORE PROTOCOL GOVERNANCE

The study administration structure is outlined in the Core Protocol. As outlined in the Core Protocol, a Pandemic Working Group (PWG) is established and works in conjunction with the International Trial Steering Committee (ITSC), to take responsibility for the Pandemic Appendix to the Core Protocol (PAtC) and to advise on operational aspects following emergence of a pandemic.

#### 4.1. Pandemic Working Group

The responsibility of the PWG is to maintain and update this PAtC and to advise the ITSC regarding application of the PAtC during a pandemic. The PWG will liaise with individuals and organizations that are external to REMAP-CAP as required. Membership of the PWG is flexible. The core membership is listed but additional members can be added at any time and as required.

**Chair:** The Chair of the ITSC will Chair the Pandemic Working Group

**Members:**
Prof. Derek Angus
Prof. Yaseen Arabi
Prof. Richard Beasley
A/Prof. Scott Berry
Prof. Frank Brunkhorst
Dr. Lennie Derde

Not for IRB submission

Dr. Robert Fowler

Prof. Anthony Gordon

Mr. Cameron Green

Dr. Ed Litton

Prof. John Marshall

Dr. Colin McArthur

A/Prof Bryan McVerry

Dr. Srinivas Murthy

Prof. Alistair Nichol

Ms. Jane Parker

Prof. Kathy Rowan

Prof. Tim Uyeki

Prof. Steve Webb

##### 4.2. Contact Details

**Chair:** Professor Steve Webb

Department of Epidemiology and Preventive Medicine

School of Public Health and Preventive Medicine, Monash University

Level 3, 533 St Kilda Road

Melbourne, Victoria, 3004

AUSTRALIA

Not for IRB submission

### 5. PANDEMIC WORKING GROUP AUTHORISATION

The Pandemic Working Group have read the appendix and authorize it as the official Pandemic Appendix to the Core Protocol for the study entitled REMAP-CAP. Signed on behalf of the committee,

**Chair** 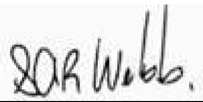 Date 18th May, 2020

Steve Webb

### 6. BACKGROUND AND RATIONALE

#### *6.1. Introduction*

It is reasonable to presume that any pandemic respiratory infection of major significance to public health will manifest as life-threatening respiratory infection including Severe Acute Respiratory illness and severe Community Acquired Pneumonia (CAP) with concomitant admission to hospital, and for some patients, admission to an Intensive Care Unit (ICU). Previous pandemics and more localized outbreaks of respiratory emerging infections have resulted in severe CAP and ICU admission[1-3]. A pandemic of respiratory infection is much more likely to be caused by a virus than a bacterium and, among viruses a distinction should be drawn between influenza, which is known to result in periodic but unpredictable pandemics, and other viruses, such as Coronaviruses, that may have pandemic potential, as the features of trial design may be different.

Previous pandemics and outbreaks of emerging infectious diseases have outlined the urgent need for evidence, preferably from Randomized Controlled Trials (RCTs), to guide best treatment. However, there are substantial challenges associated with being able to organize such trials when the time of onset of a pandemic and its exact nature are unpredictable[4-6]. As an adaptive platform trial that enrolls patients during the interpandemic period, REMAP-CAP is ideally positioned to adapt, in the event of a respiratory pandemic, to evaluate existing treatments as well as novel approaches.

One of the challenges associated with planning clinical trials during a pandemic is that the precise nature of the infecting organism, clinical consequences, and suitable interventions (particularly those that are pathogen-specific) cannot be reliably known in advance. The speed of clinical progression, from initial infection to life-threatening severe respiratory infection is another feature

Not for IRB Submission

that cannot be reliably known in advance. It is likely that a proportion of patients will present with severe CAP but other patients may present to medical attention with illness that is less severe, but remain at risk of progression to severe illness. Patients who require hospital admission, but have less severe illness are a particularly important group, because early intervention at this stage of illness may prevent progression to life-threatening illness. It is also possible that proposed treatment interventions may have differential treatment effect depending on the level of illness severity at the time that treatment is commenced, including treatment effects that are divergent. Nevertheless, a range of scenarios can be anticipated and used to provide direction and guidance regarding the most appropriate research response.

The most likely organism responsible for a respiratory pandemic is a novel influenza virus that has undergone antigenic shift[7]; the most recent influenza pandemic occurred during 2009-2010. In recent years, there have been outbreaks of severe Community Acquired Pneumonia due to novel Coronaviruses which resulted in the Severe Acute Respiratory Syndrome (SARS) outbreak in 2003 and the Middle-Eastern Respiratory Syndrome Coronavirus (MERS-CoV) outbreak that commenced in 2012. SARS-CoV-2 is the cause of a pandemic of severe respiratory disease (COVID-19), including pneumonia, that commenced in 2019. The pre-specified adaptations to REMAP-CAP will need to be different for influenza in comparison to a non-influenza pandemic pathogen.

#### 6.2. Pandemic research preparedness

##### 6.2.1. Introduction

The conceptual approach to pandemic preparedness has been influenced substantially by the occurrence of the 2009 Influenza A H1N1(2009)pdm pandemic, outbreaks of SARS and MERS-CoV, the Zika pandemic, and Ebola virus disease outbreaks in West Africa[8]. A broad conclusion from these outbreaks is that it is likely that high quality research can change the incidence and consequences of the epidemic but that such research is extremely difficult because planning of research only commences after the discovery of the epidemic. As a consequence, researchers and organizations interested in developing improved processes for research have identified three key elements to facilitate time-critical research about an epidemic. These elements are that the research must be pre-planned, pre-approved, and practiced[9,10]. REMAP-CAP and, in particular, the PAtC, is an attempt to establish these pre-requisites and to guide treatment for patients who may be critically ill with pneumonia as a consequence of infection with a pandemic organism.

The World Health Organization (WHO) has recommended establishing and strengthening outbreak-ready, multi-center clinical research networks in geographically diverse regions to facilitate research

Not for IRB Submission

during pandemics.[11] It has also recommended testing of protocols during interpandemic periods and stressed the value of such clinical research consortia in collecting and distributing information during a future pandemic.

##### 6.2.2. Pre-planned

Pre-planned means that the trial protocol is written and that the trial processes related to project management, screening, recruitment, delivery of interventions, data collection, data management, analysis, and reporting are all in place. The PAtC, in conjunction with the existing REMAP-CAP protocol documents and trial processes, will mean that all aspects that can be pre-planned have been.

##### 6.2.3. Pre-approved

The PAtC is a key component of the of the pre-approval strategy. The availability of this document allows ethics review boards, hospital research governance staff, existing and potential sites to understand and approve the study processes that would be implemented during a pandemic. Where different options need to exist, depending on the nature of the pandemic, these are pre-specified, as much as possible. Any unanticipated substantive deviation from this Appendix would be subject to an amendment, hopefully expedited, in the event of a pandemic. The PAtC, like the Core Protocol, does not specify any interventions that are evaluated within the REMAP. It is highly likely that one or more research questions (in domains already approved during the interpandemic period) will be relevant specifically in patients with severe respiratory disease including pneumonia caused by the pandemic infection. The PAtC allows these questions to continue to be answered specifically in patients with pandemic infection, where appropriate, using Bayesian prior probabilities derived from patients already enrolled during the interpandemic period. It is proposed to develop 'sleeping domains', which could be activated if appropriate during a pandemic, as well as retain the option of developing one or more completely new domains following the emergence of pandemic, which would require separate ethical approval and contracts with participating sites.

This strategy, as part of the study design, offers an ethically, clinically and legally acceptable mechanism for research in the context of a pandemic that can be initiated rapidly.

There are two further aspects relevant to ethical approval of the PAtC. The first is that existing or pandemic-specific domains of REMAP-CAP may include an intervention that specifies no treatment within that domain (noting that the Core Protocol specifies that all additional standard care is provided with treatment decisions being made by the treating clinician). This is clinically and

Not for IRB Submission

ethically appropriate as the response of critically ill patients to a range of different treatments has proven to be unpredictable. There are many examples of treatments that have resulted in harm[12] and situations in which surrogate outcome measures were not reliable indicators of improvement in patient-centered outcomes. As such, there should not be any presumption that it is better for patients to receive active interventions.

The second is the capacity to apply Response Adaptive Randomization (RAR) within the REMAP. As outlined in the Core Protocol, RAR results in an increasing proportion of patients being allocated to any intervention within a domain that has a higher probability of being superior with that proportion increasing as statistical confidence accrues. Participants within REMAP-CAP during a pandemic may be able to benefit from information about the relative effectiveness of interventions that is not in the public domain and not available to patients who are not participants in REMAP-CAP. As outlined in the Core Protocol, any intervention confirmed to be superior within the REMAP is then implemented by application of a RAR proportion that is equal to 100%. RAR will be implemented for pandemic patients as soon as sufficient data have accrued and operational implementation is feasible.

##### 6.2.4. Practiced

REMAP-CAP will be recruiting during the interpandemic period in multiple countries in both Southern and Northern Hemispheres with the support of several Regional Coordinating Centers. This research activity, during the interpandemic period, ensures that sites, site training, project management, data management, analysis processes, and trial governance are functional and practiced. Furthermore, the eligibility process and delivery of trial interventions are optimized for embedding which allows study processes to occur within minimal disruption to the delivery of clinical care, which may well be under substantial strain during a pandemic. There is already extensive experience with the Case Report Form (CRF) that is used and will continue to be used during a pandemic.

##### 6.2.5. Implications of REMAP design during a pandemic

###### *6.2.5.1. Time-critical generation of evidence*

A pandemic will likely result in a large number of affected persons with cases occurring over a short period of time, perhaps as short as a few months. Conventional clinical trials that utilize frequentist statistical techniques require a fixed sample size with limited capacity to analyze the results of the trial until recruitment is completed. The setting of the sample size requires an estimate of the size of the treatment effect and it is known that the assumptions that are made in setting the size of the

Not for IRB Submission

treatment effect are often incorrect[13,14]. A frequentist trial that over-estimates the size of the treatment effect may conclude without reaching a valid conclusion, whereas one that under-estimates the size of the treatment effect is delayed in providing time-critical information that the treatment is even more effective than estimated.

REMAP-CAP utilizes Bayesian statistical methods which allow frequent adaptive analyses to occur. This will ensure that time-critical information about the effectiveness of treatment interventions is not delayed unnecessarily. The REMAP design is particularly suited to pandemics because it requires no pre-trial assumptions about the size of the treatment effect and will allow dissemination of evidence as soon as possible. Furthermore, as the trial progresses during a pandemic the Data Safety Monitoring Board (DSMB) has access to information from adaptive analyses that may not achieve thresholds to allow reporting as a Platform Conclusion but may be relevant to public health which, under appropriate circumstances, can be shared with public health authorities and the DSMBs of other trials evaluating the same or similar interventions without threatening the scientific validity of the ongoing trial.

###### 6.2.5.2. *Multifactorial design and evaluation of interactions*

If there are multiple interventions, each of which may have independent effects on outcome, the multi-factorial nature of a REMAP allows these to be evaluated simultaneously, rather than in series or in separate parallel trials (see *Figure 1*). This design feature contributes to efficiency and is also anticipated to result in more clinical evidence being generated more rapidly during a time-critical pandemic.

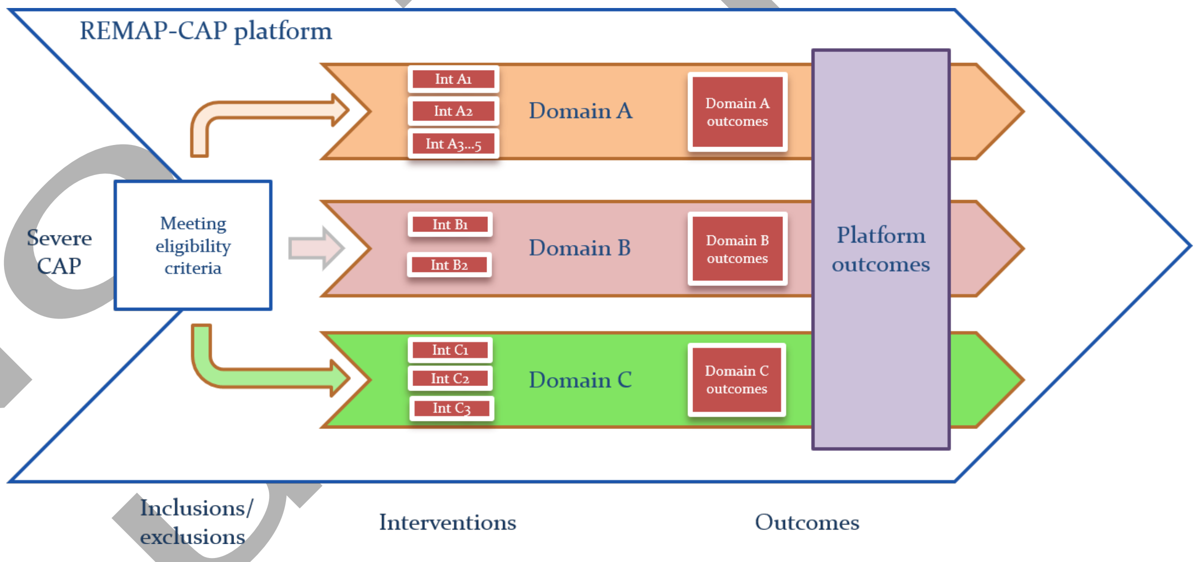

*Figure 1. The multifactorial structure of REMAP-CAP*

Not for IRB Submission

Furthermore, where pre-specified, the statistical model utilized in REMAP-CAP will allow estimation of treatment effect of interventions that may be contingent on other treatment assignments within the pandemic component of the REMAP. For example, it is plausible that the effectiveness of an intervention for immune modulation is dependent on co-delivery of an agent that is effective at inhibiting growth or replication of the pathogen. Conventional trials, in which only a single domain of treatment is evaluated, are not capable of detecting this type of treatment-by-treatment interaction, and thereby unable to identify the best overall treatment strategy for these patients.

###### 6.2.6. Setting of research priorities

In 2017, the WHO outlined the research priorities for a pandemic that was caused by a novel strain of influenza. These priorities were:

- Research on the effectiveness of empirical treatment with oseltamivir and other neuraminidase inhibitors (NAI) in critically ill patients, including placebo-controlled trials during seasonal as well as pandemic influenza.
- Investigating alternative strategies to NAI monotherapy to increase antiviral potency and improve clinical outcomes.
- Research on immune-modulatory strategies in severe influenza, including corticosteroids and macrolides.
- A need for high quality data on the effectiveness of most aspects of supportive care related to influenza.
- A need to assess the roles of virologic factors (e.g. replication sites, duration and viral load levels) in larger numbers of patients (including critically ill patients) in causing severe disease and associated complications, linking them to clinical outcomes.

REMAP-CAP is not able to meet all of these requirements but is well suited to evaluate the effectiveness of antiviral therapies active against influenza, immune modulatory strategies and different aspects of supportive care[15]. Identical or similar research questions would exist for any pandemic caused by an organism that was not influenza and REMAP-CAP has also similar capabilities in this scenario.

##### *6.3. WHO endorsement*

REMAP-CAP has been designated by the WHO as a Pandemic Special Study. Under this designation, it has been tasked with helping answer crucial questions during a declared pandemic, as listed above. This designation ensures that knowledge translation of clinical trial results can occur directly

Not for IRB Submission

with policymakers and public health officials for rapid implementation around the globe. It ensures that results generated from REMAP-CAP during a declared pandemic can be translated in an efficient and transparent manner to benefit affected patients.

### 7. ADAPTATION OF REMAP-CAP DURING A PANDEMIC

This PAtC supplements the Core Protocol during a pandemic including deactivation at the conclusion of a pandemic. Decisions regarding the operationalization of the Pandemic Appendix to the Core Protocol are made by the ITSC with advice from the PWG (see Section 8.1). The Appendix sets out all potential adaptations of the Core Protocol and unless otherwise specified, all other aspects of the Core Protocol remain active. Activation of the PAtC will be advised to the DSMB with specification of the selected operational characteristics.

#### *7.1. Objectives*

The primary objective of this REMAP during a pandemic is to identify the effect of a range of interventions to improve outcome for adult patients admitted to hospital with acute illness due to suspected or proven pandemic infection, as defined by the pandemic primary end-point.

The secondary objective is to determine the effect of a range of interventions on additional endpoints, including endpoints developed by the World Health Organization and adopted core outcome sets.

#### *7.2. Study setting: definition of an ICU and relationship of setting to severity of illness*

During the interpandemic period, the REMAP recruits only participants who are admitted to an ICU, and a combination of admission to ICU as well as provision of treatments to support failed organs is used to define severity and eligibility. During a pandemic, there are several factors that may influence the relationship between admission to an ICU and severity of illness. Firstly, there may be insufficient ICU beds available to care for all critically ill patients. This may result in provision of advanced organ support occurring in locations that do not usually provide ICU-level care. During a pandemic, such a location is referred to as a re-purposed ICU. However, a re-purposed ICU needs to be distinguished from a usual hospital ward that is capable of providing some forms of organ support, such as non-invasive ventilation. During a pandemic, there may be substantial delays in transferring a patient from an emergency department to either a ward or an ICU (or a re-purposed ICU). Patients in an emergency department who have been accepted for admission to an ICU are regarded as being admitted to an ICU. Patients in an emergency department who have been

Not for IRB Submission

accepted for admission to a ward are regarded as being admitted to a ward. Secondly, patients who are not critically ill may be treated on an ICU for reasons that are not related to severity of illness, such as access to single rooms to achieve objectives related to infection control and prevention. This can influence both admission as well as discharge practices. Thirdly, the threshold at which support for failed organs is provided may be influenced by infection control practices. For example, some forms of respiratory support may be withheld because of concerns related to the risk that staff who are caring for patients may acquire the infection.

To minimize these issues, during a pandemic, the primary determinant of severity is the provision of ICU-level care, which can be interpreted in conjunction with the physical location in which care is being provided. Determination of severity may also take into account a decision to withhold some form of organ failure support that would otherwise have be provided. Where a definition of an ICU is needed, at sites at which the pandemic stratum (see below) has been activated, an area within the hospital that is repurposed so as to be able to deliver one or more of the qualifying organ failure supports specified in the Core Protocol (non-invasive ventilation, invasive ventilation, and vasopressor therapy) will meet the definition of an Intensive Care Unit. It is preferred in such circumstances that the patient is under the care of a specialist who is trained in the provision of critical care, but this is not an essential requirement. A respiratory or other ward that provides non-invasive ventilation (including oxygen therapy delivered by high flow nasal cannula) and continues to do so during a pandemic, will not, generally, meet the definition of an ICU, particularly if the patient is not under the care of a specialist who is trained in the provision of critical care.

In some DSAs, an exclusion criteria is applied to only permit enrollment during a time-window that commences with ICU admission. For the reasons noted above, this may be operationalized using a time-window, of the same duration, that commences with the provision of sustained organ failure support.

#### *7.3. Eligibility criteria*

Platform-level eligibility criteria may be modified if necessary to accommodate a published case definition, to align with criteria specified in guidelines, such as the ATS/IDSA guidelines on CAP[16], or to accommodate necessary modifications to the online eligibility system used for enrollment. In previous epidemics of community-based infection, nosocomial acquisition has been well described. Relaxation of the requirement for community acquisition or organ failure criteria or both may be appropriate. In this regard, Version 2.0 of this Appendix modifies the organ failure support criteria so that these no longer apply as a platform-level inclusion criteria, permitting the enrollment of

Not for IRB submission

patients into the platform who are admitted to hospital or an ICU, either with or without organ failure support criteria. In association with the removal of the organ failure requirement, the requirement for a patient to meet criteria for pneumonia may be replaced with a requirement for acute illness due to suspected or proven pandemic infection. All changes to eligibility criteria would apply only to patients in the pandemic stratum (see section 7.4).

As such, the modified platform-level inclusion and exclusion criteria are:

In order to be eligible to participate in the pandemic aspects of REMAP-CAP, a patient must meet the following criteria:

1. Adult patient admitted to hospital with acute illness due to suspected or proven pandemic infection

A potentially eligible patient who meets any of the following criteria will be excluded from participation in this trial:

1. Death is deemed to be imminent and inevitable during the next 24 hours AND one or more of the patient, substitute decision maker or attending physician are not committed to full active treatment
2. Patient is expected to be discharged from hospital today or tomorrow
3. More than 14 days have elapsed while admitted to hospital with symptoms of an acute illness due to suspected or proven pandemic infection
4. Previous participation in this REMAP within the last 90 days

This extension of the platform-level inclusion criteria can apply to patients admitted to an ICU or a ward. In association with the involvement of different clinical teams, the domains and interventions that are available for patients admitted to a ward compared with those admitted to an ICU are permitted to be, but do not have to be, different.

#### 7.4. Pandemic stratum

##### 7.4.1. Introduction

As outlined in the Core Protocol, a pre-specified stratum of the REMAP is the presence or absence of suspected or proven pandemic infection. This is maintained as a ‘passive stratum’ during the interpandemic period that can become active during a pandemic. It consists of two exclusive strata categories: pandemic infection is neither suspected nor proven (PINSNP) and pandemic infection is

Not for IRB Submission

either suspected or proven (PISOP) at baseline. At times when the PAtC is not activated, i.e. during the interpandemic period, all participants are categorized as PINSNP.

###### 7.4.2. Activation and deactivation of the PAtC and PISOP stratum

In response to a pandemic (see section 8.1), the PISOP stratum is activated using a two-step process. First there is a decision of the ITSC to open the PISOP stratum for the platform. The second step is site-by-site activation of the PISOP stratum, requiring agreement of both the site and the Regional Coordinating Centre (RCC). This allows variation in activity of the pandemic infection to be accommodated with sites only open for PISOP recruitment when there is active pandemic infection locally. Switching-on of the stratum can occur at any time and expected to always be available with less than 24 hours lead time. The capacity to enroll patients into the PISOP stratum can be switched-off on a site-by-site basis, but the ITSC can switch off the PISOP stratum for all sites if it is believed that a pandemic is no longer ongoing. The REMAP applies a new and separate statistical model for participants in the PISOP stratum which can utilize, where appropriate, informative priors derived from pre-pandemic PINSNP participants.

It should be noted that for sites in which the pandemic stratum is open, that the REMAP allows for continued recruitment of patients into the REMAP who are in the PINSNP stratum. For example, during an influenza pandemic, PINSNP would include patients with infection that has been proven to be a non-pandemic strain of influenza. During a pandemic, patients who are in the PINSNP stratum continue to be analyzed using the interpandemic statistical model (see below). As such, there are two categories of PINSNP participants- those included during the interpandemic phase and those included during a pandemic. Both categories of patients contribute to the interpandemic model for all active domains.

The PAtC is activated and deactivated for a site at the same time as the PISOP stratum is opened and closed. If a pandemic commences prior to ethical and governance approval of the PAtC, the PISOP stratum can be activated using approvals for the Core Protocol, and the PAtC would be activated as soon as ethical approval is obtained.

##### 7.5. *The pandemic statistical model*

###### 7.5.1. Introduction

The model that is active during the pandemic and includes only PISOP patients (for some or all domains) is referred to as the ***pandemic model***. The model that is active before (and after) the

Not for IRB Submission

pandemic, which includes PINSNP patients during the pandemic and may include some PISOP patients for some domains, is referred to as the ***interpandemic model*** (see *Figure 2*).

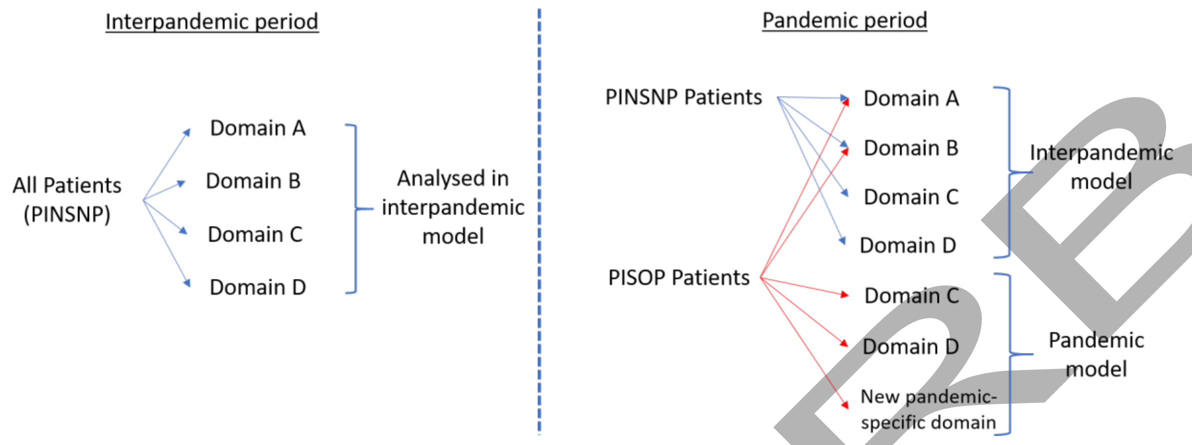

*Figure 2. Diagram of the interpandemic and pandemic models*

The pandemic model is only used for PISOP participants and only for those domains selected by the ITSC. A PISOP patient can contribute to both the pandemic and interpandemic model in different domains but each patient's contribution to a model is mutually exclusive with respect to each domain. The ITSC will select the domains to be included in the pandemic model where a differential treatment effect is postulated in the presence of pandemic infection or the need exists to learn about the outcome quickly, or both. The extension of this platform-level entry criteria does not apply to domains that are analyzed exclusively within the interpandemic statistical model.

A consequence of the application of two separate statistical models is that treatment-by-treatment interactions can only be evaluated for those domains that are in the same model. The principal advantages of the use of two models are:

- that this is necessary where the pandemic model requires a different primary end-point
- the platform is able to continue recruitment of patients with CAP who are neither suspected nor proven to have pandemic infection
- where appropriate informative priors can be included at commencement of the pandemic model
- where appropriate thresholds for a Statistical Trigger can be modified
- only those domains that are relevant to the pandemic are included within the pandemic model.

During the interpandemic period, it is intended that there may be some domains, for example the Ventilation Domain, that will utilize a separate domain-specific statistical model. It should be noted

Not for IRB Submission

that during the interpandemic period, such a domain is not part of the interpandemic statistical model. During a pandemic any such domain would continue to be evaluated with its own domain-specific statistical model. During a pandemic, the operating characteristics of the domain-specific statistical model may be modified in the same way that the pandemic model is modified from the interpandemic model. For example, PISOP patients may be analyzed within a pandemic version of the domain specific statistical model utilizing a modified primary end-point, with application of informative priors derived from the interpandemic time period.

##### 7.5.2. Pre-specification of trial parameter options

There are many clinical features of a respiratory pandemic that cannot be predicted in advance. For several parameters related to trial design and statistical analysis, this Appendix pre-specifies a range of options from which the actual modifications will be chosen at the commencement of a pandemic. The appendix provides guidance regarding the principles that would guide selection from within the available options and often provides the planned default option. The provision of flexibility regarding limited aspects of trial design provides the capacity to tailor aspects of the trial to the characteristics of the pandemic. For these decisions, the ITSC has decision-making responsibility, with advice from the PWG. These decisions would be regarded as operational and, unless otherwise specified (5.3.4), will be made prior to the conduct of the first adaptive analysis using the pandemic model and would be made only from within the range of options pre-specified in this Appendix. It is not intended that the selected parameters would be modified in any way during the pandemic unless advised to do so by the DSMB. The selected trial parameters would be placed in the public domain, on the study website, and provided as an update to participating sites and relevant ethical review bodies prior to the first adaptive analysis of the PISOP stratum. These parameters are set out in a document termed Operating Characteristics and this document applies to both REMAP-CAP core protocol documents as well as the REMAP-COVID Core Protocol, to the extent that is necessary. It is also acknowledged that specification in a new domain, may influence a pre-existing domain, such as specification of evaluation of an interaction between domains. In this situation, the DSA for the pre-existing domain will not necessarily be amended immediately with the most recently approved or amended DSA serving to specify the inter-relationship between the two domains.

##### 7.5.3. Application of other strata specified in the Core Protocol in the pandemic model

The shock strata may be applied to the PISOP stratum. The default position is that the shock strata will not be applied to the PISOP stratum.

Not for IRB submission

If the pandemic is caused by a novel strain of influenza the pre-existing influenza strata is not applied in the pandemic model. For PINSNP patients, the "influenza present" stratum would continue to apply and would be used to differentiate patients infected with a non-pandemic strain of influenza from patients in the "influenza not present" stratum. Membership of PISOP and influenza present stratum are mutually exclusive. It is anticipated that the influenza present stratum would apply only to patients with infection due to a proven non-pandemic strain of influenza at baseline. Patients in whom influenza was suspected, but the results of strain-specific diagnostic tests were not available at the time of assessment of eligibility, will be allocated to the PISOP stratum at sites where the stratum is active.

##### 7.5.4. Strata within the PISOP stratum

A strata applied within the PISOP stratum is the confirmation status of pandemic infection, defined in two categories, present or absent, based on the results of microbiological tests for the pandemic organism. Any patient with clinically suspected pandemic infection who is not tested or the result is not yet known will be deemed positive.

The availability and interpretation of microbiological tests are likely to change during the pandemic and an operational document will be used to specify how different tests are interpreted. It is noted that pandemic infection confirmed status is defined by the final results of testing for the pandemic organism which may include analysis of samples collected after enrollment where it is reasonable to presume that the sample reflected pandemic infection status at time of enrollment.

The sensitivity of microbiological testing for the pandemic organism may not be known at the beginning or even during the pandemic[17]. It is anticipated that initial analysis of the pandemic model will occur without application of this pandemic confirmation status strata but this would be applied when there was sufficient confidence about the operating characteristics of diagnostic tests in patients who are critically ill. If the pandemic confirmation status is applied, the probabilities derived from patients who have confirmed pandemic infection will be used to determine the RAR proportions for patients receiving treatment assignments in the pandemic specific domains within the PISOP stratum. Borrowing is permitted between the pandemic infection confirmed stratum and the pandemic infection not present stratum, using the methods outlined in the Core Protocol (with gamma = 0.15).

If eligibility criteria were modified to allow inclusion of a wider spectrum of illness severity, two or more states, related to severity of illness, may be applied within the PISOP stratum to distinguish current versus extended severity of illness.

Not for IRB Submission

##### 7.5.5. States within the PISOP stratum

The Core Protocol defines 'state' as a set of mutually exclusive and exhaustive categories, defined by characteristics of a patient within the REMAP, that are capable of changing over time for a single patient at different time-points during the patient's participation in the REMAP (i.e. they can be dynamic). During the pandemic, and only for patients in the PISOP stratum, two or more states may be defined, depending on illness severity. The default categorization of severity will be into two categories:

- Severe State, defined by receiving organ failure support in an ICU
- Moderate State, defined by
    - Not being admitted to an ICU, or
    - Admitted to an ICU but not receiving organ failure support

Organ failure supports that qualify a patient as severe are aligned to those that previously determined eligibility to the platform (i.e. the Severe State corresponds to the previous platform eligibility criteria). These criteria are:

- Provision of invasive mechanical ventilation
- Provision of non-invasive mechanical ventilation (including high flow nasal cannula with a flow rate of at least 30 litres per minutes and a fractional inspired oxygen concentration of 40% or higher)
- Receiving infusion of vasopressor or inotropes or both

Where states are defined, eligibility for domains or selected interventions within a domain, may be specified according to state. As such, a domain may be available in one or more states. Where a domain is available in two or more states, the interventions available in that domain in each state are permitted to vary. States can also be utilized within the statistical model to define the unit-of-analysis, with declaration of Platform Conclusions, independently in one or more states, with borrowing permitted between states.

A single patient can move between states, one or more times, during a period of time which the patient is potentially eligible within the REMAP. For the purposes of assessment of eligibility for one or more domains, state is 'instantaneous' as at the time of that assessment. A patient who has previously received non-invasive ventilation or an infusion of vasopressor or inotrope or both, but is not receiving either of those therapies at time of assessment is deemed to be in the Moderate State. A patient who has been in the Severe State, as a consequence of receiving invasive mechanical

Not for IRB Submission

ventilation in an ICU, cannot re-enter the Moderate State for the purposes of assessment of eligibility. A patient who receives an assignment in the REMAP while in the Severe State cannot receive any subsequent assignments in the Moderate State. Where trial related processes, such as reveal of assignment or obtaining consent, create a time gap between initial assessment of eligibility and awareness of the patient's assignment, the state in which the patient is analyzed is that which occurred at the time of assessment, not the time of reveal of the assignment.

A patient enrolled while in the Moderate State, if reassessed for eligibility for additional domains having progressed to the Severe State, may have new microbiological information that has accumulated during this interval of time. This could result in a patient with suspected pandemic infection having information that results in pandemic infection being excluded, at the time of reassessment. In this situation, the patient is analyzed in the pandemic model, as enrolled, in the Moderate State and is not eligible for enrollment in new domains in the Severe State (including domains evaluated in the interpandemic model). It is also noted that, for a patient who is enrolled in both states, that other time-varying basline variables may have changed between each enrolment. For such aptients, potentially time-varying baseline variables will be collected in reference to enrolment in the Moderate State and again in reference to enrolment in the Severe State.

##### 7.5.6. Domains incorporated in the pandemic model and use of informative priors derived from the interpandemic model

The domains that will be included within the pandemic model will be determined at the onset of a pandemic by the ITSC with advice from the PWG. Where appropriate and prior to the first adaptive analysis that is undertaken after activation of the PAtC, informative priors, derived from the interpandemic model (comprising patients enrolled in the REMAP prior to the pandemic), may be applied. If informative priors are applied, this is done by the Statistical Analysis Committee (SAC) who review the frequent adaptive analyses (and communicate these results to the DSMB on a regular basis). This will occur without knowledge of the values of the priors by the ITSC or any other investigator. The amount of influence that priors apply and how quickly priors are applied in combination with accruing new data will be specified by the ITSC. Coding that specifies the weighting of priors will be done by statisticians who are separate to the SAC and blind to results from adaptive analyses. With regard to selection of domains and the use of informative priors, the following principles will be applied.

###### *7.5.6.1. Non-influenza pandemic organism*

If the pandemic organism is not influenza, the following domains are intended to be included within the pandemic model:

Not for IRB submission

- Corticosteroid Domain, without application of informative priors.
- Macrolide Duration Domain, without application of informative priors.
- New domains, as appropriate for the pandemic organism, without application of informative priors.

The Influenza Antiviral Domain (which includes only antiviral agents active against influenza) would not be applied in the pandemic model. It is noted that a patient at baseline could have suspected influenza and suspected pandemic infection which could lead to enrollment in the influenza domain (evaluated in the interpandemic model) and enrollment in other domains (evaluated in the pandemic model). It is not anticipated that the Antibiotic Domain is evaluated in the pandemic model, though this may be revised if the pandemic was caused by a bacterial pathogen. In this situation only those antibiotics that are known to be active against the pandemic organism would be available within the Antibiotic Domain for patients in the PISOP stratum.

###### *7.5.6.2. Influenza pandemic*

If the pandemic organism is influenza, the following domains are intended to be included within the pandemic model:

- Corticosteroid Domain, using informative priors derived from the influenza present stratum.
- Antiviral domain, using informative priors derived from the influenza positive stratum but with exclusion of any antiviral interventions that are clinically inappropriate because of the resistance profile of the pandemic strain of influenza. If there were no antiviral agents to which the pandemic strain of influenza was susceptible the Antiviral domain would not be applied in the PISOP stratum. During the pandemic if the pandemic strain of influenza acquired resistance to antiviral agents in the Antiviral Domain, these agents would be withdrawn from the domain at affected sites.
- Macrolide Duration Domain using informative priors derived from the unit-of-analysis of the Macrolide Duration Domain in the interpandemic model.
- New domains, as appropriate, without application of informative priors.

A number of other domains, related to organ failure support may be operative at the time of a pandemic. Domains such as oxygen saturation and hemodynamic targets would be expected to remain active during a pandemic. The default plan is that during a pandemic, patients in the PISOP and PINSNP strata will be eligible to receive an assignment in these domains and will be analyzed in the interpandemic model which will continue to be analyzed for statistical triggers and platform conclusions. Patients with pandemic infection will have their treatment assignments in such domains

Not for IRB Submission

weighted according to RAR as specified by the interpandemic model which will continue to be updated during a pandemic.

The ventilation domain, which utilizes a statistical model that applies only to that domain, is expected to continue during a pandemic. If appropriate, the pandemic strata may be applied to this domain. If so, the PISOP stratum would apply informative priors.

Any new domain that is initiated during a pandemic will be submitted for ethical review and require ethical approval prior to commencement.

###### 7.5.7. Use of informative priors derived from information available from outside the REMAP

The default position is that informative priors derived from information that is external to the REMAP will not be utilized. However, if appropriate, based on high quality evidence, informative priors may be applied. The decision to apply informative priors lies with the ITSC and must involve consultation with relevant external stakeholders, the DSMB, and appropriate statistical advice regarding the potential implications for the use of informative priors.

##### 7.6. Endpoints

###### 7.6.1. Pandemic primary endpoint

Specified domains, for patients in the PISOP stratum, will be analyzed using a separate statistical model, for which the primary endpoint is called the "pandemic primary endpoint". The default pandemic primary endpoint will be an ordinal scale that is a composite end-point that comprises mortality during the acute hospital admission and the number of whole and part study days for which the patient is alive and not requiring organ failure support while admitted to an ICU up until the end of study day 21. All patients who die before discharge from an acute hospital, irrespective of whether this occurs before or after D21, will be coded as –1 day. All patients who never receive organ failure support while admitted to an ICU will be coded as 22. Patients who die between D21 and discharge from an acute hospital will be updated at the time of the next adaptive analysis. All whole and part days after discharge from an acute hospital and before D21 will be counted as being not admitted to an ICU. Hospital readmission that included a new admission to ICU between first discharge from an acute hospital and D21 will not contribute to the primary end-point.

If appropriate, based on an understanding of clinical and biological factors, as well as operational factors, an alternative pandemic primary end-point may be specified at the time of activation of the PAtC or at any time prior to the first interim analysis using the pandemic statistical model. Other

Not for IRB Submission

possible primary end-points include days alive and outside the ICU with alternative durations of follow up or the use of an alternative composite based on admission to ICU. The pandemic primary endpoint will be used for the adaptive analyses that inform the RAR and for Statistical Triggers.

If the primary end-point includes a time-based outcome measure, assignment to one or more domains will occur at different time-points if the patient receives assignments in one or more domains while in the Moderate State and one or more domains in the Severe State. The commencement of the period of observation commences at the time of assignment, which can lead to the same patient having different values for different domains, as determined by the state in which enrollment occurred. This can be accommodated because there are separate statistical models for each state. Where a patient is eligible for two or more domains in a state, assignment can only occur at a single time-point, i.e. it is not possible to have more than one time of assignment for different domains in the same state.

##### 7.6.2. Secondary endpoints

All secondary endpoints that are specified in the Core Protocol and active DSAs will continue to be active. The primary end-point specified in the Core Protocol (all-cause mortality at day 90) is a secondary end-point in the PISOP stratum.

#### 7.7. Principles of the statistical analysis

##### 7.7.1. Adaptive analyses

Adaptive analyses may be conducted more frequently and with varying cadence during a pandemic. For analyses conducted in the pandemic model and the PISOP stratum of the ventilation model, data from all available patients will be utilized using, where appropriate, modelling to impute missing data. Adaptive analyses may be conducted at different frequency for the PISOP and PINSNP stratum.

##### 7.7.2. Response adaptive randomization

For PISOP patients, RAR proportions for domains that are analyzed using the pandemic model will be derived from the pandemic model and the RAR proportions for domains that are analyzed using the interpandemic model will be derived from the interpandemic model. For PINSNP patients, the RAR proportions for all qualifying domains will be derived from the interpandemic model.

If feasible, the option of allowing sites to start with imbalanced RAR proportions may be utilized. During a pandemic, issues related to equipoise for sites to participate may be facilitated by allowing sites to select from a range of starting RAR proportions that are imbalanced. Being able to

Not for IRB Submission

implement this would be dependent on logistic feasibility as well as evaluation to exclude any adverse impact on inference.

Within the PISOP stratum, and only for domains with five or more interventions, the minimum RAR proportion may be decreased to less than 10% but will not be decreased to less than 5%.

##### 7.7.3. Unit-of-analysis

###### *7.7.3.1. Application of additional strata*

Patients within the PISOP stratum may be further stratified dependent on whether pandemic infection is confirmed or not confirmed by microbiological testing. Additional strata may be applied and this can be specified in a DSA. Any or all of these strata can be utilized to determine eligibility for a domain or an intervention within a domain. These strata can also be used to define a unit-of-analysis in the pandemic statistical model.

###### *7.7.3.2. Application of state*

The state, at time of first enrollment, can also be used to determine eligibility or be used to define a unit-of-analysis or both. Where specified in the statistical model, the treatment effect of an intervention is allowed to vary between different states. A Bayesian Hierarchical Model (BHM) is used for all treatment-by-state interactions. In the BHM a hyperprior is used for the differing treatment effects across states. The standard deviation of the hyperprior, gamma, is a modelling starting estimate for the variation in the magnitude of the difference in treatment effects between states. By default, the starting estimate of the difference is zero. The gamma parameter influences the extent to which the treatment effect of interventions is permitted to vary between states. At the commencement of a model, the gamma parameter must be set, for each domain-state pair.

In this REMAP, only three options are permitted with respect to specifying the gamma parameter for each domain-state pair. Firstly, gamma may be set to zero. The effect of this is that treatment effect of an intervention is assumed proportional between specified states. The unit-of-analysis is not sub-divided according to state. If gamma is set to zero for all states for a domain, the unit of analysis is all patients randomized in that domain. Secondly, and at the opposite extreme, gamma can be set to infinity. In this situation treatment effect is evaluated separately and independently in each state (with no borrowing between states). Thirdly, gamma may be set to a defined number between zero and infinity. This parameter value cannot be varied for different domain-state pairs, a global REMAP value has been selected. This specified value for gamma places a constraint on the variance of the difference in treatment effect in different states but permits the model to estimate treatment effect for patients enrolled in one state by borrowing from patients enrolled in one or more adjacent

Not for IRB Submission

states. Borrowing occurs to the extent that it is supported by the accumulated data, but the setting of gamma influences the amount of borrowing and how quickly borrowing is able to occur. The value of gamma that has been chosen has been determined by simulations to achieve a compromise between type I and type II error in baseline scenarios that assume either equivalence or superiority. Where a value for gamma is specified in the pandemic statistical model, in this REMAP the value of gamma will be 0.15.

A patient who is enrolled in a defined state, may have a clinical course that evolves with the patient entering a new state. Progression from one state, to another, may trigger eligibility for one or more domains. Where this occurs and the change in state defines a new unit-of-analysis, the RAR proportions may be different in each state. In this situation the RAR proportions that are relevant to that patient's state will be applied. In this regard, randomization to one or more domains in an initial state will occur, using RAR proportions that apply to that state with a separate subsequent randomization to one or more domains occurring if the patient enters a new state, with RAR proportions that apply to that state. When a new state commences there may be insufficient patients to determine valid RAR proportions for that domain in the new state. In this situation either RAR proportions are balanced or RAR proportions from an adjacent state are applied (unless otherwise specified in a DSA).

The RAR proportions that apply when state is used to define a unit-of-analysis are derived from all patients who receive an assignment in a domain in that state, irrespective of whether the patient was assigned an intervention in a different domain in a different state.

###### *7.7.3.3. Analyses for combinations of therapies*

Unless otherwise specified in a DSA, a Platform Conclusion can be reached for combinations of treatments that are being evaluated within the platform. This applies to interventions within a domain as well as interventions in different domains. As such, all of the following can be reported as Platform Conclusions: an interaction between interventions in different domains and that the treatment effect of more than one active intervention is different to a no treatment (standard of care) intervention. A domain that contains two or more treatments, each of which is assigned against a no treatment control in a factorial manner (i.e. the N x N table of yes / no for n treatments) will be analyzed as an N x N factorial. Structuring the analysis in this way allows the model to learn more quickly about the effectiveness of each treatment, recognizing common treatment exposure across intervention assignments.

Not for IRB Submission

##### 7.7.4. Thresholds for statistical triggers

###### *7.7.4.1. Introduction*

The Core Protocol specifies thresholds for Statistical Triggers that apply to superiority, inferiority, and equivalence. For PISOP patients, different thresholds for Statistical Triggers may apply during a pandemic. The decision to modify a statistical threshold will be made by the ITSC prior to the first adaptive analysis of the pandemic model. Different thresholds may be applied to different domains. Thresholds can also be specified that are asymmetric for example less stringent for inferiority than superiority. Factors that the ITSC will take into account in considering whether to modify a threshold include whether the interventions being evaluated are comparative effectiveness options (i.e. interventions that are available as part of standard care and available outside the platform) or experimental interventions with uncertain safety and risk profile that may be available only within the platform.

All decisions regarding thresholds for Statistical Triggers will be communicated to participating sites and placed in the public domain on the study website. Once specified, thresholds cannot be modified unless recommended by the DSMB.

The default thresholds are outlined in the following sections.

###### *7.7.4.2. Intervention Superiority Statistical Trigger*

At any adaptive analysis, if a single intervention has at least a 0.99 posterior probability of being a member of the optimal regimen, for that unit-of-analysis, then that intervention will be deemed as being superior to all other interventions in that domain in that target population.

The declaration of a Platform Conclusion by the DSMB for superiority will result in application of 100% RAR (see section 7.6.4). Following implementation of 100% RAR, the posterior probability will continue to be updated and evaluated by the DSMB who are empowered to act if they have concerns regarding the validity of a Platform Conclusion.

###### *7.7.4.3. Intervention Efficacy Statistical Trigger*

For any domain that has (or had) a non-active control intervention, statistical triggers for efficacy of other interventions can be determined. At any adaptive analysis, if a single intervention has at least a 0.99 posterior probability of being superior to the inactive control intervention, for that unit-of-analysis, then that intervention will be deemed as being effective in that domain in that target population. At any adaptive analysis, if a single intervention has a greater than 90% probability of

Not for IRB Submission

being harmful, compared to an inactive control intervention, for that unit-of-analysis, then the intervention will be deemed as being harmful in that domain in that target population.

The declaration of a Platform Conclusion by the DSMB for efficacy may not result in any actions and may occur after the non-active intervention has been removed. This Platform Conclusion mathematically would occur simultaneously to Superiority in a 2-intervention domain. If a determination of efficacy for an intervention with a currently randomized non-active control then the non-active control should be dropped and the RAR set to 0. In contrast, declaration of a Platform Conclusion for harm will result in removal of that intervention from the platform for that unit-of-analysis, together with Public Disclosure.

###### 7.7.4.4. Intervention Inferiority Statistical Trigger

At any adaptive analysis, if a single intervention has less than a 0.01 posterior probability of being a member of the optimal regimen, for a unit-of-analysis, then that intervention will be deemed as being inferior to other interventions in the domain for that target population. The 0.01 threshold is reduced as a function of how many units-of-analysis are available for the inferiority calculation (divided by the number of units minus 1). An asymmetrical inferiority statistical trigger may be set, particularly if an active intervention was being evaluated against an intervention that specifies no active treatment in that domain.

###### 7.7.4.5. Equivalence and futility

The equivalence boundary (delta) for different endpoints selected for the PISOP stratum may be changed depending on the clinical impact of the delta for the chosen endpoint. The default delta for the Core Protocol will be used to select clinically similar effects on the chosen primary endpoint. If a mortality or 21-day ICU- or organ support-free day endpoint is selected the 20% proportional odds equivalency delta will be the default.

Alternatively, a DSA may specify a futility boundary (delta) with respect to the primary end-point. For the pandemic primary end-point, the default futility boundary for an intervention will be set as a posterior probability of less than 0.05 for at least a 20% odds-ratio improvement. This rule corresponds to the one-sided equivalency region.

###### 7.7.4.6. Statistical thresholds for early phase interventions

During the pandemic there may be need to test multiple candidate interventions that are at an early phase of development, identifying those interventions that are most promising to be retained within the platform. Such interventions may be evaluated after a fixed recruitment against a ‘stop-go’

Not for IRB Submission

criteria for retention, and expansion, within the platform. The default threshold for retention and expansion of an intervention will be a posterior probability of 0.5 or more that there is at least a 30% benefit in odds ratio.

###### 7.7.5. Actions when a Statistical Trigger is achieved

The actions that occur when a statistical trigger is achieved are those which are specified in the Core Protocol. At the time of a Platform Conclusion that is relevant to public health or clinical management of patients with suspected or proven pandemic infection, the DSMB and ITSC are empowered to liaise directly with relevant public health authorities prior to public presentation or publication of results.

###### 7.7.6. Pre-specified subgroup analyses after achievement of a platform conclusion

Pre-specified subgroup analyses that will be conducted after a Platform Conclusion are outlined in each DSA. If a DSA does not specify a sub-group analysis related to the pandemic strata such analysis is permitted if the PISOP stratum has been open.

###### 7.7.7. Closure of the PISOP stratum and incorporation of data from pandemic statistical model into the interpandemic statistical model

The ITSC is permitted to close or suspend the PISOP stratum. At this time, evaluation of new patients within the pandemic model will cease. After the permanent closure of the PISOP stratum, the information related to domains that have been analyzed for PISOP patients within the pandemic model will be added to the interpandemic model retaining, if appropriate, a co-variate or stratum status, to reflect that the patient was enrolled in the PISOP stratum.

###### 7.7.8. Domains with their own statistical model

It is intended that domains with their own statistical model (e.g. as anticipated for the ventilation domain) will continue to be analyzed using the separate statistical model. If the PISOP stratum was applied to such a domain it is intended that a pandemic version of the separate model would be commenced and enroll only patients in the PISOP stratum. This model would utilize the pandemic primary end-point and would use informative priors derived from the preceding model. An operational decision may be made to apply an end-point that is different to the pandemic primary end-point in a domain with its own model.

Not for IRB submission

### 8. GOVERNANCE, ETHICAL, AND OPERATIONAL CONSIDERATIONS IN A PANDEMIC

#### *8.1. Decision to activate pandemic stratum*

The decision to open the pandemic stratum lies with the ITSC. In deciding to activate the pandemic stratum the ITSC should take into account, but is not dependent on, declaration of a pandemic by the WHO and decisions about pandemic activation by regional pandemic preparedness consortia.

The decision to open will be communicated to RCCs and participating sites as an operational document. Each RCC will maintain a log of the dates for which sites were activated for the PISOP stratum.

#### *8.2. Safety Monitoring and Reporting*

During the interpandemic period, the platform evaluates solely or predominantly interventions that are in widespread clinical use for severe CAP and for which the safety profile of the intervention is well described. During a pandemic, the platform may evaluate therapeutic agents that have been repurposed or are an Investigational Medical Product. Such therapeutic agents may not have an established safety profile or an established safety profile when used in critically ill patients. Where an intervention is not regarded as having an established safety profile, this will be specified in the DSA. For this type of interventions more specific or more detailed SAE reporting will be required that is specified in the Core Protocol, as follows.

This may include Adverse Events of Special Interest (AESI). SAEs that might be attributable to specific interventions are included as secondary endpoints in each DSA but are recorded only for participants who are enrolled in that domain. If more detailed SAE or AE/AESI reporting is required for an intervention, then this additional safety reporting requirement will be specified in the relevant DSA and recorded only for participants who are enrolled in that domain. The following arrangements apply to such

When submitting the SAE form the local site PI should determine if the SAE is attributable to one or more study interventions in this trial. The local PI will assess if it is possible, probable, or certain that there is a direct link between a trial intervention and the SAE (a Serious Adverse Drug Reaction, SADR).

Not for IRB submission

The regional / country project manager should review the SAE form for completeness and query any missing data with the site. Preliminary SAE report forms should be submitted as soon as the site becomes aware. It is recognised that follow-up information may be available later.

The regional lead investigator, or medically qualified designee, should review the SAE to assess expectedness and causality. The regional lead investigator or delegate cannot downgrade the site's assessment of expectedness and causality. The following requirements are specified:

- The regional Sponsor should be made aware of the SAE within 24 hours of the SAE being reported.
- All SAEs must be followed-up until resolution, or end of trial if this is sooner.
- SAEs will be reported to the relevant ethics committee and competent authority according to local regulations and requirements.

All SAEs, pooled from all regions, will be reported to the DSMB at intervals agreed by the REMAP-CAP investigators and the DSMB. This may vary depending on the specific intervention being evaluated. The DSMB may request additional specialist review of safety data for certain interventions.

If drugs have been supplied by a pharmaceutical company, then reporting of safety data to the company may be required. The details of this reporting will be included in individual Safety Data Exchange Agreements (SDEA).

On an annual basis a Developmental Safety Update Report (DSUR) will be produced including all SAE data from all regions in REMAP-CAP and will be submitted to the relevant competent authorities as required. This may be shared with pharmaceutical companies as part of the SDEA.

If an SAE is determined to be unexpected (not previously described in the Summary of Product Characteristics / Investigator Brochure / Protocol) and related to the study medication then it is considered a SUSAR. In these cases, the following steps should also be undertaken, in addition the performing the steps described above for handling SAEs:

- The relevant competent authorities and ethics committees should be notified of the SUSAR by the Sponsor or designee in each region.
- A SUSAR that results in death or is life-threatening, should be reported to the aforementioned bodies within 7 days of the Sponsor (or designee) becoming aware of the

Not for IRB Submission

event. Further relevant information should be sought and a follow-up report completed as soon as possible and submitted within 8 additional days.

A SUSAR which does not result in death or is not life-threatening should be reported within 15 days of the Sponsor (or designee) becoming aware of the event or in accordance with the local regulatory requirements. Further relevant information should be given as soon as possible. The regional / country project managers should notify all investigators at all sites that a SUSAR has occurred. The REMAP-CAP DSMB should be notified that a SUSAR has occurred.

It may be necessary to take appropriate urgent safety measures in order to protect research participants against any immediate hazard to their health or safety, without prior authorisation from a regulatory body. If this occurs the regulatory bodies should be notified as soon as possible and in any event within three days, in the form of a substantial amendment, that such measures have been taken and the reasons why.

SAEs reported will be coded using the currently available version of the Medical Dictionary for Regulatory Activities (MedDRA). Coding will be to lowest level terms. The preferred term, and the primary system organ class will be listed. Summaries of all SAEs by treatment group will include:

- The number and percentage of patients with at least 1 SAE by system organ class and preferred term
- The number of SAEs by relationship to treatment (related, not related), presented by system organ class and preferred term

#### 8.3. *Data collection and management*

A pandemic is likely to result in a substantial increase in clinical workload for sites participating in REMAP-CAP. This is acknowledged by the REMAP-CAP management, as is the primacy of patient care. The importance of contemporaneous data collection, particularly with respect to variables that are needed for adaptive analyses will be emphasized to sites. RCCs will seek to support sites as much as possible, including with requests to healthcare systems, public health authorities, and funding agencies to provide resources that allow sites to maintain data collection that is timely and complete.

#### 8.4. *Role of the DSMB*

In a pandemic the role of the DSMB is modified, taking into account the public health importance of clinical evidence during a pandemic. In meeting the requirements of their Charter during a pandemic

Not for IRB Submission

the DSMB should consider issues of public health in addition to the well-being of participants and the scientific integrity of the platform. The in-principle views of the DSMB may be obtained by the ITSC with regard to the setting of modified thresholds for statistical triggers.

While the PISOP stratum is open the DSMB is also permitted to liaise with public health authorities, regulatory authorities, or the DSMBs of trials evaluating the same or similar interventions regarding the results and appropriate interpretation of adaptive analyses in keeping with prevailing international standards. If the DSMB communicates with external groups the ITSC may be informed that such communication has occurred but the content of that communication may remain confidential between the DSMB and the relevant group. The DSMB may recommend to the ITSC that public reporting of posterior probabilities that have not attained a threshold for a Statistical Trigger should occur.

The workload of the DSMB may be substantial during a pandemic and, if requested by the DSMB, the ITSC will appoint additional members.

#### 8.5. Communication of trial results

Any Platform Conclusion that is relevant to public health that occurs during a pandemic will be presented or published as soon as possible, noting that additional work to report baseline status and secondary end-points will need to occur prior to presentation and publication of results.

#### 8.6. Funding of the trial

The trial is currently funded as described in the Core Protocol.

During the interpandemic period and during a pandemic, additional funding will be sought to provide resources for activities that exceed those that will be occurring during the interpandemic period. Possible sources of additional resources include, but are not limited to, healthcare systems, pharmaceutical companies, public health authorities, and local and international research funding bodies.

A section of the Core Protocol indicates that “the trial will not enter into a contract with a commercial organization unless the contract specifies that, among other clauses, “that all data are owned by the trial and the commercial organization has no authority to access data”. This clause should not be interpreted as indicating that access to data by a commercial organization is not permitted. Such as access can be agreed, for example, by licensing access to data, if agreed by both a commercial partner and trial sponsors.

Not for IRB Submission

#### 8.7. Monitoring

It is acknowledged that during a pandemic site monitoring may be delayed for logistical reasons. The operational monitoring plan may be updated to reflect issues that are specific to a pandemic. As outlined in Core Protocol, the DSMB will take into account intensity of monitoring and time of consideration of a Platform Conclusion. If appropriate, the contribution of data that has not been monitored as per the non-pandemic monitoring plan will be acknowledged in the public reporting of Platform Conclusions.

Not for IRB Submission

Not for IRB Submission

### Domain-Specific Appendix: COVID-19 Therapeutic Anticoagulation

#### REMAP-CAP: Randomized, Embedded, Multifactorial Adaptive Platform trial for Community-Acquired Pneumonia

Therapeutic Anticoagulation Domain-Specific Appendix Version 2.0 dated 24th June 2020

Not for IRB submission

#### Summary

In this domain of the REMAP-CAP trial, participants meeting the platform entry criteria with suspected or microbiological testing-confirmed COVID-19 infection will be randomized to one of two interventions:

- Local standard venous thromboprophylaxis
- Therapeutic anticoagulation with intravenous unfractionated heparin or subcutaneous low molecular weight heparin

At this participating site the following interventions have been selected within this domain:

☐ Local standard venous thromboprophylaxis

☐ Therapeutic anticoagulation with intravenous unfractionated heparin or subcutaneous low molecular weight heparin

This DSA applies to the following states and stratum:

| | | | | |
|---|---|---|---|---|
| Stratum | Pandemic infection suspected or proven (PISOP) | | | Pandemic infection neither suspected nor proven (PINSNP) |
| Core protocol documents | REMAP-CAP Core Protocol + Pandemic Appendix, or REMAP-COVID Core Protocol | | | REMAP-CAP Core Protocol |
| Illness Severity State | Moderate State | | Severe State | Severe State |
| Interventions specified in this DSA | Local VT<br>Therapeutic anticoagulation | | Local VT<br>Therapeutic anticoagulation | Not available |
| Interventions submitted for approval in this jurisdiction | ☐ Local VT<br>☐ Therapeutic anticoagulation | | ☐ Local VT<br>☐ Therapeutic anticoagulation | Not available |
| Interventions offered at this site | Ward | ICU | ICU | ICU |
| | ☐ Local VT<br>☐ Therapeutic anticoagulation | ☐ Local VT<br>☐ Therapeutic anticoagulation | ☐ Local VT<br>☐ Therapeutic anticoagulation | Not available |

Not for IRB Submission

| REMAP-CAP: COVID-19 Therapeutic Anticoagulation Domain Summary | |
|---|---|
| Interventions | • Local standard venous thromboprophylaxis<br>• Therapeutic anticoagulation with intravenous unfractionated heparin or subcutaneous low molecular weight heparin |
| Unit of Analysis, Strata, and State | This domain is analyzed only in the pandemic statistical model.<br><br>The pandemic statistical model includes only patients who are in the Pandemic Infection Suspected or Proven (PISOP) stratum. Within this stratum, the unit-of-analysis is defined by illness severity state at time of enrollment, defined as either Moderate State or Severe State. Unit-of-analysis may also be defined by SARS-CoV-2 infection or d-dimer strata or both. Borrowing is permitted between states and strata. If the SARS-CoV-2 strata is applied in analysis, Response Adaptive Randomization will be applied to all PISOP patients, in each illness severity state, using probabilities derived from the SARS-CoV-2 confirmed stratum. Response Adaptive Randomization may also be applied according to D-dimer strata status. |
| Evaluable treatment-by-treatment Interactions | No interaction will be evaluated with any other domain. |
| Nesting | None |
| Timing of Reveal | Randomization with Immediate Reveal and Initiation or Randomization with Deferred Reveal if prospective agreement to participate is required. |
| Inclusions | Patients will be eligible for this domain if:<br>• COVID-19 infection is suspected by the treating clinician or has been confirmed by microbiological testing<br>• Microbiological testing for SARS-CoV-2 infection of upper or lower respiratory tract secretions or both has occurred or is intended to occur |
| Domain-Specific Exclusions | Patients will be excluded from this domain if they have any of the following:<br>• More than 48 hours has elapsed since ICU admission (noting that this may be operationalized as more than 48 hours has elapsed since commencement of organ failure support)<br>• Clinical or laboratory bleeding risk or both that is sufficient to contraindicate therapeutic anticoagulation, including intention to continue or commence dual anti-platelet therapy<br>• Therapeutic anticoagulation is already present due to prior administration of any anticoagulant agent that is known or likely to still be active or a clinical decision has been made to commence therapeutic anticoagulation<br>• Enrolment in a trial evaluating anticoagulation for proven or suspected COVID-19 infection, where the protocol of that trial requires continuation of the treatment assignment specified in that trial<br>• Known or suspected previous adverse reaction to UFH or LMWH including heparin induced thrombocytopenia (HIT).<br>• The treating clinician believes that participation in the domain would not be in the best interests of the patient |
| Intervention-Specific Exclusions | None |

Not for IRB Submission

| | |
|---|---|
| Outcome measures | Primary REMAP endpoint: refer to REMAP-CAP Core Protocol + Pandemic Appendix and REMAP-COVID Core Protocol<br>Secondary REMAP endpoints: refer to REMAP-CAP Core Protocol + Pandemic Appendix and REMAP-COVID Core Protocol<br><br>Secondary domain-specific endpoints (during hospitalization censored 90 days from the date of enrollment):<br>• Confirmed deep venous thrombosis<br>• Confirmed pulmonary embolism<br>• Confirmed ischemic cerebrovascular event<br>• Total red cell blood cell units transfused between randomization and the end of study day 15<br>• Acute myocardial infarction<br>• Peak troponin<br>• Major bleeding<br>• Other thrombotic events including mesenteric ischemia and limb ischemia<br>• Serious Adverse Events (SAE) as defined in relevant core protocol documents and this DSA |

Not for IRB Submission

8.3.4. Discontinuation of study intervention ... 25
8.3.5. COVID-19 anticoagulation strategy in patients negative for COVID-19 infection ... 25
**8.4. Concomitant care ... 26**
**8.5. Endpoints ... 26**
8.5.1. Primary endpoint ... 26
8.5.2. Secondary endpoints ... 26
9. *TRIAL CONDUCT ... 27*
**9.1. Microbiology ... 27**
**9.2. Domain-specific data collection ... 27**
**9.3. Criteria for discontinuation ... 28**
**9.4. Blinding ... 28**
9.4.1. Blinding ... 28
9.4.2. Unblinding ... 28
10. *STATISTICAL CONSIDERATIONS ... 28*
**10.1. Domain-specific stopping rules ... 28**
**10.2. Unit-of-analysis and strata ... 28**
**10.3. Timing of revealing of randomization status ... 29**
**10.4. Interactions with interventions in other domains ... 29**
**10.5. Nesting of interventions ... 30**
**10.6. Threshold probability for superiority and inferiority ... 30**
**10.7. Threshold odds ratio delta for equivalence ... 30**
**10.8. Informative priors ... 30**
**10.9. Post-trial sub-groups ... 31**
11. *ETHICAL CONSIDERATIONS ... 31*
**11.1. Data Safety and Monitoring Board ... 31**
**11.2. Potential domain-specific adverse events ... 31**
**11.3. Domain-specific consent issues ... 32**
**11.4. Relationship to Antiplatelet Domain ... 32**
12. *GOVERNANCE ISSUES ... 32*
**12.1. Funding of domain ... 32**
**12.2. Funding of domain interventions and outcome measures ... 32**
**12.3. Domain-specific declarations of interest ... 33**
13. *REFERENCES ... 34*

Not for IRB Submission

Not for IRB Submission

### 1. ABBREVIATIONS

| | |
|---|---|
| ACE2 | Angiotensin-Converting Enzyme 2 |
| aPTT | Activated partial thromboplastin time |
| ARDS | Acute Respiratory Distress Syndrome |
| CCP | Clinical Characterization Protocol |
| DSA | Domain-Specific Appendix |
| DIC | Disseminated Intravascular Coagulation |
| DSMB | Data Safety and Monitoring Board |
| DSWG | Domain-Specific Working Group |
| HIT | Heparin Induced Thrombocytopenia |
| ICU | Intensive Care Unit |
| ISIG | International Statistics Interest Group |
| ITSC | International Trial Steering Committee |
| LMWH | Low Molecular Weight Heparin |
| MERS-CoV | Middle East respiratory syndrome coronavirus |
| PAtC | Pandemic Appendix to the Core Protocol |
| PE | Pulmonary Embolus |
| PISOP | Pandemic infection is suspected or proven |
| RCT | Randomized controlled trial |
| REMAP-CAP | Randomized, Embedded, Multifactorial, Adaptive Platform trial for Community-Acquired Pneumonia |
| RSA | Region-Specific Appendix |
| SAE | Serious Adverse Event |
| SARS | Serious Acute Respiratory Syndrome |
| UFH | Unfractionated heparin |

Not for IRB submission

| | |
|---|---|
| VTE | Venous Thromboembolism |
| WHO | World Health Organization |

Not for IRB submission

### 2. PROTOCOL APPENDIX STRUCTURE

The structure of this protocol is different to that used for conventional trials because this trial is highly adaptive and the description of these adaptations is better understood and specified using a 'modular' protocol design. While all adaptations are pre-specified, the structure of the protocol is designed to allow the trial to evolve over time, for example by the introduction of new domains or interventions or both (see glossary, Section 1.2 Core Protocol for definitions of these terms) and commencement of the trial in new geographical regions.

The protocol has multiple modules, in brief, comprising a Core Protocol (overview and design features of the study); a Statistical Analysis Appendix (details of the current statistical analysis plan and models); Simulations Appendix (details of the current simulations of the REMAP); multiple Domain-Specific Appendices (DSA) (detailing all interventions currently being studied in each domain); and multiple Region-Specific Appendices (RSA) (detailing regional management and governance).

The Core Protocol contains all information that is generic to the trial, irrespective of the regional location in which the trial is conducted and the domains or interventions that are being tested. The Core Protocol may be amended but it is anticipated that such amendments will be infrequent.

The Core Protocol does not contain information about the intervention(s), within each domain, because one of the trial adaptations is that domains and interventions will change over time.
Information about interventions within each domain is covered in a DSA. These Appendices are anticipated to change over time, with removal and addition of options within an existing domain, at one level, and removal and addition of entire domains, at another level. Each modification to a DSA will be subject to a separate ethics application for approval.

The Core Protocol does not contain detailed information about the statistical analysis or simulations, because the analytic model will also change over time in accordance with the domain and intervention trial adaptations but this information is contained in the Statistical Analysis and Simulations Appendices. These Appendices are

Not for IRB submission

anticipated to change over time, as trial adaptations occur. Each modification will be subject to approval from the International Trial Steering Committee (ITSC) in conjunction with advice from the International Statistics Interest Group (ISIG) and the Data Safety and Monitoring Board (DSMB).

The Core Protocol also does not contain information that is specific to a particular region in which the trial is conducted, as the locations that participate in the trial are also anticipated to increase over time. Information that is specific to each region that conducts the trial is contained within a RSA. This includes information related to local management, governance, and ethical and regulatory aspects. It is planned that, within each region, only that region's RSA, and any subsequent modifications, will be submitted for ethical review in that region.

The current version of the relevant Core Protocol (either REMAP-CAP Core Protocol +/- Pandemic Appendix or REMAP-COVID Core Protocol), DSAs, RSAs, and the Statistical Analysis Appendix is listed in the Protocol Summary and on the study website (www.remapcap.org).

### 3. COVID-19 THERAPEUTIC ANTICOAGULATION DOMAIN-SPECIFIC APPENDIX VERSION

The version of the COVID-19 Therapeutic Anticoagulation Domain-Specific Appendix is in this document's header and on the cover page.

#### 3.1. Version history

Version 1: Approved by the COVID-19 Therapeutic Anticoagulation Domain-Specific Working Group (DSWG) on 20th April 2020.

Version 2: Approved by the COVID-19 Therapeutic Anticoagulation DSWG on 24th June 2020

### 4. COVID-19 THERAPEUTIC ANTICOAGULATION THERAPY DOMAIN GOVERNANCE

#### 4.1. Domain members

**Chair:** Dr. Ryan Zarychanski

**Deputy Chair:** Dr. Ewan Goligher

Members:

Prof. Derek

Not for IRB Submission

Angus Dr. Scott Berry Dr. Shailesh Bihari D

r. Charlotte
Bradbury Prof.
Marc Carrier

Prof. Dean
Fergusson Prof.
Robert Fowler
A/Prof. Timothy
Girard Prof.
Anthony Gordon
A/Prof. Ghady
Haidar
A/Prof. Christopher
Horvat Prof. David
Huang
Prof. Beverley
Hunt Prof.
Anand Kumar
Prof. Mike
Laffan Dr.
Patrick Lawler
Dr. Patrick
Lawless Dr.
Sylvain Lother
Dr. Peter
MacCallum Dr.
Colin McArthur
A/Prof. Bryan
McVerry Prof. John
Marshall Prof.
Saskia Middeldorp
Dr. Zoe McQuilten
A/Prof. Matthew
Neal Prof. Alistair
Nichol Prof. John
Pasi
A/Prof. Christopher

Not for IRB Submission

Seymour Prof. Roger

Schutgens

Prof. Simon

Stanworth Dr.

Alexis Turgeon

Prof. Steve Webb

A/Prof. Alexandra Weissman

#### 4.2. *Contact Details*

**Chair:** Dr. Ryan Zarychanski

ON4005 – 675 McDermot Ave

Winnipeg, Manitoba, Canada.

R3M 3M6 Email:

##### *4.3. Interaction with ATTACC trial*

ATTACC is a trial that also evaluates the treatment effect of therapeutic anticoagulation in patients with COVID-19. There is overlap between the leadership of the ATTACC trial and the leadership of this domain. This domain and ATTACC have been designed to be complementary with pre-specified plans in relation to methods of analysis. It is intended that data from ATTACC will be incorporated into the pandemic statistical model of REMAP-CAP. The protocol, governance, and data management of ATTACC are separate from REMAP-CAP, but the REMAP-CAP DSMB will also serve the ATTACC trial.

#### 5. COVID-19 THERAPEUTIC ANTICOAGULATION THERAPY DOMAIN-SPECIFIC WORKING GROUP AUTHORIZATION

The COVID-19 Domain-Specific Working Group have read the appendix and authorize it as the official COVID-19 Therapeutic Anticoagulation Domain-Specific Appendix for the study entitled REMAP-CAP. Signed on behalf of the committee,

**Chair**
Dr. Ryan Zarychanski

Date 24th June 2020

#### 6. BACKGROUND AND RATIONALE

##### *6.1. Domain definition*

This is a domain within the REMAP-CAP platform to test the effectiveness of therapeutic anticoagulation versus local venous thromboprophylaxis for

Not for IRB Submission

patients with acute illness due to suspected or proven COVID-19.

#### 6.2. *Domain-specific background*

##### 6.2.1. COVID-19 infection

The first report of infection with COVID-19 occurred in Wuhan, China, in late 2019. Since that time, and as of the time of writing of this DSA, there have been over 1 million reported cases across the world with a range of severity, approximately 60,000 deaths and sustained human-human transmission. On January 30th 2020, the World Health Organization (WHO) declared this outbreak a Public Health Emergency of International Concern (https://www.who.int/news-room/detail/30-01-2020-statement-on-the-second-meeting-of-the-international-health-regulations-(2005)-emergency- committee-regarding-the-outbreak-of-novel-coronavirus-(2019-ncov)). Given past history with novel coronaviruses, such as Severe Acute Respiratory Syndrome (SARS) and Middle East respiratory syndrome coronavirus (MERS-CoV), public health agencies have responded aggressively to the urgent need to acquire knowledge regarding this emerging infection. An important component of this urgently needed knowledge includes understanding the effectiveness of alternative treatment strategies in patients with suspected or proven infection. It should also be noted that clinical guidance issued by the WHO indicates that unproven therapies should be administered preferably only as part of a clinical trial (https://www.who.int/docs/default-source/coronaviruse/clinical- management-of-novel-cov.pdf).

Estimates of the burden of critical illness among patients infected with COVID-19 vary, with estimates of case-fatality and proportion of patients who become critically ill being unstable. Several factors contribute to this uncertainty including differential timing between diagnosis and development of critical illness or death, the true incidence of infection being uncertain because of possible under-reporting of asymptomatic or mild cases driven largely by limitations in the number of diagnostic tests that can be performed.

The first case descriptions of COVID-19 disease were communicated by Chinese investigators. These reports describe a progressive severe pneumonia, with a significant proportion of patients requiring mechanical ventilation and some reports of multi-organ dysfunction. In a study of 41 hospitalized patients with laboratory-confirmed COVID-19 infection, 13 (32%) patients were admitted to an ICU and six

Not for IRB submission

(15%) died. Invasive mechanical ventilation was required in four (10%) patients, with two patients (5%) receiving extracorporeal membrane oxygenation as salvage therapy (Huang et al.). In another study of 99 hospitalized patients with COVID-19 pneumonia, 23 (23%) were admitted to ICU, 17 (17%) developed acute respiratory distress syndrome (ARDS), three (3%) acute renal failure and four (4%) septic shock. In a study of 138 patients with COVID-19 infection, 36/138 (26%) required ICU care. Patients admitted to ICU were older and were more likely to have underlying comorbidities. In the ICU, four patients (11% of those admitted to ICU) received high-flow oxygen and 15 (44.4%) received noninvasive ventilation. Invasive mechanical ventilation was required in 17 patients (47.2%), four of whom received extracorporeal membrane oxygenation as rescue therapy. A total of 13 patients received vasopressors and two patients received kidney replacement therapy (Wang et al., 2020a). In a study from the Chinese Centers for Disease Control that reported on 72,314 patients, 49% of patients defined as critically ill died before hospital discharge (1,023 of 2,087) (Wu and McGoogan, 2020).

As with the other major coronaviruses that have circulated in outbreaks in recent decades, SARS and MERS-CoV, no specific therapy, or an element of supportive care, has been formally evaluated in randomized controlled trials with sufficient statistical power to identify changes in patient-centered outcomes.

Interim recommendations from the WHO for clinical care of infected patients focus upon supportive care, including organ support as needed, prevention of complications, with any specific therapy to only be provided as part of a research protocol (https://www.who.int/docs/default- source/coronaviruse/clinical-management-of-novel-cov.pdf).

##### 6.2.2. Clinical trials for COVID-19 infection

###### 6.2.2.1. *Current clinical trials and interventions being evaluated*

As of 24th February 2020, more than 150 clinical studies from China had been registered on trial registration sites. Many of these trials are single center and with sample sizes that are unlikely to be sufficient to detect plausible treatment effects, with some studies being uncontrolled or observational. There is also a rapid decline in incidence of new infection in China and many clinical trials are unlikely to achieve their planned sample size.

A wide range of interventions are being evaluated in trials that have been registered including arbidol, lopinavir/ritonavir, darunavir/cobicistat, remdesivir, favipiravir, baloxavir, chloroquine, intravenous immunoglobulin, inhaled and parenteral interferon-α or interferon-β glucocorticoids (different agents and doses),

Not for IRB Submission

mesenchymal and other stem cells, microbiota transplantation, and a range of traditional Chinese medicines.

WHO has provided guidance regarding both trial design and prioritization of candidate therapies. With regards to trial design, WHO notes that there are no treatments with proven efficacy in patients with COVID-19. As such, WHO guidance is that trials should utilize a 'standard of care' comparator, that is, a control group that does not receive an agent intended to be active against COVID-19 infection, its associated immune response or other complications (https://apps.who.int/iris/bitstream/handle/10665/330694/WHO-HEO-RDBlueprintnCoV-2020.4- eng.pdf?ua=1).

This Therapeutic Anticoagulation Domain will evaluate the effect of therapeutic anticoagulation with intravenous unfractionated heparin (UFH) or subcutaneous low molecular weight heparin (LMWH) compared to standard venous thromboprophylaxis (delivered according to local practice in each region) in critically ill patients with COVID-19.

###### 6.2.2.2. *Need for evidence in patients who are critically ill as well as hospitalized patients*

There is need to evaluate interventions for COVID-19 infection in patients who are critically ill or hospitalized and not critically ill, separately, because of the possibility of differential treatment effect, depending on illness severity. The number of current studies that are focused on patients who are critically ill is uncertain and, for those studies that are enrolling hospitalized patients, it is unclear if stratification by severity is a design feature.

Among trials that evaluate interventions in patients who are critically ill it is common for the results of the trial to be different to that which was predicted based on a prior understanding of mechanism of action combined with known mechanism of disease (Landoni et al., 2015, Webb, 2015). This observation reinforces the importance of not necessarily relying on extrapolation of results (both positive and negative) from patients who are not critically ill. It is also possible different disease mechanisms apply at different levels of illness severity and that this may also influence balance between beneficial and adverse effects of a particular intervention. This reinforces the importance of obtaining estimates of treatment effect dependent on the level of illness severity.

###### 6.2.3. Intervention strategy for this domain

Not for IRB submission

This domain will test the potential benefits of different approaches to achieving therapeutic anticoagulation compared to usual care, comprising local standard-of-care venous pharmacological thromboprophylaxis.

If at, any stage, evidence of harm or definitive evidence of absence of effectiveness in critically ill or ward patients or both emerges for one or more interventions specified in this domain, the ITSC, as advised by the DSWG, may remove the intervention(s) prior to declaration of a Platform Conclusion. If this occurs, presentation and publication of results that relate to the intervention will occur, so as to contribute additional weight of evidence in the public domain.

##### 6.2.4. Rationale for therapeutic anticoagulation in COVID-19

Although respiratory mechanics in COVID-19-associated ARDS has not yet been systematically described, there are widespread reports that patients exhibit surprisingly high respiratory compliance despite profoundly impaired gas exchange and radiological opacities. The gas exchange impairment characteristically involves severe hypoxemia but also markedly elevated physiological dead space and elevated respiratory drive (Liu et al., 2020).

Severe illness from COVID-19 seems to be characterized by important derangements in coagulation resulting in a hypercoagulable state. These derangements are strongly associated with poor clinical outcomes and various lines of evidence suggest that the prothrombotic state is causally related to poor outcomes. In a series of 183 patients, patients who died (11%) exhibited markedly elevated D- dimers and elevated fibrin degradation products; 15 of the patients who died met criteria for disseminated intravascular coagulation (DIC), whereas only 1 survivor developed DIC (Tang et al., 2020b). Similar derangements in hemostasis were documented in a separate case series of 94 patients (Lippi and Plebani, 2020). Development of DIC correlated with clinical deterioration.
Ischemic injury of the fingers and toes has also been reported in patients with severe COVID-19 (Li et al., 2020). In multiple large case series, elevated D-dimer is consistently associated with a higher risk of developing ARDS and death (Wu et al., 2020, Zhou et al., 2020). Reports of acute cardiovascular collapse with echocardiographic evidence of right heart strain has also been reported. In a consecutive case series of 184 COVID-19 positive patients admitted to a Dutch teaching hospital, the incidence of a composite outcome comprised of symptomatic PE, deep-vein thrombosis, ischemic stroke, myocardial infarction, or systemic arterial embolism occurred in 31% of patients (Klok et al., 2020).

The exact mechanism of coagulopathy and DIC is uncertain. SARS-CoV-2 can bind angiotensin- converting enzyme 2 (ACE2) and infect and injure endothelium,

Not for IRB submission

leading to tissue factor expression, endothelial activation and activation of the coagulation cascade (Zhang et al., 2020).

Endothelial dysfunction and microvascular thrombosis could explain the constellation of pulmonary findings in severe COVID-19—high dead space and impaired oxygenation in the absence of significant increase in pulmonary elastance (Liu et al., 2020). These features suggest that the pathophysiology of severe COVID-19 is quite different from typical ARDS, where shunt and dead space increase in proportion to the loss of lung volume and resulting increase in elastance. The limited autopsy data suggest a constellation of pulmonary pathological findings including thrombus in pulmonary microvessels. Endothelial dysfunction and microvascular thrombosis could also account for the high rate of cardiac injury with elevated Troponin-I and arrhythmia—both associated with poor outcome (Guo et al., 2020).

The SARS-CoV-2 spike protein has been shown to interact with UFH and LMWH. Upon binding heparin, the spike protein undergoes significant conformational change that may prevent it from binding ACE2 (https://www.biorxiv.org/content/10.1101/2020.02.29.971093v1). Heparin has been shown to prevent cellular invasion by SARS-CoV-1 (Vicenzi et al., 2004, Lang et al., 2011), and is known to inhibit attachment and entry of other enveloped viruses such as Human Immunodeficiency Virus and Herpes Simplex Virus (Moulard et al., 2000). Thus, heparin may exert a direct antiviral effect to prevent invasion of pulmonary epithelium, myocardium, and vascular endothelium, as well as potentially act to counteract complications that arise because of a hypercoagulable state.

Independent of its role as an anticoagulant, UFH has been shown to neutralize endotoxin and increase serum tumor necrosis factor binding protein-I, thus limiting both activation of coagulation and inflammation (Anastase-Ravion et al., 2003). UFH is also a known inhibitor of complement and of adhesion molecule expression in the microvasculature, which may serve to limit hemolysis and decrease neutrophil adhesion in the setting of sepsis (Lever et al., 2000). More recently, UFH has been shown to modulate HDL and reduce oxidant induced cellular damage (Wu et al., 2004), likely by abrogating histone-mediated cytotoxicity (Wildhagen et al., 2014).

There are anecdotal reports of anticoagulation with UFH being used in the treatment of COVID-19 disease in many locations. As such, it is of substantial importance that the treatment effect of UFH is established in randomized controlled trials (RCTs).

###### 6.2.5. Evidence of effect for anticoagulation in sepsis and COVID-19 disease

Animal data suggest a benefit of heparin in models of sepsis. UFH administration reduces activation of coagulation and increases survival in endotoxin-equivalent

Not for IRB Submission

models (including live organism infusion) of septic shock (du Toit et al., 1991). A meta-analysis of studies in animal models of sepsis found that UFH reduced the odds of death (odds ratio 0.27, 95%CI 0.16 to 0.46; n = 10 studies) (Cornet et al., 2007).

In a propensity matched retrospective cohort study of patients with septic shock therapeutic dose UFH was associated with reduced 28-day when administered within 48 hours of ICU admission

(Zarychanski et al., 2008). Subgroup analyses from 3 randomized trials studying natural anticoagulants (rhAPC, antithrombin, and tissue factor pathway inhibitor) in sepsis suggest a survival advantage associated with prophylactic dose heparin when administered as a co-intervention, independent of the study drug under investigation or whether the study drug was received (OR 0.69, 95%CI 0.56 to 0.85) (Polderman and Girbes, 2004). In a meta-analysis of RCTs conducts conducted in patients with sepsis and septic shock, compared to placebo or no intervention heparin was associated with a reduction in the odds of death (odd ratio 0.88 (95% CI, 0.77 to 1.00; $I^2$ = 0%) (Polderman and Girbes, 2004). Evidence of potential benefit was not dependent on the presence of DIC or coagulopathy. In a second meta-analysis that evaluated the effects of LMWH in Chinese trials that evaluated LWMH in sepsis, LMWH was associated with reduced 28-day mortality (Fan et al., 2016). In patients with septic shock, therapeutic UFH is currently being evaluated in an international phase II/III RCT (www.halointernational.org, NCT03378466).

Specific to COVID-19 disease, in an observational study of 449 hospitalized patients from Wuhan, China, among 99 patients who received heparin (primarily LMWH, but also UFH) at prophylactic doses, heparin was associated with reduced 28-day mortality in patients with sepsis-induced coagulopathy or who had d-dimers that were greater than 6-fold the upper limit of normal (Tang et al., 2020a).

High troponin has been reported to strongly be associated with poor outcomes in patients with COVID-19 disease (Inciardi et al., 2020, Wang et al., 2020b). Reports of arterial events in critically ill COVID-19 patient, including myocardial infarction and stroke occurring in COVID-19 positive patients have also been forwarded. Platelet activation is known to occur in infection, DIC and hemophagocytic syndrome (de Stoppelaar et al., 2014). While the majority of interventional trials of anti-thrombotics in sepsis have focused on parenteral anticoagulants, the role of anti-platelet agents in sepsis and in COVID-19 patients remains to be evaluated.

###### 6.2.6. Intravenous unfractionated heparin

UFH is a naturally occurring glycosaminoglycan that exerts its anticoagulant effect by enhancing antithrombin mediated inactivation of factors Xa and IIa, but also factors IXa, XIa, and XIIa (Gans, 1975). Because its size, activity, and pharmacokinetics are variable, its anticoagulant effect requires close monitoring in

Not for IRB submission

hospital settings. Chains of UFH varies in length and molecular weights from 5,000 to over 40,000 Daltons.

###### 6.2.7. Low molecular weight heparin

LMWH represent, on average, shorter chains of UFH with an average molecular weight less than 8,000 Daltons. LMWH is obtained by various methods including fractionation or depolymerization of polymeric heparin. LMWHs exert the majority of their anticoagulant effect through factor X compared to its effect on factor II (thrombin).

###### 6.2.8. Safety of unfractionated heparin and Low molecular weight heparin

UFH and LMWH are anticoagulants and as such are associated with major and clinically relevant minor bleeding. Therapeutic anticoagulation has been studied extensively across diverse patient populations, including both critically ill and ward patients, and favorable safety data is available. Therapeutic anticoagulation is commonly used in hospitalized patients for the treatment of venous thromboembolic disease, acute coronary syndromes, and stroke prevention in patients with atrial fibrillation (Tiryaki et al., 2011). The dosing and management of both unfractionated heparin and low molecular weight heparin are very familiar to clinicians. Overall, patients receiving therapeutic anticoagulation with these agents have a 1-5% risk of major bleeding, depending on underlying risk and duration of exposure (Mismetti et al., 2005, Petersen et al., 2004, Crowther and Warkentin, 2008).

Patients with an underlying systemic hypercoagulable state (such as COVID-19), in whom therapeutic anticoagulation is being given to offset this, may intuitively have a lower risk of bleeding. For example, in cancer-associated venous thromboembolisms
– an underlying hypercoagulable state
– the estimated rate of major bleeding was reported to be 3.2% over a 6 months period (Lee et al., 2015, Li et al., 2019).

In the PROTECT trial, a multi-national thromboprophylaxis RCT comparing UFH to LMWH in critically ill patients (n=3764), the major bleeding rate was 5.6% (Group et al., 2011). In this trial, no relationship was detected between use of therapeutic heparin and the activated partial thromboplastin time (aPTT) (p = 0.41) (Lauzier et al., 2013).

In patients receiving therapeutic anticoagulation for the treatment of venous thromboembolism (VTE), the rate of major hemorrhage typically reported ranges

Not for IRB submission

from 2-3%. Rates of major hemorrhage in patients randomized to receive UFH or LMWH appear to be similar (Dolovich et al., 2000). In patients therapeutically anticoagulated for treatment of acute coronary syndrome, rates of major hemorrhage in patients receiving UFH + a glycoprotein IIb/IIIa inhibitor is approximately 6% and similar (6%) in patients receiving LMWH (Navarese et al., 2015).

In the HALO pilot randomized trial (n = 76), where patients with septic shock were randomized to receive therapeutic dose IV UFH for the treatment of VTE or dalteparin for venous thromboprophylaxis, two patients (6%, 95%CI 1 to 11%) randomized to IV UFH and 1 patient (3%, 95%CI 1 to 7%) randomized to dalteparin experienced major bleeding. None of these bleeding events were adjudicated to contribute to morbidity or mortality.

Overall, the rate of bleeding may not be significantly different between unselected critically ill patients receiving low dose thromboprophylaxis and selected patients receiving therapeutic dose heparin or LMWH.

The incidence of heparin-induced thrombocytopenia with LMWH and UFH when administered to general medical-surgical ICU patients is approximately 0.3 to 0.6% (Group et al., 2011). Heparin- induced thrombocytopenia occurs significantly less often in patients receiving low molecular weight heparin compared with UFH (RR 0.22, 95% CI 0.06 to 0.84) (Junqueira et al., 2017). The overall incidence of HIT is 0.2–0.5%, and is higher in patients receiving therapeutic doses of UFH (0.79%) compared to those receiving prophylactic doses (<0.1%) (Creekmore et al., 2006, Smythe et al., 2007).

### 7. DOMAIN OBJECTIVES

The objective of this domain is to determine the effectiveness of therapeutic anticoagulation for patients with acute illness due to suspected or proven pandemic infection.

We hypothesize that the probability of the occurrence of the primary endpoint specified in the relevant core protocol documents will differ based on allocation to different anticoagulation strategy. The following interventions will be available:

- Local standard venous thromboprophylaxis
- Therapeutic anticoagulation with intravenous unfractionated heparin or subcutaneous low molecular weight heparin

We hypothesize that the treatment effect of therapeutic anticoagulation is different

Not for IRB submission

depending on whether SARS-CoV-2 infection is confirmed to be present or absent.

We hypothesize that the treatment effect of therapeutic anticoagulation is different depending on the illness severity state at the time of enrollment.

We hypothesize that the treatment effect of therapeutic anticoagulation is different depending on D-dimer strata status.

### 8. TRIAL DESIGN

This domain will be conducted as part of the REMAP-CAP trial. Treatment allocation will be based on response adaptive randomization, as described in the core protocol documents.

#### 8.1. Population

The REMAP enrolls patients with acute illness due to suspected or proven COVID-19 admitted to hospital, including patients admitted to ICU.

#### 8.2. Eligibility criteria

Patients are eligible for this domain if they meet all of the platform-level inclusion and none of the platform-level exclusion criteria as specified in either the REMAP-CAP Core Protocol + Pandemic Appendix or the REMAP-COVID Core Protocol. Patients eligible for the REMAP may have conditions that exclude them from this specific COVID-19 Therapeutic Anticoagulation Domain.

This domain is available for patients who have acute illness due to suspected or proven pandemic infection in both the Moderate State and the Severe State.

##### 8.2.1. Domain inclusion criteria

Patients are eligible for this domain if:

- COVID-19 infection is suspected by the treating clinician or has been confirmed by microbiological testing (i.e. PISOP stratum)
- Microbiological testing for SARS-CoV-2 of upper or lower respiratory tract secretions or both has occurred or is intended to occur

##### 8.2.2. Domain exclusion criteria

Patients will be excluded from this domain if they have any of the following:

- More than 48 hours has elapsed since ICU admission (noting that this may be

Not for IRB Submission

operationalized as more than 48 hours has elapsed since commencement of sustained organ failure support)

- Clinical or laboratory bleeding risk or both that is sufficient to contraindicate therapeutic anticoagulation, including intention to continue or commence dual anti-platelet therapy
- Therapeutic anticoagulation is already present due to prior administration of any anticoagulant agent that is known or likely to still be active or a clinical decision has been made to commence therapeutic anticoagulation
- Enrolment in a trial evaluating anticoagulation for proven or suspected COVID-19 infection, where the protocol of that trial requires continuation of the treatment assignment specified in that trial
- Known or suspected previous adverse reaction to UFH or LMWH including heparin induced thrombocytopenia (HIT).
- The treating clinician believes that participation in the domain would not be in the best interests of the patient

###### 8.2.3. Intervention exclusion criteria

Nil.

#### *8.3. Anticoagulant Interventions*

###### 8.3.1. Anticoagulation interventions

Patients will be randomly assigned to receive either of the following open-label strategies. The interventions will be commenced immediately after allocation status is revealed.

☐ Local standard venous thromboprophylaxis

☐ Therapeutic anticoagulation with intravenous unfractionated heparin or subcutaneous low molecular weight heparin

Administration of venous thromboprophylaxis is based on local practice and is mandatory.

###### 8.3.2. Local standard venous thromboprophylaxis

Standard venous thromboprophylaxis that complies with local guidelines or usual practice will be administered for 14 days following randomization or until hospital discharge, whichever occurs first. The dose of agent that is chosen should not be sufficient to result in therapeutic anticoagulation.

After 14 days decisions regarding thromboprophylaxis and anticoagulation are at the discretion of the treating clinician.

Not for IRB Submission

###### 8.3.2.1. *Use of therapeutic anticoagulation in patients randomized to local standard venous thromboembolism*

Any patient who develops an accepted clinical indication for anticoagulation can have this treatment commenced by the treating clinician. Such indications include, but are not limited, to proven deep venous thrombosis, proven PE, acute coronary syndrome, systemic embolic event, intermittent hemodialysis or sustained low-efficiency daily dialysis.

Systemic therapeutic anticoagulation for continuous renal replacement therapy is not permitted, unless there is an additional indication for anticoagulation. Regional citrate, heparin priming and low-dose heparin administration (without measurable systemic anticoagulation) are permitted for continuous renal replacement therapy. If regional low-dose heparin administration is used to facilitate continuous renal replacement therapy, the dose may be increased as necessary to prevent clotting of the filter, however the dose of heparin should be minimized as much as possible.

##### 8.3.3. Therapeutic Anticoagulation

The patient will be administered either UFH or LMWH to achieve systemic anticoagulation. Either agent may be used and the same patient may be switched between UFH and LMWH at the discretion of the treating clinician

###### 8.3.3.1. *Unfractionated heparin*

If UFH is used, this is commenced, administered, and monitored according to local hospital policy, and guidelines that are used for the treatment of VTE (i.e. not for acute coronary syndrome). The target aPTT should typically be in the range of 1.5 to 2.5 times the upper limit of normal at the participating site. Alternately, therapeutic anti-Xa values (i.e. values targeted for the treatment of acute VTE) can be targeted based on local practice. If UFH is used, the availability of a local hospital policy that has specifies an aPTT target in this range or an anti-Xa value is a requirement. Based on an assessment of risk of administration of a loading dose, an initial bolus of UFH may be withheld at the discretion of the treating clinician.

###### 8.3.3.2. *Low molecular weight heparin*

Not for IRB Submission

LMWH is commenced, administered, and monitored according to local hospital policy, practice and guidelines that pertain to treatment of VTE (i.e. not thromboprophylactic doses). The dose selected should be based on measure or estimated weight of the patient.

Adjustment for impairment of renal function should be according to local practice and policy.

###### 8.3.3.3. *Duration of therapeutic anticoagulation*

The duration of therapeutic anticoagulation is 14 days. For patients who are discharged from hospital before 14 days, therapeutic anticoagulation should be ceased prior to hospital discharge. For patients admitted to an ICU therapeutic anticoagulation may be ceased before 14 days at the discretion of the treating clinician at ICU discharge but, during the 14 day period, all patients receiving invasive mechanical ventilation should receive therapeutic anticoagulation until at least 24 hours after cessation of mechanical ventilation.

After 14 days decisions regarding thromboprophylaxis and anticoagulation are at the discretion of the treating clinician.

###### 8.3.4. Discontinuation of study intervention

Anticoagulation or local standard venous thromboprophylaxis should be discontinued if there is clinical bleeding or other complication sufficient to warrant cessation in the opinion of the treating clinician. Major bleeding, including death due to bleeding, is an SAE. Anticoagulation or local standard venous thromboprophylaxis may be recommenced if deemed appropriate by the treating clinician.

Occurrence of laboratory proven HIT must result in cessation UFH or LMWH without recommencement regardless of treatment assignment. Use of an acceptable alternative agent is required in this instance as clinically indicated. Occurrence of laboratory proven HIT is an SAE.

The study interventions can be discontinued at any time by the treating clinician if doing so is regarded as being in the best interests of the patient. Temporary cessation – for the shortest period of time possible, but not longer than 24 hours - such as to allow surgical or other procedures is not a protocol deviation.

Temporary or permanent cessation of the study interventions for bleeding is not a protocol deviation.

Not for IRB submission

###### 8.3.5. COVID-19 anticoagulation strategy in patients negative for COVID-19 infection

In patients with suspected COVID-19 infection who receive an allocation status to receive active anticoagulation but who subsequently test negative for COVID-19 infection may have treatment ceased unless the treating clinician believes that doing so is not clinically appropriate. This decision should take into account the known or suspected local population incidence of COVID-19 infection among critically ill patients and sensitivity of testing for COVID-19 infection.

#### 8.4. Concomitant care

Additional agents, other than those specified in the platform, that are intended to modify the patient's coagulation function as a treatment for COVID-19 infection should not be administered. A patient who receives an agent that act to inhibit platelet function as a usual medication may have this medication continued. Commencement of any new agent that inhibits platelet function is not permitted unless there is an accepted clinical indication such as an acute coronary syndrome, ischemic stroke or transient ischemic event or the agent that inhibits platelet function has been specified in another domain of this platform.

All other treatment that is not specified by assignment within the platform will be determined by the treating clinician.

#### 8.5. Endpoints

##### 8.5.1. Primary endpoint

The primary endpoint for this domain is the primary outcome specified in the REMAP-CAP Core Protocol + Pandemic Appendix or REMAP-COVID Core Protocol.

##### 8.5.2. Secondary endpoints

All secondary endpoints as specified in the REMAP-CAP Core Protocol + Pandemic Appendix or REMAP-COVID Core Protocol.

The domain-specific secondary outcome measures (from randomization, during the index hospitalization, censored 90 days after enrollment) will be:

- Serial detection of SARS-CoV-2 in upper or lower respiratory tract specimens (using only specimens collected for routine clinically indicated testing)
- Confirmed deep venous thrombosis
- Confirmed pulmonary embolism

Not for IRB Submission

- Confirmed ischemic cerebrovascular event
- Total red cell blood cell units transfused between randomization and the end of study day 15
- Confirmed acute myocardial infarction

- Peak troponin between randomization and the end of study day 15
- Major bleeding
- Other confirmed thrombotic event including mesenteric ischemia and limb ischemia
- SAE as defined in Core Protocol and this DSA below

### 9. TRIAL CONDUCT

#### 9.1. Microbiology

Microbiological testing will be performed as per local practice, including bacterial and viral testing to guide clinical care. Results of these tests will be collected but no additional testing is specified in this protocol.

Sites that are participating in this domain are encouraged to also participate in the Clinical Characterization Protocol (CCP) for patients with COVID-19 that has been established by the International Severe Acute Respiratory and Emerging Infectious Consortium (https://isaric.tghn.org/CCP/). This protocol specifies the collection of biological samples from patients with COVID-19. Samples collected in patients who are enrolled in the CCP may be made available to REMAP-CAP investigators to evaluate aspects of host or pathogen biology associated with assignment in this domain. Ethical approval at such sites and agreement from patients to undertake the CCP will be obtained separately.

#### 9.2. Domain-specific data collection

Additional domain-specific data will be collected.

- Baseline measures of coagulation including d-dimer
- Administration of anticoagulant agents
- Administration of agents that inhibit platelet function
- Transfusion of red cells
- Peak troponin
- Acute myocardial infarction (using fourth international definition)
- Major bleeding (using the International Society on Thrombosis and Haemostasis definition)
- Mesenteric Ischemia, limb ischemia, and other clotting events

Not for IRB submission

##### 9.3. Criteria for discontinuation

Refer to relevant core protocol documents for criteria for discontinuation of participation in the REMAP-CAP trial.

##### 9.4. Blinding

###### 9.4.1. Blinding

All medication will be administered on an open-label basis.

###### 9.4.2. Unblinding

Not relevant.

#### 10. STATISTICAL CONSIDERATIONS

##### 10.1. Domain-specific stopping rules

The Platform Conclusion of equivalence in this domain will not be evaluated. Instead a Platform Conclusion of Futility will be considered. If the posterior probability of at least a 20% odds-ratio increase for therapeutic anticoagulation is less than 5% then therapeutic anticoagulation will be declared Futile as a Platform Conclusion. This rule corresponds to the one-sided equivalency region.

In all other respects the stopping rules for this domain are those outlined in the relevant core protocol documents.

##### 10.2. Unit-of-analysis and strata

This domain is analyzed only in the pandemic statistical model and includes only patients who are in the pandemic suspected or proven stratum, as specified in the REMAP-CAP Pandemic Appendix and corresponding to the eligibility criteria specified in the REMAP-COVID Core Protocol. Within this stratum, the unit-of-analysis is defined by illness severity state at time of enrollment, defined as either Moderate State or Severe State. Unit-of-analysis may also be defined by SARS-CoV-2 infection or d-dimer strata or both. The D-dimer strata will contain 3 stratum, the breakpoints of which will be determined not later than the first interim analysis

Not for IRB submission

using data derived from patients enrolled in REMAP-CAP as well as any other trials that may utilize the same statistical model. Borrowing is permitted between states and strata. If the SARS-CoV-2 strata is applied in analysis, Response

Adaptive Randomization will be applied to all PISOP patients, in each illness severity state, using probabilities derived from the SARS-CoV-2 confirmed stratum. Response Adaptive Randomization may also be applied according to D-dimer strata status. The decision to apply the SARS-CoV-2 and D- dimer strata will be operational.

At the time of a Platform Conclusion, results will be reported for all randomized patients, patients in whom COVID-19 infection is confirmed by microbiological testing, microbiological tests do not detect or isolate COVID-19 infection, and testing is not performed.

The shock strata will not contribute to unit-of-analysis for this domain, as this strata is not applied in the Pandemic Statistical Model.

The influenza strata will not contribute to unit-of-analysis for this domain.

##### 10.3. *Timing of revealing of randomization status*

The timing of the revealing of allocation status and administration of interventions is specified to be Randomization with Immediate Reveal and Initiation or Randomization with Deferred Reveal if prospective agreement to participate is required for this domain (see relevant core protocol documents).

##### 10.4. *Interactions with interventions in other domains*

An *a priori* interaction with the Antibiotic Domain is not able to be evaluated as analysis occurs in different statistical models.

An *a priori* interaction with the Macrolide Duration Domain is not considered possible will not be incorporated into the statistical models used to analyze this domain.

An *a priori* interaction with the Influenza Antiviral Domain is not able to be evaluated as analysis occurs in different statistical models.

An *a priori* interaction with the Corticosteroid Domain is not considered possible will not be incorporated into the statistical models used to analyze this domain.

Not for IRB Submission

An *a priori* interaction with the COVID-19 Immune Modulation Domain is not considered possible and will not be incorporated into the statistical models used to analyze this domain.

An *a priori* interaction with the COVID-19 Antiviral Domain is not considered possible and will not be incorporated into the statistical models used to analyze this domain.

An *a priori* interaction with the COVID-19 Statin Domain is not considered possible and will not be incorporated into the statistical models used to analyze this domain.

An *a priori* interaction with the Vitamin C Domain is either not considered possible and will not be incorporated into the statistical model used to evaluate this domain in the pandemic statistical model or is not able to be evaluated for PINSNP patients as analysis occurs in different statistical models.

No interaction is evaluable between the Ventilation Domain and this domain.

##### 10.5. Nesting of interventions

Nesting is not applicable to this domain.

##### 10.6. Threshold probability for superiority and inferiority

The threshold odds ratio delta for superiority and inferiority in this domain are those specified in the Operating Characteristics document derived from Pandemic Appendix and the REMAP-COVID Core Protocol. It is noted that the threshold for superiority and inferiority in the current model has been modified from 0.95 to 0.99 to provide adequate control of type I error, following the evaluation of simulations. It is also noted that asymmetric probabilities may be specified for harm, to allow early cessation and declaration of a Platform Conclusion for interventions that are unlikely to be effective and may be harmful. If so, this will be specified in the Operating Characteristics document which is placed in the public domain.

##### 10.7. Threshold odds ratio delta for equivalence

The Platform Conclusion of equivalence will not be evaluated in this domain. The same odds ratio delta as specified in the relevant core protocol documents for equivalence will be used for futility. This will be applied in a one-sided analysis for futility of therapeutic anticoagulation

##### 10.8. Informative priors

Not for IRB Submission

This domain will launch with priors that are not informative for main effects.

##### 10.9. *Post-trial sub-groups*

Domain-specific post-hoc sub-groups will be used in analysis following the conclusion of one or more interventions within the domain. The *a priori* patient sub-groups of interest are:

- Proven concomitant bacterial co-infection, defined as having isolation or detection of a known pathogen that causes CAP from blood, pleural fluid, or lower respiratory tract specimen
- Whether therapeutic anticoagulation is initiated with UFH or LMWH
- Shock strata
- Receiving invasive mechanical ventilation at baseline
- Baseline troponin
- All remaining potentially evaluable treatment-by-treatment interactions with other domains

#### 11. ETHICAL CONSIDERATIONS

##### 11.1. *Data Safety and Monitoring Board*

The DSMB should be aware that the superiority, efficacy, inferiority, or futility of different interventions with respect to the primary endpoints are possible.

The DSMB should take into account the public health, as well as clinical significance, of the analyses of this domain and are empowered to discuss results with relevant international and national public health authorities, with rapid dissemination of results to the larger community being the goal.

Safety secondary outcomes will be reported to the DSMB who are empowered to require additional analyses regarding these outcomes as required.

##### 11.2. *Potential domain-specific adverse events*

For patients assigned to any intervention, occurrence of any of the following should be reported as an SAE

- Laboratory proven heparin-induced thrombocytopenia

Not for IRB submission

Other SAEs should be reported only where, in the opinion of the site-investigator, the event might reasonably have occurred as a consequence of a study intervention or study participation (see relevant core protocol documents).

##### 11.3. Domain-specific consent issues

As noted in the background, and endorsed by the WHO, in the absence of evidence of effectiveness of anticoagulation for COVID-19, the use of a usual care control is both appropriate and ethical.

Both forms of anticoagulation are being used, off-trial, and typically without consent, for patients with proven or suspected COVID-19 infection. Clinicians may choose not to enroll individual patients if they feel that participation is not in patient's best interests, and safety criteria are used to exclude patients from this domain for appropriate clinical reasons.

Where all interventions that are available at a participating site and are regarded as being part of the acceptable spectrum of standard care and given the time imperative necessary to evaluate these interventions, entry to the study, for participants who are not competent to consent, is preferred to be via waiver-of-consent or some form of delayed consent.

During a pandemic, visiting by relatives of affected patients may not be possible. In such situations, alternative methods for confirming consent including electronic and telephone communication, as permitted by an appropriate ethical review body, may be acceptable methods for confirming agreement to participate in this (and other) domains of the platform.

##### 11.4. Relationship to Antiplatelet Domain

An Antiplatelet Domain of REMAP-CAP is being planned currently. If such a domain is implemented, it is intended that the Antiplatelet Domain and the Therapeutic Anticoagulation Domain will be analyzed as a 2 x N factorial, with N interventions being available within the Antiplatelet Domain.

#### 12. GOVERNANCE ISSUES

##### 12.1. Funding of domain

Funding sources for the REMAP-CAP trial are specified in the Core Protocol Section 2.5. This domain has not received any additional domain-specific funding but such

Not for IRB submission

funding, from any source, may be obtained during the life-time of the domain.

#### *12.2. Funding of domain interventions and outcome measures*

All anticoagulant agents will be provided by participating hospitals. The cost of all agents specified in this domain are known to be inexpensive.

##### 12.3. *Domain-specific declarations of interest*

All investigators involved in REMAP-CAP maintain a registry of interests on the REMAP-CAP website. These are updated periodically and publicly accessible on the study website.

Not for IRB submission

Not for IRB submission

2019 infection. *Clin Chem Lab Med*.

LIU, X., LIU, X., XU, Y., XU, Z., HUANG, Y., CHEN, S., LI, S., LIU, D., LIN, Z. & LI, Y. 2020. Ventilatory Ratio in Hypercapnic Mechanically Ventilated Patients with COVID-19 Associated ARDS. *Am J Respir Crit Care Med*.

MISMETTI, P., QUENET, S., LEVINE, M., MERLI, G., DECOUSUS, H., DEROBERT, E. & LAPORTE, S. 2005. Enoxaparin in the treatment of deep vein thrombosis with or without pulmonary embolism: an individual patient data meta-analysis. *Chest,* 128**,** 2203-10.

MOULARD, M., LORTAT-JACOB, H., MONDOR, I., ROCA, G., WYATT, R., SODROSKI, J., ZHAO, L., OLSON, W., KWONG, P. D. & SATTENTAU, Q. J. 2000. Selective interactions of polyanions with basic surfaces on human immunodeficiency virus type 1 gp120. *J Virol,* 74**,** 1948-60.

NAVARESE, E. P., ANDREOTTI, F., KOLODZIEJCZAK, M., SCHULZE, V., WOLFF, G., DIAS, S., CLAESSEN, B., BROUWER, M., TARANTINI, G., ILICETO, S., BROCKMEYER, M., KOWALEWSKI, M., LIN, Y., EIKELBOOM, J., MUSUMECI, G., LEE, L., LIP, G. Y., VALGIMIGLI, M., BERTI, S. & KELM, M. 2015. Comparative efficacy and safety of anticoagulant strategies for acute coronary syndromes. Comprehensive network meta-analysis of 42 randomised trials involving 117,353 patients. *Thromb Haemost,* 114**,** 933-44.

PETERSEN, J. L., MAHAFFEY, K. W., HASSELBLAD, V., ANTMAN, E. M., COHEN, M., GOODMAN, S. G., LANGER, A., BLAZING, M. A., LE-MOIGNE-AMRANI, A., DE LEMOS, J. A., NESSEL, C. C., HARRINGTON, R. A., FERGUSON, J. J., BRAUNWALD, E. & CALIFF, R. M. 2004. Efficacy and
bleeding complications among patients randomized to enoxaparin or unfractionated heparin for antithrombin therapy in non-ST-Segment elevation acute coronary syndromes: a systematic overview. *JAMA,* 292**,** 89-96.

POLDERMAN, K. H. & GIRBES, A. R. 2004. Drug intervention trials in sepsis: divergent results. *Lancet,*
363**,** 1721-3.

SMYTHE, M. A., KOERBER, J. M. & MATTSON, J. C. 2007. The incidence of recognized heparin- induced thrombocytopenia in a large, tertiary care teaching hospital. *Chest,* 131**,** 1644-9.

TANG, N., BAI, H., CHEN, X., GONG, J., LI, D. & SUN, Z. 2020a. Anticoagulant treatment is associated with decreased mortality in severe coronavirus disease 2019 patients with coagulopathy. *J Thromb Haemost*.

TANG, N., LI, D., WANG, X. & SUN, Z. 2020b. Abnormal coagulation parameters are associated with poor prognosis in patients with novel coronavirus pneumonia. *J Thromb Haemost,* 18**,** 844- 847.

TIRYAKI, F., NUTESCU, E. A., HENNENFENT, J. A., KARAGEANES, A. M., KOESTERER, L. J., LAMBERT, B.
L. & SCHUMOCK, G. T. 2011. Anticoagulation therapy for hospitalized patients: patterns of use, compliance with national guidelines, and performance on quality measures. *Am J Health Syst Pharm,* 68**,** 1239-44.

VICENZI, E., CANDUCCI, F., PINNA, D., MANCINI, N., CARLETTI, S., LAZZARIN, A., BORDIGNON, C.,
POLI, G. & CLEMENTI, M. 2004. Coronaviridae and SARS-associated coronavirus strain HSR1.
*Emerg Infect Dis,* 10**,** 413-8.

WANG, D., HU, B., HU, C., ZHU, F., LIU, X., ZHANG, J., WANG, B., XIANG, H., CHENG, Z., XIONG, Y.,
ZHAO, Y., LI, Y., WANG, X. & PENG, Z. 2020a. Clinical Characteristics of 138 Hospitalized Patients With 2019 Novel Coronavirus–Infected Pneumonia in Wuhan, China. *JAMA*.

WANG, L., HE, W. B., YU, X. M., LIU, H. F., ZHOU, W. J. & JIANG, H. 2020b. [Prognostic value of
myocardial injury in patients with COVID-19]. *Zhonghua Yan Ke Za Zhi,* 56**,** E009.

WEBB, S. A. 2015. Putting Critical Care Medicine on Trial. *Crit Care Med,* 43**,**

Not for IRB submission

1767-8. WILDHAGEN, K. C., GARCIA DE FRUTOS, P., REUTELINGSPERGER,

C. P., SCHRIJVER, R., ARESTE, C., ORTEGA-GOMEZ, A., DECKERS, N. M., HEMKER, H. C., SOEHNLEIN, O. & NICOLAES, G. A. 2014. Nonanticoagulant heparin prevents histone-mediated cytotoxicity in vitro and improves survival in sepsis. *Blood,* 123**,** 1098-101.

WU, A., HINDS, C. J. & THIEMERMANN, C. 2004. High-density lipoproteins in sepsis and septic shock: metabolism, actions, and therapeutic applications. *Shock,* 21**,** 210-21.

WU, C., CHEN, X., CAI, Y., XIA, J., ZHOU, X., XU, S., HUANG, H., ZHANG, L., ZHOU, X., DU, C., ZHANG, Y., SONG, J., WANG, S., CHAO, Y., YANG, Z., XU, J., ZHOU, X., CHEN, D., XIONG, W., XU, L., ZHOU, F., JIANG, J., BAI, C., ZHENG, J. & SONG, Y. 2020. Risk Factors Associated With Acute Respiratory Distress Syndrome and Death in Patients With Coronavirus Disease 2019 Pneumonia in Wuhan, China. *JAMA Intern Med*.

WU, Z. & MCGOOGAN, J. M. 2020. Characteristics of and Important Lessons From the Coronavirus Disease 2019 (COVID-19) Outbreak in China: Summary of a Report of 72314 Cases From the Chinese Center for Disease Control and Prevention. *JAMA*.

ZARYCHANSKI, R., DOUCETTE, S., FERGUSSON, D., ROBERTS, D., HOUSTON, D. S., SHARMA, S., GULATI, H. & KUMAR, A. 2008. Early intravenous unfractionated heparin and mortality in septic shock. *Crit Care Med,* 36**,** 2973-9.

ZHANG, H., PENNINGER, J. M., LI, Y., ZHONG, N. & SLUTSKY, A. S. 2020. Angiotensin-converting enzyme 2 (ACE2) as a SARS-CoV-2 receptor: molecular mechanisms and potential therapeutic target. *Intensive Care Med,* 46**,** 586-590.

ZHOU, F., YU, T., DU, R., FAN, G., LIU, Y., LIU, Z., XIANG, J., WANG, Y., SONG, B., GU, X., GUAN, L., WEI, Y., LI, H., WU, X., XU, J., TU, S., ZHANG, Y., CHEN, H. & CAO, B. 2020. Clinical course and risk factors for mortality of adult inpatients with COVID-19 in Wuhan, China: a retrospective cohort study. *Lancet,* 395**,** 1054-1062.

Not for IRB Submission

### 14. APPENDIX 1. OVERVIEW OF DESIGN AND INITIAL RESULTS FOR THE THERAPEUTIC ANTICOAGULATION DOMAIN

#### 14.1. *Introduction*

This document describes the statistical design and analysis of the testing of therapeutic anticoagulation with intravenous UFH or subcutaneous LMWH compared to local standard venous thromboprophylaxis in the COVID-19 appendix as part of the REMAP-CAP trial. Our goal is to investigate whether this is independently beneficial in increasing the number of ICU- free days for patients with COVID-19.

##### 14.1.1. Treatment Arms

The main effect for therapeutic anticoagulation in this domain will be modeled as specified in the PAtC.

##### 14.1.2. Primary Endpoint

The primary efficacy endpoint is as specified in the PAtC, the ordinal endpoint, ICU-free days through 21 days with the classification of in hospital death as the worst outcome.

#### 14.2. *Primary Analysis Model*

The primary analysis is based on a Bayesian cumulative logistic regression assuming proportional odds for intervention effects (reference the PAtC stats document??).

##### 14.2.1. Domain Platform Conclusions.

The Platform Conclusions of Superiority and Inferiority are as specified in the PAtC and are unchanged.

This domain substitutes a Platform Conclusion of Futility in place of Equivalence for this domain as demonstration of equivalence in not relevant but a conclusion of Futility of therapeutic anticoagulation is relevant. If the probability of at least a 20% odds ratio improvement for therapeutic anticoagulation is less than 5% then the

Not for IRB Submission

Statistical Trigger for Futility will have been met. This Futility trigger is the one-sided extension of the equivalence rule in PAtC. That is, Futility of therapeutic anticoagulation will be declared if $Pr(OR_1 > 1.2) < 0.05$, where $OR_1$ refers to the odd ratio for therapeutic anticoagulation compared to SOC for this domain.

#### 14.3. Simulation Details

In this section, we outline the simulations conducted for understanding the performance of this domain. Simulations were conducted separately assuming only this domain, as there are no interactions with any other domains.

##### 14.3.1. Standard-of-Care Rates and therapeutic anticoagulation effect assumptions

We created possible standard-of-care rates across the 23 levels of the outcome. We worked within a few clinically guided expected parameters: 20% mortality rate, 10% of patients are in the ICU 21 days, and median number of days in the ICU is 7 amongst those that did not die. Figure 1 shows the assumed rates for the ICU-free day endpoint in the left panel.

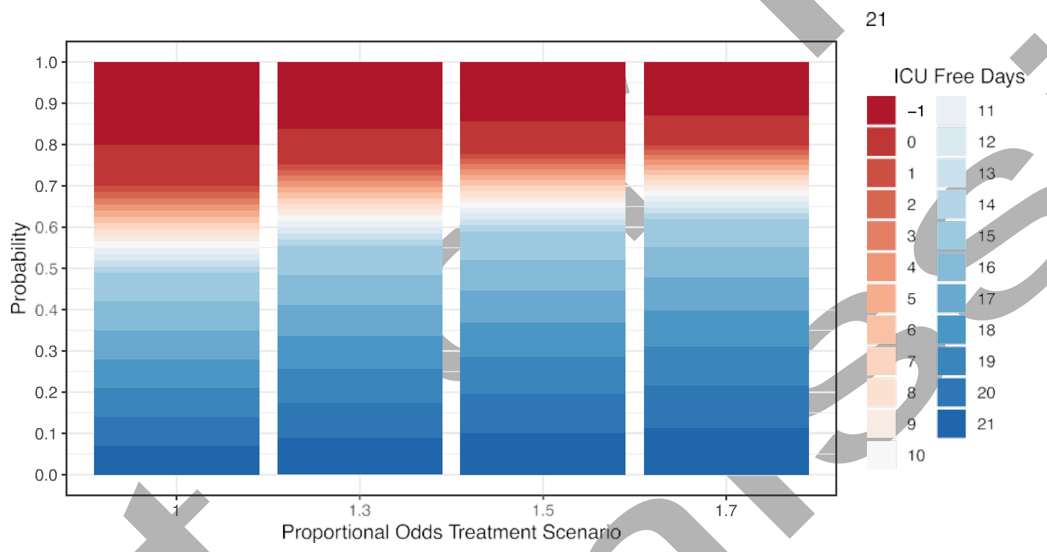

*Figure 1. Control outcome probabilities for the ICU-free day end point (left panel) and then the probabilities for treatment effects of odds ratios of 1.3, 1.5, and 1.7.*

For the simulations in this section interim analyses are assumed to occur at 200, 400, 600, 800, 1000, 1500, 2000, 2500, and 3000 patients enrolled in this domain.

#### 14.4. Operating Characteristics

Figure 2 presents the cumulative power to determine that therapeutic anticoagulation is superior to the standard-of-care intervention as a function of the total number of patients enrolled (x-axis) and the assumed effect sizes (1.3, 1.5, and 1.7).

Not IRB Submission

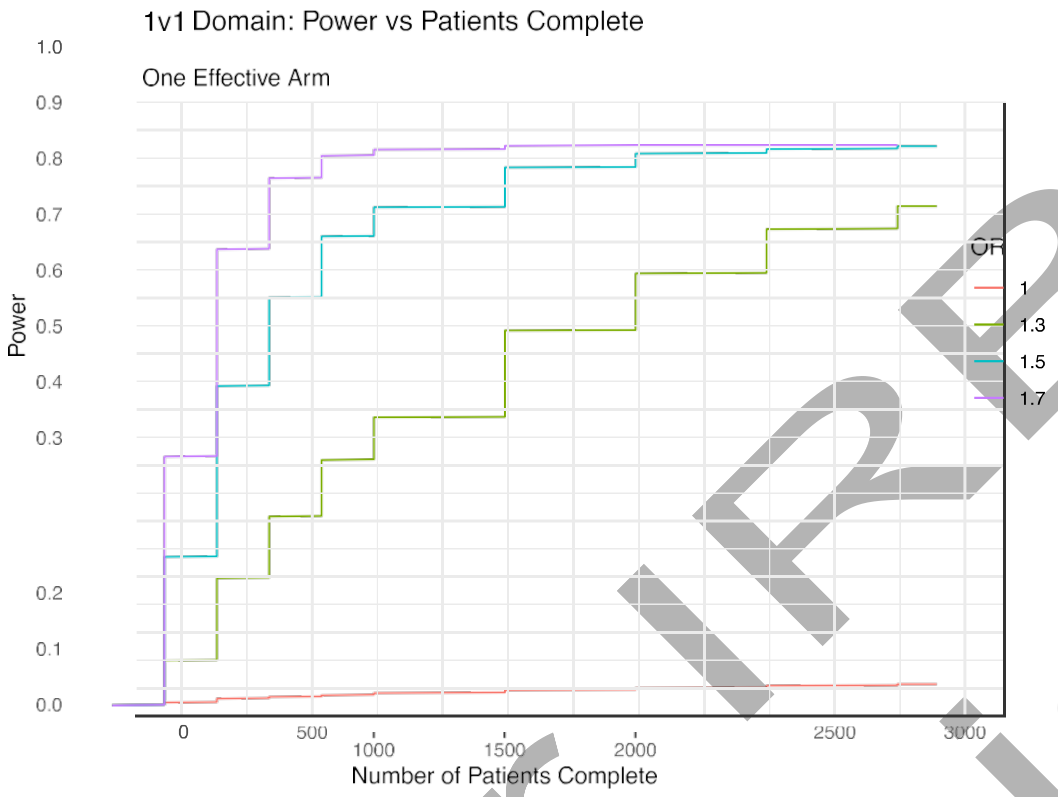

*Figure 2: The cumulative power for each of the explored treatment effects (odds ratios of 1.3, 1.5, and 1.7). The cumulative type I error is shown as the red line (effect size of 1).*

#### 14.5. Summary

The domain is designed to provide high-level evidence. The domain has 80% power to demonstrate superiority of therapeutic anticoagulation to standard-of-care by 400 patients enrolled assuming an odds ratio effect size of 1.7. For an effect size of 1.5 the power is 80% for 800 patients enrolled. The cumulative type I error through 3000 patients is less than 5%.

Not for IRB Submission

### Protocol 2: Antithrombotic Therapy to Ameliorate Complications of COVID-19 (ATTACC)

#### ANTITHROMBOTIC THERAPY TO AMELIORATE COMPLICATIONS OF COVID-19 (ATTACC), IN COLLABORATION WITH ACCELERATING COVID-19 THERAPEUTIC INTERVENTIONS AND VACCINES (ACTIV-4)

Protocol Version #: 3.0

Protocol Date: 29-Sep-2020

Study Number: OZM-113

Sponsor: Dr. Ryan Zarychanski, University of Manitoba

Sponsor's Address: ON 2051 – 675 McDermot Avenue
Winnipeg, Manitoba, Canada R3E 0V9


Co-Principal Investigators: Ryan Zarychanski, MD MSc
Patrick R. Lawler, MD MPH
Ewan Goligher, MD PhD

Lead Investigator, United States; Robert Rosenson

Lead Investigator, Brazil; Jose Nicolau

Lead Investigator, Mexico: Jorge Escobedo

Steering Committee: Charlotte Bradbury, MD
Marc Carrier, MD, MSc
Vlad Dzavik, MD
Michael Farkouh, MD
Dean Fergusson, PhD MHA
Robert Fowler, MD MSc
Emily Gibson McDonald, MD MSc
Peter Gross, MD MSc
Brett L Houston, MD

Mansoor Husain, MD
Susan Kahn, MD MSc
Anand Kumar, MD
John Marshall, MD
Srinivas Murthy, MD
Arthur Slutsky, MD MSc

Tobias Tritschler MD
Alexis Turgeon, MD

Patient Partners : Suzanne Dubois
Margaret Ostrowski

Protocol History:

Version 1.0; dated 23-APR-2020
Version 2.0; dated 11-MAY-2020

Clinical Trial Management/Clinical Trials Specialist

Lindsay Bond
Ozmosis Research Inc. 65 Queen Street West Suite 2020, Toronto, ON M5H 2M5
Main Line: 416-634-8300


#### Investigator Initiated Sponsor's Agreement to Protocol Version 3.0, 29-Sep-2020

Name of Authorized Personnel (Print) ______________________________

Title of Authorized Personnel (Print) ______________________________
______________________________
______________________________

Signature of Authorized Personnel: ______________________________

Date of Approval: ______________________________
DD-MMM-YYYY

### 1 SYNOPSIS

| | |
|---|---|
| Study Title: | **A**nti**T**hrombotic **T**herapy to **A**meliorate **C**omplications of **COVID**-19 (**ATTACC**) in collaboration with Accelerating COVID-19 Therapeutic Interventions and Vaccines (ACTIV-4) |
| Study Design: | A phase III prospective, open-label, adaptive multi-platform randomized controlled trial |
| Primary Objective/ Endpoint: | The primary endpoint in the trial is days alive and free of organ support at day 21. This endpoint is defined as the number of days that a patient is alive and free of organ support through the first 21 days after trial entry. |
| Secondary Objectives: | Secondary Safety Endpoints:<br>- Laboratory confirmed heparin induced thrombocytopenia **(HIT)**<br>- **Major bleeding**, defined according to the International Society on Thrombosis and Haemostasis (ISTH)/Scientific and Standardization Committee (SSC) definitions and bleeding assessment tool in non-surgical patients (Schulman *J Thromb Haemost* 2005): fatal bleeding; and/or symptomatic bleeding in a critical area or organ, such as intracranial, intraspinal, intraocular, retroperitoneal, intraarticular or pericardial, or intramuscular with compartment syndrome; and/or bleeding causing a fall in hemoglobin level of ≥20 g/L, or leading to transfusion of 2 or more units of whole blood or red cells.<br><br>Secondary Efficacy Endpoints:<br>- A composite endpoint of death, deep vein thrombosis, pulmonary embolism, systemic arterial thromboembolism, myocardial infarction, or ischemic stroke collected during hospitalization or at 28 days and 90 days after enrollment (whichever is earlier)<br>- Ordered categorical endpoint with three possible outcomes based on the worst status of each patient through day 30 following randomization: no invasive mechanical ventilation, invasive mechanical ventilation, or death. |

| | |
|---|---|
| | - All cause mortality assessed at 28 and 90 days following randomization<br>- All cause mortality during initial hospitalization (includes death after 28 days)<br>- Intubation assessed at 30 days following randomization<br>- Ventilator-free days (days alive not on a ventilator) assessed at 28 days following randomization<br>- Hospital-free days (days alive outside hospital assessed at 28 days following randomization)<br>- Vasopressor-free days (days alive not on a vasopressor) assessed at 28 days following randomization<br>- Renal replacement free days (days alive not on renal replacement) assessed at 28 days following randomization<br>- Hospital re-admission within 28 days<br>- Symptomatic proximal venous thromboembolism (DVT or PE) assessed at 28 and 90 days following randomization<br>- Myocardial infarction assessed at 28 and 90 days following randomization<br>- Ischaemic stroke assessed at 28 and 90 days following randomization<br>- Acute kidney injury as defined by KDIGO criteria<br>- Systemic arterial thrombosis or embolism assessed at 28 and 90 days following randomization<br>- Use of extracorporeal membrane oxygenation (ECMO) support<br>- Mechanical circuit (dialysis or ECMO) thrombosis<br>- WHO ordinal scale (peak scale over 28 days, scale at 14 days, and proportion with improvement by at least 2 categories compared to enrollment, at 28 days) |
| Duration: | The duration of accrual on this study will be ongoing in nature during the COVID-19 pandemic, following outcomes for each patient up to a maximum of 90 days. |

| | |
|---|---|
| Planned Total Sample Size: | The trial is a Bayesian adaptive design and as such is not predicated on a fixed *a priori* sample size. This design was chosen given uncertainty regarding anticipated event rates and potential treatment effect sizes. Approximately 350 to a maximum of 3000 evaluable patients are anticipated to be enrolled in this adaptive trial in combination with the ACTIV 4 and REMAP-CAP trials, with anticipated reprioritization of key subgroups (including D-dimer defined) as the trial is undertaken. |
| Drug Administration: | Participants randomized to the <u>investigational arm</u> will receive therapeutic anticoagulation for 14 days (or until hospital discharge or liberation from the need for supplemental oxygen, whichever comes first) with preference for low-molecular weight heparin (LMWH), or alternative unfractionated heparin (UFH).<br><br>Participants randomized to the <u>control arm</u> will receive usual care, which is anticipated to include thromboprophylactic dose anticoagulation according to local practice. |
| Inclusion/Exclusion Criteria: | Inclusions:<br>1. Patients ≥18 years of age providing (possibly through a substitute decision maker) informed consent who require hospitalization anticipated to last ≥72 hours, for microbiologically-confirmed COVID-19, enrolled < 72 hours of hospital admission **or** of COVID-19 confirmation<br><br>Exclusions:<br>1. Requirement for chronic mechanical ventilation via tracheostomy prior to hospitalization<br>2. Patients for whom the intent is to not use pharmacologic thromboprophylaxis<br>3. Active bleeding<br>4. Risk factors for bleeding, including:<br>a. intracranial surgery or stroke within 3 months;<br>b. history of intracerebral arteriovenous malformation;<br>c. cerebral aneurysm or mass lesions of the central nervous system; |

| | |
|---|---|
| | d. intracranial malignancy<br>e. history of intracranial bleeding<br>f. history of bleeding diatheses (e.g., hemophilia)<br>g. history of gastrointestinal bleeding within previous 3 months<br>h. thrombolysis within the previous 7 days<br>i. presence of an epidural or spinal catheter<br>j. recent major surgery <14 days<br>k. uncontrolled hypertension (sBP >200 mmHg, dBP >120 mmHg)<br>l. other physician-perceived contraindications to anticoagulation<br>5. Platelet count <50 x10$^9$/L, INR >2.0, or baseline aPTT >50<br>6. Hemoglobin <80 g/L (to minimize the likelihood of requiring red blood cell transfusion if potential bleeding were to occur)<br>7. Acute or subacute bacterial endocarditis<br>8. History of heparin induced thrombocytopenia (HIT) or other heparin allergy including hypersensitivity<br>9. Current use of dual antiplatelet therapy<br>10. Patients with an independent indication for therapeutic anticoagulation<br>11. Patients in whom imminent demise is anticipated and there is no commitment to active ongoing intervention<br>12. Anticipated transfer to another hospital that is not a study site within 72 hours<br>13. Enrollment in other trials related to anticoagulation or antiplatelet therapy |
| Study Assessments: | Study assessments are depicted in the study schedule. |
| Safety Variables & Analysis: | The safety of therapeutic anticoagulation with LWMH or intravenous UFH infusion will be evaluated by AE reports. Treatment-related AEs include bleeding and HIT. |
| Efficacy Assessments & Analysis | The efficacy of therapeutic-dose parenteral anticoagulation with subcutaneous LMWH or |

| | |
|---|---|
| | intravenous UFH will be evaluated in comparison to usual care. |
| Reasons for premature discontinuation of therapy: | Treatment will continue until any of the following occurs:<br>• Heparin induced thrombocytopenia or other heparin allergy/hypersensitivity<br>• Thrombocytopenia (platelet count <50 $x10^9$/L)<br>• Major bleeding, defined based closely on the International Society on Thrombosis and Haemostasis (ISTH)/Scientific and Standardization Committee (SSC) definitions and bleeding assessment tool in non-surgical patients<br>• Coagulopathy associated with an elevated INR (e.g., >2.0) or hypofibrinogemia<br>• Following invasive procedures where heparin is deemed unsafe to re-institute<br>• Patients requiring systemic fibrinolytic therapy<br>• Treating physician discretion |
| Statistical Analysis: | Data will be analyzed by an intention to treat analysis for the primary analysis; a per-protocol analysis will also be completed as a secondary analysis. Patients who receive at least one dose of drug will be evaluable for safety and efficacy. Response-adaptive randomization based on D-dimer subgroups is embedded. |

### Table of Contents

**1 SYNOPSIS 4**

**2 List of Abbreviations 12**

**3 Background 14**

**4 Study Rationale15**

4.1 *ATTACC Trial Rationale Overview 15*

4.2 *Risk Stratification in Patients with COVID-19 15*

4.3 *Hypercoagulability in Patients with COVID-19 16*

4.4 *Managing Hypercoagulability in Patients with COVID-19 17*

4.5 *Other Potential Therapeutic Benefits of Heparin in COVID-19: Anti-viral and Anti-inflammatory Effects 17*

4.6 *Safety of Heparin 18*

4.7 *Background Summary 19*

**5 THE TRIAL 21**

5.1 *Study Design 21*

5.2 *Patient Population 21*

5.3 *Primary Endpoint 21*

5.4 *Secondary Endpoints 21*

**6 Patient Eligibility 23**

6.1 *Inclusion Criteria 23*

6.2 *Exclusion Criteria 23*

6.3 *Patient Consent 24*

6.4 *Patient Enrollment 24*

**7 Study Plan 25**

7.1 *Anticoagulation Administration 25*

7.2 *Study Schedule 25*

**8 Concomitant Medications 28**

8.1 *Drug Interactions 28*

**9 Medicinal Product 29**

9.1 *Characterization of Investigation Medicinal Product 29*

9.2 *Accountability and Destruction 30*

9.3 *Dose Adjustments* 30
9.4 *Subject Compliance* 30
9.5 *Premature Withdrawal/ Discontinuation Criteria* 30
**10 OPTIONAL BIOREPOSITORY 32**
**11 Safety and Reporting Requirements 33**
11.1 *Adverse Event (ONLY study drug related AEs will be reported)* 33
11.2 *Adverse Event Documentation* 33
11.3 *Serious Adverse Event (SAE) due to the study drug* 33
11.4 *Reporting Serious Adverse Events (Only Study Drug Related SAEs will be reported)* 34
11.5 *Procedure for Expedited Reporting* 34
11.6 *Study Management and Governance* 34
**12 Statistical Analysis 36**
12.1 *Study Population* 36
12.2 *Evaluation of Safety* 36
12.3 *Trial Design Introduction* 36
12.4 *Primary Endpoint* 36
12.5 *Primary Analysis* 37
**13 Adaptive Design 39**
13.1 *Subgroup Conclusions* 39
**14 Publication polices and Disclosure of Data 40**
14.1 *Intellectual Property* 40
14.2 *Data Sharing* 40
**15 Ethics 41**
*Ethics Board Approval* 41
**16 Record Access and Maintenance of Study Records 42**
16.1 *Documentation of Subject's Participation* 42
16.2 *Regulatory Requirements* 42
16.3 *Subject Confidentiality and Access to Source Data/Documents* 42
16.4 *Confidentiality of the Study* 43
16.5 *Registration of Clinical Trial* 43
16.6 *Data Reporting* 43

16.7 *Maintenance of Study Records* *43*

17 Quality Assurance and Quality Control 45

17.1 Monitoring / Auditing 45

18 Administrative Procedures 46

18.1 Amendments to this Protocols 46

18.2 Protocol Deviations and Violations 46

18.3 Premature Discontinuation of the Study 46

19 References 47

20 Appendix A – FDA Guidance on Clinical Trial Conduct During COVID-19 Pandemic 51

21 Appendix B – KDIGO Criteria for acute kidney injury 52

22 Appendix C – WHO Ordinal scale for clinical improvement 53

#### 2 LIST OF ABBREVIATIONS

Ensure all abbreviations used within the protocol are listed here

| | |
|---|---|
| AE | Adverse Event |
| ACTIV | Accelerating COVID-19 Therapeutic Interventions and Vaccines |
| ATTACC | Antithrombotic Therapy to Ameliorate Complications of COVID-19 |
| aPTT | activated partial thromboplastin time |
| CCC | Clinical Coordinating Centre |
| CRF | Case Report Form |
| CRO | Clinical Research Organization |
| CTA | Clinical Trial Application |
| dBP | diastolic Blood Pressure |
| DCC | Data Coordinating Centre |
| DIC | Disseminated Intravascular Coagulation |
| DVT | Deep Vein Thrombosis |
| eCRF | Electronic Case Report Form |
| FDA | Food and Drug Administration |
| $FiO_2$ | Fraction of Inspired Oxygen |
| GCP | Good Clinical Practice |
| HIT | Heparin-Induced Thrombocytopenia |
| ICF | Informed Consent Form |
| ICH | International Council for Harmonisation of Technical Requirements for Pharmaceuticals for Human Use |
| ICU | Intensive Care Unit |
| IL-6 | Interlukin-6 |
| INR | International Normalized Ratio |
| IRB | Institutional Review Board |
| ISTH | International Society on Thrombosis and Haemostasis |

| | |
|---|---|
| LAR | Legally Acceptable Representative |
| LMWH | Low-Molecular Weight Heparin |
| PE | Pulmonary Embolus |
| REB | Regulatory Ethics Board |
| REMAP- CAP | A Randomized, Embedded, Multi-factorial, Adaptive Platform Trial for Community-Acquired Pneumonia |
| SAE | Serious Adverse Event |
| SARS-CoV-2 | Severe Acute Respiratory Syndrome Coronavirus 2 |
| sBP | systolic Blood Pressure |
| $SpO_2$ | Oxygen Saturation |
| SSC | Scientific and Standardization Committee |
| UFH | Unfractionated Heparin |
| VTE | Venous Thromboemboembolism |
| WHO | World Health Organization |

### 3 BACKGROUND

At the end of 2019, an outbreak of severe respiratory infection had surged in Wuhan, China and, since then, it has rapidly spread across the globe. A novel coronavirus was identified as the cause of this outbreak (Zhu *N Engl J Med* 2020). The World Health Organization declared this new infection a global pandemic on March 11, 2020. This disease has since been known as COVID-19 and the virus that causes COVID-19 was named severe acute respiratory syndrome coronavirus 2 (SARS-CoV-2) (Zhu *N Engl J Med* 2020; Guan *N Engl J Med* 2020). As of September 2020, more than 30 million people have been diagnosed with COVID-19 and approximately 1 million have died. Currently in Canada, >150,000 patients have been infected and approximately 10,000 have died thus far (Government of Canada 2020).

The clinical spectrum of COVID-19 is extremely variable and not yet completely understood. The incubation period is thought to be within 14 days after exposure, most commonly in the first 5 days (Guan *N Engl J Med* 2020; Wu *JAMA* 2020). Most infections are mild, and a large proportion of infected people likely develop no or very mild symptoms. Approximately 15% of symptomatic patients progress to severe pneumonia, with the need for hospitalization, and 5% develop respiratory failure, shock and multi-organ dysfunction (Wu *JAMA* 2020; Wu *JAMA Inter Med* 2020; Yang *Lancet Respir Med* 2020). The case fatality rate is extremely variable, most likely a function of differences in population demographics and density, diagnostic screening criteria, and death reports among countries (Spychalski *Lancet Infect Dis* 2020). In Wuhan, the case-fatality rate was approximately 5.8%. In Italy, the estimated case-fatality rate in March was 7.2%, while in South Korea it is currently 1.73% (WHO-China 2020; Onder *JAMA* 2020; Korea Centers for Disease Control and Prevention 2020).

There are limited proven treatments for COVID-19, including dexamethasone (Horby NEJM 2020) and possibly hydrocortisone in critically ill patients (Angus *JAMA* 2020) as well as remdesivir (Wiersinga *JAMA* 2020). The current management is supportive (Alhazzani *Crit Care Med* 2020). A number of other therapies are currently under investigation and have been used anecdotally in clinical practice, particularly in those with severe forms of COVID-19. Examples are antiviral drugs, antimalarials (e.g., chloroquine and hydroxychloroquine), alone or in combination with azithromycin, and IL- 6 inhibitors (e.g., tocilizumab) (Alhazzani *Crit Care Med* 2020) – these therapies have been or are now being studied in trials. To date, however, there remains a strong unmet clinical need for effective therapeutic approaches.

### 4 STUDY RATIONALE

#### 4.1 ATTACC Trial Rationale Overview

Several lines of evidence support the potential efficacy of therapeutic parenteral anticoagulation with heparin for the treatment of COVID-19. First, COVID-19 can be associated with a hypercoagulable state, and many patients may experience significant cardiac and pulmonary macro-and micro-vascular thrombotic complications contributing to clinical deterioration. COVID-19 is associated with an unusually high incidence of venous thromboembolic events (Klok et al. *Thrombosis Research* 2020; Cui *J Thromb Haemost* 2020). Second, heparin directly induces conformation changes in the SARS-CoV-2 receptor spike protein, and may limit cellular invasion into the pulmonary epithelium, myocardium, and vascular endothelium (MyCroft-West bioRxiv pre-print). Third, heparin has direct anti-inflammatory effects that may reduce the severity of organ injury and hemodynamic collapse. Given its ubiquitous availability, heparin may be rapidly translatable to clinical care globally if found to be effective (Thachil *J Thromb Haemost* 2020), at a time when immediately implementable solutions are urgently needed. Nonetheless bleeding concerns are present. D-dimer identifies higher risk COVID-19 patients and may be both a prognostic and predictive enrichment marker, possibly stratifying the benefit of heparin. For example, a recent observational study from Wuhan observed that when D-dimer exceeded 3.0 ug/mL, prophylactic-dose heparin use was associated with an approximately 20% absolute risk reduction in 28- day mortality (32.8% vs 52.4%, p=0.017) (Tang *J Thromb Haemo* 2020), although mortality remained high. More intensive anticoagulation strategies may provide even further event reduction. The present study therefore aims to evaluate the efficacy of therapeutic-dose parenteral heparin versus usual care in hospitalized COVID-19. Given that D-dimer may stratify the benefit of heparin, response adaptive randomization is implemented based on D-dimer cut points, enabling the trial to determine where therapeutic benefit exists across a range of D-dimer levels. The adaptive design is also appropriate for the pandemic, as information to guide sample size estimations is limited.

#### 4.2 Risk Stratification in Patients with COVID-19

Advanced age and underlying comorbidities have been associated with increased likelihood of severe illness (Zhou *Lancet* 2020). Patients with cardiovascular disease or known cardiovascular disease risk factors, such as diabetes and hypertension, are at a particularly high risk of an unfavorable disease course (Wu *JAMA* 2020). Biomarkers are also associated with worse prognosis, including D-dimer and troponin (Zhou *Lancet* 2020). In a retrospective cohort from Wuhan, Shi and collaborators reported that, among 416 consecutive hospitalized patients with COVID-19, 20% had elevated troponin at hospital admission (Shi *JAMA Cardiol* 2020; Figure 1). Compared to patients with normal troponin, these patients had a markedly increased risk of complications, including acute respiratory distress syndrome (ARDS), acute kidney injury and in-hospital mortality (adjusted HR for mortality: 3.41; 95% CI: 1.62 to 7.16; p=0.001), with 50% of these troponin-positive patients dying (Shi *JAMA Cardiol* 2020). In another observational study, Zhou and collaborators included 191 COVID-19 patients from two hospitals in Wuhan. In unpublished analyses, we have observed that most patients with elevated troponin also have elevated D-dimer, which may mark an upstream coagulopathy producing end-organ injury. Indeed, patients with **D-dimer** greater than 1mcg/mL at admission were at increased risk of in-hospital death (adjusted OR: 18.42; 95% CI: 2.64 to 128.55; p=0.0033). When comparing 113 patients with COVID-19 who died versus 161 who survived, Chen and colleagues found that troponin I and D-dimer levels were markedly higher in deceased patients (median troponin I levels: 40.8 pg/mL (IQR: 14.7-157.8) vs. 3.3 (IQR: 1.9-7.0); median d-dimer levels: 4.6 mcg/mL (IQR: 1.3-21.0) vs. 0.6 (IQR: 0.3-1.3) (Chen *BMJ* 2020). These results are consistent with emerging U.S. reports (Petrilli *bioRxiv* 2020). These biomarkers may be a valuable aid to risk stratification and guidance on resource allocation among hospitalized patients and have been routinely recommended in some institutional protocols (e.g., www.Covidprotocols.org). These observations highlight the potential use of biomarkers to guide treatment decision-making both as **prognostic** and **predictive risk markers**. However, more work is needed to understand the optimal biomarker cut-offs that align treatment with benefit. Furthermore, biomarkers that reflect downstream processes may be later markers, and treatment benefit may be realized with earlier intervention. An adaptive clinical trial with pre-defined biomarker subgroups based on D-dimer level, could address these questions while evaluating therapeutic efficacy.

#### 4.3 Hypercoagulability in Patients with COVID-19

Severe illness from COVID-19 is associated with important derangements in coagulation resulting in a hypercoagulable state. These derangements are strongly associated with poor clinical outcomes and various lines of evidence suggest that the prothrombotic state may be causally related to poor outcomes. Elevated D-dimers may be a biomarker of this pathway (Zhou *Lancet* 2020). In a series of 183 patients, those who died 11% exhibited markedly elevated D-dimers and fibrin degradation products; 15 of the patients who died met the criteria for disseminated intravascular coagulation (DIC), whereas only 1 survivor developed DIC (Tang *J Thromb Haemost* 2020). Similar derangements in hemostasis were documented in a separate case series of 94 patients (Lippi *Clin Chem Lab Med* 2020). Markers of DIC correlate with clinical deterioration including ischemic injury in the fingers and toes (Li *Emerg Microbes Infect* 2020).

Macrovascular embolic events seem frequent. In a case series, pulmonary embolism was identified in 10 of the 25 patients who underwent CT pulmonary angiography (pre-print data, SSRN id: 3548771). The limited autopsy data suggest a constellation of pathological findings including thrombus in pulmonary microvessels (Fox *bioRxiv* 2020; Yao *Chin J Pathol* 2020).

The specific drivers of coagulopathy and DIC in COVID-19 disease are uncertain. SARS-CoV-2 can bind ACE2 and infect and injure endothelium, leading to tissue factor expression, endothelial activation and activation of the coagulation cascade (endotheliopathy). Strikingly, respiratory mechanics in patients requiring mechanical ventilation markedly differs from conventional mechanics seen in ARDS, and in COVID-19 is characterized by high dead space and impaired oxygenation in the absence of significant increase in pulmonary elastance, suggestive of pulmonary vascular occlusion. Similarly, reports of rapid deterioration in left ventricular systolic function also support both coronary and cardiac micro-thrombotic occlusive processed. Coronary events are well known to be increased early in patients with other forms of viral pneumonia, and this may explain the 3 to 4 fold increased in mortality hazard in patients with elevated troponin.

#### 4.4 Managing Hypercoagulability in Patients with COVID-19

Many institutional guidelines, as well as the International Society of Thrombosis and Hemostasis (ISTH), recommend routine venous thromboprophylaxis in patients hospitalized with COVID-19 (e.g., www.Covidprotocols.org; Thachil *J Thromb Haemost* 2020). However, based on the above there is a compelling rationale to administer therapeutic heparin earlier in the disease course, which may also have pleiotropic benefit. In a recently published case-control study from Wuhan, 99 out of 449 consecutive patients received heparin (primarily thromboprophylactic dose heparin) for 7 days or longer (Tang *J Thromb Haemost* 2020). Although no difference in 28-day mortality was observed between patients receiving (or not) heparin (adjusted OR: 1.65; 95% CI: 0.93 to 2.92; p=0.088), among patients with elevated D-dimer, those treated with heparin, compared to those not treated with heparin, experienced lower 28-day mortality (OR for D-dimer> 3mcg/mL: 0.44; 95% CI: 0.27 to 0.87; p=0.017) (Tang *J Thromb Haemost* 2020). Based on clinical experience, an increasing understanding of the pathobiology of COVID-19, and emerging observational evidence, administering therapeutic anticoagulation in patients with COVID-19 has become the practice in some centers who have cared for a large number of these patients, but equipoise remains and currently this is not part of standard of care.

#### 4.5 Other Potential Therapeutic Benefits of Heparin in COVID-19: Anti-viral and Anti-inflammatory Effects

In addition to its antithrombotic benefit, heparin has two additional potential beneficial effects in COVID-19: anti-viral effects that may limit viral invasion and anti-inflammatory effects. Very recently, the SARS-CoV-2 spike protein has been shown to interact with heparin. Upon binding heparin, the spike protein undergoes significant conformational changes that may prevent it from binding ACE2 (Mycroft-West *bioRxiv* 2020). We have discussed these data with the investigators in the U.K., and these anti-viral effects are present for both unfractionated heparin and some low molecular weight heparins.

Heparin had previously also been shown to prevent cellular invasion by SARS-CoV-1 (Vicenzi *Emerg Infect Dis* 2004; De Haan *J Virology 2005*), and is known to inhibit attachment and entry of other enveloped viruses such as HIV and HSV (Moulard *J Virol* 2000). Thus, heparin may exert a direct antiviral effect to prevent viral invasion of pulmonary epithelium, myocardium, and vascular endothelium.

Unfractionated heparin (UFH) also has potentially beneficial anti-inflammatory effects. UFH is a known inhibitor of complement and adhesion molecule expression in the microvasculature (Lever *Br J Pharmacol* 2000). UFH administration prevents acute lung injury and increases survival in various models of septic shock (Gans *Surgery* 1975). In a propensity-matched retrospective cohort study of patients with septic shock, heparin was associated with reduced 28-day mortality (Zarychanski *Crit Care Med* 2008). In a systematic review and meta-analysis of 6 randomized trials of heparin enrolling 2,477 patients with sepsis, the pooled odds ratio for mortality was 0.88 (95% CI: 0.77 to 1.00; I2 = 0%) (Zarychanski *Crit Care Med* 2015).

#### 4.6 Safety of Heparin

The most frequent complication of anticoagulation use is bleeding. However, anticoagulation with parenteral anticoagulation (low molecular weight heparin or unfractionated) has been studied extensively across diverse patient populations and favourable safety data is available. Therapeutic parenteral anticoagulation is commonly used in hospitalized patients for the prevention and treatment of venous thromboembolic disease, acute coronary syndromes, and stroke prevention in patients with atrial fibrillation (Tiryaki AmJ Heal Pharm 2011). Its dosing and management of heparin is thus very familiar to clinicians. Overall, patients receiving therapeutic heparin have a 1-5% risk of major bleeding, depending on underlying risk and duration of exposure (Mismetti Chest 2005; Petersen JAMA 2004; Crowther Blood 2008).

When explored as an intervention for reducing mortality in patients with septic shock requiring intensive care unit admission, unfractionated compared to placebo or usual care demonstrated no significant differences in bleeding (gastrointestinal, central nervous system, epistaxis, hematuria) or need for blood product transfusion (Jaimes Crit Care Med 2009; Zarychanski Crit Care Med 2008). Subsequent meta-analysis of studies comparing UFH or LMWH with placebo or usual care in patients with sepsis admitted to the ICU suggests no increase the risk of major hemorrhage (RR 0.79, 95% CI 0.53–1.17), although its use is associated with a modest increase in minor hemorrhage (RR 1.49, 95% CI 1.07–2.07) (Zarychanski Crit Care Med 2015). In critically-ill patients, increased doses of UFH are not associated with increased clinically significant bleeding (0.2% with higher UFH dosing compared to 0.3% with standard unfractionated dosing, p=0.059) (Reynolds J Hum Pharmacol Drug Ther 2019). When compared to unfractionated, low molecular weight heparin — the preferred anticoagulant of choice in this study in the absence of any clinical contraindications — may exhibit a reduced risk of bleeding (OR 0.43; 95% CI 0.22 to 0.83; p=0.01) (Alikhan

Cochrane Database Syst Rev 2014). In a randomized-controlled trial, no difference in bleeding has been observed with enoxaparin 1.5 mg/kg subcutaneous once daily versus 1 mg/kg twice daily (Merli Annals Internal Med 2001). In this trial of 900 patients with venous thromboembolic disease, the incidence of major haemorrhage did not differ among those receiving unfractionated heparin (2.1%) once-daily enoxaparin (1.7%) or twice-daily enoxaparin group (1.3%).

Patients with an underlying systemic hypercoagulable state, in whom heparin is being given to offset this, may intuitively have a lower risk of bleeding. For example, in cancer-associated venous thromboembolisms (VTEs) – an underlying hypercoagulable state – with an estimated rate of major bleeding of 3.2% over 6 months' follow-up (Lee JAMA 2015; Li Thromb Res 2019).

Major bleeding will be a secondary safety endpoint and will be monitored by the DSMB in frequent interim analyses. Premature discontinuation of therapy related to bleeding, need for procedures requiring >48 hours of interruption, or physician discretion will also be monitored. A minimum hemoglobin cut-off of 80 g/L will be applied to reduce the risk of requiring blood transfusion in patients who experience bleeding.

Heparin-induced thrombocytopenia is another adverse event which can accompany heparin therapy, occurring significantly less often in patients receiving low molecular weight heparin compared with UFH (RR 0.22, 95% CI 0.06 to 0.84) (Junqueira Cochrane Database Syst Rev 2017). The overall incidence of HIT is 0.2–0.5%, and is higher in patients receiving therapeutic doses of UFH (0.79%) compared to those receiving prophylactic doses (<0.1%). (Creekmore J Hum Pharmacol Drug Ther 2006; Smythe Chest 2007).

Injection site adverse effects may include pain, mild local irritation, hard inflammatory nodules and injection site hematomas may follow the subcutaneous injection of a low molecular weight heparin.

Other rare adverse reactions reported with the use of unfractionated heparin and LMWH are hypersensitivity and allergic reactions, hepatic enzymes increase, hypercalcemia, urticaria, pruritus, erythema.

#### 4.7 Background Summary

Taken together, these observations provide a compelling rationale for the use of therapeutic dose heparin in patients with COVID-19. These beneficial effects reflect antithrombotic properties, direct SARS-CoV2 antiviral properties, and anti-inflammatory properties (**Figure 1**). Risks of bleeding from prior studies are small. Risk/benefit may be best aligned using D-dimers. The clinical trial proposed herein looks to prevent clinical deterioration and improve survival in patients hospitalized with COVID-19 with therapeutic-dose heparin treatment, using an adaptive design that allows various D-dimer cut-offs to be studied objectively.

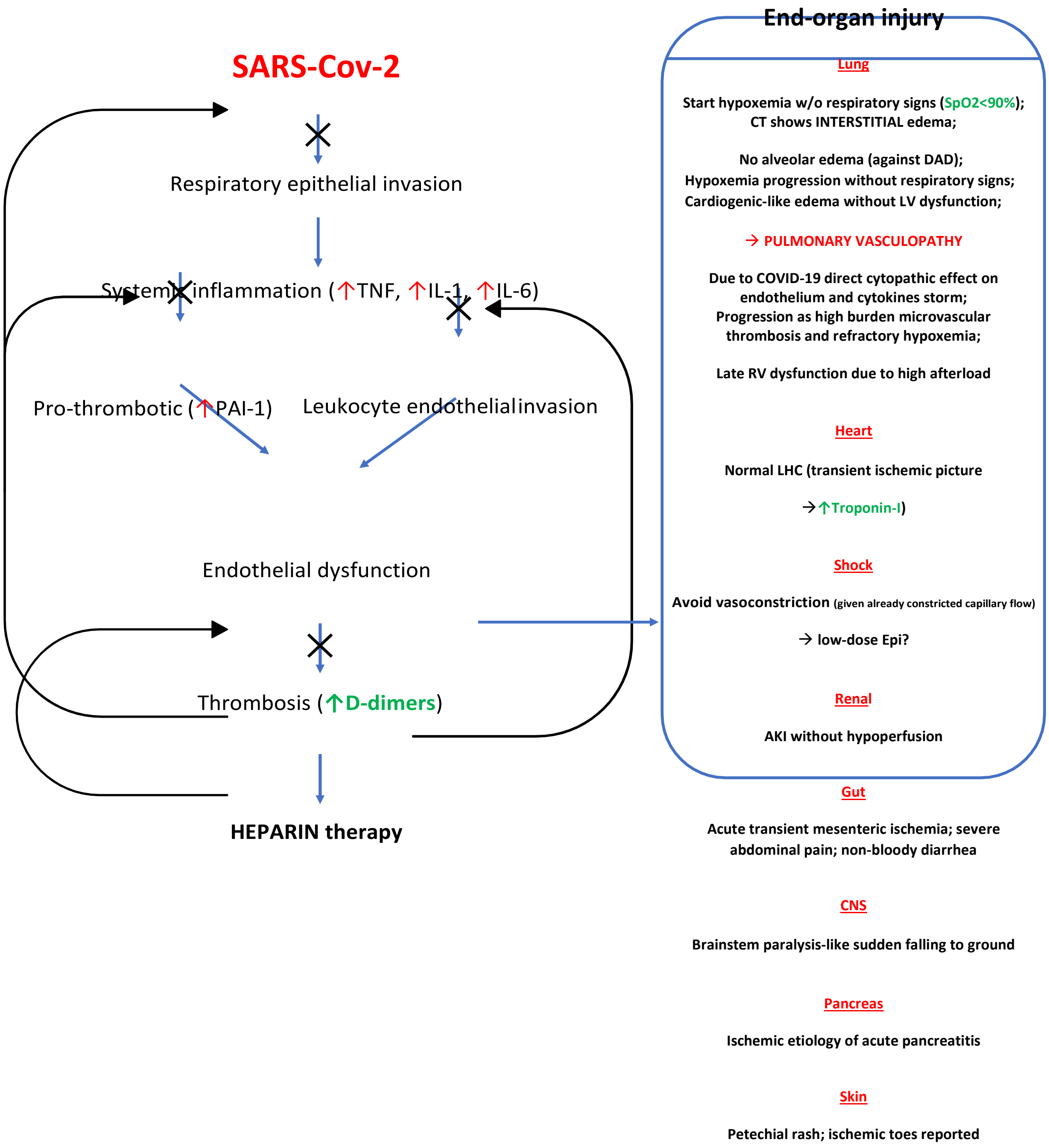

**Figure 1:** Proposed pathophysiologic pathways in COVID-19 upon which heparin may act.

### 5 THE TRIAL

#### 5.1 Study Design

A phase III prospective, open-label, adaptive multi-platform randomized controlled trial.

#### 5.2 Patient Population

Participants with laboratory confirmed COVID-19 requiring hospitalization anticipated to last ≥72 hours will be enrolled into this study.

#### 5.3 Primary Endpoint

The primary endpoint in the trial is days alive and free of organ support at day 21. This endpoint is defined as the number of days that a patient is alive and free of organ support through the first 21 days after trial entry. Organ support is defined as receipt of invasive or non-invasive mechanical ventilation, high flow nasal oxygen (>30 L/min), vasopressor therapy, or ECMO support. Death at any time (including beyond 21 days) during the index hospital stay is assigned the worst possible score of –1.

#### 5.4 Secondary Endpoints

Secondary safety endpoints: (determined to occur after enrollment)

- Laboratory-confirmed heparin induced thrombocytopenia **(HIT)**
- Major bleeding, defined according to the International Society on Thrombosis and Haemostasis (ISTH)/Scientific and Standardization Committee (SSC) definitions and bleeding assessment tool in non-surgical patients (Schulman *J Thromb Haemost* 2005): fatal bleeding; and/or symptomatic bleeding in a critical area or organ, such as intracranial, intraspinal, intraocular, retroperitoneal, intraarticular or pericardial, or intramuscular with compartment syndrome; and/or bleeding causing a fall in hemoglobin level of ≥20 g/L, or leading to transfusion of 2 or more units of whole blood or red cells

Secondary efficacy endpoints:

- A composite endpoint of death, deep vein thrombosis, pulmonary embolism, systemic arterial thromboembolism, myocardial infarction, or ischemic stroke collected during hospitalization or at 28 days and 90 days after enrollment (whichever is earlier)
- All cause mortality assessed at 28 and 90 days following randomization
- All cause mortality during initial hospitalization (includes death after 28 days)
- Intubation assessed at 30 days following randomization
- Ordered categorical endpoint with three possible outcomes based on the worst status of each patient through day 30 following randomization: no invasive mechanical ventilation, invasive mechanical ventilation, or death

- Ventilator-free days (days alive not on a ventilator) assessed at 28 days following randomization
- Hospital-free days (days alive outside hospital) assessed at 28 days following randomization
- Vasopressor-free days (days alive not on a vasopressor) assessed at 28 days following randomization
- Renal replacement free days (days alive not on renal replacement) assessed at 28 days following randomization
- Hospital re-admission within 28 days
- Symptomatic proximal venous thromboembolism (DVT or PE) assessed at 28 and 90 days following randomization
- Myocardial infarction assessed at 28 days and 90 days following randomization
- Ischaemic stroke assessed at 28 and 90 days following randomization
- Acute kidney injury as defined by KDIGO criteria
- Systemic arterial thrombosis or embolism assessed at 28 and 90 days following randomization
- Use of extracorporeal membrane oxygenation (ECMO) support
- Mechanical circuit (dialysis or ECMO) thrombosis
- Organ support-free days at 28 days following randomization
- WHO ordinal scale (peak scale over 28 days, scale at 14 days, and proportion with improvement by at least 2 categories compared to enrollment, at 28 days)

### 6 PATIENT ELIGIBILITY

This trial will be conducted in compliance with the protocol and GCP. Any questions about eligibility criteria must be addressed prior to patient registration.

#### 6.1 Inclusion Criteria

1. Patients ≥18 years of age providing (possibly through a substitute decision maker) informed consent who require hospitalization anticipated to last ≥72 hours for microbiologically-confirmed COVID-19, enrolled < 72 hours of hospital admission **or** of COVID-19 confirmation

#### 6.2 Exclusion Criteria

1. Requirement for chronic mechanical ventilation via tracheostomy prior to hospitalization
2. Patients for whom the intent is to not use pharmacologic thromboprophylaxis
3. Active bleeding
4. Risk factors for bleeding, including:
    a. intracranial surgery or stroke within 3 months;
    b. history of intracerebral arteriovenous malformation;
    c. cerebral aneurysm or mass lesions of the central nervous system;
    d. intracranial malignancy
    e. history of intracranial bleeding
    f. history of bleeding diatheses (e.g., hemophilia)
    g. history of gastrointestinal bleeding within previous 3 months
    h. thrombolysis within the previous 7 days
    i. presence of an epidural or spinal catheter
    j. recent major surgery <14 days
    k. uncontrolled hypertension (sBP >200 mmHg, dBP >120 mmHg)
    l. other physician-perceived contraindications to anticoagulation
5. Platelet count <50 x$10^9$/L, INR >2.0, or baseline aPTT >50
6. Hemoglobin <80 g/L (to minimize the likelihood of requiring red blood cell transfusion if potential bleeding were to occur)
7. Acute or subacute bacterial endocarditis
8. History of heparin induced thrombocytopenia (HIT) or other heparin allergy including hypersensitivity
9. Current use of dual antiplatelet therapy
10. Patients with an independent indication for therapeutic anticoagulation
11. Patients in whom imminent demise is anticipated and there is no commitment to active ongoing intervention
12. Anticipated transfer to another hospital that is not a study site within 72 hours

13. Enrollment in other trials related to anticoagulation or antiplatelet therapy

#### 6.3 Patient Consent

Sample English consent forms for the trial will be provided. A copy of the initial full board REB/IRB approval and approved consent form must be sent to Ozmosis Research. The subject/LAR must sign consent prior to registration or may provide consent as per FDA guidance document Coronavirus (COVID-19) Update: FDA Issues Guidance for Conducting Clinical Trials (Appendix A) or REB/IRB recommendations.

Patients admitted to hospital that have microbiologically-confirmed SARS-CoV-2 will be assessed for eligibility. Screening will include a review of inclusion and exclusion criteria. Study staff will discuss with the treating clinical team a potential subject's suitability to be approached for the trial. Information about the study will be presented to potential subjects (or legally authorized representative, which can include a substitute decision-maker in cases of incapacity). Given the potential for viral transmission and the nature of the studied intervention, consent will be obtained as per the REB/IRB recommendations or FDA Guidance (Appendix A). This is anticipated to include verbal consent and consent by telephone.

#### 6.4 Patient Enrollment

Patient enrollment and randomization will occur through the eCRF system (eSOCDAT).

### 7 STUDY PLAN

#### 7.1 Anticoagulation Administration

Participants randomized to the <u>therapeutic arm</u> will be given therapeutic-dose parenteral anticoagulation daily, up to 14 days or until *recovery*, defined as hospital discharge or liberation from supplemental oxygen >24 hours (provided supplemental oxygen was originally required), whichever comes first. If patient was on oxygen pre-hospitalization, recovery is defined as return to their baseline oxygen requirement, or hospital discharge (whichever comes first).

Participants randomized to the <u>control arm</u> will receive usual care, which is expected to include thromboprophylactic dose anticoagulation according to local practice. To ensure adequate separation between the study groups the dose of heparin/LMWH used in the usual care arm should not equal more than half of the approved therapeutic dose for that agent for the treatment of venous thromboembolism.

*See Section 9 (Medicinal Product) for further details.*

#### 7.2 Study Schedule

Subjects will be followed until hospital discharge, after which time telephone contact will be undertaken to ascertain vital status following hospital discharged. (Schedule days refer to post-randomization days.) All post-discharge follow-up is telephone-/remote.

| Investigations | Pre-Treatment (Baseline)[6] | Day 1 | Day 3 | Day 5 | Day 7 | Day 10 | Day 14 | Day 21 | Day 30 | Day 90 |
|---|---|---|---|---|---|---|---|---|---|---|
| Windows | -72 hours | - 3 days | | | | | | | + 7 days | |
| Consent & Randomization | X | | | | | | | | | |
| Demographics | X | | | | | | | | | |
| Medical History | X | | | | | | | | | |
| Weight | X | | | | | | | | | |
| SOC Vitals documented (SpO2 and FiO2, heart rate, blood pressure, respiratory rate, temperature)[1] | X | X | X | | X | | X | | | |
| Hematology bloodwork (SOC)[1] | X | X | X | X | X | X | X | | | |

| | | | | | | | |
|---|---|---|---|---|---|---|---|
| Biochemistry bloodwork (SOC)[1] | X | X | X | X | X | X | X |

| | | | | | | | | | | |
|---|---|---|---|---|---|---|---|---|---|---|
| Troponin (SOC)[1] | X | X | X | X | X | X | X | | | |
| D-dimer[2] | X | | | | | | | | | |
| D-dimer (SOC)[1] | | X | X | | X | | X | | | |
| Optional Biorepository (blood samples) | X | X | X | | X | | X | | | |
| Anticoagulant Administration | | X | | | | | | | | |
| Organ-free support outcome | | | | | | | | X | | |
| Primary and secondary outcomes[3,] | | X | | | | | | | | |
| Survival, DVT, PE, MI (by phone)[3] | | | | | | | | | X | |
| WHO ordinal assessment | | | | | | | | | | X |
| Adverse events[5] | | X | | | | | | | | |
| Concomitant medications[5] | | X | | | | | | | | |

**Footnotes:**

[1]As per routine standard of care, collected while on therapy (until discharge or up to day 14 or recovery). Record the values closest to the day of the assessment. **Values from - 3 days may be used provided the values are temporally after the previous assessment.** If more than 1 value exists, the value closest to the day of study assessment will be used. Exceptions to this rule; troponin (collect highest troponin since last assessment), hemoglobin (collect lowest hemoglobin since last study assessment) and creatinine (collect highest creatinine since last study visit).

[2]**D-dimer is to be collected at baseline for all patients**. This is part of the current standard of care at most institutions, but where it is not, it will be collected as part of the trial protocol at baseline. If possible, it should be reported (i.e., a result available) prior to randomization, so that participants may benefit from response-adaptive randomization; however, patients are still able to be randomized if the D-dimer result is not available prior to randomization, in which case randomization will proceed 1:1.
Nonetheless, a level is required to be drawn at baseline in all cases if one is not already available within 72 hours of randomization. If there is a site that is not able to collect D-dimer for all patients at baseline, this will be discussed with the sponsor on a case-by-case basis.

[3]All post-discharge follow-up is telephone-/remote.

[4]Refer to 'THE TRIAL' section for a list of study outcomes

[5]Treatment-related adverse events are assessed only while on therapy. Concomitant medications are assessed from the time of consent and for the duration of therapy.

[6]The laboratory values closest to randomization should be recorded. Values up to 72 hours prior to randomization can be used for baseline values if the test is not available at the time of randomization and can not be repeated.

### 8 CONCOMITANT MEDICATIONS

Concomitant medications representing experimental COVID-19 treatments (as long as not related to anticoagulation) and anti-platelet agents, will be collected for the study from time of consent to 14 days or time of pre-defined treatment discontinuation (whichever comes first).

Potential Drug Interactions: refer to the next section and to the current product monographs for up to date interactions.

#### 8.1 Drug Interactions

Heparin should be used with caution in patients receiving non-steroidal anti- inflammatory drugs, thrombolytic agents, glycoprotein IIb/IIIa antagonists, acetylsalicylic acid, platelet inhibitors, vitamin K antagonists, activated protein C, direct factor Xa and IIa inhibitors because of increased risk of bleeding.

### 9 MEDICINAL PRODUCT

#### 9.1 Characterization of Investigation Medicinal Product

There is a preference for subcutaneous low molecular weight heparin, given ease of administration and resultant reductions in the need for clinical staff-patient interactions, if no contraindication is present. Enoxaparin is the preferred low molecular weight heparin given emerging data supporting potential viral inhibitory properties (Mycroft- West bioRvix preprint and Mark Skidmore, Keele University, personal communication), although tinzaparin or dalteparin are also acceptable, if available. Alternatively, intravenous unfractionated heparin infusion may be also used. The therapy may be switched within a subject during the course of the trial at the discretion of the treating physician. Anticoagulants used in the trial, whether as part of the intervention arm or as part of usual care/control arm, will be sourced, stored and dispensed by participating hospitals according to current practice and local policy.

This is a pragmatic trial of therapeutic anticoagulation, and hence the treating physician should determine what is the most appropriate parenteral anticoagulant for the patients to receive.
For pregnant women, use of non-tinzaparin (Innohep) product is preferred. If tinzaparin is the only product available, then only pre-filled syringes (without benzyl alcohol) will be administered as per the product monograph.

Low molecular weight heparin (LMWH)

LMWH is commenced, administered, and monitored according to local hospital policy, practice and guidelines that pertain to treatment of venous thromboembolism (i.e. not thromboprophylactic doses). The dose selected should be based on measured or estimated weight of the patient.

Adjustment for impairment of renal function should be according to local practice and policy.

Generally accepted dosing regimens for enoxaparin include: 1.5 mg/kg subcutaneous once daily or 1 mg/kg subcutaneous twice daily, assuming no dose adjustment is required. Alternatively, other subcutaneous low molecular weight heparins may be used, including tinzaparin, if available, (generally given at a dose of 175 anti-Xa IU/kg subcutaneous once daily if no dose adjustment is required) or dalteparin (200 IU/kg subcutaneous once daily or 100 IU/kg subcutaneous twice a day if no dose adjustment is required), as available.

Unfractionated heparin (UFH)

If UFH is used, this is commenced, administered, and monitored according to local hospital policy, and guidelines that are used for the treatment of venous thromboembolism (i.e. not for acute coronary syndrome). An intravenous infusion ofunfractionated heparin, is typically dosed according to total body weight and pragmatically adjusted according to local institutional policy to achieve an activated partial thromboplastin time (aPTT) of 1.5-2.5x the reference value. If UFH is used, the availability of a local hospital policy that specifies an aPTT target in this range or an anti-Xa value is a requirement.

#### 9.2 Accountability and Destruction

This standard-of-care study drug for thromboprophylaxis will be provided from standard hospital supply and dosed according to local policy and practice. Sites will be responsible for assuring adequate local supply in coordination with local pharmacy.

#### 9.3 Dose Adjustments

Dosing and dose adjustment of anticoagulants used for therapeutic anticoagulation should conform to local practices and policies. Examples are provided below.

*Renal Impairment:* In patients with acute or chronic severe renal impairment (creatinine clearance <30 mL/min), dose-adjustments or changes in therapy in patients receiving therapeutic dose LMWH) is typically required. Dose adjustments and continued use of LMWH in this setting (vs. switching to UFH) is at the discretion of the treating medical team.

*Liver Impairment:* Low molecular weight heparin should be used with caution in patients with hepatic insufficiency.

#### 9.4 Subject Compliance

Daily drug administration including name of drug, dose, and route of administration will be recorded in the source documents and captured into an eCRF.

#### 9.5 Premature Withdrawal/ Discontinuation Criteria

Treating physicians may choose to discontinue therapy at their discretion. A premature discontinuation of treatment will be defined as an interruption in study drug for >26 hours. Temporary, shorter interruptions, for example to safely facilitate invasive procedures, are not considered interruptions or discontinuations in therapy provided the interruption does not exceed 48 hours. Reasons for treatment discontinuation may include but are not limited to:

- Heparin induced thrombocytopenia or other heparin allergy/hypersensitivity
- Thrombocytopenia if platelet count <50x$10^9$/L
- Major Bleeding, defined based closely on the ISTH/SSC definitions and bleeding assessment tool in non-surgical patients (below)

- Coagulopathy associated with an elevated INR (e.g. >2.0) or hypofibrinogemia (fibrinogen < 1 g/L)
- Following invasive procedures where heparin is deemed unsafe to re-institute
- Patients requiring systemic anticoagulation or fibrinolytic therapy
- Treating physician discretion

Temporary interruptions in therapy for ≤26hours (e.g., to facilitate invasive procedures) will not be considered a premature treatment discontinuation or a protocol violation.

Change in choice of anticoagulant or dose will not be considered discontinuation of therapy or a protocol violation. Time on unfractionated heparin will be determined as the time during which the infusion was administered, not determined by aPTT level.

Occurrence of HIT must result in cessation UFH or LMWH without recommencement regardless of treatment assignment. Use of an acceptable alternative agent is required in this instance as clinically indicated. Occurrence of HIT is an AE.

### 10 OPTIONAL BIOREPOSITORY

Blood sample collection will occur at the time points indicated in the Study Plan for patients that have provided informed consent on the optional consent for blood sample collection. It is not mandatory for all institutions to participate in the collection of correlative samples and each institutions willingness and ability to participate will be discussed on a case-by-case basis with the sponsor.

Refer to the Laboratory Manual for details on correlative sample collection.

### 11 SAFETY AND REPORTING REQUIREMENTS

#### 11.1 Adverse Event (ONLY study drug related AEs will be reported)

As an open label trial of UFH or LMWH, adverse events captured in this trial are events that are **plausibly related** to the investigational agent. Adverse events **plausibly related** to heparin or low molecular weight heparin include major bleeding and HIT.

#### 11.2 Adverse Event Documentation

**Treatment-related** AEs must be recorded in the eCRFs. Documentation must be supported by an entry in the subject's file.

#### 11.3 Serious Adverse Event (SAE) due to the study drug

SAEs must be serious events that are believed to be **plausibly related** to the study drug and will include:

- Major bleeding (outlined the definition here of the ISTH)
- Laboratory confirmed heparin-induced thrombocytopenia

As per ICH guidelines, a **Serious Adverse Event** is any AE occurring at any dose that

#### (we will only report study drug related SAEs):

- Results in death
- Is life-threatening
- Requires prolongation of existing hospitalization
- Results in persistent or significant disability/incapacity
- Is a congenital anomaly / birth defect

**Life-threatening**: The term "life-threatening" in the definition of "serious" refers to an adverse event in which the subject was at risk of death at the time of the event. It does not refer to an adverse event that hypothetically might have caused death if it were more severe.

**Disability** means a substantial disruption of a person's ability to conduct normal life's functions.

**Important medical event**: Any adverse event may be considered serious because it may jeopardize the subject and may require intervention to prevent another serious condition.

Any death (regardless of cause) that occurs from the time of administration of the first dose of study therapy until 28 days after the final administration of the study drug, and any death occurring after this time that is judged at least possibly related to prior treatment with the study drug, will be promptly reported.

#### 11.4 Reporting Serious Adverse Events (Only Study Drug Related SAEs will be reported)

All serious adverse events (SAE) defined as per ICH guidelines (see above) and other adverse events that are **plausibly related** to the study drug (see section 11: HIT and major bleeding) must be recorded on the eCRF. Any collected event that is deemed serious must be reported through eSOCDAT within 24 hours of the site becoming aware of the event.

#### 11.5 Procedure for Expedited Reporting

Responsibility for Reporting Serious Adverse Events to Health Canada

Ozmosis Research will provide expedited reports of SAEs to Health Canada according to applicable guidelines and regulations (including the 7-day notification for death and life-threatening events), i.e. events which are BOTH serious AND unexpected, AND which are thought to be related to protocol treatment (or for which a causal relationship with protocol treatment cannot be ruled out).

Responsibility for Reporting Serious Adverse Events to Sponsor

Ozmosis Research will be responsible for submitting SAE reports (Initial and/or Follow-up reports) received from the sites, to the Sponsor within 24hrs after receipt of the SAE form at Ozmosis Research.

Reporting Serious Adverse Events to Local Research Ethics Boards

Ozmosis Research will notify all Investigators on this study of all Serious Adverse Events that are reportable to regulatory authorities in Canada. This includes all serious events that are unexpected and related to protocol treatment. Investigators must notify their Research Ethics Boards (REB/IRBs) and file the report with their Investigator Site File. Documentation that serious adverse events (SAEs) have been reported to REB/IRBs must be kept on file at Ozmosis Research.
Documentation can be any of the following:

- letter from the REB/IRB acknowledging receipt
- stamp from the REB/IRB, signed and dated by REB/IRB chair, acknowledging receipt
- letter demonstrating the SAE was sent to the board

All expedited serious adverse events occurring within a centre should also be reported to local REB/IRBs.

#### 11.6 Study Management and Governance

Clinical Coordination Centre

The Clinical Coordinating Centre (CCC) will be Ozmosis Research who will be responsible for overall study management.

Data Coordinating Centre

The Data Coordinating Centre (DCC) is SOCAR Research in Switzerland. SOCAR will receive statistical support from Berry Consultants, who will perform interim analyses based on shared limited datasets from SOCAR to Berry. Data may also be shared with external research consortia (including other trials) to facilitate pooled analyses and more rapid/timely dissemination of results.

Executive Committee and Steering Committee

The Executive Committee will consist of the Principal Investigators, as well as representatives from the CCC and SOCAR. The Executive Committee is responsible for the execution of the trial according to the study protocol. A Steering Committee will be responsible for providing clinical and methodological guidance, including overall study design, execution, analysis, and publication of the main study results. The Steering Committee will oversee the management of the clinical trial sites and will also act as the Publication Committee. While the study is ongoing, the Committee will approve any protocol amendment that may become necessary and is responsible for maintaining the scientific integrity of the study.

Collaboration with ACTIV

The ACTIV trial platform is a U.S. science initiative that has collaborated with ATTACC and REMAP. The trials are operationally distinct but are working as harmonized trials that will perform primary analysis together.

### 12 STATISTICAL ANALYSIS

#### 12.1 Study Population

Data from ATTACC, ACTIV and REMAP will be analyzed together as a single multiplatform randomized controlled trial. Each of the three platforms operate individually but in coordination, and with harmonized protocols and DSMB oversight. The adaptive design does not require specification of a sample size a priori. This design was chosen given uncertainty about effect sizes and event rates given the lack of historical data amidst the emerging pandemic.. The trial will be discontinued when pre- specified criteria for superiority or futility are met according to regular interim analyses, including in subgroups of the overall trial (defined based on biomarkers and clinical parameters). We anticipate enrolling between 350 and 2,000 patients, which gives 90% power to detect an odds ratio ≥ 1.5 for avoiding organ-support or death. Data will be analyzed primarily with intention-to-treat.

#### 12.2 Evaluation of Safety

The safety of therapeutic anticoagulation will be evaluated by means of AE reports.

#### 12.3 Trial Design Introduction

The trial design is an adaptive trial comparing therapeutic anticoagulation with UFH or LMWH vs. usual care. The effect of therapeutic anticoagulation is modeled as potentially different within prespecified patient subgroups based on the baseline D- dimer levels (moderate patients only) or severe status (receiving organ support at baseline). Each patient is classified by their baseline D-dimer levels as high (defined as ≥2-fold increase above the local site's upper limit of normal range of values), and low (below this threshold).

The effect of therapeutic anticoagulation is modeled as a potential function of the patient D-dimer subgroup. Each conclusion for therapeutic anticoagulation is by subgroup with a statistical model that borrows the effect across subgroups.

The adaptive aspects of the trial include response adaptive randomization within each of the 2 D-dimer subgroups as well as any potential conclusions (superiority, futility) for the effect of therapeutic anticoagulation within each of these 2 subgroups.

Interim analyses will be conducted monthly. The details of the trial design rules are presented in the Adaptive Design Section.

#### 12.4 Primary Endpoint

The primary endpoint is an ordinal scale that is a composite end-point that comprises mortality during the acute hospital admission and the number of whole and part study days for which the patient is alive and not requiring organ failure support while admitted to an ICU up until the end of study day 21. All patients who die before discharge from an acute hospital, irrespective of whether this occurs before or after day 21, will be coded as –1 day. All patients who never receive organ failure support will be coded as 22. Organ support is defined as mechanical ventilation, high flow nasal oxygen >30 L/min, or the use of vasopressors.

#### 12.5 Primary Analysis

Let $Y_i = \{-1, 0, 1, ..., 21, 22\}$ denote the ordinal outcome (OSFD) for patient $i$. The probability of patient $i$ observing $y$ OSFD or less is denoted as $\pi_{iy} = \Pr(Y_i \leq y)$. The model is a proportional odds model, where the log odds-ratio parameters in the model are structured so that a value > 0 implies treatment benefit, and an odds-ratio > 1 implies treatment benefit. The primary analysis model is formulated as follows:

$$\log\left(\frac{\pi_{iy}}{1-\pi_{iy}}\right) = \alpha_{y,s} - [\gamma_P + \nu_{Site,s} + \lambda_{Time,s} + \theta_{a,s:d} + \beta_{Age,s} + \beta_{Sex,s} + \beta_d]$$

1. Each "platform," *P*, has a covariate adjustment in the model, *P*=1 is REMAP- CAP, *P*=2 is ATTACC, and *P*=3 is ACTIV.
2. The "site" variable is the clinical site within the trial. These will be set as distinct sites across all three trials. The site effects are estimated separately within the severe and moderate disease states, but do not vary by d-dimer level.
3. The "time" variable is an indicator of the time epoch in which a patient was enrolled in the trial, numbered increasing from the first most recent epoch to the earliest time for the analysis. Time epochs are two-week time periods, Time=1, and then every 2-week epoch, moving back in time throughout the enrollment in the trial, Time=2,3,4,…. The time effects are modeled separately within the severe and moderate disease states, but do not vary by d-dimer level.
4. The "arm" to which a patient is randomized is labeled as *a* where *a*=1 is the control and *a*=2 is the treatment arm. The effects of arm vary by the disease state and the d-dimer level. The treatment effects for arm *a* within subtype *s*:*d* are modeled with the $\theta_{a,s:d}$ parameters.
5. The "age" variable is a categorical classification of age as ≤ 39, 40-49, 50-59, 60- 69, 70-79, and 80+. The age effects will be estimated separately within the moderate and severe disease states, but do not vary by d-dimer level.
6. The "sex" variable is sex at birth. The sex effects will be estimated separately within the moderate and severe disease states, but do not vary by d-dimer level.
7. The additive effects of d-dimer levels are modeled with the $\beta_d$ for d = 0, 1, 2, 3.
8. The $\alpha_{y,s}$ parameters determine the baseline rates of the ordinal outcome, which are modeled separately by disease state but not d-dimer level.

### 13 ADAPTIVE DESIGN

The trial design is adaptive. A sequence of frequent interim analyses will be conducted as a function of enrollment rate. The anticipation is to conduct interim analyses on the 1st day of each month.

In the multiplatform analysis that includes data from ATTACC, each rule is separately evaluated within the three subtypes: moderate state: low d-dimer, moderate state: high d-dimer, and severe state. At each interim analysis the trial could reach a trial conclusion within any of the subgroups which would stop randomization in that subgroup in favor of the control (standard of care) or therapeutic anticoagulation. If no conclusion within a subgroup is reached and randomization continues the randomization probabilities will be set based on a response adaptive randomization algorithm distinctly within the subgroup. At this time, only ATTACC is utilizing responsive adaptive randomization based on probability of treatment successes within a subgroup of d-dimer (moderate state) or within the severe state.

#### 13.1 Subgroup Conclusions

A subgroup may stop for **superiority of therapeutic anticoagulation** . This conclusion would be reached at any interim analysis in which the probability that therapeutic anticoagulation is more effective (OR > 1.5) than control in the subgroups is 99% or greater. That is a subgroup will stop for superiority of therapeutic anticoagulation if:

The trial may stop for **futility of therapeutic anticoagulation.** If the probability of at least a 20% improvement in the odds-ratio (OR > 1.2) is less than 10% then the trial will stop for futility.

The trial will continue as long as there are subgroups that have not reached a conclusion.

### 14 PUBLICATION POLICES AND DISCLOSURE OF DATA

#### 14.1 Intellectual Property

For publications, the first or senior authors will include the Principal Investigators of the study. Additional authors will be those who have made the most significant contribution to the overall success of the study. This contribution will be assessed, in part but not entirely, in terms of subjects enrolled and will be reviewed at the end of the trial by the Principal Investigator.

#### 14.2 Data Sharing

Data sharing with external parties for collaborative research and analysis will be permitted. Data will be entered in the electronic data capture SOCAR. This information is accessible to investigators.

### 15 ETHICS

#### Ethics Board Approval

Each participating centre will have on file with Ozmosis Research, a list indicating the composition of its IRB/REB consistent with Canadian regulatory guidelines. This list will be updated as appropriate.

A Health Canada REB Attestation Form must be completed and signed by the REB representative. Alternatively, an attestation may be included in the signed local ethics approval document. This documentation must be received by Ozmosis Research before the centre can be locally activated.

#### Each sub study will be reviewed by IRB/REB for determination if full board review is needed or not.

Initial approval: All study sites are required to obtain local ethics approval of the protocol and consent form by the appropriate REB/IRB prior to commencement of the clinical trial at each site.

Continuing approval: Annual (or as required by the REB/IRB) re-approval may be required for as long as subjects are being followed on protocol. It will be investigator's responsibility to apply for and obtain the re-approval.

Amendment: All protocol amendments will be confirmed in writing and submitted, as appropriate, for review by the REB/IRB and health authorities. Amendments will be reviewed and approved by applicable regulatory authorities prior to central implementation of the amendment, and by REB/IRBs prior to local implementation, EXCEPT when the amendment eliminates an immediate hazard to clinical trial subjects or when the change(s) involves only logistical or administrative aspects of the trial.

REB/IRB Refusals: If an REB/IRB refuses to approve this protocol (or an amendment/revision to this protocol), Ozmosis Research must be notified immediately of the date of refusal and the reason(s) for the refusal. Notification will then be made to Health Canada.

### 16 RECORD ACCESS AND MAINTENANCE OF STUDY RECORDS

#### 16.1 Documentation of Subject's Participation

A statement acknowledging the participation of a subject in this clinical trial must be documented in the subject's medical records along with the signed ICF.

#### 16.2 Regulatory Requirements

Health Canada approval is required for this protocol.

The following documents are required:

For participating Canadian centres:

- All Investigators must complete and sign the Health Canada Qualified Investigator Undertaking form.
- All applicable regulatory documents as listed in the Protocol Activation Checklist provided by Ozmosis Research to the sites.
- Ozmosis Research will submit via email to Health Canada a completed Health Canada Clinical Trial Site Information Form after local activation of each participating Canadian centre.

For participating U.S.A centres:

- This study is IND exempt.
- All Investigators must also submit to Ozmosis Research Inc. an up-to-date (current to within 2 years of the study start) curriculum vitae.
- Laboratory certification / accreditation and normal ranges for local lab(s).
- Consent forms, reviewed by Ozmosis Research Inc. before submission to the local IRB.
- A completed site delegation list.
- A copy of the initial full board approval letter from the local IRB. Continuing approval (full board) will be obtained at least yearly until follow-up on patients is completed and no further data is being obtained for research purpose.

#### 16.3 Subject Confidentiality and Access to Source Data/Documents

Any research information obtained about the subject in this study will be kept confidential. A subject will not be identified by name, only by his/her initials. The subject's name or any identifying information will not appear in any reports published as a result of this study.

However, information obtained from individual subject's participation in the study may be disclosed with his/her consent to the health care providers for the purpose of obtaining appropriate medical care. The subject's medical records/charts, tests will be made available to Ozmosis Research, University of Manitoba, its potential partners, Health

Canada, the REB/IRB and any other regulatory authorities. This is for the purpose of verifying information obtained for this study. Confidentiality will be maintained throughout the study within the limits of the law.

A subject's name will not be given to anyone except the researchers conducting the study, who have pledged an oath of confidentiality. All identifying information will be kept behind locked doors, under the supervision of the study Principal Investigator and will not be transferred outside of the hospital.

A subject may take away his/her permission to collect, use and share information about him/her at any time. If this situation occurs, the subject will not be able to remain in the study. No new information that identifies the subject will be gathered after that date. However, the information about the subject that has already been gathered and transferred may still be used and given to others as described above in order to preserve the scientific integrity and quality of the study.

#### 16.4 Confidentiality of the Study

Data generated as a result of this study are to be available for inspection on request by local health authority auditors, the Sponsor's Study Monitors and other personnel (as appropriate) and by the REB/IRB. The Investigator shall permit sponsor, authorized agents of the sponsor, CRO and regulatory agency employees to enter and inspect any site where the study intervention or records pertaining to the study intervention are held, and to inspect all source documents, unless there are entry restrictions into the hospital sites due to the pandemic. The protocol and other study documents contain confidential information and should not be shared or distributed without the prior written permission of sponsor.

#### 16.5 Registration of Clinical Trial

Prior to the first subject being registered/enrolled into this study, the Sponsor will be responsible for ensuring that the clinical trial is registered appropriately to remain eligible for publication in any major peer-reviewed journal, adhering to the guidelines put forth by the International Committee of Medical Journal Editors (ICMJE).

#### 16.6 Data Reporting

The data will be entered into the SOCAR database.

#### 16.7 Maintenance of Study Records

To enable evaluations and/or audits from Regulatory Authorities, Ozmosis Research or the Sponsor, the Investigator agrees to keep records, including the identity of all participating subjects (sufficient information to link records, CRFs and hospital records), all original signed informed consent forms, source documents, and detailed records of treatment disposition. The Investigator should retain these records for the duration of time required by the applicable regulatory body.

If the investigator relocates, retires, or for any reason withdraws from the study, then the Sponsor should be prospectively notified. The study records must be transferred to an acceptable designee, such as another investigator, another institution, or to the Sponsor. The investigator must obtain the Sponsor's written permission before disposing of any records.

### 17 QUALITY ASSURANCE AND QUALITY CONTROL

As per the ICH Guidelines of Good Clinical Practice, the sponsor will be responsible for implementing and maintaining quality assurance and quality control systems.

#### 17.1 Monitoring / Auditing

Ozmosis Research will organize monitoring of this study to be conducted as per Monitoring Plan, which may include delegating monitoring responsibilities to other research organizations. This may involve remote monitoring if it is not feasible to monitor on-site due to hospital restrictions during this pandemic.

As this trial is conducted under a CTA with Health Canada, your site may be subject to an inspection by Health Canada. Other audits may be conducted by the study sponsor or Ozmosis Research.

### 18 ADMINISTRATIVE PROCEDURES

#### 18.1 Amendments to this Protocols

Modification of this protocol is only possible by approved protocol amendments authorized by the Sponsor. Where required, all protocol amendments will be approved by the REB and Health Canada (for Canadian sites) and by the IRB and FDA (For U.S.A sites if FDA exemption is not granted) prior to implementation. The Investigator must not implement any deviation from, or change to the protocol, except where it is necessary to eliminate an immediate hazard to trial subject or when the change(s) involves only logistical or administrative aspects of the trial.

#### 18.2 Protocol Deviations and Violations

All violations or deviations are to be reported to the site's REB/IRB (as per REB/IRB guidelines) for each sub-study, as applicable. All REB/IRB correspondence is to be forwarded to Ozmosis Research. The site must notify Ozmosis Research and/or sponsor immediately of any protocol violations.

#### 18.3 Premature Discontinuation of the Study

The Sponsor reserves the right to discontinue any trial for any reason but intends only to exercise this right for valid scientific or administrative reasons. After such a decision, the Investigators must contact all participating subjects immediately after notification. Follow- up for subjects will be assured and, where required by the applicable regulatory requirement(s), the relevant regulatory authority(ies) will be informed.

The REB/IRB will be informed promptly and provided with a detailed written explanation for the termination or suspension.

As directed by the Sponsor, all study materials must be collected and all CRFs completed to the greatest extent possible.

### 20 APPENDIX A – FDA GUIDANCE ON CLINICAL TRIAL CONDUCT DURING COVID-19 PANDEMIC

FDA U.S. FOOD & DRUG
ADMINISTRATION

**FDA Issues Updated Guidance on Clinical Trial Conduct During the COVID-19 Pandemic**

On March 18, 2020, FDA issued "FDA Guidance on Conduct of Clinical Trials of Medical Products during COVID-19 Pandemic" to provide general considerations to assist sponsors in assuring the safety of trial participants, maintaining compliance with good clinical practice (GCP), and minimizing risks to trial integrity during the COVID-19 pandemic. On March 27, 2020, FDA amended the guidance to include an appendix to further explain those general considerations by providing answers to questions about conducting clinical trials that the Agency has received during the COVID-19 pandemic. For the updated guidance, please see: https://www.fda.gov/regulatory-information/search-fda-guidance-documents/fda-guidance-conduct-clinical-trials-medical-products-during-covid-19-pandemic.

For additional questions regarding clinical trial conduct during the COVID-19 pandemic, please.

U.S. Food and Drug Administration
10903 New Hampshire Avenue, Silver Spring, MD 20993
1-888-INFO-FDA (1-888-463-6332)

Privacy Policy | www.fda.gov
Manage Preferences or Unsubscribe from this List | Unsubscribe from all Email Lists

### 21 APPENDIX B – KDIGO CRITERIA FOR ACUTE KIDNEY INJURY

Acute kidney injury after enrollment is defined by KDIGO criteria for Acute Kidney Injury in the setting of not meeting these criteria upon enrollment:

Three Stages:

**Stage 1:** Serum Cr 1.5-1.9 times baseline, OR ≥ mg/dl increase in serum Cr

**Stage 2:** Serum Cr 2.0-2.9 times baseline

**Stage 3:** Serum Cr ≥ 3.0 times baseline, OR increase in serum creatinine to ≥ 4.0mg/dl, OR initiation of renal replacement therapy

### 22 APPENDIX C – WHO ORDINAL SCALE FOR CLINICAL IMPROVEMENT

(https://www.who.int/blueprint/priority-diseases/keyaction/COVID-19_Treatment_Trial_Design_Master_Protocol_synopsis_Final_18022020.pdf)

| Patient State | Score | Descriptor |
|---|---|---|
| Uninfected | 0 | No clinical or virological evidence of infection |
| Ambulatory | 1 | No limitation of activities |
| | 2 | Symptomatic: Limitation of activities |
| Hospitalized: Mild disease | 3 | Hospitalized; no oxygen therapy |
| | 4 | Hospitalized; oxygen by mask or nasal prongs |
| Hospitalized: Severe disease | 5 | Non-invasive ventilation or high-flow oxygen |
| | 6 | Intubation & Mechanical ventilation |
| | 7 | Ventilation and additional organ support – pressors, RRT, ECMO |
| Death | 8 | Death |

### Protocol 3: A Multicenter, Adaptive, Randomized Controlled Platform Trial of the Safety and Efficacy of Antithrombotic Strategies in Hospitalized Adults with COVID-19 (ACTIV-4 Acute)

#### A Multicenter, Adaptive, Randomized Controlled Platform Trial of the Safety and Efficacy of Antithrombotic Strategies in Hospitalized Adults with COVID-19

**Short Title: ACTIV-4 ACUTE (AC-INPT)**

**ClinicalTrials.gov Number:** NCT04505774

Supported by:

**National Heart Lung and Blood Institutes (NHLBI), National Institute of Neurological Disorders (NINDS), ACTIV (ACTIV IV), National Institutes of Health (NIH), Biomedical Advanced Research and Development Authority (BARDA), Operation Warp Speed (OWS),**

U.S. Department of Health & Human Services (HHS)

Version Number: 1.0

**21AUG2020**

| | |
|---|---|
| **Trial Chair** | Judith Hochman, MD<br>Harold Snyder Family Professor & Associate Director of Cardiology<br>Senior Associate Dean for Clinical Sciences<br>Co-Director, NYU-HHC Clinical and Translational Science Institute<br>NYU School of Medicine<br>530 First Avenue, Skirball 9R<br>New York, NY 10016<br>Tel 212-263-6927<br> |
| **Coordinating Center** | Stephen Wisniewski, PhD<br>Professor of Epidemiology<br>Vice Provost for Budget and Analytics<br>University of Pittsburgh<br><br><br>Matthew Neal, MD<br>Roberta G. Simmons Assistant Professor of Surgery<br>Attending Surgeon, Division of Trauma and Acute Care Surgery<br>Assistant Professor of Clinical and Translational Science and Critical Care Medicine Departments of Surgery, Critical Care Medicine, and the Clinical and Translational Science Institute (CTSI), University of Pittsburgh University of Pittsburgh Medical Center<br>F1271.2 PUH 200 Lothrop Street<br>Pittsburgh, PA 15213<br>Tel: 412-647-1158<br>Fax: 412-647-1448<br> |
| **Trial Biostatisticians** | Scott Berry, PhD<br>President, Senior Statistical Scientist<br>Berry Consultants<br>Tel 979-575-6280<br><br><br>Eric Leifer, PhD<br>NHLBI<br><br><< >> |
| **NHLBI Representative** | Andrei Kindzelski, MD<br> |
| **HHS Representative** | Rachel Harrigan<br> |
| **IND** | Waiver |
| **ClinicalTrials.gov Identifier** | NCT04505774 |

#### Statement of Compliance

In the United States this study will be conducted in accordance with the Code of Federal Regulations on the Protection of Human Subjects (45 CFR Part 46), 21 CFR Parts 50, 56, 312, and 812 as applicable, any other applicable US government research regulations, and institutional research policies and procedures. The International Conference on Harmonisation ("ICH") Guideline for Good Clinical Practice ("GCP") (sometimes referred to as "ICH-GCP" or "E6") and the General Data Protection Regulations (GDPR) will be applied only to the extent that it is compatible with FDA and DHHS regulations.

Outside of the United States this study will be conducted according to local legal and regulatory requirements and regulations, ICH guidelines, and GDPR guidelines as applicable.

The Principal Investigator will assure that no deviation from, or changes to, the protocol will take place without prior agreement from the sponsor and documented approval from the Institutional Review Board (IRB), except where necessary to eliminate an immediate hazard(s) to the trial participants. All personnel involved in the conduct of this study have completed Human Subjects Protection Training.

The signature below provides the necessary assurance that this study will be conducted according to all stipulations of the protocol including statements regarding confidentiality, and according to local legal and regulatory requirements, US federal regulations (if applicable), and ICH E6(R2) GCP guidelines.

Version Date: Aug. 2020

________________________________ ________________________

Signature of Principal Investigator Date

________________________________

Printed Name of Principal Investigator

________________________________

Name of Facility

________________________________

Location of Facility (City, Country)

**List of Abbreviations**

| | |
|---|---|
| AE | Adverse Event/Adverse Experience |
| ARDS | Acute Respiratory Distress Syndrome. |
| AT | Arterial Thrombosis |
| CFR | Code of Federal Regulations |
| CHF | Congestive Heart Failure |
| CrCl | Creatinine Clearance |
| COVID-19 | Coronavirus Disease |
| CRF | Case Report Form |
| CSOC | Clinical Study Oversight Committee |
| DCC | Data Coordinating Center |
| DHHS | Department of Health and Human Services |
| DIC | Disseminated Intravascular Coagulation |
| DSMB | Data and Safety Monitoring Board |
| DVT | Deep Vein Thrombosis |
| ECMO | Extracorporeal Membrane Oxygenation |
| eGFR | Estimated Glomerular Filtration Rate |
| FDA | Food and Drug Administration |
| FFR | Federal Financial Report |
| FWA | Federal Wide Assurance |
| GCP | Good Clinical Practice |
| GI | Gastrointestinal |
| HFNO | High-flow (≥30L/min) Nasal Oxygen |
| HIPAA | Health Insurance Portability and Accountability Act |
| HIT | Heparin Induced Thrombocytopenia |
| ICF | Informed Consent Form |
| ICH | International Conference on Harmonisation |
| ICMJE | International Committee of Medical Journal Editors |
| IRB | Institutional Review Board |
| ISM | Independent Safety Monitor |
| ISTH | International Society on Thrombosis and Haemostasis |
| ITT | Intent to Treat |
| KDIGO | Kidney Disease Improving Global Outcomes |
| LAR | Legally Authorized Representative |

| | |
|---|---|
| LOS | Length of Stay |
| MI | Myocardial Infarction |
| MOP | Manual of Procedures |
| N | Number (typically refers to participants) |
| NIH | National Institutes of Health |
| NIV | Non-invasive ventilation |
| OHRP | Office for Human Research Protections |
| OHSR | Office of Human Participants Research |
| OSFD | Organ Support Free Days |
| PE | Pulmonary Embolism |
| PI | Principal Investigator |
| PRBC | Packed Red Blood Cells |
| PTT | Partial Thromboplastin Time |
| QA | Quality Assurance |
| QC | Quality Control |
| SAE | Serious Adverse Event/Serious Adverse Experience |
| sICH | Symptomatic Intracranial or Intracerebral Hemorrhage |
| SOC | Standard of Care |
| SOP | Standard Operating Procedure |
| US | United States |
| VTE | Venous thromboembolism |
| WHO | World Health Organization |

#### Master Protocol Summary

| | |
|---|---|
| Title | A Multicenter, Adaptive, Randomized, Open Label Controlled Platform Trial of the Safety and Efficacy of Antithrombotic Strategies in Hospitalized Adults with COVID-19 |
| Short Title | ACTIV-4 ACUTE |
| Brief Summary | This is a randomized, open label, adaptive platform trial to compare the effectiveness of antithrombotic strategies for prevention of adverse outcomes in COVID-19 positive inpatients |
| Objectives | **1.** To determine the most effective antithrombotic strategy for increasing the number of days free of organ support and reducing death.<br><br>**2.** To determine the most effective antithrombotic strategy on the composite endpoint of death, deep vein thrombosis (DVT), pulmonary embolism (PE), myocardial infarction (MI), ischemic stroke, or other systemic arterial thrombosis (AT).<br><br>**3.** To assess the safety of antithrombotic strategies through the endpoint of major bleeding as defined by ISTH.<br><br>**4.** To compare the effect of antithrombotic strategies on the endpoint of all-cause mortality in the study population.<br><br>Assessment of efficacy and safety will yield information of the net clinical benefit of different antithrombotic strategies in the study population. It will also yield information on outcomes specific to under- represented minority populations, specifically African- and Hispanic-descent persons. |
| Methodology | Adaptive Randomized Platform Trial |

| | |
|---|---|
| Endpoints | Primary Endpoint: 21 Day Organ Support Free Days, which is defined as the number of days that a patient is alive and free of organ support through the first 21 days after trial entry. Organ Support is defined as receipt of invasive or non-invasive mechanical ventilation, high flow nasal oxygen, vasopressor therapy, or ECMO support, with death at any time (including beyond 21 days) during the index hospitalization assigned -1 days.<br><br>Key Secondary Endpoint: Composite endpoint of death, pulmonary embolism, systemic arterial thromboembolism, myocardial infarction, or ischemic stroke at hospital discharge or 28 days, whichever occurs first.<br><br>Other Secondary Endpoints: Composite endpoint of death, deep vein thrombosis, pulmonary embolism, systemic arterial thromboembolism, myocardial infarction, or ischemic stroke at hospital discharge or 28 days, whichever occurs first. Acute kidney injury defined by KDIGO criteria, Individual endpoints comprising the key secondary endpoint, death during hospitalization, 28 Day Ventilator-Free Days, 28 Day Vasopressor Free Days, 28 Day Renal Replacement Free Days, WHO clinical scale, 28 Day Hospital Free Days, 28 day organ support free days, and all-cause mortality at 90 days.<br><br>Primary Safety Endpoint: Major bleeding (as defined by the ISTH)<br>Secondary Safety Endpoint: Confirmed heparin induced thrombocytopenia (HIT) |
| Study Duration | Approximately 1 year |
| Participant Duration | Hospital duration with periodic contact at post-discharge, including at 90 days, with potential contact up to 1 year |
| Duration of assigned treatment strategy | During hospitalization (unless otherwise specified in description of arm) |
| Population | Adult patients hospitalized for COVID-19 |
| Study Sites | Approximately 400 sites |
| Number of participants | The sample size is described in each arm-specific appendix. |
| Description of Study Agents | Randomized arms- see appendix<br><br>This platform trial allows for multiple therapies to be investigated in this trial over time. The trial is governed by a Master Protocol that describes the trial design, endpoint collection, primary endpoint, and inclusion/exclusion criteria. Different therapies, referred to as arms, are detailed in arm-specific appendices. These arm-specific appendices work in a modular fashion as arms are removed and added to the platform trial. |
| Key Procedures | Observation during hospitalization, contact at 90 days post-enrollment, and collection of standard of care laboratory results. Ancillary biobanking will be completed in consenting patients at capable centers. |

| | |
|---|---|
| Statistical Analysis | Inferences in this trial are based on a Bayesian statistical model, which considers the variation in outcomes by site, disease state, time, and arm of the trial. The specific analyses for each arm, including interim analysis schedule, are specified in each arm-specific appendix. |

### 1 Introduction, Background Information and Scientific Rationale

#### 1.1 *Background Information, Significance and Relevant Literature*

The severe acute respiratory syndrome coronavirus 2, which causes the highly contagious coronavirus disease 2019 (COVID-19), has resulted in a global pandemic.

The clinical spectrum of COVID-19 infection is broad, encompassing asymptomatic infection, mild upper respiratory tract illness, and severe viral pneumonia with respiratory failure and death. The risk of thrombotic complications is increased, even as compared to other viral respiratory illnesses, such as influenza (1-4). A pro-inflammatory cytokine response as well as induction of procoagulant factors associated with COVID-19 has been proposed to contribute to thrombosis as well as plaque rupture through local inflammation (5). Patients with COVID-19 are at increased risk for arterial and vein thromboembolism(6), with high rates observed despite thromboprophylaxis (7). Autopsy reports have noted micro and macro vascular thrombosis across multiple organ beds consistent with an early hypercoagulable state (8).

Notably, in COVID-19, data in the U.K. and U.S. document that infection and outcomes of infection are worse in African and Hispanic descent persons than in other groups. The reasons for this are uncertain.

**Viral Infection and Thrombosis**

A large body of literature links inflammation and coagulation; altered hemostasis is a known complication of respiratory viral infections (9-11). Procoagulant markers are severely elevated in viral infections. Specifically, proinflammatory cytokines in viral infections upregulate expression of tissue factor, markers of thrombin generation, platelet activation, and down-regulate natural anticoagulant proteins C and S (11).

Studies have demonstrated significant risk of deep venous thrombosis (DVT), pulmonary embolism (PE), and myocardial infarction (MI) associated with viral respiratory infections (10,12). In a series of patients with fatal influenza H1N1, 75% had pulmonary thrombi on autopsy (a rate considerably higher than reported on autopsy studies among the general intensive care unit population (13).
Incidence ratio for acute myocardial infarction in the context of Influenza A is over 10 (14). Severe acute respiratory syndrome coronavirus-1 (SARS CoV-1) and influenza have been associated with disseminated intravascular coagulation (DIC), endothelial damage, DVT, PE, and large artery ischemic stroke (11,15). Obi et al. found that patients with Influenza H1N1 and acute respiratory distress syndrome (ARDS) had a 23.3-fold higher risk for pulmonary embolism, and a 17.9-fold increased risk for deep vein thrombosis (16). Compared to those treated with systemic anticoagulation, those without treatment were 33 times more likely to suffer a VTE (16).

Thrombosis, both microvascular and macrovascular, is a prominent feature in multiple organs at autopsy in fatal cases of COVID-19 (8). Thrombosis may thus contribute to respiratory failure, renal failure, and hepatic injury in COVID-19. The number of megakaryocytes in tissues is higher than in other forms of ARDS, and thrombi are platelet-rich based on specific staining. Thrombotic stroke has been reported in young COVID-19 patients with no cardiovascular risk factors (17). Both arterial and venous thrombotic events have been seen in increasing numbers of hospitalized patients infected with COVID-19. The incidence of thrombosis has ranged from 10 to 30% in hospitalized patients; however, this varies by type of thrombosis captured (arterial or vein) and severity of illness (ICU level care, requiring mechanical ventilation, etc.).

D-dimer, a biomarker of fibrin formation and degradation, is elevated in conditions associated with thrombosis, and has been strongly associated with increased mortality among patients with COVID- 19 (1, 2, 3, 6, 7). In a retrospective analysis of 191 patients with COVID-19, Zhou et al. found that non-survivors were more likely to have D-dimer levels > 1 ug/mL than survivors (81% v 24%) (1).
Similarly, in a study of 183 patients, Tang et al. noted that non-survivors had significantly higher D- dimer values on admission than survivors (2.12 v 0.61 ug/mL, P < 0.001) (2). In a retrospective study, patients with COVID-19 and D-dimer values > 6-fold upper limit of normal had lower 28-day mortality when treated with prophylactic anticoagulation compared with no anticoagulation (32.8% v 52.4%, p=0.017) (8). Data suggest a strong association between D-dimer and the outcomes of ICU intubation and all-cause mortality, and the association between D-dimer and (1) mortality, (2) critical illness, (3) acute kidney injury, and (4) thrombotic risk is increased at a D-dimer between 1X to 2X the upper limit of normal. Thrombosis is also increased in those with elevated inflammation indexed by C-reactive protein level (20). Preliminary data suggest that platelet activity is increased in COVID-19(18) and that biomarkers of platelet activity correlate with the incidence of death or thrombosis in hospitalized patients with COVID-19. Platelet-fibrin thrombi have been observed in alveolar capillaries, where they may affect gas exchange (8), and in the renal peri-tubular capillaries, where they may contribute to acute tubular necrosis and renal dysfunction. Consistently, autopsy findings demonstrate an increase in the number of circulating megakaryocytes outside the bone marrow and lung. Finally, thrombotic events have been noted – even among patients treated with full dose anticoagulation.

There may be racial and ethnic differences in response to COVID 19 infection. It is hypothesized that antithrombotic interventions being tested will benefit all patients, including those who are disproportionately affected. (21–25, 26).

<u>The ACTIV-4 ACUTE investigators postulate that an antithrombotic regimen will improve clinical outcomes in COVID-19 patients.</u> This protocol intends to define the optimal regimen in an adaptive randomized trial of patients hospitalized with COVID-19 at risk for adverse clinical outcomes. The primary outcome will be the number of days free of organ support within 21 days after randomization. This primary outcome was selected because thrombosis is thought to contribute to the pathogenesis of multi-organ failure in COVID-19, because it is pragmatic and yet clinically relevant, and to align with ongoing studies that may or may not involve antithrombotic therapy, in a time frame relevant to acute illness. Organ support free days is defined by days in which patient is not on invasive or non-invasive mechanical ventilation, high flow nasal oxygen, vasopressor therapy, or ECMO support (see Appendix 2), with death assigned the value of –1 days.

##### 1.1.1 Adaptive Design

This platform trial will have multiple arms, which may be dropped or added as the platform trial progresses. Sample size will be flexible: the trial will be stopped for efficacy or futility based on pre- determined statistical thresholds as defined in the arm-specific appendix (Appendix 3 and 4). Each arm will have an adaptive component for determinations of futility or success.

##### 1.2 *Potential Risks & Benefits*

See arm-specific Appendices for details

#### 2 Study Design

##### 2.1 *Overall Study Design*

This trial design is built as a process – with the possibility of multiple interventions being investigated. The trial is designed to be flexible, and these flexible aspects are planned as part of the protocol. This trial may incorporate a flexible number of interventions, and the number of interventions may evolve as the science evolves. Intervention arms will be added or dropped based on criteria defined in arm-specific appendices. Co-enrollment in other trials is permitted as long as the other trial does not test agents with antithrombotic properties and there is no other scientific contraindication.

##### 2.2 *Randomization*

Randomization assignments are at the participant level and are assigned at baseline. Randomization will be stratified by enrolling site and may also be stratified by severity of illness and/or other arm-specific criteria. In general, allocation will be equally distributed across arms for which the participant is eligible, but may be altered with future arm-specific appendices.

#### 3 Objectives and Purpose

The overarching objective of this adaptive platform design is to iteratively learn which antithrombotic strategy is the best for reducing the primary, secondary, and safety outcomes. Additional alternative strategy(-ies) will be compared to the current standard of care arm, which may trigger new standard of care designated arms as appropriate based on interim analysis results and evolving literature.

This process will continue until no new strategies replace the standard of care or potential options for additional antithrombotic interventions are exhausted.

### 4 Study Design and Endpoints

#### 4.1 *Description of Study Design*

This trial design is built as a process – with the possibility of multiple interventions being investigated. This is an open label randomized trial of patients hospitalized for COVID-19 who are assigned to different antithrombotic regimens.

#### 4.2 *Study Endpoints*

##### 4.2.1 Primary Study Endpoint

21 Day Organ-Support free-days. The primary endpoint is the number of days that a patient is alive and free of organ support through 21 days after trial entry. Organ support is defined by receipt of invasive or non-invasive mechanical ventilation, high flow nasal oxygen, vasopressor therapy, or ECMO support. If the patient dies at any time (including beyond 21 days) during the index hospital stay, they are assigned the worst possible score of –1.

##### 4.2.2 Secondary Endpoints

- **Key Secondary Endpoint**: A composite endpoint of death, pulmonary embolism, systemic arterial thromboembolism, myocardial infarction, or ischemic stroke during hospitalization or at 28 days after enrollment (whichever is earlier)

###### Other Secondary Endpoints:

- A composite endpoint of death, deep vein thrombosis, pulmonary embolism, systemic arterial thromboembolism, myocardial infarction, or ischemic stroke during hospitalization or at 28 days after enrollment (whichever is earlier)
- 28 Day Hospital free days (non-ICU level patients)
- 28 Day Ventilator-Free Days (ICU level patients)
- 28 Day Vasopressor-Free Days (ICU level patients)
- 28 Day Renal Replacement Free Days
- Hospital readmission within 28 days
- Acute kidney injury as defined by KDIGO criteria
- Deep vein thrombosis
- Pulmonary embolism
- Systemic arterial thrombosis or embolism
- Myocardial infarction
- Ischemic stroke
- Use of extracorporeal membrane oxygenation (ECMO) support
- Mechanical circuit (dialysis or ECMO) thrombosis
- All-cause mortality at 28 days
- Organ support free days at 28 days
- All-cause mortality during initial hospitalization (includes death after 28 days)
- WHO ordinal scale (peak scale over 28 days, scale at 14 days, and proportion with improvement by at least 2 categories compared to enrollment, at 28 days)
- All-cause mortality at 90 days

##### 4.2.3 All-cause mortality at 90 days Additional Study Endpoints

- Individual endpoints of the thrombotic endpoint
- Length of Hospital stay
- Exploratory endpoints (subset of sites)
  - Cardiac injury (e.g., troponin)
  - Trajectories of biomarkers related to COVID-19
  - DIC

See arm-specific Appendices for additional tertiary endpoints of interest specific to arm.

##### 4.2.4 Safety Endpoints

- Major Bleeding (as defined by the ISTH)
- Symptomatic intracranial or intracerebral hemorrhage (evaluated as a separate endpoint from other major bleeding) (19)
- Confirmed Heparin induced thrombocytopenia (laboratory confirmed by anti-PF4 test or Serotonin Release Assay (SRA))

### 5 Study Enrollment

#### 5.1 *Inclusion Criteria*

In order to be eligible to participate in this study, an individual must meet all of the following criteria:

- ≥ 18 years of age
- Hospitalized for COVID-19*
- Enrolled within 72 hours of hospital admittance or 72 hours of positive COVID test
- Expected to require hospitalization for > 72 hours
- (See arm-specific Appendices for additional criteria and details)

*It is strongly recommended to confirm SARSCoV2 with a positive microbiological test prior to randomization. At centers where there is delay in confirming the diagnosis, a sufficiently high clinical suspicion is sufficient to proceed with randomization as long as confirmation is expected within 24 hours.

#### 5.2 *Exclusion Criteria*

- Imminent death
- Requirement for chronic mechanical ventilation via tracheostomy prior to hospitalization
- Pregnancy
- See arm-specific appendices.

#### 5.3 *Vulnerable Subjects*

Critically ill patients with COVID-19 may not have capacity to provide consent. This trial will include participants who have no capacity to consent only if their legal proxy is able to consent on their behalf. It has become increasingly apparent that individuals with COVID-19 are at risk for thrombotic (and bleeding) events. Patients without the capacity to consent for themselves will have a potential for direct benefit by being part of the trial.

Participation in this trial is expected to facilitate careful monitoring of both thrombotic and bleeding endpoints, which may benefit participants.

Capacity assessment will be conducted by the treating physician or an independent medical provider with appropriate expertise based on the standard clinical assessment of capacity and communicated to the study team. Surrogate consent will be provided by the subject's Legally Authorized Representative as defined by local policies and state/country regulations.

Consent will be obtained from the LAR before any study related procedures begin. Participants' capacity will be monitored throughout the study by working with the treatment team. Once the participant regains the capacity to consent, they will be informed of their participation in the study and will have an opportunity to withdraw from further participation in the study. The enrollment of patients without capacity is important because critically ill patients, especially those who are not ambulatory, are at higher risk of developing clotting complications.

#### 5.4 *Strategies for Recruitment and Retention*

Listings of patients admitted to the participating sites with COVID-19 may be reviewed for eligibility by the study team, to identify and recruit potential participants, until study enrollment goals have been met. The study team should communicate with the inpatient care team. All treating physicians will be informed of the study and will have the option to advise of any conditions that would preclude any individual patient being approached.

#### 5.5 *Duration of Study Participation*

Duration of study participation is, 90 days from enrollment. Participants may be contacted for follow-up for approximately one year.

**Total Number of Participants**

The total sample size for the Platform trial is not pre-determined. The sample size for each arm will be set in the arm-specific appendix and will incorporate an adaptive design. There will be interim monitoring to allow early stopping for futility, efficacy, or safety. If one strategy proves to be efficacious, then this strategy may become the reference arm for comparison(s) with new experimental treatment(s). New arms can be introduced according to scientific and public health needs. Some arms may not relate solely to antithrombotic therapy.

#### 5.6 *Participant Withdrawal or Termination*

##### 5.6.1 Reasons for Withdrawal or Termination

Participants are free to withdraw from participation in the study at any time upon request. Discontinuation of a study agent, regardless of the reason, e.g. patient or physician request, or adverse event, does not constitute study withdrawal. Patient data will still be collected as planned and analyzed as intent to treat unless the participant withdraws consent for continued follow-up. An investigator may terminate participation in the study if:

- Any situation occurs such that continued participation in the study would not be in the best interest of the participant

#### 5.7 *Premature Termination or Suspension of Study*

All deaths and DSMB-specified severe adverse events within the study period will be reviewed by the DSMB. The decision to stop or suspend the study, or an arm of the study, will be made by the DSMB after considering the totality of the data and the benefit-risk of continuing the study.

This study, or an arm of the study, may be temporarily suspended or prematurely terminated if there is sufficient reasonable cause.

Circumstances that may warrant termination or suspension include, but are not limited to:

- Determination of unexpected, significant, or unacceptable risk to participants in a strategy, such as excess mortality and/or major bleeding (this will be determined by the oversight data safety monitoring plan; See 7.4.7)
- Demonstration of efficacy or lack thereof that would warrant stopping (See 7.4.7)
- Insufficient compliance to protocol requirements
- Data that are not sufficiently complete and/or evaluable
- Determination of futility

The study may resume once concerns about safety, protocol compliance, data quality are addressed and satisfy the sponsor, IRB and/or FDA.

### 6 Study Agent and Procedural Intervention

#### 6.1 *Study Agents*

Each arm in this platform trial will include different treatment strategies. Information about the treatment strategies for a given arm can be found in the arm -specific appendices.

#### 6.2 *Duration of Therapy*

Once participants are randomized to a treatment strategy (arm), they will remain on treatment for the duration specified by the relevant appendix. However, if a participant randomized to one arm develops an indication for a different strategy (e.g., thrombotic event, worsening clinical status), the participant will be treated based on institutional guidelines with any measures required by local clinical judgment.

### 7 Study Procedures and Schedule

#### 7.1 *Study Schedule*

| **Activity** | **Screening/ Enrollment** | **Hospital Duration** | **28 days and/or hospital discharge*** ** | **90-days post randomization** |
|---|---|---|---|---|
| **Eligibility** | | | | |
| Consent | X | | | |
| Demographic and Medical History | X | | | |
| Assessment of Inclusion/Exclusion criteria | X | | | |
| Self-reported race/ethnicity and gender | X | | | |
| **Study Drug Administration** | | | | |

| | | | | |
|---|---|---|---|---|
| Randomization | X | | | |
| Study treatment | X | X | | |
| **Study Procedures** | | | | |
| Height | X | | | |
| Weight | X | | | |
| Vital signs | X | | | |
| Concomitant medications | X | X | | |
| WHO ordinal assessment | X | X | X | X |
| Outcomes Assessment | | X | X | X |
| **SOC Laboratory Assessments** | | | | |
| Chemistry panel | X | X | | |
| Hematology panel | X | X | | |
| See arm-specific appendix for additional measures | | | | |
| D-dimer* | X | | | |
| Blood Group** | X | | | |

*D-dimer is strongly recommended for measurement in all participants as close to the time of randomization as feasible.
**Blood group will come from hospital record or self report if available. Biospecimens see appendix 4.
***Assessments indicated in the table above will be ascertained at discharge, or at 28 days, whichever comes first. Participants must be followed for vital status until discharged from the hospital or another care facility (if transferred on organ support) up to 90 days. To maximize retention, participants will be contacted intermittently (e.g. at one and two months post-discharge)

**Laboratory Procedures/Evaluations**

See arm specific appendices.

All analyses will be performed on SOC labs and procedures done for usual care. The standard operating procedures for samples to be collected for research purposes are included as Appendix
5. All research samples will be timed with clinical lab draws to limit provider exposure. Collection of research samples as outlined in Appendix 5 is strongly encouraged where safe and feasible.

##### 7.1.1 Visit 1 and Hospitalization Visits (see arm-specific appendices for details)

*Visit 1 (Screening and Randomization)*

1. Informed consent obtained
2. Assessment of inclusion/exclusion criteria assessed
3. Screening, consisting of reviewing participant medical history and information in their chart such as height, weight, vital signs, and normal clinically performed laboratory assessments, including pregnancy test for all women of childbearing age.
4. If confirmed eligible, following randomization, initiation of treatment with the assigned strategy

##### *Hospitalization Visits*

1. Recording of specifics of study treatment according to assigned arm
2. Laboratory assessments as part of standard of care
3. Daily WHO ordinal assessment
4. Ongoing daily outcomes and safety assessment

##### 7.1.2 28 days and/or Date of Hospital Discharge

1. Recording of outcomes and safety assessments as reported by participant or observed by investigator
2. WHO Ordinal Assessment
3. Recording of vital status and ascertainment of events
4. Recording of participant's adherence to treatment strategy, if patient is in hospital

These assessments will be ascertained at discharge, or at 28 days, whichever comes first. Participants must be followed for vital status until discharged from the hospital or another care facility (if transferred on organ support) up to 90 days.

Participants may be contacted by a research contact and/or by the participating hospital study team periodically for longer term follow-up for approximately a year. To maximize retention, participants will be contacted intermittently (e.g. at one and two months post-discharge). Discharge visits must be completed.

#### 7.2 *Concomitant Medications, Treatments, and Procedures*

Concomitant medications taken during study participation will be recorded on the case report forms (CRFs). Concomitant medications to be recorded are:

- Other antithrombotics (e.g., aspirin and other antiplatelet agents)
- Any medications used for the treatment of COVID-19 infection (e.g., remdesivir, steroids, IL- 6 inhibitor such as tocilizumab)
- Others specified in arm-specific appendices

#### 7.3 *Expedited Critical and Major Event Reporting*

All efficacy and safety outcome events will be assessed and documented in the participants' study records. The ACTIV-4 Platform will have a uniform policy for reporting adverse events to ensure that all events are assessed quickly and are submitted to the DSMB, IRB(s), and other groups as needed (e.g., FDA), following each group's reporting guidelines and timelines. Events meeting the independent DSMB-specified criteria will be reported immediately and within the time frames specified by the DSMB.

Sites are required to follow their local reporting guidelines.

#### 7.4 *Data and Safety Monitoring Plan and Study Halting Rules*

The ACTIV-4 Platform will have a uniform Data and Safety Monitoring Plan, encompassing all research carried out within the Platform.

### 8 Statistical Considerations

#### 8.1 *Statistical and Analytical Plans (SAP)*

There will be a formal Statistical Analysis Plan (SAP) and each arm added to the trial will have its own arm-specific SAP. This will include the primary analysis, the primary comparison, futility and success rules, and interim analysis schedule. The SAP will be created prior to the first interim analysis for the study and each arm-specific SAP will be created before the first interim analysis for that arm.

#### 8.2 *Statistical Modeling for the Primary Analysis*

Inferences in this trial are based on a Bayesian statistical model for the ordinal primary outcome, organ-support free-days (OSFD). There is a single Bayesian model for the primary outcome across each arm and subpopulation. The Bayesian model is an ordinal cumulative logistic regression model described below.

Let $Y_i = \{-1, 0, 1, ..., 21, 22\}$ denote the ordinal outcome (OSFD) for patient $i$. The probability of patient $i$ observing $y$ OSFD or less is denoted as $\pi_{iy} = \Pr(Y_i \leq y)$. The parameters in the model are structured so that a value > 0 implies treatment benefit, and hence an odds-ratio > 1 implies treatment benefit. The generic primary analysis model is formulated as follows:

$$\log\left(\frac{\pi_{iy}}{1-\pi_{iy}}\right) = \alpha_{y,s} - [\nu_{Site,s} + \lambda_{Time,s} + \theta_{a,s:d} + \beta_{Age,s} + \beta_{Sex,s} + \beta_d]$$

1. The “subtype” variable, $s$, corresponds to the two patient subgroups defined by disease severity:
    a. subtype = 1 is non-ICU level care
    b. subtype = 2 is ICU-level care
2. The d-dimer level for a patient, d, is classified for a patient as
    a. d=1 is a low or unknown d-dimer level
    b. d=2 is a high d-dimer

The d-dimer level is only used for non-ICU (s=1) patients. We use the notation s:d to imply the parameterization would be s=1, d=1 (non-ICU level care, low d-dimer); s=1, d=2 (non-ICU level care, high d-dimer); and s=2 (ICU care).

3. The “site” variable is the clinical site within the trial. These will be site effects estimated separately within the non-ICU and ICU level of case disease states, but not varying by d- dimer levels.
4. The “time” variable is an indicator of the month of enrollment in the trial, numbered decreasing from the first enrollment to the last enrollment for the analysis. The time effects will be estimated separately within the non-ICU and ICU level of case disease states, but not varying by d-dimer levels.
5. The “arm” the patient is randomized to is labeled as *a*. The effects of arm are modeled by both the disease state and the d-dimer level.
6. The “age” variable is a categorical classification of age as ≤ 39, 40-49, 50-59, 60-69, 70-79, and 80+. The age effects will be estimated separately within the non-ICU and ICU level of case disease states, but not varying by d-dimer levels.
7. The “sex” variable is sex at birth. The sex effects will be estimated separately within the non-ICU and ICU level of case disease states, but not varying by d-dimer levels.

If additional covariates (e.g. race and ethnicity) are added to the model they will by default, unless otherwise specified, vary by disease state, but not d-dimer levels.

The $\alpha_{y,subtype}$ parameters are the baseline rates of the ordinal outcome, which are modeled separately by disease subtype. The additive effects of d-dimer levels are modeled with the $\beta_d$.

#### 8.3 *Model Priors*

The treatment effects for arm *a*, within disease subtype *s* and d-dimer level *d* are modeled with the $\theta_{a,s:d}$ parameters. The $\beta$ parameters model any covariate effects included in the model. The $\lambda$ parameters model the effect of time within the pandemic.

The ordinal endpoint rates are modeled using an inverse Dirichlet model where the individual probabilities for the 24 outcomes are based on 10 patients weight on real-world evidence-based outcomes (details in the SAP).

$$logit(\alpha_{y,s}) \sim Dirichlet(10 * P), where \ldots$$

The site effects, $\nu_{Site}$, are modeled using a hierarchical model where site is nested within the country of the site:

$$\nu_{Site,s} \sim N(\mu_{country,s}, \tau^2_{country,}\ ), site = 2, \ldots, N_{Site}$$

$$\mu_{country,s} \sim N(0,1);\ \tau^2_{country,} \sim IG(0.25,0.1),\ s = 1,2$$

A referent site, expected to be the largest enrolling site, will be set such that $\nu_{Site} \equiv 0$. The hyper- parameters of the site hierarchical model are separate by disease state *s*.

The effect of time (*T*) is modeled using a second-order normal dynamic linear model separately by disease state, *s*. The most recent two time periods are modeled as the referent time epochs with the time parameters set to 0. The preceding time epochs are modeled as a normal dynamic linear model as:

$$\lambda_1 = \lambda_2 \equiv 0$$

$$\lambda_3 \sim N(0, 0.15^2)$$

$$\lambda_T - 2\lambda_{T-1} + \lambda_{T-2} \sim N(0, \tau^2_{Time}),\ T \geq 4$$

The treatment effect parameters are set against a control arm, which will be labeled in the arm- specific appendix. The treatment effect for the control arm, labeled as arm *a* = 1, will be set to 0 for each of the disease subtype and d-dimer level:

$$\theta_{1,subtype,d} \equiv 0$$

The effect of each other arm introduced will be modeled hierarchically across disease subtype and d-dimer level. The treatment effects for each arm, *a*, are modeled as:

$$\theta_{a,s:d} \sim N(\mu_a, \tau^2_a),\ s:d = 1{:}1, 1{:}2, 2, \qquad a = 1, \ldots, N_{arms}$$

$$\mu_a \sim N(0,1);\ \tau^2_a \sim IG(0.25, 0.1)\ \text{(TBD)}$$

Any additional covariates included in the model will have independent N(0,1) priors unless otherwise specified.

#### 8.4 *Assessing Effectiveness*

The treatment effect parameters, $\theta$, represent the log-odds ratio, of the treatment, for the cumulative logistic for the ordinal model. In this parametrization an odds ratio > 1, or a log-odds ratio > 0, signifies improved outcomes relative to the referent control treatment. The odds-ratio parameter $\exp(\theta)$, labeled OR, will be used to summarize the treatment effect relative to control or $\exp(\theta_{a_1} - \theta_{a_2})$ for the odds-ratio between arms $a_1$ and $a_2$. The posterior mean, median, standard deviation, and 95% credible intervals for the odds-ratio will be used to summarize relative treatment effects.

The posterior probability that an arm, $a_1$, is superior to another arm, say, $a_2$, is:

$$\Pr(\theta_{a_1} > \theta_{a_2}).$$

This probability will be used for triggers of superiority of one arm to another arm.

The posterior probability that an arm, $a_1$, is superior to another arm, say, $a_2$, by a specified difference on the odds-ratio scale is:

$$\Pr(\exp(\theta_{a_1}) > \exp(\theta_{a_1}) + \delta).$$

This probability will typically be used for futility. If the probability is small that a treatment has benefit above a control of some specified amount ($\delta$), the arm may be dropped for futility.

#### 8.5 *Analysis Datasets*

The intention-to-treat (ITT) analysis dataset will be the source of data for primary analyses. This will include all randomized participants regardless of actual receipt or compliance with therapy. The safety analysis set will consist of all participants who received at least one dose of study medication. The per protocol analysis will be conducted based on adherence to assigned treatment; this dataset will support sensitivity analyses to complement the primary ITT analyses.

The ITT group for an arm consists of the participants that were randomized in the platform that were eligible to be randomized to that arm. This may vary from the platform ITT population, which consists of all participants randomized.

Participants who are randomized to receive one strategy may in fact be treated with another strategy based on health status and provider discretion. Exploratory analyses will estimate the causal effect of the treatment for these participants using marginal structural modelling techniques. These techniques use inverse probability weighting methods that are based on patient-level covariates to create comparable groups for the analysis.

##### 8.5.1 Safety Analyses

Monitoring for safety will be conducted continuously. For each arm-specific appendix potential adverse events of importance will be identified. A Bayesian monitoring rule will be used to summarize the adverse event rates across all arms for the adverse events of importance within each arm-specific appendix. A Bayesian prior distribution of a beta (0.1, 0.9) will be used to model the likelihood of each adverse event of importance. For each adverse event of importance, the posterior mean event rates, the posterior mean of the difference between each arm, and the 95% credible intervals for the risk-difference and odds-ratio will be summarized.

##### 8.5.2 Adherence and Retention Analyses

The primary analysis is by intention to treat. Per protocol analysis will be conducted based on adherence to assigned treatment. For any scheduled follow-up post hospital discharge every effort will be made to recontact participants who are unreachable. Due to the short trial participation timeline, excellent patient retention is anticipated.

##### 8.5.3 Baseline Descriptive Statistics

All variables will be summarized using mean, median, standard deviation, and range (for continuous variables) and frequency (for categorical variables). Treatment groups will be compared with respect to baseline characteristics to verify randomization balance.

##### 8.5.4 Planned Interim Analysis

An independent data safety and monitoring board (DSMB) will review all interim analyses prepared by an unblinded statistical analysis committee.

###### 8.5.5 Safety Review

Monitoring for safety will be conducted continuously. The DSMB will be monitoring safety for each arm-specific appendix. The DSMB monitoring plan includes guidance on stopping specific arms for safety concerns.

###### 8.5.6 Tabulation of Individual Response Data

The composite outcome evaluated will be tabulated and broken down by component (e.g., death, pulmonary embolus, symptomatic DVT, myocardial infarction, etc.). Note that some participants may experience more than one component of the primary endpoint.

###### 8.5.7 Exploratory Analyses

Exploratory analyses will be conducted in a subset of participants on whom additional clinical and basic science assays are performed. These will be descriptive and hypothesis-generating.

##### 8.6 *Sample Size*

Sample size for the platform trial is not pre-determined. The platform trial will run as long as there is a need and there are investigational arms enrolling. The sample size for each arm will be specified in the arm-specific appendix. Interim analyses for each arm will take place in the platform trial and detailed in the arm-specific appendix. Conclusions of futility or superiority may be drawn specific to a patient subtype. Effort will be taken to conduct all interim analyses at the same time in the platform trial since there is a single Bayesian model of the efficacy of all arms conducted. If one strategy proves to be efficacious, then this strategy may become the reference arm for comparison(s) with new experimental treatment(s). New arms can be introduced according to scientific and public health needs.

Generic sample size calculations for an ordinal endpoint of 21-day OSFD with a maximum sample size of 1000 for an investigational arm, compared to a second control arm with 1000 participants (2000 participants total), yields over 80% power for an odds-ratio change of 1.25 on the OSFD endpoint. An odds ratio of 1.5 has approximately 90% power for 400 participants per arm. An odds-ratio of 2 results in more than 90% power for the first interim analysis of 200 participants per arm.

The following figure presents the assumptions for the ordinal outcome of organ-support free days for the control arm and the distribution under each assumed treatment effect for the cumulative odds-ratio used for these power calculations. The second figure presents the power for each assumed effect size for each fixed sample size (the x-axis is the total number on the two arms).

#### 9 Measures to Minimize Bias

##### 9.1 *Enrollment/Randomization*

**Enrollment**

1. Patients hospitalized for COVID-19 are screened daily within the eligibility time window for inclusion/exclusion criteria. Any patient who meets all inclusion criteria and no exclusion criteria will be approached for enrollment.

2. Patients remain in the intention-to-treat group if they meet the criterion for another treatment strategy after randomization.

### 10 Randomization

Randomization assignments are performed for participants at baseline. Randomization will be equal across all arms a patient is eligible. Randomization stratification will be done by site, and disease subtype (ICU and non-ICU level care).

### 11 Source Documents and Access to Source Data/Documents

The ACTIV-4 Platform will have uniform policies describing what source documents are, how to make corrections, and who can access them.

### 12 Quality Assurance and Quality Control

The ACTIV-4 Platform will have uniform policies for quality assurance at the data entry level and site monitoring.

### 13 Ethics/Protection of Human Subjects

#### 13.1 *Ethical Standard*

The investigator will ensure that this study is conducted in full conformity with Regulations for the Protection of Human Subjects of Research codified in 45 CFR Part 46, 21 CFR Part 50, 21 CFR Part 56, and/or the ICH E6.

#### 13.2 *Institutional Review Board*

The protocol, informed consent form(s), recruitment materials, and all participant materials will be submitted to the IRB for review and approval. Approval of both the protocol and the consent form must be obtained before any participant is enrolled. Any amendment to the protocol will require review and approval by the IRB before the changes are implemented to the study. All changes to the consent form will be IRB approved; a determination will be made regarding whether previously consented participants need to be re-consented.

#### 13.3 *Informed Consent Process*

##### 13.3.1 Consent/Assent and Other Informational Documents Provided to Participants

Consent forms describing in detail the study agent, study procedures, and risks are given to the participant, and written documentation of informed consent is required prior to starting intervention/administering study product.

A written consent will be sought from every participant via a face to face consenting process or remotely by using an e-consent option as per IRB approved method.

##### 13.3.2 Consent Procedures and Documentation

Informed consent is a process that is initiated prior to the individual's agreeing to participate in the study and continues throughout the individual's study participation. Informed consent will be obtained following institutional COVID policy to protect study staff.

An extensive discussion of risks and possible benefits of participation will be provided to the participants and their families. Consent forms will be IRB-approved and the participant will be asked to read and review the document. The investigator will explain the research study to the participant and answer any questions that may arise. Participants will have the opportunity to carefully review the written consent form and ask questions prior to signing. The participants should have the opportunity to discuss the study with their surrogates or think about it prior to agreeing to participate. The participant will sign the consent document prior to any procedures being done specifically for the study. The participants may withdraw consent at any time throughout the course of the trial. A copy of the signed informed consent document will be provided to participants. The rights and welfare of the participants will be protected by emphasizing to them that the quality of their medical care will not be adversely affected if they decline to participate in this study.

Participants who have no capacity to consent for themselves will have a surrogate consenting process via legally authorized representative.

##### 13.4 *Posting of Clinical Trial Consent Form*

The informed consent form will be posted on the Federal website after the clinical trial is closed to recruitment, and no later than 60 days after the last study visit by any subject, as required by the protocol.

##### 13.5 *Participant and Data Confidentiality*

The ACTIV-4 Platform will have uniform policies for protecting the privacy of participants and maintaining confidentiality. These policies will adhere to the requirements of the Health Insurance Portability and Accountability Act of 1996 (HIPAA).

#### 14 Data Handling and Record Keeping

##### 14.1 *Data Collection and Management Responsibilities*

The ACTIV-4 Platform will have uniform policies for data management.

##### 14.2 *Study Records Retention*

The ACTIV-4 Platform will have uniform policies for records retention.

##### 14.3 *Protocol Deviations*

A protocol deviation is any noncompliance with the clinical trial protocol, GCP, or Manual of Procedures (MOP) requirements. The noncompliance may be either on the part of the participant, the investigator, or the study site staff. As a result of deviations, corrective actions are to be developed by the site and implemented promptly.

It is the responsibility of the site PI/study staff to use continuous vigilance to identify and report deviations.

Protocol deviations must be reported to the local IRB per their guidelines. The site PI/study staff is responsible for knowing and adhering to their IRB requirements. Further details about the handling of protocol deviations will be included in the MOP.

##### 14.4 *Publication and Data Sharing Policy*

The ACTIV-4 Platform will have uniform policies for publications and data sharing.

#### 15 Study Finances

##### 15.1 *Funding Source*

National Institutes of Health

##### 15.2 *Costs to the Participant*

Participant health insurance may be billed for the costs of medical care during this study since these expenses would have happened even if the participant were not in the study. If the participant's insurance does not cover these costs or the participant does not have insurance, these costs will be participant's responsibility.

#### 16 Conflict of Interest Policy

The ACTIV-4 Platform will have uniform policies for identifying and disclosing potential conflicts of interest.

#### Appendix 1: Criteria for Addition and Replacement of Arms

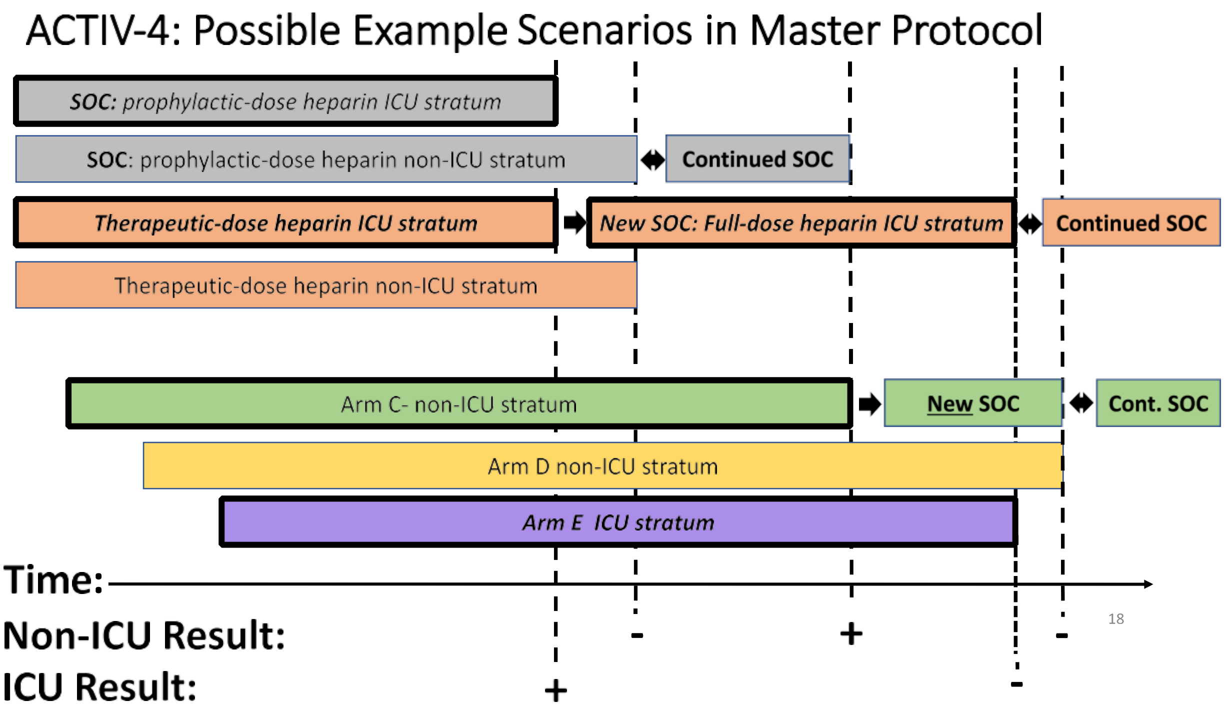

### Appendix 2: Definition and Determination of Outcomes

#### *A2.1 Approach to ascertainment and verification of outcomes*

Outcomes are assessed locally and will not be centrally adjudicated in this pragmatic trial platform, except as specified in the arm-specific appendix. Outcomes should be assessed by a local investigator or other qualified study team member who is blinded to treatment assignment, using the definitions below.

#### *A2.2 Outcome definitions*

**21 Day Organ-Support Free-Days (OSFD)**

Defined as the number of days that a patient is alive and free of organ support through 21 days after trial entry. Organ support is defined by receipt of invasive or non-invasive mechanical ventilation, high flow nasal oxygen, vasopressor therapy, or ECMO support. If the patient dies at any time (including beyond 21 days) during the index hospital stay, they are assigned the worst possible score of –1.

- Non-invasive mechanical ventilation is defined as BIPAP or CPAP when used for acute respiratory support (the use of BIPAP or CPAP at night or when sleeping for sleep apnea is not considered organ support).
- High Flow Nasal Cannula Oxygen is defined as delivery of oxygen through a system that typically delivers oxygen at 30 to 60 liters per minute (but may be as low as 20 liters per minute) with a titratable FiO2.
- Invasive mechanical ventilation is defined as positive pressure ventilation through endotracheal tube or tracheostomy.
- Vasopressor support includes infusion of any vasopressor or inotropic medication.
- A patient must be extubated and not receiving mechanical ventilation for at least 2 days before being considered free of mechanical ventilation. If a patient was extubated and re- intubated and placed back on mechanical ventilation within 1 or 2 days, the patient is considered to be on mechanical ventilation during those 1 or 2 days before re-intubation.
- Any patient dying in the acute hospital stay (even if beyond day 21) are assigned 21 Day Organ-Support Free Days of –1.
- If there is intervening time in which a patient is free of organ support but goes back on organ support the intervening time does not count toward the organ support free days endpoint. Only time before organ support and after the last use of organ support are counted as "free days."
- If a patient was discharged alive without mechanical ventilation prior to Day 21, the patient is assumed to be free of organ support after hospital discharge for the remainder of the 21 days.
- If a patient was discharged alive on mechanical ventilation prior to Day 21, a call to the discharge facility is needed to confirm ventilation status on Day 21 and the last day on mechanical ventilation.

#### Primary Endpoint

Days free of organ support within 21 days after randomization. Organ support free days (OSFD) is defined as days in which patient is not on invasive or non-invasive mechanical ventilation, high flow nasal oxygen, or vasopressor therapy or ECMO support. If the patient dies at any time (including beyond 21 days) during the index hospital stay, they are assigned the worst possible score of –1.

To be specific about which organ support was affected, secondary outcomes include: ventilator free days, renal replacement free days, vasopressor free days.

Justification for use of OSFD:

- Pragmatic
- Can be calculated from WHO ordinal scores
- Incorporates clinically important need for organ support but also duration of organ support
- No additional data collection is necessary to calculate secondary outcomes of vent free days, renal replacement free days, and vasopressor free days to understand which organ support was most impacted
- Incorporates mortality as the worst possible outcome

#### Deep vein thrombosis

Deep vein thrombosis will be diagnosed by venous ultrasound or point-of-care ultrasound (POCUS) or other imaging modality and documented in a note, and performed for clinical indications. A positive ultrasound test is defined by a noncompressible or partially noncompressible venous segment and should be reported. Thrombosis may involve the cerebral venous sinus or any venous bed, including the upper extremities. Routine screening for deep vein thrombosis is not recommended. If deep vein thrombosis is diagnosed and treated without imaging due to imaging availability concerns or risk of exposure to SARS CoV-2, this will be classified as probable deep vein thrombosis. Later imaging is preferable in these cases when possible.

#### Pulmonary embolism

Pulmonary embolism will be confirmed by chest CT with PE protocol, pulmonary angiography or ventilation-perfusion scan. Events may also be defined without this imaging by the care team, as evidenced by, for example, "clot in transit" on echocardiogram. If PE is diagnosed and treated without imaging due to imaging availability concerns or risk of exposure to SARS CoV-2, this will be classified as probable PE. Later imaging is preferable in these cases when possible.

#### Stroke/ Peripheral Arterial Systemic Thromboembolism

Stroke or systemic embolism as diagnosed by imaging (i.e., head CT, lower extremity CT angiogram) or deemed "highly-likely" by the provider based on physical examination (i.e., acute hemiplegia thought to be due to stroke, acute distal lower extremity hypoperfusion). Systemic thromboembolism may involve the retinal artery, spinal cord or other vascular beds. Classification of ischemic vs. other etiologies is based on neuroimaging. Venous sinus thrombosis will be included in the category of vascular occlusion/ischemic stroke on the venous side. Primary CNS hemorrhage: Intracerebral hemorrhage, Subarachnoid hemorrhage, Subdural hematoma, rarely- epidural hematoma, spinal hematoma. Secondary hemorrhagic stroke: Ischemic infarct containing blood - often subclassified by size- PH1, PH2, PH3.

#### ICU Level of care disease state

Defined as planned admission to ICU or receipt of organ support as defined in the 21-day organ support free days.

#### Myocardial infarction

Myocardial infarction is defined according to the universal definition of MI, which excludes myocardial injury e.g., isolated elevation of cardiac troponin. MI must include rise and fall of cardiac troponin above the 99th percentile with at least one of the following: symptoms of acute ischemia, ECG changes consistent with ischemia, new/presumed new wall-motion abnormalities or other imaging evidence of MI, abnormal coronary angiography (e.g. identification of a coronary thrombus).

#### Acute Kidney Injury

Acute kidney injury after enrollment is defined by KDIGO criteria for Acute Kidney Injury in the setting of not meeting these criteria upon enrollment:

THREE STAGES:

- Stage 1: Serum Cr 1.5–1.9 times baseline, OR ≥ 0.3 mg/dl increase in serum Cr
- Stage 2: Serum Cr 2.0–2.9 times baseline
- Stage 3: Serum Cr ≥ 3.0 times baseline, OR Increase in serum creatinine to ≥ 4.0mg/dl, OR Initiation of renal replacement therapy

#### Disseminated Intravascular Coagulation (DIC) (Overt) – DIC score ≥ 5

1. Platelet count ≥ 100 K (0); 50–100K (1 point); < 50K (2 points)
2. Elevated D-dimer: no increase (0 points); moderate increase (1 point); severe increase (3 points) according to local criteria.
3. Prolonged PT < 3 seconds (0 points); 3–6 seconds (1 point); ≥ 6 seconds (2 points)
4. Fibrinogen level ≥ 100 (0 points); < 100 (1 point) mg/dL

#### ISTH Defined Major Bleeding

Bleeding that:

1. Resulted in death,
2. Occurred in a critical organ (intracranial, intraspinal, intraocular, retroperitoneal, intraarticular, intramuscular with compartment syndrome, or pericardial), or
3. Associated with either a decrease in the hemoglobin level of at least 2 g per deciliter or a transfusion of at least 2 units of packed red cells

#### Symptomatic Intracranial or Intracerebral Hemorrhage (sICH)

sICH is defined as any acute extravasation of blood into the brain parenchyma, subarachnoid space, subdural space, or epidural space as demonstrated by imaging or autopsy, associated with any clinical deterioration or death

**WHO ordinal scale for clinical improvement** (https://www.who.int/blueprint/priority-diseases/key-action/COVID-19_Treatment_Trial_Design_Master_Protocol_synopsis_Final_18022020.pdf)

| Patient State | Score | Descriptor |
|---|---|---|
| Uninfected | 0 | No clinical or virological evidence of infection |
| Ambulatory | 1 | No limitation of activities |
| | 2 | Symptomatic: Limitation of activities |
| Hospitalized: Mild disease | 3 | Hospitalized; no oxygen therapy |
| | 4 | Hospitalized; oxygen by mask or nasal prongs |
| Hospitalized: Severe | 5 | Non-invasive ventilation or high-flow oxygen |

| | | |
|---|---|---|
| disease | 6 | Intubation & Mechanical ventilation |
| | 7 | Ventilation and additional organ support – pressors, RRT, ECMO |
| Death | 8 | Death |

**Appendix 3: Therapeutic-dose Anticoagulation (Arm A)**

Any of the following strategies are recommended for therapeutic-dose anticoagulation:

#### *A3.1 Therapeutic Dose Anticoagulation***

<table>
<tr><th>CrCl</th><th>BMI</th><th>Enoxaparin</th><th>Dalteparin</th><th>Tinzaparin</th><th>Heparin</th></tr>
<tr><td rowspan="2">≥30</td><td><40</td><td>1 mg/kg SC q12h<br>OR<br>1.5 mg/kg SC q24h</td><td>200 units/kg SC q24h<br>OR<br>100 units/kg SC q12h</td><td rowspan="2">175 units/kg SC q24h</td><td rowspan="2">IV bolus, with continuous infusion to titrate to anti-Xa 0.3-0.7 IU/mL or corresponding aPTT values*</td></tr>
<tr><td>≥40</td><td>1 mg/kg SC q12h</td><td>100 units/kg SC q12h</td></tr>
<tr><td rowspan="2"><30</td><td><40</td><td colspan="4" rowspan="2">Heparin IV bolus, with continuous infusion to titrate to anti-Xa 0.3-0.7 IU/mL or corresponding aPTT values*</td></tr>
<tr><td>≥40</td></tr>
</table>

* Initial bolus dose determined by sites, encouraging use of dosing algorithm designed for treatment of VTE. UFH anti-Xa titration is preferred over aPTT if available because achieving a therapeutic aPTT may be challenging in patients with a pro-inflammatory state such as COVID-19.

Note: Tinzaparin commonly used in Canada

Note: Fondaparinux not advised in this setting due to its long half life

**These drugs are considered standard of care as an anticoagulants (1). Different drugs are used in different regions, countries, and hospital formularies. In this pragmatic trial of antithrombotic therapy in COVID-19, sites will use the anticoagulant that they typically use in the hospital setting.

It is recommended that participants be given therapeutic-dose parenteral anticoagulation daily for at least 14 days or until hospital discharge, whichever comes first. Treatment may continue beyond 14 days at the discretion of the most responsible physician. At the time of treatment discontinuation, standard of care antithrombotic prophylaxis should be administered.

If there is transfer to ICU level care, continue assigned treatment unless there are contraindications.

#### *A3.2 Discontinuation of study intervention:*

Patients randomized based on suspicion of COVID 19 whose tests do not confirm SARS CoV2 infection should not continue to receive study assigned therapeutic dose anticoagulation.

Anticoagulation should be discontinued if there is clinical bleeding or other complications sufficient to warrant cessation in the opinion of the treating clinician. Major bleeding, including death due to bleeding, is an SAE. Assigned treatment may be resumed if deemed appropriate by the treating clinician.

Occurrence of HIT must result in the cessation of UFH or LMWH without recommencement regardless of treatment assignment. The use of an acceptable alternative agent is required in this instance as clinically indicated. Occurrence of HIT is an SAE.

Study interventions can be discontinued at any time by the treating clinician if doing so is regarded as being in the best interests of the patient. Temporary cessation – for the shortest period of time possible, but not longer than 24 hours – such as to allow surgical or other procedures is not a protocol deviation.

Temporary or permanent cessation of study intervention for bleeding is not a protocol deviation.

##### *A3.3 Study Schedule*

| **Activity** | **Screening/ Enrollment** | **Hospital Duration** | **28 days and/or hospital discharge+** | **90 days post randomization** |
|---|---|---|---|---|
| **Eligibility** | | | | |
| Consent | X | | | |
| Demographic and Medical History | X | | | |
| Assessment of Inclusion/Exclusion criteria | X | | | |
| Self-reported race/ethnicity and sex | X | | | |
| Pregnancy Test, for women of childbearing potential | X | | | |
| **Study Drug Administration** | | | | |
| Randomization | X | | | |
| Study treatment | X | X | | |
| **Study Procedures** | | | | |
| Height | X | | | |
| Weight | X | | | |
| Vital signs | X | X | | |
| Concomitant medications | X | X | | |
| WHO ordinal assessment | X | X | X | X |
| Outcomes assessment | | X | X | X++ |
| **SOC Laboratory Assessments** | | | | |
| Chemistry panel | X | X | X^ | |
| CBC with platelet count | X | X | X^ | |
| Blood Group* | X | | | |
| PT, PTT if known | X | X | | |
| Anticoagulation Monitoring (e.g., PTT/ Antifactor Xa level)** | X | X (site-specific) | | |
| D-dimer*** | X | X | X^ | |
| Troponin**** | X | X | X^ | |
| Coagulation and inflammatory markers***** | X | X | X^ | |
| Optional Biorepository | X | X | | |

*Blood group taken from hospital record or self report if that is not available.

** Frequency and mode (Anti-factor Xa/aPTT) of testing will be based on site routine. Anti-factor Xa monitoring is preferred over PTT

***Baseline D-dimer is required with SOC labs (sample needs to be obtained prior to randomization, but results do not need to be available at the time of randomization). All values collected should be recorded

****Strongly recommended as part of routine care

***** Optional, listed in case report form

+Assessments indicated in the table above will be ascertained at discharge, or at 28 days, whichever comes first. Participants must be followed for vital status until discharged from the hospital or another care facility (if transferred on organ support) up to 90 days. To maximize retention, participants will be contacted intermittently (e.g. at one and two months post-discharge)

++Participants will be contacted to ascertain vital status.functional status and quality of life. (Instruments detailed in the manual of operations)

^ May be collected at hospital discharge and at 28 days in participants who remain in hospital at that time

#### *A3.4 Potential Risks & Benefits*

##### A3.4.1 Known Potential Risks

Participants are monitored as per standard of care to minimize the risk of bleeding or developing clots. The therapeutic dose anticoagulation group will receive potent anticoagulation and thus may be at higher risk of bleeding.

##### A3.4.2 Known Potential Benefits

A recent study from patients with COVID-19 hospitalized in China found that patients with elevated D-dimer had the benefit of prophylactic dose anticoagulation versus no anticoagulation. Thus, there is a direct benefit of decreased clotting events in both arms – although this trial hypothesizes that the benefit in terms of decreasing adverse events will be superior in this therapeutic dose anticoagulation. All participants will be closely monitored by the study team and any changes will be discussed with the treating physicians and/or clinical team. There is a potential direct benefit of identifying clots or bleeding more rapidly based on this monitoring. This trial will contribute to the body of generalizable knowledge about the best anticoagulation strategy to use to minimize the risk of clotting in patients with COVID-19.

**A3.5 *Study Enrollment***

##### A3.5.1 Inclusion Criteria

Same as the Master Protocol.

##### A3.5.2 Exclusion Criteria

Exclusion criteria are as follows:

- Contraindication to anticoagulation, including but not limited to:
    - known bleeding within the last 30 days requiring emergency room presentation or hospitalization

    - known history of an inherited or active acquired bleeding disorder
    - known history of heparin induced thrombocytopenia
    - recent ischemic stroke
- Indication for therapeutic anticoagulation in the case that it cannot be stopped
- Platelet count < 50x $10^9$/L
- Hemoglobin < 8 g/dL
- Pregnancy
- Patient on dual antiplatelet therapy, when one of the agents cannot be stopped safely

##### A3.6 Event Adjudication

A subset of thrombotic events will be centrally adjudicated, with the proportion adjusted as needed based on agreement between the site and the event committee.

**A3.7 *Safety Analyses***

The safety event of importance for the therapeutic dose anticoagulation is major bleeding. The rates of ISTH major bleeding, ICH and fatal bleeds, and mortality will be monitored. The rates of bleeding will be directly compared to the control arm (prophylactic dose anticoagulation) as well as to any additional arms added to the platform trial subsequently if this arm continues. For ISTH major bleeding, ICH and fatal bleeds, and all cause mortality the DSMB will review the number of events, the event rates, and the posterior mean and 95% credible intervals for the event rates, difference between arms, and odds-ratios between arms will be summarized.

##### A3.8 *Statistical Analyses*

The therapeutic dose anticoagulation arm will be compared to the control arm (prophylactic dose anticoagulation) for efficacy on the primary analysis and the secondary endpoints. In addition, this arm will be compared to the control and may be stopped for futility if it does not improve the primary clinical outcome.

The primary Bayesian statistical model (see Master protocol) will be used for modeling this arm.

##### A3.8.1 Interim Analysis Schedule

Interim analyses will take place for this arm starting when 200 participants have been randomized to this arm and have 21-day follow-up. Analyses will continue with every 200 participants enrolled to this arm, at sample sizes of 400, 600, 800, and 1,000.

##### A3.9 *Adaptive Decision Rules*

At each analysis the following rules will be carried out:

1. **Superiority**: Within each subtype, if the posterior probability of superiority of the therapeutic dose anticoagulation arm compared to the prophylactic dose (control) anticoagulation arm is greater than 0.99, then the therapeutic dose anticoagulation arm will be declared superior to prophylactic dose anticoagulation in that subtype. If the prophylactic dose anticoagulation arm has not been discontinued in that disease subtype, then this result trigger should discontinue randomization within that stratum to the prophylactic dose anticoagulation arm.

If the therapeutic dose anticoagulation arm reaches 1,000 total participants enrolled without a declaration of superiority or futility within a subtype, this arm may be discontinued within these remaining subtypes. If the steering committee decides it is important to keep this arm in the platform trial, it may expand beyond this 1,000 participants randomized and this plan will be amended.

##### A3.10 *Operating Characteristics*

See Arm-specific SAP for the operating characteristics for the therapeutic dose anticoagulation arm.

### Appendix 4: Prophylactic Dose Anticoagulation (Arm B)

Any of the following strategies are recommended for prophylactic dose anticoagulation:

#### A4.1 *Prophylactic Dose Anticoagulation**

| CrCl | BMI | Enoxaparin | Dalteparin | Tinzaparin | Fondaparinux | Heparin |
|---|---|---|---|---|---|---|
| ≥30 | <40 | 40 mg SC q24h | 5000 units SC q24h | 4500 units SC q24h | 2.5 mg SC q24h | 5000 units SC q8-12h |
| | ≥40 | 40 mg SC q12h | 5000 units SC q12h | 9000 units SC q24h | NA | 7500 units SC q8h |
| <30 | <40 | Heparin 5000 units SC q8-12h | | | | |
| | ≥40 | Heparin 7500 units SC q8h | | | | |

*All drugs are considered standard of care as an anticoagulant (1–2). Different drugs are used in different regions, countries, and hospital formularies. As a pragmatic trial of antithrombotic therapy in COVID-19, sites will use the anticoagulant that they typically use in the hospital setting.

It is recommended that participants be given prophylactic-dose parenteral anticoagulation daily for at least 14 days or until hospital discharge, whichever comes first. Treatment may continue beyond 14 days at the discretion of the most responsible physician.

The following measures are recommended during CRRT: regional citrate, heparin priming and prophylactic dose heparin administration (without measurable systemic anticoagulation)

**Full therapeutic dose anticoagulation (therapeutic dose UFH or LMWH) is permitted as rescue therapy in the event of suspected or confirmed deep vein thrombosis, pulmonary embolism, systemic arterial thromboembolism, acute coronary syndrome, circuit or line thrombosis during continuous renal replacement therapy (despite regional measures) or sustained low-efficiency daily dialysis. Full therapeutic dose anticoagulation is also acceptable for intermittent hemodialysis. These are adherent to protocol.** Full therapeutic anticoagulation is not recommended solely for clinical deterioration involving transfer to ICU-level care in this setting in the absence of suspected PE. If the team changes to therapeutic dose for other reasons (e.g., increasing D-dimer; team is not comfortable with prophylactic dose, minor increase in oxygen support), then this is not adherent to protocol and site PI will need to discuss this with the clinical team.

#### A4.2 *Discontinuation of study intervention*

Anticoagulation should be discontinued if there is clinical bleeding or another complication sufficient to warrant cessation in the opinion of the treating clinician. Major bleeding, including death due to bleeding, is an SAE. Assigned treatment may be resumed if deemed appropriate by the treating clinician.

Occurrence of HIT must result in the cessation of UFH or LMWH without recommencement regardless of treatment assignment. Use of an acceptable alternative agent is required in this instance as clinically indicated. Occurrence of HIT is an SAE.

Study interventions can be discontinued at any time by the treating clinician if doing so is regarded as being in the best interests of the patient. Temporary cessation – for the shortest period of time possible, but not longer than 24 hours – such as to allow surgical or other procedures is not a protocol deviation.

Temporary or permanent cessation of study intervention for bleeding is not a protocol deviation.

**A4.3 *Study Schedule***

| **Activity** | **Screening/ Enrollment** | **Hospital Duration** | **28 days and/or hospital discharge⁺** | **90 days post randomization⁺⁺** |
|---|---|---|---|---|
| **Eligibility** | | | | |
| Consent | X | | | |
| Demographic and Medical History | X | | | |
| Assessment of Inclusion/Exclusion criteria | X | | | |
| Self-reported race/ethnicity and sex | X | | | |
| Pregnancy Test, for women of childbearing potential | X | | | |
| **Study Drug Administration** | | | | |
| Randomization | X | | | |
| Study treatment | X | X | | |
| **Study Procedures** | | | | |
| Height | X | | | |
| Weight | X | | | |
| Vital signs | X | X | | |
| Concomitant medications | X | X | | |
| WHO ordinal assessment | X | X | X^ | X |
| Outcomes assessment | | X | X | X⁺⁺ |
| **SOC Laboratory Assessments** | | | | |
| Chemistry panel | X | X | X^ | |
| CBC with platelet count | X | X | X^ | |
| Blood Group* | X | | | |
| PT, PTT if known | X | X | | |
| Anticoagulation Monitoring (ex, PTT/ Antifactor Xa level)** | X | X (site-specific) | | |
| D-dimer*** | X | X | X^ | |
| Troponin**** | X | X | X^ | |
| Coagulation and inflammatory markers***** | X | X | X^ | |
| Optional Biorepository | X | X | | |

*Blood group taken from hospital record or self report if that is not available.

** Frequency and mode (Anti-factor Xa/PTT) of testing will be based on site routine. Anti-factor Xa monitoring is preferred over PTT

*** Baseline D-dimer is required with SOC labs (sample needs to be obtained prior to randomization, but results do not need to be available at the time of randomization). All values collected should be recorded.

****Strongly recommended as part of routine care, all values collected should be recorded

*****Optional, listed in case report form

+Assessments indicated in the table above will be ascertained at discharge, or at 28 days, whichever comes first. Participants must be followed for vital status until discharged from the hospital or another care facility (if transferred on organ support) up to 90 days. To maximize retention, participants will be contacted intermittently (e.g. at one and two months post-discharge)

++Participants will be contacted to ascertain vital status and functional status and quality of life. (Instruments detailed in the manual of operations)

^ May be collected at hospital discharge and at 28 days in participants who remain in hospital at that time

#### A4.4 *Potential Risks & Benefits*

##### A4.4.1 Known Potential Risks

Participants are monitored as per standard of care to minimize the risk of bleeding or developing clots.

##### A4.4.2 Known Potential Benefits

A recent study from patients with COVID-19 hospitalized in China found that patients with elevated D-dimer had a benefit of prophylactic dose anticoagulation versus no anticoagulation. Thus, there is a direct benefit of decreased clotting events in both arms – although this trial hypothesizes that the benefit in terms of decreasing adverse events will be superior in this therapeutic dose anticoagulation. All participants will be closely monitored by the study team and any changes will be discussed with the treating physicians and/or clinical team. There is a potential direct benefit of identifying clots or bleeding more rapidly based on this monitoring. This trial will contribute to the body of generalizable knowledge about the best anticoagulation strategy to use to minimize the risk of clotting in patients with COVID-19.

**A4.5 *Study Enrollment***

##### A4.5.1 Inclusion Criteria

Same as the Master Protocol.

##### A4.5.2 Exclusion Criteria

Exclusion criteria are as follows:

- Contraindication to anticoagulation, including but not limited to
  - known bleeding within the last 30 days requiring emergency room presentation or hospitalization
  - known history of a bleeding disorder of an inherited or active acquired bleeding

disorder
  - known history of heparin induced thrombocytopenia
  - recent ischemic stroke
- Indication for therapeutic anticoagulation in the case that it cannot be stopped safely
- Platelet count < 50x $10^9$/L
- Hemoglobin < 8 g/dL
- Pregnancy
- Patient on dual antiplatelet therapy when one of the agents cannot be stopped safely

#### A4.6 Event Adjudication

A subset of thrombotic events will be centrally adjudicated, with the proportion adjusted as needed based on agreement between the site and the event committee.

**A4.7 *Safety Analyses***

The safety event of importance for the prophylactic dose anticoagulation is serious thrombotic events. The risk is that with a sub therapeutic dose there may be elevated thrombotic events. The rates of serious thrombotic events and mortality will be monitored. The rates of serious thrombotic events will be directly compared to the therapeutic dose anticoagulation arm as well as to any additional arms added to the platform trial subsequently if this arm continues. For serious thrombotic events the DSMB will review the number of events, the event rates, and the posterior mean and 95% credible intervals for the event rates, difference between arms, and odds-ratios between arms will be summarized.

#### A4.8 *Statistical Analyses*

The prophylactic anticoagulation arm is intended as the initial control arm in the platform trial. This arm will be the referent arm in the Bayesian statistical model. This arm will not have any efficacy or futility stopping rules. This arm will function as a comparator within each of the subtypes and will continue in each until an arm demonstrates efficacy compared to this arm and it is discontinued in that subtype. This prophylactic dose anticoagulation arm is likely to be the safest of the arms in this platform trial and hence arms will have a need to demonstrate statistical superiority to this arm for it to be discontinued.

#### A4.9 Number of Participants

Approximately 1,000 participants will be randomized to this arm and have 90-day follow-up.

### Appendix 5: ACTIV-4 Blood Sampling – proposed samples and times for sites participating in mechanistic studies and biorepository

The goal of the Mechanistic Studies Center and the Biorepository/Central Lab is to add significant value to the clinical trials by collecting high-quality blood samples for studies aimed at elucidating underlying disease mechanisms and insights into how the therapy modifies these underlying disease processes. A goal is to identify biomarkers that can identify pathological mechanisms, predict outcomes, direct therapy, and/or identify higher-risk patient subpopulations.

**A5.1 *Inpatient sampling***

##### *Blood collection times for inpatients:*

- Days 1 (time of enrollment), 3, 7 and 14
- Samples should be coordinated with clinical lab blood draws when possible.

##### *Standard samples to be collected & volumes at each time point:*

- Citrate plasma
  - Two 4.5 mL Citrate tubes (BD # 369714)
- EDTA plasma
  - One 10 mL EDTA tube (BD# 366643)
- Serum
  - One 5.0 mL Serum tube (BD # 367814)

***Note 1:*** *We anticipate that some sites may not be able to collect & process all the samples and time points listed above. We plan to work with those sites to identify more limited time points and/or discard samples that could be collected, processed and sent to the biorepository.*

***Note 2:*** *We anticipate that some high-functioning sites may, in addition to the sample collections noted above, also participate in enhanced collections & studies, which may include:*

- Additional blood collection tubes such as:
  - HTI SCAT-144 plasma
  - Paxgene RNA whole blood
  - Cell Prep Tube (CPT)
- Whole blood assays:
  - Viscoelastic assays (thromboelastography or thromboelastometry)
  - Platelet aggregometry
  - Whole blood genomics

**A5.2 *Sample processing***

A detailed Manual of Operations (MOP) will provide instructions to clinical lab and research personnel regarding sample processing including centrifugation, processing, freezing, storing, & shipping samples. Also, the following will be provided: training materials; sample processing kits with prelabeled transport and/or storage vials; sample tracking software; shipping materials.

#### A5.3 *Biorepository/Central Lab*

The Biorepository will archive biosamples from the clinical sites, and distribute them to the labs doing ACTIV-4 approved mechanistic studies and other research. If ACTIV-4 biosamples cannot be shipped to the Biorepository for some reason, the information will be captured and used to form a "Virtual Biorepository", so that those samples can contribute to the mechanistic studies as well.

#### Appendix 6. Additional data inclusion from other trials merged under activ-4 platform

There are several clinical trials that have been testing safety and efficacy of Arm A and B regiments. Data collected in these trials will be included in the data analysis under this protocol provided that the subjects consented for the data to be shared or a waiver of consent and authorization had been granted by the reviewing IRB. The data will be labeled with subject ID and only include dates which are necessary to assess safety and efficacy endpoint events. All other private health information (PHI) will be removed. The data will be stored at the study coordinating center, University of Pittsburg, in HIPAA compliant electronic system and only coordinating center staff will have access to the data. The statistical analysis plan will account for this additional data.

### SECONDARY ENDPOINT ADJUDICATION: ANTICOAGULATION AND ANTIPLATELET DOMAINS

1 Introduction ... 482
2 Interaction with ATTACC and ACTIV-IV ... 482
3 Endpoints for adjudication ... 482
4 Secondary endpoint definitions ... 483
4.1 Domain-specific secondary endpoints ... 483
4.1.1 Major bleeding ... 483
4.1.2 Acute Myocardial Infarction (AMI) ... 483
4.1.3 Confirmed deep vein thrombosis ... 484
4.1.4 Confirmed pulmonary embolus ... 484
4.1.5 Confirmed ischemic cerebrovascular event ... 484
4.1.6 Mesenteric ischemia ... 484
4.1.7 Limb ischemia ... 485
4.2 Derived endpoints ... 485
4.2.1 Systemic arterial thrombosis or embolism ... 486
4.2.2 Major thrombotic event ... 486
4.2.3 All thrombotic events ... 486
4.3 Heparin induced thrombocytopenia (HIT) ... 485
5 Data items to be used for adjudication ... 486
6 Adjudication process ... 486
6.1 Data extraction ... 486
6.2 Blinded Adjudication ... 487
6.3 Additional data collection ... 487
6.4 Resolution of discordant adjudication assessments ... 488

#### Introduction

This document outlines the process for adjudication of key secondary domain-specific endpoints for the Anticoagulation, Antiplatelet and Immunoglobulin domains. This includes the methodology and adjudication criteria to be used specifically for major bleeding, thrombosis outcomes and heparin-induced thrombocytopenia (HIT).

#### Interaction with ATTACC and ACTIV-IV

Both ATTACC and ACTIV-IV are platform trials also evaluating the treatment effect of therapeutic anticoagulation in patients with COVID-19. The REMAP-CAP Anticoagulation domain, ATTACC and ACTIV-IV, have been designed to be complementary with pre-specified plans in relation to methods of analysis. It is intended that data from this domain, ATTACC and ACTIV-IV will be incorporated into a statistical mode that is separate from the pandemic statistical model of REMAP-CAP. The protocol, governance, and data management of ATTACC and ACTIV-IV are separate from REMAP-CAP. The REMAP-CAP DSMB will have overlapping membership with the DSMB for the ATTACC trial; ACTIV-IV functions with a separate independent DSMB. The adjudication process for secondary endpoints in REMAP-CAP is separate to ATTACC and ACTIV-IV, but has been designed to align with these platform trials. The outcome measures and definitions were aligned across all three platform trials.

#### Endpoints for adjudication

Secondary domain-specific endpoints to be adjudicated are:

- Major bleeding
- Confirmed deep venous thrombosis
- Confirmed pulmonary embolism
- Confirmed ischemic cerebrovascular event
- Acute myocardial infarction
- Other thrombotic events, including mesenteric ischemia and limb ischemia
- Major thrombotic events
- Heparin induced thrombocytopenia (HIT)

These endpoints may be collected within more than one domain, including the Anticoagulation, Antiplatelet and Immunoglobulin domains.

#### Secondary endpoint definitions

The following are the definitions for the outcomes reported by site investigators, as outlined in the Domain Specific Appendix (DSA) and/or the Case Report Form Data Completion Guide.

##### Domain-specific secondary endpoints

###### Major bleeding

Fatal bleeding, symptomatic or clinically manifest bleeding in a critical are or organ (such as intracranial, intraspinal, intraocular, retroperitoneal, intraarticular, pericardial, or intramuscular with compartment syndrome), OR
Blood loss above 300mls or bleeding causing a fall in haemoglobin of ≥2g/dL (20g/L, 1.24mmol/L), or leading to the transfusion of 2 or more whole blood or red cell units

###### Acute Myocardial Infarction (AMI)

The definition of an AMI requires detection of rise and fall or just a fall of cardiac biomarkers, such as any form of troponin assay, with at least one value above the upper reference limit (URL) PLUS evidence of myocardial ischemia with at least one of the following:

- Symptoms of cardiac ischemia
- ECG changes indicative of new ischemia (new ST-T changes or new LBBB)*
- Development of pathological Q waves in the ECG**
- Imaging evidence of new loss of viable myocardium or new regional wall motion abnormality

*ECG manifestation of acute myocardial ischemia (in the absence of LVH and LBBB):

- ST Elevation - New ST elevation at the J-point in two contiguous leads with the cut-off points of ≥ 0.2 mV in men or ≥ 0.15 mV in women in leads V2-V3 and/or ≥ 0.1 mV in other leads.
- ST depression and T-wave changes – New horizontal or down-sloping ST depression ≥ 0.05 mV in two contiguous leads; and/or T inversion ≥ 0.1 mV in two contiguous leads with prominent R waves or R/S ratio >1.

**Pathological Q waves:

- Any Q-wave in leads V2-V3 ≥ 0.02 seconds or QS complex in leads V2 and V3

- Q-wave ≥ 0.03 seconds and ≥ 0.1 mV deep or QS complex in leads I, II, aVL, aVF, or V4-V6 an any two leads of a contiguous lead grouping (I, aVL, V6; V4-V6; II, III, aVF; V7-V9).
- R-wave ≥ 0.04 s in V1–V2 and R/S ≥ 1 with a concordant positive T-wave in the absence of a conduction defect

##### Confirmed deep vein thrombosis

Proximal deep vein thrombosis is a thrombus located in axillary vein or more proximal, including the internal jugular vein, and a thrombus located in popliteal vein or more proximal. Confirmation requires imaging with techniques that include ultrasound or CT scan.

##### Confirmed pulmonary embolus

Segmental or multi-sub-segmental pulmonary emboli that is confirmed using CT pulmonary angiography or has a high probability ventilation: perfusion lung scan

##### Confirmed ischemic cerebrovascular event

An acute ischemic stroke is defined as central nervous system infarction defined as brain, spinal cord, or retinal cell death attributable to ischemia, based on:

- Pathological, imaging, or other objective evidence of cerebral, spinal cord, or retinal focal ischemic injury in a defined vascular distribution; OR
- Clinical evidence of cerebral, spinal cord, or retinal focal ischemic injury based on symptoms persisting ≥24 hours or until death, and other etiologies excluded. (Note: CNS infarction includes types I and II hemorrhagic infarctions) OR
- Infarction or hemorrhage in the brain, spinal cord, or retina because of thrombosis of a cerebral venous structure. Symptoms or signs caused by reversible oedema without infarction or hemorrhage do not qualify as stroke.

##### Mesenteric ischemia

Mesenteric Ischemia for arterial or venous mesenteric ischemia diagnosed on contrast imaging by CT or angiography or diagnosed at laparotomy or via laparoscopy.

#### Limb ischemia

Limb ischemia if evidence of acute limb ischemia sufficient to require surgical revascularization including bypass procedure, intraarterial thrombolysis, or embolectomy; amputation of a limb due to acute ischemia; or decision to withdraw or limit treatment because of acute limb ischemia. It is not sufficient for there to be evidence of limb ischemia that does not result in surgical intervention or determine a decision to institute palliative care. Ischemia attributed to vasopressor medication is insufficient unless also meets the above definition.

#### Heparin induced thrombocytopenia (HIT)

*Definite HIT:*

- Positive Serotonin Release Assay (SRA) or equivalent functional HIT confirmatory test
- Positive Enzyme-linked immunosorbent assay (ELISA) AND treated as HIT
- Positive quantitative rapid immunoassay (RIA) AND treated as HIT

*Possible HIT:*

- Positive ELISA or quantitative RIA AND any heparin/LMWH anticoagulation stopped
- 4T score >3 (high or intermediate), no Laboratory testing for HIT done, AND treated as HIT

*No HIT:*

- Negative HIT ELISA, or Negative Quantitative RIA
- Negative SRA, or Negative equivalent functional confirmatory assay
- No laboratory testing for HIT done AND not treated as HIT (heparin continued or discontinued for other reason and uneventful course - defined as absence of thrombotic event and platelet count recovery during follow-up).

#### Derived endpoints

The following are the definitions for the endpoints outlined in the Statistical Analysis Plan for the Anticoagulation Domain which are derived from the domain-specific secondary endpoints:

##### Systemic arterial thrombosis or embolism

Clinical evidence of sudden significant worsening of organ or limb perfusion and either confirmation of arterial obstruction (e.g. by imaging, hemodynamics, intraoperative findings or pathology evaluation) or requirement for intervention (thrombolysis, thrombectomy or urgent bypass).

##### Major thrombotic event

A composite dichotomous endpoint of pulmonary embolism, ischemic cerebrovascular event, myocardial infarction, or systemic arterial thromboembolism diagnosed at any time during the index hospitalization

##### All thrombotic events

A composite dichotomous endpoint of deep vein thrombosis, pulmonary embolism, ischemic cerebrovascular event, myocardial infarction, or systemic arterial thromboembolism diagnosed at any time during the index hospitalization or death in hospital

#### Data items to be used for adjudication

Data are collected and recorded in the REMAP-CAP eCRF, with definitions of secondary endpoints provided in the CRF Data Completion Guide. In addition, these endpoints may be reported as a Serious Adverse Events (SAE) as outlined in the Core Protocol and DSA. The data items extracted and made available for adjudication are:

- Individual data items for occurrence of each of the specified secondary outcomes
- Free text fields used to describe Major Bleeding
- Free text fields used to describe “Other thromboembolic events”
- SAE report fields, including free text fields

#### Adjudication process

##### Data extraction

Patients that have a report of any of the specified secondary endpoints above, have data entered into free text field of Major Bleeding or Other Thromboembolic event field or have one or more SAEs reported are identified by an unblinded data manager. The data item fields outlined above are extracted for these patients, along with a newly generated unique patient identifier. The data extraction does not include any identifying information (site or participant) or information on treatment allocation. The unblinded data manager will screen the SAE text boxes and where feasible, references to treatment allocation in free text fields will be removed prior to adjudication. This deidentified data extract is then provided to the blinded adjudicators.

#### Blinded Adjudication

Individual patient data will be independently reviewed by two adjudicators with relevant clinical haemostasis/thrombosis expertise and clinical trial experience who are blinded to treatment allocation. Adjudicators review the data and determine:

1. Major Bleeding – review free text and SAE reports to determine if the description meets the definition as outlined above: – Yes, No or More information required
2. Other thromboembolic events – confirms the description meets the definition of a thromboembolic event: - Yes, No or More information required
3. Other thromboembolic event – determines if the thrombosis is Arterial, Venous or More information required
4. Other thromboembolic event – determine if the event meets definition of Systemic Arterial Thromboembolic event: Yes, No or More information required
5. SAE reports – does the described event meet the definition of any of the specified secondary endpoints: Yes, No or More information required
6. SAE reports – if the report meets a secondary endpoint definition, and has not been reported, then classify according to the definitions above
7. Major thrombotic event – Yes, No or More information required

#### Additional data collection

Where insufficient data is available for one or both adjudicators to complete these fields, additional information will be requested from sites using source documents. Once the additional information has been provided to REMAP-CAP, updated data will be returned to the adjudicators to repeat the process as outlined above.

#### Resolution of discordant adjudication assessments

For outcomes where there is a discordant result between the two blinded adjudicators, a third reviewer will independently review the endpoint. Once the third adjudication is complete, the three adjudicators will meet to determine a consensus decision for the endpoint.

### Clinical Events Committee (CEC) Charter

*ANTITHROMBOTIC THERAPY TO AMELIORATE COMPLICATIONS OF COVID-19 (ATTACC trial)*

CONFIDENTIAL

Version 1.1

Effective Date: January 7, 2021

Last Updated Date: January 7, 2021

### CEC CHARTER APPROVAL DOCUMENTATION

**Study** ANTITHROMBOTIC THERAPY TO AMELIORATE COMPLICATIONS OF COVID-19 (ATTACC)

**Version: 1.0** **Last Updated: December 28th 2020**

The signatures below confirm that the signees have reviewed the aforementioned Charter thoroughly, and its content is acceptable in quality and form for the purpose of this study.

### REQUIRED SIGNATURES

**CEC Chair**

| Print Name | Signature | DDMMMYYYY |
|---|---|---|
| Brendan Everett, MD, MPH | | 7-Jan-2021 |

**Co-Principal Investigator, on behalf of Trial Principal Investigators**

| Print Name | Signature | DDMMMYYYY |
|---|---|---|
| Patrick Lawler, MD, MPH | | January 7th, 2020 |

#### CEC CHARTER DOCUMENT MANAGEMENT

| Version | Author(S) | Date | Version Description |
|---|---|---|---|
| 1.0 | PRL, BE, BAK | December 28th, 2020 | First charter version developed in collaboration with CanVECTOR and Brigham and Women's adjudication center, aligning with definitions in ACTIV-4a |

#### ABBREVIATIONS

| | |
|---|---|
| AE | Adverse Event |
| AMI | Acute Myocardial Infarction |
| BARC | Bleeding Academic Research Consortium |
| CABG | Coronary artery bypass grafting |
| CAPA | Corrective and preventive actions |
| CEC | Clinical Event Committee |
| CRO | Contract Research Organization |
| CV | Cardiovascular |
| ECG | Electrocardiogram |
| eCRF | electronic Case Report Form |
| eTMF | electronic Trial Master File |
| GCP | Good Clinical Practice |
| IEC/IRB | Independent Ethics Committee / Institutional Review Board |
| MI | Myocardial Infarction |
| LVOT | Left ventricular outflow tract |
| URL | Upper Reference Limit |
| NSTEMI | Non-ST-elevation myocardial infarction |
| NT-proBNP | N-terminal pro-B-type natriuretic peptide |
| PCI | Percutaneous coronary intervention |
| PHI | Protected health information |
| PI | Principal Investigator |
| QC | Quality Control |

| | |
|---|---|
| SAE | Serious Adverse Event |
| SOP | Standard Operating Procedure |
| STEMI | ST-elevation myocardial infarction |
| TC | Teleconference |
| ULN | Upper limit of normal |
| VF | Ventricular fibrillation |
| VT | Ventricular tachycardia (VT) |

#### 1 INTRODUCTION

The Clinical Event Committee (CEC) will be responsible to for the conduct of the endpoint adjudication operations for the study in collaboration with SOCAR Research, Avanti, and Ozmosis. This Charter will set out the methodology and adjudication criteria to be used when adjudicating the following secondary outcomes and adverse events occurring up to 90 days following randomization.

a. Secondary efficacy endpoints:
   - Venous thromboembolic events (VTE), including deep venous thrombosis and pulmonary embolism
   - Acute myocardial infarction (MI)
   - Stroke
   - Systemic arterial thrombosis or embolism

b. Safety averse events (all events submitted adjudicated; sites instructed to submit adverse events occurring during the treatment window, which is up to 14 days following randomization):
   - Major bleeding
   - Heparin induced thrombocytopenia (HIT) Adjudicators will

remain blinded to the study treatment assignment.

#### 2 CHARTER AMENDMENTS

This Charter will be reviewed and formally approved before event adjudication commences.

The Charter may require revision during the study for various reasons. This determination will be made by the CEC Chairpersons together with the three trial Principal Investigators (PIs).

#### 3 PROTOCOL OVERVIEW

The ATTACC trial is a prospective, open-label, randomized, multicentre, adaptive-design clinical trial examining the efficacy of full dose heparin compared with usual care at reducing the need for organ support or death in patients hospitalized with COVID-19.

##### 3.1 Primary efficacy endpoint

The primary outcome is days alive and free of organ support at day 21. The primary outcome will not be adjudicated by the CEC.

##### 3.2 Secondary efficacy endpoint

The full list of secondary endpoints is available in the trial protocol. Among these, the following secondary efficacy endpoints will be adjudicated by the CEC:

- Venous thromboembolism (DVT or PE) assessed at 28 and 90 days following randomization
- Myocardial infarction assessed at 28 and 90 days following randomization
- Ischemic stroke assessed at 28 and 90 days following randomization
- Systemic arterial thrombosis or embolism assessed at 28 and 90 days following randomization

The following secondary safety events will be adjudicated by the CEC (sites instructed to report events occurring during the intervention window as defined in the protocol):

- Laboratory confirmed HIT
- Major bleeding, defined according to the International Society on Thrombosis and Haemostasis (ISTH)/Scientific and Standardization Committee (SSC) definitions and bleeding assessment tool in non-surgical patients (Schulman *J Thromb Haemost* 2005).

#### 4 PROJECT TIMELINE

The study has established a high-level timeline that will span an estimated 6 months for all the activities to meet the goals outlined in this Charter, although this timeline may be extended pending study progress and the pandemic's evolution.

#### 5 CEC ORGANIZATION

All events adjudicated will be under the supervision of the CEC chair, presiding over the CEC. Safety event adjudication will be supported by CanVECTOR members who are specialists in thrombosis and anticoagulant complications, including HIT and major bleeding. Secondary efficacy endpoints related to venous thrombosis will also be adjudicated by members of CanVECTOR, under the guidance of the CEC chair. Arterial thrombotic events will be adjudicated by cardiologists (MI) and neurologists (stroke) serving on the CEC. The CEC will receive logistical support from the SOCAR CEC Coordination Team aswell as Ozmosis and Avanti. Source documents in non-English, non-French languages will be translated; Avanti will oversee the translation of Portuguese and Spanish source documents which will be completed by a third party contracted to Avanti. Avanti will be responsible for uploading both original and translated source documents to the *e*SOCDAT™ web-based adjudication module.

#### 6 CEC OVERVIEW

SOCAR Research (SOCAR) will be responsible for CEC operations and logistics management and oversight. The SOCAR team will create and maintain the CEC Charter in close collaboration with the CEC chair, Steering Committee and the CEC adjudicators. Key responsibilities of the SOCAR CEC Coordination Team will be:

- Support drafting of the CEC Charter and managing its revisions
- Develop the adjudication eCRF forms in collaboration with the CEC and Steering Committee members
- Configure the eSOCDAT adjudication module to the requirements of the ATTACC adjudication
- Coordinate event adjudication
- Train independent CEC adjudicators

Events will be adjudicated using the *e*SOCDAT™ web-based adjudication module. The events will be adjudicated remotely, and independently by adjudicators who are not otherwise operationally involved in the ATTACC trial.

### 7 ROLES AND RESPONSIBILITIES

#### 7.1 CEC Chairperson

The CEC Chairperson will oversee all aspects of this CEC project in conjunction with the SOCAR CEC Coordination Team. Additional responsibilities of the CEC Chairperson may include, but are not limited to the following:

- Assist in selection of CEC adjudicators
- Preside over CEC adjudication conference calls and meetings
- Finalize endpoint criteria definitions and adjudication criteria in consultation with the study Steering Committee
- Approve the CEC Charter.
- Suggest/approve any Charter amendments that may be necessary during the course of the study

#### 7.2 CEC Adjudicators

CEC adjudicators are board certified or board eligible physicians who are not otherwise operationally involved in the trial. The CEC adjudicators will not be involved in participant recruitment or follow-up, be a member on the Data Monitoring Committee, or be involved with the safety reporting or study monitoring. The CEC adjudicators will review all events that are to be adjudicated as they become available and adjudicate the events in a consistent and unbiased manner according to event definitions in this Charter. Source documents will be redacted to both patient identifiers and randomized treatment allocation (to the most complete extent reasonably feasible), so that adjudicators may remain blinded to study treatment assignment throughout the adjudication process and for the duration of the study. Further responsibilities of the CEC adjudicators may include, but are not limited, to the following:

- Contribute toward the development of the CEC processes and provide input to the development of this charter and study adjudication forms and definitions
- Review clinical Event Dossier and complete event adjudication forms
- Attend training sessions, and disagreement meetings as required

##### 7.2.1 CEC Adjudicator Qualifications

CEC adjudicator qualifications will include clinical expertise in the field of cardiovascular medicine, neurology, thrombosis, hematology, internal medicine, and bleeding, as well as knowledge of clinical study methodology and CEC processes, and commitment to study timelines. The members must disclose all financial relationships with organizations or institutions at the beginning of the study and annually, collected by the CEC coordinating office staff. The financial disclosures will be reviewed by the CEC Chairperson and if any disclose a potential conflict, a review with SOCAR and the study Steering Committee will take place and decision made regarding continued membership on the committee.

The SOCAR Coordination Team will maintain documentation of CEC members' qualifications up-to-date, including at least a current Curriculum Vitae which shall be made available for audits and/or inspections.

##### 7.2.2 CEC Adjudicator Membership

CEC adjudicator membership will be for the duration of the study. Although each CEC adjudicator will be expected to serve for the duration, in the event a CEC adjudicator is unable to fulfill his/her responsibilities, the reason(s) will be documented, and a replacement will be proposed by the CEC Chairpersons.

### *8 CEC Adjudicator Training*

All CEC adjudicators will receive training on the study protocol, CEC processes, eSOCDAT™ web-based adjudication module and event definitions. Proof of training will be documented and archived in TMF.

#### 8.1 CEC Adjudicator Confidentiality

The CEC adjudicators will have an obligation to maintain the confidentiality of all study information. The CEC members must take appropriate measures to ensure that confidentiality will be maintained.

#### 8.2 SOCAR CEC Coordination Team

The SOCAR CEC Coordination Team will consist of the SOCAR CEC Project Manager and the SCCR CEC Coordinator(s).

#### 8.3 Disbandment of CEC

The Endpoint Adjudication Committee will disband 6 months after the final database closure of the clinical study.

### 9 CEC OPERATIONS

The flow of data from event reporting at site until adjudication outcome is summarized in the sections hereafter.

#### 9.1 Event Identification

##### 9.1.1 Primary Method

The primary method for event identification in this study will be reporting by Site Investigators of suspected events on the eCRF event forms. The following will be reported:

- Bleeding and HIT: mandatory reporting as adverse event (AE) considered secondary safety endpoint. Bleeding-related AEs not reported by sites as meeting criteria for ISTH major bleeding may be adjudicated to assure that they do not meet ISTH criteria, as judged by information provided to the data coordinating centre. HIT is ascertained by site reporting similarly as an AE, and sites have been instructed to only report laboratory-confirmed HIT. Sites have been instructed only to report events during the active treatment period, which lasts up to 14 days, or until recovery (defined as hospital discharge or liberation from supplemental oxygen, provided it was originally required, for >24 hours).
- Arterial thrombotic events: all events with onset on or after randomization and up to 90 days after randomization.
- Venous thrombotic events: all events with onset on or after randomization and up to 90 days after randomization.

The procedures / process to collect information for an event suggestive of any of the events just cited above is depicted in **Figure 1**.

Figure 1: PROCESS FOR THE COLLECTION OF SOURCE DOCUMENTATION FOR EVENTS REQUIRING ADJUDICATION

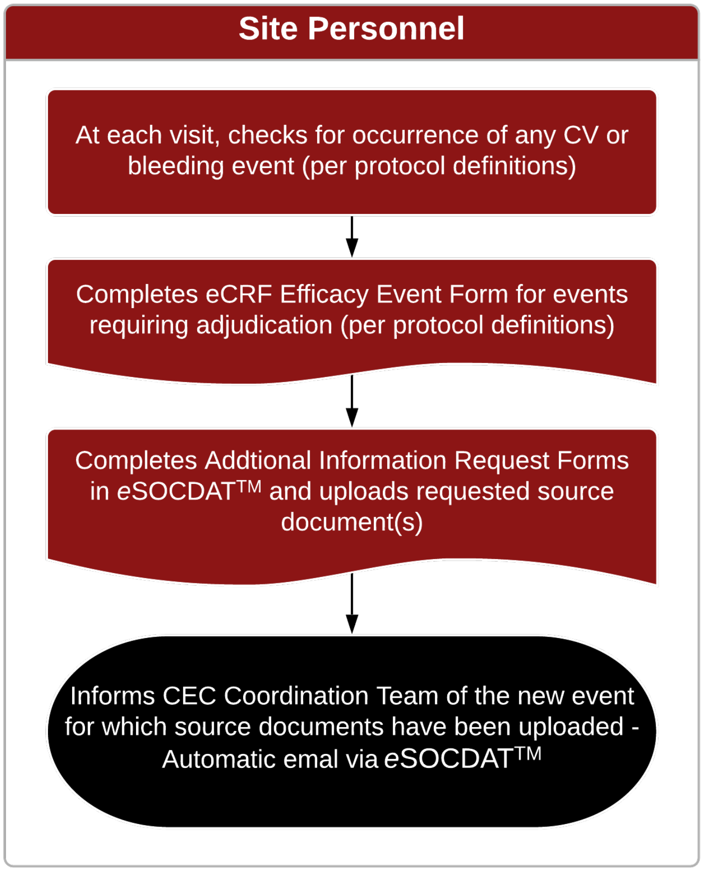

##### 9.1.2 Supplementary Methods

Some potential events may also be reported in the narratives received but not reported in the eCRF; SOCAR will query and follow-up with sites to ensure that the Event eCRFs are appropriately completed.

#### 9.2 Multiple Events occurring in The Same Subject

Multiple events in the same subject will be adjudicated as separate events. However, multiple events occurring during the same hospitalization will be considered a single case. They will be assigned the same case number and will be adjudicated together. For each hospitalization, a distinct case number will be assigned.

#### 9.3 Redaction

Sites will be requested to ensure appropriate redaction of personal identifiers (PHI) source documents provided to the CEC.

If CEC Coordination Team sees information during case preparation that should be redacted but is not, then the CEC Coordination Team will redact the information but also send a note to Ozmosis or Avanti (as appropriate) about the site failure to redact the information in case additional site training is needed. In the instance that any of the CEC adjudicators see any information during event adjudication that should have been redacted but is not, they will notify the CEC Coordination Team about the situation. The CEC staff will recall the event packet from the other reviewer, redact the information and resubmit to an adjudicator for adjudication. A note will also be sent to the Ozmosis or Avanti teams concerning the site failure to redact the information in case additional site training is needed.

Randomized treatment allocation will be redacted to the most complete extent possible. The trial is open label, and the investigators recognize that complete redaction may, in some cases, not be possible. To preserve blinding of the investigators, as noted above, CEC members are not permitted to hold roles in directing trial operations – so that in the event that blinding is lost and adjudicators inadvertently become aware of potential treatment effects, trial operations are not influenced.

#### 9.4 CEC Coordination Team Review

The CEC Coordinator(s) will log into the *e*SOCDAT™ and review all the information and documents associated with the event as established in the approved Charter and depicted in **Figure** .

Where there is insufficient information (i.e., missing documents or clarifications needed), the CEC Coordinator will post a query for the site to provide the missing information. For further details on the query management process please refer to Section 9.5 and **Figure 2**.

Once the CEC Coordinator considers that there is sufficient information to progress the event, he/she will complete the applicable form in *e*SOCDAT™ to initiate the adjudication process.

Where the source documents require translation (in Spanish and Portuguese), Avanti will oversee source document translation be qualified medial translators prior to document upload. The CEC Coordinator will then upload the translated source documents once translation is complete.

The CEC Coordinator will also ensure additional documentation including lab reports and patient profiles have been uploaded by the site.

#### 9.5 Dossier Creation and Event Assignment

Once the CEC Coordinator agrees that there is sufficient information to progress the event for adjudication, he/she will confirm the completion of the Event Dossier in eSOCDAT™. The Event Dossier will include all submitted source documents and notes relevant for adjudication. eSOCDAT™ will then randomly assign the event to the CEC adjudicators as per the process outlined in Section 9.7. When multiple events are being adjudicated at the same time, eSOCDAT™ will distribute the workload evenly among the CEC adjudicators.

Figure 2: COLLECTION AND MANAGEMENT OF SOURCE DOCUMENTS TO COMPILE EVENT DOSSIER

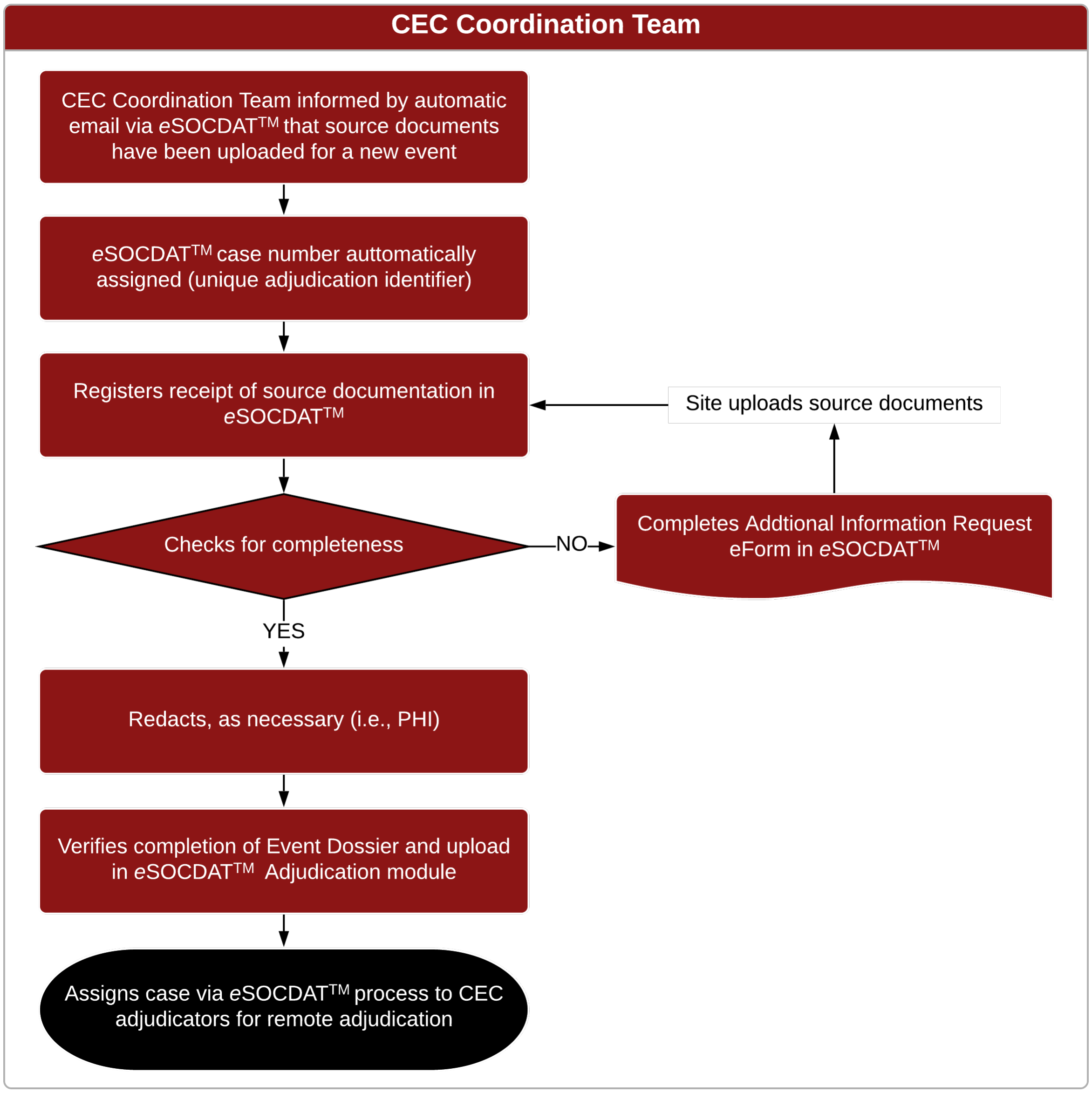

#### 9.6 Adjudication Timeline

The goal turnaround time for CEC adjudicators to return an adjudication decision in eSOCDAT™ will be approximately five (5) to seven (7) days starting from the date of being assigned a case for adjudication. Electronic notifications from eSOCDAT™ will notify the adjudicators in advance of sensitive timelines.

#### 9.7 CEC Adjudicator Review Process

The process for the case assigned to the CEC adjudicators and the subsequent adjudication process is depicted in **Figure** .

Figure 3: CASE ASSIGNMENT AND ADJUDICATION PROCESS

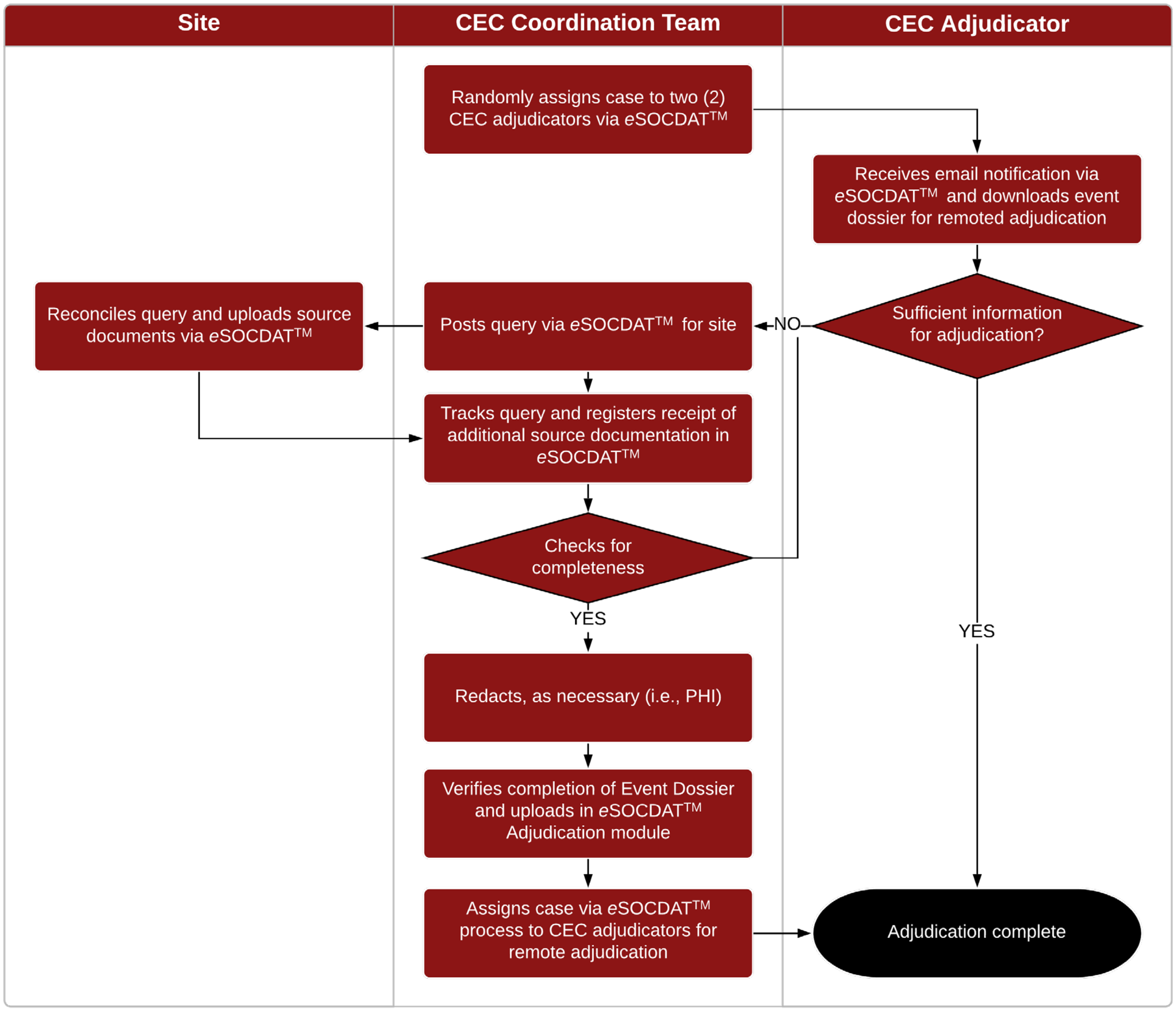

##### 9.7.1 Phase I Review

Phase I review (event adjudication) will consist of a remote, independent adjudication of each event using the criteria listed in the Charter by two CEC adjudicators. The CEC adjudicators will adjudicate the event based on the information submitted in the Event Dossier remotely and independently of each other **Figure** .

If the CEC adjudicator determines that the Event Dossier contains insufficient information for an appropriate adjudication, the CEC adjudicator may request additional information via *e*SOCDAT™. The CEC adjudicator indicate this in the eSOCDAT adjudication module. The SOCAR CEC coordinator will request the site to provide the requested information. Once this requested information is made available in *e*SOCDAT™ from the site, it will be re-distributed to both CEC adjudicators.

The decisions of the two CEC adjudicators will be captured in real time in *e*SOCDAT™. Reports can be generated in *e*SOCDAT™ to display the adjudication status and adjudication outcomes for each case submitted.

If the CEC adjudicators are in agreement, then the adjudication process will be considered complete and the adjudication of the case will be considered final. If the CEC adjudicators are in disagreement, the Phase II review process will be initiated [Section 9.7.2].

**Figure 4: PHASE I ADJUDICATION PROCESS**

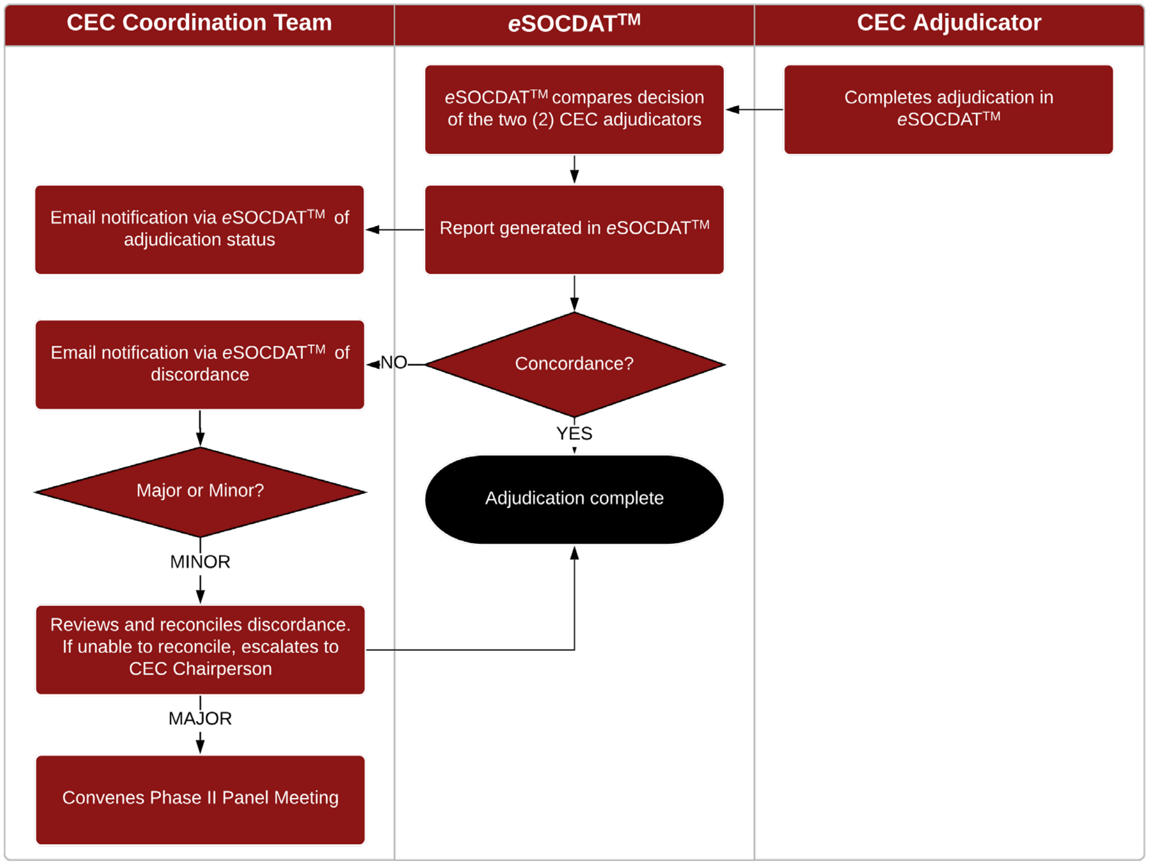

###### 9.7.1.1 Agreements

Once a case has the two decisions submitted and signed by the CEC adjudicators, eSOCDAT™ will automatically compare the forms and when there is 100% consensus, the adjudication will be considered final. The CEC chair will select which data elements require concordance – it is anticipated that minor discordance of some ancillary adjudication details may not be judged sufficiently important to trigger committee review; this will be determined *a priori*.

###### 9.7.1.2 Disagreements

Where there is disagreement, the CEC Coordinator will be notified so that he/she can review the discrepancies to ensure that the latter are not typographical errors.

##### 9.7.2 Phase II Review

Phase II committee (panel) review will be required when the two Phase I CEC adjudicators are in disagreement. Phase II review consists of a disagreement meeting at which at least 3 physicians from the committee are in attendance [**Figure** ]. The disagreement meeting will be convened by teleconference. The final decision is achieved by consensus, which is then documented and entered in *e*SOCDAT™ by the CEC Chairperson(s) or designee.

**Figure 5: PHASE II ADJUDICATION PROCESS**

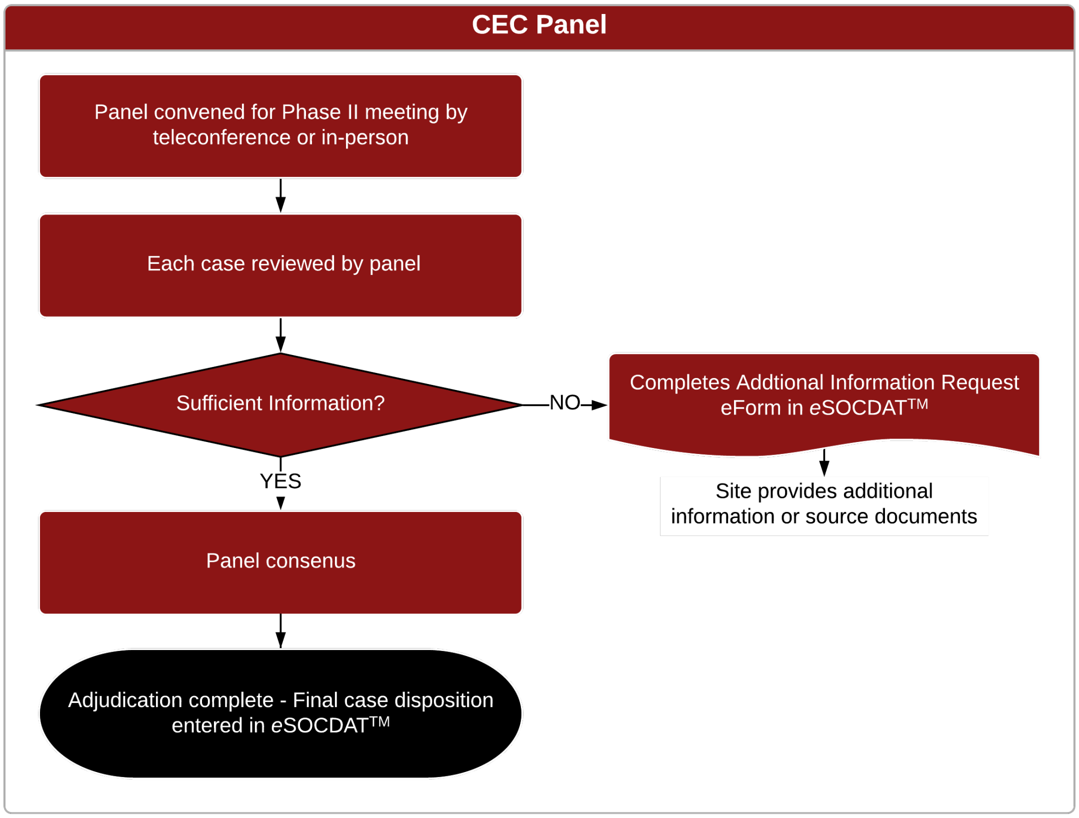

If the Phase II committee determines that insufficient information is available for an appropriate adjudication, the CEC Chairperson(s) or designee may request additional information via *e*SOCDAT™. Once this requested information is made available in *e*SOCDAT™ from the site, it will be re- distributed to the Phase II committee.

The CEC Coordination Team will document all disagreement meeting discussions which consist of the following contents: date of the meeting, member attendance, and a list of the case numbers reviewed. The documentation will be archived.

The turnaround time for Phase II will be approximately five (5) to seven (7) business days starting from the date the second of the two adjudicators completes the Phase I adjudication. However, the Phase II review meetings may be scheduled at a convenient time to combine the review of several cases with disagreement. This may result, for some cases, in a turnaround time of more than 10 days. Eventually, the turnaround time for a given case should not exceed 1 month.

##### 9.7.3 Data Changes after the Event Adjudication

Once an event has completed adjudication, additional information about that event will not be routinely sought. However, if additional or corrected data become available, the CEC Coordinator will be informed via eSOCDAT and will perform a preliminary review of the additional or corrected data to determine if the new data may affect the adjudication result. If the new data is not significant (i.e., logistical error such as transcription error of a date), the event will not be re-adjudicated. If the additional or corrected data is clinical in nature, the CEC Coordinator will discuss the new data with the CEC Chairperson to see if this may affect the adjudication result. If this is the case, the event will be re-sent to the original reviewers for re-adjudication. The case will subsequently be updated in *e*SOCDAT™ to reflect any changes to the initial adjudication.

If the CEC Chairperson or CEC Coordinator concludes that the new data does not require re-review of the event, the event will not be submitted for re- review. The decision and its rationale will be documented in eSOCDAT.

#### 9.8 CEC Quality Assessment

The SOCAR CEC Coordination Team will be responsible for implementing and maintaining Quality Assessment of the endpoint assessment and adjudication procedures.

##### 9.8.1 Quality Assessment review and documentation

After each quality assessment milestone [Section 9.8], a quality assessment report will be generated in *e*SOCDAT™ and submitted to the CEC Chairperson for review. Once approved by the CEC Chairperson, the report will be submitted to Study Chair. All quality assessment findings will be reviewed by the CEC Chairperson who will decide what action should be taken in case of discrepancies. Such action may include (but are not limited to):

- re-training of CEC adjudicators,
- call a CEC meeting,
- increase the number of cases to be included in the quality assessment step,
- re-adjudicate events.

The quality assessment report will also be used to determine reproducibility and reliability of methods used in the adjudication process.

Based on the Quality Assessment report, the CEC Operations Team may perform the following:

- Make no modification to the adjudication process
- Modify the adjudication process
- Provide additional training on the adjudication process

#### 9.9 Committee Meetings

The CEC Coordination Team may conduct Committee meetings to review the operational processes throughout the course of the study, and on an *ad hoc* basis.

The Committee meetings will be convened preferably in the form of video conference. The CEC Coordination Team will draft the agenda in collaboration with the CEC Chair and CEC adjudicators.

The committee meetings will address topics such as (but not limited to):

1. General Study Update
2. Discuss Phase I & II adjudication discrepancy and review the event
3. Discuss CEC quality assessment report
4. Discuss changes in event definitions

5. Provide feedback on any other issues that arise in the adjudication process

The CEC Coordination Team will document all meeting discussions which consist of the following contents: attendance, key discussions and inputs, and decisions made by the attendees. The contents will be documented in the form of meeting minutes, meeting agenda, and/or email exchanges.

#### 9.10 *Ad hoc* Meetings

Ad-hoc Adjudication Committee meetings may be organized at the request of the Adjudication Committee chairperson, in case new information becomes available from the clinical study or other ongoing, related trials that may have an impact on the adjudication of the data, or for any other reason.

### *10 DATA HANDLING*

All endpoint adjudication data and documentations will be retained in *e*SOCDAT™. At the end of the project or at a mutually agreed upon time, the stored data will be transferred to the Sponsor.

### *11 DOCUMENT RETENTION*

#### 11.1 Administrative Documents

All administrative study documents that do not contain subject details will be maintained by the CEC chair's office. Documents may include but is not limited to the Protocol, Plans, Manuals, and Training Records.

#### 11.2 Documents for Events Adjudication

All event packages, query communications, and adjudication forms will be stored in *e*SOCDAT™. At the end of the study or a mutually agreed upon time, the complete event packets (adjudication forms, outcome event supportive documents, and queries) will be returned as directed by the trial Sponsor.

#### 11.3 Document archival and study closure

The SOCAR CEC Coordinator will make the Adjudication Committee meeting minutes and the final Adjudication Committee report available to the Adjudication Committee members.

The Curriculum Vitae of the Adjudication Committee members, including description of experience relevant for the clinical study and conflict of interest disclosure, will be archived in the eTMF.

### 12 EVENT DEFINITIONS

#### 12.1 Deep Venous Thromboembolism

The diagnosis of <u>definite</u> symptomatic deep venous thromboembolism (DVT) requires symptoms of venous thromboembolism with at least one of the following:

- Abnormal compression ultrasound consistent with DVT or abnormal flow pattern or direct clot visualization in veins not amenable to compression.
- One or more new filling defects by venography, CT venography, or MR venography.
- Abnormal compression ultrasound where compression had been normal or, if known to be non-compressible, a substantial increase (≥4mm) in the diameter of a previously non-compressible venous segment.
- Point-of-care ultrasound (POCUS) performed by a provider and documenting DVT in a note.
- An extension of an intraluminal filling defect, or a new intraluminal filling defect, or an extension of non-visualization of veins in the presence of a sudden cut-off on venography.
- Proximal DVT is defined as clot at or proximal to the trifurcation of the popliteal vein (in the lower extremity) OR clot at or proximal to the axillary vein segment (in the upper extremity).
- Distal DVT is defined as clot distal to the trifurcation of the popliteal vein (in the lower extremities) OR clot at or distal to the brachial vein segment (in the upper extremities).
- Non-limb venous thrombosis includes thrombosis of the cerebral, portal, mesenteric, hepatic, gonadal, splenic, renal, or retinal veins, or thrombosis of the superior or inferior vena cava. In aggregated

analyses with other platforms, cerebral sinus thrombosis may be considered in the stroke category; however, for adjudication purposes it is adjudicated as a venous thrombosis event in ATTACC.

The diagnosis of presumed deep venous thromboembolism requires the following:

- In the absence of objective testing, high pre-test probability according to investigator assessment
  - OR adjudicator's gestalt
  - OR Wells score
- AND a treatment plan for DVT was initiated (initiation of anticoagulation, or escalation of anticoagulation dose, frequency, or duration).

#### 12.2 Pulmonary Embolism

The diagnosis of definite pulmonary embolism requires at least one of the following:

- New intraluminal filling defect at CT pulmonary angiography in a subsegmental or larger vessel.
- New intraluminal filling defect, or an extension of an existing defect, or a new sudden cut-off of vessels > 2.5 mm in diameter at pulmonary angiogram,
- Inconclusive CT pulmonary angiography, pulmonary angiography, or VQ scan evidence of a new or recurrent PE with demonstration of a new or recurrent DVT in the lower extremities by compression ultrasonography or venography. [2, 3]
- New clot or intraluminal filling defect noted in the right heart ("clot in transit") or the pulmonary vasculature at echocardiogram
- High probability (revised PIOPED criteria) on planar ventilation/perfusion (V/Q) scan OR positive PE on SPECT ventilation perfusion (V/Q) scan.
- Pulmonary embolism found at autopsy

The diagnosis of presumed pulmonary embolism requires the following:

Clinical signs and symptoms of pulmonary embolism, including but not limited to: dyspnea, cough, hypoxemia, tachycardia, appropriate electrocardiographic changes, or evidence of right heart strain on echocardiogram; AND chest CT or pulmonary angiography are unable to be performed AND therapeutic dose anticoagulation or fibrinolytic therapy is prescribed by a physician

#### 12.3 Systemic arterial thrombosis or embolism

The diagnosis of systemic arterial thrombosis or embolism is defined as the following:

- A clinical history and presentation consistent with a sudden significant worsening of end organ or limb perfusion AND

EITHER

- Confirmation of arterial obstruction by imaging, hemodynamics, intraoperative findings, or pathological evaluation

OR

- Requirement for thrombolysis, thrombectomy, or urgent bypass.

Note that this endpoint includes both acute *in situ* thrombotic events and acute embolic events. Note that while ischemic stroke and myocardial infarction can be arterial thromboembolic events, those events will be adjudicated according to the separate standardized criteria included below.

#### 12.4 Myocardial Infarction

COVID-19 patients are well known to have elevations in cardiac troponin concentrations, and these elevations often do not represent arterial thrombosis and

**Figure 6**: Myocardial injury/infarction.

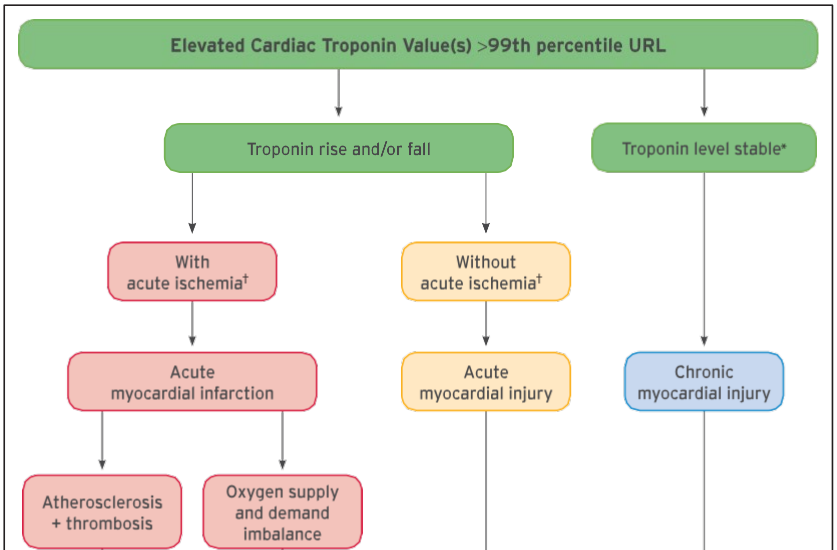

downstream myocardial ischemia. Therefore, the CEC will make an effort to distinguish true myocardial infarction from coronary artery obstruction, typically from atherothrombosis (usually considered a “type 1 myocardial infarction”) from myocardial infarction due to demand ischemia (usually defined as a “type 2 myocardial infarction”) and myocardial injury (an elevation in cardiac troponin typically without symptoms of chest pain or signs of arterial thrombosis) (**FIGURE 6**). These definitions will be consistent with the 4th Universal Definition of Myocardial Infarction (**FIGURE 7**) and will take into considerations suggestions made about classification of certain conditions as type 1 as compared to type 2 myocardial infarction. Regional coronary venous thrombosis with associated regional myocardial infarction has been reported in COVID-19. If this mechanism is documented, these will be considered and type 1 MI. The trial and CEC are focused on ascertaining and adjudicating cases of acute myocardial injury and acute myocardial infarction, and classifying those cases as described below. COVID-19 also causes microvascular thrombi which are associated with patchy myocardial necrosis. These will be grouped with myocardial injury.

**Figure 7**: Fourth Universal Definition of Myocardial Infarction

#### 2. UNIVERSAL DEFINITIONS OF MYOCARDIAL INJURY AND MYOCARDIAL INFARCTION: SUMMARY

| Universal definitions of myocardial injury and myocardial infarction |
|---|
| **Criteria for myocardial injury** |
| The term myocardial injury should be used when there is evidence of elevated cardiac troponin values (cTn) with at least 1 value above the 99th percentile upper reference limit (URL). The myocardial injury is considered acute if there is a rise and/or fall of cTn values. |
| **Criteria for acute myocardial infarction (types 1, 2 and 3 MI)** |
| The term acute myocardial infarction should be used when there is acute myocardial injury with clinical evidence of acute myocardial ischemia and with detection of a rise and/or fall of cTn values with at least 1 value above the 99th percentile URL and at least 1 of the following:<br>• Symptoms of myocardial ischemia;<br>• New ischemic ECG changes;<br>• Development of pathological Q waves;<br>• Imaging evidence of new loss of viable myocardium or new regional wall motion abnormality in a pattern consistent with an ischemic etiology;<br>• Identification of a coronary thrombus by angiography or autopsy (not for types 2 or 3 MIs).<br>Postmortem demonstration of acute atherothrombosis in the artery supplying the infarcted myocardium meets criteria for *type 1 MI*. Evidence of an imbalance between myocardial oxygen supply and demand unrelated to acute atherothrombosis meets criteria for *type 2 MI*. Cardiac death in patients with symptoms suggestive of myocardial ischemia and presumed new ischemic ECG changes before cTn values become available or abnormal meets criteria for *type 3 MI*. |
| **Criteria for coronary procedure–related myocardial infarction (types 4 and 5 MI)** |

| |
|---|
| Percutaneous coronary intervention (PCI)–related MI is termed *type 4a MI*. |
| Coronary artery bypass grafting (CABG)–related MI is termed *type 5 MI*. |
| Coronary procedure–related MI ≤48 hours after the index procedure is arbitrarily defined by an elevation of cTn values >5 times for *type 4a MI* and >10 times for *type 5 MI* of the 99th percentile URL in patients with normal baseline values. Patients with elevated preprocedural cTn values, in whom the preprocedural cTn level are stable (≤20% variation) or falling, must meet the criteria for a >5 or >10 fold increase and manifest a change from the baseline value of >20%. In addition with at least 1 of the following:<br>• New ischemic ECG changes (this criterion is related to *type 4a MI* only);<br>• Development of new pathological Q waves;<br>• Imaging evidence of loss of viable myocardium that is presumed to be new and in a pattern consistent with an ischemic etiology;<br>• Angiographic findings consistent with a procedural flow-limiting complication such as coronary dissection, occlusion of a major epicardial artery or graft, side-branch occlusion-thrombus, disruption of collateral flow or distal embolization. |
| Isolated development of new pathological Q waves meets the *type 4a MI* or *type 5 MI* criteria with either revascularization procedure if cTn values are elevated and rising but less than the prespecified thresholds for PCI and CABG. |
| Other types of 4 MI include *type 4b MI* stent thrombosis and *type 4c MI* restenosis that both meet *type 1 MI* criteria. |
| Postmortem demonstration of a procedure-related thrombus meets the *type 4a MI* criteria or *type 4b MI* criteria if associated with a stent. |
| **Criteria for prior or silent/unrecognized myocardial infarction** |
| Any 1 of the following criteria meets the diagnosis for prior or silent/unrecognized MI:<br>• Abnormal Q waves with or without symptoms in the absence of nonischemic causes.<br>• Imaging evidence of loss of viable myocardium in a pattern consistent with ischemic etiology.<br>• Patho-anatomical findings of a prior MI. |

CABG indicates coronary artery bypass grafting; cTn, cardiac troponin; ECG, electrocardiogram; MI, myocardial infarction; PCI, percutaneous coronary intervention; URL, upper reference limit.

**Myocardial Injury:** The increasing sensitivity of cardiac troponin (cTn) assays means that ongoing myocardial injury is frequently detected.

Myocardial injury is a prerequisite for myocardial infarction (MI), but as noted below, criteria in addition to myocardial injury are necessary to make the diagnosis of MI. Adjudicators must distinguish between acute myocardial injury that is not secondary to ischemia but may be due to other

conditions (**TABLE 1**).

<u>Criteria for Myocardial Injury:</u> Detection of an elevated cTn value above the 99$^{th}$ percentile upper reference limit (URL) is defined as myocardial injury. The injury is considered acute if there is a rise and/or fall of cTn values.

<u>Criteria for Procedure Related Myocardial Injury:</u> Cardiac procedural myocardial injury is arbitrary defined by increased in cTn values (>99$^{th}$ percentile URL) in patients with normal baseline values (<99$^{th}$ percentile URL) or a rise of cTn values >20% of the baseline value when it is the above the 99$^{th}$ percentile URL but is stable or falling.

**Table 1.** Example causes of myocardial injury.

Heart failure; Myocarditis; Cardiomyopathy; Takotsubo syndrome; Coronary revascularization procedure; Cardiac procedure other than revascularization; Catheter ablation; Defibrillator shocks; Cardiac contusion; Sepsis, infectious disease; Chronic kidney disease; Stroke, subarachnoid hemorrhage; Pulmonary embolism, pulmonary hypertension; Infiltrative disease, e.g., amyloidosis, sarcoidosis

Chemotherapeutic agents; Critically ill patients; Strenuous exercise Other

**Myocardial Infarction Type 1**: Detection of rise and/or fall of cardiac biomarkers with at least one value above the 99th percentile of the upper reference limit (URL) together with evidence of myocardial ischemia with at least one of the following:

- Symptoms of ischemia
- New ischemic ECG changes indicative of new ischemia (new ST-T changes or new LBBB)*
- Development of pathological Q waves in the ECG**
- Imaging evidence of new loss of viable myocardium or new regional wall motion abnormality in a pattern consistent with an ischemic etiology
- Identification of a coronary thrombus by angiography including intracoronary imaging or by autopsy†
- *ECG manifestation of acute myocardial ischemia (in the absence of LVH and LBBB):
    - ST Elevation: New ST elevation at the J-point in two contiguous leads with the cut-point: ≥ 1 mm in all leads other than leads V2-V3, where the following cut-points apply: ≥2mm in men ≥40 years; ≥2.5 mm in men <40 years; or ≥ 1.5 mm in women regardless of age.
    - ST-depression and T-wave changes: New horizontal or down- sloping ST depression ≥ 0.5 mm in 2 contiguous leads and/or T inversion ≥ 1 mm in two

contiguous leads with prominent R waves or R/S ratio >1.

- **Pathological Q waves:
  - Any Q-wave in leads V2-V3 >0.02 seconds or QS complex in leads V2-V3
  - Q-wave ≥ 0.03 seconds and ≥1 mm deep or QS complex in leads I, II, aVL, aVF, or V4-V6 in any 2 leads of a contiguous lead grouping (I, aVL;V1-V6; II, III, aVF; V7-V9).
  - R-wave ≥ 0.04s in V1–V2 and R/S ≥ 1 with a concordant positive T-wave in the absence of a conduction defect
- †Postmortem demonstration of an atherothrombus in the artery supplying the infarcted myocardium, or a macroscopically large circumscribed area of necrosis with or without intramyocardial hemorrhage meets the type 1 MI criteria regardless of cTn values.
- Consideration will be given to recent proposals to modify myocardial infarction type 1 to include coronary obstruction by spontaneous coronary artery dissection, coronary embolism, or coronary vasospasm or microvascular dysfunction.[4]

**Myocardial Infarction Type 2:** Detection of a rise and/or fall of cTn values with at least 1 value above the $99^{th}$ percentile URL, and evidence of imbalance between myocardial oxygen supply and demand unrelated to coronary atherothrombosis, requiring at least 1 of the following:

- Symptoms of acute myocardial ischemia
- New ischemic ECG changes
- Development of pathological Q waves;
- Imaging evidence of new loss of viable myocardium or new regional wall motion abnormality in a pattern consistent with ischemic etiology

**Myocardial Infarction Type 3:** Patients who suffer cardiac death, with symptoms suggestive of myocardial ischemia accompanied by presumed new ischemic ECG changes or ventricular fibrillation but die before blood samples for biomarkers can be obtained, or before increases in cardiac biomarkers can be identified, or MI is detected by autopsy examination.

**Myocardial infarction Type 4a and 4b (myocardial infarction associated with percutaneous coronary intervention):** Criteria for percutaneous coronary intervention (PCI)-related MI ≤48 hours after the index procedure are as follows: Coronary intervention-related MI is arbitrarily defined by an elevation of cTn values >5 times the 99the percentile URL in patients with normal baseline values. In patients with elevated preprocedural cTn in whom the cTn levels are stable (≤20% variation) or falling, the post procedure cTn must rise by >20%. However, the absolute procedural value must still be at least 5 times the 99$^{th}$ percentile URL. In addition, 1 of the following elements is required:

- New ischemic ECG changes
- Development of new pathological Q waves; note that the development of new pathological Q waves meets the criteria for procedure-related MI if the cTn values are elevated and rising but <5 times the 99$^{th}$ percentile URL.
- Imaging evidence of new loss of viable myocardium or new regional wall motion abnormality in a pattern consistent with an ischemic etiology
- Angiographic findings consistent with a procedural flow-limiting complication such as coronary dissection, occlusion of a major epicardial artery or side branch occlusion/thrombus, disruption of collateral flow, or distal embolization.
- Type 4a MI is an MI associated with PCI
- Type 4b MI is an MI associated with stent/scaffold thrombosis

**Myocardial Infarction Type 4c**: A type 4c MI is an MI associated with restenosis associated with prior PCI. Possible Type 4c MI is evaluated using the same criteria as Type 1 MI.

**Myocardial Infarction Type 5**: Criteria of coronary artery bypass grafting (CABG)-related MI ≤ 48 hours after the index procedure. CABG-related MI is arbitrarily defined as elevation of cTn values >10 times the 99$^{th}$ percentile URL in patients with normal baseline cTn values. In patients with elevated preprocedural cTn in whom cTn are stale (≤20% variation) or falling, the post procedure cTn must rise by >20%. However, the absolute postprocedural values must still be >10 times the $99^{th}$ percentile URL. In addition, 1 of the following elements is required:

- Development of new pathological Q waves; note that the development of new pathological Q waves meets the criteria for procedure-related MI if the cTn values are elevated and rising but <10 times the $99^{th}$ percentile URL.
- Angiographically documented new graft occlusion or new native coronary artery occlusion;
- Imaging evidence of new loss of viable myocardium or new regional wall motion abnormality in a pattern consistent with an ischemic etiology

**Special or unusual circumstances:** Further guidance on distinguishing myocardial injury from myocardial infarction in the context of non-cardiac surgery, heart failure, myocarditis, Takotsubo syndrome, kidney disease, and in critically ill patients, and myocardial infarction nonobstructive coronary arteries is included in the $4^{th}$ Universal Definition of MI.[1]

#### 12.5 Stroke

The definition of stroke used here is drawn from the definitions proposed by Hicks et al. and Sacco et al.[6, 7] Stroke is defined as the acute onset of focal neurological dysfunction caused by brain, spinal cord, or retinal vascular injury as a result of hemorrhage or infarction. Note that while all strokes will be adjudicated, only ischemic stroke is part of the primary efficacy endpoint. Hemorrhagic stroke is considered a major bleed (see section 6.7).

A stroke is the acute onset of a new persistent neurological deficit attributed to an obstruction in cerebral blood flow with no apparent nonvascular cause (e.g. tumor, trauma, infection). Available neuroimaging studies will be considered to support the clinical impression and to determine if there is a demonstrable lesion compatible with an acute stroke. To the extent possible, all strokes will be classified as ischemic, hemorrhagic or unknown. While all types of strokes will be adjudicated by the CEC, only ischemic strokes will be included in the primary endpoint.

For the diagnosis of stroke, the following criteria should be fulfilled:

**1.** Rapid onset of a focal neurological deficit not related to any other known non-cerebrovascular process with at least one of the following:

- Change in level of consciousness
- Hemiplegia
- Hemiparesis
- Numbness or sensory loss affecting one side of the body
- Dysphasia/aphasia
- Hemianopia
- Other new neurological sign/symptom(s) consistent with stroke
- If the timing of onset is uncertain, a diagnosis of stroke may be made provided that there are no plausible non-stroke causes for the clinical presentation.

AND

**2.** Duration of a focal/global neurological deficit that is:

- EITHER ≥ 24 hours,
- OR < 24 hours if:
    - Resolution of symptoms is due to least one of the following interventions:
        1. Pharmacologic: intravenous or intraarterial thrombolysis
        2. Non-pharmacologic: (i.e. neuro-interventional procedure

such as intracranial angioplasty)

- OR available MRI clearly documents a new hemorrhage or infarct
- OR available head CT clearly documents a new hemorrhage or infarct or excludes a mimic of stroke
- OR the neurological deficit results in death.

Ideally, at least one of should be present to confirm the diagnosis of stroke:

- Confirmation by neurology or neurosurgery specialist
- Brain imaging procedure (at least one of the following): CT scan, MRI scan, or cerebral vessel angiography
- Lumbar puncture (i.e. spinal fluid analysis diagnostic of intracranial hemorrhage)

If the acute focal signs represent a worsening of a previous deficit, these signs must persist for more than 24 hours and be accompanied by an appropriate new MRI or CT scan finding.

Strokes are sub-classified as follows:

*Ischemic (non-hemorrhagic):* An acute episode of focal cerebral, spinal, or retinal dysfunction caused by infarction of central nervous system tissue. Hemorrhage may be a consequence of ischemic stroke. In this situation, the stroke is an ischemic stroke with hemorrhagic transformation and not a hemorrhagic stroke; such events should nonetheless also be considered as major bleeding events as they meet ISTH criteria based on intracranial bleeding.

*Hemorrhagic:* An acute episode of focal or global cerebral or spinal dysfunction caused by intraparenchymal, intraventricular, or subarachnoid hemorrhage. Hemorrhage in the brain is documented by neuroimaging or autopsy or lumbar puncture. Note that subdural hematomas are intracranial hemorrhagic events and not strokes.

*Undetermined:* An acute episode of focal or global neurological dysfunction caused by presumed brain, spinal cord, or retinal vascular injury as a result of hemorrhage or infarction but with insufficient information to allow categorization as either ischemic or hemorrhagic.

#### 12.6 Major Bleeding

Major bleeding is defined as acute clinically overt bleeding associated with one or more of the following (as per ISTH guidelines for non-surgical patients):[8, 9]

- Decrease in hemoglobin of 2 g/dL or more;
- Transfusion of 2 units or more of packed red blood cells;
- Bleeding that occurs in at least one of the following critical sites:
    - Intracranial
    - Intraspinal
    - Intraocular (within the corpus of the eye. A conjunctival bleed is not an intraocular bleed)
    - Pericardial
    - Intraarticular
    - Retroperitoneal
    - Intramuscular with compartment syndrome
- Bleeding that leads to death (primary cause of death or contributes directly to death)

#### 12.7 Heparin-Induced Thrombocytopenia

The committee will use the definition of HIT recently proposed by McGowan *et al.* (McGowan *Blood* 2016), adapted for the present study ("possible HIT" definition added). An optical density ≥0.4 will be used as the threshold for the anti-PF4 ELISA test. A quantitative rapid immunoassay (RIA) ≥1 unit/ml will be considered as positive. The following categorization of site-reported HIT events will be made:

● *Definite HIT*:

- Positive SRA or equivalent functional HIT confirmatory test

- Positive ELISA AND treated as HIT

- Positive quantitative RIA AND treated as HIT

● *Possible HIT*:

- Positive ELISA or quantitative RIA AND any heparin/LMWH AC stopped

- 4T score high or intermediate (at least >3), no Laboratory testing for HIT done, AND treated as HIT

● *No HIT*

- Negative HIT ELISA, or Negative Quantitative RIA

- Negative SRA, or Negative equivalent functional confirmatory assay

- No laboratory testing for HIT done AND not treated as HIT (heparin continued and uneventful course (defined as absence of thrombotic event, platelet count recovery during follow-up).

#### Appendix A: ATTACC Events Adjudication Committee Membership

| **Events Adjudication Committee** | **Professional Role(s)** |
|---|---|
| CEC Chairman | Brendan Everett, MD, MPH |
| CEC Co-Chairman | Sean van Diepen, MD, MSc** |
| CEC Adjudicator #1 | Gregoire Le Gal, MD* |
| CEC Adjudicator #2 | Deborah Siegal, MD* |
| CEC Adjudicator #3 | Jean-Philippe Galanaud, MD* |
| CEC Adjudicator #4 | Sheila Hegde, MD** |
| CEC Adjudicator #5 | Yuri Kim, MD** |
| CEC Adjudicator #6 | Natalia Rost, MD*** |
| CEC Adjudicator #7 | Aneesh Singhal, MD*** |

*Adjudicator for VTE, bleeding, and HIT.
**Adjudicator for arterial thrombotic events, excluding stroke.
*** Adjudicator for stroke.
The CEC hold final discretion over adjudicator roles based on expertise and events.

#### Appendix B: Conflict of Interest

Before being confirmed in their position, candidates for membership of the Clinical Events Committee shall notify the CEC Chairperson(s) in writing *(using TPL-100648)* of any actual or potential conflict of interest between their personal interests and their role in the Clinical Events Committee, including without limitation:

- financial arrangements that could be affected by the outcome of the clinical study, or any other financial interest in the outcome of the clinical study; or
- any financial relationship, including without limitation research grants, fees for services rendered and retainers for consultancy services, but not including payments solely for participation in the Clinical Events Committee; or
- an equity interest (including without limitation shares, stock options, dividends, profits, returns on investment and/or other derivatives) by the candidate and/or his/her immediate family, in the owner of the study drug, or in any other company whose share value could reasonably be affected by the outcome of the clinical study; or
- involvement in the study other in a leadership capacity, for example as a steering committee member, site investigator, or other role which could be potentially conflicted through inadvertent loss of blinding in the process of adjudication, or which may be perceived to present a conflict as an adjudicator where an investigator may present biased interest in assessment of events; or
- any involvement, in clinical studies or other projects concerning the same indication, or concerning the same, related or competing study drug.

#### Appendix C: Source Documents Requested from Site, by Event Type

**Types of Source Documents Required**

In all cases, study staff are asked to provide a narrative of the event as understood by the study staff.

*** Those documents essential for adjudication are identified with an "*".** Other documents should be uploaded if performed.

| **Myocardial Infarction** |
|---|
| **Clinical documentation:** |
| *Admission history and physical |
| *Discharge summary |
| *Cardiac biomarker laboratory values and normal ranges (troponin values from 3 days before and after reported date of MI) |
| ECG (baseline, pre-event) |
| *ECG (event) |
| Imaging reports (echo/stress echo, radionuclide imaging, angiography, CT, MRI etc.) |
| Procedure notes (PCI, CABG, other) |
| Autopsy report |
| Event summary signed by the Principal Investigator if there are no source documents available |
| **Stroke** |
| *Admission history and physical |
| *Discharge summary |
| *Relevant progress notes |
| *Neurology and/or neurosurgical consult (if available) |
| *Imaging reports (CT, MRI, cerebral angiogram, carotid echo, cardiac echo) |
| *Procedure notes (cerebral angio and/or intervention) |
| Lumbar puncture results (if available) |
| Autopsy report |
| Event summary signed by the Principal Investigator if there are no source documents available |
| **Deep Venous Thromboembolism** |
| *Admission history and physical (including VTE pre-test probability if assessed) |
| *Discharge summary |
| Pretest probability if assessed |
| Outpatient clinic notes, if applicable |
| *Consulting notes from hematology/thrombosis/cardiology consultant/ER |
| Venous ultrasound report |
| CT, MR or invasive venography report (lower limb, abdominal/chest/head CT/MRI) |
| *Point-of-care ultrasound with documentation of deep vein thrombus in a note |
| *D-dimer laboratory values from 3 days before and after reported date of DVT |

| |
|---|
| Document of change in therapy in response to diagnosis |
| Autopsy report, if applicable |
| Event summary signed by the Principal Investigator if there are no source documents, or only minimal source documents available. |
| **Pulmonary Embolism** |
| *Admission history and physical (including VTE pre-test probability if assessed) |
| *Discharge summary |
| *Consulting notes from hematology/thrombosis/cardiology consultant/ER |
| *Pulmonary angiography report |
| *CT pulmonary angiography report |
| *Ventilation/perfusion (V/Q) scan report |
| *D-dimer laboratory values from 3 days before and after reported date of PE |
| Cardiac echo |
| Autopsy report, if applicable |
| Clinical documentation to support evidence of hemodynamic instability with acute right ventricular dysfunction with a corresponding therapeutic intervention (anticoagulation and/or fibrinolysis) |
| Clinical documentation demonstrating signs and symptoms of pulmonary embolism with evidence of therapeutic anticoagulation |
| Event summary signed by the Principal Investigator if there are no source documents, or only minimal source documents available. |
| **Arterial Thromboembolism** |
| *Admission history and physical |
| *Discharge summary |
| Outpatient clinic notes, if applicable |
| *Imaging demonstrating arterial obstruction |
| *Operative reports, if applicable |
| Pathologic evaluation, if applicable |
| Medication administration records |
| Autopsy report, if applicable |
| Event summary signed by the Principal Investigator if there are no source documents, or only minimal source documents available. |
| **Major Bleeding** |
| *Admission history and physical |
| *Discharge summary |
| Outpatient clinic notes or inpatient physician/nursing notes including specialist consultant notes (as appropriate) |
| *Laboratory results demonstrating change in hemoglobin if the hemoglobin criteria alone was used to support ISTH Major Bleeding |
| *Clinical documentation of bleeding |
| Operative or procedural (including diagnostic) reports |
| Documentation of blood transfusion (notes, transfusion records, physician orders, etc.) |
| Autopsy report |
| Imaging reports (e.g. abdominal US, head CT scan) |
| **Heparin-induced thrombocytopenia** |
| *Admission history and physical |

| |
|---|
| *Discharge summary and/or other pertinent narrative notes |
| *Lab records with platelet (PLT) counts within 3 days before and after event |
| *Confirmatory testing: ELISA, SRA and any lab test considered to be relevant by the investigator (beyond platelet count) |
| Document of change in therapy in response to diagnosis |
| Imaging report if thrombosis was diagnosed or suspected<br>- lower or upper extremity DVT: US or CT<br>- PE: US, V/Q scan, chest CTPA, echocardiogram<br>- non-limb deep vein thrombosis: abdominal CT/MRI/US, head CT/MRI |

* These documents are essential for adjudication

**CLINICAL ENDPOINTS COMMMITTEE (CEC) ADJUDICATION PROCEDURES**

### ACTIV-4 ACUTE (AC-INPT)

**A Multicenter, Adaptive, Randomized Controlled Platform Trial of the Safety and Efficacy of Antithrombotic Strategies in Hospitalized Adults with COVID-19**

**ClinicalTrials.gov Number: NCT04505774**

**Sponsor:**

**National Heart Lung and Blood Institutes (NHLBI), National Institute of Neurological Disorders (NINDS), ACTIV (ACTIV IV), National Institutes of Health (NIH), Biomedical Advanced Research and Development Authority (BARDA), Operation Warp Speed (OWS), U.S. Department of Health & Human Services (HHS)**

**Version 1.2**

**January 4, 2021**

TABLE OF CONTENTS

### Signature Page

| REQUIRED SIGNATURES |
|---|

**CEC Chair**

| | | |
|---|---|---|
| Brendan Everett, MD, MPH | 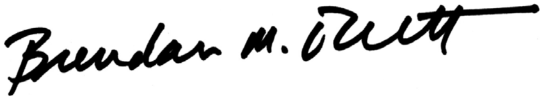 | 01/07/2021 |
| Print Name | Signature | DDMMMYYYY |

**Trial Chair**

| | | |
|---|---|---|
| Judith Hochman, MD | 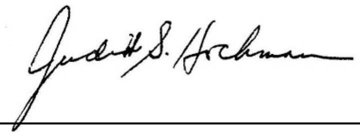 | 01/07/2021 |
| Print Name | Signature | DDMMMYYYY |

### INTRODUCTION

#### 1.0. MISSION OF THE CLINICAL ENDPOINT COMMITTEE

The CEC adjudication committee ("The Committee") is an independent clinical endpoint adjudication committee, convened to evaluate and adjudicate, in a blinded fashion, specified endpoints for the trial.

The ACTIV-4 Acute trial primary endpoint is the number of days that a patient is alive and free of organ support through 21 days after trial entry. Organ support is defined by receipt of invasive or non-invasive mechanical ventilation, high flow nasal oxygen, vasopressor therapy, or ECMO support. If the patient dies at any time (including beyond 21 days) during the index hospital stay, they are assigned the worst possible score of –1. The Clinical Endpoints Committee is NOT adjudicating the ACTIV-4 Acute trial primary endpoint.

The ACTIV-4 Acute trial Secondary Endpoint is a composite of death, pulmonary embolism, systemic arterial thromboembolism, myocardial infarction, or ischemic stroke during hospitalization or at 28 days after enrollment (whichever is earlier).

Other ACTIV-4 Secondary Endpoints include:

- A composite endpoint of death, deep vein thrombosis, pulmonary embolism, systemic arterial thromboembolism, myocardial infarction, or ischemic stroke during hospitalization or at 28 days after enrollment (whichever is earlier)
- 28 Day Hospital free days (non-ICU level patients)
- 28 Day Ventilator-Free Days (ICU level patients)
- 28 Day Vasopressor-Free Days (ICU level patients)
- 28 Day Renal Replacement Free Days
- Hospital readmission within 28 days
- Acute kidney injury as defined by KDIGO criteria
- Deep vein thrombosis
- Pulmonary embolism
- Systemic arterial thrombosis or embolism
- Myocardial infarction
- Ischemic stroke
- Use of extracorporeal membrane oxygenation (ECMO) support
- Mechanical circuit (dialysis or ECMO) thrombosis
- All-cause mortality at 28 days
- Organ support free days at 28 days
- All-cause mortality during initial hospitalization (includes death after 28 days)
- WHO ordinal scale (peak scale over 28 days, scale at 14 days, and proportion with improvement by at least 2 categories compared to enrollment, at 28 days)
- All-cause mortality at 90 days

The specified endpoints that will be adjudicated by the Clinical Endpoints Committee are:

- **Pulmonary embolism**
- **Deep vein thrombosis**
- **Arterial thromboembolism**
- **Myocardial infarction**
- **Ischemic stroke**
- **Major bleeding**

The Committee will fulfill its mission through ad hoc meetings, teleconferences and mailings, in order to allow an ongoing and timely evaluation of events.

#### 2.0. MEMBERS OF THE CLINICAL ENDPOINT COMMITTEE

The CEC is an independent group consisting of physicians (MDs) who have expertise in the clinical adjudication of thrombotic and thromboembolic events, including venous thromboembolism, pulmonary embolus, coronary and cerebrovascular events, bleeding, and death. The following experts form the Clinical Endpoint Committee for CIRT:

| Expert | Institution | City, Country | Subspecialty |
|---|---|---|---|
| Brendan Everett, MD (CEC Chair) | Brigham and Women's Hospital | Boston, USA | Cardiology |
| Sean Van Diepen, MD | University of Alberta Hospital | Alberta, Canada | Cardiology |
| Sheila Hegde, MD | Brigham and Women's Hospital | Boston, USA | Cardiology |
| Yuri Kim, MD | Brigham and Women's Hospital | Boston, USA | Cardiology |
| Natalia Rost, MD | Massachusetts General Hospital | Boston, USA | Neurology |
| Aneesh Singhal, MD | Massachusetts General Hospital | Boston, USA | Neurology |
| Gregoire Le Gal, MD, PhD | University of Ottawa | Ottawa, Canada | Hematology |
| Deborah Siegal, MD, MSc | University of Ottawa | Ottawa, Canada | Hematology |
| Jean-Philippe Galanaud, MD, PhD | Sunnybrook Health Sciences Centre | Toronto, Canada | Hematology |

Appointments to the CEC will be made according to the required expertise and will accommodate the workflow of events collected in the trial and referred to the Committee for adjudication. Any such appointments would be made with the consent of the Chair of the CEC. CEC members will not participate directly in the recruitment of patients for the trial.

#### 3.0. RESPONSIBILITIES LIST

**Chairperson of the CEC:**

The Chairperson of the CEC, a Cardiologist, is primarily responsible for overseeing the operations of the CEC in accordance with the CEC Charter. The Chairperson of the CEC will:

- Take part in adjudication process as an Adjudicator and receive relevant trainings
- Provide oversight to the CEC
- Finalize and approve the CEC charter
- Liaise with the NHLBI (the trial funder), the study chair and co-chair, and the Data and Clinical Coordinating Centers of the trial.
- Provide the necessary training to CEC Members.
- Attend CEC Meetings
- Complete or delegate all administrative tasks related to the CEC Consensus Meetings
- Ensure consistent adjudication throughout the program
- Communicate issues related to the adjudication of clinical events to the Data Coordinating Center and study chair.

**CEC Adjudicator:**

CEC Adjudicators are selected based on clinical expertise. They must be board-certified or board-eligible in the appropriate subspecialty (e.g. cardiovascular medicine, hematology/oncology, neurology) or licensed according to local legislation. An Adjudicator is free of conflict of interest. One of adjudicators will also be the CEC Chairperson.

The CEC Adjudicators are responsible to:

- Complete self-training on the clinical program protocols, CEC Charter and Electronic Adjudication System usage.
- Independently adjudicate clinical events according to the definitions in the CEC Charter and consistent with the clinical study protocols, and in a uniform, unbiased, and confidential manner.
- Communicate procedural concerns to the CEC Chairperson and CEC Coordinator.
- Provide queries for additional information to the Data Coordinating Center.

**Data Coordinating Center:**

- Draft Endpoint Management Plan and Endpoint Reporting Manual
- Develop the appropriate case report forms to ascertain and track reported potential events, collect supporting source documents for each event dossier, and review the event dossier for quality, accuracy and completeness of data and source documentation to allow for adjudication.
- Review and approve clinical event dossiers to be submitted to the CEC adjudicators for adjudication.

- Receive queries for additional information from the CEC

#### 4.0. PROCEDURES OF THE CLINICAL ENDPOINT COMMITTEE

The CEC will consider for adjudication all cases of the following:

- **Deep venous thrombosis**
- **Pulmonary embolism**
- **Venous thromboembolism**
- **Arterial thromboembolism**
- **Myocardial infarction**
- **Ischemic stroke**
- **Major bleeding**

A detailed definition for each of these clinical endpoints can be found in Section 6.0.

5.1. Identification and Reporting of Clinical Events to the CEC
Clinical events are identified for adjudication in three different ways:

1. Possible events are identified based on investigator reports through electronic case record forms.

2. Possible events are identified by CEC adjudicators as they review case records sent for adjudication for other events. If CEC member identifies a possible as yet unreported event, he or she may manually add another event in the electronic adjudication system.

3. Listings of adverse events (AEs) will be periodically reviewed by staff at the Data Coordinating Center. If the reviewer is concerned that the AE under review is a possible clinical endpoint, s/he can query the individual site or local principle investigator regarding the possible event. The site may then report the AE as a potential endpoint for adjudication or may offer a rationale for declining to do so. If the site refuses to enter the AE as a potential endpoint, the DCC may manually enter the event in the electronic adjudication system for referral to and adjudication by the CEC.

4. Database programs that query for potential missed events based on algorithms.

5.1. Assembly of Clinical Dossiers for CEC Review
Clinical records of possible endpoints will be collected and complied into a dossier by the Data Coordinating Center. The specific components requested by the CEC prior to adjudicator review are including the Manual of Procedures/Endpoint Management Plan. Each clinical event will be assigned for review by 2 members of the CEC as noted in Section 5.3.

5.2. CEC Adjudication Process

Each complete clinical event dossier will be assigned to two CEC Adjudicators based on the event type. The cardiovascular events (e.g., venous thrombosis, pulmonary embolism, arterial embolism, myocardial infarction) will be assigned to 2 cardiologist CEC adjudicators who will independently review the complete clinical event dossier and render an adjudication decision in the electronic adjudication system. These events may also be assigned to hematologist/oncologist adjudicators. The strokes will be assigned to two neurologist CEC adjudicators who will independently review the complete clinical event dossier and render an adjudication decision in the electronic adjudication system. The bleeding events will be assigned to 2 adjudicators with expertise in bleeding (hematologist/oncologists or cardiologists) who will render an adjudication decision in the electronic adjudication system. All deaths will be assigned and adjudicated by two cardiologist adjudicators, aside from those deaths reported by the submitting investigator to be due to stroke. Deaths potentially related to stroke will generally be assigned and adjudicated by two neurologist adjudicators, although they may also be reviewed and adjudicated by cardiologist adjudicators.

If the adjudication result determined by each independent CEC adjudicator agrees, then the adjudication of that clinical event is considered to be complete and a final adjudication is rendered automatically. If the adjudication result determined by each independent CEC adjudicator disagrees or is discordant, then the CEC adjudicators will discuss the potential clinical event at a CEC meeting or teleconference. In most cases, adjudicators are able to reach consensus. If they are unable to arrive at consensus, a third CEC member will adjudicate the case and serve as the tiebreaker in the voting process. The case may also be reviewed by the CEC Chairperson, who will then render the final decision.

The CEC adjudicators may request additional information from the site in order to adjudicate the clinical event. The CEC Adjudicators' queries for additional information will be submitted in the endpoint adjudication system to the Data Coordinating Center for resolution. If, after all reasonable measures to collect additional documentation regarding a clinical event fail, then the CEC Adjudicators will review and render an adjudication based on best-available data.

**5.3. CEC Meetings**

CEC Meetings will be held to address any cases requiring Committee review. The CEC will document the Committee decision by submitting an updated decision in the EAS.

**5.4. Management of New Information Received after Adjudication**

CEC Adjudicators will be provided with required clinical data prior to clinical event adjudication as feasible. However, data can be updated by the site at any time and additional clinical data may become available subsequent to the adjudication of a clinical event. Any new information entered into the clinical database or submitted by the site will be uploaded into the endpoint adjudication system to be processed accordingly and reviewed by the CEC Chair. The CEC Chair will be responsible for determining if the new information could materially impact the adjudication results. If the Chair believes the new clinical information could impact the adjudication results, the clinical event dossier with the new information will be submitted for re-adjudication. The new data that prompted the second event review will be identified on the case report form included with the clinical source material in the event dossier. The standard review process will be followed. The CEC adjudicators will submit an updated decision, and the CEC adjudication form will subsequently be updated to reflect any changes to the initial adjudication.

##### 5.5. Archiving of Adjudication Documentation and Outcome Results

CEC adjudication outcome results will be stored in the electronic adjudication system and may be extracted periodically for scheduled safety analyses and project status reports.

At study closeout, all files stored on the database will be transferred to the Data Coordinating Center.

#### 5.0. CLINICAL EVENT DEFINITIONS

The CEC will consider for adjudication all cases of the following:

- **Deep venous thromboembolism**
- **Pulmonary embolism**
- **Arterial thromboembolism**
- **Myocardial infarction**
- **Stroke**
- **Major bleeding**
- **Death due to cardiovascular, non-cardiovascular, and undetermined cause**

##### 6.1. Deep Venous Thromboembolism

The diagnosis of <u>definite</u> symptomatic deep venous thromboembolism (DVT) requires symptoms of venous thromboembolism with at least one of the following:

- Abnormal compression ultrasound consistent with DVT or abnormal flow pattern or direct clot visualization in veins not amenable to compression.
- One or more new filling defects by venography, CT venography, or MR venography.
- Abnormal compression ultrasound where compression had been normal or, if known to be non-compressible, a substantial increase (≥4mm) in the diameter of a previously non-compressible venous segment.
- Point-of-care ultrasound (POCUS) performed by a provider and documenting DVT in a note.
- An extension of an intraluminal filling defect, or a new intraluminal filling defect, or an extension of non-visualization of veins in the presence of a sudden cut-off on venography.
- Proximal DVT is defined as clot at or proximal to the trifurcation of the popliteal vein (in the lower extremity) OR clot at or proximal to the axillary vein segment (in the upper extremity).
- Distal DVT is defined as clot distal to the trifurcation of the popliteal vein (in the lower extremities) OR clot at or distal to the brachial vein segment (in the upper extremities).

- Non-limb venous thrombosis includes thrombosis of the cerebral, portal, mesenteric, hepatic, gonadal, splenic, renal, or retinal veins, or thrombosis of the superior or inferior vena cava.

The diagnosis of presumed deep venous thromboembolism requires the following:

- In the absence of objective testing, high pre-test probability according to investigator assessment
    - OR adjudicator's gestalt
    - OR Wells score
- AND a treatment plan for DVT was initiated (initiation of anticoagulation, or escalation of anticoagulation dose, frequency, or duration).

##### 6.2. Pulmonary Embolism

The diagnosis of definite pulmonary embolism requires at least one of the following:

- New intraluminal filling defect at CT pulmonary angiography in a subsegmental or larger vessel.
- New intraluminal filling defect, or an extension of an existing defect, or a new sudden cut-off of vessels > 2.5 mm in diameter at pulmonary angiogram,
- Inconclusive CT pulmonary angiography, pulmonary angiography, or VQ scan evidence of a new or recurrent PE with demonstration of a new or recurrent DVT in the lower extremities by compression ultrasonography or venography. [2, 3]
- New clot or intraluminal filling defect noted in the right heart ("clot in transit") or the pulmonary vasculature at echocardiogram
- High probability (revised PIOPED criteria) on planar ventilation/perfusion (V/Q) scan OR positive PE on SPECT ventilation perfusion (V/Q) scan.
- Pulmonary embolism found at autopsy

The diagnosis of presumed pulmonary embolism requires the following:

Clinical signs and symptoms of pulmonary embolism, including but not limited to: dyspnea, cough, hypoxemia, tachycardia, appropriate electrocardiographic changes, or evidence of right heart strain on echocardiogram; AND chest CT or pulmonary angiography are unable to be performed AND therapeutic dose anticoagulation or fibrinolytic therapy is prescribed by a physician

##### 6.3. Arterial Thromboembolism

The diagnosis of arterial thromboembolism is defined as the following:

- A clinical history and presentation consistent with a sudden significant worsening of end organ or limb perfusion AND

EITHER

- Confirmation of arterial obstruction by imaging, hemodynamics, intraoperative findings, or pathological evaluation

OR

- Requirement for thrombolysis, thrombectomy, or urgent bypass.

Note that arterial thromboembolism includes both acute *in situ* thrombotic events and acute embolic events. Note that while ischemic stroke and myocardial infarction can be arterial thromboembolic events, those events will be adjudicated according to the separate standardized criteria included below.

##### 6.4. Myocardial Infarction

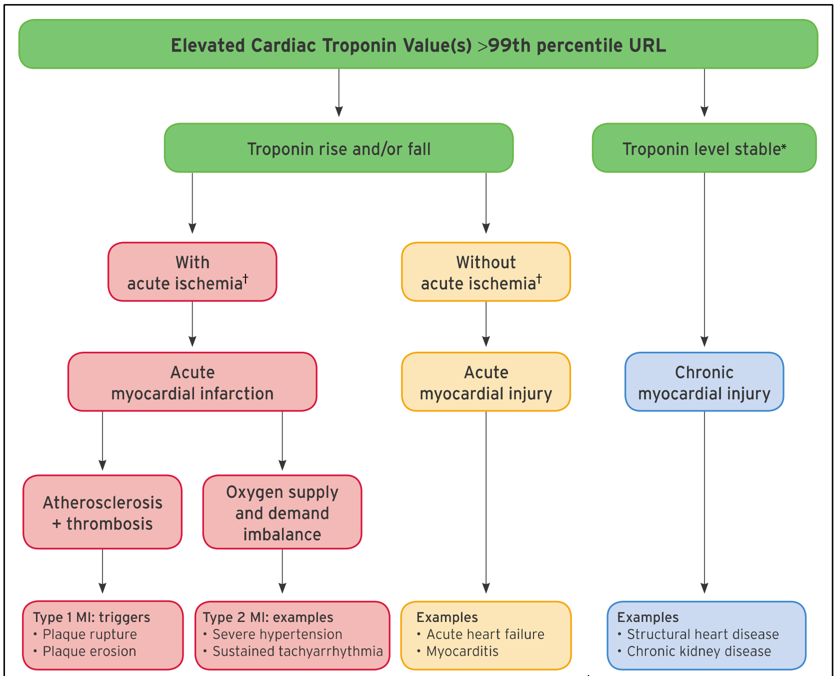

**Figure 1**. Model for interpreting myocardial injury.[1]

COVID-19 patients are well known to have elevations in cardiac troponin concentrations, and these elevations often do not represent arterial thrombosis and downstream myocardial ischemia. Therefore, the CEC will make an effort to distinguish true myocardial infarction from coronary artery obstruction, typically from atherothrombosis (usually considered a "type 1 myocardial infarction") from myocardial infarction due to demand ischemia (usually defined as a "type 2 myocardial infarction") and myocardial injury (an elevation in cardiac troponin typically without symptoms of chest pain or signs of arterial thrombosis). These definitions will be consistent with the 4th Universal Definition of Myocardial Infarction and will take into considerations suggestions made about classification of certain conditions as type 1 as compared to type 2 myocardial infarction.[1, 4] Regional coronary venous thrombosis with associated regional myocardial infarction has been reported in COVID. If this mechanism is documented, these will be considered an type 1 MI. The trial and CEC are focused on ascertaining and adjudicating cases of acute myocardial injury and acute myocardial infarction, and classifying those cases as described below. COVID also causes microvascular thrombi which are associated with patchy myocardial necrosis. These will be grouped with myocardial injury.

#### 2. UNIVERSAL DEFINITIONS OF MYOCARDIAL INJURY AND MYOCARDIAL INFARCTION: SUMMARY

| Universal definitions of myocardial injury and myocardial infarction |
|---|
| **Criteria for myocardial injury** |
| The term myocardial injury should be used when there is evidence of elevated cardiac troponin values (cTn) with at least 1 value above the 99th percentile upper reference limit (URL). The myocardial injury is considered acute if there is a rise and/or fall of cTn values. |
| **Criteria for acute myocardial infarction (types 1, 2 and 3 MI)** |
| The term acute myocardial infarction should be used when there is acute myocardial injury with clinical evidence of acute myocardial ischemia and with detection of a rise and/or fall of cTn values with at least 1 value above the 99th percentile URL and at least 1 of the following:<br>• Symptoms of myocardial ischemia;<br>• New ischemic ECG changes;<br>• Development of pathological Q waves;<br>• Imaging evidence of new loss of viable myocardium or new regional wall motion abnormality in a pattern consistent with an ischemic etiology;<br>• Identification of a coronary thrombus by angiography or autopsy (not for types 2 or 3 MIs).<br>Postmortem demonstration of acute atherothrombosis in the artery supplying the infarcted myocardium meets criteria for *type 1 MI*. Evidence of an imbalance between myocardial oxygen supply and demand unrelated to acute atherothrombosis meets criteria for *type 2 MI*. Cardiac death in patients with symptoms suggestive of myocardial ischemia and presumed new ischemic ECG changes before cTn values become available or abnormal meets criteria for *type 3 MI*. |
| **Criteria for coronary procedure–related myocardial infarction (types 4 and 5 MI)** |
| Percutaneous coronary intervention (PCI)–related MI is termed *type 4a MI*.<br>Coronary artery bypass grafting (CABG)–related MI is termed *type 5 MI*.<br>Coronary procedure–related MI ≤48 hours after the index procedure is arbitrarily defined by an elevation of cTn values >5 times for *type 4a MI* and >10 times for *type 5 MI* of the 99th percentile URL in patients with normal baseline values. Patients with elevated preprocedural cTn values, in whom the preprocedural cTn level are stable (≤20% variation) or falling, must meet the criteria for a >5 or >10 fold increase and manifest a change from the baseline value of >20%. In addition with at least 1 of the following:<br>• New ischemic ECG changes (this criterion is related to *type 4a MI* only);<br>• Development of new pathological Q waves;<br>• Imaging evidence of loss of viable myocardium that is presumed to be new and in a pattern consistent with an ischemic etiology;<br>• Angiographic findings consistent with a procedural flow-limiting complication such as coronary dissection, occlusion of a major epicardial artery or graft, side-branch occlusion-thrombus, disruption of collateral flow or distal embolization.<br>Isolated development of new pathological Q waves meets the *type 4a MI* or *type 5 MI* criteria with either revascularization procedure if cTn values are elevated and rising but less than the prespecified thresholds for PCI and CABG.<br>Other types of 4 MI include *type 4b MI* stent thrombosis and *type 4c MI* restenosis that both meet *type 1 MI* criteria.<br>Postmortem demonstration of a procedure-related thrombus meets the *type 4a MI* criteria or *type 4b MI* criteria if associated with a stent. |
| **Criteria for prior or silent/unrecognized myocardial infarction** |
| Any 1 of the following criteria meets the diagnosis for prior or silent/unrecognized MI:<br>• Abnormal Q waves with or without symptoms in the absence of nonischemic causes.<br>• Imaging evidence of loss of viable myocardium in a pattern consistent with ischemic etiology.<br>• Patho-anatomical findings of a prior MI. |

CABG indicates coronary artery bypass grafting; cTn, cardiac troponin; ECG, electrocardiogram; MI, myocardial infarction; PCI, percutaneous coronary intervention; URL, upper reference limit.

**Figure 2**. Table from the 4$^{th}$ Universal Definition of Myocardial Infarction summarizing the different definitions of myocardial injury and infarction.[1]

**Myocardial Injury:** The increasing sensitivity of cardiac troponin (cTn) assays means that ongoing myocardial injury is frequently detected. Myocardial injury is a prerequisite for myocardial infarction (MI), but as noted below, criteria in addition to myocardial injury are necessary to make the diagnosis of MI. Adjudicators must distinguish between acute myocardial injury that is not secondary to ischemia but may be due to other conditions (Table 2).

<u>Criteria for Myocardial Injury:</u> Detection of an elevated cTn value above the 99$^{th}$ percentile upper reference limit (URL) is defined as myocardial injury. The injury is considered acute if there is a rise and/or fall of cTn values.[1]

Criteria for Procedure Related Myocardial Injury: Cardiac procedural myocardial injury is arbitrary defined by increased in cTn values (>99th percentile URL) in patients with normal baseline values (<99th percentile URL) or a rise of cTn values >20% of the baseline value when it is the above the 99th percentile URL but is stable or falling.

| **Table 2.** Causes of non-ischemic myocardial injury[1, 5] |
|---|
| Heart failure |
| Myocarditis |
| Cardiomyopathy |
| Takotsubo syndrome |
| Coronary revascularization procedure |
| Cardiac procedure other than revascularization |
| Catheter ablation |
| Defibrillator shocks |
| Cardiac contusion |
| Sepsis, infectious disease |
| Chronic kidney disease |
| Stroke, subarachnoid hemorrhage |
| Pulmonary embolism, pulmonary hypertension |
| Infiltrative disease, e.g., amyloidosis, sarcoidosis |
| Chemotherapeutic agents |
| Critically ill patients |
| Strenuous exercise |
| Other |

**Myocardial Infarction Type 1**: Detection of rise and/or fall of cardiac biomarkers with at least one value above the 99th percentile of the upper reference limit (URL) together with evidence of myocardial ischemia with at least one of the following:

- Symptoms of ischemia
- New ischemic ECG changes indicative of new ischemia (new ST-T changes or new LBBB)*
- Development of pathological Q waves in the ECG**
- Imaging evidence of new loss of viable myocardium or new regional wall motion abnormality in a pattern consistent with an ischemic etiology
- Identification of a coronary thrombus by angiography including intracoronary imaging or by autopsy†
- *ECG manifestation of acute myocardial ischemia (in the absence of LVH and LBBB):
    - ST Elevation: New ST elevation at the J-point in two contiguous leads with the cut-point: ≥ 1 mm in all leads other than leads V2-V3, where the following cut-points apply: ≥2mm in men ≥40 years; ≥2.5 mm in men <40 years; or ≥ 1.5 mm in women regardless of age.
    - ST-depression and T-wave changes: New horizontal or down-sloping ST depression ≥ 0.5 mm in 2 contiguous leads and/or T inversion ≥ 1 mm in two contiguous leads with prominent R waves or R/S ratio >1.
- **Pathological Q waves:
    - Any Q-wave in leads V2-V3 >0.02 seconds or QS complex in leads V2-V3
    - Q-wave ≥ 0.03 seconds and ≥1 mm deep or QS complex in leads I, II, aVL, aVF, or V4-V6 in any 2 leads of a contiguous lead grouping (I, aVL;V1-V6; II, III, aVF; V7-V9).
    - R-wave ≥ 0.04s in V1–V2 and R/S ≥ 1 with a concordant positive T-wave in the absence of a conduction defect
- †Postmortem demonstration of an atherothrombus in the artery supplying the infarcted myocardium, or a macroscopically large circumscribed area of necrosis with or without intramyocardial hemorrhage meets the type 1 MI criteria regardless of cTn values.
- Consideration will be given to recent proposals to modify myocardial infarction type 1 to include coronary obstruction by spontaneous coronary artery dissection, coronary embolism, or coronary vasospasm or microvascular dysfunction.[4]

**Myocardial Infarction Type 2:** Detection of a rise and/or fall of cTn values with at least 1 value above the $99^{th}$ percentile URL, and evidence of imbalance between myocardial oxygen supply and demand unrelated to coronary atherothrombosis, requiring at least 1 of the following:

- Symptoms of acute myocardial ischemia
- New ischemic ECG changes
- Development of pathological Q waves;
- Imaging evidence of new loss of viable myocardium or new regional wall motion abnormality in a pattern consistent with ischemic etiology

**Myocardial Infarction Type 3:** Patients who suffer cardiac death, with symptoms suggestive of myocardial ischemia accompanied by presumed new ischemic ECG changes or ventricular fibrillation but die before blood samples for biomarkers can be obtained, or before increases in cardiac biomarkers can be identified, or MI is detected by autopsy examination.

**Myocardial infarction Type 4a and 4b (myocardial infarction associated with percutaneous coronary intervention):** Criteria for percutaneous coronary intervention (PCI)-related MI ≤48 hours after the index procedure are as follows: Coronary intervention-related MI is arbitrarily defined by an elevation of cTn values >5 times the 99the percentile URL in patients with normal baseline values. In patients with elevated preprocedural cTn in whom the cTn levels are stable (≤20% variation) or falling, the post procedure cTn must rise by >20%. However, the absolute procedural value must still be at least 5 times the $99^{th}$ percentile URL. In addition, 1 of the following elements is required:

- New ischemic ECG changes
- Development of new pathological Q waves; note that the development of new pathological Q waves meets the criteria for procedure-related MI if the cTn values are elevated and rising but <5 times the $99^{th}$ percentile URL.
- Imaging evidence of new loss of viable myocardium or new regional wall motion abnormality in a pattern consistent with an ischemic etiology
- Angiographic findings consistent with a procedural flow-limiting complication such as coronary dissection, occlusion of a major epicardial artery or side branch occlusion/thrombus, disruption of collateral flow, or distal embolization.
- Type 4a MI is an MI associated with PCI
- Type 4b MI is an MI associated with stent/scaffold thrombosis

**Myocardial Infarction Type 4c**: A type 4c MI is an MI associated with restenosis associated with prior PCI. Possible Type 4c MI is evaluated using the same criteria as Type 1 MI.

**Myocardial Infarction Type 5**: Criteria of coronary artery bypass grafting (CABG)-related MI ≤ 48 hours after the index procedure. CABG-related MI is arbitrarily defined as elevation of cTn values >10 times the $99^{th}$ percentile URL in patients with normal baseline cTn values. In patients with elevated preprocedural cTn in whom cTn are stale (≤20% variation) or falling, the post procedure cTn must rise by >20%. However, the absolute postprocedural values must still be >10 times the $99^{th}$ percentile URL. In addition, 1 of the following elements is required:

- Development of new pathological Q waves; note that the development of new pathological Q waves meets the criteria for procedure-related MI if the cTn values are elevated and rising but <10 times the $99^{th}$ percentile URL.
- Angiographically documented new graft occlusion or new native coronary artery occlusion;
- Imaging evidence of new loss of viable myocardium or new regional wall motion abnormality in a pattern consistent with an ischemic etiology

**Special or unusual circumstances:** Further guidance on distinguishing myocardial injury from myocardial infarction in the context of non-cardiac surgery, heart failure, myocarditis, Takotsubo syndrome, kidney disease, and in critically ill patients, and myocardial infarction nonobstructive coronary arteries is included in the $4^{th}$ Universal Definition of MI.[1]

##### 6.5. Stroke

The definition of stroke used here is drawn from the definitions proposed by Hicks et al. and Sacco et al.[6, 7] Stroke is defined as the acute onset of focal neurological dysfunction caused by brain, spinal cord, or retinal vascular injury as a result of hemorrhage or infarction. Note that while all strokes will be adjudicated, only ischemic stroke is part of the primary efficacy endpoint. Hemorrhagic stroke is considered a major bleed (see section 6.7).

A stroke is the acute onset of a new persistent neurological deficit attributed to an obstruction in cerebral blood flow with no apparent nonvascular cause (e.g. tumor, trauma, infection). Available neuroimaging studies will be considered to support the clinical impression and to determine if there is a demonstrable lesion compatible with an acute stroke. To the extent possible, all strokes will be classified as ischemic, hemorrhagic or unknown. While all types of strokes will be adjudicated by the CEC, only ischemic strokes will be included in the primary endpoint.

For the diagnosis of stroke, the following criteria should be fulfilled:

1. Rapid onset of a focal neurological deficit not related to any other known non-cerebrovascular process with at least one of the following:

    - Change in level of consciousness
    - Hemiplegia
    - Hemiparesis
    - Numbness or sensory loss affecting one side of the body
    - Dysphasia/aphasia
    - Hemianopia
    - Other new neurological sign/symptom(s) consistent with stroke
    - If the timing of onset is uncertain, a diagnosis of stroke may be made provided that there are no plausible non-stroke causes for the clinical presentation.

AND

**2.** Duration of a focal/global neurological deficit that is:

- EITHER ≥ 24 hours,
- OR < 24 hours if:
    - Resolution of symptoms is due to least one of the following interventions:
        1. Pharmacologic: intravenous or intraarterial thrombolysis
        2. Non-pharmacologic: (i.e. neuro-interventional procedure such as intracranial angioplasty)
- OR available MRI clearly documents a new hemorrhage or infarct
- OR available head CT clearly documents a new hemorrhage or infarct or excludes a mimic of stroke
- OR the neurological deficit results in death.

Ideally, at least one of should be present to confirm the diagnosis of stroke:

- Confirmation by neurology or neurosurgery specialist
- Brain imaging procedure (at least one of the following): CT scan, MRI scan, or cerebral vessel angiography
- Lumbar puncture (i.e. spinal fluid analysis diagnostic of intracranial hemorrhage)

If the acute focal signs represent a worsening of a previous deficit, these signs must persist for more than 24 hours and be accompanied by an appropriate new MRI or CT scan finding.

Strokes are sub-classified as follows:

*Ischemic (non-hemorrhagic):* An acute episode of focal cerebral, spinal, or retinal dysfunction caused by infarction of central nervous system tissue. Hemorrhage may be a consequence of ischemic stroke. In this situation, the stroke is an ischemic stroke with hemorrhagic transformation and not a hemorrhagic stroke but would also be listed as a major bleeding safety event.

*Hemorrhagic:* An acute episode of focal or global cerebral or spinal dysfunction caused by intraparenchymal, intraventricular, or subarachnoid hemorrhage. Hemorrhage in the brain is documented by neuroimaging or autopsy or lumbar puncture. Note that subdural hematomas are intracranial hemorrhagic events and not strokes.

*Undetermined:* An acute episode of focal or global neurological dysfunction caused by presumed brain, spinal cord, or retinal vascular injury as a result of hemorrhage or infarction but with insufficient information to allow categorization as either ischemic or hemorrhagic.

#### 6.6. Major Bleeding

Major bleeding is defined as acute clinically overt bleeding associated with one or more of the following (as per ISTH guidelines):[8, 9]

- Decrease in hemoglobin of 2 g/dL or more;
- Transfusion of 2 units or more of packed red blood cells;
- Bleeding that occurs in at least one of the following critical sites:
  - Intracranial
  - Intraspinal
  - Intraocular (within the corpus of the eye. A conjunctival bleed is not an intraocular bleed)
  - Pericardial
  - Intraarticular
  - Retroperitoneal
  - Intramuscular with compartment syndrome
- Bleeding that leads to death (primary cause of death or contributes directly to death)

#### 6.7. Definitions of Cardiovascular, Non-cardiovascular, and Undetermined Cases of Death

Definitions of Cardiovascular, Non-Cardiovascular, and Undetermined Cases of Death
The classifications for death are drawn from Hicks et al.[6] Death is classified into one of three categories: cardiovascular, non-cardiovascular, and undetermined cause of death. The intent is to identify one of these categories as the underlying cause of death. The key priority is differentiating between cardiovascular and non-cardiovascular causes of death. Death attribution can be difficult, particularly for sudden death, even when witnessed. While all deaths will be adjudicated by the CEC, only those deemed to be of cardiovascular cause will be included as part of the trial primary composite endpoint. Deaths of non-cardiovascular cause will not be included in the primary efficacy endpoint. Deaths of undetermined cause will not be part of the primary efficacy endpoint of PROMINENT.

**Cardiovascular death** can be due to acute myocardial infarction (MI), sudden cardiac death, heart failure, stroke, pulmonary embolism, a cardiovascular procedure, cardiovascular hemorrhage, or other cardiovascular cause.

**Cardiovascular death due to acute MI:** Death by any cardiovascular mechanism (arrhythmia, sudden death, heart failure, stroke, pulmonary embolus, PAD) within 30 days after an acute MI, related to the immediate consequences of the MI, such as progressive heart failure or recalcitrant arrhythmia. While there may be assessable (attributable) mechanisms of cardiovascular death during this time period, for simplicity, if the cardiovascular death occurs within 30 days of an acute MI, it will be considered a death due to MI.

Note: Acute MI should be verified to the extent possible by the diagnostic criteria outlined for acute MI or by autopsy findings showing recent MI or recent coronary thrombosis. Death resulting from a procedure to treat an MI (PCI or CABG), or to treat a complication resulting from MI, should also be considered death due to acute MI. Death resulting from an elective coronary procedure to treat myocardial ischemia (i.e., chronic stable angina) or death due to an

MI that occurs as a direct consequence of a cardiovascular investigation/ procedure/operation should be considered as a death due to a cardiovascular procedure.

**Cardiovascular death due to sudden cardiac death**: Death that occurs unexpectedly

and not within 30 days of an acute MI. Sudden cardiac death includes the following scenarios:

- Death witnessed and occurring without new or worsening symptoms
- Death witnessed within 60 min of the onset of new or worsening cardiac symptoms unless the symptoms suggest acute MI
- Death witnessed and attributed to an identified arrhythmia (e.g., captured on an electrocardiographic recording, witnessed on a monitor, or unwitnessed but found on ICD review)
- Death after unsuccessful resuscitation from cardiac arrest (e.g., ICD unresponsive sudden cardiac death, pulseless electrical activity arrest)
- Death after successful resuscitation from cardiac arrest and without identification of a specific cardiac or non-cardiac etiology
- Unwitnessed death in a subject seen alive and clinically stable ≤24 h before being found dead without any evidence supporting a specific non- cardiovascular cause of death (information about the patient's clinical status preceding death should be provided if available)

Unless additional information suggests an alternate specific cause of death (e.g., Death due to Other Cardiovascular Causes), if a patient is seen alive ≤24 h before being found dead, sudden cardiac death should be recorded. For patients who were not observed alive within 24 h of death, undetermined cause of death should be recorded (e.g., a subject found dead in bed but who had not been seen by family members for >24 h).

**Cardiovascular death due to heart failure (HF):** Death associated with clinically worsening symptoms and/or signs of HF, regardless of HF etiology. Note: Deaths due to HF can have various etiologies, including single or recurrent MIs, ischemic or nonischemic cardiomyopathy, hypertension, or valvular disease.

**Death due to stroke:** Death after a stroke that is either a direct consequence of the stroke or a complication of the stroke. Note: acute stroke should be verified to the extent possible by the diagnostic criteria outlined for stroke.

**Cardiovascular death due to cardiovascular procedure:** Death caused by the immediate complication(s) of a cardiovascular procedure.

**Cardiovascular death due to cardiovascular hemorrhage:** Death related to hemorrhage such as a non-stroke intracranial hemorrhage (e.g., subdural hematoma) nonprocedural or non-traumatic vascular rupture (e.g., aortic aneurysm), or hemorrhage causing cardiac tamponade.

**Cardiovascular death due to other cardiovascular causes:** Cardiovascular death not included in the above categories with specific, known cause (e.g., PE, PAD).

**Definition of Non-cardiovascular Death:** When death is due to a non-cardiovascular cause, a cardiovascular cause of death is excluded.

- Pulmonary (excludes malignancy)
- Renal
- Gastrointestinal (disease of the esophagus, stomach, or intestines (excludes malignancy)
- Hepatobiliary (disease of the liver, gall bladder, or biliary ducts (excludes malignancy)
- Pancreatic (disease of the pancreas (excludes malignancy)
- Infection (including sepsis)
- Inflammatory/immune (death attributable to an inflammatory or immune-mediated disease or process, including systemic inflammatory response syndrome (SIRS), immunological, and autoimmune disease and disorders.  Includes anaphylaxis from environmental allergies)
- Hemorrhage (bleeding that is not considered cardiovascular hemorrhage or stroke
- Non-CV procedure or surgery (death caused by the immediate complications of a non-cardiovascular procedure or surgery)
- Trauma (death attributable to trauma.  Includes homicide)
- Suicide
- Nonprescription drug reaction or overdose
- Prescription drug reaction or overdose (includes anaphylaxis)
- Neurological (excludes malignancy, as well as death from ischemic stroke, hemorrhagic stroke, or undetermined cause of stroke, or cardiovascular hemorrhage of central nervous system)
- Malignancy (leukemia, lymphoma, or other malignancy)
- Other (death attributable to a cause other than those listed in this classification; specify organ system)

**Undetermined cause of death:** Causality may be difficult to determine if information available from the time of death is minimal or nonexistent. Deaths of undetermined cause will not be considered presumed cardiovascular, and thus will not be part of the primary efficacy endpoint of PROMINENT.

#### 8.0. EXPERT SIGNATURE PAGE

Clinical Events Committee Charter
CEC Charter Version 1.0

Name: ____________________________________________________________

Address: __________________________________________________________

I hereby approve the Charter of the Clinical Events Committee.

Signature: ________________________________________________________

Date: ___________________________________
