## Supplementary Appendix Anticoagulation mpRCT Severe State for "Therapeutic Anticoagulation in Critically Ill Patients with Covid-19 – Preliminary Report"

#### Multi-Platform Randomized Controlled Trial (mpRCT) Therapeutic anticoagulation in patients with severe Covid-19

REMAP-CAP

ACTIV-4
Acute Inpatient Anti-Thrombotic Study

ATTACC

### Section 1 - mpRCT Investigators and Collaborators

#### 1.1 Data Safety and Monitoring Board Members

##### 1.1.1 REMAP-CAP

Victoria Manax, MD (Chair); Jason Connor, PhD (Deputy Chair); Julian Bion, MD; Simon Gates, PhD; John Reynolds, PhD; Tom van der Poll, PhD

##### 1.1.2 ATTACC

Jason Connor, PhD (Chair); Victoria Manax, MD (Deputy Chair); Julian Bion, MD; Simon Gates, PhD; John Reynolds, PhD; James Douketis, MD; Damon Scales, MD, PhD, Peter Nickerson, MD (sponsor representative)

##### 1.1.3 ACTIV-4a

Richard C. Becker, MD (Chair); Gregory del Zoppo, MD; Peter Henke, MD; Richard Holubkov, PhD; Kim Kerr, MD; Agnes Lee, MD; Fedor Lurie, MD, PhD; Sara K. Vesely, PhD

#### 1.2 REMAP-CAP

**International Trial Steering Committee:**

Farah Al-Beidh, Derek Angus, Djillali Annane, Yaseen Arabi, Abi Beane, Wilma van Bentum-Puijk, Scott Berry, Zahra Bhimani, Marc Bonten, Charlotte Bradbury, Frank Brunkhorst, Meredith Buxton, Allen Cheng, Lennie Derde, Lise Estcourt, Herman Goossens, Anthony Gordon, Cameron Green, Rashan Haniffa, Francois Lamontagne, Patrick Lawler, Kelsey Linstrum, Edward Litton, John Marshall, Colin McArthur, Daniel McAuley, Shay McGuinness, Bryan McVerry, Stephanie Montgomery, Paul Mouncey, Srinivas Murthy, Alistair Nichol, Rachael Parke, Jane Parker, Kathryn Rowan, Marlene Santos, Christopher Seymour, Alexis Turgeon, Anne Turner, Frank van de Veerdonk, Steve Webb (Chair), Ryan Zarychanski

**Regional Management Committees**

***Australia and New Zealand***

Yaseen Arabi, Lewis Campbell, Allen Cheng, Lennie Derde, Andrew Forbes, David Gattas, Cameron Green, Stephane Heritier, Peter Kruger, Edward Litton, Colin McArthur (Deputy Executive Director), Shay McGuinness (Chair), Alistair Nichol, Rachael Parke, Jane Parker, Sandra Peake, Jeffrey Presneill, Ian Seppelt, Tony Trapani, Anne Turner, Steve Webb (Executive Director), Paul Young

***Canadian Regional Management Committee***

Zahra Bhimani, Brian Cuthbertson, Rob Fowler, Francois Lamontagne, John Marshall (Executive Director), Venika Manoharan, Srinivas Murthy (Deputy Executive Director), Marlene Santos, Alexis Turgeon, Ryan Zarychanski

***Critical Care Asia (CCA) Regional Management Committee***

Diptesh Aryal, Abi Beane (Chair), Arjen M Dondrop, Cameron Green, Rashan Haniffa (Executive Director), Madiha Hashmi, Deva Jayakumar, John Marshall, Colin McArthur, Srinivas Murthy, Timo Tolppa, Vanessa Singh, Steve Webb

***European Regional Management Committee***

Farah Al-Beidh, Derek Angus, Djillali Annane, Wilma van Bentum-Puijk, Scott Berry, Marc Bonten (Co-Executive Director), Nicole Brillinger, Frank Brunkhorst, Maurizio Cecconi, Lennie Derde (Co-Executive Director and Chair), Stephan Ermann, Bruno Francois, Herman Goossens, Anthony Gordon, Cameron Green, Sebastiaan Hullegie, Rene Markgraff, Colin McArthur, Paul Mouncey, Alistair Nichol, Mathias Pletz, Pedro Povoa, Gernot Rohde, Kathryn Rowan, Lorraine Parker, Irma Scheepstra-Beukers, Steve Webb

***United States Regional Management Committee***
Brian Alexander, Derek Angus (Executive Director), Kim Basile, Meredith Buxton (Chair), Timothy Girard, Christopher Horvat, David Huang, Kelsey Linstrum, Florian Mayr, Bryan McVerry, Stephanie Montgomery, Christopher Seymour

**Regional Coordinating Centers**
***Australia, CCA region, and Saudi Arabia:*** The Australia and New Zealand Intensive Care Research Centre (ANZIC-RC), Monash University
***Canada:*** St. Michael's Hospital, Unity Health Toronto
***Europe:*** University Medical Center Utrecht (UMCU)
***New Zealand:*** The Medical Research Institute of New Zealand (MRINZ)
***United States:*** Global Coalition for Adaptive Research (GCAR), and University of Pittsburgh Medical Center
**CRIT Care Asia (CCA):** NICS MORU.

**Domain-Specific Working Groups**
***Antibiotic and Macrolide Duration Domain-Specific Working Group***
Richard Beasley, Marc Bonten, Allen Cheng (Chair), Nick Daneman, Lennie Derde, Robert Fowler, David Gattas, Anthony Gordon, Cameron Green, Peter Kruger, Colin McArthur, Steve McGloughlin, Susan Morpeth, Srinivas Murthy, Alistair Nichol, Mathias Pletz, David Paterson, Gernot Rohde, Steve Webb

***Corticosteroid Domain-Specific Working Group***
Derek Angus (Chair), Wilma van Bentum-Puijk, Lennie Derde, Anthony Gordon, Sebastiaan Hullegie, Peter Kruger, Edward Litton, John Marshall, Colin McArthur, Srinivas Murthy, Alistair Nichol, Bala Venkatesh, Steve Webb

***Influenza Antiviral Domain-Specific Working Group***
Derek Angus, Scott Berry, Marc Bonten, Allen Cheng, Lennie Derde, Herman Goossens, Sebastiaan Hullegie, Menno de Jong, John Marshall, Colin McArthur, Srinivas Murthy (Chair), Tim Uyeki, Steve Webb

***COVID-19 Antiviral Domain-Specific Working Group***
Derek Angus, Yaseen Arabi (Chair), Kenneth Baillie, Richard Beasley, Scott Berry, Marc Bonten, Allen Cheng, Menno de Jong, Lennie Derde, Eamon Duffy, Rob Fowler, Herman Goossens, Anthony Gordon, Cameron Green, Thomas Hills, Colin McArthur, Susan Morpeth, Srinivas Murthy, Alistair Nichol, Katrina Orr, Rachael Parke, Jane Parker, Asad Patanwala, Kathryn Rowan, Steve Tong, Tim Uyeki, Frank van de Veerdonk, Steve Webb

**COVID-19 Immune Modulation Domain-Specific Working Group**
Derek Angus, Yaseen Arabi, Kenneth Baillie, Richard Beasley, Scott Berry, Marc Bonten, Frank Brunkhorst, Allen Cheng, Nichola Cooper, Olaf Cremer, Menno de Jong, Lennie Derde (Chair), Eamon Duffy, James Galea, Herman Goossens, Anthony Gordon, Cameron Green, Thomas Hills, Andrew King, Helen Leavis, John Marshall, Florian Mayr, Colin McArthur, Bryan McVerry, Susan Morpeth, Srinivas Murthy, Mihai Netea, Alistair Nichol, Kayode Ogungbenro, Katrina Orr, Jane Parker, Asad Patawala, Ville Pettilä (Deputy Chair), Emma Rademaker, Kathryn Rowan, Manoj Saxena, Christopher Seymour, Wendy Sligl, Steven Tong, Tim Uyeki, Frank van de Veerdonk, Steve Webb, Taryn Youngstein

**COVID-19 Immune Modulation -2 Domain-Specific Working Group**
Derek Angus, Scott Berry, Lennie Derde, Cameron Green, David Huang, Florian Mayr, Bryan McVerry, Stephanie Montgomery, Christopher W. Seymour (Chair), Steve Webb

***Therapeutic Anticoagulation Domain-Specific Working Group***
Derek Angus, Diptesh Aryal, Scott Berry, Shailesh Bihari, Charlotte Bradbury, Marc Carrier, Dean Fergusson, Robert Fowler, Ewan Goligher (Deputy Chair), Anthony Gordon, Christopher Horvat, David Huang, Beverley Hunt, Devachandran Jayakumar, Anand Kumar, Mike Laffan, Patrick Lawler, Sylvain Lother, Colin McArthur, Bryan McVerry, John Marshall, Saskia Middeldorp, Zoe McQuilten, Matthew Neal, Alistair Nichol, Christopher Seymour, Roger Schutgens, Simon Stanworth, Alexis Turgeon, Steve Webb, Ryan Zarychanski (Chair)

***Vitamin C Domain-Specific Working Group***
Neill Adhikari (Chair), Derek Angus, Djillali Annane, Matthew Anstey, Yaseen Arabi, Scott Berry, Emily Brant, Angelique de Man, Lennie Derde, Anthony Gordon, Cameron Green, David Huang, Francois Lamonagne (Chair), Edward Litton, John Marshall, Marie-Helene Masse, Colin McArthur, Shay McGuinness, Paul Mouncey, Srinivas Murthy, Rachael Parke, Alistair Nichol, Tony Trapani, Andrew Udy, Steve Webb

***COVID-19 Immunoglobulin Domain-Specific Working Group***
Derek Angus, Donald Arnold, Phillipe Begin, Scott Berry, Richard Charlewood, Michael Chasse, Mark Coyne, Jamie Cooper, James Daly, Lise Estcourt (Chair, UK lead), Dean Fergusson, Anthony Gordon, Iain Gosbell, Heli Harvala-Simmonds, Tom Hills (New Zealand lead), Christopher Horvat, David Huang, Sheila MacLennan, John Marshall, Colin McArthur (New Zealand lead), Bryan McVerry (USA lead), David Menon, Susan Morpeth, Paul Mouncey, Srinivas Murthy, John McDyer, Zoe McQuilten (Australia lead), Alistair Nichol (Ireland lead), Nicole Pridee, David Roberts, Kathryn Rowan, Christopher Seymour, Manu Shankar-Hari (UK lead), Helen Thomas, Alan Tinmouth, Darrell Triulzi, Alexis Turgeon (Canada lead), Tim Walsh, Steve Webb, Erica Wood, Ryan Zarychanski (Canada lead)

***Simvastatin Domain-Specific Working Group***
Derek Angus, Yaseen Arabi, Abi Beane, Carolyn Calfee, Anthony Gordon, Cameron Green, Rashan Haniffa, Deva Jayakumar, Peter Kruger, Patrick Lawler, Edward Litton, Colin McArthur, Daniel McAuley (Chair), Bryan McVerry, Matthew Neal, Alistair Nichol, Cecilia O'Kane, Murali Shyamsundar, Pratik Sinha, Taylor Thompson, Steve Webb, Ian Young

***Antiplatelet Domain-Specific Working Group***
Derek Angus, Scott Berry, Shailesh Bihari, Charlotte Bradbury (Chair), Marc Carrier, Timothy Girard, Ewan Goligher, Anthony Gordon, Ghady Haidar, Christopher Horvat, David Huang, Beverley Hunt, Anand Kumar, Patrick Lawler, Patrick Lawless, Colin McArthur, Bryan McVerry, John Marshall, Zoe McQuilten, Matthew Neal, Alistair Nichol, Christopher Seymour, Simon Stanworth, Steve Webb, Alexandra Weissman, Ryan Zarychanski

***Mechanical Ventilation Domain***
Derek Angus, Wilma van Bentum-Puijk, Lewis Campbell, Lennie Derde, Niall Ferguson, Timothy Girard, Ewan Goligher, Anthony Gordon, Cameron Green, Carol Hodgson, Peter Kruger, John Laffey, Edward Litton, John Marshall, Colin McArthur, Daniel McAuley, Shay McGuinness, Alistair Nichol (Chair) Neil Orford, Kathryn Rowan, Ary Neto, Steve Webb

***ACE-2 RAAS Domain***
Rebecca Baron, Lennie Derde, Slava Epelman, Claudia Frankfurter, David Gattas, Frank Gommans, Anthony Gordon, Rashan Haniffa, David Huang, Edy Kim, Francois Lamontagne, Patrick Lawler (Chair), David Leaf, John Marshall, Colin McArthur, Bryan McVerry, Daniel McAuley, Muthiah Vaduganathan, Roland van Kimmenade, Frank van de Veerdonk, Steve Webb

**Statistical Analysis Committee**
Michelle Detry, PhD, Mark Fitzgerald, PhD, Roger Lewis, MD, PhD (Chair), Anna McGlothlin, PhD, Ashish Sanil, PhD, Christina Saunders, PhD

**Statistical Design Team**
Lindsay Berry, PhD, Scott Berry, PhD, Elizabeth Lorenzi, PhD

**Data Coordinating Team**
Adrian Buzgau, Cameron Green, Alisa Higgins, MPH, PhD

**Project Management**
***Australia and Saudi Arabia:*** Jane Parker, Vanessa Singh, Claire Zammit
***Canada:*** Zahra Bhimani, Marlene Santos
***CCA:*** Abi Beane, Rashan Haniffa, Timo Tolppa
***Europe:*** Wilma van Bentum-Puijk, Lorraine Parker, Irma Scheepstra-Beukers, Erika Groeneveld, Svenja Peters, Clementina Okundaye, Denise van Hout, Albertine Smit, Linda Rikkert, Sara Bari, Kik Raymakers, Marion Kwakkenbos-Craanen, Sophie Post, Gerwin Schreuder.
***Germany:*** Nicole Brillinger, Rene Markgraf
***Global:*** Cameron Green
***Ireland:*** Kate Ainscough, Kathy Brickell, Peter Doran
***New Zealand:*** Anne Turner
***United Kingdom:*** Farah Al-Beidh, Aisha Anjum, Janis-Best Lane, Elizabeth Fagbodun, Lorna Miller, Paul Mouncey, Karen Parry-Billings, Sam Peters, Alvin Richards-Belle, Michelle Saull, Stefan Sprinckmoller, Daisy Wiley

***United States of America:*** Kim Basile, Meredith Buxton, Kelsey Linstrum, Stephanie Montgomery, Renee Wunderley

**Database Providers**

***Research Online:*** Marloes van Beurden, Evelien Effelaar, Joost Schotsman,
***Spinnaker Software:*** Craig Boyd, Cain Harland, Audrey Shearer, Jess Wren
***University of Pittsburgh Medical Center:*** Giles Clermont, William Garrard, Christopher Horvat, Kyle Kalchthaler, Andrew King, Daniel Ricketts, Salim Malakoutis, Oscar Marroquin, Edvin Music, Kevin Quinn
***NICS MORU: on behalf of CCA*****:** Udara Attanayaka, Abi Beane, Sri Darshana, Rashan Haniffa, Pramodya Ishani, Issrah Jawad, Upulee Pabasara, Timo Tolppa, Ishara Udayanga.

**Clinical Trials Groups**

The REMAP-CAP platform is supported by the Australian and New Zealand Intensive Care Society Clinical Trials Group, the Canadian Critical Care Clinical Trials Group, the Irish Critical Care Clinical Trials Network, the UK Critical Care Research Group and the International Forum of Acute Care Trialists.

REMAP-CAP was supported in the UK by the NIHR Clinical Research Network and we acknowledge the contribution of Kate Gilmour, BSc (Hons), Karen Pearson, MSc, Chris Siewerski, MSc, Sally-Anne Hurford, MSc, Emma Marsh, FdSC, Debbie Campbell, Penny Williams, MSc, Kim Shirley, LLB Hons, Meg Logan, NVQ, Jane Hanson, Becky Dilley, BSc, Louise Phillips, CIM, Anne Oliver, MSc, Mihaela Sutu, MSc, Sheenagh Murphy, PGDip, Latha Aravindan, PhD, Joanne Collins, MRes, Holly Monaghan, Adam Unsworth, NVQ, Seonaid Beddows, MSc, Laura Ann Dawson, LLM, Sarah Dyas, Adeeba Asghar, MSc, Kate Donaldson, BA, Tabitha Skinner, BSc, Nhlanhla Mguni, BSc (Hons), Natasha Muzengi, BSc, Ji Luo, PhD, Joanna O'Reilly, BSc (Hons), Chris Levett, MSc, Alison Potter, David Porter, PhD, Teresa Lockett, MSc, Jazz Bartholomew, MSc, Clare Rook, MSc, Rebecca McKay, Hannah Williams, MSc, Alistair Hall, FRCP, Hilary Campbell, BSc (Hons), Holly Speight, BA, Sandra Halden, Susan Harrison, Mobeena Naz, Kaatje Lomme, MA, Paula Sharratt, MSc, Johnathan Sheffield, FRCP, William Van't Hoff, FRCPCH, James D Williamson, PhD, Alex Barnard, BSc (Hons), Catherine Birch, BA, Morwenna Brend, PhD, Emma Chambers, PhD, Sarah Crawshaw, PhD, Chelsea Drake, BSc, Hayley Duckles-Leech, PhD, Justin Graham, BSc (Hons), Heather Harper, BSc (Hons), Stephen Lock, PGCert, Nicola McMillan, PhD, Cliodhna O'Flaherty, Eleanor OKell, PhD, Amber Hayes, Sally Sam, BSc, Heather Slade, MSc, Susan Walker, PhD, Karen Wilding, MRes, Jayne Goodwin, Helen Hodgson, Yvette Ellis, Dawn Williamson, Madeleine Bayne, MSc, Shane Jackson, Rahim Byrne, Sonia McKenna, Alsion Clinton, NIHR Urgent Public Health Group: https://www.nihr.ac.uk/documents/urgent-public-health-group-members/24638#Members

REMAP-CAP was supported in France by the CRICS-TRIGGERSEP network

REMAP-CAP was supported in Ireland by the Irish Critical Care Clinical Trials Network and we acknowledge the contribution of Kate Ainscough, Kathy Brickell and Peter Doran.

REMAP-CAP was supported in the Netherlands by the Research Collaboration Critical Care the Netherlands (RCC-Net).

REMAP-CAP was supported in Canada but the Canadian Institutes of Health Research, St. Michael's Unity Health, and the Canadian Critical Care Trials Group

**Site Investigators and Research Coordinators**

***Australia:***

*The Alfred Hospital*: Andrew Udy, PhD, Phoebe McCracken, MPH, Meredith Young, MPH, Jasmin Board, MPH, Emma Martin, BPharmSc;

*Ballarat Health Services*: Khaled El-Khawas, FCICM, Angus Richardson, FCICM, Dianne Hill, BN GradCert(CritCare), Robert J Commons, PhD, Hussam Abdelkharim, FCICM;

*Bendigo Hospital*: Cameron Knott, MBBS(Hons) GDipClinUS MClinEd FCICM FRACP, Julie Smith, GDipN(CritCare), Catherine Boschert, PGDip;

*Caboolture Hospital*: Julia Affleck, MBiotech, Yogesh Apte, FCICM, Umesh Subbanna, FCICM, Roland Bartholdy, FCICM, Thuy Frakking, PhD;

*Campbelltown Hospital*: Karuna Keat, MB BS(Hons), Deepak Bhonagiri, MD, Ritesh Sanghavi, MBBS, Jodie Nema, B.App.Sc.(Ag), Megan Ford, BSc;

*Canberra Hospital*: Harshel G. Parikh, FCICM, Bronwyn Avard, MLMEd, Mary Nourse, PGCert(CritCare);

*Concord Repatriation General Hospital*: Winston Cheung, MBChB, Mark Kol, MBBS, Helen Wong, RN, Asim Shah, MBChB, Atul Wagh, MBBS;

*Eastern Health (Box Hill, Maroondah & Angliss Hospitals)*: Joanna Simpson, FCICM FANZCA, Graeme Duke, FCICM, Peter Chan, FCICM, Brittney Carter, PGCert(CritCare), Stephanie Hunter, MN;

*Flinders Medical Centre*: Shailesh Bihari, PhD, Russell D Laver, FCICM, Tapaswi Shrestha, MN, Xia Jin, MN;

*Fiona Stanley Hospital*: Edward Litton, FCICM, Adrian Regli, FCICM, Susan Pellicano, PGCert, Annamaria Palermo, BA, Ege Eroglu, BSc(Hons);

*Footscray Hospital*: Craig French, MBBS, Samantha Bates, RN GDipCritCare, Miriam Towns, MPH, Yang Yang, MBBS, Forbes McGain, PhD;

*Gold Coast University Hospital*: James McCullough, FCICM, Mandy Tallott, MN;

*John Hunter Hospital*: Nikhil Kumar, MBBS, Rakshit Panwar, FCICM, Gail Brinkerhoff, PGCert, Cassandra Koppen, BPharm, Federica Cazzola, MBBS;

*Launceston General Hospital*: Matthew Brain, FCICM, FRACP, DDU; Sarah Mineall, MN;

*Lyell McEwin Hospital*: Roy Fischer, FCICM, Vishwanath Biradar, FCICM, Natalie Soar, BSN;

*Logan Hospital*: Hayden White, PhD, Kristen Estensen, MBBS, Lynette Morrison, PGCert(CritCare), Joanne Sutton, RN, Melanie Cooper, PGCert(CritCare);

*Monash Health (Monash Medical Centre, Dandenong Hospital & Casey Hospital):* Yahya Shehabi, PhD, Wisam Al-Bassam, MBChB, Amanda Hulley, BSc; Umesh Kadam, MSc, Kushaharan Sathianathan, MSc;

*Nepean Hospital*: Ian Seppelt, PhD, Christina Whitehead, MN, Julie Lowrey, BN, Rebecca Gresham, BN, Kristy Masters, BN;

*Princess Alexandra Hospital*: Peter Kruger, PhD, James Walsham, FCICM, Mr Jason Meyer, BN, Meg Harward, MN, Ellen Venz, PGCert(CritCare);

*The Prince Charles Hospital*: Kara Brady, MPharm, Cassandra Vale, GDipClinPharm, Kiran Shekar, PhD, Jayshree Lavana, FCICM, Dinesh Parmar, FCICM;

*The Queen Elizabeth Hospital*: Sandra Peake, PhD, Patricia Williams, BN, Catherine Kurenda, RN;

*Rockhampton Hospital*: Helen Miles, MBBS MPH FCICM, Antony Attokaran, MBBS FCICM FRACP;

*Royal Adelaide Hospital*: Samuel Gluck, MD, Stephanie O'Connor, MNSc, Marianne Chapman, PhD, Kathleen Glasby, CCRN;
*Royal Darwin Hospital*: Lewis Campbell, FCICM, Kirsty Smyth, PGCert(CritCare), Margaret Phillips, MN;
*Royal Melbourne Hospital*: Jeffrey Presneill, PhD, Deborah Barge, CCRN, Kathleen Byrne, MNSc, Alana Driscoll GDipN(CritCare), Louise Fortune, MNSc;
*Royal North Shore Hospital*: Pierre Janin, MD, Elizabeth Yarad, MN, Frances Bass, MSc, Naomi Hammond, PhD, Anne O'Connor, RN;
*Royal Perth Hospital*: Sharon Waterson, PGCert(CritCare), Steve Webb, PhD, Robert McNamara, BMBS;
*Royal Prince Alfred Hospital*: David Gattas, MMed(ClinEpid), Heidi Buhr, MScMed(ClinEpid), Jennifer Coles, GDipN(CritCare);
*Sir Charles Gardiner Hospital*: Sacha Schweikert, FCICM, Bradley Wibrow, FCICM, Matthew Anstey, FCICM, Rashmi Rauniyar, MPH;
*St George Hospital*: Kush Deshpande, FCICM, Pam Konecny, FRACP, Jennene Miller, BN, Adeline Kintono, BN, Raymond Tung B.Pharm, MPS, MSHP
*St. John of God Midland Public and Private Hospitals*: Ed Fysh, PhD, Ashlish Dawda, MD, Bhaumik Mevavala, MHA;
*St. John of God Hospital, Murdoch*: Annamaria Palermo, BA, Adrian Regli, FCICM, Bart De Keulenaer, FCICM;
*St. John of God Hospital, Subiaco*: Ed Litton, PhD, Janet Ferrier, BSc;
*St. Vincent's Hospital (NSW)*: Priya Nair, PhD, Hergen Buscher, FCICM, Claire Reynolds, MClinNurse, Sally Newman, PGCert(CritCare);
*St. Vincent's Hospital (VIC)*: John Santamaria, MDBS, Leanne Barbazza, PGCert(CritCare), Jennifer Homes, PGCert(CritCare), Roger Smith, MPH;
*Sunshine Coast University Hospital*: Peter Garrett, FCICM, Lauren Murray, MSc, Jane Brailsford, PGCert(CritCare), Loretta Forbes, PGCert(CritCare), Teena Maguire, BA;
*Sunshine Hospital*: Craig French, MBBS, Gerard Fennessy, MBBS, John Mulder, MBBS, Rebecca Morgan, RN PGCert(CritCare), Rebecca McEldrew, RN PGCert(CritCare);
*The Sutherland Hospital*: Anas Naeem, MClinEd, Laura Fagan, GDipN(CritCare), Emily Ryan, PGCert(CritCare);
*Toowoomba Hospital*: Vasanth Mariappa, FCICM, Judith Smith, PGCert;
*University Hospital Geelong*: Scott Simpson, MBBS, Matthew Maiden, PhD, Allison Bone, GDipN(CritCare), , Michelle Horton, MN, Tania Salerno, PGCert;
*Wollongong Hospital*: Martin Sterba, PhD, Wenli Geng, MN;

***Belgium:***
*Ghent University Hospital*: Pieter Depuydt, PhD, Jan De Waele, PhD, Liesbet De Bus, PhD, Jan Fierens, MD, Stephanie Bracke, BSc, Joris Vermassen, MD, Daisy Vermeiren, BSc;

***Canada*:**
*Brantford General Hospital:* Brenda Reeve, MD, William Dechert, MSc;
*Centre de recherche de l'Institut universitaire de cardiologie et de pneumologie de Québec*: Francois Lellouche, Patricia Lizotte
*Centre Hospitalier de l'Universite de Montreal:* Michaël Chassé, PhD, François Martin Carrier, MSc, Dounia Boumahni, BSc, Fatna Benettaib, MSc., Ali Ghamraoui, BSc;

*CHU de Québec – Université Laval*: Alexis Turgeon, MSc, David Bellemare, BSc, Ève Cloutier, Rana Daher, François Lauzier, MSc, Charles Francoeur, MSc;
*Centre Hospitalier Universitaire de Sherbrooke:* François Lamontagne, MD, Frédérick D'Aragon, MD, Elaine Carbonneau, BACC, Julie Leblond, BACC;
*Grace Hospital:* Gloria Vazquez-Grande, Nicole Marten
*Grand River Hospital (Kitchener)*: Theresa Liu, Atif Siddiqui
*Health Sciences Centre, Winnipeg:* Ryan Zarychanski, MD, Gloria Vazquez-Grande, MD, Nicole Marten, RN, Maggie Wilson, MSc;
*Hôpital du Sacré Coeur de Montréal:* Martin Albert, MD, Karim Serri, MD, Alexandros Cavayas, MD, Mathilde Duplaix, MSc, Virginie Williams, PhD;
*Juravinski Hospital:* Bram Rochwerg, MD, Tim Karachi, MD, Simon Oczkowski, MD, John Centofanti, MD, Tina Millen, RRT
*McGill University Health Centre*: Kosar Khwaja, Josie Campisi
*Niagara Health (St. Catherine's Hospital):* Erick Duan, MD, Jennifer Tsang, MD, Lisa Patterson, BA;
*Regina General Hospital*: Eric Sy, Chiraag Gupta, Sandy SHA Kassir
*Royal Alexandra Hospital*: Demetrios Kutsogiannis, Patricia Thompson
*Sunnybrook Health Sciences Centre*: Rob Fowler, Neill Adhikari, Maneesha Kamra, Nicole Marinoff
*St. Boniface General Hospital*: Ryan Zarychanski, Nicole Marten
*St. Joseph's Healthcare Hamilton*: Deborah Cook, Frances Clarke
*St. Mary's General Hospital (Kitchener)*: Rebecca Kruisselbrink, Atif Siddiqui
*St. Michael's Hospital*: John Marshall, MD, Laurent Brochard, MD, Karen Burns, MD, Gyan Sandhu, RN, Imrana Khalid, MD;
*The Ottawa Hospital:* Shane English, MSc, Irene Watpool, BScN, Rebecca Porteous, BSN, Sydney Miezitis, BSc, Lauralyn McIntyre, MSc;
*University Health Network:* Elizabeth Wilcox, Lorenzo del Sorbo, Hesham Abdelhady, Tina Romagnuolo
*University of Alberta:* Wendy Sligl, Nadia Baig, Oleksa Rewa MD, Sean Bagshaw MD
*William Osler Health System:* Alexandra Binnie, MD, Elizabeth Powell, MD, Alexandra McMillan, MD, Tracy Luk, MD, Noah Aref, MSc

**Critical Care Asia:**
*India Apollo Speciality Hospital - OMR, Chennai*: Devachandran Jayakumar, MD, R Pratheema, MD, Suresh Babu BSc;
*Apollo Main Hospital, Chennai*: C Vignesh, MD, Bharath Kumar TV, MD, N Ramakrishnan, MD, Augustian James BSc, Evangeline Elvira, BE

*Apollo Speciality Vanagaram, Vanagaram, Chennai*; R Ebenezer MD, S Krishnaoorthy MD, Lakshmi Ranganathan PhD, Manisha, MD, Madhu Shree BSc.
*Apollo First Med Hospital, Chennai:* Ashwin Kumar Mani, MD, Meghena Mathew, MD, Revathi BSc

*Nepal Grande International Hospital*: Sushil Khanal, MD, Sameena Amatya, RN;
*HAMS Hospital*: Hem Raj Paneru, MD, Sabin Koirala, MD, Pratibha Paudel ,RN;
*Nepal Mediciti Hospital*: Diptesh Aryal, MD, Kanchan Koirala, RN, Namrata Rai, RN, Subekshya Luitel, BSc;
*Tribhuvan University Teaching Hospital:* Hem Raj Paneru,MD, Binita Bhattarai, RN;

*Pakistan Ziauddin Hospital Clifton Campus:* Prof Madiha Hashmi, MD; Dr Ashok Panjwani, MD; Dr Zulfiqar Ali Umrani, Dr Shoaib Siddiq; Mr Mohiuddin Shaikh;
*National Institute of Cardiovascular Diseases Pakistan*: Nawal Salahuddin, MD, Sobia Masood;

***Croatia:***
*General Hospital Pozega*: Zdravko Andric, Sabina Cviljevic, Renata Đimoti, Marija Zapalac, Gordan *Mirković;*
*University Hospital of Infectious Diseases "Dr Fran Milhajevid"*: Bruno Baršić, Marko Kutleša, Viktor Kotarski;
*University Hospital of Zagreb*: Ana Vujaklija Brajković, Jakša Babel, Helena Sever, Lidija Dragija, Ira Kušan;

***Finland:***
*Helsinki University Hospital*: Suvi Vaara, PhD, Leena Pettilä, Jonna Heinonen, Ville Pettilä, PhD;
*Tampere University Hospital*: Anne Kuitunen, PhD, Sari Karlsson, PhD, Annukka Vahtera, PhD, Heikki Kiiski, PhD, Sanna Ristimäki***;***

***France:***
*Ambroise Pare Hospital*: Amine Azaiz, Cyril Charron, MD, Mathieu Godement, MD, Guillaume Geri, Antoine Vieillard-Baron;
*Centre Hospitalier de Melun*: Franck Pourcine, Mehran Monchi;
*Centre Hospitalier Simone Veil, Beauvais*: David Luis, MD, Romain Mercier, MD, Anne Sagnier, MD, Nathalie Verrier, MD, Cecile Caplin, MD, Jack Richecoeu, MD, Daniele Combaux, MD;
*Centre Hospitalier Sud Essonne*: Shidasp Siami, PhD, Christelle Aparicio, Sarah Vautier, Asma Jeblaoui, Delphine Lemaire-Brunel;
*Centre Hospitalier Tenon*: Muriel Fartoukh, MD, Laura Courtin, Vincent Labbe, MD , Guillaume Voiriot, MD, Sara Nesrine Salhi;
*Centre Hospitalier Victor Dupouy*: Gaetan Plantefeve, MD, Cécile Leparco, RN, Damien Contou, MD ;
*CHR d'Orleans*: Grégoire Muller, MD, Mai-Anh Nay, MD, Toufik Kamel, MD, Dalila Benzekri, MD, Sophie Jacquier, MD, Isabelle Runge, MD, Armelle Mathonnet, MD, François Barbier, MD, Anne Bretagnol, MD;
*CHRU Tours Hopital Bretonneau*: Emmanuelle Mercier, MD, Delphine Chartier, Charlotte Salmon, MD, Pierre-François Dequin, PhD, Denis GAROT, MD;
*CHU Dupuytren, Limoges;*
*Hôpital Civil, Hôpitaux Universitaires de Strasbourg;*
*Hôpital de Hautepierre, Hôpitaux Universitaires de Strasbourg*: Francis Schneider, PhD, Vincent Castelain, PhD, Guillaume Morel, MD, Sylvie L'Hotellier, MSc;
*Hospital Nord Franche-Comté*: Julio Badie, MD, Fernando Daniel Berdaguer, MD, Sylvain Malfroy, MD, Chaouki Mezher, MD, Charlotte Bourgoin, MSc, Guy Moneger, MD, Elodie Bouvier, MSc;
*Lariboisière Hospital*: Bruno Megarbane, PhD, Sebastian Voicu, PhD, Nicolas Deye, PhD, Isabelle Malissin, MD, Laetitia Sutterlin, MD, Aymen Mrad, MD, Adrien Pépin Lehalleur, MD, Giulia Naim, MD, Philippe Nguyen, MD, Jean-Michel Ekhérian, MD, Yvonnick Boué, MD, Georgios Sidéris, PhD, Dominique Vodovar, PhD, Emmanuelle Guérin, MD, Caroline Grant, MD;

*Le Mans Hospital*: Christophe Guitton, PhD, Cédric Darreau, MD, Mickaël Landais, MD, Nicolas Chudeau, MD, Alain Robert, PhD, Patrice Tirot, MD, Jean Christophe Callahan, MD, Marjorie Saint Martin, MD, Charlène Le Moal, MD, Rémy Marnai, MD, Marie Hélène Leroyer;

*Raymond Poincaré Hospital*: Djillali Annane, MD, Pierre Moine, MD, Nicholas Heming, MD, Virginie Maxime, MD, Isabelle Bossard, MSc, Tiphaine Barbarin Nicholier, MD, Bernard Clair, MD, David Orlikowski, MD, Rania Bounab, MD, Lilia Abdeladim, MD;

*Vendee Hospital*: Gwenhael Colin, MD, Vanessa Zinzoni, Natacha Maquigneau, Matthieu Henri-Lagarrigue, MD, Caroline Pouplet, MD;

***Germany:***

*Carl-Thiem-Klinikum Cottbus gGmbH*: Jens Soukup, PD, Richard Wetzold, Madlen Löbel, Dr. Ing, Lisa Starke, Patrick Grimm;

*Charité - Universitätsmedizin Berlin*: André Finn, MD, Gabriele Kreß, Uwe Hoff, MD, Carl Friedrich Hinrichs, MD, Jens Nee, MD;

*Jena University Hospital*: Mathias W. Pletz, PhD, Stefan Hagel, PhD, Juliane Ankert, MSc, Steffi Kolanos, BSc, Frank Bloos, PhD;

*Klinikum Dortmund gGmbH*: Daniela Nickoleit-Bitzenberger, MD , Bernhard Schaaf, MD, Werner Meermeier, MD, Katharina Prebeg, Harun Said Azzaui, Martin Hower, Klaus-Gerd Brieger, Corinna Elender, Timo Sabelhaus, Ansgar Riepe, Ceren Akamp, Julius Kremling, Daniela Klein, Elke Landsiedel-Mechenbier;

*University Hospital of Leipzig*: Sirak Petros, MD, Kevin Kunz, MD, Bianka Schütze, BSc;

*Universitätsklinikum Hamburg-Eppendorf*: Stefan Kluge, MD, Axel Nierhaus, MD, Dominik Jarczak, MD, Kevin Roedl, MD;

*University Hospital of Frankfurt*: Gernot Gerhard Ulrich Rohde, MD, Achim Grünewaldt, MD, Jörg Bojunga, MD;

*University Hospital of Wuerzburg*: Dirk Weismann, MD, Anna Frey, MD; Maria Drayss, MD, M.E. Goebeler, MEG, Thomas Flor, Gertrud Fragner, Nadine Wahl, Juliane Totzke, Cyrus Sayehli, MD;

*Vivantes Klinikum Neukölln*: Lorenz Reill, Michael Distler, MD, Astrid Maselli;

***Hungary:***

*Almási Balogh Pál Hospital, Ózd*: János Bélteczki, István Magyar, Ágnes Fazekas, Sándor Kovács, Viktória Szőke;

*Jósa András County Hospital, Nyíregyháza*: Gábor Szigligeti, János Leszkoven;

***Ireland:***

*Beacon Hospital Dublin*: Daniel Collins, MRCPI, Kathy Brickell, RGN, Liadain Reid, MPH, Michelle Smyth, PgDip, Patrick Breen, MB FJFICMI, Sandra Spain, RGN;

*Beaumont Hospital*: Gerard Curley; PhD, Natalie McEvoy, MSc, Pierce Geoghegan, MB, Jennifer Clarke, MB;

*Galway University Hospitals*: John Laffey, MD, Bairbre McNicholas, PhD, Michael Scully, MD, Siobhan Casey, RN, Maeve Kernan, RN, Aoife Brennan, PhD, Ritika Rangan, PhD, Riona Tully, MB, Sarah Corbett, MB, Aine McCarthy, MB, Oscar Duffy, MB, David Burke, MB;

*St Vincent's University Hospital, Dublin*: Alistair Nichol, PhD, Kathy Brickell, RGN, Michelle Smyth, PGDip, Leanne Hayes, PhD, Liadain Reid, MPH, Lorna Murphy, RGN, Andy Neill, MB, Bryan Reidy, MSc, Michael O'Dwyer, PhD, Donal Ryan, MD, Kate Ainscough, PhD;

***Netherlands:***

*Canisius Wilhelmina Ziekenhuis*: Oscar Hoiting, MD, Marco Peters, MD, Els Rengers, MD, Mirjam Evers, RN, Anton Prinssen, RN;

*Deventer Hospital*: Huub L.A. van den Oever, MD, Arriette Kruisdijk-Gerritsen, CCRN;

*Jeroen Bosch Ziekenhuis*: Koen Simons, PhD, Tamara van Zuylen, RN, Angela Bouman, RN;

*Meander Medisch Centrum*: Laura van Gulik, PhD;

*Radboud University Medical Center Nijmegen*: Jeroen Schouten, PhD, Peter Pickkers, PhD, Noortje Roovers, BSc, Margreet Klop-Riehl, BSc, Hetty van der Eng, BSc;

*UMC Leiden*: Evert de Jonge, PhD, Jeanette Wigbers, RN, Michael del Prado, RN;

*UMC Utrecht*: Marc Bonten, PhD, Olaf Cremer, PhD, Lennie Derde, MD, PhD, Jelle Haitsma Mulier, MD, Anna Linda Peters, PhD, Birgit Romberg, MD; Helen Leavis, PhD; Roger Schutgens, PhD

*Ziekenhuis Gelderse Vallei*: Sjoerd van Bree, MD, Marianne Bouw-Ruiter, RN, Barbara Festen, MD, Fiona van Gelder, MD, Mark van Iperen, MD, Margreet Osinga, RN, Roel Schellaars, MD, Dave Tjan, MD, Ruben van der Wekken, MD, Max Melchers, MD, Arthur van Zanten, MD;

*Haga Ziekenhuis*: Kees van Nieuwkoop, PhD; Thomas Ottens MD; Yorik Visser, RC

*Onze Lieve Vrouwe Gasthuis*: Nicole Juffermans, PhD; Matty Koopmans, RC

***New Zealand*:**

*Auckland City Hospital, Cardiothoracic and Vascular ICU:* Shay McGuinness, MBChB, Rachael Parke, PhD, Eileen Guilder, MA, Magdalena Butler, RN, Keri-Anne Cowdrey, RN, Melissa Woollett, BHSc (Nurs);

*Auckland City Hospital, DCCM:* Colin McArthur, FJFICM, Thomas Hills, DPhil, Lynette Newby, MN, Yan Chen, MN, Catherine Simmonds, PGDipHSc, Rachael McConnochie, MN, Caroline O'Connor;

*Christchurch Hospital:* Jay Ritzema Carter, PhD, Seton Henderson, MD, Kymbalee Van Der Heyden, BSc, Jan Mehrtens, PGCert, Anna Morris, BN, Stacey Morgan, BN;

*Middlemore Hospital:* Tony Williams, BMedSc, Alex Kazemi**,** BMedSc, Susan Morpeth, PhD, Rima Song, PGDip, Vivian Lai, MHSc, Dinuraj Girijadevi, PGCert;

*North Shore Hospital:* Robert Everitt, FACEM, Robert Russell, BSc(Hons), Danielle Hacking, PGDipNurs;

*Rotorua Hospital:* Ulrike Buehner, FCARCSI, Erin Williams, MSc;

*Tauranga Hospital:* Troy Browne, FCICM, Kate Grimwade, FRACP, Jennifer Goodson, RN, Owen Keet, FANZCA, Owen Callender, FANZCA;

*Waikato Hospital:* Robert Martynoga, FCICM, Kara Trask, PGCert, Amelia Butler, PGCert,

*Wellington Hospital:* Paul Young, PhD, Chelsea Young, PGDip, Eden Lesona, MNSc, Shaanti Olatunji, MClinIm, Leanlove Navarra, BSc (Nurs), Raulle Sol Cruz, BSc (Nurs)

*Whangarei Hospital:* Katherine Perry, FJFICM, Ralph Fuchs, FANZCA, Bridget Lambert, PGDip;

*Taranaki Base Hospital:* Jonathan Albrett, FCICM, Carolyn Jackson, RN, Simon Kirkham, PGDip;

***Portugal*:**

*Hospital de Abrantes:* Nuno José Teodoro Amaro dos Santos Catorze, MD, Tiago Nuno Alfaro Lima Pereira, MD, Ricardo Manuel Castro Ferreira, RN, Joana Margarida Pereira Sousa Bastos, PharmD, Teresa Margarida Oliveira Batista, RN;

***Romania*:**

*"Dr. Victor Babes" Clinical Hospital of Infectious and Tropical Diseases Bucharest:* Simin Aysel Florescu, PhD, Delia Stanciu, MD, Mihaela Florentina Zaharia, MD, Alma Gabriela Kosa, MD, Daniel Codreanu;

***Saudi Arabia*:**

*King Abdulaziz Medical City- Riyadh:* Yaseen M Arabi, MD, Eman Al Qasim, RN, Haytham Tlayjeh, MD, Lolowa Alswaidan, MSc, Brintha Naidu, RN

***Spain*:**

*Hospital del Mar:* Rosana Muñoz-Bermúdez, MD, Judith Marin-Corral, PhD, Anna Salazar Degracia, PhD, Francisco Parrilla Gómez, MD, Maria Isabel Mateo López;

*Reina Sofia University Hospital:* Rafael León López, MD, Jorge Rodriguez, PhD, Sheila Cárcel, Rosario Carmona, MD, Carmen de la Fuente, MD, Marina Rodriguez, MD;

***United Kingdom*:**

*Aberdeen Royal Infirmary:* Callum Kaye, MBChB, Angela Allan, PGDip;

*Addenbrooke's Hospital:* Charlotte Summers, PhD, Petra Polgarova MSc;

*Alder Hey Children's NHS Foundation Trust:* Stephen J McWilliam, PhD, Daniel B Hawcutt, MD, Laura Rad, BSc(Hons), Laura O'Malley, BSc(Hons), Jennifer Whitbread, BSc(Hons);

*Alexandra Hospital Redditch:* Olivia Kelsall, MBChB, Nicholas Cowley MD, Laura Wild, BSc(Hons), Jessica Thrush, RGN, Hannah Wood, BSc(Hons), Karen Austin, RGN;

*Altnagelvin Hospital:* Adrian Donnelly, FFICM, Martin Kelly, MD, Naoise Smyth MB ChB, Sinéad O'Kane, BSc(Hons), Declan McClintock, MSc, Majella Warnock, MPharm, Ryan Campbell BSc, Edmund McCallion MPharm;

*Antrim Area Hospital:* Paul Johnson, FFARCSI, Shirley McKenna, MSc, Joanne Hanley, MSc, Andrew Currie, MSc, Barbara Allen, MPharm, Clare Mc Goldrick, MPhil, Moyra Mc Master, RGN, ;

*Barnet Hospital:* Rajeev Jha, MD, Michael Kalogirou, MD, Christine Ellis, PhD, Vinodh Krishnamurthy, PhD, Vashish Deelchand, MSc, Aibhilin O'Connor, MSc;

*Basildon Universty Hospital: Dipak Mukherjee, MD, Agilan Kaliappan, MD, Anirudda Pai, MD,Mark Vertue, Anne Nicholson, Joanne Riches, Gracie Maloney, Lauren Kittridge, Amanda Solesbury, Kezia Allen*

*Belfast Health and Social Care Trust (Belfast City Hospital, Mater Infirmorium, Royal Victoria Hospital):* Jon Silversides, PhD, Peter McGuigan, MBBCh, Kathryn Ward, BSc, Aisling O'Neill, BSc, Stephanie Finn, BSc;

*Brighton and Sussex University Hospitals Trust:* Barbara Phillips, Laura Oritz-Ruiz de Gordoa, BSc;

*Bristol Royal Infirmary:* Jeremy Bewley, MBChB, Matthew Thomas, MBChB, Katie Sweet, BSc(Hons), Lisa Grimmer, BSc(Hons), Rebekah Johnson, BSc(Hons);

*Calderdale and Huddersfield Foundation Trust:* Jez Pinnell, MD, Matt Robinson, BSc(Hons), Lisa Gledhill, MSc, Tracy Wood, BSc(Hons);

*Cardiff and Vale University Health Board:* Matt Morgan, PhD, Jade Cole, BSc, Helen Hill, BSc, Michelle Davies, BN, Angharad Williams, BSc, Emma Thomas, BSc, Rhys Davies, BSc, Matt Wise, DPhil;

*Charing Cross Hospital:* David Antcliffe, PhD, Maie Templeton, MSc, Roceld Rojo, BSN, Phoebe Coghlan, MA, Joanna Smee, BSc;

*Chesterfield Royal Hospital:* Euan Mackay, MD, Jon Cort, MD, Amanda Whileman, BSc, Thomas Spencer, Nick Spittle, Sarah Beavis, MD, Anand Padmakumar, MD, Katie Dale, BSc, Joanne Hawes, BSc, Emma Moakes, BSc, Rachel Gascoyne, BSc, Kelly Pritchard, BSc, Lesley Stevenson, BSc, Justin Cooke, MD, Karolina Nemeth-Roszpopa, MD;

*The Christie NHS Foundation Trust:* Vidya Kasipandian, FFICM, Amit Patel, Suzanne Allibone, Roman Mary-Genetu, BSc;

*Colchester Hospital:* Mohamed Ramali, FRCA, Ooi HC, MRCEM, Alison Ghosh, RN, Rawlings Osagie, PharmD, Malka Jayasinghe Arachchige, MBBS, Melissa Hartley, MBBS;

*Countess of Chester Hospital:* Peter Bamford, FFICM, Emily London, MBChB, Kathryn Cawley, MRes, Maria Faulkner, BSc, Helen Jeffrey, DipNS;

*Croydon University Hospital:* Ashok Sundar Raj, MD, Georgios Tsinaslanidis, MD, Reena Nair Khade, BSc, Gloria Nwajei Agha, BSc, Rose Nalumansi Sekiwala;

*Cumberland Infirmary:* Tim Smith, FRCA, Chris Brewer, BPharm(Hons), Jane Gregory, BSc(Hons);

*Darlington Memorial Hospital:* James Limb, FRCA, Amanda Cowton, BSc(Hons), Julie O'Brien, DipNurs, Kelly Postlethwaite, DipNurs;

*Derriford Hospital:* Nikitas Nikitas, PhD, Colin Wells, MSc, Liana Lankester, PGCert, Helen McMillan, MSc;

*Dorset County Hospital:* Mark Pulletz, FFICM, Patricia Williams, AdDip, Jenny Birch, BA, Sophie Wiseman, Mpharm, Sarah Horton, BA(Hons);

*East Kent Hospitals (Queen Elizabeth the Queen Mother Hospital):* Ana Alegria, CCT, Salah Turki, MBBch, Tarek Elsefi, MRCP, Nikki Crisp, BSc, Louise Allen, BSc;

*East Lancashire Hospitals NHS Trust (Royal Blackburn Hospital):* Matthew Smith, MD, Sri Chukkambotla, MD, Wendy Goddard, BSc, Stephen Duberley BSc;

*Freeman Hospital and Royal Victoria Infirmary, Newcastle upon Tyne*: Iain J McCullagh, FRCA, Philip Robinson, MSc, Bijal Patel, MSc, Sinead Kelly, PGDip;

*Frimley Health NHS Foundation Trust:* Omar Touma, MD, Susan Holland, Christopher Hodge, Holly Taylor, Meera Alderman, Nicky Barnes, Joana Da Rocha, BSc, Catherine Smith, BSc, Nicole Brooks, Thanuja Weerasinghe, BSc, Julie-Ann Sinclair, Yousuf Abusamra, MD, Ronan Doherty, MD, Joanna Cudlipp, MD, Rajeev Singh, MD, Haili Yu, MD, Admad Daebis, MD, Christopher NG, MD, Sara Kendrick, MD, Anita Saran, MD, Ahmed Makky, MD, Danni Greener, MD, Louise Rowe-Leete, Dip, Alexandra Edwards, Dip, Yvonne Bland, BSc, Rozzie Dolman, BSc, Tracy Foster, BSc;

*Gateshead Health NHS Trust:* Vanessa Linnett, MD, Amanda Sanderson, Jenny Ritzema, Helen Wild;

*George Eliot Hospital:* Divya Khare, FRCA, Meredith Pinder, BSN, Selvin Selvamoni, MSc, Amitha Gopinath, MBA;

*Glan Clwyd Hospital:* Richard Pugh, FFICM, Daniel Menzies, FRCP, Richard Lean, MBChB, Xinyi Qiu, MBChB, Jeremy James Scanlon, MBChB;

*Glasgow Royal Infirmary:* Kathryn Puxty, MD, Susanne Cathcart, BSc, Chris Mc Govern, MBChB, Samantha Carmichael, MRPharms, Dominic Rimmer, BSc;

*Glenfield Hospital Leicester:* Hakeem Yusuff, FFICM, Graziella Isgro, FFICM, Chris Brightling, PhD, Michelle Bourne, BSc(Hons), Michelle Craner, DipHE, Rebecca Boyles, BSc (Hons);

*Grange University Hospital:* Tamas Szakmany, PhD, Shiney Cherian, BSc, Gemma Williams, BSc, Christie James, MSc, Abby Waters, MSc;

*Great Western Hospitals NHS Foundation Trust:* Malcolm Watters, MBBCh, Rachel Prout, MBChB, Louisa Davies, BSc, Suzannah Pegler, BSc(Hons), Lynsey Kyeremeh, BPharm, Aiman Mian, MBBS;

*Guy's & St Thomas' NHS Foundation Trust:* Manu Shankar-Hari, PhD, Marlies Ostermann, PhD, Marina Marotti, BSc, Neus Grau Novellas, BSc, Aneta Bociek, BSc;

*Hammersmith Hospital:* Stephen Brett, MD, Sonia Sousa Arias, BSc, Rebecca Elin Hall, BN;

*Homerton University Hospital NHS Foundation Trust:* Susan Jain, MD, Abhinav Gupta MD, Catherine Holbrook

*James Cook University Hospital:* Jeremy Henning, MB, Stephen Bonner, BSc, Keith Hugill, BSc, Emanuel Cirstea, MSc, Dean Wilkinson, BSc, Jessica Jones, BSc;

*James Paget University Hospitals:* Michal Karlikowski, MD, Helen Sutherland, BSc(Hons), Elva Wilhelmsen, DipHE, Jane Woods, BSc, Julie North, BSc(Hons);

*Kettering General Hospital:* Dhinesh Sundaran, FFICM, Laszlo Hollos, FFICM, Susan Coburn, PGCert, Anna Williams, BSc, Samantha Saunders, BTEC;

*King's College Hospital (Denmark Hill site):* Phil Hopkins, MD, John Smith, RN, Harriet Noble, RN, Maria Theresa Depante, RN, Emma Clarey, RN;

*Lancashire Teaching Hospitals NHS Foundation Trust:* Shondipon Laha, FFICM, Mark Verlander, MBA, Alexandra Williams, MSc;

*Leeds Teaching Hospitals Trust:* Elankumaran Paramasivam, FRCP, Elizabeth Wilby, BSc (Hons), Bethan Ogg, BSc (Hons), Clare Howcroft, BSc (Hons), Angelique Aspinwall, BSc (Hons), Sam Charlton, BSc (Hons), Richard Gould, MBBS, Deena Mistry, MPharm, Sidra Awan, MPharm, Caroline Bedford, MPharm;

*Leicester General Hospital:* Andrew Hall, MRCP, Jill Cooke, RGN, Caroline Gardiner-Hill, RGN, Carolyn Maloney, Nigel Brunskill, PhD;

*Leicester Royal Infirmary:* Hafiz R Qureshi, MRCPI, Neil Flint, MBChB, Sarah Nicholson, Sara Southin, Andrew Nicholson, Amardeep Ghattaoraya, MSc;

*Lewisham and Greenwich NHS Trust:* Dr Daniel Harding, MD, Sinead O'Halloran , Amy Collins Emma Smith , Estefania Trues;

*Liverpool Foundation Trust Aintree:* Barbara Borgatta, PhD, Ian Turner-Bone, DipHE, Amie Reddy, Laura Wilding, DipHE;

*Liverpool Heart and Chest Hospital:* Craig Wilson, Zuhra Surti;

*Luton and Dunstable University Hospital:* Loku Chamara Warnapura, FFICM, Ronan Agno, BSc, Prasannakumari Sathianathan, MSc, Deborah Shaw, FFICM, Nazia Ijaz, FFICM, Dean Burns, MD, Mohammed Nisar, MD, Vanessa Quick, MD, Craig Alexander, BSc, Sanil Patel BSc, Nafisa Hussain, Yvynne Croucher, BSc, Eva-Maria Lang, MD, Banu Rudran, MD, Syed Gilani, MD, Talia Wieder, MD, Margaret Louise Tate, BSc;

*Maidstone and Tunbridge Wells NHS Trust:* David Golden, FFICM, Miriam Davey, PGDip, Rebecca Seaman BSc (Hons);

*Manchester Royal Infirmary:* Tim Felton, FFICM, Jonathan Bannard-Smith, FFICM, Joanne Henry, Richard Clark, DiPHE, Kathrine Birchall, BSc(Hons), Joanne Henry, MA, Fiona Pomeroy, BSc (Hons), Rachael Quayle, Dip.HE, Katharine Wylie, MSc, Anila Sukuraman, BSc, John McNamarra, MD;

*Medway Maritime Hospital:* Arystarch Makowski, PhD, Beata Misztal, PhD, Iram Ahmed, PhD, Kevin Neicker, MBA, Sam Millington, BMBS, Rebecca Squires, BSc, Masroor Phulpoto, MBBS;

*Milton Keynes University Hospital:* Richard Stewart, Esther Mwaura, BSc, Louise E Mew, BSc(Hons), Lynn Wren, BSc(Hons), Felicity Willams, PhD;

*Mid & South Essex NHS Foundation Trust:* Aneta Oborska, FRCA, Rino Maeda, MBBS, Selver Kalchko-Veyssal, MD, Raji Orat Prabakaran, BSc, Bernard Hadebe, MSc, Eric Makmur, MBBS, Guy Nicholls, MBBS;

*Musgrove Park Hospital:* Richard Innes, MBBCh, Patricia Doble, BSc(Hons), Libby Graham, RN, Charmaine Shovelton, RN;

*Nevill Hall Hospital:* Vincent Hamlyn, MBBCh, Nancy Hawkins, PhD, Anna Roynon-Reed, MSc, Sean Cutler, MSc, Sarah Lewis, MBChB;

*Newham University Hospital:* Juan Martin Lazaro, PhD, Tabitha Newman, MSc;

*Ninewells Hospital:* Pauline Austin, MBChB, Susan Chapman, MBChB, Louise Cabrelli, BSc;

*Norfolk and Norwich University Hospital:* Simon Fletcher, FFICM, Jurgens Nortje, FFICM, Deirdre Fottrell-Gould, Dip, Georgina Randell, Dip, Katie Stammers, BSc;

*Northampton General Hospital:* Mohsin Zaman, MRCP, Einas Elmahi, MPhil, Andrea Jones, PhD, Kathryn Hall, Dip;

*Northern General Hospital, Sheffield:* Gary H Mills, PhD, Kim Ryalls, RegPharmTech, Kate Harrington RCN, Helen Bowler, BSc, Jas Sall, BSc, Richard Bourne, PhD;

*North Manchester General Hospital:* Zoe Borrill, MD, Tracy Duncan, MD, Thomas Lamb, MD, Joanne Shaw, BSc, Claire Fox, BSc, Kirstie Smith, BSc, Sarah Holland, Bethany Blackledge, BSc, Liam McMorrow, BSc, Laura Durrans, Jade Harris;

*North Middlesex University Hospital:* Jeronimo Moreno Cuesta, MD, Kugan Xavier, MD, Dharam Purohit, EDIC, Munzir Elhassan, MBBS, Anne Haldeos, BSc, Rachel Vincent, DipHE, Marwa Abdelrazik, MBBCH, Samuel Jenkins, BMBS, Arunkumar Ganesan, MD, Rohit Kumar, DA, David Carter, MBBS, Dhanalakshmi Bakthavatsalam, BSc;

*Oxford University Hospitals:* Matthew Rowland, FFICM, Paula Hutton, PGCert, Archana Bashyal, MSc, Neil Davidson, BSc, Clare Hird, MSc, Sally Beer, MSc;

*Pilgrim Hospital Boston:* Manish Chhablani, FFICM, Gunjan Phalod, MPharm, Amy Kirkby, BSc(Hons), Simon Archer, BSc(Hons), Kimberley Netherton, RGN;

*Princess Royal Hospital:* Barbara Philips, MD, Dee Mullan, BSc, Denise Skinner, BSc, Jane Gaylard, BSc, Julie Newman, BSc;

*Princess of Wales Hospital:* Sonia Arun Sathe, MD, Lisa Roche, BSc, Ellie Davies, BSc, Keri Turner;

*Poole Hospital:* Henrik Reschreiter, FFICM, Julie Camsooksai, PGDE, Sarah Patch, BSc(Hons), Sarah Jenkins, BSc(Hons), Charlotte Humphrey, BSc (Hons);

*Queen Alexandra Hospital Portsmouth:* David Pogson, MSc, Steve Rose, BSc, Zoe Daly, BSc, Lutece Brimfield, BN, Angie Nown;

*Queen Elizabeth Hospital, Birmingham:* Dhruv Parekh, PhD, Colin Bergin, BSc, Michelle Bates, BSc, Christopher McGhee, BSc, Daniella Lynch, BSc, Khushpreet Bhandal, Dip, Kyriaki Tsakiridou, MSc, Amy Bamford, BSc, Lauren Cooper, MSc, Tony Whitehouse, MD, Tonny Veenith, MD;

*Queen Elizabeth University Hospital, Glasgow:* Malcolm A.B. Sim, MD, Sophie Kennedy Hay, BN, Steven Henderson, MPH, Maria Nygren, MSc, Eliza Valentine, HNC;

*Queen's Hospital, Burton:* Amro Katary, MD, Gill Bell, BSc, Louise Wilcox, BSc, Katy English BSc, Ann Adams;

*Queen's Hospital, Romford:* Mandeep-Kaur Phull, MBBS, Abbas Zaidi, MBBS, Tatiana Pogreban, BN, Lace Paulyn Rosaroso, BN;

*Queens Medical Centre and Nottingham City Hospital:* Daniel Harvey, BMBS, Benjamin Lowe, BMBS, Megan Meredith, BSc(Hons), Lucy Ryan, MNSc, DREEAM Research Team;
*The Rotherham NHS Foundation Trust:* Anil Hormis, FRCA, Rachel Walker, BA, Dawn Collier, BSc, Sarah Kimpton, MSc, Susan Oakley;
*Royal Alexandra Hospital:* Kevin Rooney, MBChB, Natalie Rodden, BSc, Emma Hughes, BSc, Nicola Thomson, BSc(Hons), Deborah McGlynn, BSc, Charlotte Clark, Dip, Patricia Clark, BSc;
*Royal Berkshire Hospital:* Andrew Walden, FFICM, Liza Keating, MBChB, Matthew Frise, DPhil, Tolu Okeke, BSc, Nicola Jacques, MSc, Holly Coles, BSc, Emma Tilney, BSc, Emma Vowell, DipHE;
*Royal Bournemouth and Christchurch Hospitals:* Martin Schuster-Bruce, FRCA, Sally Pitts, BSc, Rebecca Miln, ADipHE, Laura Purandare, MBA, Luke Vamplew, BSc;
*Royal Brompton Hospital:* Brijesh Patel, FRCA, Debra Dempster, Mahitha Gummadi, Natalie Dormand, Shu Fang Wang;
*Royal Cornwall NHS Trust:* Michael Spivey, FFICM, Sarah Bean, RN, Karen Burt, RN, Lorraine Moore, MPharm;
*Royal Devon and Exeter NHS Foundation Trust:* Christopher Day, MD, Charly Gibson, MBChB, Elizabeth Gordon, BSc, Letizia Zitter, BSc, Samantha Keenan, BSc;
*Royal Glamorgan Hospital:* Jayaprakash Singh, MD, Ceri Lynch, MD, Lisa Roche, Justyna Mikusek, Bethan Deacon, Keri Turner;
*Royal Gwent Hospital*: Tamas Szakmany, PhD, Evelyn Baker, MSc, John Hickey, MSc, Shreekant Champanerkar, MBBS, Lindianne Aitken, MSc, Lorraine Lewis Prosser, MSc;
*Royal Hallamshire Hospital, Sheffield:* Gary H Mills, PhD, Ajay Raithatha, FFICM, Kris Bauchmuller, FFICM, Norfaizan Ahmad, FFICM, Matt Wiles FFICM, Jayne Willson, RN;
*Royal Hampshire Hospitals:* Irina Grecu, MD, Jane Martin, Caroline Wrey Brown, Ana-Marie Arias, Emily Bevan;
*Royal Infirmary of Edinburgh:* Thomas H Craven, PhD, David Hope, PGDip, Jo Singleton, BN, Sarah Clark, MNurs, Corrienne McCulloch, PhD;
*Royal Liverpool University Hospital:* Ingeborg D Welters, PhD, David Oliver Hamilton, BMBS, Karen Williams, RGN, Victoria Waugh, BA, David Shaw, DipHE, Suleman Mulla, MBChB, Alicia Waite, PhD, Jaime Fernandez Roman, BSc, Maria Lopez Martinez, BSc;
*Royal London Hospital:* Zudin Puthucheary, PhD, Timothy Martin, BA(Hons), Filipa Santos, RN, Ruzena Uddin, MSc(Hons), Maria Fernandez, MSc, Fatima Seidu, MSc, Alastair Somerville, MSc, Mari Lis Pakats, MSc, Priya Dias, PhD, Salam Begum, BSc, Tasnin Shahid, BSc;
*The Royal Free Hospital:* Sanjay Bhagani, FRCP, Mark De Neef, MSc, Helder Filipe, BSc, Sara Mingos, BSc, Amitaa Maharajh, BA, Glykeria Pakou, BA, Aarti Nandani, MPharm;
*The Royal Marsden NHS Foundation Trust:* Kate Colette Tatham, PhD, Shaman Jhanji, PhD, Ethel Black, BSNurs, Arnold Dela Rosa, BSNurs, Ryan Howle, FRCA, Ravishankar Rao Baikady, FRCA;
*The Royal Oldham Hospital:* Redmond P Tully, FFICM, Andrew Drummond, FFICM, Joy Dearden, BSc, Jennifer E Philbin, MSc, Sheila Munt, SRN;
*The Royal Wolverhampton NHS Trust:* Shameer Gopal, MBBCh, Jagtar- Singh Pooni, MBBS, Saibal Ganguly, MBBS, Andrew Smallwood, RGN, Stella Metherell, RGN;
*Royal Papworth Hospital:* Alain Vuylsteke, MD, Charles Chan, FRCA, Saji Victor, COVID Research Team, Papworth Hospital;
*Royal Stoke Hospital:* Ramprasad Matsa, FRCP, Minerva Gellamucho, BSN, Michelle Davies, NVQ;

*Royal Surrey County Hospital:* Ben Creagh-Brown, PhD, Joe Tooley, MSc, Laura Montague, BSc, Fiona De Beaux, BSc, Laetitia Bullman, MBChB;

*Royal United Hospital Bath:* Ian Kerslake, FFICM, Carrie Demetriou, RN, Sarah Mitchard, MBBS, Lidia Ramos, RN, Katie White, MSc;

*Russells Hall Hospital:* Michael Reay, FFICM, Steve Jenkins, MD, Caroline Tuckwell, Angela Watts, BSc, Eleanor Traverse, Stacey Jennings;

*Salisbury NHS Foundation Trust:* Phil Donnison, FFICM, Maggie Johns, RGN, Ruth Casey, BSc, Lehentha Mattocks, Dip, Sarah Salisbury;

*Salford Royal NHS Foundation Trust:* Paul Dark, PhD, Alice Harvey, BSc, Reece, Doonan, BSc, Liam McMorrow, BA (Hons), Karen Knowles, BA (Hons);*Sandwell and West Birmingham NHS Trust:* Jonathan Hulme, FFICM, Santhana Kannan, FFICM, Sibet Joseph, BSc, Fiona Kinney, RGN, Ho Jan Senya, BPharm;

*Sherwood Forest Hospitals NHS Foundation Trust:* Valli Ratnam, MD, Mandy Gill, Jill Kirk, Sarah Shelton

*South Tyneside District Hospital:* Christian Frey, MD, Riccardo Scano, MD, Madeleine McKee, BSc, Peter Murphy, BSc;

*Southmead Hospital:* Matt Thomas, FFICM, Ruth Worner, RGN, Beverley Faulkner, RGN, Emma Gendall, BSc, Kati Hayes, BSc, Hayley Blakemore, BSc, Borislava Borislavova, MSc;

*St. Bartholomew's Hospital:* Colin Hamilton-Davies, MBBS, Carmen Chan, BSc, Celina Mfuko, BSc, Hakam Abbass, MSc, Vineela Mandadapu, MSc;

*St. George's Hospital:* Susannah Leaver, MRCP, Kamal Patel, MRCP, Sarah Farnell-Ward, MSc, Romina Pepermans Saluzzio, BSc, John Rawlins, MBBS;

*St. Mary's Hospital:* Anthony Gordon, MD; Dorota Banach, BSc, Ziortza Fernández de Pinedo Artaraz, BN, Leilani Cabreros, BSN;

*St. Peter's Hospital, Chertsey:* Ian White, FFICM, Maria Croft, BSc(Hons), Nicky Holland, BN(Hons), Rita Pereira, MPharm;

*Stepping Hill Hospital, Stockport:* Ahmed Zaki, PhD, David Johnson, MPhil, Matthew Jackson, MBChB, Hywel Garrard, BMBS, Vera Juhaz, MD, Louise Brown BSc (Hons);

*Sunderland Royal Hospital:* Alistair Roy, MBChB, Anthony Rostron, PhD, Lindsey Woods, BSc, Sarah Cornell, BSc;

*Swansea Bay University Health Board:* Suresh Pillai, FFCIM, Rachel Harford, RN, Helen Ivatt, FRCA, Debra Evans, BN, Suzanne Richards, BN, Eilir Roberts, MBBCh, James Bowen, MBBCh James Ainsworth, MBBS;

*Torbay and South Devon NHS Foundation Trust:* Thomas Clark, MBChB, FRCA, FFICM, Angela Foulds, BSc, Stacey Atkins, Dip RN;

*United Lincolnshire NHS Trust:* Kelvin Lee, PhD, Russell Barber, FRCA, Anette Hilldrith, RGN, Claire Hewitt, RGN, Gunjan Phalod, MPharm;

*University Hospitals Coventry & Warwickshire NHS Trust:* Pamela Bremmer, BSc, Geraldine Ward, MA, Christopher Bassford, PhD;

*University Hospital of North Tees:* Farooq Brohi, FFARCSI, Vijay Jagannathan, FRCA, Michele Clark, MA, Sarah Purvis, Dip, Bill Wetherill, MSc;

*University Hospital Southampton NHS Foundation Trust:* Ahilanandan Dushianthan, PhD, Rebecca Cusack, MD, Kim de Courcy-Golder, PGDip, Karen Salmon, MSc, Rachel Burnish, Simon Smith, BN, Susan Jackson, BSc, Winningtom Ruiz, BSc (Hons), Zoe Duke BSc (Hons) Magaret Johns, BA, Michelle Male, Dip, Kirsty Gladas, BSc (Hons), Satwinder Virdee, MPharm, Jacqueline Swabe, MPharm, Helen Tomlinson;

*Warwick Hospital:* Ben Attwood, MBBCh, Penny Parsons, BSc, Bridget Campbell, BSc, Alex Smith, BSc;
*Watford General Hospital:* Valerie J Page, MBBCh, Xiao Bei Zhao, BSc (Hons), Deepali Oza, BPharm, Gail Abrahamson, DPhil, Ben Sheath, BSc (Hons), Chiara Ellis, BSc (Hons);
*Western General Hospital, Edinburgh:* Jonathan Rhodes, PhD, Thomas Anderson, MBChB, Sheila Morris;
*Whipps Cross Hospital:* Charlotte Xia Le Tai, MBChB, Amy Thomas, MSc, Alexandra Keen, MSc;
*Whiston Hospital*: Dr Ascanio, Tridente, Karen Shuker, Jeanette Anders, Sandra Greer, Paula Scott, Amy Millington, Philip Buchanan, Jodie Kirk
*Wirral University Teaching Hospital NHSFT:* Craig Denmade, MBChB, Girendra Sadera, MBBS, Reni Jacob, BSc, Cathy Jones, BSc, Debbie Hughes;
*Worcester Royal Hospital:* Stephen Digby, MBBS, Nicholas Cowley, MD, Laura Wild, BSc(Hons), Jessica Thrush, RGN, Hannah Wood, BSc(Hons), Karen Austin, RGN;
*Wrexham Maelor Betsi Cadwaladr University Hospital*: David Southern, FFICM, Harsha Reddy, FFICM, Sarah Hulse, BSc, Andy Campbell, FFICM, Mark Garton, Claire Watkins, PGDip, Sara Smuts, BN;
*Wrightington, Wigan and Leigh Teaching Hospitals NHS Foundation Trust:* Alison Quinn, MD, Benjamin Simpson, MD, Catherine McMillan, MD, Cheryl Finch, BSc, Claire Hill BSc, Josh Cooper;
*Wye Valley NHS Trust:* Joanna Budd, MBBS, Charlotte Small, PhD, Ryan O'Leary, MBBS, Janine Birch, RN, Emma Collins, BSc (Hons);
*Wythenshawe Hospital*: Peter D G Alexander, FFICM, Tim Felton, FFICM, Susan Ferguson, BSc, Katharine Sellers, BSc, Joanne Bradley-Potts, BSc;
*York Teaching Hospital*: David Yates, FCRA, Isobel Birkinshaw, BSc(Hons), Kay Kell, BSc(Hons), Zoe Scott, BN, Harriet Pearson, BSc;

***United States of America***
*University of Pittsburgh Research Staff*: CRISMA Center—Kelsey Linstrum, Stephanie Montgomery, Kim Basile, Dara Stavor, Dylan Burbee, Amanda McNamara, Renee Wunderley, Nicole Bensen, Aaron Richardson; MACRO Center—Peter Adams, Tina Vita, Megan Buhay, Denise Scholl, Matthew Gilliam, James Winters, Kaleigh Doherty, Emily Berryman

*UPMC Hospital Champions*: UPMC Altoona—Mehrdad Ghaffari, UPMC East—Meghan Fitzpatrick; UPMC Jameson and Horizon —Kavitha Bagavathy; UPMC Mercy—Mahwish Hussain, Chenell Donadee; UPMC Williamsport—Emily Brant; UPMC McKeesport—Kayla Bryan-Morris, John Arnold and Bob Reynolds; UPMC Hamot—Gregory Beard; UPMC Presbyterian—Bryan McVerry, David Huang; UPMC Pinnacle—Janice Dunsavage, Salim Saiyed, Erik Hernandez, John Goldman, Cynthia Brown, Susan Comp, James Raczek, Jenny Lynne Morris, Jesus Vargas Jr., Daniel Weiss, Joseph W. Hensley, Erik Kochert, Chris Wnuk, Christopher Nemeth, Brent Mowery, Christina Hutchinson, Lauren Winters

*UPMC Pharmacy*: Erin McCreary, Elise Martin, Ryan Bariola, Alex Viehman, Jessica Daley, Alyssa Lopus, Mark Schmidhofer, UPMC Directors of Pharmacy

*UPMC ICU Service Center*: Rachel Sackrowitz, Chenell Donadee, Aimee Skrtich

*UPMC Wolff Center*: Tami Minnier, Mary Kay Wisniewski, Katelyn Mayak

*UPMC eRecord Team*: Richard Ambrosino, Sherbrina Keen, Sue Della Toffalo, Martha Stambaugh, Ken Trimmer, Reno Perri, Sherry Casali, Rebecca Medva, Brent Massar, Ashley Beyerl, Jason Burkey,

Sheryl Keeler, Maryalyce Lowery, Lynne Oncea, Jason Daugherty, Chanthou Sevilla, Amy Woelke, Julie Dice, Lisa Weber, Jason Roth, Cindy Ferringer, Deborah Beer, Jessica Fesz, Lillian Carpio

*Data Collection/Curation Team*: Salim Malakouti (Computer Science, University of Pittsburgh), Edvin Music and Dan Ricketts (CRISMA Center), Andrew King (Biomedical Informatics, University of Pittsburgh), Gilles Clermont (Critical Care Medicine), Robert Bart (UPMC Health Services Division)

*UPMC Clinical Analytics*: Oscar Marroquin, Kevin Quinn, William Garrard, Kyle Kalchthaler

*UPMC Office of Healthcare Innovation*: Derek Angus

*Department of Emergency Medicine*: Alexandra Weissman, Donald Yealy, David Barton, Nadine Talia

*Department of Critical Care Medicine*: David Huang, Florian Mayr, Andrew Schoenling, Mark Andreae, Varun Shetty, Emily Brant, Brian Malley, Chenell Donadee, Derek Angus, Christopher Horvat, Christopher Seymour, Timothy Girard, Gilles Clermont, Rachel Sackrowitz, Robert Bart

*Division of Infectious Diseases*: Ghady Haidar

*Division of Pulmonary, Allergy, and Critical Care Medicine*: Bryan McVerry, William Bain, Ian Barbash, Meghan Fitzpatrick, Christopher Franz, Georgios Kitsios, Kaveh Moghbeli, Brian Rosborough, Faraaz Shah, Tomeka Suber

*Berry Consultants*: Roger Lewis, Michelle Detry, Anna McGlothlin, Christina Saunders, Mark Fitzgerald, Ashish Sanil, Scott Berry

*Global Coalition for Adaptive Research (GCAR):* Meredith Buxton, Brian Alexander, Tracey Roberts

#### 1.3 ATTACC

**Lead Principal Investigators**
Ryan Zarychanski, Patrick Lawler, Ewan Goligher

**International Trial Steering Committee**
Ryan Zarychanski, Chair (University of Manitoba, Canada), Patrick Lawler (University of Toronto, Canada), Ewan Goligher (University of Toronto, Canada), Robert Rosenson (Mount Sinai New York) U.S. Country Lead, Jose Nicolau (University of Sao Paulo) Brazil Country Lead, Jorge Escobedo (Unidad de Investigación en Epidemiología Clínica) Mexico Country Lead, Michael Farkouh (University of Toronto, Canada), Dean Fergusson (Ottawa Hospital Research Institute, Canada), Anand Kumar (University of Manitoba, Canada), Nicole Marten (St Boniface Hospital, Wpg, Canada), John Marshall (University of Toronto, Canada), Alexis Turgeon (Université Laval, Québec, Canada), Charlotte Bradbury (University of Bristol, United Kingdom), Marc Carrier (University of Ottawa, Canada), Vlad Dzavik (University of Toronto, Canada), Rob Fowler (University of Toronto, Canada), Emily Gibson McDonald (McGill University, Canada), Peter Gross (McMaster University, Canada), Brett Houston (University of Manitoba, Canada), Mansoor Hussain (University of Toronto, Canada), Susan Kahn (McGill University, Canada), Srinivas Murthy (University of British Columbia, Canada), Arthur Slutsky (University of Toronto, Canada), Tobias Tritschler (University of Ottawa). Patient Partners: Margaret Ostrowski and Suzanne Dubois.

**Global Project Management**
*Ozmosis Research Inc.*: Lindsay Bond, Jackie Amaral, Vanessa Wareham, Karlee Trafford, Mila Khanna
*University of Manitoba:* Nicole Marten, Dayna Solvason

**Global Data Coordinating Center**
*SOCAR Research:* Bridget-Anne Kirwan, Sophie de Brouwer, David La Framboise

**Regional Project Management**
***Brazil***
*Avanti:* Andrea Martinez, Pedro Ohara, Juliana Bacca, Natalia de Jesus, Sandra Zier, Douglas Assis
*InCor:* Jose Nicolau, Natassja Huemer, Neury Martins, Fabiana Nakajima

***Mexico***
*UNAM:* Jorge Escobedo

**Adjudication Committee**
Brendan Everett, MD, MPH, Sean van Diepen, MD, MSc, Gregoire Le Gal, MD, Deborah Siegal, MD, Jean-Philippe Galanaud, MD, Sheila Hegde, MD, Yuri Kim, MD, Natalia Rost, MD, Aneesh Singhal, MD

**Statistical Analysis Committee**
Roger Lewis, Michelle Detry, Anna McGlothlin (Chair), Mark Fitzgerald, Christina Saunders, Maria Brooks

**Laboratory Committee**
Peter Gross, Rita Selby, Patrick Lawler

**Site Principal Investigators and Research Coordinators**
***Canada***
*Health Sciences Centre:* Brett Houston, Soumya Alias, Rhonda Silva

*St. Boniface General Hospital:* Vi Dao, Nicole Marten, Maureen Hutmacher, Lisa Rigaux, Quinn Tays, Hessam Kashani
*Grace Hospital*: Glen Drobot, Nicole Marten Maureen, Hutmacher, Nora Choi
*University Health Network:* Ewan Goligher, Richard Dunbar-Yaffe, Mohammad Shafiee, Jenna Wong
*Hamilton Health Sciences:* Peter Gross, Michele Zondag
*Ottawa Hospital:* Lana Castellucci, Penny Philips, Moussa Meteb, Irene Watpool, Rebecca Porteous
*Centre Hospitalier Universitaire de Québec – Université Laval*: Alexis Turgeon, David Bellemare, Olivier Costerousse
*Centre Hospitalier de l'Université de Montréal:* Emmanuelle Duceppe, Roberta-Daila Carling, Madeleine Durand
*Jewish General Hospital:* Vicky Tagalakis, Elena Shulikovsky, Sophie Florencio
*McGill University Health Centre:* Emily McDonald, Sarah Elsayed, Kristen Moran
*Institut Universitaire de Cardiologie et de Pneumologie – Université Laval:* Francois Lellouche, Patricia Lizotte
*Regina General Hospital:* Andrea Lavoie, Kendra Townsend
*Victoria General Hospital:* Daniel Ovakim, Deborah Parfett, Fiona Auld
*St. Joseph's Healthcare Hamilton:* Peter Gross, Michele, Zondag
*Hôpital Montfort:* Marc Carrier, Mélodie Carrier
*Centre Hospitalier Universitaire de Sherbrooke:* Francois Lamontagne, Elaine Carbonneau

***United States***
*University of Chicago Medicine:* Jonathan Paul, Cynthia Arevalo, Karli Molignoni
*Ochsner Medical Center*: Mark B. Effron, Sarah Cohen, Hunter McDaniel
*William Beaumont Hospital:* Girish B. Nair, Tammy Osentoski
*St. Louis Veterans Affairs Medical Centre:* Martin Schoen, Kristin Courtright, Kelly Reno
*Maine Medical Centre:* Daniel Meyer, Terilee Gerry
*Vanderbilt University Medical Centre:* Aaron Aday, Emily Shardelow, Michaela Burton
*Henry Ford University:* Scott Kaatz, Stacy Ellsworth
*Emory University Hospital Midtown:* Bryan Wells, Claudia Merlin, Amanda Fieback
*Mayo Clinic:* Vivek Iyer, Matthew Johnson
*Saint Barnabas Medical Center:* Nirav Mistry, Amber Turner
*Cooper University Health Care*: Nitin Puri, Christa Schorr
*Hackensack University Medical Center*: Ronaldo Go, Peter Canino
*Montefiore Health System:* Henny Billett, Ervin Mazniku

***Brazil***
*Instituto do Coração (InCor) :* Felipe Gallego Lima, Alexandra Vieira, Renata Maia, Anna Mostachio
*Instituto de Infectologia Emilio Ribas:* Walter Braga, Sunamita Lima
*Hospital 9 de Julho:* Fernanda Santos
*Hospital das Clínicas da Faculdade de Medicina da Universidade de São Paulo*: Rinaldo Siciliano
*Sociedade Beneficente Israelita Hospital Albert Einstein:* Remo Furtado, Fernanda Ferraz Assir, Beatriz Moraes
*Instituto Goiano de Oncologia e Hematologia (INGOH):* Mayler Santos, Luciana Barros
*Instituto de Cardiologia de Santa Catarina:* Artur Herdy, Vera Pereira
*Santa Casa de Votuporanga:* Mauro Hernandes, Renée Amorim, Milena Bandeira
*AngioCor Clínica Médica em Blumenau:* Adrian Kormann, Jádina Spricigo, Sabrina Zimmerman
*Hospital São Vicente de Paulo:* Rogerio Tumelero, Andressa Giordani, Flavia Ghizzoni
*Instituto de Medicina Vascular*: Euler Manenti, Karen Ruschel, Aline Borba

*Instituto de Pesquisa Clínica de Campinas (IPECC):* José Saraiva, Carla Vicente, Marina Silva
*Hospital Agamenon Magalhães: Cardiologia:* Joao Moraes Jr, Shylla Ribeiro, Thais Barros
*Centro de Pesquisas Clínicas Humap - UFMS:* Delcio Silva Jr, Paula Serafin, João Xavier
*Instituto de Cardiologia do Rio Grande do Sul:* Oscar Dutra, Andrea Brum
*Hospital Felicio Rocho:* Ana Procopio, Maria Alves
*Praxis Pesquisa Médica:* Anete Grumach, Lara Bertolini
*Hospital Universitário Pedro Ernesto:* Carmem Porto, Samara Oliveira
*Casa de Saúde Santa Marcelina:* Marcelo Burihan, Monica Santos
*UNIMED Campo Grande:* Delcio Silva Jr, Erica Nery, Paula Serafin
*Instituto de Moléstias Cardio Vasculares de Tatuí:* Wladmir Saporito, Thais Pereira, Bruna Mancini
*Santa Casa de Misericórdia de Itabuna:* Eduardo Kowalski Neto, Bruno Andrade, Jessica Santos
*Clínica de Campo Grande S/A:* Mauricio Pompilio, Rebeca Pompilio
*Parana Medical Research Center:* Sergio Grava, Karen Koga
*Hospital das Clínicas da UFPR:* Miguel Silva, Debora Lemos

***Mexico***
*1 Carlos MacGregor Sánchez Navarro:* Jorge Escobedo, Betriz Villegas
*Hospital de Infectologia del Centro Médico Nacional La Raza:* Eduardo Mateos García, Miguel Ángel Cortés Vázquez, Yessica Sara Pérez González, Paulina Carreño Pérez
*Hospital General regional 2 El Marqués:* Julieta Valenzuela, Juan Anwar Santillán

#### 1.4 ACTIV-4a

**Lead Investigators**

Judith S. Hochman MD, Matthew D. Neal MD, Jeffrey S. Berger MD

**Protocol Development Committee**

Judith S. Hochman MD (Chair), Matthew D. Neal MD (co-Chair), Jeffrey S. Berger MD (co-PI)
Mary Cushman, MD, MSc, Lisa Baumann Kreuziger, MD , Scott Berry, PhD, Michael Farkouh, MD, Michelle N. Gong, MD, Kristin Hudock, MD, MSTR, Keri S. Kim, PharmD, Lucy Z. Kornblith, MD, Patrick R. Lawler, MD, MPH, Eric Leifer, PhD, Bryan J. McVerry, MD, Harmony R. Reynolds, MD, Jennifer G. Wilson, MD

**NYU Grossman School of Medicine/ NYU Langone Health Clinical Coordinating Center and Chairs Office**

Judith Hochman (Chair); Jeffrey Berger (Co-PI), Harmony Reynolds (Co-Investigator), Aira Contreras, Stephanie Mavromichalis, Margaret Gilsenan, Anna Naumova, Arlene Roberts

**University of Pittsburgh Data Coordinating Center**

Matthew Neal (Co-Chair), Stephen Wisniewski, Christine Leeper, Derek Angus, Heather Eng, Maria Brooks, Kelsey Linstrum, Christopher Seymour, Timothy Girard, Scott Berry, Stephanie Montgomery, Mary Martinez, Jake Schreiber, Joshua Froess, Zhuxuan Fu, Yongqi Zhong, Ashita Vadlamudi, Frank Sciurba, Alison Morris

**SOCAR Research:** Bridget-Anne Kirwan, Sophie de Brouwer, Emilie Perrin, Caroline Gombault, Sandra Bula, Michael Nelson, Céline Daelemans, Robin Wegmüller, David La Framboise

**National Heart Lung and Blood Institute, National Institutes of Health**

Program: W. Keith Hoots, MD, Andrei Kindzelski, MD, PhD, Traci Mondoro, PhD, Antonello Punturieri, MD, PhD, Gail Weinmann, MD

**Statistics:** Eric Leifer, PhD, James F. Troendle, PhD

**RTI International:** Amy S. Kendrick, RN, MSN, Tracy L Nolen, DrPH, Sonia Thomas, DrPH

**Network Coordinating Centers**

*ISCHEMIA/MINOCA-HARP/EPPIC-NET:* Harmony Reynolds (Lead), Aira Contreras, Stephanie Mavromichalis, Margaret Gilsenan, Anna Naumova, Danielle Sin, Elhaji Diene, Ewelina Gwiszcz, Isabelle Hogan, Alair Holden
*PETAL:* Michelle Gong (Lead), Nancy Ringwood, Laura Fitzgerald, John Sharer, Daniel Ceusters, Carolyn Hintlian
*Multi-Net*: Lucy Kornblith (Lead), Brenda Nunez-Garcia, Valerie Uribe, Carolyn Hendrickson, Christen Barua, M. Margaret Knudson, John Park, Ana Gonzalez
*FIBHULP*: Jose Lopez-Sendon (Lead), Paloma Moraga Alapont, Paula Prieto, Victoria Hernandez
*RAPID*: Mary Cushman, Lisa Baumann Kreuziger, Shannon Broaddrick
*IlliNet*: Keri Kim, Sean Quigley
*REMAP-CAP*: Bryan McVerry (Lead), David Huang, Meredith Buxton, Tracey Roberts, Kelsey Linstrum
*StrokeNet*: Hooman Kamel, Pooja Khatri, Jamey Frasure, Amy Silken

**Site Investigators and Research Coordinators**

***Spain***

*Hospital Universitario Ramon Y Cajal*: Jose Luis Lopez-Sendon Moreno, Raquel Morillo Guerrero, Sebastian Garcia Madrona, Almudena Molinera, Otilia Navarro Carrion, Raquel Besse Diaz, Sergio Diz Farina, Fernando Hidalgo Salinas, Paula Gonzalez Ferrandiz, Svetlana Zhilina, Macarena Alpanes Buesa, Andres Gonzalez Garcia

*Hospital Clinico Universitario de Salamanca*: Miguel Marcos Martin, Monica Sanchez, Juan Hernandez, Felipe Alvarez Navid, Moncef Belhassen Garcia, Cristina Carbonell Munoz, Guillermo Hernandez Perez, Amparo Lopez Bernus, Jose Angel Martin Oterino

***United States***

*NYU Langone Health*: Jeffrey Berger, Norma Keller, Eugene Yuriditsky, Tania Ahuja, James Horowitz, Alexander Hindenburg, Tamta Chkhikvadze, Sam Parnia, Zelda Moran, Maja Fadzan, Julia Levine, Stanley Cobos, Arline Roberts, Lia Mamistvalova, Michela Garabedian, Farzana Ahmed, Gabriela Zapata, Marsha Robinson

*University of Illinois at Chicago Health*: John Quigley, Keri Kim, Jeff Jacobson, Neha Atal

*Montefiore Medical Center*: Michelle Gong, Daniel Ceusters, Omowunmi Amosu, Hiwet Tzehaie, Rahul Nair, Brenda Lopez, Manuel Hache Marliere, Daniel Fein, Obiageli Offor, Michael Kiyatkin, Sweta Chekuri, Benjamin Galen, Aram Hambardzumyan, Aditi Desai, Mahmuda Akhter, Hammad Aleem, Sahil Virdi, Roshni Shah, Aluko Hope, Jen-Ting Chen, Amira Mohamed

*Zuckerberg San Francisco General Hospital*: Lucy Kornblith, John Park, Carolyn Hendrickson, M. Margaret Knudson, Aaron Kornblith, India Shelley, Biniam Ambachew, Brenda Nunez-Garcia

*UPMC Presbyterian*: David Huang, Bryan McVerry, Kelsey Linstrum, Nicole Bensen, Dylan Burbee, Aaron Richardson, Amanda McNamara, Dara Stavor, Menna Abaye, Denise Scholl, Renee Wunderley, Anne Yang, Sher Shah Amin, Emily Berryman, Matthew Gilliam**,** Kim Basile, Giles Clermont, William Garrard, Christopher Horvat, Kyle Kalchthaler, Andrew J. King, Daniel Ricketts, Salim Malakouti, Oscar Marroquin, Edvin Music, Kevin Quinn, Mark Andreae, William Bain, Ian Barbash, Emily Brant, David Barton, Meghan Fitzpatrick, Christopher A Franz, Ghady Haidar, Mahwish Hussain, Georgios D Kitsios, Florian B Mayr, Brian Malley, Erin McCreary, Kaveh Moghbeli, Brian Rosborough, Andrew Schoenling, Faraaz A Shah, Varun U Shetty, Tomeka Suber, Nadine Talia, Alexandra Weissman, Caitlin Schaefer, Michael Muir, Kelly Lynn Urbanek

*Rutgers New Jersey Medical School*: Yonatan Greenstein, Randall (Randy) Teeter, Michael Plump, Olga Kovalenko, Eliana Obando, Yanille Taveras, Brittany Fanka, Nipun Suri, Sunil Patel, Maninderpal Kaur

*University of Cincinnati Medical Center*: Kristin Hudock, Robert Hite, Tammy Roads, Adamsegd Gebremedhen, Simra Kiran, Harshada More

*UC San Diego Hillcrest*: Todd Costantini, Terry Curry, Emmer Trinidad, MaryBeth Tyler, Allison Berndtson, Matthew Allison, Harpreet Bhatia, Julie Denenberg, Brennan Marsh-Armstrong, Ilya Verzhbinsky, Timothy Morris, Timothy Fernandes, Ann Elliott, Amelia Eastman

*Ronald Reagan UCLA Medical Center*: George Lim, Gregory Hendey, Steven Chang, Nida Qadir, Rebecca Beutler, Trisha Agarwal, Julia Vargas, Jason Singer, David Haase, James Murphy, Agatha Brzezinski, Anna Yap, Dong Han Yao, Claudie Bolduc, Catherine Antonuk, Hannah Spungen, Ashley Vuong

*Stanford University Medical Center*: Jennifer Wilson, Angela Rogers, Joseph Levitt, Rosemary Vojnik, Jonasel Roque, Cynthia Perez

*Oregon Health and Science University*: Akram Khan, Olivia Krol, Kinjal Mistry, Kelly Nguyen, Zhengchun Lu, Milad Karami Jouzestani, Atinderpal Singh, Madeline Mcdougal, Andrew Salar, Simeon Florea, Raya Adi, Chandni Anadkat, Emmanuel Mills, Zachary Zouyed, Rupali Deshmukh, Catherine Hough

*Baylor Scott and White Medical Center-Temple*: Robert Widmer, Wanda Fikes, Elizabeth Kiesle

*University of Michigan*: Robert Hyzy, Pauline Park, Shijing Jia, Jakob McSparron, Bonnie Wang, Sinan Hanna, Kelli McDonough, Amanda Melvin, Kristine Nelson, Norman Olbrich

*Sarah Cannon and HCA Research Institute*: Andrew Goodman, Hallie E. Hank, Drew Quillen, Abdullah Shamsuddin, Logan Michl, Molly Harper, Mariah Phipps, Charita Braker

*Duke University Hospital*: Lana Wahid, Oluwayemisi Mohammed, Stephen Gazda, Jacob Craven, Ryan Jackson, Kira Abuchowski, Rowena Dolor, Thomas Ortel, Maria Manson, Lorraine Vergara, Gloria Pinero, Stephanie Freel

*MetroHealth System:* Vidya Krishnan, Cindy Newman, Pete Leo, Carla Greenwood, Andrew Wright, Edward L. Warren, John Daryl Thornton, Calen Frolkis

*UCSF San Francisco*: Michael Matthay, Kirsten, Kangelaris, Kathleen Liu, Carolyn Calfee, Hanjing Zhuo, Brian Daniel, Kimberly Yee, Alejandra Jauregui, Rajani Ghale, Suzanna Chak, Katherine Wick, Emily Siegel, Chayse Jones, Kimia Ashktorab

*Kansas University Medical Center*: Lewis Satterwhite, Penelope Harris, Kimberly Lovell, Mohamed Mourad, Charles Bengtson, Tahani Atieh, Kyle Brownback, Carolina Aguiar, Megan White, Karisa Deculus, Lawrence Scott, Lindsey English, Stephanie Greer, Sharon Murry, Lisa Woodring, Usman Nazir, Amanda Truong, Nelda Mallett, Shereesa Williams, Heidi Hellwig, Michael Burton

*University of Texas Southwestern Medical Center*: Ambarish Pandey, Christopher Bates, Bienka Lewis, Jesse Tarbutton, Nitin Kondamudi, Ruth Giselle Huet, Xiaohang Xu, Marielle Berger-Nagele, Erika Molina

*Cleveland Clinic Foundation*: Abhijit Duggal, Simon Mucha, Omar Mehkri, Alexander King, Bryan Poynter, Kiran Ashok, Nirosshan Thiruchelvam, Debasis Sahoo, Alice Goyanes, Matthew Siuba, Ravi Sunderkrishnan, Steven Minear, Jaime Hernandez-Montfort, Jinesh Mehta, Carla McWilliams, Chinwe Anekwe, Amy Van, Andrea Calderon, Luz Arazo, Syed Sohaib Nasim, Camila De Carvalho Teixeira, Delmy Zelaya

*Cook County Health*: Saurabh Malhotra, Arlet Nedeltcheva, Katayoun Rezai, Michael Hoffman, Ruben Hernandez Acosta, Juan Sarmiento, Shreeyala Uday

*Ascension St. John Clinical Research Institute*: Nicholas Hanna, Anuj Malik, Stacie Merritt, Julie Davenport, Kathryn Mears, Jane Bryce, Melanie Arnold, Joy Norwood , Cheryl Urias

*University of Mississippi Medical Center*: Matthew Kutcher, James Galbraith, Alan Jones, Utsav Nandi, Vishnu Garla, Rebekah Peacock, Jenna Davis, Emily Grenn, Taylor Shaw, Morgan Moore

*Hennepin County Medical Center*: Matt Prekker, Michael Puskarich, Brian Driver, Jason Baker, Anne Frosch, Adam Kolb, Laura Hubbard, Alex Dunn, Audrey Hendrickson, Ellen Maruggi, Tayne Andersen, William Miller, Abagail Raiter, Radhika Edpuganti, Quinn Ehlen, Grace Leland, Michael Roth, Tyler Scharber, Walker Tordsen, Mackenzie Reing, Ann Isaksen, Heidi Erickson

*University of Wisconsin Hospital; Meriter Hospital*: John Sheehan, Sarah Stewart, Kraig Kumfer, Rafael Veintimilla, Chris Roginski, Nicole Bonk, Scott Ensminger, Muhammad Shahzeb Munir, Jashan Octain, Ann Sheehy, Alexis Waters, Scott Wilson

*Boston University*: Naomi Hamburg, Erika Teresa Minetti, Karla Damus, Robert Eberhardt, Elizabeth Klings, Rena Zheng, Leili Behrooz, Anna Gao

*Denver Health and Hospital Authority:* Mitchell Cohen, Caitlin Robinson, Andrew Byars, Marcela Fitzpatrik-Wilson, Katherine Ling, Tiffany Bendelow, James Wallace, Ivor Douglas

*University of Alabama*: Sheetal Gandotra, Mark Dransfield, Elizabeth Westfall, Micah Whitson, Donna Harris, Derek Russell, Siddharth Patel

*VA New York Harbor Healthcare System*: Binita Shah, Leandro Maranan, Alana Choy-Shan, Nathaniel Smilowitz, Robert Donnino, Jeffrey Lorin, Mary Keary

*Penn State Health Milton S. Hershey Medical Center:* Steven Moore, Kunal Karamchandani, Pauline Go, Anthony Bonavia, Lonnie Fender, Nancy Campbell, Judie Howrylak, Kevin Gardner, Lisa Fox, Paula Trump, Katie Loffredo, McKenna Snyder, Sharon O`Brien, Lisa Schultz, Shane Kinard

*Washington University School of Medicine, ACCS Research*: Grant Bochicchio, Kelly Bochicchio, Stacey Reese, Ricardo Fonseca, Bryan Sato, Kristen Ferguson, Chris Machica, Jennifer McCarthy, Jose Aldana, Rohit Rasane, Melissa Canas, Hussain Afzal, Tiffany Osborn, Mark Hoofnagle, Jennifer Leonard, Jason Snyder, Douglas Schuerer, Melissa Stewart, Piroska Kopar, Kelly Vallar, Jessica Kramer, Isaiah Turnbull

#### 1.5 Funding Agencies

##### 1.5.1 REMAP-CAP

Supported by the European Union — through FP7-HEALTH-2013-INNOVATION: the Platform for European Preparedness Against (Re-)emerging Epidemics (PREPARE) consortium (602525), and Horizon 2020 research and innovation program: the Rapid European Covid-19 Emergency Research response (RECOVER) consortium (101003589) — and by the Australian National Health and Medical Research Council (APP1101719), the Health Research Council of New Zealand (16/631), a Canadian Institutes of Health Research Strategy for Patient-Oriented Research Innovative Clinical Trials Program Grant (158584), the U.K. NIHR and the NIHR Imperial Biomedical Research Centre, the Health Research Board of Ireland (CTN 2014-012), the UPMC Learning While Doing Program, the Breast Cancer Research Foundation, the French Ministry of Health (PHRC-20-0147), the Minderoo Foundation, Amgen, Eisai, the Global Coalition for Adaptive Research, and the Wellcome Trust Innovations Project (215522). Dr. Gordon is funded by an NIHR Research Professorship (RP-2015-06-18), and Dr. Shankar-Hari by an NIHR Clinician Scientist Fellowship (CS-2016-16-011).

##### 1.5.2 ATTACC

The ATTACC platform was supported by grants from the Canadian Institutes of Health Research, LifeArc Foundation, Thistledown Foundation, Research Manitoba, Ontario Ministry of Health, and the Peter Munk Cardiac Centre.

##### 1.5.3 ACTIV-4a

The ACTIV-4a platform was sponsored by the National Heart, Lung, and Blood Institute, National Institutes of Health, Bethesda, MD and administered through OTA-20-011.

##### 1.5.4 Disclaimer

The views expressed in this publication are those of the author(s) and not necessarily those of the National Health Service (UK), the National Institute for Health Research (UK), the Department of Health and Social Care (UK), or of the National Institutes of Health (USA).

#### Section 2 – Supplemental Methods

As described in the Statistical Analysis Plan (Protocol Appendix pp 34-112), the primary analysis was conducted on all patients with confirmed Covid-19. According to a Bayesian hierarchical approach, the estimation of treatment effect in patients with severe Covid-19 included the potential for statistical borrowing from patients with moderate patient groups of Covid-19 (high baseline D-dimer - >2 times the upper limit of normal, low baseline D-dimer - ≤2 times the upper limit of normal, or missing baseline D-dimer) thereby allowing maximal incorporation of all information. Borrowing between patient groups in the model only occurs where the observed effects were similar between groups. As the primary model included information about assignment in patient groups where randomization was ongoing and blinded to the investigators, the primary analysis was run by the fully unblinded Statistical Analysis Committee (SAC), who conduct all protocol-specified trial update analyses and report results to the DSMB.

In addition to the primary analysis conducted by the SAC, multiple sensitivity analyses were conducted by the investigator team who remain blinded to results of on-going randomization in the moderate patient groups. A sensitivity analysis of the primary outcome was repeated in a second model using only data from patients in the severe Covid-19 patient group (i.e. excluding patients with moderate Covid-19, Table S2). For this analysis there was no adjustment for borrowing from the moderate state participants.

Further sensitivity analyses assessed whether results were modified by including patients who were randomized as suspected Covid-19 but ultimately did not document SARS-CoV-2 PCR test positivity, or whether results were modified by excluding site and time effects from the model. As described in the mpRCT statistical design plan, the analytical model includes site and time covariate terms to account for variability in treatment effect according to site (random effect) and to variation in the endpoint over time (fixed effect).

As the incidence of major bleeding is an important secondary outcome, and one of the platforms in the mpRCT (REMAP-CAP) was testing the additive benefit of antiplatelet therapy concomitant with the anticoagulation domain, an additional sensitivity analysis was performed excluding patients who were receiving an antiplatelet agent at baseline or who were randomized to either treatment or control in the antiplatelet domain of REMAP-CAP.

To determine the extent to which interpretation of the trial results would be affected by one's prior beliefs and given the widely accepted belief that therapeutic anticoagulation would be beneficial in severe Covid-19, we applied a post hoc prior for enthusiasm for therapeutic anticoagulation to the model. This prior specified a 50% prior probability that the odds ratio would be ≥ 1.75 and a 10% probability of an odds ratio < 1 (inferior), consistent with fairly confident belief that the treatment was beneficial. Assuming a baseline mortality rate of 35%, an odds ratio of 1.75 is equivalent an approximately 10% absolute risk reduction, consistent with reasonably strong enthusiasm for meaningful clinical benefit. In addition, we applied a post hoc prior for skepticism against therapeutic anticoagulation in the model. This prior specified a 50% prior probability that the odds ratio would be < 1 (inferior) and a 66% probability of futility (OR ≤ 1.2) consistent with reasonable skepticism against a benefit of therapeutic anticoagulation.

For per protocol analyses, protocol adherence was defined according to pre-specified dosing classifications for each form of heparin (see Protocol Appendix page 40 - Statistical Analysis Plan). For the therapeutic anticoagulation arm, management was defined as “per protocol” if the initial anticoagulants over the first 48 hours after randomization were administered at therapeutic or subtherapeutic doses. In the usual care arm, management was defined as “per protocol” if the initial anticoagulants over the first 48 hours after randomization was administered at low or intermediate doses.

Treatment effect was examined in pre-specified subgroup analyses (See Protocol Appendix page 45 - Statistical Analysis Plan) including age, sex, and requirement for invasive mechanical ventilation at baseline. To assess for potential heterogeneity of treatment effect according to venous thromboprophylaxis dosing in the usual care arm, subgroups were defined by whether patients were randomized at a site that used “intermediate dose” venous thromboprophylaxis in more than 50% of usual care patients, or less than 50% of usual care patients. For each subgroup analysis a separate treatment effect is modeled by subgroup. The posterior distribution for each subgroup effect and 95% credible intervals are reported.

There was no imputation of missing outcomes in either primary or secondary analyses. Cases were excluded on an analysis-by-analysis basis, i.e. patients missing outcome data were included in treatment compliance and safety analyses. The secondary outcomes were also analysed in the unblinded ITT model. The primary safety analysis compared the proportion of patients who developed one or more serious adverse thrombotic or major bleeding events across groups.

### Section 3 – Supplemental Results

#### Site Participation in the Severe State Therapeutic Heparin Trial

During the study period, April 21, 2020 to December 19, 2021, 393 sites in 10 countries (U.K., Ireland, Netherlands, Australia, New Zealand, Saudi Arabia, Canada, United States, Brazil and Mexico) were open for enrolment in the severe state therapeutic heparin multiple platform trial.

#### Additional Analyses

Provided below are the sensitivity analyses of the primary outcome (days alive and free of organ support) and secondary outcomes of hospital mortality, death or major thrombotic events, and major bleeding where appropriate. Table S1 presents proportion of patients receiving specific drugs and treatment regimens. Table S2 provides the odds ratio (OR) and 95% Credible Interval (CrI) as well as posterior probabilities for futility, inferiority, and superiority for each parameter in the model as described. Table S3 details the breakdown of major thrombotic events by treatment group.

The additional analyses specified in the statistical analysis plan will be presented with the final report when more detailed long term outcome data are available.

### Section 4 – Supplemental Tables

#### Table S1 – Additional Results

| | Therapeutic anticoagulation N=532 | Usual care venous thromboprophylaxis N=557 |
|---|---|---|
| Anticoagulant drug – n (%)[a] | N=443 | N=465 |
| Enoxaparin | 207 (46.7) | 242 (52.0) |
| Dalteparin | 154 (34.8) | 166 (35.7) |
| Tinzaparin | 32 (7.2) | 26 (5.6) |
| Subcutaneous unfractionated heparin | 1 (0.2) | 21 (4.5) |
| Intravenous unfractionated heparin | 47 (10.6) | 2 (0.4) |
| Fondaparinux | 0 (0) | 0 (0) |
| Direct oral anticoagulant | 0 (0) | 0 (0) |
| None | 2 (0.5) | 8 (1.7) |
| Other | 0 (0) | 0 (0) |
| Post-randomization dosage equivalents [a] | N=412 | N=433 |
| Low dose thromboprophylaxis | 15 (3.6) | 179 (41.3) |
| Intermediate dose thromboprophylaxis | 45 (10.9) | 222 (51.3) |
| Subtherapeutic dose anticoagulation | 32 (7.8) | 9 (2.1) |
| Therapeutic dose anticoagulation | 320 (77.7) | 23 (5.3) |
| Enrollment by platform | | |
| REMAP-CAP | | |
| Confirmed Covid-19 | 452 | 466 |
| Suspected Covid-19 | 47 | 44 |
| ATTACC [b] | 18 | 20 |
| ACTIV-4a [b] | 62 | 71 |

[a] Data reported reflects those in whom specific dosing information was available at the time the dataset was locked for analysis

[b] Platforms exclusively enrolled patients with confirmed Covid-19

#### Table S2 – Sensitivity Analyses

| | Therapeutic anticoagulation | Usual care pharmacological thromboprophylaxis |
|---|---|---|
| **Patients with confirmed severe Covid-19[a]** | N=529 | N=545 |
| **Organ support-free days** | | |
| Median, (IQR) | 3 (-1, 16) | 5 (-1, 16) |
| Adjusted odds ratio (95% CrI) | 0.85 (0.68, 1.06) | 1 (Reference) |
| Probability of futility[b], % | 99.8 | - |
| Probability of superiority[c], % | 7.9 | - |
| Probability of inferiority[d], % | 92.1 | - |
| **Hospital survival** | | |
| No. of patients/total no. (%) | 340/529 (64.3) | 356/545 (65.3) |
| Adjusted odds ratio (95% CrI) | 0.88 (0.66, 1.16) | 1 (Reference) |
| Probability of superiority[c], % | 17.5 | - |
| Probability of inferiority[d], % | 82.5 | - |
| **Organs support-free days: assuming enthusiastic prior in favor of therapeutic anticoagulation** | | |
| Adjusted odds ratio (95% CrI) | 0.89 (0.72, 1.10) | 1 (Reference) |
| Probability of futility[b], % | 99.7 | - |
| Probability of superiority[c], % | 14.2 | - |
| Probability of inferiority[d], % | 85.8 | - |
| **Organ support free days: assuming skeptical prior in favor of therapeutic anticoagulation** | | |
| Adjusted odds ratio (95% CrI) | 0.86 (0.69, 1.07) | 1 (Reference) |
| Probability of futility[b], % | 99.9 | - |
| Probability of superiority[c], % | 8.7 | - |
| Probability of inferiority[d], % | 91.3 | - |
| **Patients with either confirmed or suspected Covid-19[a]** | N=574 | N=583 |
| **Organ support-free days** | | |
| Median, (IQR) | 4 (-1, 16) | 7 (-1, 16) |
| Adjusted odds ratio (95% CrI) | 0.88 (0.71, 1.08) | 1 (Reference) |
| Probability of futility[b], % | 99.8 | - |
| Probability of superiority[c], % | 11.3 | - |
| Probability of inferiority[d], % | 88.7 | - |
| **Hospital survival** | | |
| Adjusted odds ratio (95% CrI) | 0.89 (0.68, 1.16) | 1 (Reference) |
| Probability of superiority[c], % | 19.1 | - |
| Probability of inferiority[d], % | 80.9 | - |

| | Therapeutic anticoagulation | Usual care pharmacological thromboprophylaxis |
|---|---|---|
| Site and time effects excluded from model[a] | N=529 | N=545 |
| Organ support-free days | | |
| Adjusted odds ratio (95% CrI) | 0.88 (0.71, 1.09) | 1 (Reference) |
| Probability of futility[b], % | 99.7 | - |
| Probability of superiority[c], % | 12.3 | - |
| Probability of inferiority[d], % | 87.7 | - |
| Hospital survival | | |
| Adjusted odds ratio (95% CrI) | 0.88 (0.67, 1.16) | 1 (Reference) |
| Probability of superiority[c], % | 17.4 | - |
| Probability of inferiority[d], % | 82.6 | - |
| Excluding concomitant antiplatelet agents or randomization to antiplatelet domain of REMAP-CAP[a] | N=424 | N=427 |
| Organ support-free days | | |
| Adjusted odds ratio (95% CrI) | 0.89 (0.69, 1.14) | 1 (Reference) |
| Probability of futility[b], % | 99.1 | - |
| Probability of superiority[c], % | 17 | - |
| Probability of inferiority[d], % | 83 | - |
| Hospital survival | | |
| Adjusted odds ratio (95% CrI) | 0.94 (0.68, 1.29) | 1 (Reference) |
| Probability of superiority[c], % | 35 | - |
| Probability of inferiority[d], % | 65 | - |
| Major bleeding | | |
| Adjusted odds ratio (95% CrI) | 1.46 (0.61, 3.49) | 1 (Reference) |
| Probability of superiority[c], % | 20.1 | - |
| Probability of inferiority[d], % | 79.9 | - |
| Per Protocol Analysis[e] | N=304 | N=341 |
| Organ support-free days | | |
| Median, (IQR) | 7 (-1, 16) | 7 (-1, 16) |
| Adjusted odds ratio (95% CrI) | 0.94 (0.70, 1.25) | 1 (Reference) |
| Probability of futility[b], % | 95.4 | - |
| Probability of superiority[c], % | 32.3 | - |
| Probability of inferiority[d], % | 67.7 | - |
| Hospital survival | | |
| No. of patients/total no. (%) | 68.6 | 64.5 |
| Adjusted odds ratio (95% CrI)[a] | 1.09 (0.75, 1.58) | 1 (Reference) |
| Probability of superiority[c], % | 32.3 | - |
| Probability of inferiority[d], % | 67.7 | - |

| | Therapeutic anticoagulation | Usual care pharmacological thromboprophylaxis |
|---|---|---|
| Major thrombotic events or death | | |
| Adjusted odds ratio (95% CrI) | 0.92 (0.64, 1.36) | 1 (Reference) |
| Probability of superiority[c], % | 65.7 | - |
| Probability of inferiority[d], % | 34.3 | - |

Probabilities represent posterior probabilities of therapeutic anticoagulation compared to thromboprophylaxis
Odds ratios represent the posterior median; IQR – Interquartile Range; CrI – Credible Interval

[a] Sensitivity analyses exclude moderate patient groups from the model

[b] Futility is defined as odds ratio <1.2

[c] For organ support-free days and hospital survival, superiority is defined as odds ratio >1; for major bleeding and major thrombotic events, superiority is defined as odds ratio <1

[d] For organ support-free days and hospital survival, inferiority is defined as odds ratio <1; for major bleeding and major thrombotic events, inferiority is defined as odds ratio >1

[e] Per protocol analyses conducted on REMAP-CAP and ATTACC platform participants only

#### Table S3 – Major Thrombotic Events

| | Therapeutic anticoagulation | Usual care pharmacological thromboprophylaxis |
|---|---|---|
| Number of patients in whom major thrombotic event outcome was available as of January 28th, 2021 | 471 | 476 |
| Number of patients with a major thrombotic event | 27 | 49 |
| Major thrombotic event breakdown – n[a] | | |
| Total Events | 32 | 53 |
| Pulmonary Embolism | 13 | 32 |
| Myocardial Infarction | 6 | 8 |
| Ischemic Cerebrovascular Event | 8 | 9 |
| Systemic Arterial Thromboembolism | 5 | 4 |

[a] Some patients had more than one event; total events are reported here

#### Section 5 – Supplemental Figures

##### Figure S1

Subgroup analyses on the primary endpoint (organ support-free days to day 21) in patients with severe Covid-19 (without borrowing from moderate Covid-19 patient groups). PTP - usual care pharmacological thromboprophylaxis, TAC - therapeutic anticoagulation.

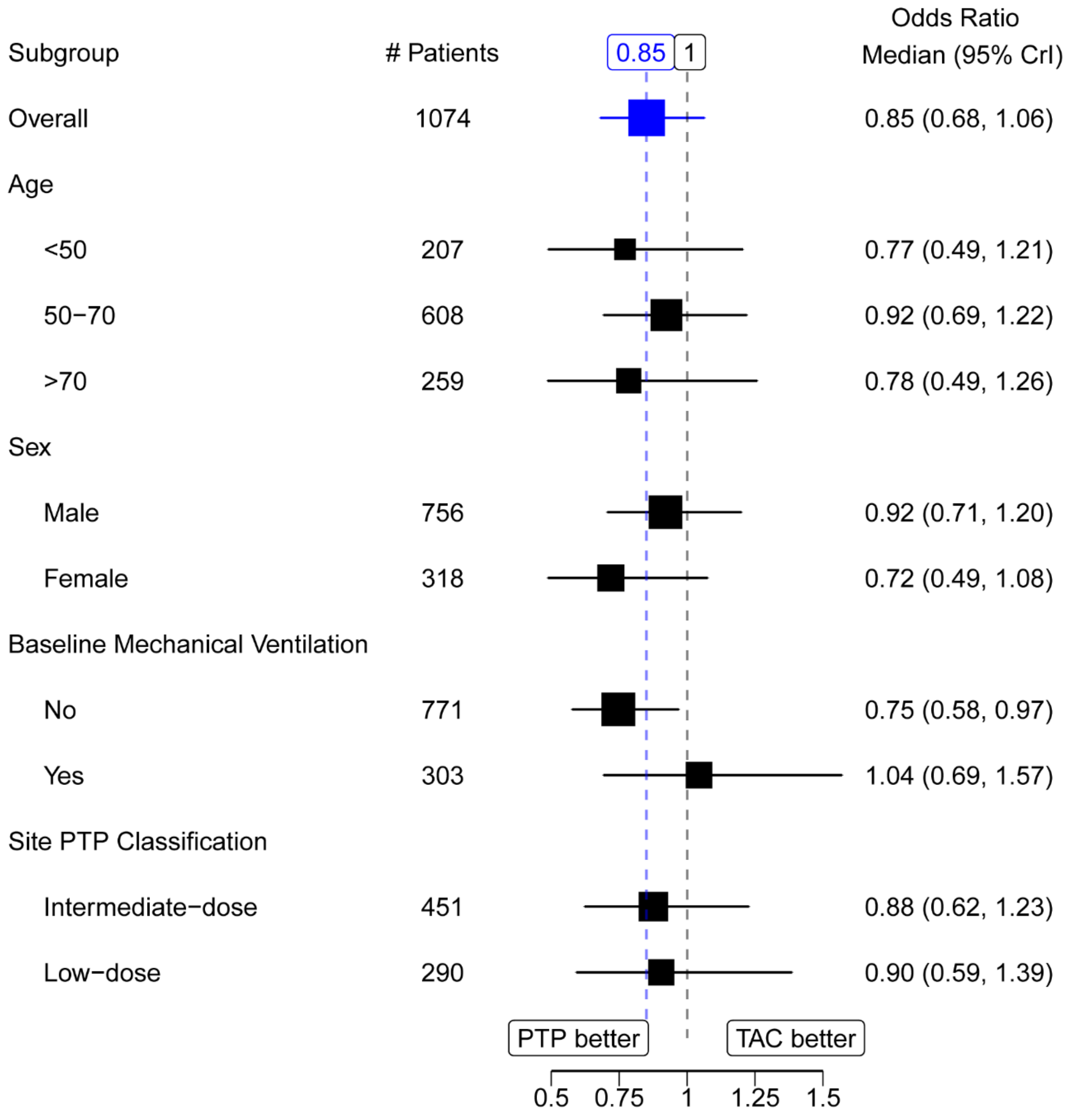
